# Screening for Lung Cancer with Computed Tomography: Systematic Reviews on Effectiveness and Patient Preferences

**DOI:** 10.64898/2026.03.24.26349227

**Authors:** Jennifer Pillay, Samantha Guitard, Sholeh Rahman, Guylène Thériault, Donna Reynolds, Jason E. Pagaduan, Lisa Hartling

## Abstract

**Purpose:** We systematically reviewed the evidence for three questions on screening for lung cancer with computed tomography (CT): benefits (from randomized trials) and harms of screening versus no screening/minimal intervention or alternative screening approaches (e.g., selection criteria, screening intervals); relative importance that informed patients place on the potential benefits and harms of screening (patient preferences); and comparative effects from observational studies of different screening selection criteria (using risk prediction models) or nodule classification systems compared with those used in the screening trials.

**Methods:** A working group of primary care and specialist clinicians (previously members of Canadian Task Force on Preventive Health Care) and topic experts provided input into the eligibility criteria and key potential effect moderators, rated outcomes for their importance to decision making, and developed decision thresholds for use when making conclusions and assessing certainty of the evidence. Critical outcomes of screening effects included all-cause mortality, lung-cancer mortality, and overdiagnosis (via excess cancer incidence from screening). For patient preferences, we sought direct preference data via (i) disutilities of relevant health states (measuring their impact on one’s health-related quality of life on a scale of 0 [perfect health] to 1 [death], mainly using EQ-5D), and (ii) other preference-based data, such as outcome trade-offs, as well as indirect preference data via (iii) the relative importance of benefits versus harms inferred from attitudes, intentions, and behaviors towards screening among eligible patients informed with estimates of the outcomes. For screening benefits and harms and for patient preferences, we searched three databases (MEDLINE, Embase, and Central & MEDLINE, Scopus, and EconLit, respectively) to July 11, 2025. For screening studies published prior to 2015 we relied on searches for other reviews, and for patient preferences our search was limited to 2012-onwards. For comparative effects, we searched MEDLINE and Embase from 2019 to September 23, 2025, with reliance on other reviews for studies published 2012-2018. Reference lists were scanned and trial registries searched. For the main searches, two independent reviewers screened titles and abstracts and then full texts; for search updates we applied AI to assist with title and abstract screening. Data extraction and analysis were undertaken by single reviewers, with verification; risk of bias and GRADE certainty assessments were undertaken independently by at least two reviewers. Data were pooled where suitable using random effects methods appropriate to the outcome metric and prevalence. Subgroup analyses explored heterogeneity (e.g., sex, number of rounds, type of comparator, sensitivity of nodule management, type of utility measurement). When not pooled (e.g., patient preferences based on screening intentions) data were analyzed by grouping studies based on factors such as population, setting, exposure, and outcomes, with consideration of study size and risk of bias. Conclusions and certainty assessments for screening effects were based on estimates of absolute effects.

**Results:** We included 85 studies (N=640,537; 13 trials) on screening benefits and harms, 59 on patient preferences (33 [N=42,219] on disutilities and 26 [N=10,829] other studies), and 16 for comparing trial (National Lung Screening Trial [NLST]) and LungRADs nodule management, either directly (2 studies, N=26,978) or indirectly (14 studies, N=1,102,285). *Screening benefits and harms*: Findings from nine trials (N=94,530) examining low-dose CT (LDCT) screening on all-cause (RR 0.97, 95% CI 0.93 to 1.01; 3.7 fewer [8.5 fewer to 1.2 more] per 1000) and lung-cancer mortality (RR 0.87, 95% CI 0.79 to 0.96; 4.0 fewer [1.2 to 6.4 fewer] per 1000) offered low and moderate certainty, respectively, that screening previous/current 20-30 pack-year smokers 50-74 years old 3-4 times will probably result in at least 1 (all-cause) and 2 (cause-specific) fewer deaths per 1000 screened after 10-12 years. The absolute effects may not apply to participants at the lowest baseline risk for lung-cancer incidence (e.g., <1.5% over 6 years) or death. Seven trials (N=35,161) contributed to meta-analysis for overdiagnosis (RR 1.19, 95% CI 1.03 to 1.37; 8.4 [1.3 to 16.3] per 1000), and our certainty was moderate that LDCT screening 3-4 times will probably result in at least 2.5 cases of overdiagnosis per 1000 screened over 10 years. For important outcomes, we had high certainty that screening 3-4 times results in at least 75 people per 1000 screened (and probably at least 225) having at least one benign biopsy/false positive, 150 having one or more incidental findings (likely at least 450), and 50 (probably at least 100) having a clinically significant/actionable incidental finding, but probably does not have an important impact on major complications or death from invasive testing among those without cancer. Though undergoing a LDCT scan probably causes little-to-no psychosocial harm, having a positive screening result likely causes at least a small degree of harm (i.e., 4-8% change from baseline), especially for the 10-15% having to undergo invasive procedures where some may experience moderate harm. Among those without cancer, these effects may last for several months while the diagnostic process is underway, though moderate certainty evidence found little-to-no effects remaining after 6 months from diagnostic resolution. *Comparative effects*: Findings from applying different baseline predicted risks for lung-cancer incidence or mortality to the trial populations (i.e., alternative selection criteria) were considered with the effects from screening benefits and harms. Using LungRADs instead of NLST nodule management (among NLST eligible people) probably reduces the false positive rate substantially (about half), though the number of false positives still exceeded the decision threshold of 75 per 1000 and the effects for benefits or other harm outcomes are not known. *Patient preferences*: Findings showed little-to-no disutility (i.e., <0.04) from a positive screening test (moderate certainty) or a false positive result (low certainty). Low-certainty evidence found there may be little-to-no disutility from a stage I-IIIA cancer diagnosis (before treatment) but small but important disutilities from a stage IIIB/IV diagnosis, during first-line treatment of any stage (though possibly moderate disutility of about 0.09 for stage IIIB/IV), and after treatment for stage IIIB/IV but not stage I-IIIA (without progression) where effects were inconsistent but indicated that any disutility may not be long-lasting. Findings for stage I-IIIA are likely most relevant for understanding the consequences of overdiagnosis. For stated preferences between outcomes, there was low certainty evidence that a small majority (51-75%) of people may accept 69-122 false positives and at least 1.3 cases of overdiagnosis per prevented lung-cancer death, and think that the reduction in lung-cancer mortality is more important than experiencing one of the relevant harms. After being informed about anticipated benefits and harms from screening (with the largest screening effects shown to those at higher baseline risk), progressively more people preferred screening (mainly via intentions) as the “net benefit” of screening improved from low [25-50% preferred] to moderate [51-75%] to high [>75%].

**Conclusions:** This review provides contemporary data on the benefits and harms of LDCT screening after at least a decade of follow-up and makes conclusions based on absolute effects while considering thresholds for decision making. Across the reviews, findings indicate that screening those aged 50-74 years with 3-4 rounds of LDCT will lead to benefits and harms for which a majority, but not all, eligible people probably find acceptable and worthwhile. While current nodule management using LungRADs likely reduces false positives, whether it impacts the benefits of screening is less certain and worth further research. Further, comparative prospective studies are lacking to determine the effects from screening for those not meeting the minimum age (50 years) and smoking history criteria in the trials, despite having an equivalent risk for lung cancer.

## Introduction

Although lung cancer has been declining over the past two decades in Canada, it is the most common cancer when excluding non-melanoma skin cancers (1). Approximately 1 in 14 men and women in Canada will develop lung cancer in their lifetime (2) with an overall annual incidence of about 66 cases per 100,000 and rates of 12.8, 5.0, 12.0, and 30.5 for stages I, II, III, and IV, respectively (3). Lung cancer is also the leading cause of cancer deaths in Canada. Five-year survival rates across all ages for stages I, II, III, and IV are about 62%, 39%, 16%, and 3%, respectively (4). Lung cancer is one of the most costly cancers in the country, with estimated costs in 2024 to the healthcare system of $5.6 billion and to patients and their caregivers of $1.2 billion (5); approximately $190,000 is spent on health system costs over a lifetime per patient.

A large majority (86%) of lung cancer cases are attributed to modifiable risk factors, especially tobacco smoking (72%) (6). Other major factors include residential radon, asbestos, outdoor air pollution, physical inactivity, and certain occupational exposures such as construction, mining, and transportation (7). Nonmodifiable, independent risk factors include sex, age, a personal or family history of lung cancer, a personal history of lung disease, previous radiation therapy to the chest, and a weakened immune system (3, 8, 9). In Canada, 98% of lung cancers are diagnosed among those aged 50 and older (8% among 50-59 year-olds) and 93% of lung cancer deaths are among those aged 60 and older with about half among those 55-74 years (2). Five-year survival for lung cancer is higher among females compared with males (26% vs. 19%) and decreases with age (e.g., 24-29% for those aged 45-74 years versus 19% and 11% for those aged 75-84 and 85-99 years) (2). Across Canadian provinces, lung cancer incidence (years 2012–2016) and mortality (2013–2017) rates are highest in Nunavut, the Northwest Territories, Quebec (mortality data only), and to a lesser extent, the Atlantic provinces (3). Through systemic, economic, and geographic barriers, people living in rural or remote communities and/or having a lower income experience inequities in lung cancer incidence (e.g., 6% more diagnoses at later stages), access to care (e.g., 26% fewer curative surgeries), and outcomes (e.g., 13–25% lower survival at 3 years, depending on stage) (10). Disparities in lung cancer incidence (e.g., 20–40% higher in First Nations people in Ontario), mortality, and risk factors exist for Indigenous peoples in Canada with many contributing factors among a variety of social determinants of health (11–13). For example, First Nations, Inuit, and Métis populations have lower rates of cancer screening than non-Indigenous people in Canada (14). Few studies exist describing inequities in lung cancer risk and prognosis for other racial or ethnic groups in Canada (15). One cohort using 2006 census data found lower rates of diagnosis among most visible minority groups versus non-visible minorities (e.g., odds ratios for Black and South Asians 0.45 and 0.39 [p<0.0001], respectively, with exception for Chinese [odds ratio 1.0]), and similar rates among groups for diagnostic or therapeutic procedures in individuals with a lung cancer diagnosis (16).

A majority of lung cancers are diagnosed at a late stage because early cancers are often asymptomatic (3). Screening aims to detect earlier lung cancers when treatment is more likely to be curative. Low-dose computed tomography (LDCT) is the most common screening modality used in current clinical practice, largely based on results in 2011 of the National Lung Cancer Screening Trial (NLST) showing a 20% reduction in lung-cancer mortality (absolute risk reduction 3.12 per 1000; number needed to screen 320) at a median 6.5 years follow-up from three annual screens with LDCT compared to chest X-ray among current or former smokers aged 55 to 74 years with at least a 30-pack year smoking history within the past 15 years (17). However, the net benefit of screening may differ after longer-term follow-up (where baseline risks are higher) and it is unclear what magnitude of benefits and harms are important to patients. The net benefit may also vary by participant selection and nodule management used during screening.

Various models exist to predict lung cancer incidence or prognosis. Using data from current and former smokers aged 55–74 years in two large screening trials (Prostate, Lung, Colorectal, and Ovarian [PLCO] Cancer Screening Trial [studying screening with chest X-ray] and the NLST), the PLCO_M2012_ model is widely used and predicts cancer incidence over 6 years if unscreened (18). This multivariable model has shown to have higher sensitivity to predict lung cancer than the eligibility criteria used in the NLST, and may have the potential to reduce disparities for those (e.g., Black people) at high risk despite age or smoking history (19). Predicted risk may also help determine which patients benefit the most from screening. Performance characteristics of the nodule classification system will affect the number of benign positive tests (considered “false-positives” [FPs] for this review) and missed cancers, which may impact early detection while avoiding possible psychosocial harms from a FP or physical harms from investigations for benign lesions. The NLST trial for example, used a nodule management system where test positivity was based on a fairly small nodule size (4 mm and only accounting for growth at the 3rd screening round). This resulted in a considerably high (about 34%) number of participants having one or more FPs across the three screening rounds. Other randomized controlled trials (RCTs) have used different approaches to classify nodules (e.g., larger nodule size and accounting for growth in all subsequent screens) and since the initiation of existing RCTs, the American College of Radiology developed the Lung-RADS™ classification system in attempt to standardize LDCT screening (20). LungRADs is currently being used/considered by most provinces (except British Columbia (21)) with existing or proposed screening programs (22). Examination of evidence comparing these methods in terms of patient-important outcomes is warranted to help determine the ideal selection criteria and nodule management for screening.

Preferences for or against screening are influenced by the relative importance patients place on the expected or experienced outcomes (22–25). Preference data can be elicited directly through comparing the disutilities of different health states, measured on a scale of 0 (no disutility) to 1 (similar to death) and a value of about 0.04 considered important among the Canadian public (26). A disutility measures the impact of the relevant outcome on one’s health-related quality of life (HRQoL) and can be measured using generic multi-attribute utility instruments such as the EQ-5D or via direct choice-based utility elicitation methods such as standard gamble (SG) or time tradeoff (TTO) (i.e., determining what people would be willing to risk or give up to avoid living in that health state). Other preference-based data, such as trade-offs/ratings/rankings between outcomes, also capture preferences. Indirectly, the relative importance of benefits versus harms (i.e., the net benefit) of screening can be inferred from attitudes, intentions, and behaviors towards screening among informed patients provided with estimates of the magnitudes of benefit(s) and harm(s). Evidence on how patients weigh the relevant outcomes is important to inform decision makers when considering the balance of benefits and harms and determining whether this balance might vary across different individuals (27).

### Purpose and scope of review

In 2016, focusing on short-term results of the NLST the Canadian Task Force on Preventive Health Care (CTFPHC) recommended screening adults 55–74 years of age who have at least a 30-pack-year smoking history and who smoked or quit smoking less than 15 years ago, annually for 3 years with LDCT (weak recommendation, low-quality evidence) (28). Other existing guidance in Canada is also outdated (29, 30). Since the CTFPHC guidance, extended follow-up from the NLST (31) and at least one large RCT (NELSON (32)) have been published. Further, a synthesis of evidence related to the use of selection criteria and nodule management practices that are either currently being used or under consideration for use in Canada was warranted. We updated the previous review conducted for the CTFPHC (33) on the benefits and harms of screening with LDCT with various modifications. Screening with chest X-ray was not a focus for this update. We also conducted a de novo review comparing different selection criteria and nodule management approaches with those used in the existing RCTs, as well as a review on patient preferences. Three key questions (KQs) were answered.

### Key questions

1. What are the benefits and harms of screening for lung cancer in adults aged 18 years and older?
2. What is the relative importance people place on the potential benefits and harms of screening for lung cancer?
3. a) What are the comparative benefits and harms of risk prediction models compared with trial-based criteria to identify eligibility for lung cancer screening? b) What are the comparative benefits and harms of alternate nodule classification systems compared with nodule classification systems used in lung cancer screening trials?

## Methods

We followed methods outlined in the CTFPHC manual (34). The protocol was previously published (35) and any deviations are reported herein. This review is reported in accordance with current standards (36). A working group consisting of CTFPHC members and clinical experts (see acknowledgements) contributed to the development of the KQs and PICOTS (population, intervention(s) or exposure(s), comparator(s), outcomes, timing [of outcome measurement], setting, and study design) elements. During the protocol stage, the working group also chose the outcomes that were considered critical (rated at 7 or above on a 9-point scale) or important (rated 4-6) for clinical decision-making, according to methods of Grading of Recommendations Assessment, Development, and Evaluation (GRADE) (37). Ratings by the clinical experts were solicited to ensure acceptable alignment with the views of CTFPHC working group members, who determined the final ratings. During the conduct of the review but while blinded to results and key study identifiers including trial name, country and sample size, the working group determined decision thresholds to use when interpreting the findings (38, 39), provided input to refine some of the outcomes, and informed refinement to the analytic plan especially around defining the potentially moderating variables that may account for varying results for different populations and interventions. They were not involved in the selection of studies, extraction of data, appraisal of the risk of bias, synthesis of data, or assessment of the certainty of evidence. Two members of the working group (GT, DR) also contributed to the writing of this paper and met authorship criteria. Since starting these reviews conducted, the CTFPHC’s mandate has concluded and the Public Health Agency of Canada has initiated a renewed task force that may or may not issue recommendations on this topic or use these finding for their recommendations. Nonetheless, the findings remain important for use by clinicians and other interest holders in Canada and elsewhere.

### Eligibility criteria

Supplemental tables S1.1, S1.2, and S1.3 outline the eligibility criteria for each KQ (Additional file 1). For KQ1, we included studies of adults in whom lung cancer was not suspected, among a general adult population or people meeting eligibility criteria associated with increased risk for lung cancer, as defined by authors. In revised criteria, we excluded studies that only enrolled select populations, such as those with HIV or other cancers. The critical outcomes were lung-cancer mortality, all-cause mortality (both benefit outcomes), and overdiagnosis (harm outcome). Important outcomes were HRQoL (benefit outcome), and the harms of FPs, incidental findings (any as well as only clinically significant/actionable), major complications and death from invasive diagnostic procedures undertaken as a result of screening (primarily among those without cancer), and psychosocial harms from the screening process. The final classification of benefit or harm for all outcomes was based on the effects observed. RCTs comparing CT with no screening or an alternative screening modality (e.g., chest X-ray) or strategy (e.g., pre-screening with biomarkers, different eligibility criteria, classification of findings, screening interval) were eligible. Apart from overdiagnosis where trial data with a no screening arm is most valid and was known to be available (40), for harms from screening we also included nonrandomized studies and uncontrolled studies using the same diagnostic procedure (i.e., nodule classification) as the RCTs. We examined the evidence on harms for a particular modality of screening when there was at least low certainty of some benefit for one or more benefit outcomes from RCTs.

For KQ2 on values and preferences, individuals may or may not have experienced screening, lung cancer or one or more of the critical or important outcomes (i.e., heath states). For participants without experience of a relevant health state (e.g., those deciding whether to screen), we required that a large majority received and reviewed structured information with numerical data about the potential benefits and harms from screening with LDCT. Study designs were any quantitative design measuring preferences between different outcomes either directly using health-state utility values (HSUVs) or trade-offs/ratings/rankings, or indirectly, allowing inferences about relative importance between benefits and harms based on attitudes/intentions/uptake of screening. Health states of interest for capturing HSUVs included those related to screening (e.g., undergoing the screening test, getting positive results, receiving a FP result) as well as after diagnosis and during and after first-line treatment (by cancer stage). The disutilities for diagnosis and treatment for lung cancer (particularly stage I-IIIA) were provided to help understand the impacts of an overdiagnosed cancer. Measures of values and preferences related to the critical and important outcomes were based on the GRADE methodology (24).

For KQ3, we included nonrandomized controlled studies (including modelling studies) comparing the benefits and harms between RCT (included in KQ1) (KQ3a) screening selection criteria or (KQ3b) nodule classification systems and different (KQ3a) selection based on (externally validated) risk prediction models or (KQ3b) nodule classification systems. RCTs of these comparisons would have been captured in KQ1. Because of sparse eligible data, especially for the LungRADs nodule classification system, we revised our eligibility to allow for indirect comparisons between relevant findings (e.g., FPs when using NLST nodule management) in studies from KQ1 and studies reporting on use of LungRADs in an uncontrolled manner, as long as the eligibility criteria were similar (i.e., within 2-3 years of age) to the NLST or NELSON trials.

For most relevance to Canada, for all KQs we only included studies conducted in countries listed as very high according to the Human Development Index (41) and having full texts in English or French. For KQs 2 and 3, we limited studies to those published during or after 2012. Based on clinical input and working group discussion, utility-based outcomes will have changed over time because treatments and their impact have changed quite dramatically, and the best indirect measurements, for example, based on decision aids, would be based on contemporary estimates of the effect of CT screening since the publication of NLST trial in 2011. This date also represents the publication of the major risk prediction models and the emergence of studies comparing different screening protocols.

### Searching the literature

For KQ1 we located all full texts from the previous CTFPHC review (33). The previous review’s final search date for studies was March 31, 2015 (33), so for this KQ we searched for studies published from 2015 onwards (first on October 11, 2022 and then updated on July 11, 2025) in MEDLINE and Embase, via Ovid, and Cochrane Central including the Central Register for Controlled Trials. The previous searches for benefits and harms were modified slightly to increase their sensitivity, with a more sensitive filter applied for RCTs, and broaden their scope, such as adding controlled vocabulary and keywords for incidental findings and psychosocial harms into the harms concept. For KQ2 we searched in April 2023 (and updated on July 11, 2025) Ovid MEDLINE, Scopus, and EconLit from 2012 using two searches: one for utility-based studies, focusing on relevant preference-based instrument/methodology terms and the relevant outcomes, and another for decision-making/acceptance/attitudes about lung-cancer screening.

For KQ3, in December 2022 we searched MEDLINE and Embase. For MEDLINE we relied on the 2019 search performed by the authors of a review on this question to inform the United States Preventive Services Task Force (USPSTF) (and screened all of their included studies for eligibility) (42) and updated the search using a de novo search strategy; for Embase we ran the de novo search from 2012. For our update on September 23, 2025, we restricted the KQ3 search to studies focusing on selection criteria using the PLCO risk prediction model and nodule classification using LungRADs or the Pan-Canadian Early Detection of Lung Cancer model to narrow the scope to that most applicable to Canada. Searches were developed in collaboration with an information specialist and peer-reviewed by another using the PRESS 2015 checklist (43). The final searches are located in Additional file 1. We scanned reference lists of included studies and relevant reviews. We searched ClinicalTrials.gov and the World Health Organization International Clinical Trials Registry Platform for results data for published and unpublished trials (past 2 years).

We exported the results of database searches to an EndNote library (Clarivate Analytics, Philadelphia, USA, 2018) for record-keeping and removed duplicates. We documented our supplementary search process, for any study not originating from the database searches, and entered these citations into EndNote individually.

### Selecting studies

Records retrieved from the database searches were uploaded to DistillerSR (Evidence Partners Inc., Ottawa, Canada) for screening. For our initial searches, titles and abstracts of all citations retrieved from the database searches were screened by two reviewers independently. For title/abstract screening of our search updates in 2025, DistillerAI was used to continually re-prioritize records based on screening decisions amongst two reviewers and after at least 50% of the citations were screened, and DistillerAI estimated that ≥95% of included studies were found, we switched to a single reviewer. Full texts of any citation from the searches considered potentially relevant by either reviewer were retrieved. In all instances, two reviewers independently reviewed all full texts including the studies from the previous reviews against a structured eligibility form, and a consensus process was used for any full text not included by both reviewers. Where necessary, a third reviewer and/or author contact was used to arbitrate decisions. The screening and full-text forms were pilot-tested with a sample of 100 abstracts and 20 full texts, respectively. Screening additional studies located from reference lists, trial registries, and websites were conducted by one experienced reviewer, with two reviewers reviewing full texts. We documented the flow of records through the selection process, with reasons provided for all full-text exclusions, and present these in flow diagrams (36) (Figures S2.1, S3.1, and S4.1) and the appended excluded studies list.

### Data extraction

One reviewer extracted data and another verified all data for accuracy and completeness, except for results data for the critical outcomes in KQ1 which were extracted in duplicate, with decisions based on consensus or arbitration by a third reviewer. Each data extraction form was piloted with a sample of at least five studies. Sufficient data was collected to allow for description and analyses on specific populations and intervention/exposure characteristics of interest and for assessment of risk of bias and reporting/missing outcome biases.

For most of the outcomes in KQ1, the denominators used were the population enrolled in the relevant arm/group(s) in the study (i.e., intention-to-screen). One exception was for psychosocial harms where sub-populations (e.g., those receiving a positive screening result) were considered. Another exception was for overdiagnosis. We calculated estimates of overdiagnosis by the relative (40) and absolute (excess) risk of cumulative lung-cancer incidence through follow-up in the screening compared with no screening group, and by the excess risk of cancer from screening among those (i) having cancer diagnosed in the screening arm, and (ii) having cancer diagnosed through screening in the screening arm.

For mortality outcomes and overdiagnosis, per protocol we used crude data on the cumulative number of events from the longest follow-up time point unless there was substantial contamination after a previous time point (>20% in the no screening group receiving screening). Because of poor reporting on contamination, we revised this to be based on overall risk of bias for the outcome (i.e., used longest follow-up unless a shorter follow-up had lower risk of bias). For incidence (cumulative detection from baseline) rates, used when calculating overdiagnosis, we required follow-up beyond the active phase of screening and a control group receiving no screening (vs. chest X-ray). For HRQoL, we extracted the mean baseline and endpoint or change scores (at longest follow-up without substantial contamination), standard deviations (SDs), or other measures of variability, and the number analyzed in each group.

All other outcomes (except in some cases for psychosocial harms) are reported only for those receiving LDCT screening; very few, if any, studies reported on these outcomes among the no screening arms, and risks among those receiving chest X-ray were not of interest. Our protocol stated that for binary events that could occur more than once we would use a hierarchy, prioritizing proportions (i.e., the proportion of individuals having one or more events) over events (i.e., the number of events across individuals). For more meaningful interpretation and because many studies reported both results, we revised our methods to keep these outcomes separate but considered the proportion data for our main conclusions. Further, because most of the non-randomized studies only reported on results of one round of screening, we used data after one round in addition to that across rounds from the RCTs.

For the outcome of major complications (defined through clinical input rather than as reported by study authors) from invasive testing as a result of screening, we captured events among those who later received a negative diagnosis (FPs) as well as for anyone receiving invasive testing (i.e., those with lung cancer and FPs). For incidental findings, we captured all incidental findings and clinically significant incidental findings, as well as several specific incidental findings of interest to the working group. Apart from specific incidental findings, studies were required to report findings in at least two different body systems to reduce the possibility of selective outcome reporting.

For FPs and psychosocial harms from screening, we examined results for i) anyone receiving a recommendation for early recall (e.g., indeterminate result with repeat CT screening at 3 or 6 months) or for a diagnostic follow-up (e.g., result suspicious of lung cancer) as well as ii) only those recommended to have diagnostic follow-up. Because screening results can take many months to resolve, even though we required at least 6 months’ follow-up after screening we made sure to record the proportion of individuals with a positive screening test that had diagnostic resolution, for consideration in our risk of bias assessment. For psychosocial harms, we extracted the mean baseline and endpoint or change scores, standard deviations (SDs) or other measures of variability, and the number analyzed (by group if a control group was used). Results considered consistent with the outcome of psychosocial harms included data from patient-reported outcome measurements of symptoms of anxiety, depression, distress, and concern about lung cancer. In cases where composite scores meeting these concepts were available, we used these and not subscales. Single-question items were not eligible. Subscales of overall HRQoL scales (e.g., mental health) were considered to measure psychosocial harm if other tools measuring the same symptoms were not reported. For this outcome we extracted data at all reported time points during the active phase of screening to primarily capture these harms from undertaking screening itself and from receiving a FP result. For data from non-randomized studies (including subgroup analysis from RCTs), we relied, when possible, on results adjusted for potential confounders.

For KQ2, studies reporting on HSUVs, data using the most commonly used measurement tool (e.g., EQ-5D), using tariffs (valuation) from the same country was prioritized in studies reporting on more than one measure. For cancer-related health states, rather than just before and after treatment as per protocol, we refined this extraction to three time points: a) at diagnosis, or before treatment initiation (treatment naïve); b) on first-line treatment; c) after first-line treatment completion and/or on maintenance therapy.

For studies reporting on other data related to patient preferences (e.g., outcome ratings, intentions to screen), we only extracted results relevant to our review. For example, when studies compared screening intentions among those receiving a decision aid or other materials (meeting our criteria with numerical estimates provided for screening outcomes) versus usual care or material not meeting our criteria, we only extracted data from the eligible arm. While not an outcome for this review, we extracted results from knowledge tests given to participants to help assess risk of bias concerns related to understanding of the information provided. We also focused on results among those eligible for screening, when those ineligible (with awareness of this) were also participating in the study.

For missing results data for any outcome, including additional information on interventions, exposures (e.g., contents of decision aids in KQ2 studies), or measures of variance, we contacted authors by email with two reminders over 1 month. If variance measures were not reported or obtained from authors, we computed missing SDs or standard errors (SEs) from other study data, or as a last resort, imputed based on other studies in the review. When computing SDs for change from baseline values, we assumed a correlation of 0.5 (44), unless other information was present in the study that allowed us to compute it more precisely. We used available software (i.e., Plot Digitizer, http://plotdigitizer.sourceforge.net/) to estimate effects from figures where no numerical values were provided.

### Risk of bias

We assessed the risk of bias by outcome, including its timing of assessment. For RCTs we used the Cochrane risk of bias (ROB 2.0) tool, assessing the effect of assignment to the intervention for each relevant outcome (cancer-specific and all-cause mortality, cancer incidence [to calculate overdiagnosis] and HRQoL) (45). For nonrandomized studies (including single-arm data on harms from RCTs) in KQ1 and 3, we used a checklist from JBI (46). For KQ2, we used items as per GRADE guidance, about the choice/selection of representative participants; appropriate administration and choice of instrument; instrument-described health-state presentation (for HSUVs among those without experience of health state) or valid descriptions of screening outcomes in information provided to those eligible for screening (for other preference studies); patient understanding; and valid analysis and reporting of results (e.g., lack of variance measures) (23). The specific questions and our rules for determining overall risk of bias for KQ2 are presented in Additional file 3.

Two reviewers independently assessed the studies and came to a consensus on the final risk of bias assessment for each question using a third reviewer where necessary. Each risk of bias tool was piloted with a sample of at least five studies, using multiple rounds until agreement on all elements was high. Regardless of the tool, we used the language of “low”, “some concerns”, and “high” to rate individual domains and overall risk of bias for the related outcome. These assessments were incorporated into our assessment of the risk of bias across studies when rating the certainty of the evidence for each outcome using GRADE.

### Data analysis for screening benefits and harms (KQ1)

When two or more outcome comparisons were sufficiently similar, we pooled their data. The decision to pool studies was not based solely on statistical heterogeneity; the *I*^2^ statistic was reported, but it is recognized that the *I*^2^ is influenced by the number of studies and the magnitude and direction of effects(47). Rather, we relied on interpretations of the clinical and methodological differences between studies. For meta-analysis, if there were large differences in trial sizes and potentially small-study bias (e.g., missing small studies with null effects) or within-study bias in smaller studies (48), our main analyses would employ a fixed-effects model, but if these factors were not apparent we planned to use a random-effects model. We used the DerSimonian Laird method when events were not rare (>1%), which applied for all results using comparative data. For dichotomous outcomes, we analyzed and reported data using risk ratios (RRs) and their 95% confidence intervals (95% CIs). Proportions were pooled using the Freeman-Tukey double-arc sine transformation method; exact confidence interval adjustment was used in analyses with rare or zero events (49).

Meta-analysis was not undertaken for HRQoL (only reported in two RCTs) or psychosocial harms data due to variation in measurement scales and differences in design (between-arms and before-after data) and timepoints. We transformed all data into a standardized measure by calculating the percentage difference or change (e.g., difference between arms of 2.0 on a scale with a range of 0-20 would be 10%). For psychosocial harms data, we grouped data based on which screening outcome would be represented by timing and populations. We then compared and contrasted findings based on the risk of bias, measurement tool, and sample characteristics. Within the included studies there were few reported thresholds for a minimally important difference, and rationale was only applicable to the specific tool. Based on clinical input, we interpreted a difference of 5% to <10% to be a small difference and of 10% to 20% to be a moderate difference. For each analysis we used our best estimate of whether the data across studies would meet these thresholds.

Analyses were performed using Microsoft Excel, Review Manager (version 5.3), and STATA (version 17.0). Relative effects were transformed into absolute effects. When meta-analysis was performed, we used the relative effects together with an unweighted average of control events rates across the included studies for calculating absolute effects (50).

#### Unit of analysis issues

There were no cluster-randomized trials, therefore we did not have to account for unit-of-analysis errors when reporting their findings and/or incorporating them into meta-analysis.

#### Assessment of heterogeneity

Sensitivity analysis was performed removing the high risk of bias studies. We conducted subgroup (stratified) analyses, using variables associated with the population, the intervention, and (for harms) the source data (i.e., data from RCTs vs. nonrandomized studies). The specific categorization of each variable was decided after charting out study characteristics, but before results were extracted. For the population, we compared findings based on control event rates for cancer incidence across the studies (above and below 2% and 3% 6-year risk of cancer, assuming rates in a control group would be similar across years when making any calculations). For interventions we used the following variables to create strata: i) comparator (no screening vs. chest X-ray), ii) nodule classification (highly sensitive [>4 mm nodule and no growth accounted for until third round] vs. not highly sensitive [others which had quite similar systems]), iii) number of rounds (1 vs. 2-3 vs. 4-5), and iv) screening interval (only one round vs. annually vs. less than annually). We also extracted results from within-study analyses related to our specified variables of interest, and we attempted to pool this data where possible. For sex where there were multiple studies reporting findings by sex as well as at least one study only enrolling men, we pooled the within-study findings using a ratio of relative risks (51) and also compared findings among males versus females across studies.

Subgroups were tested for statistical significance, but credibility of the results was interpreted based on available guidance (52) and considering absolute effects in comparison with our decision thresholds for each outcome based on GRADE guidance on inconsistency (53) (see Rating certainty of the evidence). Ideally, findings would be consistent across multiple studies. We prioritized within-study subgroup findings because of their lower propensity to be influenced by other confounding factors (e.g., differences between studies by other variables). Additionally, subgroup findings for participant characteristics accounting simultaneously for multiple risk factors (e.g., for calculating 5-year lung cancer mortality risk) were preferred over those related to single factors (e.g., age or smoking duration).

#### Reporting bias

When meta-analyses of trials contained at least 10 studies of varying size for comparative outcomes, we planned to test for small study bias (54, 55). We also explored missing outcome bias (i.e., data missing from an analysis due to selective non-reporting by other studies), especially harms data that came from observational studies without a protocol outlining pre-planned outcome measurement. For major complications and deaths from invasive testing, we examined whether at least half of the total sample size of all studies reporting FPs also reported these outcomes, as those attending diagnostic work-up after a positive scan would incur these complications at this time or within a short window of time afterwards.

### Data analysis for patient preferences (KQ2)

When pooling mean HSUVs (or disutilities) we used a random-effects model with weighting by the inverse of variance. Some studies provided data for the disutility of one or more relevant health states, comparing (subtracting) the utility of the health state from that of an “unaffected population” (e.g., eligible but unscreened). To estimate disutilities for other studies without unaffected population values, we took an unweighted average of the utility values for the unaffected populations reported in the other included studies and subtracted from this the pooled utility value of the health state of interest (i.e., a between-study comparison). Early stage (stages I-IIIA) and advanced stage (stages IIIB-IV) lung cancers were analyzed as separate populations due to differences in management and prognosis (56). Based on clinical input and results on minimally important disutility in Canada (26), for each health state a disutility of 0.04 to <0.08 was considered small-but-important and ≥0.08 was considered moderate. Where we were not able to use a study’s data in a meta-analysis (e.g., only *p* value and direction reported), we considered these findings and compared them with the results of the meta-analysis. Sensitivity analysis was performed removing the high risk of bias studies, measurement tools other than the EQ-5D, or studies for which we needed to impute measures of variance. Analyses were performed using Microsoft Excel and Review Manager (version 5.3).

For studies not measuring HSUVs in KQ2, we separated data into groupings for analysis. We first categorized the study outcomes based on whether they directly measured the relative importance of screening outcomes of interest to the working group (e.g., utility scores from conjoint analysis, trade-offs, or ranking/ratings focusing on the outcomes) (i.e., “direct, preference-based data”) or indirectly measured the relative importance of the overall benefits versus harms of screening, via attitudes, intentions and screening uptake after being provided with information of the estimated effects. This indirectness relates to two main issues: (i) the information provided, often in decision aids, did not usually only focus of the outcomes of interest to the working group (e.g., also presented data on false negatives, radiation exposure); and (ii) the main outcomes of intentions and uptake are influenced not only by the importance of the outcomes (with the presented risk of each) from screening but also by additional factors such as provider recommendations, perceived cancer risk, accessibility of screening etcetera. We then mapped out the study participant and exposure characteristics to determine meaningful additional groupings. For study participants, we considered how applicable the study sample was to the main target population for the guideline, that is, people accessing primary care who are eligible for and currently considering whether or not to screen. For the exposures, we grouped direct data by which screening outcomes were being compared, and for indirect data we compared the information provided about the effects of screening to make categories based on the relative “net benefit” of screening portrayed across the studies (regardless of whether we judged the effects to be valid or not). Our synthesis further considered the risk of bias of each data point, and to some extent the size of the study since very small samples (n≤30) were assumed to have less validity than larger studies. We would have considered meta-analysis if a large majority of studies in each grouping had similarity in their populations, exposures and outcome measures.

For synthesis we created four categories of study findings, using quartiles, to reflect a large majority preferring (i.e., >75%) (or a small minority [<25%] not preferring) or a small majority preferring (51-75%) (or a large minority [25-50%] not preferring) screening (or one outcome preferred over another within a direct comparison). This classification allowed us to use data from studies that did not use binary (yes/no) outcome measurement, for example when a scale was used (e.g., mean score 7 ± 2 on a 11-point scale on intentions) and we could make a good estimate of the category of results (e.g., 50-75% had intentions) even though an exact percentage of participants could not be calculated. We could also perform some sensitivity analysis when studies allowed for answers of “uncertain” intentions, where we made an assumption that about 50% of these respondents would state a preference for screening if forced. Further, for the direct data, when possible, we converted results into trade-offs even if these were not directly elicited in the study. For example, if the results indicated a large majority (>75%) had positive attitudes about screening when provided with estimates of 3 fewer lung-cancer deaths and 300 FPs, we concluded that a large majority would accept *at least* 100 FPs per prevented death. However, most studies did not directly elicit trade-offs or examine a range of estimates for the outcomes being compared so the acceptable upper limit of the harm (e.g., FPs) is not known. Findings were presented in harvest plots.

Analyses were performed using Microsoft Excel and R Statistical Software (v4.5.0; R Core Team 2025).

### Data analysis for comparative effects of selection criteria and nodule management (KQ3)

For KQ3a, studies that compared selection criteria in the trials to that using various risk prediction models were analyzed as subgroup data in KQ1. These studies focused on retrospective analysis of NLST data. Meta-analysis was not performed as all included studies re-analyzed outcomes from the NLST trial by applying different risk prediction models to its participants. No modelling studies were included as none met our eligibility criteria.

For KQ3b, there were no studies identified with prospective comparison of trial and different nodule management protocols among people meeting the eligibility for the large trials in KQ1 (NLST and NELSON). There were only retrospective designs that recategorized nodules, using NLST nodule management, according to LungRADs criteria. Due to this limited number of within-study comparisons, we also made an indirect comparison between weighted averages of uncontrolled data using LungRADs versus NLST management. Analyses were performed using Microsoft Excel and Review Manager (version 5.3).

### Rating the certainty of the evidence

We used GRADE methods to assess the certainty of evidence for each main outcome (23, 25, 53, 57, 58). We did not assess the certainty for harms data using events/rates, proportions of FPs only requiring diagnostic follow-up, specific incidental findings, or measures of overdiagnosis apart from excess incidence. For harms data reported by first and multiple rounds, we used all data to rate our certainty about effects over 3-4 rounds of screening. Two reviewers independently assessed the certainty of evidence for each outcome and agreed on the final assessments. A third reviewer arbitrated if necessary.

We assessed the certainty of evidence (very low, low, moderate, or high) based on five main domains (study limitations/risk of bias, inconsistency, indirectness, imprecision, and reporting biases [small study bias and missing outcome data]). Within each, we rated down once when we had serious and twice for very serious concerns. All outcomes started at high certainty, including harms from uncontrolled studies because the outcomes were mostly attributed to the screening intervention (59). Psychosocial outcomes, which may not be attributed to screening, also started at high certainty as most data came from comparisons in RCTs though we rated down for risk of bias when needing to rely mostly on nonrandomized comparisons. Studies measuring HSUVs and other preferences also started at high certainty since the ideal study design for these outcomes was used.

We applied an approach whereby the certainty for each outcome was rated based on whether the effect would be large enough to exceed one or more decision thresholds. These decision thresholds were determined by consensus among the working group before they saw the findings, based on a survey approach (38, 39) where they agreed on a magnitude of effect judged to reflect where a majority (>50%) of informed patients would think the effects were at least small but important (all among 1000 screened and considering 3-4 rounds of screening and about 10 years follow-up): lung-cancer mortality (1.0 fewer [or more if harmful]), all-cause mortality (0.5 and 1.0 fewer/more), overdiagnosis (2.5), FPs (75), incidental findings (any finding: 100 and 150; clinically significant: 50 and 100), and major complications (2.5) or deaths (among FPs) (0.1) from invasive diagnostic procedures. Thresholds for patient-reported outcomes were also created though did not employ this survey approach (see Data analysis for KQ1).

Because some of the point estimates differenced substantially from the thresholds and guideline panels/decision makers are required to make judgements about the magnitude of effects when applying GRADE guidance, in a *post hoc* manner we identified magnitudes of effect larger than the decision thresholds (or smaller for harms outcomes) for which we had moderate- or high-certainty evidence. The findings of certainty should be interpreted in relation to these thresholds rather than the magnitude of the pooled estimates, for which we would usually have less certainty.

We present tables with our summary of findings, by outcome for each comparison, and provide explanations for all decisions. We adopted standard GRADE wording to describe our findings, using the word “may” together with the direction of effect to describe findings of low certainty, “probably/likely” for those of moderate certainty, and without any modifier for high certainty (60). When our certainty in the evidence was very low, we describe the evidence only as uncertain without any associated direction or data.

## Results

### Key question 1

#### Study characteristics

For this key question we included 85 studies (31, 32, 61–143) with 43 associated reports (17, 18, 144–188) (Table S2.1 and 2.2 in Additional file 2). There were 11 RCTs reporting benefit outcomes (nine also used for harms), and two additional RCTs and 72 observational studies used for harms outcome data. The mean age across studies was 61.4 years (range 49.5 to 67.7) and the total number of participants was 640,537 (range 72 to 152,918). The average proportion of male participants was 63.0% (range 41.6% to 100%), and among the third of studies reporting on race or ethnicity there was an average of 79.0% Caucasian participants. The largest trial (NLST (31)) enrolled 91% Caucasian, 4.4% Black, and 4.8% “other” individuals. Of the included studies, 39 came from the United States (31, 66, 68, 69, 74–76, 78–82, 84–86, 89, 92, 93, 99, 102, 103, 105–107, 110, 118–124, 127, 129, 136, 137, 139, 142, 143), nine from the United Kingdom (63, 70, 73, 97, 101, 125, 131, 133, 135, 187, 189), seven from Italy (62, 83, 113, 115–117, 138), seven from South Korea (87, 88, 94, 95, 114, 128, 130), five from Canada (72, 77, 90, 104, 134), three from Poland (109, 112, 126), three from multiple countries (32, 65, 132), two from Australia (100, 108), two from France (64, 96), and one study each from Denmark (141), Germany (98), Spain (67), Sweden (61), Finland (140), Japan (111), Estonia (91), and Lithuania (71). More detailed characteristics and findings are divided by screening modality (LDCT alone [11 RCTs and all nonrandomized studies] vs. targeted screening with LDCT after positive results on another screening test [2 RCTs]) and outcome.

#### Low-dose chest tomography screening strategies: critical outcomes

For lung-cancer mortality, all-cause mortality and overdiagnosis (Table 1), there were nine RCTs that offered LDCT to all participants randomized to the intervention arm (31, 32, 73, 74, 83, 98, 113, 116, 141). The mean age across studies was 61 years, all participants were current or former smokers (with the exception of one trial that included <1% non-smoking participants that met other lung cancer risk criteria (141)), and the total number of participants randomized was 94,530 (range 2,450 to 53,454). Seven trials reported all outcomes across both sexes (31, 73, 74, 98, 113, 116, 141), with an average 63.3% male, one trial enrolled exclusively male participants (83), and one trial (NELSON) reported on both male and female participants for mortality outcomes (83.6% male), but only males for overdiagnosis (32). Race or ethnicity were only reported in two trials (NLST and UKLS), each enrolling over 90% Caucasian participants (31, 73).

**Table 1.**
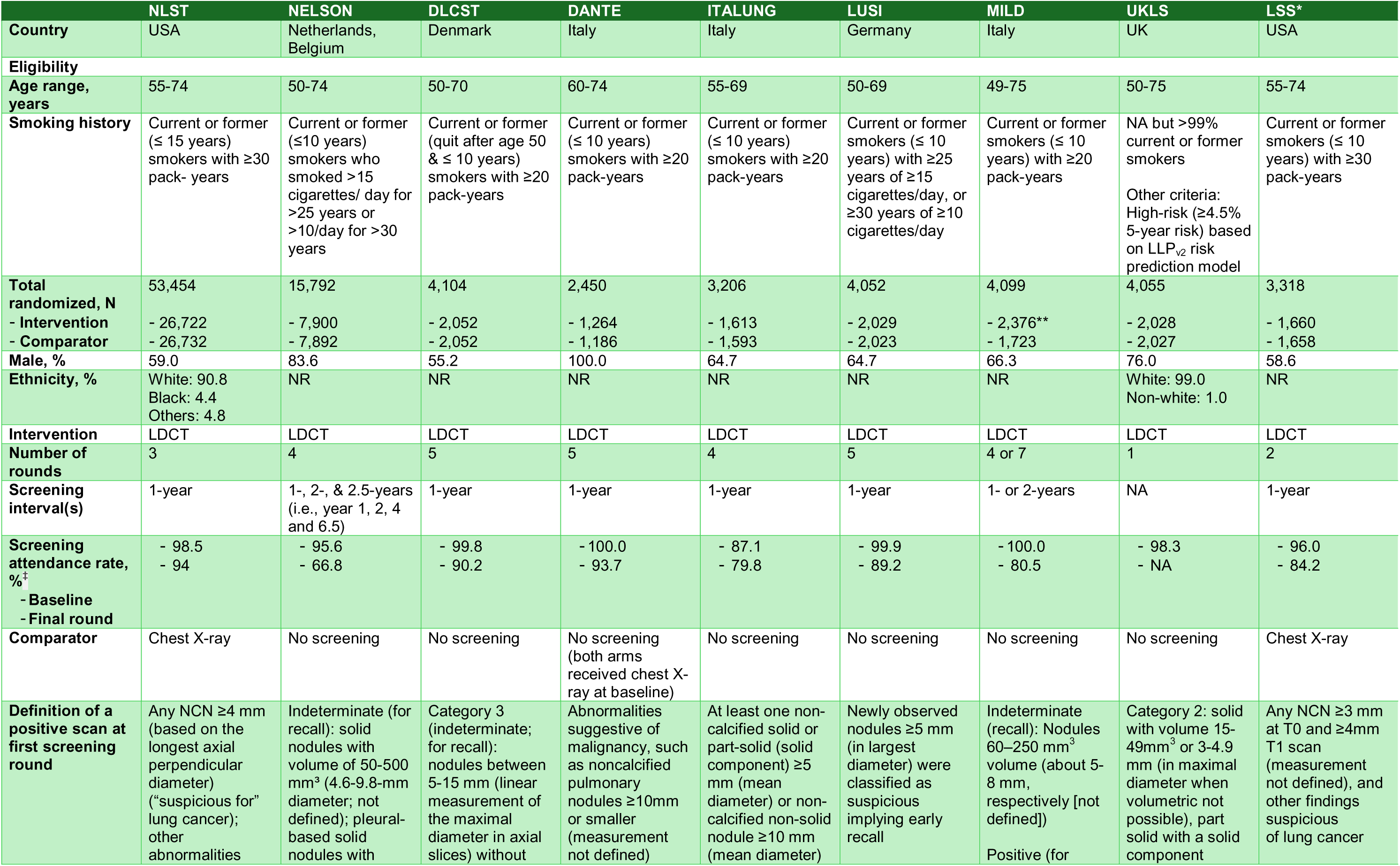
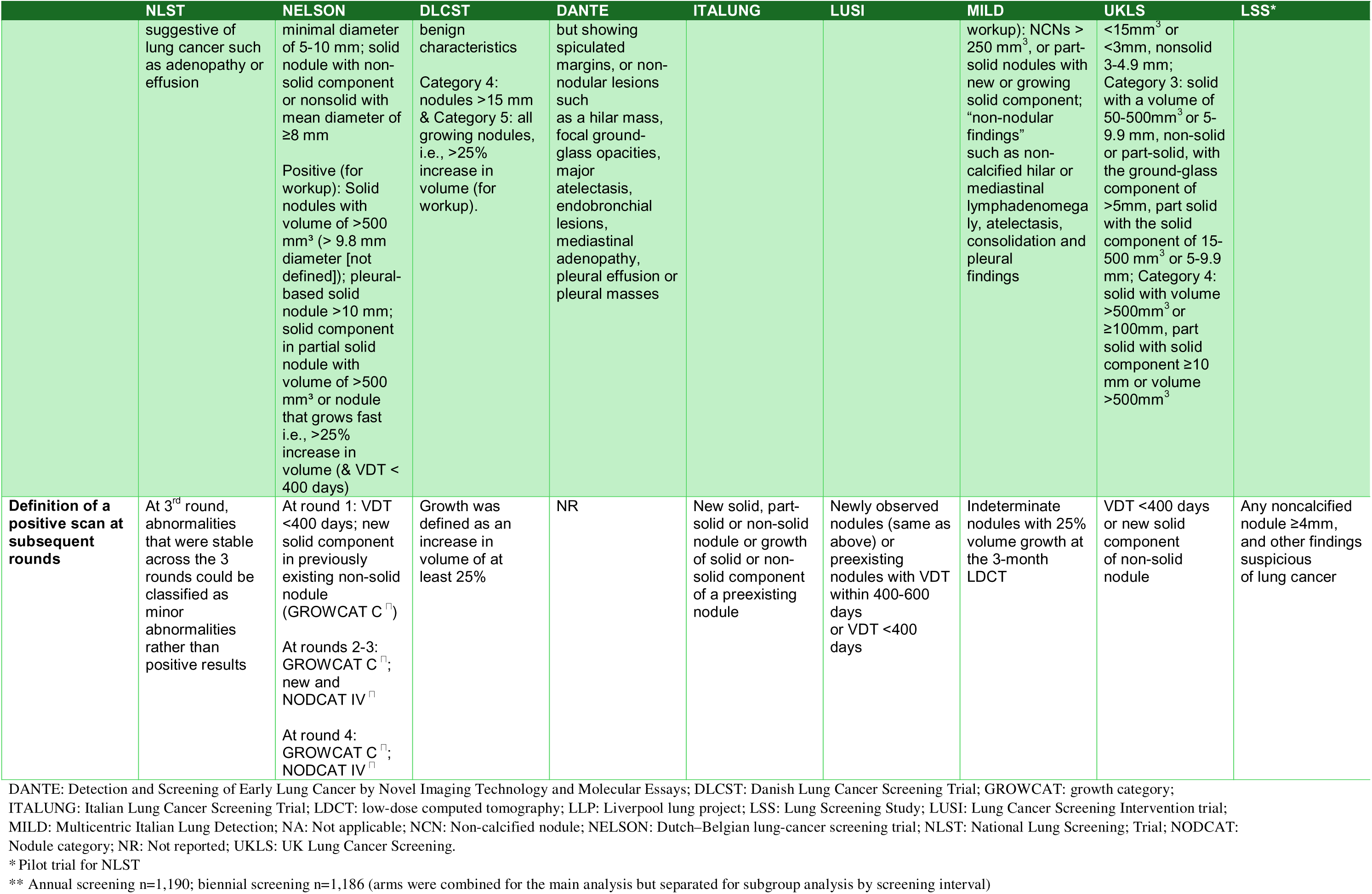
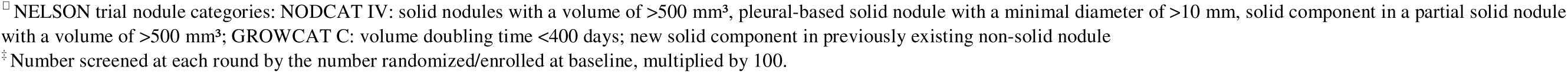
Characteristics of trials on benefits and harms of screening for lung cancer with low-dose computed tomography.

Seven trials were not rated at high risk of bias for any critical outcome at any timepoint (31, 32, 73, 74, 98, 113, 141) (Table S2.3). However, all had some concerns across one or more domains, most commonly for concerns of deviations from intended interventions where contamination was rarely reported. Other domains commonly rated to have some concerns were the randomization process (e.g., unclear allocation concealment but demonstration of similarity between groups), outcome measurement (e.g., unblinded adjudication of lung-cancer mortality though not thought to impact conclusions), and selection of the reported results (e.g., uncertain if timing of follow-up in results was prespecified). The MILD trial (116) was at high risk of bias for overdiagnosis as there were concerns that the follow-up time (after the screening phase) for outcome measurement was insufficient (i.e., <3.6 years follow-up (40)). The DANTE trial (83) was at high risk of bias for all three critical outcomes due to concerns that randomization was done before formal enrolment without adequate details on how concealment was maintained or how groups compared.

Two trials used a chest X-ray comparator at each screening round (31, 74), and six did not offer any form of screening in the control arm (32, 73, 98, 113, 116, 141). One trial offered chest X-ray to both trial arms at baseline (83). Most trials offered smoking cessation regardless of trial allocation. Six trials offered at least 4 rounds of screening (32, 83, 98, 113, 116, 141), two offered 2-3 rounds (31, 74), and one had a single round (73). Of the trials with multiple rounds, a majority screened at 1-year intervals (31, 74, 83, 98, 113, 141) and two screened less frequently (e.g., NELSON screened at baseline then after 1, 2 and 2.5 years) (32, 116). Our estimated 6-year lung-cancer diagnosis rate in the control arm was >2% for eight trials (31, 73, 74, 83, 98, 113, 116), and >3% for four trials (31, 74, 83, 113). Nodule management was considered highly sensitive for the LSS and NLST trials because a positive scan was defined as a non-calcified nodule ≥3mm (at LSS baseline scan) or ≥4mm (NLST, and LSS at round 2), or other findings that were deemed suspicious of lung cancer (31, 74). Additionally, the NLST did not account for nodule growth starting in the second round. The seven other trials used less sensitive criteria with all using larger nodules, and with the exception of the UKLS trial (73), accounted for new nodules or nodule growth starting at round two (32, 83, 98, 113, 116, 141). All trials were eligible for analysis of lung-cancer mortality and all-cause mortality, whereas data from seven trials were used to calculate overdiagnosis (32, 73, 83, 98, 113, 116, 141). The NLST and LSS trials were ineligible for overdiagnosis; they both compared LDCT with chest X-ray, which may have had a small impact on incidence rates, and the LSS did not report cancer incidence after the screening phase. The main sensitivity and subgroup analyses for lung-cancer mortality are reported in Table 2 (with associated figures in Additional file 2), whereas results of all other sensitivity and subgroup analyses can be found in Table S2.10. Post hoc, we performed sensitivity analysis including the NLT trial for overdiagnosis. There were fewer than 10 trials, therefore small-study bias was not explored.

**Table 2.** Lung-cancer mortality, main sensitivity and subgroup analyses.

| Comparison | N trials | Sample size | Relative effects (RR unless otherwise) [95% CI] | Mean control event rate | I<sup>2</sup> | Absolute difference [95% CI] per 1,000 |
| --- | --- | --- | --- | --- | --- | --- |
| Methodological sensitivity and subgroup analyses |  |  |  |  |  |  |
| Sensitivity analysis: longest follow-up vs. longest follow-up <10 years |  |  |  |  |  |  |
| Longest follow-up | 9 | 94,530 | 0.87 [0.79, 0.96] | 0.03053 | 14% | -3.97 [-6.41, -1.22] |
| Longest follow-up <10 years for NLST (6.5 years follow-up) | 9 | 94,530 | 0.83 [0.76, 0.91] | 0.02769 | 0% | -4.71 [-6.65, -2.49] |
| Sensitivity analysis: risk of bias (Fig. S2.2) |  |  |  |  |  |  |
| All trials | 9 | 94,530 | 0.87 [0.79, 0.96] | 0.03053 | 14% | -3.97 [-6.41, -1.22] |
| Some concerns of bias trials | 8 | 92,080 | 0.86 [0.77, 0.95] | 0.02855 | 21% | -4.00 [-6.57, -1.43] |
| Between-study subgroup: comparator strategy (test for subgroup differences, p=0.07) (Fig. S2.3) |  |  |  |  |  |  |
| No screening | 7 | 37,758 | 0.80 [0.71, 0.90] | 0.03041 | 0% | -6.08 [-8.82, -3.04] |
| CXR screening | 2 | 56,772 | 0.95 [0.82, 1.10] | 0.03096 | 11% | -1.55 [-5.57, 3.10] |
| Between-study subgroup: number of screening rounds (test for subgroup differences, p=0.20) (Fig. S2.4) |  |  |  |  |  |  |
| 1 round | 1 | 4,055 | 0.76 [0.51, 1.15] | 0.02516 | NA | -6.04 [-12.33, 3.77] |
| 2-3 rounds | 2 | 56,772 | 0.95 [0.82, 1.10] | 0.03096 | 11% | -1.55 [-5.57, 3.10] |
| 4+ rounds | 6 | 33,703 | 0.80 [0.70, 0.91] | 0.03129 | 0% | -6.26 [-9.39, -2.82] |
| Between-study subgroup: screening interval (test for subgroup differences, p=0.08) (Fig. S2.5) |  |  |  |  |  |  |
| Annual | 7 | 72,636 | 0.92 [0.86, 0.99] | 0.03117 | 0% | -2.49 [-4.36, -0.31] |
| >Annual | 2 | 17,840 | 0.75 [0.63, 0.90] | 0.02731 | 0% | -6.83 [-10.11, -2.73] |
| Only one round | 1 | 4,055 | 0.76 [0.51, 1.15] | 0.02516 | NA | -6.04 [-12.33, 3.77] |
| Between-study subgroup: ≤2% vs >2% 6-year lung cancer detection rate in comparator group (test for subgroup differences, p=0.004) (Fig. S2.6) |  |  |  |  |  |  |
| >2% | 8 | 90,426 | 0.86 [0.78, 0.95] | 0.03204 | 22% | -4.49 [-7.05, -1.60] |
| ≤2% | 1 | 4,104 | 1.03 [0.66, 1.60] | 0.01852 | NA | 0.56 [-6.30, 11.11] |
| Between-study subgroup: ≤3% vs >3% 6-year lung cancer detection rate in comparator group (test for subgroup differences, p=0.02) (Fig. S2.7) |  |  |  |  |  |  |
| >3% | 5 | 62,428 | 0.77 [0.67, 0.89] | 0.03869 | 0% | -8.90 [-12.77, -4.26] |
| ≤3% | 4 | 32,102 | 0.93 [0.86, 1.00] | 0.02401 | 0% | -1.68 [-3.36, 0.00] |
| Between-study subgroup: nodule management strategy (test for subgroup differences, p=0.07) (Fig. S2.8) |  |  |  |  |  |  |
| Highly sensitive | 2 | 56,772 | 0.95 [0.82, 1.10] | 0.03096 | 11% | -1.55 [-5.57, 3.10] |
| Less sensitive | 7 | 37,758 | 0.80 [0.71, 0.90] | 0.03041 | 0% | -6.08 [-8.82, -3.04] |
| Between-study subgroup analysis for demographic and clinical history |  |  |  |  |  |  |
| Between-study subgroup: sex (test for subgroup differences, p=0.31) (Fig. S2.9) |  |  |  |  |  |  |
| Men | 5 | 52,833 | 0.88 [0.76, 1.02] | 0.034 | 37% | -4.06 [-8.12, 0.68] |
| Women | 4 | 26,965 | 0.75 [0.56, 1.00] | 0.029 | 30% | -7.18 [-12.63, 0.00] |
| Within-study subgroup analyses for demographic and clinical history |  |  |  |  |  |  |
| Within-study subgroup: sex (test for subgroup differences, p=0.14) (Fig. S2.10) |  |  |  |  |  |  |
| Men vs. women | 4 | 77,348 | Ratio of RRs: 1.21 [0.93, 1.57] | NA | 45.4% | NA |
| Within-study subgroup: race (test for subgroup differences: p=0.29) |  |  |  |  |  |  |
| White | 1 (NLST) | 47,902 | Adjusted HR: 0.86 [0.75, 0.99] | NR | NA | NR |
| Black | 1 (NLST) | 2,361 | Adjusted HR: 0.61 [0.37, 1.01] | NR | NA | NR |
| Other/missing race | 1 (NLST) | 3,189 | Adjusted HR: 0.72 [0.53, 0.98] | NR | NA | NR |
| Within study subgroup: 5-year lung cancer death risk quintile (relative effects test for trend, p=0.80; absolute effects trend, p=0.01) |  |  |  |  |  |  |
| Risk quintile 1 to 5<br>5-year risk of lung-cancer death: 0.15 to 0.55% in quintile 1 to more than 2.00% in quintile 5 | 1 (NLST) | 53,158 total; ~10,000 per quintile | Lung cancer mortality ratio (ratio of expected to observed lung cancer deaths): Quintile 1: 0.97, Quintile 2: 0.78, Quintile 3: 0.75, Quintile 4: 0.70, Quintile 5: 0.84; p=0.80 for trend<br>Overall rate of death from lung cancer: 24.6 per 10,000 person-years vs. 30.9 per 10,000 person years<br>77 of 88 CT-prevented lung-cancer deaths (88%) occurred among the 60% of participants with a 5-year risk of lung-cancer death of 0.85% or more (i.e., in quintiles 3 through 5), whereas only 1% of prevented lung-cancer deaths occurred among the 20% of participants at lowest risk (i.e., in quintile 1). |  |  | Per 10,000 person years:<br>Quintile 1: 0.2<br>Quintile 2: 3.5<br>Quintile 3: 5.1<br>Quintile 4: 11.0<br>Quintile 5: 12.0 p=0.01 for trend<br>NNS:<br>Quintile 1: 5276<br>Quintile 2: 531<br>Quintile 3: 415<br>Quintile 4: 171<br>Quintile 5: 161<br>p<0.001 for trend |
| Within study subgroup: PLCO 6-year cancer risk (p-value for interaction p=0.61) |  |  |  |  |  |  |
| Quartiles 1 to 4<br>Quartiles based on PLCO<sub>M</sub>2012 **6-year risk for cancer** (risks not reported) | 1 (NLST) | 53,202 total; roughly equally sized groups per quartile | HRs:<br>Quartile 1: 0.86 [0.50, 1.48];<br>Quartile 2: 0.71 [0.49, 1.04];<br>Quartile 3: 0.70 [0.53, 0.91]; |  |  |  |
|  |  |  | Quartile 4: 0.88 [0.73, 1.06]; p-value for interaction=0.61<br>At all four levels of risk, the screening effect was protective. Random variation may explain differences in hazard ratios according to risk quartiles. |  |  |  |
| Within study subgroup: Sex, and PLCO 6-year cancer risk (test for interaction, p=NR) |  |  |  |  |  |  |
| Women vs men, quintiles 1 to 5<br>Quintiles based on PLCO<sub>M</sub>2012 **6-year risk for cancer** | 1 (NLST) | 53,452 | Odds ratios (men as reference group), p-value:<br>Quintile 1: 1.63 (95% CI 0.79 to 2.49), p=0.15<br>Quintile 2: 0.78 (95% CI 0.59 to 1.37), p=0.46<br>Quintile 3: 1.38 (95% CI 0.93 to 1.82), p=0.11<br>Quintile 4: 1.09 (95% CI 0.73 to 1.47), p=0.62<br>Quintile 5: 1.45 (95% CI 1.17 to 1.72), p=0.002<br>OR (95% CI) data estimated from a figure |  |  |  |
| Within-study subgroup: deciles of PLCO<sub>m2012</sub> lung cancer risk among PLCO trial participants applied to NLST population (test for subgroup difference, p=NR) |  |  |  |  |  |  |
| Risk percentile: 0 to 10 | 1 (NLST) | 1 | NA (zero in denominator) | NR | NA | NR |
| Risk percentile: >10 to 20 | 1 (NLST) | 8 | NA (zero in denominator) | NR | NA | NR |
| Risk percentile: >20 to 30 | 1 (NLST) | 154 | NA (zero in denominator) | NR | NA | NR |
| Risk percentile: >30 to 40 | 1 (NLST) | 997 | 1.76 [0.09, 103.71] | NR | NA | 2.48 [-7.74, 12.68] per 10,000 py |
| Risk percentile: >40 to 50 | 1 (NLST) | 3,094 | 0.73 [0.18, 2.68] | NR | NA | -1.82 [-8.50, 4.87] per 10,000 py |
| Risk percentile: >50 to 60 | 1 (NLST) | 5,940 | 0.62 [0.28, 1.36] | NR | NA | -3.68 [-9.31, 1.95] per 10,000 py |
| Risk percentile: >60 to 70 | 1 (NLST) | 8,581 | 1.01 [0.61, 1.66] | NR | NA | 0.14 [-5.74, 6.01] per 10,000 py |
| Risk percentile: >70 to 80 | 1 (NLST) | 10,819 | 0.74 [0.52, 1.04] | NR | NA | -6.06 [-12.74, 0.62] per 10,000 py |
| Risk percentile: >80 to 90 | 1 (NLST) | 10,997 | 0.76 [0.58, 0.99] | NR | NA | -9.38 [-18.07, -0.70] per 10,000 py |
| Risk percentile: >90 to 100 | 1 (NLST) | 11,374 | 0.92 [0.76, 1.10] | NR | NA | -5.94 [-18.17, 6.30] per 10,000 py |
| At PLCO<sub>m2012</sub> risk ≥0.0151 (1.5%), the 65<sup>th</sup> percentile of risk, the NLST CT arm mortality rates are consistently below the CXR arm's rates. The NNS to prevent one lung cancer death in the 65th to 100th percentile risk group was 255 (95% CI 143 to 1,184; ARD 6.43 per 10,000 person-years), and in the 30th to <65th percentile risk group was 963 (95% CI 291 to 2754; ARD 1.60 fewer per 10,000 person-years); the NNS could not be estimated in the <30<sup>th</sup> percentile risk group because of absence of lung cancer deaths. |  |  |  |  |  |  |

ARD=adjusted risk difference; HR=hazard ratio; CT=computed tomography; CXR=chest X-ray; NA=not applicable; NNS=number needed to screen; NR=not reported; PY=person-years

##### Lung-cancer mortality

Data on lung-cancer mortality came from results at longest follow-up from all trials (N=94,530; absolute difference of 4.0 fewer per 1,000 [95% CI, 1.2 to 6.4 fewer] over 10-12 years (31, 32, 73, 74, 83, 98, 113, 116, 141)) (Figure 1). No result from a shorter time-point was at lower risk of bias. Further, a random-effects model was chosen because although there was a large difference in sample sizes there was no suspicion of missing small studies and the results of the high risk of bias trial (DANTE n=2,450 (83)) did not impact findings. The NLST (n=53,454), had some risk of bias concerns for lung-cancer mortality at both a median of 6.5 and 12.3 years from randomization (31, 170). As a *post-hoc* sensitivity analysis, we explored whether using the shorter timepoint for this trial would impact the findings as it contributed the most weight to the meta-analysis (RR 0.83 [95% CI, 0.76 to 0.91]; 4.7 fewer per 1000). Although the relative effects indicate less benefit at longer-term follow-up, absolute findings were very similar between analyses as the pooled control event rate/baseline risk increased with duration of follow-up (31 vs. 28 per 1000).

**Figure 1.**
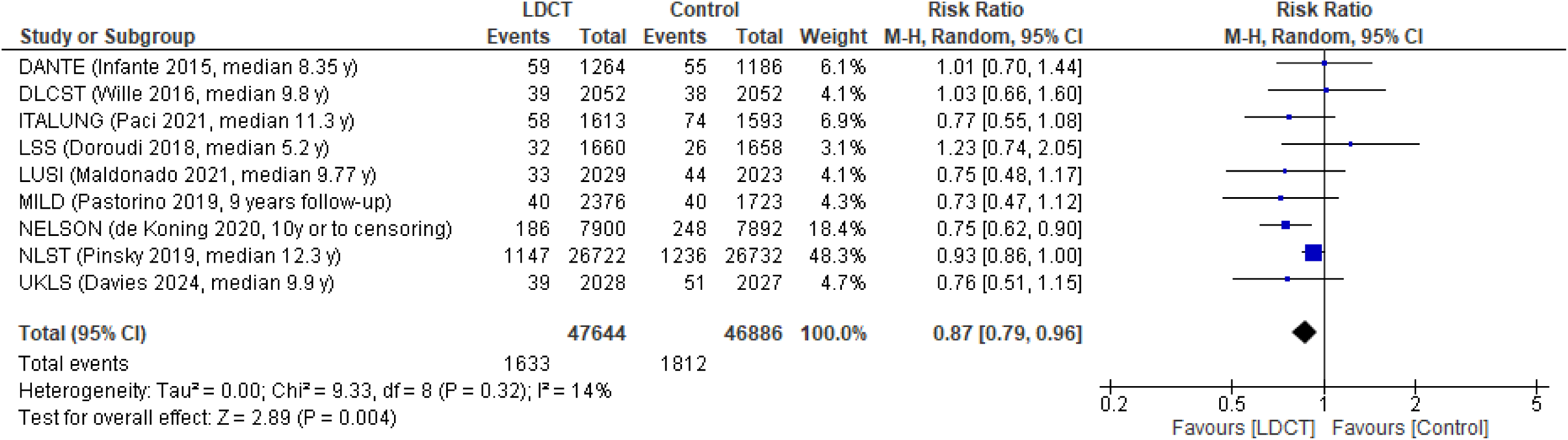
Effect on lung-cancer mortality from LDCT

The credibility of most subgroup findings was low. Across most of our between-study/stratified analyses (Table 2), while there were some differences in magnitude of relative and absolute effects, the point estimates of the absolute effects generally remained above our decision threshold of 1 fewer lung-cancer death per 1,000 screened. Further, the heterogeneity within strata and multiple differences across trials may confound possible subgroup effects (e.g., NLST had both a higher sensitivity for its nodule classification and used a chest X-ray comparator).

Using data reported by sex and a trial only enrolling men, the between-study analysis did not show a significant difference in the effects from screening between men and women, however absolute findings showed a potentially important difference with greater effects for women (7.2 fewer per 1000 vs. 4.1 fewer for men). The within-study analysis using a ratio of ratios yielded similar results (men vs. women, ratio of RRs 1.21 [95% CI, 0.93 to 1.57]; Figure S2.10). An analysis by the authors of the NELSON trial also found larger effects for females versus males (ratio of ratio 1.34) when limiting data to those also eligible for the NLST (32). Of note, none of these findings were adjusted for potential confounders. The effects based on sex did not lead to serious inconsistency in the main analysis across sexes, or separately rating certainty by sex, because both findings exceeded the outcome’s decision threshold.

Several studies reported other within-study analyses for participant characteristics, though we did not pool any apart from sex due to differences in definitions and effect measures. Findings for race or ethnicity from a single trial (NLST) enrolling few non-Caucasians support a possibility of greater benefit for non-Caucasian participants, though absolute risks were not reported (Table 2).

Findings by predicted risk for lung-cancer incidence and mortality at baseline were also only reported for NLST participants. One report categorized participants into five quintiles of 5-year lung-cancer death risk, each having roughly the same sample size. Across quintiles, the lung-cancer mortality ratios were not significantly different; however, a difference was seen in the absolute findings per 10,000 person-years (p=0.01) (160). Lung-cancer mortality reductions increased with 5-year risk, from 0.2 in quintile 1 (i.e., approximately equivalent to 0.2 per 1,000 over 10 years) to 12.0 in quintile 5. Another report used quartiles based on PLCOm2012 6-year risk of cancer among NLST participants and found similar protective effects of screening across quartiles (18). A third report explored categorizing NLST participants into deciles by PLCO_m2012_ 6-year risk of cancer (179). The predicted risks among the NLST participants were fitted into the deciles created from data for the smoking participants in the main trial (PLCO) used to develop this model. At and above the 65^th^ percentile of risk (i.e., about a 1.5% 6-year risk), mortality rates in the LDCT screening arm were consistently lower than the chest X-ray arm. A limitation to this last analysis is that the deciles had unequal numbers of NLST participants with many fewer participants in the lower-risk deciles. Overall, we assessed findings as moderately credible that the relative effects from screening are consistent across baseline risks, though absolute findings may vary greatly as expected especially over longer-term follow-up. Other within-study subgroup findings for age, smoking status (current vs. former and based on pack-years), socioeconomic status, and coexisting conditions (chronic-obstructive pulmonary disease) were reported in single studies or showed inconsistent effects across studies possibly due to other confounding variables (Table S2.7). There was some consistency using data from three trials indicating that there may be little-to-no effect from LDCT screening (adjusted relative effects 1.01 in NLST (190) and 1.32 in DANTE and MILD combined (158)) for former smokers (i.e., quit ≤15 years in NLST and ≤10 years in others). No data were located examining effects related to 2SLGBTQ+ status or homelessness.

Overall, we concluded with moderate certainty that among people 50-74 years of age who are current or former smokers with 20-30 pack-years’ smoking history, screening with LDCT 3-4 times probably reduces lung-cancer mortality by at least 1 per 1,000 screened after about 10-12 years follow-up (Table 3). We did not have serious concerns about inconsistency, with >80% of the weight in the analysis coming from trials showing effects surpassing our threshold of 1 per 1,000 screened. There were serious concerns about indirectness from the within-study subgroup findings suggesting those at lower risk (despite meeting age and pack-year history) will fall under this threshold. The certainty for this outcome was also moderate for effects of at least 2 fewer per 1,000, though noting some imprecision from the lower limit of the 95% CI for the absolute effects.

**Table 3.** Summary of findings for LDCT screening: Critical outcomes.

| Outcome<br>No. participants<br>(studies)<br>Threshold(s) | Relative<br>effects, RR<br>(95 % CI) | Anticipated absolute effects |  |  | Certainty** | What happens? |
| --- | --- | --- | --- | --- | --- | --- |
|  |  | With screening | Without screening* | Difference |  |  |
| Lung cancer mortality<br>(Fig. 1)<br>Follow-up: 10-12 years<br>N=94,530 (9 RCTs)<br>Threshold: 1 fewer per 1000 | RR 0.87 (0.79 to 0.96) | 26.6 per 1000<br>(24.1 to 29.3) | 30.5 per 1000 | 4.0 fewer (1.2 to 6.4 fewer) per 1000 | **MODERATE<sup>c</sup>**<br>⊕⊕⊕⊖ | Among people 50-74 years of age who are current or former smokers, screening with LDCT 3-4 times probably reduces lung-cancer mortality by at least 1 per 1000 screened after 10-12 years follow-up.<br>The certainty is moderate for effects of at least 2 fewer per 1000 screened. |
| All-cause mortality<br>(Fig. 2)<br>Follow-up: 10-12 years<br>N=94,530 (9 RCTs)<br>Thresholds: 0.5 and 1 fewer per 1000 | RR 0.97 (0.93 to 1.01) | 118.2 per 1000<br>(113.3 to 123.1) | 121.8 per 1000 | 3.7 fewer (8.5 fewer to 1.2 more) per 1000 | **LOW<sup>b, d</sup>**<br>⊕⊕⊖⊖ | Among people 50-74 years of age who are current or former smokers, screening with LDCT 3-4 times may reduce all-cause mortality by at least 1 per 1000 screened after 10-12 years.<br>The same certainty rating applies for the threshold of 0.5 fewer per 1000. |
| Overdiagnosis (excess cancer incidence)<br>(Fig. 3)<br>Follow-up: Median 10 years from randomization (majority 3-5 years after screening)<br>N=35,161 (7 RCTs)<br>Threshold: 2.5 more per 1000 | RR 1.19 (1.03 to 1.37) | 52.6 per 1000<br>(45.5 to 60.5) | 44.2 per 1000 | 8.4 more (1.3 to 16.3 more) per 1000 | **MODERATE<sup>c</sup>**<br>⊕⊕⊕⊖ | Among people 50-74 years of age who are current or former smokers, screening with LDCT 3-4 times probably causes at least 2.5 per 1000 to be overdiagnosed after 10 years. |

\*Mean unweighted control event rate.
\*\* Reasons for rating down certainty: a=risk of bias, b=inconsistency, c=indirectness, d=imprecision. Explanations are within the text.

**Table 4.** Summary of findings for patient preferences, disutilities.

| Number of included studies;<br>Sample size | Certainty* | What does the evidence say? |
| --- | --- | --- |
| **Unaffected eligible population comparator (FOR USE TO CALCULATE DISUTILITIES OF OTHER HEALTH STATES)** |  |  |
| 5 studies; N=23,466 (EQ-5D: 2 studies, N=1,237; SF-6D: 2 studies, N=4,918; 15D: 1 study, n=4,191)<br>Alberta norms; n=19,859 | **LOW<sup>b, c</sup>**<br>⊕⊕⊖⊖ | The utility value for an unaffected eligible population comparator may be 0.81. |
| **Disutility of positive screening result (results not known; among those without cancer)** |  |  |
| Within study, EQ-5D: 2 studies; N=263<br>Between study: 3 studies; N=778 (EQ-5D: 2 studies, N=263; SG: 1 study, n=515) | **MODERATE<sup>d</sup>**<br>⊕⊕⊕⊖ | There is probably little-to-no disutility (0.01) of receiving a positive screening result. |
| **Disutility of false positive result (results known)** |  |  |
| Within study, EQ-5D: 1 study; N=246 (1 month and 12 months after results)<br>Between study: 2 studies; N=371 (EQ-5D: 1 study, N=168; TTO: 1 study; N=203; up to 1 month after diagnostic workup) | **LOW<sup>c, d</sup>**<br>⊕⊕⊖⊖ | There may be little-to-no disutility (0.00) of a false positive result. |
| **Disutility of lung cancer at diagnosis/treatment naïve, stage I-IIIa** |  |  |
| Between study:<br>8 studies; N=1,572 (EQ-5D: 8 studies, N=1,057; SG: 1 study; n=515) | **LOW<sup>c, d</sup>**<br>⊕⊕⊖⊖ | There may be little-to-no disutility (0.01) for stage I-IIIa lung cancer at diagnosis/treatment naïve. |
| **Disutility of lung cancer at diagnosis/treatment naïve, stage IIIB-IV** |  |  |
| Between study:<br>10 studies; N=2,243<br>(EQ-5D: 9 studies, N=1,728;<br>SG: 1 study, n=515) | **LOW<sup>c, d</sup>**<br>⊕⊕⊖⊖ | There may be a small, but important disutility (0.07) for stage IIIB-IV lung cancer at diagnosis/treatment naïve. |
| **Disutility of lung cancer, on first-line treatment (none only receiving surgery), stage I-IIIa** |  |  |
| Between study:<br>EQ-5D: 3 studies; N=609 | **LOW<sup>c, d</sup>**<br>⊕⊕⊖⊖ | There may be a small, but important disutility (0.04) for stage I-IIIa lung cancer during first-line treatment. |
| **Disutility of lung cancer, on first-line treatment, stage IIIB-IV** |  |  |
| Between study:<br>13 studies; N=2,635 (EQ-5D: 12 studies, N=2,335; TTO: 1 study, n=300) | **MODERATE<sup>c</sup>**<br>⊕⊕⊕⊖<br>**LOW<sup>b, c</sup>**<br>⊕⊕⊖⊖ | There is probably a small, but important disutility (at least 0.04) for stage IIIB-IV lung cancer on first-line treatment.<br>There may be a moderate disutility (0.09) for stage IIIB-IV lung cancer on first-line treatment. |
| **Disutility of lung cancer, after first-line treatment completion and non-progressive (post-surgery, on maintenance therapy, or survivors), stage I-III A** |  |  |
| Within study:<br>15D: 1 study; n=230<br>Between study:<br>11 studies; N=13,848 (EQ-5D: 10 studies, N=13,618; 15D: 1 study, n=230) | **LOW<sup>b, c</sup>**<br>⊕⊕⊖⊖ | There may be little-to-no disutility (0.03) for stage I-III A lung cancer after first-line treatment completion without progression. |
| **Disutility of lung cancer, after first-line treatment completion and non-progressive (on maintenance therapy, or survivors) stage IIIB-IV** |  |  |
| Between study:<br>8 studies; N=1,158 (EQ-5D: 7 studies, N=1,083;<br>TTO: 1 study, n=75) | **LOW<sup>c, d</sup>**<br>⊕⊕⊖⊖ | There may be a small, but important disutility (0.06) for stage IIIB-IV lung cancer after first-line treatment completion without progression. |

\*Reasons for rating down certainty: a=risk of bias, b=inconsistency, c=indirectness, d=imprecision. Explanations are within the text and Additional file 3. All figures are located in Additional file 3.

##### All-cause mortality

Based on evidence from nine trials (N=94,530 (31, 32, 73, 74, 83, 98, 116, 141, 172)), among people 50-74 years of age who are current or former smokers, screening with LDCT 3-4 times may reduce all-cause mortality by at least 1 per 1,000 after 10-12 years (Table 3 and Figure 2; low certainty). The main analysis found an absolute difference of 3.7 fewer per 1,000 (95% CI, 8.5 fewer to 1.2 more). We used a random-effects model as no small study bias was detected, and sensitivity analysis, excluding the one high risk of bias trial (n=2,450 (83)), yielded similar absolute findings that passed the threshold of effect (Table S2.7). We had serious concerns about inconsistency in the absolute effects between trials for both of our thresholds of 0.5 and 1.0 fewer per 1,000. The inconsistency not was explained by any of the subgroup findings, including differences between trials based on the comparator. We also had serious concerns about imprecision, as the upper limit of the 95% CI crossed both thresholds. The certainty was also low for effects of at least 0.5 fewer per 1,000 for the same reasons. There may be similar concerns as for lung-cancer mortality about applicability to those at lower baseline risk, though we did not rate down the certainty further for this possibility.

**Figure 2.**
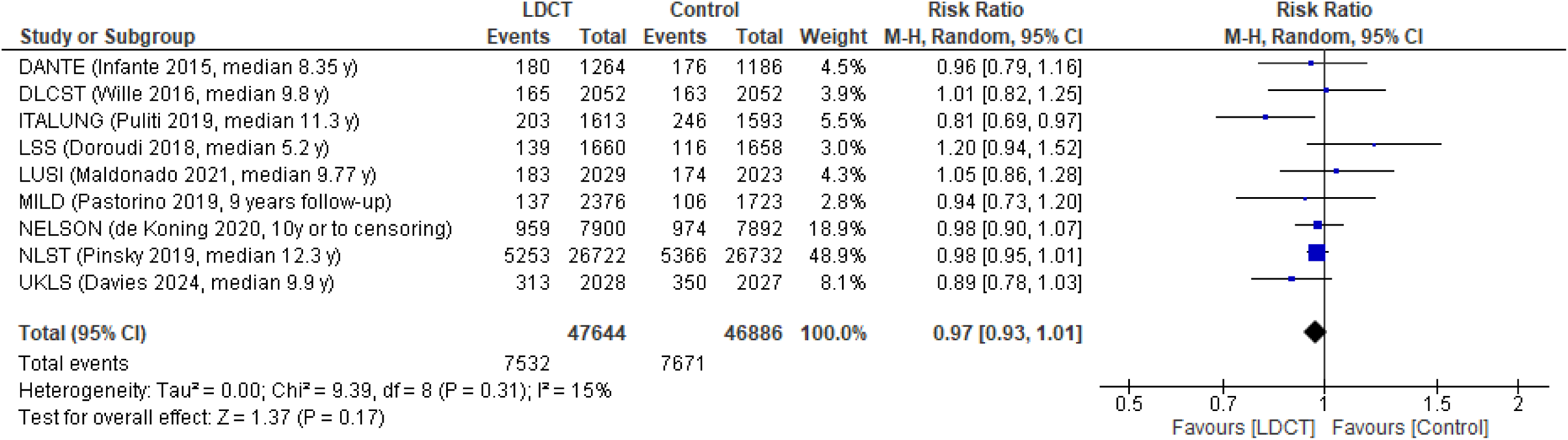
Effect on all-cause mortality from LDCT

**Figure 3.**
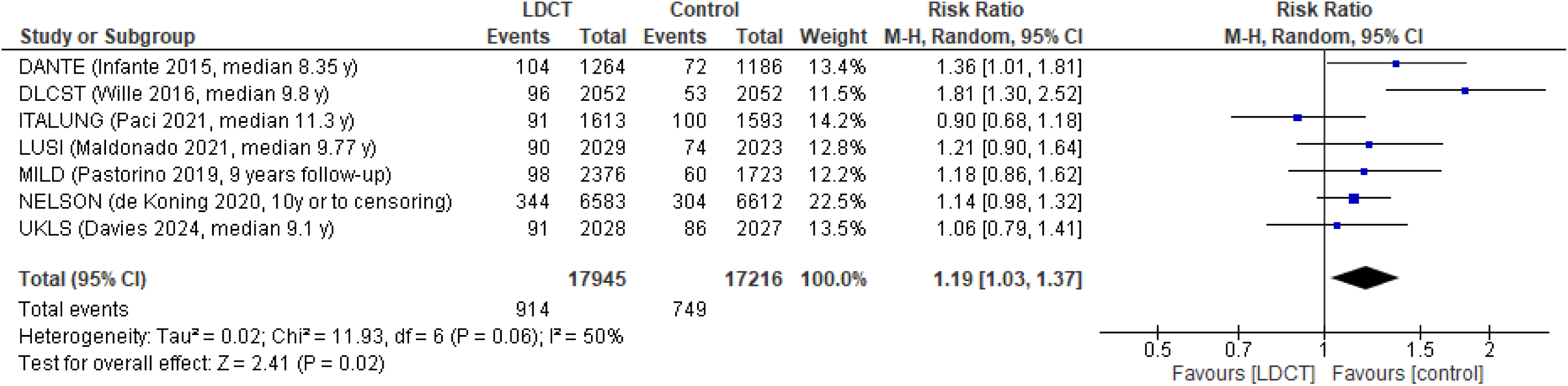
Effect on lung cancer diagnoses from LDCT, to calculate overdiagnosis

##### Overdiagnosis

Data from seven trials were used to estimate overdiagnosis (excess cancer diagnoses) (N=35,161 (32, 73, 83, 98, 113, 116, 141)) which was assessed using a threshold of 2.5 per 1,000 screened (Table 2). There was moderate certainty evidence based on an absolute difference of 8.4 per 1,000 screened (95% CI, 1.3 to 16.3). We used a random-effects model as there was an absence of small study effects (Tables S2.7). There were no serious concerns about risk of bias, as sensitivity analysis resulted in similar absolute findings whether the two trials at high risk of bias were excluded (83, 116). There were serious concerns of indirectness, because though we did not have a minimum follow-up duration after screening completion the duration of follow-up in most RCTs (3 to 5 years) was considered short (modelling suggests lead time may be at least 9 years (191)) and thus findings may overestimate the true effects. We did not have serious concerns about inconsistency or imprecision. Between-study subgroup findings consistently showed results that all passed the threshold for absolute effects. Our post-hoc sensitivity analysis, adding results from the NLST trial with its chest X-ray comparator (RR 1.01 95% CI, 0.95-1.09), also passed our threshold of effect (RR 1.15 [95% CI, 1.02 to 1.30]; 7.0 more [95% CI, 0.9 to 14.0 more] per 1000). Within-study and between-study analyses exploring sex differences showed no statistical differences. Absolute differences for both sexes surpassed the threshold of effect, with women potentially experiencing more overdiagnosis (ratio of RRs 0.93 [95% CI, 0.65 to 1.33]; men: 7.8 per 1,000 vs. women: 8.2 per 1,000 (32, 83, 98, 187)). There are several other definitions of overdiagnosis that were explored, of which calculations can be found in Table S1.4 and findings reported in Table S2.8. On average, the proportion of screen-detected cancers that might be overdiagnosed was 24%.

#### Low-dose chest tomography screening strategies: important outcomes

We included 11 RCTs (17, 64, 125, 145, 152, 155, 157, 163, 165, 167, 168) and 72 observational studies (61–63, 65–72, 75–82, 84–97, 99–112, 114, 115, 117–124, 126–130, 132, 134–140, 142, 143) that reported on at least one of the important outcomes. False positives, incidental findings, major complications and deaths from invasive testing were only extracted for those receiving LDCT, whereas psychosocial harms could be extracted for both groups and HRQoL (a benefit outcome) required data from both arms in RCTs. The eligibility criteria for observational studies were generally similar to the trials, and rarely included individuals at lower risk (never-smokers and individuals under 50 years of age). Mean age across all studies was 61.4 years, and the median sample size was 1,715 (range 72 to 152,918). On average, 63.3% of the participants were male, and across 31 studies reporting race or ethnicity, 19 included ≥80% Caucasian individuals. Other races or ethnicities were rarely reported. A full description of the characteristics of trials and observational studies are reported in Tables S2.1 and S2.2 (Additional file 2). All relevant sensitivity and subgroup analyses based on differences between and within studies are reported in Table S2.10 and Table S2.9 contains the summary of findings and certainty of evidence for outcomes rated as important for this review.

The risk of bias assessments are reported in Tables S2.3, S2.4, S2.5 and S2.6. The most frequent reasons that comparative data (HRQoL and psychosocial harms) were at high risk of bias were selective reporting of data among multiple timepoints or missing outcome data/attrition that was unbalanced between groups. Proportion data from people receiving screening were most commonly at high risk of bias due to attrition (e.g., insufficient number with resolved diagnostic workup), lack of detailed outcome definitions (e.g., unclear exclusion for incidental findings of pre-existing conditions), and insufficient methods of assessing outcomes (e.g., single radiologist without a structured template for classifying findings).

Additional data that were not evaluated with GRADE, including all event/rate data, proportions of FPs requiring diagnostic follow-up, deaths from invasive diagnostic testing including those with cancer, and specific incidental findings of interest are reported in Table S2.11.

##### False positives

Data on the proportion of individuals with a FP result (requiring recall for imaging or a diagnostic follow-up) was grouped based on a single round of screening (11 trials, N=48,888 (17, 64, 125, 145, 152, 155, 157, 162, 165, 167, 168)); 18 observational, N=40,757 (63, 67, 70, 81, 92, 96, 97, 100, 104, 108, 109, 112, 115, 134, 138, 140, 142, 185)), or cumulative effects across 2-3 rounds (2 trials, N=28,382 (17, 147); 8 observational, N=55,308 (67, 81, 88, 97, 104, 126, 140, 142)) or 4-5 rounds (2 trials, N=2,877 (83, 162); 1 observational, n=1,267 (105)). Pooled findings for 1, 2-3 and 4-5 rounds of screening were 190 per 1,000 (95% CI, 160 to 220), 310 per 1,000 (95% CI 180 to 440), and 350 per 1,000 (95% CI, 220 to 480), respectively. Considering all data, there was high certainty evidence that for people 50-74 years of age who are current or former smokers, screening with LDCT 3-4 times causes at least 75 people to have one or more FP results per 1,000 screened. Findings were similar between trials and observational studies. There was high risk of bias in six studies at the first screening round (17, 63, 92, 96, 109, 152, 162) and both studies over 2-3 rounds (17, 147), though findings were similar when excluding these studies in sensitivity analysis. Subgroups based on study design and nodule management, and within-study subgroups based on age, sex, and risk quintile all surpassed the threshold of effect. Our certainty would be moderate for effects up to 225 people per 1,000 after 3-4 rounds because we would have serious concerns about inconsistency among studies. A large majority of FPs only require recall and not invasive diagnostic procedures; though not reported in detail here or assessed for its certainty, the proportions requiring diagnostic follow-up were between 20 and 30 per 1000 screened (e.g., <10% of FPs) from one or across multiple rounds (Table S2.11).

##### Incidental findings

At round 1, no trials and 13 observational studies (N=14,549 (65, 67, 76, 84, 91–93, 97, 106, 117, 122, 129)) had findings of 550 incidental findings per 1,000 (95% CI, 370 to 730). Across 2-3 rounds a single trial (n=26,722 (151)) reported 570 per 1,000 (95% CI 560 to 570), and across 4-5 rounds there were two trials (N=2,877 (83, 172)) and one observational study (n=519 (117)) with findings of 400 per 1,000 (95% CI, 50 to 750). We concluded that for individuals 50-74 years of age who are current or former smokers, screening with LDCT 3-4 times causes at least 150 people to have one or more incidental finding per 1,000 screened. We had high certainty in these effects, as well as for the lower threshold of 100 per 1,000 screened. Sensitivity analysis showed that findings were similar and continued to surpass the threshold of effect after excluding six high risk of bias studies at round 1 (65, 76, 84, 92, 93, 102, 117, 122) and one study over 4-5 rounds (117). A between-study subgroup analysis by study design yielded similar results for both trials and observational studies. The certainty was moderate for effects of at least 450 people having an incidental finding per 1,000 screened as we would have serious concerns of inconsistency among studies contributing to analyses at both round 1 and across 4-5 rounds of screening.

##### Clinically significant incidental findings

Clinically significant incidental findings were those having high suggestibility that further review or intervention was required. Round 1 included two trials (N=27,582 (151, 161)), and 26 observational studies (N=81,843 (71, 72, 78, 82, 85, 86, 89–93, 97, 101, 102, 106, 108, 117, 118, 121, 122, 124, 129, 136, 144)), whereas over 2-3 rounds there was one trial (n=26,722 (151)), and over 4-5 rounds there were four observational studies (N=12,026 (99, 117, 121, 174)) that reported on this outcome (Table S2.9).

From 1, 2-3, and 4-5 rounds, absolute findings were 220 per 1,000 (95% CI 180 to 260), 340 per 1,000 (95% CI 330 to 340), and 80 per 1,000 (95% CI, 30 to 120), respectively. Findings indicate that among people 50-74 years of age who are current or former smokers, screening with LDCT 3-4 times causes at least 50 (high certainty), and probably causes at least 100 (moderate certainty) people to have one or more clinically significant incidental finding per 1,000 screened. For our larger threshold of 100 per 1,000 we rated down once due to serious concerns about inconsistency among studies contributing to analyses at round 1 and across 4-5 rounds of screening. We compared our overall findings to a sensitivity analysis excluding 24 studies at round 1 (71, 72, 78, 85, 86, 89, 90, 92, 93, 101, 102, 108, 117, 118, 121, 122, 124, 136, 144, 161), and three studies over 4-5 rounds (99, 117, 121) at high risk of bias. The sensitivity analysis resulted in similar absolute findings that passed both thresholds of effect; further, any impact from risk of bias was considered to contribute to inconsistency for which we rated down. Additionally, between- and within-study subgroup analysis findings were consistent with overall findings at each timepoint.

##### Major complications from invasive diagnostic testing

Major complications that occurred during invasive diagnostic testing after a positive LDCT screen were analyzed for those who either received a FP result or a lung cancer diagnosis as well among only those with a FP result. Using clinical input, we identified reported complications and determined which were considered “major”. We did not find evidence of missing outcome bias for these outcomes, as they were reported among 64% (87,804/136,997) of the total sample size of studies contributing to a FP analysis.

Major complications from invasive diagnostic testing were reported at round 1 (4 trials, N=9,829 (32, 64, 152, 157); 7 observational, N=19,795 (75, 92, 94, 107, 108, 123, 139)), across 2-3 rounds (1 trial, N=26,722 (156); 4 observational, n=48,344 (88, 99, 100, 126)) and across 4-5 rounds (2 trials, n=8,196 (32, 162); 1 observational, n=2,471 (124)). Results showed 0.7 per 1,000 (95% CI, 0.0 to 2.40) for round 1, 3.0 per 1,000 (95% CI 1.3 to 5.1) over 2-3 rounds, and 1.8 per 1,000 (95% CI, 0.0 to 7.9) over 4-5 rounds. Based on this data we have low certainty evidence that screening with LDCT 3-4 times may not cause at least 2.5 people to have at least one major complication from invasive diagnostic testing per 1,000 screened. We rated down twice due to very serious concerns of inconsistency within and between the analyses at each timepoint. Between-study subgroups by study design were generally consistent, and within-study subgroup findings were based on single studies. Two studies at round 1 were high risk of bias (92, 94), but findings were consistent whether these studies were included or not.

Considering only major complications in participants with a FP result, we included five trials (N=10,751 (32, 64, 125, 152, 157)) and six observational studies (N=13,040 (68, 75, 92, 100, 107, 123)) at round 1, one trial (n=26,722 (156)) and two observational studies (n=2,259 (99, 100)) across 2-3 rounds, and one trial (n=6,583 (32)) and one observational study (n=2,471 (124)) across 4-5 rounds. Events were low, with 0.0 per 1,000 (95% CI, 0.0 to 0.3) at round 1, 0.0 per 1,000 (95% CI, 0.0 to 0.2) over 2-3 rounds, and 0.1 per 1,000 (95% CI, 0.0 to 0.4) over 4-5 rounds. Findings were similar between trials and observational study designs, and when removing from round 1 data the study at high risk of bias (92). We found that screening with LDCT 3-4 times does not cause at least 2.5 people to have a major complication from invasive testing among those without cancer per 1,000 screened. We had no serious concerns in any GRADE domain. The certainty would be moderate for a threshold of fewer than 1 per 1,000 screened, where we would have serious concerns about inconsistency among studies.

##### Deaths from invasive diagnostic testing among those without cancer

For deaths as a result of invasive diagnostic testing among FPs, at round 1 (3 trials, N=3,309 (64, 83, 152); 3 observational, N=44,123 (88, 94, 164)) absolute findings were 0.0 per 1,000 (95% CI, 0.0 to 0.0), over 2-3 rounds (1 trial, n=26,722 (17); 1 observational, N=2,003 (99)) findings were 0.1 per 1,000 (95% CI, 0.0 to 0.3), and over 4-5 rounds (1 trial, N=1,264 (83)) findings were 0.0 per 1,000 (95% CI, 0.0 to 2.9). There were no serious concerns of missing outcome bias, as 50% (69,080/136,997) of the sample reporting on FPs also reported on this outcome. One observational study was at high risk of bias at round 1 (94), but its findings did not change conclusions. Among people 50-74 years of age who are current or former smokers, screening with LDCT 3-4 times may not cause at least 0.1 deaths from invasive testing among those without a diagnosis of lung cancer per 1,000 screened. There was moderate certainty evidence due to serious concerns of imprecision due to inadequate sample size for this very rare outcome. We did not rate own for inconsistency; the only finding above our threshold was from the NLST (0.2 per 1000) which used 60-day mortality from any cause to define the outcome which could have captured deaths attributable to other causes.

##### Health-related quality of life

Authors of two trials used subsamples to evaluate whether HRQoL was impacted by screening for lung cancer (150, 181). The NELSON trial (n=1,125 (181)) used the EQ-VAS and SF-12 mental and physical component scores (both having scales 0-100) in those randomized to CT screening or no screening at 1.5 years follow-up after the first screen. For the EQ-VAS, there was a difference in change scores of 1.39 (95% CI, -0.46 to 3.24; 1.4% increase from screening), and for the SF-12 differences of 1.05 (95% CI, - 0.13 to 2.23; 1.1% increase) and 0.87 (95% CI, -0.58 to 2.32; 0.9% increase) on the physical and mental component scores, respectively. The NLST (n=2,812 (150)) used the SF-36 tool at 6 months’ follow-up after receiving the first screen. For the SF-36 mental component score, a difference in change score of 0.07 (95% CI, -0.61 to 0.74; 0.5% increase from LDCT vs. X-ray screening) was found, whereas for the physical component score there was a difference in change score of 0.50 (95% CI -0.06, 1.07; 0.1% increase). In summary, both trials found little-to-no difference in HRQoL between trial arms over time based on our thresholds of 5% to <10% as a small, and 10% to <20% as a moderate difference between groups. The evidence was of very low certainty based on serious concerns of risk of bias and very serious concerns about indirectness. The trial arms were no longer randomized for these analyses, with the outcome measured among a small fraction (∼10-20%) of the original randomized groups. The indirectness resulted from the outcomes being measured after only one round of screening rather than across 3-4 rounds of screening (and into follow-up) as was desired, and the NLST used an active screening control rather than a no screening comparator.

##### Psychosocial harms

###### Participating in a lung cancer screening program

We first compared the psychosocial harm across the screening process between screening and control arms. Three studies (N=9,205) were included, with two using randomized trial arms (146, 176) and one using a subsample of the original randomized groups (181). All studies reported on differences between groups before and after the first round of screening was completed (e.g., roughly 1 year after baseline) and all participants knew their screening results. One trial (146) also reported on differences between groups at a shorter time-frame (only 2 weeks after screening results were known), and another (176) reported differences between groups before screening at each of 5 rounds of screening. When compared with our thresholds, studies found little-to-no difference in psychosocial harm across several different measurements. The study using the non-randomized subsample was at high risk of bias (181), but results were consistent with the two randomized comparisons only having some concerns of bias. There was moderate-certainty evidence due to serious concerns of indirectness as only one study provided results for greater than 1 round of screening.

###### Undergoing a CT scan (after screening, before results are known)

Two studies (N=1,645 (161, 182)) were included for this outcome. One study at high risk of bias (n=1,014 (161)) comparing a LDCT screening group to an unscreened community sample, matched for age and smoking history, found a small psychosocial harm in lung-cancer worry right after receiving the scan. Within the screening arm in this study (n=631), and in the screening arm of one trial subsample (N=630 (182)), there was little-to-no difference across various measurement tools before and after screening. We determined that undergoing a LDCT scan may cause little-to-no psychosocial harm. There was low-certainty evidence due to serious concerns of risk of bias and inconsistency. Only one study had a no screening comparator (161), and it was at high risk of bias, therefore, we mainly relied on data from pre-post analyses. Additionally, there were inconsistent findings between studies.

###### Positive screen (before repeat LDCT or diagnostic results known)

Seven studies (N=4,794 (66, 134, 146, 150, 175, 181, 186)) were included for this outcome. One study contributed to both a comparison with a non-screening control, and a before-after comparison among screening participants (181). Three studies with a no screening comparator (146, 175, 181), and five studies with a pre-post comparison (66, 134, 150, 181, 186) generally found small psychosocial harm when using lung-cancer specific measurements, whereas other tools were somewhat inconsistent (e.g., State-Trait anxiety subscales). Across all comparisons and tools, we had low certainty evidence that among people 50-74 years of age who are current or former smokers, undergoing a LDCT scan and receiving a positive screening result may cause small psychosocial harm. There were serious concerns of risk of bias as all studies had some concerns of bias. Additionally, there was inconsistency in findings between measurement tools.

###### Positive screen, requiring diagnostic follow-up (before diagnostic results known)

Psychosocial harm of a positive screening result, before receiving diagnostic work-up (excluding those only requiring recall for repeat LDCT) was evaluated in two studies (N=1,651 (66, 146)). One study (n=1,627 (146)) that used an unscreened comparator found moderate psychosocial harm (i.e., 18% increase in cancer worry) from a positive screen. An additional pre-post study (n=24 (66)) used three different tools, with two suggesting small psychosocial harm and the other suggesting moderate harm. For individuals 50-74 years of age who are current or former smokers, receiving a positive screening result requiring diagnostic follow-up probably causes a small, and may cause moderate psychosocial harm. We had moderate certainty for small psychosocial harm, due to risk of bias concerns, and low certainty for moderate psychosocial harm where there were additional concerns about inconsistency.

###### False positive screening result (≤6 months after diagnostic results known)

The impact of a FP on psychosocial harm was evaluated in six studies (N=1,860 (66, 134, 150, 161, 175, 186)) within 6 months from the notification of benign results. Two studies (N=791 (161, 175)) compared the findings for these participants to an unscreened comparator group. One of these studies also reported on a pre-post comparison in the screening arm (n=131 (161)), along with four other studies (N=1,069 (66, 134, 150, 186)). Across all comparisons, there was generally little-to-no psychosocial harm. There was low certainty evidence that receiving a FP screening result may cause little-to-no psychosocial harm within 6 months after the diagnostic results are known. There were serious concerns about risk of bias as four of the six studies (including one of the two comparative studies) were at high risk of bias (134, 151, 161, 186), and the majority of data came from pre-post analyses. There were also serious concerns about inconsistency as one study (161) found moderate psychosocial harm in a comparative analysis, and two studies (161, 186) found small or moderate psychosocial harm in pre-post analysis that disagreed with all other studies.

###### False positive screening result (≥6 months after diagnostic results known)

Six studies (N=4,553 (61, 66, 134, 146, 175, 181)) were included for psychosocial harm of a FP at 6 or more months after diagnostic results were known. One study had both a comparative and pre-post analysis (181), three studies had only a comparison with an unscreened control (61, 146, 175), and two only had a pre-post analysis (66, 134). Findings across all studies and comparisons consistently showed little-to-no psychosocial harm. The evidence was at moderate certainty because two of four comparative studies were at high risk (146, 181), and all other studies had some concerns of bias.

#### Targeted screening strategies

Two trials used a targeted strategy for lung-cancer screening (Table S2.1 (131, 133)). The ECLS trial (n=12,209 (133)) enrolled participants with a mean age of 60.5 years in Scotland. Intervention arm participants were randomized to receive an earlyCDT-Lung test (an enzyme-linked immunosorbent assay that measures 7 autoantibodies), and if positive they received a chest X-ray and LDCT scan. Screening continued for 5 rounds at 6-month intervals. The control arm received standard clinical care and both arms received smoking cessation advice. Follow-up time was up to 5 years. The LungSEARCH trial (n=1,568 (131)) recruited participants with mild chronic-obstructive pulmonary disease (COPD) from ten sites across the United Kingdom, with a mean age of 63 years. The intervention arm received annual sputum cytology and cytometry, and those who had a positive test received both LDCT and autofluorescence bronchoscopy. The control arm received usual care and an exit chest X-ray after 5 years from randomization if no lung cancer developed.

Both trials reported on lung-cancer and all-cause mortality. Neither trial was at high risk of bias for mortality outcomes (Table S2.3). Both trials had some concerns of bias due to insufficient reporting of contamination, and unblinded outcome measurement specifically for lung-cancer mortality. The LungSEARCH trial published mortality outcome data at a median of 5 years follow-up, and has not yet had sufficient follow-up to publish data for their pre-specified timepoint of 15 years.

##### Lung-cancer and all-cause mortality

Results from the two trials were not pooled due to differences in their screening strategies. From the ECLS trial data (133), we were very uncertain about the effects of this targeted screening strategy on lung-cancer mortality (Table S2.12). Absolute findings of 2.7 fewer per 1,000 (95% CI, 5.9 fewer to 1.6 more) were considered very indirect from an insufficient follow-up time of 5 years, and very imprecise as the upper limit of the 95% CI crossed both thresholds of fewer and more deaths. We were also very uncertain about the effects of screening on all-cause mortality (7.0 fewer per 1,000 [14.6 fewer to 1.9 more]) for the same reasons.

Evidence from the LungSEARCH trial (131) was also very uncertain for both outcomes (Table S2.12). Participants were only followed for a median of 5 years follow-up and the trial included only patients with COPD, so there were very serious concerns of indirectness. There was also very serious imprecision for both lung-cancer mortality (6.4 fewer per 1,000 [95% CI, 16.1 fewer to 12.1 more]) and all-cause mortality (33.1 fewer per 1,000 [95% CI 56.4 fewer to 3.7 fewer]), with the inadequate sample size a major consideration for the latter outcome (192)).

Per protocol, as benefit outcomes had very low certainty, the harm outcomes were not explored.

##### Patient preferences (KQ2), disutilities

###### Study characteristics

We included 33 studies reporting HSUVs (Table S3.1 in Additional file 3) (134, 193–224). Age and sex were reported in 23 studies. The mean age across studies was 63.1 years and the average proportion of male participants was 54%. Race and ethnicity were rarely reported especially for the sample we used for our analyses, but the majority of participants were Caucasian (average across 8 studies was 75.5%). The total number of participants was 42,219 (range 24 to 11,772). Eight studies came from multiple countries (196, 200, 201, 207, 213, 216, 223, 224), four from Canada (134, 205, 214, 221), four from the United States (195, 199, 212, 220), four from Japan (204, 206, 218, 219), two from Denmark (193, 202), two from Italy (194, 198), two from Turkey (197, 222), two from the United Kingdom (208, 211) and one study each from Australia (215), Finland (217), Germany (210), South Korea (203), and Thailand (209). Twenty-seven studies used the EQ-5D index score (134, 193, 194, 196–202, 204–210, 212, 214, 216, 218–224), two used the SF-6D (195, 215), and one used the 15D tool (217), all using patient samples. Three studies used direct methods, two with the TTO (211, 213), and one with the SG (203), using vignettes in public samples.

The summary of findings and certainty of evidence for all outcomes are reported in Table 3. A more detailed synthesis of the evidence, including forest plots, can be found in Additional file 3. The majority of studies reported on one or more health states before, during, or after first-line treatment among those with cancer, whereas only three reported on screening-related health states. Disutilties (range 0-1.0) of 0.04 to <0.08 were considered small but important and ≥0.08 were considered moderate.

Ten studies were at high risk of bias for at least one outcome of interest, largely due to concerns of attrition/missing data and/or not reporting a measure of variance (Table S3.2). Studies without a measure of variance had a standard deviation imputed from other included studies, prioritizing studies using similar tools within the same analysis, and were subject to sensitivity analysis. Four studies were at low risk of bias, and the rest had some concerns of bias.

###### Unaffected screen-eligible comparator

To calculate screening, diagnostic, and treatment disutilities, we required a measure of utility from unscreened but screen-eligible (i.e., unaffected) people. Across our outcomes, we prioritized within-study disutility measures (based on an unaffected group within the study). To supplement this data, we allowed for between-study (indirect) estimates of the disutility for an outcome by subtracting a reported utility value (or a pooled utility estimate) for an outcome from a pooled utility of an unaffected population from other studies. Five studies of unaffected populations were used. Using the EQ-5D tool, two studies measured utilities in screen-eligible individuals prior to receiving a LDCT scan (134, 212). The SF-6D was used in one study to evaluate a sample of NLST trial participants before LDCT screening (195). An additional study used the SF-6D to provide utilities based on five different eligibility criteria for screening (215). Finally, the 15D tool was used in a general population sample within a study also measuring treatment utilities (217).

We took an unweighted average across these five studies (N=23,466), and calculated a pooled utility of 0.81 (range 0.74 to 0.91) for the unaffected population. We had low certainty about this estimate. There were serious concerns that the measured utilities were inconsistent across different tools, especially the EQ-5D that had the highest scores (0.87). There was also serious indirectness as three of the five studies used individuals enrolled in lung-cancer screening studies, rather than a general eligible population. Two studies were at high risk of bias, though sensitivity analysis had similar results (pooled utility 0.82). If we chose to use solely the EQ-5D data (to better match the tool used by a large majority of the studies for each health state), the estimate would be much higher than available norms in Canada (0.82 across individuals 45 to 75 years in Alberta (225)), therefore we used the pooled utility of 0.81.

##### Screening health states

###### Disutility of a positive screening result (results not known, among those without cancer)

Two studies (N=263 (134, 212)) used the EQ-5D to calculate the disutility of receiving a positive CT screening test using a within-study comparator (0.01 (95% CI, -0.02 to 0.03)). Neither study was at high risk of bias for this comparison. For between-study analysis, consistent findings were found from these studies (134, 212) when pooled with a study using the SG (n=515 (203)), resulting in an estimated disutility of -0.03 (95% CI, -0.05 to 0.00) (the negative sign indicates a utility rather than disutility). Two studies were at high risk of bias for this comparison (134, 203), and one small study was not at high risk (n=24 (212)).

Excluding results from using the SG or high risk of bias studies in the between-study comparison did not impact overall conclusions. The study using the SG examined utilities by age and sex, with similar findings between groups. Overall, we found that there is probably little-to-no disutility (0.01) from receiving a positive screening result. We had moderate certainty of evidence due to imprecision. The within-study disutilities were based on small samples, and the lower bound of the 95% CI for the between-study disutility indicates a possible small, but important difference.

###### Disutility of a false-positive result (results known)

One within-study comparison with the EQ-5D tool (n=246 (134)) found that the disutility of receiving a FP result 1 and 12 months after screening were 0.00 (95% CI, -0.03 to 0.04), and -0.01 (95% CI, -0.05 to 0.02), respectively. At the 1-month timepoint the study was at high risk of bias, but results were consistent with findings at the 12-month follow-up not at the same risk. For between-study analysis, two studies (N=371 (134, 211)) found that the disutility of receiving a FP result up to 1 month after screening was -0.05 (95% CI, -0.07 to -0.03). Similar findings were observed when excluding the one study using TTO (211), or when removing the EQ-5D data at high risk of bias (134). We found low-certainty evidence that there may be little-to-no (0.00) disutility from knowledge about a FP result. We rated down for indirectness because about 12% of the sample in the within-study analysis had lung cancer. We also rated down for imprecision as the within-study analysis included only a single study and at both timepoints one of the limits of the 95% CI crossed a threshold (in one case for an important utility [-0.05]). There were no serious concerns of inconsistency as both the within-study and between-study analyses found no disutility.

##### Lung-cancer diagnosis

###### Disutility of lung cancer at diagnosis/treatment naïve, stage I-IIIA

Nine studies contributed to a between-study disutility estimate for stage I-IIIA lung cancer at diagnosis (N=1,572 (193, 202–205, 208, 216, 218, 220)). We focused on the eight studies using the EQ-5D, as subgroup analysis by utility tool found substantially different results when including the SG data (p<0.00001), which was also at high risk of bias. Using the seven EQ-5D studies (N=1,057; pooled utility 0.82 [95% CI, 0.77 to 0.87] (193, 202, 204, 205, 208, 216, 218, 220)) the estimated disutility was -0.01 (95% CI, -0.06 to 0.04). One EQ-5D study was at high risk of bias (193), but its exclusion gave similar results. Findings for stage I, I-II, and I-IIIA yielded similar utility measurements. Subgroup findings for age and sex in one study did not show any differences (203). Findings of little-to-no disutility (-0.01) for stage I-IIIA cancer at diagnosis were low certainty from serious concerns of indirectness, as we did not have any within-study disutility measurement, and for imprecision as both limits of the 95% CI crossed our thresholds for either a small utility and disutility, respectively.

###### Disutility of lung cancer at diagnosis/treatment naïve, stage IIIB-IV

Nine studies using the EQ-5D (N=1,728 (197, 198, 200, 205, 206, 220, 221, 223, 224)), and one study using the SG (n=515 (203)) were included for calculating a between-study disutility of stage IIIB-IV lung cancer at diagnosis. The study using a SG was not pooled with EQ-5D studies due to large and statistically significant differences in findings (p<0.00001); the estimated disutility using the SG was 0.46. The pooled EQ-5D utility was 0.74 (95% CI, 0.67 to 0.80) and the estimated disutility was 0.07 (95% CI, 0.01 to 0.14). Sensitivity analysis removing the high risk of bias study (200) yielded nearly identical results. Subgroup analysis based on specific stages of disease resulted in nonsignificant findings (p=0.44) between stage IIIB-IV (N=4 studies; N=662) and stage IV alone (N=5 studies; N=1,066), but with some potential for stage IIIB-IV cancer to have more disutility (0.10 vs. 0.05 for stage IV alone). EQ-5D utilities in subgroup analysis by age, gender and smoking status were only reported in one study (n=266 (197)). We found that there may be a small, but important disutility (0.07) for stage IIIB-IV lung cancer at diagnosis. We rated down for indirectness as we did not have any within-study disutilities to compare to the between-study analysis, and for imprecision as the lower bound of the confidence interval crossed our threshold.

##### Treated lung cancer

###### Disutility of lung cancer, on first-line adjuvant treatment (none only receiving surgery), stage I-IIIA

This disutility was evaluated in three studies with the EQ-5D tool (N=609 (199, 205, 220)). None of the studies had a within-study comparison. One study was at high risk of bias (199), but findings were similar in sensitivity analysis. Using the pooled utility value (0.77 [95% CI, 0.73 to 0.80]), the estimated disutility was 0.05 (95% CI, 0.02 to 0.08); therefore, there may be a small, but important disutility (0.05) for stage I-IIIA lung cancer during first-line treatment. We rated down for indirectness, as there were no within-study measurements, and for imprecision as the 95% CI included little-to-no disutility. Findings from a between-study subgroup by stage I-II versus I-IIIA studies were statistically different, but the comparison only used a single study for stage I-II, and it is not reported how many stage IIIA lung cancers were in the two studies including stage I-IIIA.

###### Disutility of lung cancer, on first-line adjuvant treatment, stage IIIB-IV

The disutility of stage IIIB-IV lung cancer during first-line adjuvant treatment was assessed using between-study findings with 13 studies (N=2,635 (196, 198, 200, 201, 205, 206, 209, 210, 213, 220–222, 224)). One study reported utilities using a TTO (n=300 (213)), and 12 studies used the EQ-5D (N=2,335 (196, 198, 200, 201, 205, 206, 209, 210, 220–222, 224)). Four studies were at high risk of bias (200, 213, 222, 224), including the study using TTO, but this did not impact conclusions. Based on the pooled estimate of 0.72 (95% CI, 0.67 to 0.76), we calculated a disutility of 0.09 (95% CI, 0.05 to 0.14) and judged that there is probably a small but important disutility (0.04), and may be a moderate disutility (0.09) for stage IIIB-IV lung cancer during first-line treatment. We rated down once for a small disutility for indirectness as there were no within-study comparisons. We also rated down a moderate disutility for inconsistency which was not explained by stage of disease or measurement tool.

###### Disutility of lung cancer, after first-line treatment completion and non-progressive (post-surgery, on maintenance therapy, or survivors), stage I-IIIA

One study using the 15D (n=230 (217)), was used for both within- and between-study analyses, and ten studies using the EQ-5D (N=13,618 (193, 194, 202, 204, 208, 214, 216, 218–220)) were added to the between-study analysis. A majority of studies measured the outcome within one year after treatment. The within-study comparison found a disutility of 0.03 (95% CI, 0.01 to 0.04) (217). This was the same as the estimate we found (0.03 [95% CI, -0.02 to 0.08]) when estimating the disutility from a pooled utility from the nine studies (0.78 [95% CI, 0.73 to 0.83]). No studies in this analysis were at high risk of bias, and no impact on the findings were found when excluding the 15D study from the between-study analysis. Subgroup findings based on cancer stage suggested more disutility for lower stages, but findings were not significant. We judged that there may be little-to-no disutility (0.03) for stage I-IIIA lung cancer after first-line treatment completion without progression. We rated down for inconsistency that was not well explained by measurement tool or subgroup findings by treatment type (e.g., measured right after surgery vs. after adjuvant therapies) or stage of disease. We also rated down for indirectness as there was only one within-study comparison.

###### Disutility of lung cancer, after first-line treatment completion and non-progressive (on maintenance therapy, or survivors), stage IIIB-IV

For this health state we included eight studies (N=1,158 (198, 200, 207, 209, 213, 214, 220, 224)) and used between-study analysis. Seven studies (N=1,083 (198, 200, 207, 209, 214, 220, 224)) used the EQ-5D tool, and one used a TTO (n=75 (213)). There were five studies at high risk of bias, including four with the EQ-5D and one with the TTO (198, 200, 207, 213, 224). Between-study findings based on stage of disease were limited as most studies only including stage IV patients. The pooled utility was 0.75 (95% CI, 0.71 to 0.78) and estimated disutility was 0.06 (95% CI 0.03 to 0.10) indicating there may be a small, but important disutility (0.06) for stage IIIB-IV after first-line treatment without progression. Sensitivity analysis excluding the five high risk of bias studies resulted in similar findings, as did removing only the TTO study. We rated down for both indirectness, because there was no within-study data, and imprecision as the lower limit of the 95% CI crossed suggested little-to-no disutility.

#### Patient preferences (KQ2), other preference data

##### Study characteristics

We included 26 studies (89, 226–251) (with one associated report (252)). Twenty-two studies were from the United States, and one each was from Australia (238), the United Kingdom (245), Italy (246), and multiple European countries (249). Across studies, the mean age was 63.3 years (range 58.0 to 67.7) and percent female 44.4% (3 studies having few females). Eighteen studies enrolled people at least 55 years of age, one limited enrollment to those age 60 years an older, and the other seven enrolled people at least 45 or 50 years old; 74 to 80 years was the typical upper age limit for participation. Among 20 studies reporting on race or ethnicity, the percent Caucasian was on average 59% (range 0-91). One study only enrolled African Americans (235), in four others a majority of participants were Black (234, 241, 247, 248), and in 16 studies there were at least 15% Black participants. No study focused on or had more than 3% Indigenous people (American Indian or Alaskan Native in the US studies). Only three studies reported on family history of lung cancer, none reported lung-cancer screening history (though this was assumed to be low), and six studies excluded those with a recent chest CT. Most studies only included people eligible for screening (typically via the criteria of the 2013 recommendations by the USPSTF criteria, of aged 55-80 years with a 30-pack year history and smoking within the last 15 years). In two studies (231, 233) we needed to rely on data from a sample of participants where some were ineligible (i.e., participants were aware of their ineligibility), though this was considered a risk of bias. Sample sizes contributing data of interest to this review ranged from 11 to 4246 (median 178, three with n≤30), though this does not include participants in study groups receiving usual care (e.g., brief explanations of screening without any numerical information (243)) because our review did not focus on the effectiveness of decision aids or other materials but rather the views of people after receiving detailed risk information.

Seven studies provided direct, preference-based data (226–228, 231, 233, 237, 243) and 25 indirectly provided inferences on the importance of the benefits versus harms via attitudes, intentions and screening uptake (six also provided direct data). Study samples were classified as i) community/public (e.g., public survey panels; n=6), ii) attenders of primary/secondary care (one clearly defined as primary care; n=7), iii) selective (e.g., patients of tobacco treatment programs, smoking quit-line participants; n=7), and iv) screening program attendees (n=6). The latter group was considered at high risk of bias, as they had already been referred to the screening program by a primary care physician which would have in many cases already involved a positive attitude by the patient and a physician’s recommendation for screening. Clinical input determined that the other three categories were reasonably applicable to the target population.

When grouping studies by the net benefit (usually after 3 screens and 7 years’ follow-up) portrayed by decision aids or other materials, a low net-benefit screening scenario provided data for an estimated 2-5 fewer lung-cancer deaths per 1000 screened, and numerical risks for FPs (approximately 250-350 per 1000), FPs requiring biopsy (14-25 in 1000), complications from invasive procedures (1-3 per 1000), and overdiagnosis (4 in 1000 screened, 10% of screen-detected cancers). Moderate net-benefit scenarios mentioned the same outcomes but did not report specific risks (but rather only some textual information of their possibility) for either, or both, complications or overdiagnosis. High net-benefit scenarios presented benefits either only in relative terms (e.g., 20% reduction in lung-cancer mortality) or as varying based on one’s personal risk for cancer (usually using modeling data incorporated into ShouldIScreen decision aid (253)). In this scenario a majority of participants were assumed to have been told there could be at least 6 (and up to 20) fewer deaths per 1000 screened based on the reported mean PLCO_M2012_ 6-year predicted risk for cancer or the participant characteristics. The harms data in the high net-benefit scenarios were quite similar to those in the low and moderate scenarios, without any reduction in risk for one or more outcomes (e.g., FPs and associated harms (160)) for those at increased risk for cancer. Across all studies, only two exposures mentioned all-cause mortality (indicating a reduction) and few reported numeric risks for incidental findings or portrayed these as having any possibility of harm. When more than one exposure in a study met our criteria and was assigned to the same grouping of net benefit, we treated the groups as one sample for our purposes. Tables S3.4 (direct data) and S3.5 (indirect data) contain study characteristics and all study results.

Risk of bias assessments by outcome and study are documented in Table S3.6 and summarized within the findings for each outcome. Many issues related to screening or study outcomes being inadequately defined (e.g., FPs portrayed as being told one has cancer, screening uptake measured at <6 months in studies that would have required a subsequent referral and appointment, benefits only described for LDCT in comparison with chest X-ray), and when the interventions were not extensively tested for understanding or findings from knowledge tests indicated the participants had poor knowledge of the risk information or outcome (e.g., 30% showing conceptual understanding of overdiagnosis (238)). We only assigned an item at high risk of bias if the issue may have impacted findings substantially; for example, if intentions were at the lower range in category, such as 52%, and the study’s poor knowledge scores were shown to positively associate with preferences (240)). Two studies where materials mentioned that the “USPSTF recommends screening” or provided an invitation letter for screening signed by the participant’s primary care physician were also considered at risk for bias. Lastly, as mentioned previously, data from samples attending screening programs or in mixed samples with a substantial portion not eligible for screening were at high risk of selection bias.

##### Direct comparisons between outcomes via preference-based data

Five of the included seven studies presented participants with decision aids and examined preferences between various benefits and harms (n=2 among community/public samples) (227, 228) or different harms (n=3 among primary care and select populations) (233, 237, 243) via ranking, ratings, or other scales measuring their relative importance. Another study evaluated three formats for presenting the same risks of lung-cancer mortality and FPs (231), whereas another employed a contingent valuation exercise where lung-cancer mortality and FPs were used as two of the screening program attributes (226). Most comparisons used data from at least two studies with broadly applicable samples. Table 5 summarizes the findings and certainty of evidence from preference-based studies where specific outcomes of interest were compared.

**Table 5.** Summary of findings for other preference data, direct comparisons between outcomes.

| Included studies;<br>Sample size | Certainty* | What does the evidence say? |
| --- | --- | --- |
| **Lung-cancer mortality vs. false positives** |  |  |
| 3 studies; N=821 | **LOW<sup>b,c</sup>**<br>⊕⊕⊖⊖ | Among patients eligible for lung-cancer screening, a small majority (51-75%) may accept at least 69-122 FPs per lung-cancer death prevented. |
| **Lung cancer mortality vs. overdiagnosis** |  |  |
| 2 studies; N=567 | **LOW<sup>b,c</sup>**<br>⊕⊕⊖⊖ | Among patients eligible for lung-cancer screening, a small majority (51-75%) may accept at least 1.3 overdiagnoses per lung-cancer death prevented. |
| **Lung cancer mortality vs. any harm** |  |  |
| 2 studies; N=567 | **LOW<sup>b,c</sup>**<br>⊕⊕⊖⊖ | Among patients eligible for lung-cancer screening, a small majority (>50-75%) may think that the reduction in lung-cancer mortality outweighs the risk of experiencing one of the harms.<br>It is unclear how people would rate the outcomes when assuming people could experience multiple harm outcomes during their participation in screening. |
| **False positives vs. overdiagnosis** |  |  |
| 4 studies; N=801 | **LOW<sup>b,c</sup>**<br>⊕⊕⊖⊖ | Among patients eligible for lung-cancer screening, a small majority (>50-75%) may think that a risk of 50-100 FPs is of similar importance to that of 1 overdiagnosed case. |
| **False positives vs. psychosocial harms from undergoing screening** |  |  |
| 1 study; n=384 | **VERY LOW<sup>a,c</sup>**<br>⊕⊖⊖⊖ | The evidence is uncertain about the relative importance of FPs versus the psychosocial harms from undergoing screening. |

\*Reasons for rating down certainty: a=risk of bias, b=inconsistency, c=indirectness, d=imprecision; capital letters indicate very serious concerns. Explanations are within the text.

###### Lung-cancer mortality versus false positives

Our analysis focused on data from three studies. One high risk of bias study (n=254; 44% never smokers) examining different risk formats to present FPs and lung-cancer mortality among outpatients of a pulmonary practice found that a large majority (86%) endorsed screening that would result in 91 FPs per lung-cancer death prevented (231). Two other studies (N=567) employed outcome rating (scale 1-5 with 1=not very important, 5=very important) and ranking in public surveys based on data indicating either 206 (227) or 365 (228) FPs and 3 fewer lung-cancer deaths per 1000 screened (i.e., 69 and 122 FPs per prevented death). There was some inconsistency between studies in rating data, with one showing a preference by at least a small majority of people for lung-cancer mortality (4.3 [SD 1.0] vs. 3.3 [SD 1.2]) (228) but one showing that FPs were preferred by at least some (4.2 [SD 1.0] vs. 3.7 [SD 1.3]) (227). In both studies, ranking data indicated a preference for lung-cancer mortality with this outcome being ranked first (among 6 outcomes) by 52% and 59% of participants whereas the proportions for FPs were 7.3% and 13.3%. In the study also reporting mean scores, the mean rank (lower being more important) was 2.5 (SD 1.9) for lung-cancer mortality and 3.7 (SD 1.7) for FPs which again indicated a preference for the benefit by at least a small majority (51-75%) (227). We judged findings across the studies to indicate that a small majority of people may think at least 69-122 FPs is acceptable to prevent one lung-cancer death. Alternatively, a small minority (>25-50%) may not find at least 69-122 FPs acceptable. An upper limit for how many FPs may be acceptable is unknown, as trade-offs were not directly elicited. The certainty of the evidence was low due to inconsistency in findings between studies and indirectness from reliance in two studies on data which may have been confounded by ratings/rankings of other outcomes presented and because explicit trade-off methodology was not employed in any of the studies. The study not contributing to our conclusions (n=210) employed a conjoint analysis experiment and reported slightly higher importance for lung-cancer mortality than for FPs (scores of 17.2 and 15.8, respectively, with a sum of 100 for each individual when rating five attributes being considered) (226). These findings are difficult to interpret considering that the levels for FPs (10%, 25% and 40% of those screened) used absolute risk whereas those for lung-cancer mortality (1%, 10%, and 20% reduction) used relative risks.

###### Lung-cancer mortality versus overdiagnosis

Rating and ranking data from the above mentioned two studies (227, 228) using public surveys were also used to determine the relative importance between lung-cancer mortality and overdiagnosis (3 fewer and 4 in 1000; ratio 1:1.3). There was some inconsistency in ratings of importance, where overdiagnosis had higher ratings of importance (4.2 on scale 1-5) than lung-cancer mortality (3.7) in one study (227) but not the other (3.6 vs. 4.3) (228). Lung-cancer mortality had the highest proportion of people ranking it as most important in both studies (52% and 59% vs. 6.3% and 6.8%) and in the study reporting mean ranks these were lower (indicating more importance) for mortality (2.5) than overdiagnosis (3.7) (227). We judged findings across the studies to indicate that a small majority of people may think at least 1.3 cases of overdiagnosis is acceptable to prevent one lung-cancer death. An upper limit for how many overdiagnosis may be acceptable for this proportion is unknown, as trade-offs were not directly elicited. As for the comparison with FPs, the certainty of the evidence is low due to inconsistency in findings and indirectness.

###### Lung-cancer mortality versus any harm

Rating and ranking data from the above mentioned two studies (227, 228) using public surveys was also used to determine the relative importance between lung-cancer mortality (3 fewer per 1000) and experiencing any harm of interest (206 or 365 FPs, 14 or 25 in 1000 FPs requiring biopsy, 4 in 1000 overdiagnosis, 3 in 1000 major complications from invasive procedures, and 200 in 1000 incidental findings [last 2 outcomes only measured in one study each]). There was some inconsistency in ratings of importance; each of the harms presented had higher ratings of importance (4.0-4.2 on scale 1-5) than lung-cancer mortality (3.7) in one study (presenting incidental findings), but not the other (3.3-3.7 vs. 4.3; presenting complications). Lung-cancer mortality had the highest proportion of people ranking it as most important in both studies (52% and 59% vs. 5.8-13% and 5.9-7.3% for other harms) and in the study reporting mean ranks these were lower (indicating more importance) for mortality (2.5) than all harms (3.2-3.7) (227). Across the four harms presented in each study, ratings and rankings were quite similar among the harms and between FPs and the other harms. One of these studies reported that Black/African-American respondents were more likely to rate overdiagnosis higher than Whites (4.0 vs. 3.5, p=0.02), though their ratings for lung-cancer mortality were not provided to know the relative ratings (228); race did not impact intentions to screen. In both studies, the authors reported that a substantial minority (45% and 34%) of participants ranked one of the harm attributes as most important, even above mortality benefit. Findings indicate that a small majority may think that reducing lung-cancer mortality is more important than experiencing one of the harms, when screening provides a relatively low net-benefit. Our certainty is low due to inconsistency in findings and indirectness from only single studies measuring two of the outcomes of interest and reliance on mean scores (and their variance) to estimate proportions. It is unclear how people would rate the outcomes when assuming people could experience multiple harm outcomes during their participation in screening.

###### Preferences among harms

Rating and ranking data from the public survey studies (N=567) (227, 228) described above indicated that there was similar importance among the harms for at least a small majority of people. Rather than providing conclusions about every possible comparison among harms, we focused on FPs versus overdiagnosis and psychosocial harms because a few additional studies provided data specific to one or more of these comparisons.

There was some inconsistency between the two public survey studies when comparing FPs and overdiagnosis (206 or 365 FPs vs. 4 overdiagnoses per 1000 [50-100 FPs per case of overdiagnosis]). Although ratings for these outcomes were similar between studies (4.2 vs. 4.2 and 3.3 vs. 3.6 on scale 1-5), and the mean ranks in the study reporting these were the same (3.7 for both outcomes), a higher proportion of people ranked FPs as most important in one study (13.3% vs. 5.8%) (227) but not the other (16% vs. 15%) (228). Additionally, two high risk of bias studies (N=234) enrolling select populations measured the relative importance between FPs and overdiagnosis after participants viewed similar decision aids (233, 243). Findings are difficult to interpret because although estimates were provided for FPs (365 per 1000 screened) no numerical information was provided for overdiagnosis (“some people may be treated for cancer that would never have harmed them”). In addition, both studies found that participants had similar scores for FPs and overdiagnosis in level of concern about the outcomes (7.0 [IQR 5] and 6.0 [IQR 5] on an 11-point scale (233); 81.4% and 83.8% concerned or very concerned (243)). However, when one study (n=204) (243) associated scores of concern for each outcome with screening uptake, concern for FPs associated with higher uptake (odds ratio 2.25 [0.65, 7.78]) but concern for overdiagnosis largely associated with lower uptake (e.g., odds ratio 0.15 [0.05, 0.53]), suggesting overdiagnosis may be more important for some people. Our conclusions relied mostly on findings from the public survey samples. A finding of a slight majority thinking the presented risks for these outcomes are of similar importance is considered to have low certainty due to inconsistency and indirectness from reliance on textual information for overdiagnosis (in two studies), on mean scores (and their variance) to estimate proportions (in two others), and because explicit trade-off methodology was not employed in any of the studies.

We used data from one study (n=348 among veterans with a majority being Caucasian [91%] and former smokers [56%]) at high risk of bias to compare the importance of FPs (365 per 1000 presented) with psychosocial harms from undergoing screening (“anxiety waiting for LDCT results”) (237). A similar proportion of people (30.8% vs. 28.0%) reported that these outcomes were very or extremely important for decision making. It is difficult to interpret the findings because while the survey asked about anxiety from waiting on the LDCT findings, the decision aid focused on the experience of waiting for a resolution of a suspicious finding. This implies that the concern was more related to expectations about the consequences of a positive screen (i.e., FP or cancer diagnosis) rather than waiting for the screening results. We judged that the findings about the importance of a FP versus psychosocial harm from undergoing screening are uncertain due to risk of bias and very serious indirectness from an unclear comparison and a single study population that may not apply broadly to the target audience.

##### Indirect data of the relative importance of the potential benefits and harms

Figures 4 and 5 contain harvest plots displaying the data on screening intentions and uptake. In each figure, data from each study (numbered, corresponding to Study# in Table S3.5) is shown as a bar within the category of the study exposure’s net-benefit scenario. The upper panel contains data from all eligible studies, whereas the lower panel represents a sensitivity analysis (“best case” scenario), i) excluding data from studies at high risk of bias for the outcome (including all studies of screening program attendees), and (for intentions), ii) increasing the intentions in one or more study based on the assumption that about 50% of people “unsure” about their intentions would, if forced to decide, prefer screening. Study 19 was designated to have two interventions, a and b, which used the same decision aid but people in one arm (19b) also received assistance for their screening appointment and attendance by a community health worker (234). We present data for each arm, in part to demonstrate how uptake of screening may be affected substantially by factors (e.g., structural barriers to screening) apart from attitudes about screening efficacy which was the intent of this review. Three studies reported on attitudes towards screening and findings are described in Table 6.

**Figure 4.**
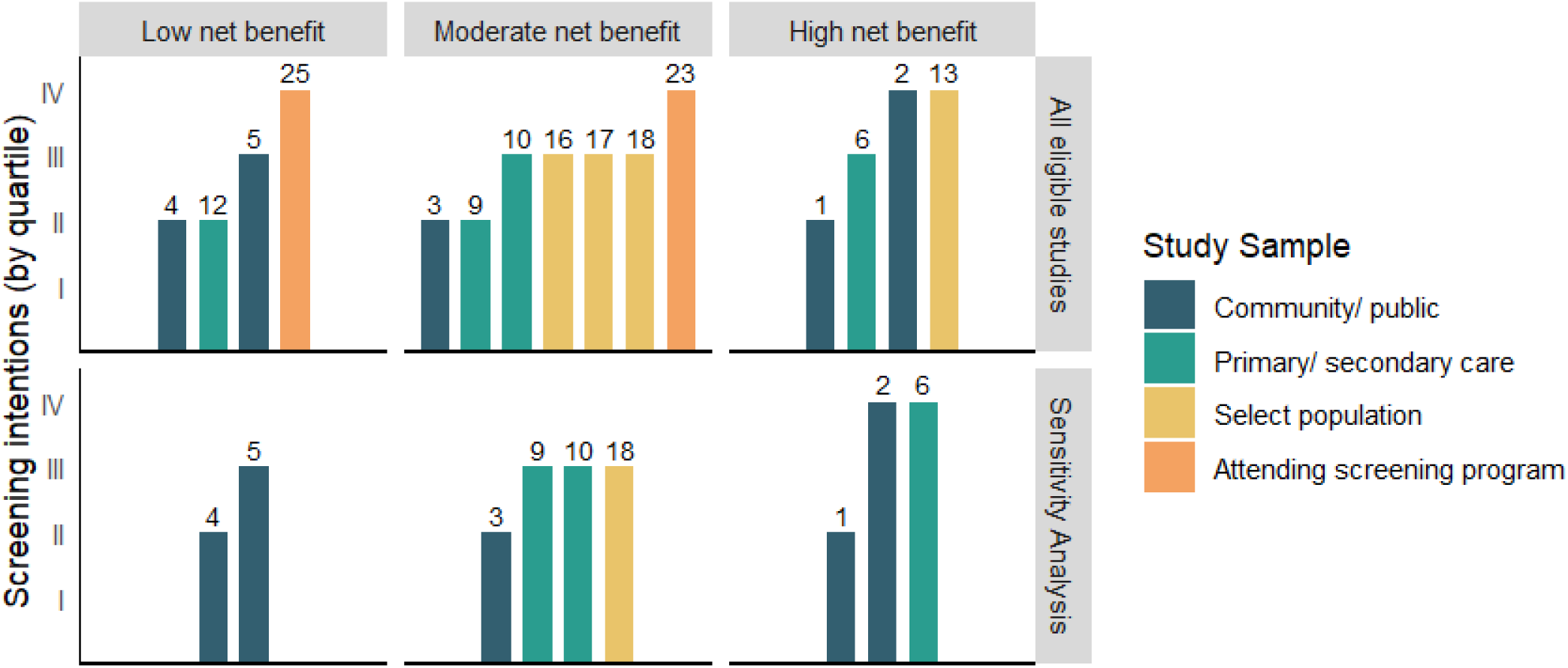
Intentions to screen, by relative net benefit of screening (low, moderate or high) and study sample (i.e., recruitment setting). Numbers represent study ID. Study findings were categorized into quartiles of 0-24% (small minority), 25-50% (large minority), 51-75% (small majority), and >75% (large majority). The lower panel/row removes data at high risk of bias and, for studies 9 and 6, assumes 50% of those reporting “unsure” intentions actually prefer screening. Study sample sizes are not reflected; study 10 is very large (n=4246) and studies 1, 2 and 17 very small n≤30).

**Figure 5.**
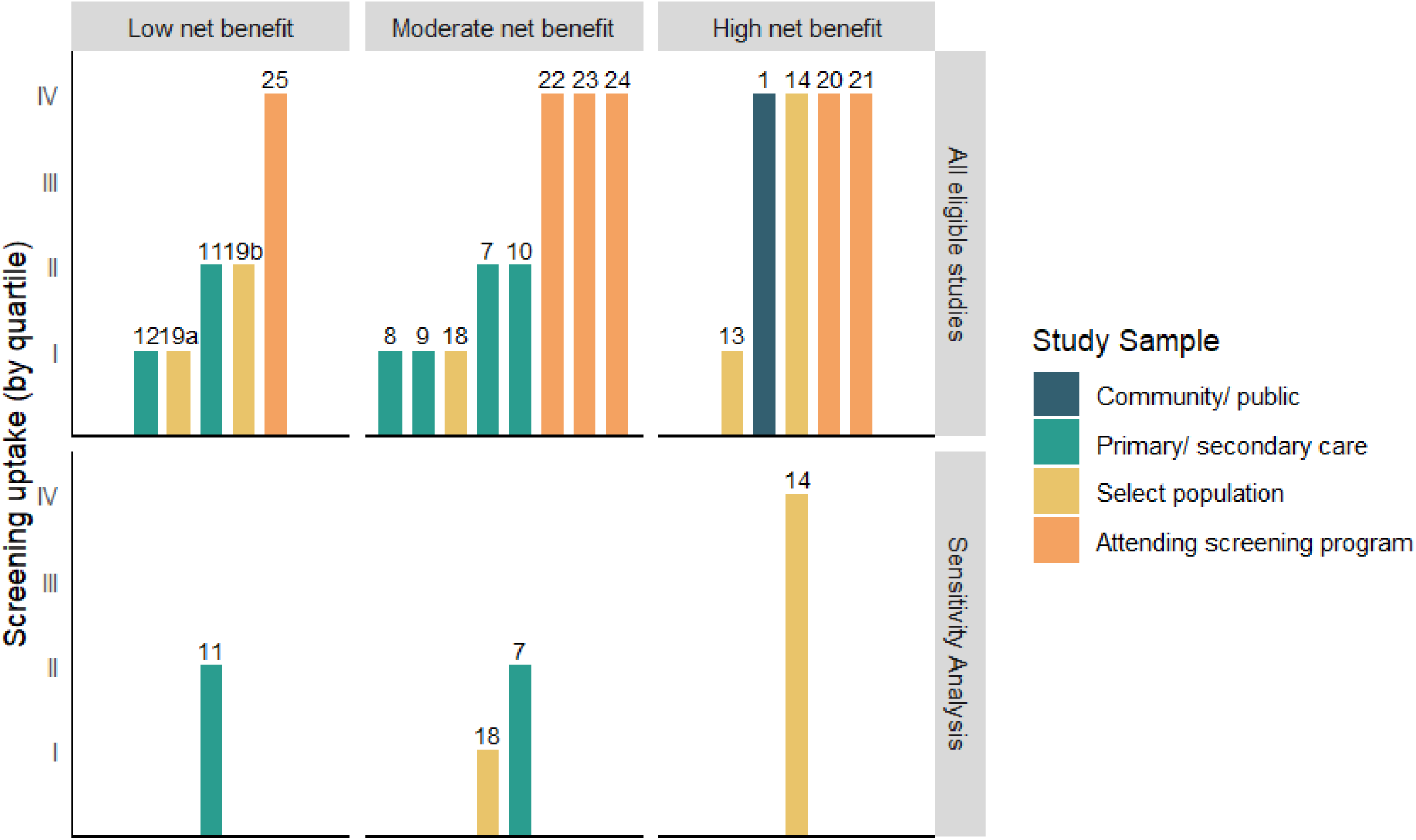
Screening uptake, by relative net benefit of screening (low, moderate or high) and study sample (i.e., recruitment setting). Numbers represent study ID. Study findings were categorized into quartiles of 0-24% (small minority), 25-50% (large minority), 51-75% (small majority), and >75% (large majority). The lower panel/row removes data at high risk of bias. Study sample sizes are not reflected; study 10 is very large (n=4246) and study 1 and 19b very small (n≤30).

**Table 6.** Summary of findings from other preference studies, indirect comparisons.

| Included studies;<br>Studies contributing most to<br>analysis (All studies) | Findings | Certainty* | What does the evidence say? |
| --- | --- | --- | --- |
| **Relatively low net-benefit scenario** |  |  |  |
| Attitudes: 0 studies<br>Intentions: 2 studies, N=567 (4 studies, N=1,267)<br>Uptake: 1 study, n=69 (4 studies, N=821) | Intentions: Figure 4<br>Uptake: Figure 5 | **LOW<sup>b, c</sup>**<br>⊕⊕⊖⊖ | In a relatively low net-benefit scenario, a large minority (25-50%) of people eligible for screening may weigh the benefits as more important than the harms from screening. |
| **Relatively moderate net-benefit scenario** |  |  |  |
| Attitudes: 1 study, n=64 (2 studies, N=208)<br>Intentions: 4 studies, N=6,034 (7 studies, 6,378)<br>Uptake: 2 studies, N=301 (8 studies, N=5,728) | Attitudes: Among 64 patients of primary care practices receiving telephone SDM, 81% had preference scores 0.51-1.0 (scale 0-1) (Study 7 in Fig 5; low risk of bias). In another study, 90% of 144 patients seeking smoking cessation services reported being “quite or a lot” in favor of screening (Study 15; high risk of bias).<br>Intentions: Figure 4<br>Uptake: Figure 5 | **MODERATE<sup>c</sup>**<br>⊕⊕⊕⊖ | In a relatively moderate net-benefit scenario, a small majority (51-75%) of people eligible for screening probably weigh the benefits as more important than the harms from screening. |
| **Relatively high net-benefit scenario** |  |  |  |
| Attitudes: 1 study, n=11 (1 study, n=11)<br>Intentions: 3 studies, N=62 (4 studies, N=208)<br>Uptake: 1 study, n=137 (5 studies, N=1,554) | Attitudes: In one study of 11 volunteers at a University (63% with at least college education) who were eligible for screening, the mean score on a 10-point scale about the effectiveness of high net-benefit screening was 8.09 (SD 2.07), indicating that a large majority (>75%) weighed the benefits as more important than the harms (also Study 2 in Figure 4; low risk of bias).<br>Intentions: Figure 4<br>Uptake: Figure 5 | **LOW<sup>c, d</sup>**<br>⊕⊕⊖⊖ | In a relatively high net-benefit scenario, a large majority (>75%) of people eligible for screening may weigh the benefits as more important than the harms from screening. |

\*Reasons for rating down certainty: a=risk of bias, b=inconsistency, c=indirectness, d=imprecision. Explanations are within the text.

For each net-benefit scenario, we used all available results to make narrative statements about the proportion of people (using quartiles) eligible for screening thinking the benefits outweigh the harms. We preferred data on attitudes and intentions over uptake because of the influence on behaviors from costs (particularly in the studies from the US where out-of-pocket costs subsequent to screening were noted in several studies as a consideration), peer/family expectations, recommendations by physicians, and fear of cancer among others. Our conclusions also generally relied on the data from studies not at high risk of bias, particularly as most of these studies included highly selective/inapplicable populations. Table 6 summarizes the findings and certainty of evidence from indirect data, by net-benefit scenario.

###### Low net-benefit scenario

Four studies (two not at high risk of bias (227, 228)) reported intentions, four (one not at high risk of bias (247)) reported uptake, and none reported attitudes about this scenario. In a relatively low net-benefit scenario, a large minority (25-50%) of people eligible for screening may weigh the benefits as more important than the harms from screening (low certainty). Alternatively stated, a small majority (51-74%) may not favor screening. Apart from indirectness of the outcomes, applicable for all outcomes in this set of data (see Data analysis in Methods), there was inconsistency in findings among the two studies (N=567; intentions of 41% (227) and 57% (228) contributing the most to the conclusions.

###### Moderate net-benefit scenario

Seven studies (four not at high risk of bias (89, 230, 243, 249)) reporting on intentions and eight (two not at high risk of bias (243, 250)) on uptake presented screening as having moderate net benefit. Uptake was lower than intentions in three of the studies reporting on both outcomes, which was likely in part due to short follow-up durations, even after including one study not at high risk of bias that only used the minimally acceptable 6-month follow-up. Data on attitudes that contributed to the conclusions showed that among 64 patients of primary care practices receiving telephone shared decision making, 81% had preference scores 0.51-1.0 (scale 0-1) (250). For this relatively higher net-benefit screening scenario, we judged the evidence to have moderate certainty that a small majority (51-75%) of people probably prefer screening (while a large minority [25-50%] likely do not). Indirectness was the only serious concern when relying on the lower risk of bias study results.

###### High net-benefit scenario

Three (N=62) of the four studies reporting on intentions under a high net-benefit scenario were not at high risk of bias (235, 236, 238). Only one (n=137) (242) of the five studies reporting uptake was not at high risk of bias in addition to the single (n=11)(236) study reporting attitudes. For this scenario, we rated the evidence as low certainty for a large majority (>75%) preferring screening. We rated down once for both indirectness and imprecision, due to a small sample (N=210) contributing to the main analysis.

No conclusions could be drawn about any differences in findings based on race or ethnicity. The five studies enrolling at least a majority of Black individuals (studies 1, 11, 13, 19 and 24 in Figures 4 and 5) had issues including very small size (n=15) (235), high risk of bias (234, 241, 248), inconsistent findings (i.e., low preferences in low and high net-benefit scenarios and high preference in moderate net-benefit scenario), and enrollment of individuals from different settings, thus invalidating comparisons with other study findings. As mentioned previously, in study 19, screening uptake was likely less influenced by the relative importance of the outcomes but more from “competing priorities and patient factors (e.g., social barriers to keeping appointments)” (234). Three other studies reported subgroup findings based on race and ethnicity. In a community/public sample (study 5, n=219) with 25% visual minorities, there were no differences between White, Black, Hispanic or Latino, and “other” groups in screening intentions (data not reported) (228). In two other high risk of bias studies among screening program attendees, one (study 21) found no differences in uptake between Black and White participants, but a lower adjusted odds of uptake among a very small sample of Asians (n=13) compared with White participants (245). The other (study 22) found a significantly lower uptake (yet still indicating a large majority) among Black (84%) but not Asian (90%), compared with White (93%) participants (229). Similar to study 19, the authors of study 22 found multiple factors unrelated to screening outcomes of interest (e.g., worries and concerns, social influence and support, susceptibility) that influenced uptake.

Sex or age were not associated with screening intentions (study 5, n=219, ages 55-80 years) (228) or uptake (study 21, n=845, ages 60-75 years) (245) in the two studies reporting on these variables (data not reported). No study reported findings based on risk for cancer or cancer mortality so it is not clear if, within a particular net-benefit scenario, those at higher risk would have different preferences for screening. Overall, we judged that there was likely not a substantial difference by race, age, or sex in terms of outcome preferences and did not rate down our certainty ratings for additional indirectness.

#### Comparative effects of trial-based selection criteria and use of risk prediction models (KQ3a)

All eligible studies re-examined NLST data and were included in subgroup analyses in KQ1. These studies retrospectively applied risk prediction models to baseline characteristics of the participants to evaluate whether differences in risk for lung-cancer incidence (e.g., PLCO_2012_ 6-year risk) or mortality impacted screening effects on lung-cancer mortality (n=4 studies (18, 160, 179, 188)), and FPs (n=2 studies (160, 169)). There was moderately credibility that absolute (but not relative) findings may be considerably higher for those in the highest versus lowest risk quartiles/quintiles (e.g., <1% 6-year risk for cancer), and that there may be little-to-no screening benefit for the lowest risk individuals.

#### Comparative effects of trial-based nodule classification and different nodule classification systems (KQ3b)

##### Study characteristics

There were 15 studies included for this key question (68, 75, 94, 106, 119, 123, 152, 254–261). The total sample size was 1,098,253 (range 256 to 1,052,591) and on average, 59.1% were male (range: 47.5% to 97.9%). Fewer studies reported on average age which was 63.8 (range: 61.0 to 66.0; N=8 studies), and percent Caucasian was 68.2% (23.0% to 99.2%; n=9 studies). The majority of studies came from the United States ((68, 75, 106, 119, 123, 152, 254–256, 258–261)), and one study came from Australia (257), and South Korea (94). One of the two studies with a within-study analysis, and five of the 14 studies used for the between-study analysis were at high risk of bias (Table S4.2). The most common reasons for rating as high risk were attrition, and an unstructured method of ascertaining the outcome. We only identified data on FPs for this KQ. Additional file 4 includes all data used for this KQ.

##### False positives

Two studies provided a within-study comparison of FPs over one round of screening by reanalyzing study data from using NLST nodule management and applying LungRADs nodule management to the LDCT scan results (N=26,968 (257, 259)). For indirect/between-study analysis we compared the proportions of FPs over one round of screening from studies using NLST nodule management (3 studies; N=28,638 (152, 257, 259)) with those in studies focused on LungRADs (14 studies; N=1,102,285 (68, 75, 94, 106, 119, 123, 254–261)).

We rated the certainty of evidence for the comparator used for the between-study analysis (i.e., 300.0 per 1000 [95% CI, 210.0 to 380.0] from use of NLST nodule management) and found that among populations meeting NLST eligibility criteria, using the NLST nodule management criteria probably results in 300 FPs per 1,000 screened once (Table 7). We had moderate certainty after rating down for inconsistency between studies, which also contributed to the width of the 95% CI for which we did not rate down.

**Table 7.** KQ3b Summary of findings for NLST eligible populations: LungRADs versus NLST nodule management criteria, one round of screening.

| Outcome No. participants (studies) | Anticipated absolute effects |  |  | Certainty of evidence (GRADE)* | What happens? |
| --- | --- | --- | --- | --- | --- |
|  | With LungRADs nodule criteria | With NLST nodule criteria | Difference |  |  |
| NLST eligibility and nodule management (for calculating between-study difference in proportions) |  |  |  |  |  |
| Proportion with a false positive at round 1 with NLST nodule management (Fig. S4.2); N=28,638 (3 studies) | Pooled proportion: 300.0 per 1,000 (210.0 to 380.0) |  |  | Moderate<sup>b</sup><br>⊕⊕⊕⊖ | Among populations meeting NLST eligibility criteria, using the NLST nodule management criteria probably results in 300 false positives per 1,000 screened. |
| LungRADs versus NLST nodule management, amongst NLST eligible |  |  |  |  |  |
| Within study analysis: N=26,978 (2 studies)(Fig S4.3) | Within study:<br>RR (95% CI): 0.48 (0.44, 0.51)<br>Proportion with LungRADs: 125.6 (115.2 to 133.5) per 1,000<br>Proportion with NLST: 261.8 per 1,000<br>**Difference in proportions (95% CI): 136.1 fewer (146.6 to 128.3 fewer) per 1,000** |  |  | Moderate<sup>c</sup><br>⊕⊕⊕⊖ | Among populations meeting NLST eligibility criteria, using LungRADs nodule management compared with NLST nodule management probably reduces FPs by about half, though the proportion with FPs will probably still be at least 75 per 1,000 screened. |
| Between study analysis: LungRADs N=1,102,285 (14 studies) (Fig S4.4) versus NLST N=28,638 (3 studies) | Between study:<br>LungRADs pooled proportion (95% CI) (Fig. X): 140.0 (120.0 to 160.0) per 1,000<br>NLST pooled proportion: 300.0 per 1,000<br>**Difference in proportions (95% CI): 160.0 fewer (140.0 to 180.0 fewer) per 1,000** |  |  |  |  |

\*Reasons for rating down certainty: a=risk of bias, b=inconsistency, c=indirectness, d=imprecision. Explanations are within the text and figures are in Additional file 4.

For our certainty assessment of the comparison between NLST and LungRADS nodule management, we used findings from both the within (136.1 fewer per 1,000 [95% CI, 146.6 to 128.3 fewer]) and between-study (160.0 fewer per 1,000 [95% CI, 140.0 to 180.0 fewer]) analysis. Moderate-certainty evidence found that among populations meeting NLST eligibility criteria, using LungRADs nodule management probably reduces FPs by about half (45-53%) though the proportion of FPs is likely still important (i.e., at least 75 per 1,000 screened). We rated down for indirectness as both the within- and between-study analyses used older versions of the LungRADs (12 of 14 LungRADs studies used version 1.0 (68, 75, 94, 106, 119, 254–259, 261), two used version 1.1 (123, 260), none used version v2022). Also, the majority of data came from between-study analysis that required an indirect comparator. We did not have serious concerns about risk of bias. For the indirect comparator, although some studies were at high risk of any differences between study findings could also be attributed to other factors (e.g., higher-risk population (257)) which relate more to directness for which we rated down. Five of the 14 LungRADs studies were at high risk of bias (68, 94, 123, 255, 258), but there were no major differences after sensitivity analysis.

There were no within- or between-study comparisons over more than one round of screening, but over multiple rounds the number of FPs would likely increase proportionally for both groups and findings are considered applicable over multiple rounds.

## Discussion

This review provides a contemporary synthesis of the benefits and harms of LDCT screening after a decade of follow-up. Conclusions were made using up-to-date guidance, with reliance on absolute effects and considering thresholds for decision making about clinical care and guidelines which were determined by clinician consensus and validated by empirical evidence on patient preferences.

Effects considered important were found for the critical outcomes of lung-cancer and all-cause mortality (at least 2 and 1 fewer per 1000 screened, respectively) and overdiagnosis (at least 2.5 per 1000 screened). Considering the populations and screening protocols in the available trials, findings are most applicable for those starting to screen at aged 50-74 years of age, having a current or past 20-30 pack-year smoking history, and after high uptake of 3-4 rounds of annual screening with 10-12 years’ follow-up.

Most subgroup analyses were of limited credibility, mainly because of multiple differences between trials that confounded results for individual variables. Screening less frequently (e.g., NELSON trial screened at baseline then at years 1, 3 and 5.5) may be effective, though findings are limited without rigorous direct comparison between intervals. Absolute effects will differ across populations at different baseline risk (e.g., via multivariable prediction tools) and the decision thresholds used may not be met for those at the lowest risk despite meeting the eligibility of the trial having the most stringent selection criteria (NLST). Within- and between-study analyses found that women may benefit, and be harmed, from screening more than men, in both relative and absolute terms. Authors of three trials suggested that findings of larger relative effects on lung-cancer mortality for females may be due to i) relatively more small and squamous cell cancers in men, for which screening appears to make little-to-no difference (145, 170), and ii) a longer screen-detectable preclinical period for women (32). In the NLST, the effects were quite similar for men and women when analyzing adenocarcinomas (RR=0.77 and 0.73, respectively), as they were for all non-small cell lung cancers excluding squamous (170). Further, absolute effects for each sex exceeded our decision thresholds for benefits and harms.

The benefits from screening need to be weighed against the harms from overdiagnosis as well as FPs and their associated consequences. For the important number of people having one or more benign positive screening tests (i.e., FPs probably for at least 225 per 1000) there will likely be a small degree of psychosocial distress for many and possibly a moderate degree of distress for the ∼10% having to undergo invasive diagnostic procedures. Though the affects appear to wane during some months after diagnostic resolution, because resolution may take several months the harm may be considerable. Psychosocial harms from a cancer diagnosis, particularly without the chance for benefit among those overdiagnosed, may also be small in magnitude but considerably longer lasting. Despite causing psychosocial harm, data from studies measuring disutilities suggests that the overall impact on HRQoL from a FP or early-stage/overdiagnosed cancer may not be substantial. As per the findings from KQ3, the use of LungRADS instead of the nodule classification protocols in the trials appears to reduce the magnitude of FPs by up to half, but the eligible studies were not designed to capture effects on mortality which may be impacted if missed cancers (or a delay in their diagnosis) are substantial. The impact of major and serious (i.e., death) harms from invasive testing among those without cancer is likely little-to-none.

The consequences from incidental findings are hard to interpret because it is difficult to predict what proportion may end up being beneficial (due to effective treatment) versus harmful (e.g., over-investigation, overdiagnosis and over-treatment of other conditions). The large reduction (from probably 450 to 100 per 1000 screened) in incidental findings when defining them as clinically significant (“actionable” with most reportable to the referring physician) indicates that patients and primary care providers may best be spared information about many findings. What is classified as actionable may impact the magnitude and management of this outcome, which could be considerably resource intensive. For example, for the Ontario lung-cancer screening project, recommendations did not support considering emphysema as actionable for the lung screening population (262), as it was in several of the studies examined in this review.

Findings from our review on patient preferences provide support for the decision thresholds used. Based on the findings on screening benefits and harms which generally align with what we considered a moderate net-benefit scenario (i.e., somewhat less harmful than described for the low net-benefit scenario), a small majority of informed and eligible people will probably prefer screening. Further, among those at highest baseline risk there may be stronger preferences. As indicated in several studies within this review and when examining levels of intentions versus uptake of screening, people may experience barriers to screening unrelated to their attitudes which would need to be addressed to attain attendance at sufficient levels among those wanting to screen.

### Comparison with previous review

There are some differences in the methods and results between this review and that previously used by the CTFPHC for their 2016 recommendations (33). We did not examine the effects from screening with X-ray since the 2016 review as well as others have found no benefit (RR 0.99 for lung-cancer mortality). The previous review did not pool the results of the NLST (and LSS) using an X-ray comparison with other studies using comparators with no screening, but we chose to pool these (and undertake subgroup analysis) when considering the lack of benefit from X-ray. We included five additional RCTs (including the NELSON trial), longer-term results for the NLST and other trials, and at least 50 more observational studies on harms and incidental findings. The latter is likely in part due to our more comprehensive search strategy (e.g., adding incidental findings and psychosocial outcomes). The point estimates for lung-cancer mortality (3 fewer per 1000), all-cause mortality (4.6 fewer) and FPs (23% [though the guideline focused on the 35% from the NLST]) are quite similar between reviews, though we make different conclusions based on newer GRADE guidance defining the targets of certainty (i.e., decision thresholds) and recognition that precision around the point estimate requires consideration from absolute effects. We also closely investigated between- and within-study findings for key variables of interest, including sex and baseline risk for cancer and mortality. Our review focused much more on incidental findings; though difficult to define in terms of potential patient-important benefit and harms, these findings will likely be a major consideration for making decisions about screening. For the outcomes of major complications and deaths from invasive procedures, we focus on events among those without cancer to better capture the harms from screening and to avoid confounding due to these deaths also being captured for lung-cancer mortality. Our review followed more recent guidance from GRADE for reviewing patient preferences, which has a strong focus on preference-based data (e.g., HSUVs) and informed decision making. We also added data for the use of newer nodule classification system (e.g., LungRADs) to understand how this may impact the benefits and harms found in the existing trials.

### Limitations of the reviews

These reviews have potential limitations. First, only English and French language studies were included, though our search was not limited by language and we did not find evidence of any large studies on screening effects that were excluded for this reason. Second, for KQs 1 and 3 we primarily relied on existing reviews for locating studies published before 2015 (KQ1) and 2019 (KQ3) and there some minor deficiencies in the previous searches (e.g., limited sensitivity for observational studies reporting harms) to locate all studies eligible for our review. Our scans of included studies lists and several systematic reviews likely mitigated this impact. Third, the DerSimonian and Laird random-effects model was used to pool trial data, which may result in 95% CIs that are relatively narrow, particularly when heterogeneity is present (263). Considering that our judgments about precision were made in comparison with our decision thresholds and not the null where this effect may be more critical, we do not feel this impacted our conclusions. Fourth, some clinicians and interest holders may choose different decision thresholds or fail to appreciate that our conclusions reflect our certainty about whether the effects meet or exceed the decision thresholds rather than the point estimates (e.g., 4.0 fewer per 1000 for lung-cancer mortality) for which we would have (sometimes substantially) lower certainty. The thresholds were chosen to reflect effects considered by a majority of patients to be small but important, and additional thresholds for moderate and large effects, particularly for some harm outcomes, may have allowed for more nuanced conclusions. This is a major reason that we added, *post hoc,* conclusions about effects differing from the thresholds when the evidence was of moderate or high certainty. Lastly, this review did not use input from patients for determining its scope or interpreting findings. We tried to circumvent this deficiency by examining several studies on patient preferences and using patient-reported outcomes. Additional considerations potentially useful for moving from evidence to recommendations (e.g., acceptability, resources, costs) (57), and for implementing screening programs (e.g., identifying eligible patients, personnel and manner to conduct shared decision-making) were not examined in this review.

### Research gaps

There are several research gaps related to these reviews. There is limited direct, empirical evidence on important questions about screening duration and intervals, screening selection criteria (e.g., screening those <50 years old but with equivalent cancer risk to others), and nodule classification. For our KQ3, we sought but did not locate data from modelling studies on specific comparisons of interest. Modelling studies that have been used by other guideline producers, such as the USPSTF in 2021 (264), when determining their recommendations about eligibility and screening frequency, may be informative. No trial has enrolled people who have never smoked tobacco, though some modelling suggests there may be some benefit (though unknown harm) from screening among this population comprising a substantial (25%) proportion of cancer cases (265). It is not known if screening for more than 3-4 rounds improves benefits, and it may add additional harm. Further, effects among populations deserving of equity considerations, including racialized and Indigenous peoples, are lacking or very limited. Few studies in the review on patient preferences examined the disutility of screening health states or attitudes about the potential for further management and possibly overdiagnosis (e.g., thyroid nodules) from incidental findings. The effects on critical benefits and harms from using LungRADs versus the nodule management protocols in the RCTs has not been directly examined.

### Conclusions

Across the reviews, findings indicate that screening those aged 50-74 years with 3-4 rounds of LDCT will likely lead to benefits and harms for which a small majority of eligible people probably find acceptable and worthwhile. There is limited direct, empirical evidence on important questions about screening duration and intervals, screening selection criteria, and nodule classification. Effects of screening among populations deserving of equity considerations, including racialized and Indigenous peoples, are lacking or very limited.

## Data Availability

All data produced in the present work are contained in the manuscript and additional files

## Supplementary Information

The online version contains supplementary material available at x.

Additional file 1. Search strategies, eligibility criteria, and overdiagnosis calculations

Additional file 2. Additional tables and figures for key question 1

Additional file 3. Additional tables and figures for key question 2

Additional file 3. Additional tables and figures for key question 3

## Acknowledgements

Ms. Maria Tan, MLIS, of the University of Alberta, developed and ran the searches. Nicole Gehring and Sabrina Saba (University of Alberta) assisted with data extraction. Canadian Task Force on Preventive Care Lung Cancer working group members assisted with developing the scope of the review, developed the decision thresholds, and provided input to help interpret the evidence: Guylène Thériault, Eddy Lang, Donna Reynolds, Ashran Sefin, Henry Siu, Nathalie Slavtcheva, Scott Klarenbach, Jennifer Flemming, Kate Miller, and (clinical/topic experts) Matthew McInnis, Natasha Leighl, and Christian Finley.

## Author contributions

JP led the development of the review scope (eligibility) and methods. JP, SG (all KQs), SR (KQ 1), and JEP (KQ2 and 3) contributed to study screening and selection, risk of bias assessments, and analysis. JP, SG and SR assessed the certainty for KQ1. SG and JP assessed the certainty of KQ2 utility data and KQ3, and JP and JEP assessed the certainty of additional data in KQ2. LH reviewed all analyses and certainty assessments. GT and DR assisted with developing the scope of the review, helped developed the decision thresholds and analytic plan, and provided input to help interpret the evidence. JP and SG drafted the manuscript and all authors reviewed it and approve of its submission. JP is guarantor of the review.

## Funding

This review was conducted for the Canadian Task Force on Preventive Health Care that ended March 31, 2026, with funding provided by the Public Health Agency of Canada (PHAC). The views expressed in this article do not necessarily represent those of PHAC or the Government of Canada. The Staff of the Global Health and Guidelines Division at PHAC (Gregory Traversy, Laure Tessier) provided input during the development of the protocol and reviewed a draft version of the manuscript, but did not take part in the selection of studies, data extraction, analysis, or interpretation of the findings. LH is funded by a Tier 1 Canada Research Chair in Knowledge Synthesis and Translation and is a Distinguished Researcher with the Stollery Science Lab supported by the Stollery Children’s Hospital Foundation.

## Availability of data and materials

The data generated during this study are available within the manuscript or its supplementary files.

## Declarations

### Ethics approval and consent to participate

Not applicable.

### Consent for publication

Not applicable

### Competing interests

The authors declare that they have no competing interests.

### Author details

^1^Alberta Research Centre for Health Evidence, Faculty of Medicine and Dentistry, University of Alberta, 11405 87 Avenue NW, Edmonton, Alberta T6G 1C9, Canada. ^2^ Université de Montréal, ^3^ University of Toronto

## Notes

### Competing Interest Statement

The authors have declared no competing interest.

