## Additional files for "Screening for Lung Cancer with Computed Tomography: Systematic Reviews on Effectiveness and Patient Preferences"

### Additional file 1. Search strategies, eligibility criteria, and overdiagnosis calculations

#### Key Question 1 Search strategies

Database(s): **Ovid MEDLINE(R) ALL** 1946, run Oct 5 2022 and July 11, 2025

| # | Searches |
|---|---|
| 1 | exp Lung Neoplasms/ |
| 2 | ((bronchial or lung or pulmonary) adj3 (adenocarcinoma or cancer* or carcinoma or malignan* or neoplasm* or tumo?r*)).ti,ab,kf. |
| 3 | 1 or 2 |
| 4 | Early Detection of Cancer/ |
| 5 | Mass Screening/ |
| 6 | Tomography, X-Ray Computed/ |
| 7 | (compute* adj2 tomograph*).ti,ab,kf. |
| 8 | (c?t adj1 (chest* or imag* or scan*)).ti,ab,kf. |
| 9 | (LDCT or LD-CT or ULDCT or ULD-CT).ti,ab,kf. |
| 10 | (low-dose adj ct*).ti,ab,kf. |
| 11 | screen*.ti,ab,kf. |
| 12 | or/4-11 |
| 13 | 3 and 12 |
| 14 | limit 13 to yr="2015 -Current" |
| 15 | exp Radiography, Thoracic/ |
| 16 | Sputum/cy [Cytology] |
| 17 | ((chest or thoracic or thorax) adj1 (radiograph* or xray* or x-ray*)).ti,ab,kf. |
| 18 | (sputum adj2 cyto*).ti,ab,kf. |
| 19 | or/15-18 |
| 20 | 3 and 19 |
| 21 | limit 20 to yr="2020 -Current" |
| 22 | 14 or 21 |
| 23 | Randomization/ |

| | |
|---|---|
| 24 | Random Allocation/ |
| 25 | Double-Blind Method/ |
| 26 | Double-Blind Studies/ |
| 27 | Single-Blind Method/ |
| 28 | Single-Blind Studies/ |
| 29 | Control Groups/ |
| 30 | allocated.ti,ab,hw. |
| 31 | (control* adj3 (group* or studies or study or trial*)).ti,ab,kf. |
| 32 | ((Danish or Dante or German or LUSI or MILD or Nelson or UK) adj5 (lung* or trial)).ti,ab. |
| 33 | (DLCST or DLST or Italung or NLST or UKLS).ti,ab. |
| 34 | ((doubl* or singl* or trebl* or triple) adj (blind* or dumm* or mask*)).ti,ab,hw,kf. |
| 35 | random*.ti,ab,hw,kf. |
| 36 | or/23-35 |
| 37 | 22 and 36 |
| 38 | exp Lung Neoplasms/ |
| 39 | ((bronchial or lung or pulmonary) adj3 (adenocarcinoma or cancer* or carcinoma or malignan* or neoplasm* or tumo?r*)).ti,ab,kf. |
| 40 | 38 or 39 |
| 41 | Early Detection of Cancer/ |
| 42 | Mass Screening/ |
| 43 | Tomography, X-Ray Computed/ |
| 44 | (compute* adj2 tomograph*).ti,ab,kf. |
| 45 | (c?t adj1 (chest* or imag* or scan*)).ti,ab,kf. |
| 46 | (LDCT or LD-CT or ULDCT or ULD-CT).ti,ab,kf. |
| 47 | (low-dose adj ct*).ti,ab,kf. |
| 48 | screen*.ti,ab,kf. |
| 49 | or/41-48 |
| 50 | 40 and 49 |

| | |
|---|---|
| 51 | limit 50 to yr="2015 -Current" |
| 52 | exp Radiography, Thoracic/ |
| 53 | Sputum/cy [Cytology] |
| 54 | ((chest or thoracic or thorax) adj1 (radiograph* or xray* or x-ray*)).ti,ab,kf. |
| 55 | (sputum adj2 cyto*).ti,ab,kf. |
| 56 | or/52-55 |
| 57 | 40 and 56 |
| 58 | limit 57 to yr="2020 -Current" |
| 59 | 51 or 58 |
| 60 | exp Anti-Anxiety Agents/ |
| 61 | exp Antidepressive Agents/ |
| 62 | Anxiety/ |
| 63 | denial, psychological/ |
| 64 | Depression/ |
| 65 | exp Fear/ |
| 66 | Mental Health/ |
| 67 | Psychological Distress/ |
| 68 | Psychometrics/ |
| 69 | exp Psychiatric Status Rating Scales/ |
| 70 | Stress, Psychological/ |
| 71 | (afraid or fear*).ti,ab,kf. |
| 72 | (antianxiety or anti-anxiety or anxiolytic* or tranquiliz*).ti,ab,kf. |
| 73 | (antidepress* or anti-depress*).ti,ab,kf. |
| 74 | (anxiet* or anxious*).ti,ab,kf. |
| 75 | BDI-II.ti,ab,kf. |
| 76 | ((cancer or worry) adj2 scale*).ti,ab,kf. |
| 77 | (consequence*-of-screening* or COS or COS-LC).ti,ab,kf. |

| | |
|---|---|
| 78 | (denial or deny).ti,ab,kf. |
| 79 | depressi*.ti,ab,kf. |
| 80 | distress*.ti,ab,kf. |
| 81 | ((emotion* or psychological or psychosocial or psycho-social) adj2 (aftereffect* or aspect* or burden* or consequence* or discomfort* or effect* or impact* or measure* or outcome* or reaction* or response* or symptom*)).ti,ab,kf. |
| 82 | HADS.ti,ab,kf. |
| 83 | (impact-of-event*-scale* or IES).ti,ab,kf. |
| 84 | mental-health.ti,ab,kf. |
| 85 | (patient*-health-questionnaire* or PHQ9 or PHQ-9).ti,ab,kf. |
| 86 | PCQ.ti,ab,kf. |
| 87 | ((psychological or psychosocial or psycho-social) adj2 (assess* or questionnaire* or scale* or screen* or test*)).ti,ab,kf. |
| 88 | (psychometr* or psycho-metr*).ti,ab,kf. |
| 89 | STAI.ti,ab,kf. |
| 90 | stress*.ti,ab,kf. |
| 91 | or/60-90 |
| 92 | Diagnostic Errors/ |
| 93 | False Positive Reactions/ |
| 94 | Incidental Findings/ |
| 95 | exp Medical Overuse/ |
| 96 | ((adverse or harm* or serious or undesir* or uninten* or unwant*) adj3 (effect* or event* or outcome* or reaction*)).ti,ab,kf. |
| 97 | ((bleed* or complication* or harm* or infection*) adj3 (biops* or death* or followup* or follow-up* or morbidity or mortality or procedure* or resect* or surg* or test* or treatment* or workup* or work-up*)).ti,ab,kf. |
| 98 | ((extrapulmonary or extra-pulmonary or incidental) adj3 (discover* or finding*)).ti,ab,kf. |
| 99 | false*-positiv*.ti,ab,kf. |
| 100 | (invasive adj2 (follow-up* or followup* or procedure* or test* or treatment* or workup* or work up*)).ti,ab,kf. |

| | |
|---|---|
| 101 | (overdetect* or over-detect* or overdiagnos* or over-diagnos* or overtreat* or over-treat*).ti,ab,kf. |
| 102 | (unnecessary adj2 (biops* or followup* or follow-up* or procedur* or resect* or surg* or test* or treatment* or workup* or work-up*)).ti,ab,kf. |
| 103 | or/92-102 |
| 104 | 59 and (91 or 103) |
| 105 | Epidemiologic studies/ |
| 106 | exp case control studies/ |
| 107 | exp cohort studies/ |
| 108 | Case control.tw. |
| 109 | (cohort adj (study or studies)).tw. |
| 110 | Cohort analy$.tw. |
| 111 | ((Follow-up or followup) adj (study or studies)).tw. |
| 112 | (observational adj (study or studies)).tw. |
| 113 | Longitudinal.tw. |
| 114 | Retrospective.tw. |
| 115 | (Cross-sectional or crosssectional).tw. |
| 116 | Cross-sectional studies/ |
| 117 | ((control and (group* or study)) or (time and factors) or program or survey* or ci or cohort or comparative-stud* or evaluation-studies or follow-up*).mp. |
| 118 | or/105-117 |
| 119 | 104 and 118 |
| 120 | 37 or 119 |
| 121 | (Case Reports.pt. or (case report? or case study or case studies).ti.) not (review* or trial*).ti,ab,kf,hw. |
| 122 | comment/ or editorial/ or (comment or editorial or news or newspaper article).pt. |
| 123 | 120 not (121 or 122) |
| 124 | remove duplicates from 123 |

Database(s): **Embase** 1974 to 2022, ran October 05 2022 and July 11 2025

| # | Searches |
|---|---|
| 1 | exp lung tumor/ |
| 2 | ((bronchial or lung or pulmonary) adj3 (adenocarcinoma or cancer* or carcinoma or malignan* or neoplasm* or tumo?r*)).ti,ab,kw. |
| 3 | 1 or 2 |
| 4 | early cancer diagnosis/ |
| 5 | Mass Screening/ |
| 6 | computer assisted tomography/ |
| 7 | (compute* adj2 tomograph*).ti,ab,kw. |
| 8 | (c?t adj1 (chest* or imag* or scan*)).ti,ab,kw. |
| 9 | (LDCT or LD-CT or ULDCT or ULD-CT).ti,ab,kw. |
| 10 | (low-dose adj ct*).ti,ab,kw. |
| 11 | screen*.ti,ab,kw. |
| 12 | or/4-11 |
| 13 | 3 and 12 |
| 14 | limit 13 to yr="2015 -Current" |
| 15 | exp thorax radiography/ |
| 16 | sputum cytodiagnosis/ |
| 17 | ((chest or thoracic or thorax) adj1 (radiograph* or xray* or x-ray*)).ti,ab,kw. |
| 18 | (sputum adj2 cyto*).ti,ab,kw. |
| 19 | or/15-18 |
| 20 | 3 and 19 |
| 21 | limit 20 to yr="2020 -Current" |
| 22 | 14 or 21 |
| 23 | randomization/ |
| 24 | double-blind procedure/ |

| | |
|---|---|
| 25 | single-blind procedure/ |
| 26 | control group/ |
| 27 | allocated.ti,ab,hw. |
| 28 | (control* adj3 (group* or studies or study or trial*)).ti,ab,kf. |
| 29 | ((Danish or Dante or German or LUSI or MILD or Nelson or UK) adj5 (lung* or trial)).ti,ab. |
| 30 | (DLCST or DLST or Italung or NLST or UKLS).ti,ab. |
| 31 | ((doubl* or singl* or trebl* or triple) adj (blind* or dumm* or mask*)).ti,ab,hw,kf. |
| 32 | random*.ti,ab,hw,kf. |
| 33 | or/23-32 |
| 34 | 22 and 33 |
| 35 | exp lung tumor/ |
| 36 | ((bronchial or lung or pulmonary) adj3 (adenocarcinoma or cancer* or carcinoma or malignan* or neoplasm* or tumo?r*)).ti,ab,kw. |
| 37 | 35 or 36 |
| 38 | early cancer diagnosis/ |
| 39 | Mass Screening/ |
| 40 | computer assisted tomography/ |
| 41 | (compute* adj2 tomograph*).ti,ab,kw. |
| 42 | (c?t adj1 (chest* or imag* or scan*)).ti,ab,kw. |
| 43 | (LDCT or LD-CT or ULDCT or ULD-CT).ti,ab,kw. |
| 44 | (low-dose adj ct*).ti,ab,kw. |
| 45 | screen*.ti,ab,kw. |
| 46 | or/38-45 |
| 47 | 37 and 46 |
| 48 | limit 47 to yr="2015 -Current" |
| 49 | exp thorax radiography/ |
| 50 | sputum cytodiagnosis/ |
| 51 | ((chest or thoracic or thorax) adj1 (radiograph* or xray* or x-ray*)).ti,ab,kw. |

| | |
|---|---|
| 52 | (sputum adj2 cyto*).ti,ab,kw. |
| 53 | or/49-52 |
| 54 | 37 and 53 |
| 55 | limit 54 to yr="2020 -Current" |
| 56 | 48 or 55 |
| 57 | exp anxiolytic agent/ |
| 58 | anxiety/ |
| 59 | denial/ |
| 60 | depression/ |
| 61 | fear/ |
| 62 | mental health/ |
| 63 | distress syndrome/ |
| 64 | psychological rating scale/ |
| 65 | mental stress/ |
| 66 | (afraid or fear*).ti,ab,kw. |
| 67 | (antianxiety or anti-anxiety or anxiolytic* or tranquiliz*).ti,ab,kw. |
| 68 | (antidepress* or anti-depress*).ti,ab,kw. |
| 69 | (anxiet* or anxious*).ti,ab,kw. |
| 70 | BDI-II.ti,ab,kw. |
| 71 | ((cancer or worry) adj2 scale*).ti,ab,kw. |
| 72 | (consequence*-of-screening* or COS or COS-LC).ti,ab,kw. |
| 73 | (denial or deny).ti,ab,kw. |
| 74 | depressi*.ti,ab,kw. |
| 75 | distress*.ti,ab,kw. |
| 76 | ((emotion* or psychological or psychosocial or psycho-social) adj2 (aftereffect* or aspect* or burden* or consequence* or discomfort* or effect* or impact* or measure* or outcome* or reaction* or response* or symptom*)).ti,ab,kw. |
| 77 | HADS.ti,ab,kw. |

| | |
|---|---|
| 78 | (impact-of-event*-scale* or IES).ti,ab,kw. |
| 79 | mental-health.ti,ab,kw. |
| 80 | (patient*-health-questionnaire* or PHQ9 or PHQ-9).ti,ab,kw. |
| 81 | PCQ.ti,ab,kw. |
| 82 | ((psychological or psychosocial or psycho-social) adj2 (assess* or questionnaire* or scale* or screen* or test*)).ti,ab,kw. |
| 83 | (psychometr* or psycho-metr*).ti,ab,kw. |
| 84 | STAI.ti,ab,kw. |
| 85 | stress*.ti,ab,kw. |
| 86 | or/57-85 |
| 87 | 56 and 86 |
| 88 | diagnostic error/ |
| 89 | incidental finding/ |
| 90 | exp medical overuse/ |
| 91 | ((adverse or harm* or serious or undesir* or uninten* or unwant*) adj3 (effect* or event* or outcome* or reaction*)).ti,ab,kw. |
| 92 | ((bleed* or complication* or harm* or infection*) adj3 (biops* or death* or followup* or follow-up* or morbidity or mortality or procedure* or resect* or surg* or test* or treatment* or workup* or work-up*)).ti,ab,kw. |
| 93 | ((extrapulmonary or extra-pulmonary or incidental) adj3 (discover* or finding*)).ti,ab,kw. |
| 94 | false*-positiv*.ti,ab,kw. |
| 95 | (invasive adj2 (follow-up* or followup* or procedure* or test* or treatment* or workup* or work up*)).ti,ab,kw. |
| 96 | (overdetect* or over-detect* or overdiagnos* or over-diagnos* or overtreat* or over-treat*).ti,ab,kw. |
| 97 | (unnecessary adj2 (biops* or followup* or follow-up* or procedur* or resect* or surg* or test* or treatment* or workup* or work-up*)).ti,ab,kw. |
| 98 | or/88-97 |
| 99 | 56 and (86 or 98) |
| 100 | exp case control study/ |
| 101 | cohort analysis/ |

| | |
|---|---|
| 102 | Case control.ti,ab. |
| 103 | (cohort adj (study or studies)).ti,ab. |
| 104 | Cohort analy$.ti,ab. |
| 105 | ((Follow-up or followup) adj (study or studies)).ti,ab. |
| 106 | (observational adj (study or studies)).ti,ab. |
| 107 | Longitudinal.ti,ab. |
| 108 | Retrospective.ti,ab. |
| 109 | (Cross-sectional or crosssectional).ti,ab. |
| 110 | ((control and (group* or study)) or (time and factors) or program or survey* or ci or cohort or comparative-stud* or evaluation-studies or follow-up*).mp. |
| 111 | or/100-110 |
| 112 | 99 and 111 |
| 113 | 34 or 112 |
| 114 | (Case Reports.pt. or (case report? or case study or case studies).ti.) not (review* or trial*).ti,ab,kf,hw. |
| 115 | comment/ or editorial/ or (comment or editorial).pt. |
| 116 | (conference abstract* or conference paper*).af. not (conference abstract* or conference paper*).ab,ga. |
| 117 | conference proceeding*.af. not conference proceeding*.ab,ga. |
| 118 | 113 not (or/114-117) |
| 119 | remove duplicates from 118 |

Cochrane Central Register of Controlled Trials, Issue 10 of 12, ran October 11 2022 and July 2025

ID Search Hits

#1 [mh "Lung Neoplasms"]

#2 ((bronchial or lung or pulmonary) NEAR/2 (adenocarcinoma or cancer* or carcinoma or malignan* or neoplasm* or tumo?r*)):ti,ab,kw

#3 #1 OR #2

#4 [mh ^"Early Detection of Cancer"]

#5 [mh ^"Mass Screening"]

#6 [mh ^"Tomography, X-Ray Computed"]

#7 (compute* NEAR/1 tomograph*):ti,ab,kw

#8 (c?t NEAR/0 (chest* or imag* or scan*)):ti,ab,kw

#9 (LDCT or LD-CT or ULDCT or ULD-CT):ti,ab,kw

#10 (low-dose NEXT ct*):ti,ab,kw

#11 screen*:ti,ab,kw

#12 #4 OR #5 OR #6 OR #7 OR #8 OR #9 OR #10 OR #11

#13 #3 AND #12 with Publication Year from 2015 to 2022, in Trials

#14 [mh "Radiography, Thoracic"]

#15 MeSH descriptor: [Sputum] this term only and with qualifier(s): [cytology - CY]

#16 ((chest or thoracic or thorax) NEXT (radiograph* or xray* or x-ray*)):ti,ab,kw

#17 (sputum NEAR/1 cyto*):ti,ab,kw

#18 #14 OR #15 OR #16 OR #17

#19 #3 AND #18 with Publication Year from 2020 to 2022, in Trials

#20 #13 OR #19

#21 [mh ^"Randomization"]

#22 [mh ^"Random Allocation"]

#23 [mh ^"Double-Blind Method"]

#24 [mh ^"Double-Blind Studies"]

#25 [mh ^"Single-Blind Method"]

#26 [mh ^"Single-Blind Method"]

#27 [mh ^"Control Groups"]

#28 allocated:ti,ab,kw

#29 (control* NEAR/2 (group* or studies or study or trial*)):ti,ab,kw

#30 ((Danish or Dante or German or LUSI or MILD or Nelson or UK) NEAR/4 (lung* or trial)):ti,ab

#31 (DLCST or DLST or Italung or NLST or UKLS):ti,ab

#32 ((doubl* or singl* or trebl* or triple) adj (blind* or dumm* or mask*)):ti,ab,kw

#33 random*:ti,ab,kw

#34 #21 OR #22 OR #23 OR #24 OR #25 OR #26 OR #27 OR #28 OR #29 OR #30 OR 331 OR #32 OR #33 1339600

#35 #20 AND #34

#36 [mh "Anti-Anxiety Agents"]

#37 [mh "Antidepressive Agents"]

#38 [mh ^"Anxiety"]

#39 [mh ^"denial, psychological"]

#40 [mh ^"Depression"]

#41 [mh "Fear"]

#42 [mh ^"Mental Health"]

#43 [mh ^"Psychological Distress"]

#44 [mh ^"Psychological Distress"]

#45 [mh ^"Psychological Distress"]

#46 [mh ^"Stress, Psychological"]

#47 (afraid or fear*):ti,ab,kw

#48 (antianxiety or anti-anxiety or anxiolytic* or tranquiliz*):ti,ab,kw

#49 (antidepress* or anti-depress*):ti,ab,kw

#50 (anxiet* or anxious*):ti,ab,kw

#51 BDI-II:ti,ab,kw

#52 ((cancer or worry) NEAR/1 scale*):ti,ab,kw

#53 ("consequence*-of-screening" OR COS OR COS-LC):ti,ab,kw

#54 (denial or deny):ti,ab,kw

#55 depressi*:ti,ab,kw

#56 distress*:ti,ab,kw

#57 ((emotion* or psychological or psychosocial or psycho-social) NEAR/1 (aftereffect* or aspect* or burden* or consequence* or discomfort* or effect* or impact* or measure* or outcome* or reaction* or response* or symptom*)):ti,ab,kw

#58 HADS:ti,ab,kw

#59 ("impact-of-event*-scale*" or IES):ti,ab,kw

#60 mental-health:ti,ab,kw

#61 ("patient*-health-questionnaire*" or PHQ9 or PHQ-9):ti,ab,kw

#62 PCQ:ti,ab,kw

#63 ((psychological or psychosocial or psycho-social) NEAR/1 (assess* or questionnaire* or scale* or screen* or test*)):ti,ab,kw

#64 (psychometr* or psycho-metr*):ti,ab,kw

#65 STAI:ti,ab,kw

#66 stress*:ti,ab,kw

#67 #36 OR #37 OR #38 OR #39 OR #40 OR #41 OR #42 OR #43 OR #44 OR #45 OR #46 OR #47 OR #48 OR #49 OR #50 OR #51 OR #52 OR #53 OR #54 OR #55 OR #56 OR #57 OR #58 OR #59 OR #60 OR #61 OR #62 OR #63 OR #64 OR #65 OR #66

#68 [mh ^"Diagnostic Errors"]

#69 [mh ^"False Positive Reactions"]

#70 [mh ^"Incidental Findings"]

#71 [mh "Medical Overuse"]

#72 ((adverse or harm* or serious or undesir* or uninten* or unwant*) NEAR/2 (effect* or event* or outcome* or reaction*)):ti,ab,kw

#73 ((bleed* or complication* or harm* or infection*) NEAR/2 (biops* or death* or followup* or follow-up* or morbidity or mortality or procedure* or resect* or surg* or test* or treatment* or workup* or work-up*)):ti,ab,kw

#74 ((extrapulmonary or extra-pulmonary or incidental) NEAR/2 (discover* or finding*)):ti,ab,kw

#75 "false*positiv*":ti,ab,kw

#76 (invasive NEAR/1 (follow-up* or followup* or procedure* or test* or treatment* or workup* or work-up*)):ti,ab,kw

#77 (overdetect* or over-detect* or overdiagnos* or over-diagnos* or overtreat* or over-treat*):ti,ab,kw

#78 (unnecessary NEAR/1 (biops* or followup* or follow-up* or procedur* or resect* or surg* or test* or treatment* or workup* or work-up*)):ti,ab,kw

#79 #68 or #69 OR #70 OR #71 OR #72 OR #73 OR #74 OR #75 OR #76 OR #77 OR #78

#80 (#67 OR #79)

#81 #20 AND #80

#82 [mh ^"Epidemiologic studies"]

#83 [mh "Case control studies"]

#84 [mh "cohort studies"]

#85 Case control:ti,ab

#86 (cohort NEXT (study or studies)):ti,ab

#87 "Cohort analysis" OR "cohort analyses":ti,ab

#88 ((Follow-up or followup) NEXT (study or studies)):ti,ab

#89 (observational NEXT (study or studies)):ti,ab

#90 Longitudinal:ti,ab

#91 Retrospective:ti,ab

#92 (Cross-sectional or crosssectional):ti,ab

#93 [mh ^"Cross-sectional studies"]

#94 ((control and (group* or study)) or (time and factors) or program or survey* or ci or cohort or comparative-stud* or evaluation-studies or follow-up*):ti,ab,kw

#95 #82 OR #83 OR #84 OR #85 OR #86 OR #87 OR #88 OR #89 OR #90 OR #91 OR #92 OR #93 OR #94

#96 #81 AND #95

#97 #35 OR #96

#### Key Question 2 Search Strategies

Database(s): Ovid MEDLINE(R) ALL 1946 to April 18, 2023

Search dates: April 20 2023 and July 11 2025

| # | Searches |
|---|---|
| 1 | exp Lung Neoplasms/ |
| 2 | ((bronchial or lung or pulmonary) adj3 (adenocarcinoma or cancer* or carcinoma or lesion* or malignan* or mass$2 or neoplasm* or tumo?r* or nodule*)).ti,ab,kf. |
| 3 | 1 or 2 |
| 4 | Early Detection of Cancer/ |
| 5 | Mass Screening/ |
| 6 | (screen* or detect*).ti,ab,kf. |
| 7 | or/4-6 |
| 8 | 3 and 7 |
| 9 | attitude to health/ |

10 Choice Behavior/

11 Consumer Behavior/

12 Decision Making/

13 Decision Making, Shared/

14 decision support techniques/

15 health knowledge, attitudes, practice/

16 "patient acceptance of health care"/

17 patient participation/

18 patient preference/

19 patient satisfaction/

20 Perception/

21 ((acceptabilit* or acceptance or acceptable or attitude* or belief* or expectation* or intent* or perceiv* or perception* or perspective* or prefer* or valuation* or valued or values or views or willing*) adj4 (elicit* or client* or consumer* or female* or health or individual* or man or men or male$1 or participant* or patient* or people* or person* or public* or respondent* or screenee* or smoker* or stated or user* or wom#n)).ti,ab,kf.

22 ((choice* or choos*) adj2 (behavio?r* or discrete or experiment*)).ti,ab,kf.

23 (conjoint adj3 (analy* or design* or evaluation or exercise* or experiment* or studies or study or survey* or task* or valuation or value* or valuing)).ti,ab,kf.

24 (contingent adj3 (analy* or design* or evaluation or valuation or value* or valuing)).ti,ab,kf.

25 (decision* adj (board$1 or certain* or conflict* or dissatisf* or satisf* or uncertain*)).ti,ab,kf.

26 (decision* adj2 (aid$1 or mak* or needs or support* or tool*)).ti,ab,kf.

27 (decision* adj5 balance).ti,ab,kf.

28 (ranking or rating).ti,ab,kf.

29 (("LCS" or screen*) adj4 (acceptabilit*or acceptance or acceptable or agree* or attitude* or behavio?r* or belief* or choice* or choos* or decid* or decision* or declin* or expectation* or inclin* or intending or intent* or perceiv* or perception* or perspective* or prefer*or propensity or pursue or values or views or willing*)).ti,ab,kf.

30 (tradeoff* or trade-off*).ti,ab,kf.

31 or/9-30

32 8 and 31

33 (Case Reports.pt. or (case report? or case study or case studies).ti.) not (review* or trial*).ti,ab,kf,hw.

34 comment/ or editorial/ or (comment or editorial or news or newspaper article).pt.

35 (exp Animals/ or Models, Animal/ or Disease Models, Animal/) not Humans/

36 ((animal or animals or canine* or dog or dogs or feline or hamster* or lamb or lambs or mice or monkey or monkeys or mouse or murine or pig or pigs or piglet* or porcine or primate* or rabbit* or rats or rat or rodent* or sheep* or veterinar*) not (human* or patient*)).mp.

37 32 not (or/33-36)

38 limit 37 to yr="2012-Current"

39 remove duplicates from 38

40 exp Lung Neoplasms/

41 ((bronchial or bronchus or lung or pulmonary) adj3 (adenocarcinoma or cancer* or carcinoma or malignan* or neoplasm* or tumo?r*)).ti,ab,kf.

42 40 or 41

43 Diagnostic Errors/

44 False Positive Reactions/

45 Incidental Findings/

46 Precancerous Conditions/

47 Solitary Pulmonary Nodule/

48 ((bronchial or bronchus or lung or pulmonary) adj3 (lesion* or mass$2 or node* or nodule*)).ti,ab,kf.

49 ((extrapulmonary or extra-pulmonary or incidental) adj3 (discover* or finding*)).ti,ab,kf.

50 (false*-positiv* or overdiagnosis or (diagnos* adj3 error*)).ti,ab,kf.

51 (indetermina* or suspect* or suspicious*).ti,ab,kf.

52 or/43-51

53 (bronchial or bronchus or lung or pulmonary).hw,ti,ab,kf.

54 52 and 53

55 Quality-Adjusted Life Years/

56 *"Quality of Life"/

57 "Value of Life"/

58 Visual Analog Scale/

59 (DALY or DALYS or QALY or QALYS).ti,ab,kf.

60 direct*-elicitation*.ti,ab,kf.

61 disabil*-weight*.ti,ab,kf.

62 ((disability or quality) adj adjusted-life).ti,ab,kf.

63 eortc-qlq-30c.ti,ab,kf.

64 (EuroQol5D or EuroQol or Euro-Qol or EuroQual5D or EuroQual or Euro-Qual or EQ5D or EQ-5D).ti,ab,kf.

65 fact-i.ti,ab,kf.

66 (health adj2 (utilit* or disutilit*)).ti,ab,kf.

67 (health* adj2 year* adj2 equivalent*).ti,ab,kf.

68 (health-stat$2 adj2 (cost* or estimat* or index or indice* or measure* or scale* or score* or valu* or weight*)).ti,ab,kf.

69 ((HRQOL or QOL or quality-of-life) adj3 (assess* or instrument*)).ti,ab,kf.

70 (HSUV or HUSVs).ti,ab,kf.

71 (HUI or HUI2 or HUI3).ti,ab,kf.

72 (HYE or HYES).ti,ab,kf.

73 (multi-attribute or multi-criteria or multiattribute or multicriteria).tw,kf.

74 (quality adj2 (wellbeing or well-being)).ti,ab,kf.

75 (sf6 or sf-6 or short-form-6 or shortform-6 or sf-six or sfsix or shortform-six or short-form-six or shortform6 or short-form6).ti,ab,kf.

76 (sf8 or sf-8 or sf-eight or sfeight or shortform-8 or shortform-8 or shortform8 or short-form8 or shortform-eight or short-form-eight).ti,ab,kf.

77 (sf12 or sf-12 or short-form-12 or shortform-12 or short-form12 or shortform12 or sf-twelve or sftwelve or shortform-twelve or short-form-twelve).ti,ab,kf.

78 (sf16 or sf-16 or short-form-16 or shortform-16 or short-form16 or shortform16 or sf-sixteen or sfsixteen or shortform-sixteen or short-form-sixteen).ti,ab,kf.

79 (sf20 or sf-20 or short-form-20 or shortform-20 or short-form20 or shortform20 or sf-twenty or sftwenty or shortform-twenty or short-form-twenty).ti,ab,kf.

80 (sf36 or sf-36 or short-form-36 or shortform-36 or short-form36 or shortform36 or sf-thirtysix or sfthirtysix or sfthirty-six or sf-thirty-six or shortform-thirtysix or shortform-thirty-six or short-form-thirtysix or short-form-thirty-six).ti,ab,kf.

81 standard gamble*.ti,ab,kf.

82 (time adj (tradeoff or trade-off)).ti,ab,kf.

83 TTO.ti,ab,kf.

84 (utility adj (cost* or estimat* or index or indice* or measure* or scale* or score* or valu* or weight*)).ti,ab,kf.

85 vignette*.ti,ab,kf.

86 visual-analog*-scale*.ti,ab,kf.

87 or/55-86

88 42 and 87

89 54 and 87

90 88 or 89

91 (Case Reports.pt. or (case report? or case study or case studies).ti.) not (review* or trial*).ti,ab,kf,hw.

92 comment/ or editorial/ or (comment or editorial or news or newspaper article).pt.

93 (exp Animals/ or Models, Animal/ or Disease Models, Animal/) not Humans/

94 ((animal or animals or canine* or dog or dogs or feline or hamster* or lamb or lambs or mice or monkey or monkeys or mouse or murine or pig or pigs or piglet* or porcine or primate* or rabbit* or rats or rat or rodent* or sheep* or veterinar*) not (human* or patient*)).mp.

95 90 not (or/91-94)

96 limit 95 to yr="2012 -Current"

97 remove duplicates from 96

98 39 or 97

Database: EconLit

Search dates: 20 Apr2023 and July 11, 2025

Search mode: Find all my search terms

| # | **Query** |
|---|---|
| S1 | ((bronchial or lung or pulmonary) N3 (adenocarcinoma or cancer* or carcinoma or lesion* or malignan* or mass or masses or neoplasm* or tumo#r* or nodule*)) AND ( screen* or detect*)) |
| S2 | (acceptabilit* or acceptance or acceptable or attitude* or belief* or expectation* or intent* or perceiv* or perception* or perspective* or prefer* or valuation* or valued or values or views or willing*) N4 (elicit* or client* or consumer* or female* or health or individual* or man or men or male$1 or participant* or patient* or people* or person* or public* or respondent* or screenee* or smoker* or stated or user* or wom?n) |
| S3 | (choice* or choos*) N2 (behavio?r* or discrete or experiment*) |

| | |
|---|---|
| S4 | (conjoint N3 (analy* or design* or evaluation or exercise* or experiment* or studies or study or survey* or task* or valuation or value* or valuing) |
| S5 | contingent N3 (analy* or design* or evaluation or valuation or value* or valuing) |
| S6 | decision* N0 (board or boards or certain* or conflict* or dissatisf* or satisf* or uncertain*) |
| S7 | decision* N1 (aid or aids or mak* or needs or support* or tool*) |
| S8 | decision* N3 balance |
| S9 | ranking or rating |
| S10 | (tradeoff* or trade-off*) |
| S11 | S2 OR S3 OR S4 OR S5 OR S6 OR S7 OR S8 OR S9 OR S10 |
| S12 | S1 AND S11 |
| S13 | ((bronchial or bronchus or lung or pulmonary) N2 (adenocarcinoma or cancer* or carcinoma or malignan* or neoplasm* or tumo#r*) |
| S14 | (bronchial or bronchus or lung or pulmonary) N2 (lesion* or mass or masses or node* or nodule*) |
| S15 | (extrapulmonary or extra-pulmonary or incidental) N2 (discover* or finding*) |
| S16 | (false*-positiv* or overdiagnosis or (diagnos* N2 error*) |
| S17 | (indetermina* or suspect* or suspicious*) |
| S18 | (bronchial or bronchus or lung or pulmonary) |
| S19 | (S14 OR S15 OR S16 OR S17) AND S18 |
| S20 | (DALY or DALYS or QALY or QALYS) |
| S21 | direct*-elicitation* |
| S22 | disabil*-weight* |
| S23 | (disability or quality) N0 adjusted-life |

| | |
|---|---|
| S24 | Eortc-qlq-30c |
| S25 | (EuroQol5D or EuroQol or Euro-Qol or EuroQual5D or EuroQual or Euro-Qual or EQ5D or EQ-5D) |
| S26 | Fact-i |
| S27 | health N1 (utilit* or disutilit*) |
| S28 | (health* N1 year* N1 equivalent*) |
| S29 | health-status N1 (cost* or estimat* or index or indice* or measure* or scale* or score* or valu* or weight*) |
| S30 | (HRQOL or QOL or quality-of-life) N2 (assess* or instrument*) |
| S31 | (HSUV or HUSVs or HUI or HUI2 or HUI3 or HYE or HYES) |
| S32 | (multi-attribute or multi-criteria or multiattribute or multicriteria) |
| S33 | (quality N1 (wellbeing or well-being) |
| S34 | (sf6 or sf-6 or short-form-6 or shortform-6 or sf-six or sfsix or shortform-six or "short-form-six" or shortform6 or short-form6) |
| S35 | sf8 or sf-8 or sf-eight or sfeight or shortform-8 or shortform-8 or shortform8 or short-form8 or shortform-eight or short-form-eight |
| S36 | (sf12 or sf-12 or short-form-12 or shortform-12 or short-form12 or shortform12 or sf-twelve or sftwelve or shortform-twelve or short-form-twelve) |
| S37 | (sf16 or sf-16 or short-form-16 or shortform-16 or short-form16 or shortform16 or sf-sixteen or sfsixteen or shortform-sixteen or short-form-sixteen) |
| S38 | sf20 or sf-20 or short-form-20 or shortform-20 or short-form20 or shortform20 or sf-twenty or sftwenty or shortform-twenty or short-form-twenty |
| S39 | (sf36 or sf-36 or short-form-36 or shortform-36 or short-form36 or shortform36 or sf-thirtysix or sfthirtysix or sfthirty-six or sf-thirty-six or shortform-thirtysix or shortform-thirty-six or short-form-thirtysix or short-form-thirty-six) |

| | |
|---|---|
| S40 | standard gamble* |
| S41 | time N0 (tradeoff or trade-off) |
| S42 | TTO |
| S43 | (utility N0 (cost* or estimat* or index or indice* or measure* or scale* or score* or valu* or weight*)) |
| S44 | vignette* |
| S45 | visual-analog*-scale* |
| S46 | S20 OR S21 OR S22 OR S23 OR S24 OR S25 OR S26 OR S27 OR S28 OR S29 OR S30 OR S31 OR S32 OR S33 OR S34 OR S35 OR S36 OR S37 OR S38 OR S39 OR S40 OR S41 OR S42 OR S43 OR S44 OR S45 |
| S47 | S13 AND S46 |
| S48 | S19 AND S46 |
| S49 | S47 OR S48 |
| S50 | S12 OR S49 |
| S51 | S12 OR S49 [Limiters Published Date: 20120101-20230431] |

Database: Scopus

Search dates: 20 Apr2023 and July 22, 2025

( ( ( TITLE ( ( bronchial OR lung OR pulmonary ) W/2 ( adenocarcinoma OR cancer* OR carcinoma OR lesion* OR malignan* OR mass OR masses OR neoplasm* OR tumor* OR tumour OR nodule* ) AND ( screen* OR detect* ) ) ) AND ( ( TITLE-ABS ( ( acceptabilit* OR acceptance OR acceptable OR attitude* OR belief* OR expectation* OR intent* OR perceiv* OR perception* OR perspective* OR prefer* OR valuation* OR valued OR values OR views OR willing* ) W/2 ( elicit* OR client* OR consumer* OR female* OR health OR individual* OR man OR men OR male OR males OR participant* OR patient* OR people* OR person* OR public* OR respondent* OR screenee* OR smoker* OR stated OR user* OR woman OR women ) ) ) OR ( TITLE-ABS ( ( choice* OR choos* ) W/2 ( behavior* OR behaviour OR discrete OR experiment* ) ) ) OR ( TITLE-ABS ( ( conjoint W/2 ( analy* OR design* OR evaluation OR exercise* OR experiment* OR studies OR study OR survey* OR task* OR valuation OR value* OR valuing ) ) ) ) OR ( TITLE-ABS ( ( contingent W/2 ( analy* OR design* OR evaluation OR exercise* OR experiment*

OR studies OR study OR survey* OR task* OR valuation OR value* OR valuing ) ) ) ) OR ( TITLE-ABS ( decision* W/1 ( aid OR aids OR mak* OR needs OR support* OR tool* OR balanc* ) ) ) OR ( TITLE-ABS ( ranking OR rating OR tradeoff* OR trade-off* ) ) ) ) OR ( ( ( TITLE ( ( bronchial OR bronchus OR lung OR pulmonary ) W/2 ( lesion* OR mass OR masses OR node* OR nodule* ) ) ) OR ( ( TITLE ( ( bronchial OR bronchus OR lung OR pulmonary ) ) ) AND ( ( TITLE-ABS ( ( false*-positiv* OR overdiagnosis OR over-diagnosis OR indetermina* OR suspect* OR suspicious* ) ) ) OR ( TITLE-ABS ( ( extrapulmonary OR extra-pulmonary OR incidental ) W/2 ( discover* OR finding* ) ) ) OR ( TITLE-ABS ( ( bronchial OR bronchus OR lung OR pulmonary ) AND ( ( false*-positiv* OR overdiagnosis OR over-diagnosis ) OR ( diagnos* W/2 error* ) ) ) ) ) ) ) AND ( ( TITLE-ABS ( direct*-elicitation ) ) OR ( TITLE-ABS ( ( daly OR dalys OR qaly OR qalys OR direct*-elicitation* OR disabil*-weight* ) ) ) OR ( TITLE-ABS ( ( disability OR quality ) W/0 adjusted-life ) ) OR ( TITLE-ABS ( eortc-qlq-30c OR euroqol5d OR euroqol OR euro-qol OR euroqual5d OR euroqual OR euro-qual OR eq5d OR eq-5d OR fact-i ) ) OR ( TITLE-ABS ( health W/1 ( utilit* OR disutilit* ) ) ) OR ( TITLE-ABS ( ( health* W/1 year* W/1 equivalent* ) ) ) OR ( TITLE-ABS ( health-status W/1 ( cost* OR estimat* OR index OR indice* OR measure* OR scale* OR score* OR valu* OR weight* ) ) ) OR ( TITLE-ABS ( ( hrqol OR qol OR quality-of-life ) W/2 ( assess* OR instrument* ) ) ) OR ( TITLE-ABS ( hsuv OR husvs OR hui OR hui2 OR hui3 OR hye OR hyes OR multi-attribute OR multi-criteria OR multiattribute OR multicriteria ) ) OR ( TITLE-ABS ( quality W/1 ( wellbeing OR well-being ) ) ) OR ( TITLE-ABS ( sf6 OR sf-6 OR short-form-6 OR shortform-6 OR sf-six OR sfsix OR shortform-six OR "short-form-six" OR shortform6 OR short-form6 OR sf8 OR sf-8 OR sf-eight OR sfeight OR shortform-8 OR shortform-8 OR shortform8 OR short-form8 OR shortform-eight OR short-form-eight OR sf12 OR sf-12 OR short-form-12 OR shortform-12 OR short-form12 OR shortform12 OR sf-twelve OR sftwelve OR shortform-twelve OR short-form-twelve OR sf16 OR sf-16 OR short-form-16 OR shortform-16 OR short-form16 OR shortform16 OR sf-sixteen OR sfsixteen OR shortform-sixteen OR short-form-sixteen OR sf20 OR sf-20 OR short-form-20 OR shortform-20 OR short-form20 OR shortform20 OR sf-twenty OR sftwenty OR shortform-twenty OR short-form-twenty OR sf36 OR sf-36 OR short-form-36 OR shortform-36 OR short-form36 OR shortform36 OR sf-thirtysix OR sfthirtysix OR sfthirty-six OR sf-thirty-six OR shortform-thirtysix OR shortform-thirty-six OR short-form-thirtysix OR short-form-thirty-six ) ) OR ( TITLE-ABS ( standard AND gamble* ) ) OR ( TITLE-ABS ( time W/0 ( tradeoff OR trade-off ) ) ) OR ( TITLE-ABS ( utility W/0 ( cost* OR estimat* OR index OR indice* OR measure* OR scale* OR score* OR valu* OR weight* ) ) ) OR ( TITLE-ABS ( vignette* OR visual-analog*-scale* ) ) ) ) AND PUBYEAR > 2011 AND PUBYEAR < 2024 ) AND NOT ( SRCTYPE ( p ) OR SRCTYPE ( n ) OR SRCTYPE ( w ) )

#### Key Question 3 Search Strategies

**Original searches:**

**Database(s):** Ovid MEDLINE(R) ALL 1946 to December 02, 2022
**Search Strategy**: Lung CA Screening - RPM_MEDLINE

**Date Run:** 5Dec2022

| # | Searches |
|---|---|
| 1 | exp Lung Neoplasms/ |
| 2 | ((bronchial or lung or pulmonary) adj3 (adenocarcinoma or cancer* or carcinoma or lesion* or malignan* or mass$2 or neoplasm* or tumo?r* or nodule*)).ti,ab,kf. |
| 3 | 1 or 2 |
| 4 | Early Detection of Cancer/ |
| 5 | Mass Screening/ |
| 6 | (screen* or detect*).ti,ab,kf. |
| 7 | or/4-6 |
| 8 | 3 and 7 |
| 9 | ((nodule adj3 threshold*) or (nodule adj3 evaluat*) or (nodule adj3 classifi*) or (nodule adj3 categor*)).ti,ab,kf. |
| 10 | (BIMC or Bayesian Inference Malignancy Calculator or "Bayesian Method" or "Brock University cancer prediction equation" or "Lung Nodule Malignancy Risk Calculator" or "McWilliams nodule malignancy score for risk prediction" or "Solitary Pulmonary Nodule Malignancy Risk Score" or "Mayo Clinic Model" or "UKLS Nodule Risk" or UKLS-NRM or uklsnrm or ("UK Lung Screen" adj3 model) or ((BTS or British Thoracic Society) adj4 nodule)).ti,ab,kf. |
| 11 | ((Brock or Herder or Mayo or McWilliams or Pan-CAN or PanCAN or Pan-Canadian Early Detection of Lung Cancer or "Veterans Administration") adj3 (calculat* or score* or risk* or tool or model)).ti,ab,kf. |
| 12 | ("Lung cancer risk assessment tool" or "Lung cancer death risk assessment tool" or LCRAT or LCDRAT or "multiscale lung nodule prediction strategy" or "Vancouver Risk Calculator" or "BTS guidelines" or "British Thoracic Society guidelines").ti,ab,kf. |

| | |
|---|---|
| 13 | ("Lung CT Screening Reporting and Data System" or lung-RADs or lungRADS or I-ELCAP or NODCAT or "Pulmonary Nodule Plasma Proteomic Classifier").ti,ab,kf. |
| 14 | or/9-13 |
| 15 | limit 14 to yr="2012 -Current" |
| 16 | exp "Predictive Value of Tests"/ |
| 17 | exp Risk/ |
| 18 | exp "Sensitivity and Specificity"/ |
| 19 | assessment-tool$1.ti,ab. |
| 20 | (micro-simulation* or microsimulation).ti,ab. |
| 21 | (predict* adj3 (clinical or model* or outcome* or rule$1 or scor*)).ti,ab. |
| 22 | (risk adj3 (assessment$1 or clinical or death or model$1 or mortality or predict* or target*)).ti,ab. |
| 23 | simulation-model$1.ti,ab. |
| 24 | Statistical model$1.ti,ab. |
| 25 | ((Bach or Hoggart or "Hunt Lung Cancer" or Kovalchik or Knoke or "Liverpool lung project risk" or LLP or "UKLS Nodule Risk" or Spitz or Mookgavkar or "Moolgavkar and Knudson" or "Two-Stage Clonal Expansion" or TSCE) adj3 (model or score)).ti,ab. |
| 26 | (PLCO adj3 (calculator or model or score or tool)).ti,ab. |
| 27 | or/16-26 |
| 28 | limit 27 to yr="2019-Current" |
| 29 | 15 or 28 |
| 30 | 8 and 29 |
| 31 | Epidemiologic studies/ |
| 32 | exp case control studies/ |
| 33 | exp cohort studies/ |
| 34 | Case control.tw. |

| | |
|---|---|
| 35 | (cohort adj (study or studies)).tw. |
| 36 | Cohort analy$.tw. |
| 37 | ((Follow-up or followup) adj (study or studies)).tw. |
| 38 | (observational adj (study or studies)).tw. |
| 39 | Longitudinal.tw. |
| 40 | Retrospective.tw. |
| 41 | (Cross-sectional or crosssectional).tw. |
| 42 | Cross-sectional studies/ |
| 43 | ((control and (group* or study)) or (time and factors) or program or survey* or ci or cohort or comparative-stud* or evaluation-studies or follow-up*).mp. |
| 44 | or/31-43 |
| 45 | exp Validation Study/ |
| 46 | exp "Sensitivity and Specificity"/ |
| 47 | (validat* or validity or sensitiv* or specific* or reliability or reliable).ti,ab,kf. |
| 48 | or/45-47 |
| 49 | 44 or 48 |
| 50 | 30 and 49 |
| 51 | (Case Reports.pt. or (case report? or case study or case studies).ti.) not (review* or trial*).ti,ab,kf,hw. |
| 52 | comment/ or editorial/ or (comment or editorial or news or newspaper article).pt. |
| 53 | (Case Reports.pt. or (case report? or case study or case studies).ti.) not (review* or trial*).ti,ab,kf,hw. |
| 54 | comment/ or editorial/ or (comment or editorial or news or newspaper article).pt. |
| 55 | (exp Animals/ or Models, Animal/ or Disease Models, Animal/) not Humans/ |
| 56 | ((animal or animals or canine* or dog or dogs or feline or hamster* or lamb or lambs or mice or monkey or monkeys or mouse or |

| | |
|---|---|
| | murine or pig or pigs or piglet* or porcine or primate* or rabbit* or rats or rat or rodent* or sheep* or veterinar*) not (human* or patient*)).mp. |
| 57 | 50 not (or/51-56) |
| 58 | remove duplicates from 57 |

**Database(s):** Embase 1974 to 2022 December 02
**Date searched:** 5Dec2022

| # | Searches | Results | Annotations |
|---|---|---|---|
| 1 | exp lung tumor/ | 453898 | |
| 2 | ((bronchial or lung or pulmonary) adj3 (adenocarcinoma or cancer* or carcinoma or lesion* or malignan* or mass$2 or neoplasm* or tumo?r* or nodule*)).ti,ab,kw. | 426978 | |
| 3 | 1 or 2 | 581292 | LungCA |
| 4 | early cancer diagnosis/ | 12003 | |
| 5 | Mass Screening/ | 60373 | |
| 6 | (screen* or detect*).ti,ab,kw. | 4419343 | |
| 7 | or/4-6 | 4438299 | Screening |
| 8 | 3 and 7 | 107257 | LungCA Screening |
| 9 | ((nodule adj3 threshold*) or (nodule adj3 evaluat*) or (nodule adj3 classifi*) or (nodule adj3 categor*)).ti,ab,kw. | 1510 | |
| 10 | (BIMC or Bayesian Inference Malignancy Calculator or "Bayesian Method" or "Brock University cancer prediction equation" or "Lung Nodule Malignancy Risk Calculator" or "McWilliams nodule malignancy score for risk prediction" or "Solitary Pulmonary Nodule Malignancy Risk Score" or "Mayo Clinic Model" or "UKLS Nodule Risk" or UKLS-NRM or uklsnrm or ("UK Lung Screen" adj3 model) or ((BTS or British Thoracic Society) adj4 nodule)).ti,ab,kw. | 2377 | |
| 11 | ((Brock or Herder or Mayo or McWilliams or Pan-CAN or PanCAN or Pan-Canadian Early Detection of Lung Cancer or "Veterans Administration") adj3 (calculat* or score* or risk* or tool or model)).ti,ab,kw. | 7777 | |
| 12 | ("Lung cancer risk assessment tool" or "Lung cancer death risk assessment tool" or LCRAT or LCDRAT or "multiscale lung nodule prediction strategy" or "Vancouver Risk Calculator" or "BTS guidelines" or "British Thoracic Society guidelines").ti,ab,kw. | 834 | |

| | | | |
|---|---|---|---|
| 13 | ("Lung CT Screening Reporting and Data System" or lung-RADs or lungRADS or I-ELCAP or NODCAT or "Pulmonary Nodule Plasma Proteomic Classifier").ti,ab,kw. | 503 | |
| 14 | or/9-13 | 12796 | |
| 15 | limit 14 to yr="2012 -Current" | 10138 | Nodule 2012-current |
| 16 | exp "Predictive Value of Tests"/ | 228207 | |
| 17 | exp Risk/ | 2932708 | |
| 18 | exp "Sensitivity and Specificity"/ | 453740 | |
| 19 | assessment-tool$1.ti,ab. | 48119 | |
| 20 | (micro-simulation* or microsimulation).ti,ab. | 2758 | |
| 21 | (predict* adj3 (clinical or model* or outcome* or rule$1 or scor*)).ti,ab. | 466660 | |
| 22 | (risk adj3 (assessment$1 or clinical or death or model$1 or mortality or predict* or target*)).ti,ab. | 483304 | |
| 23 | simulation-model$1.ti,ab. | 14260 | |
| 24 | Statistical model$1.ti,ab. | 18956 | |
| 25 | ((Bach or Hoggart or "Hunt Lung Cancer" or Kovalchik or Knoke or "Liverpool lung project risk" or LLP or "UKLS Nodule Risk" or Spitz or Mookgavkar or "Moolgavkar and Knudson" or "Two-Stage Clonal Expansion" or TSCE) adj3 (model or score)).ti,ab. | 181 | |
| 26 | (PLCO adj3 (calculator or model or score or tool)).ti,ab. | 29 | |
| 27 | or/16-26 | 3933004 | |
| 28 | limit 27 to yr="2012 -Current" | 2493843 | Other RPM: 2012-current |
| 29 | 15 or 28 | 2501050 | Nodule 2012-current OR Other RPM 2012-current |
| 30 | 8 and 29 | 16839 | LungCA screening + (nodule or RPM) |
| 31 | exp case control study/ | 214298 | |
| 32 | cohort analysis/ | 928351 | |

| | | | |
|---|---|---|---|
| 33 | Case control.ti,ab. | 195011 | |
| 34 | (cohort adj (study or studies)).ti,ab. | 424140 | |
| 35 | Cohort analy$.ti,ab. | 17786 | |
| 36 | ((Follow-up or followup) adj (study or studies)).ti,ab. | 72558 | |
| 37 | (observational adj (study or studies)).ti,ab. | 232507 | |
| 38 | Longitudinal.ti,ab. | 412719 | |
| 39 | Retrospective.ti,ab. | 1164584 | |
| 40 | (Cross-sectional or crosssectional).ti,ab. | 630666 | |
| 41 | ((control and (group* or study)) or (time and factors) or program or survey* or ci or cohort or comparative-stud* or evaluation-studies or follow-up*).mp. | 10113066 | |
| 42 | or/31-41 | 11031433 | Non-RCT study designs |
| 43 | exp validation study/ | 100292 | |
| 44 | exp "sensitivity and specificity"/ | 453740 | |
| 45 | (validat* or validity or sensitiv* or specific* or reliability or reliable).ti,ab,kw. | 7026833 | |
| 46 | or/43-45 | 7151524 | Validation studies |
| 47 | 42 or 46 | 15782229 | Non-RCTs or validation studies |
| 48 | 30 and 47 | 14296 | LungCA Screening + (nodule 2012-current OR other RPM 2012-current) + Study Designs |
| 49 | (Case Reports.pt. or (case report? or case study or case studies).ti.) not (review* or trial*).ti,ab,kf,hw. | 330984 | |
| 50 | comment/ or editorial/ or (comment or editorial).pt. | 787759 | |
| 51 | (conference abstract* or conference paper*).af. not (conference abstract* or conference paper*).ab,ga. | 5400382 | |
| 52 | conference proceeding*.af. not conference proceeding*.ab,ga. | 39397 | |
| 53 | exp review/ or meta analysis/ or consensus/ or exp practice guideline/ | 3715024 | |

| | | |
|---|---|---|
| 54 | review.pt. or (review or meta-analysis or metaanalysis or guideline*).ti. | 3532818 |
| 55 | (exp animal/ or exp animal model/) not human/ | 5292532 |
| 56 | ((animal or animals or canine* or dog or dogs or feline or hamster* or lamb or lambs or mice or monkey or monkeys or mouse or murine or pig or pigs or piglet* or porcine or primate* or rabbit* or rats or rat or rodent* or sheep* or veterinar*) not (human* or patient*)).mp. | 4846079 |
| 57 | 48 not (or/49-56) | 7083 |

Search updates for KQ3 in September 2025

Database(s): Ovid MEDLINE(R) ALL 1946 to September 22, 2025

| # | **Searches** |
|---|---|
| 1 | exp Lung Neoplasms/ |
| 2 | ((bronchial or lung or pulmonary) adj3 (adenocarcinoma or cancer* or carcinoma or lesion* or malignan* or mass$2 or neoplasm* or tumo?r* or nodule*)).ti,ab,kf. |
| 3 | 1 or 2 [LUNG CA] |
| 4 | Early Detection of Cancer/ |
| 5 | Mass Screening/ |
| 6 | (screen* or detect*).ti,ab,kf. |
| 7 | or/4-6 [SCREENING] |
| 8 | 3 and 7 [LUNG CA SCREENING] |
| 9 | ((nodule adj3 threshold*) or (nodule adj3 management) or (nodule adj3 classifi*) or (nodule adj3 categor*)).ti,ab,kf. |
| 10 | ((Pan-CAN or PanCAN or "Pan-Canadian Early Detection of Lung Cancer") adj3 (calculat* or score* or risk* or tool or model)).ti,ab,kf. |

11 ("Lung CT Screening Reporting and Data System" or "Lung Imaging Reporting and Data Systems" or "Lung Imaging Reporting and Data System" or "Lung Reporting and Data System" or lung-RADs or lungRADS).ti,ab,kf.

12 or/9-11

13 limit 12 to yr="2012 -Current" [Date limit: nodule, PAN-CAN, LungRADS]

14 ((eligib* or selecting or selection) adj3 screening).ti,ab.

15 ((PLCO* or "Prostate Lung Colorectal and Ovarian Cancer Screening Trial") adj3 (calculator or model or risk or score or tool)).ti,ab.

16 or/14-15

17 limit 16 to yr="2019-Current" [Date limit: Eligibility/PLCO]

18 13 or 17 [RPM of interest]

19 8 and 18 [LUNG CA SCREENING - RPM]

20 exp cohort studies/

21 (cohort adj (study or studies)).tw.

22 Cohort analy$.tw.

23 ((Follow-up or followup) adj (study or studies)).tw.

24 (observational adj (study or studies)).tw.

25 Longitudinal.tw.

26 Retrospective.tw.

27 ((control and (group* or study)) or (time and factors) or program or survey* or ci or cohort or comparative-stud* or evaluation-studies or follow-up*).mp.

28 or/20-27 [NON-RCT FILTER]

29 exp Validation Study/

30 exp "Sensitivity and Specificity"/

31 ((validat* or validity) adj3 (external or cohort)).ti,ab,kf.

32 or/29-31 [VALIDATION STUDY FILTER]

33 28 or 32

34 19 and 33 [LUNG CA SCREENING RPM - NON-RCTs, VALIDATION STUDIES]

35 (Case Reports.pt. or (case report? or case study or case studies).ti.) not (review* or trial*).ti,ab,kf,hw.

36 comment/ or editorial/ or (comment or editorial or news or newspaper article).pt.

37 ((animal or animals or canine* or dog or dogs or feline or hamster* or lamb or lambs or mice or monkey or monkeys or mouse or murine or pig or pigs or piglet* or porcine or primate* or rabbit* or rats or rat or rodent* or sheep* or veterinar*) not (human* or patient*)).mp.

38 34 not (or/35-37) [REMOVE ANIMALS, OPINION PIECES]

39 remove duplicates from 38

—

Database(s): Embase 1974 to 2025 September 22

| # | Searches |
|---|---|
| 1 | exp lung tumor/ |
| 2 | ((bronchial or lung or pulmonary) adj3 (adenocarcinoma or cancer* or carcinoma or lesion* or malignan* or mass$2 or neoplasm* or tumo?r* or nodule*)).ti,ab. |
| 3 | 1 or 2 [LUNG CA] |
| 4 | early cancer diagnosis/ |
| 5 | Mass Screening/ |
| 6 | (screen* or detect*).ti,ab. |
| 7 | or/4-6 [SCREENING] |
| 8 | 3 and 7 [LUNG CA SCREENING] |
| 9 | ((nodule adj3 threshold*) or (nodule adj3 management) or (nodule adj3 classifi*) or (nodule adj3 categor*)).ti,ab. |
| 10 | ((Pan-CAN or PanCAN or "Pan-Canadian Early Detection of Lung Cancer") adj3 (calculat* or score* or risk* or tool or model)).ti,ab. |
| 11 | ("Lung CT Screening Reporting and Data System" or "Lung Imaging Reporting and Data Systems" or "Lung Imaging Reporting and Data System" or "Lung-Reporting and Data System" or lung-RADs or lungRADS).ti,ab. |
| 12 | or/9-11 |
| 13 | limit 12 to yr="2012 -Current" [Date limit: nodule, PAN-CAN, LungRADS] |

14 ((eligib* or selecting or selection) adj3 screening).ti,ab.

15 ((PLCO* or "Prostate Lung Colorectal and Ovarian Cancer Screening Trial") adj3 (calculator or model or risk or score or tool)).ti,ab.

16 or/14-15

17 limit 16 to yr="2019 -Current" [Date limit: Eligibility/PLCO]

18 13 or 17

19 8 and 18 [LUNG CA SCREENING - RPM]

20 cohort analysis/

21 (cohort adj (study or studies)).ti,ab.

22 Cohort analy$.ti,ab.

23 ((Follow-up or followup) adj (study or studies)).ti,ab.

24 (observational adj (study or studies)).ti,ab.

25 Longitudinal.ti,ab.

26 Retrospective.ti,ab.

27 ((control and (group* or study)) or (time and factors) or program or survey* or ci or cohort or comparative-stud* or evaluation-studies or follow-up*).mp.

28 or/20-27 [NON-RCT FILTER]

29 exp validation study/

30 exp "sensitivity and specificity"/

31 (validat* or validity or sensitiv* or specific* or reliability or reliable).ti,ab,kw.

32 or/29-31 [VALIDATION STUDY FILTER]

33 28 or 32

34 19 and 33 [LUNG CA SCREENING RPM - NON-RCTs, VALIDATION STUDIES]

35 (Case Reports.pt. or (case report? or case study or case studies).ti.) not (review* or trial*).ti,ab,kf,hw.

36 comment/ or editorial/ or (comment or editorial).pt.

37 (conference abstract* or conference paper*).pt.

38 exp review/ or meta analysis/ or consensus/ or exp practice guideline/

39 review.pt. or (review or meta-analysis or metaanalysis or guideline*).ti.

40 ((animal or animals or canine* or dog or dogs or feline or hamster* or lamb or lambs or mice or monkey or monkeys or mouse or murine or pig or pigs or piglet* or porcine or primate* or rabbit* or rats or rat or rodent* or sheep* or veterinar*) not (human* or patient*)).mp.

41 34 not (or/35-40) [REMOVE ANIMALS, OPINION PIECES]

42 remove duplicates from 41

Table S1.1 Eligibility criteria for key question 1 on benefits and harms of screening for lung cancer with computed tomography

| | Inclusion Criteria | Exclusion Criteria |
|---|---|---|
| Population | Adults ≥ 18 years old and not suspected to have lung cancer<br>Specific population of interest may include:<br>• Age<br>• Sex<br>• Smoking history (e.g., duration, pack-years)<br>• Race and ethnicity (e.g., First Nations, Inuit and Métis)<br>• Populations for which screening access and outcomes may be inequitable (e.g., LGBTQ+, low socioeconomic status, homeless) | • Studies of people seeking care for symptoms of lung cancer or who are suspected of lung cancer<br>• Studies of people who have been previously diagnosed with lung cancer<br>• Studies of people younger than 18 years old<br>• Study population includes >25% individuals with recent abnormal screening result<br>• Revied to add studies focusing on specific populations (e.g., HIV, other cancers) |
| Intervention | Computed tomography (CT), with or without any other lung cancer screening interventions (e.g. biomarkers).<br><br>The focus is on screening interventions involving CT but we will include new RCTs (published in 2015 or later) of other screening modalities. Screening modalities (i.e. chest x-ray) examined in the last review (chest x-ray) will only be included if new RCTs are found. | |
| Comparator | • No screening/placebo/minimally or non-active intervention (e.g., smoking cessation)<br>• An alternative screening strategy/protocol (e.g., use of risk-prediction model vs risk-factor based screening criteria, different screening interval or method to classify nodules)<br>• None (only for harm outcomes except overdiagnosis where a no screening arm is required) | |
| Outcomes (by importance as rated by the WG) | Benefits:<br>1. All-cause mortality*<br>2. Lung cancer mortality*<br>3. Health-related quality of life<br><br>If there is trial evidence showing at least low certainty of benefit for 1+ of the above outcomes, then:<br><br>Harms:<br>4. Overdiagnosis of lung cancer*<br>5. Major complications or morbidity from invasive testing as a result of screening<br>6. Death from invasive testing as a result of screening (among those without cancer)<br>7. False positives (all and among those requiring invasive workup)<br>8. Psychosocial consequences of the screening process (i.e., before or right after screening, receiving a false positive result)<br>9. Incidental findings (all and clinically relevant)<br><br>*Rated as critical outcomes; others rated as important | |
| Timing | No limitation on the duration of follow-up, except for FPs where ≥12 mos follow-up after the screening result is required in ≥80% of participants (if there is no evidence found meeting this criteria we will accept ≥6 mos follow-up).<br><br>Longest follow-up will be used for mortality, quality of life, and overdiagnosis, unless there has been substantial (>20%) contamination by the control group receiving screening (revised to use lower risk of bias findings among different follow-ups). | |
| Delivery Setting | Any setting relevant to primary care | |

| | Inclusion Criteria | Exclusion Criteria |
|---|---|---|
| | Countries rated as very high on the Human Development Index 2019 (59) | |
| Study design & publication type | • RCTs (parallel designs using individual or cluster randomization)<br>• With the exception of overdiagnosis, for harms nonrandomized studies are also eligible. For false positives, the study must use similar (revised to remove section criteria and) methods of determining a positive result (e.g., diameter, volumetric) as used in an RCT. If a method used in an RCT is compared with another method in a nonrandomized study these will be considered in KQ3. For psychosocial harms there must be at least a within-group comparison relevant to the outcome e.g., before and after the screening test, between people with a negative vs. a positive test.<br><br><u>Note</u>: nonrandomized studies comparing the benefits of screening selection criteria or the nodule classification systems used in the RCTs with use of a risk prediction model or a different nodule classification system will be considered in KQ3.<br><br>Journal articles<br>Letters, abstracts and grey literature (e.g., government reports, results in trial registries) if information on study design (e.g., eligibility criteria, participant characteristics, presentation of scenarios) is sufficient to assess study and results are confirmed as final (accessible online or via author contact). | • Studies using modeling or simulation, or retrospectively applying differing screening criteria<br>• Editorials, commentaries<br>• Case reports and series (i.e. all participants have the outcome) |
| Language of full text | English or French | |
| Dates of publication | Any date | |

Table S1.2 Eligibility criteria for key question 2 on the relative importance people place on the potential benefits and harms from screening for lung cancer

| | Inclusion | Exclusion |
|---|---|---|
| **Population** | Adults aged 18 years and older (revised to 45 years and older, and eligible for screening)<br><br><u>Specific populations of interest may include:</u><br>– Age<br>– Sex<br>– Smoking history<br>– Race or ethnicity (e.g. First Nations, Inuit and Métis)<br>– Populations for which screening access and outcomes may be inequitable (e.g. LGBTQ+, low socioeconomic status, homeless)<br>– For health-state utility studies: cancer detection via screening vs. clinical presentation; stage/severity of cancer (i.e., curative vs. palliative/non-metastatic vs. metastatic; revised to stage I-IIIa vs. Stage IIIb/IV due to reporting) | Expert or healthcare providers (doctor, nurse) acting as proxy for patients and public |
| **Exposures** | **a) Direct measurements:**<br><u>Utility-based measurements</u> (e.g., health state utilities, trade-offs between outcomes)<br>i. Experience with outcome/health state (as per KQ1, adding different stages/severities of cancer)<br>ii. Exposure to clinical scenarios about the outcome(s)<br>iii. Exposure to choice sets or other risk exercises (e.g. trade-offs, balance sheet, ranking) with differing risks/magnitudes of effects on benefits versus harms from screening *(must contain 1+ benefit and 1+ harm)*<br><br><u>Quantitative non-utility studies</u> (e.g. simple ratings, rankings or trade-offs between 1+ benefit and 1+ harm)<br>i. Any exposure<br><br>**b) Indirect measurements** (allowing inferences about how many people perceive benefits as more important than harms & acceptability of screening; not only in context of critical outcomes):<br>i. Exposure to estimates of effect from screening for 1+ benefit and 1+ harm (e.g. decision aids). | |
| **Comparisons** | **a) Utility-based measurements:**<br>i. Healthy/usual state without outcome (may include screen-negative patients)<br>ii. Different outcome/health state (e.g. false positives vs. overdiagnosis; includes studies comparing different stages/severity of cancers)<br>iii. No comparison, if no other studies for the outcome comparison<br><br>**b) Quantitative non-utility studies & indirect studies**<br>i. No comparison<br>ii. Comparison with another screening strategy (e.g., having different magnitude of effects; based on comparisons evaluated in evidence found for KQ1) | Studies measuring utilities during cancer treatments that are not considered a standard first-line of care for a representative sample, by stage of cancer (will gather clinical input as needed) |
| **Outcomes** | **a) Direct measurements:**<br>i. Health-state utility values; health states may include different stages/severities of lung cancer and nodules to help estimate the utility of an overdiagnosed case<br>ii. Other utility scores (e.g. trade-offs, magnitude of coefficients/utility weights in models for each outcome)<br>iii. Non-utility, quantitative information about relative importance of different benefits and harms (ratings, rankings)<br>**b) Indirect measurements:**<br>i. Inferences about how many people perceive benefits as greater than harms<br>ii. Preference for or against screening (screening attendance, intentions, or acceptance) or preferred screening strategy based on different outcome risk descriptions (e.g. using decision aids)<br>**c) Measures of variability for all of above** (e.g. 95% CIs, proportion unwilling to trade any benefit for harm, proportion undecided from decision aids vs those confident in decision for vs. against) | |

| | Inclusion | Exclusion |
|---|---|---|
| **Timing** | Measured immediately after experiencing outcome/diagnosis (to 3 months) an longer term (after treatment): revised to a) at diagnosis or before treatment initiation (treatment naïve), b) on first-line treatment and c) after first-line treatment completion and/or on maintenance therapy. | |
| **Setting** | Any setting in Very High Human Development Index countries (59) | |
| **Study Design and Publication Status** | Any quantitative study design<br><br>Journal articles<br>Letters, abstracts and grey literature (e.g., government reports, results in trial registries) if information on study design (e.g., eligibility criteria, participant characteristics, presentation of scenarios) is sufficient to assess study and results are confirmed as final (accessible online or via author contact). | Editorials. |
| **Language** | English or French | |
| **Publication date** | 2012 – present | |

Table S1.3 Eligibility criteria for key question 3 on the comparative effects between a) trial-based selection criteria and use of risk prediction models, and b) trial-based nodule classification and different nodule classification systems

| | **Inclusion** | **Exclusion** |
|---|---|---|
| Population | Adults aged 18 years and older not suspected to have lung cancer. | Studies that focused on people under age 18 or that targeted adults 18 years and older who were either suspected of having lung cancer or were previously diagnosed with lung cancer. |
| Intervention(s) | For risk prediction models used for screening eligibility:<br>Externally validated risk prediction models intended for identifying persons who may benefit from screening.<br>**The subsequent screening process must be similar to trials included in KQ1 (e.g. LDCT, nodule classification)**<br>Note: The main purpose of the study may be to provide external validation.<br><br>For nodule classification systems: any nodule classification system.<br>**Other elements of screening must be similar to trial included in KQ1.** | Risk prediction models that include factors (laboratory tests, other assessment) that are not available or feasible in primary care.<br><br>Risk prediction models used for screening eligibility that incorporate nodule characteristics. |
| Comparison(s) | For risk prediction models:<br>Trial-based eligibility criteria (similar to that used by trials included in KQ1 (e.g. NLST, NELSON)).<br><br>For nodule classification systems:<br>Nodule classification system similar to those used by trials included in KQ1 (e.g. NLST, NELSON). | |
| Outcomes | Benefits:<br>1. All-cause mortality*<br>2. Lung cancer mortality*<br>3. Health-related quality of life<br><br>Harms:<br>4. Overdiagnosis of lung cancer*<br>5. Major complications or morbidity from invasive testing as a result of screening<br>6. Death from invasive testing as a result of screening<br>7. False positives<br>8. Psychosocial consequences of the screening process (i.e., before or right after screening, receiving a false positive result)<br>9. Incidental findings<br><br>*Rated as critical outcomes; others rated as important<br><br>Note: outcome may be reported as a relative measure between benefit(s) and harm, e.g. number of overdiagnosed cancers per prevented death | False positive selections (individual is positive for “increased risk” using model [i.e. is selected for screening with CT or other modality] but does not get cancer or die from lung cancer during follow up) |
| Delivery Setting | Any setting relevant to primary care | |
| Study design(s) & Publication status | Nonrandomized clinical trials, prospective or retrospective controlled observational studies, modelling studies<br><br>Journal articles<br>Letters, abstracts and grey literature (e.g., government reports, results in trial registries) if information on study design (e.g., eligibility criteria, participant characteristics, presentation of scenarios) is sufficient to assess study and results are confirmed as final (accessible online or via author contact). | -RCTs (captured in KQ1)<br><br>- Editorials<br>- Commentaries<br>- Case reports and series |

| | | |
|---|---|---|
| Countries | Studies conducted on populations from countries categorized as “Very High” on the 2016 Human Development Index (as defined by the United Nations Development Programme).<br><br>Priority will be given to studies relevant to the Canadian context | Studies conducted on populations from countries NOT categorized as “Very High” on the 2016 Human Development Index (as defined by the United Nations Development Programme). |
| Language of full text | English or French | Languages other than English and French. |
| Dates of publication | 2012 (publication of first RCT showing benefit of lung cancer screening was Aug 2011) | |

Table S1.4 Overdiagnosis calculations and descriptions

| Description and calculations |
|---|
| **Overdiagnosis: Excess incidence (used for main analysis and GRADE):**<br><br>[{Total no. of lung CA detected in intervention arm/ N enrolled} – {total no. of lung CA detected in control arm / N enrolled}] *100<br><br>During active screening phase and follow-up (taking into account interval cancers) |
| **Overdiagnosis: excess incidence from screening**<br><br>[{Total no. of lung CA detected by screening in intervention arm/ N enrolled} – {total no. of lung CA detected in control arm / N enrolled}] *100 |
| **Overdiagnosis: excess incidence of lung cancer from screening among those having cancer diagnosed in the screening arm**<br><br>[{Total no. of lung CA detected by screening in intervention arm/ N enrolled} – {total no. of lung CA detected in control arm / N enrolled} / total no. diagnosed with cancer in screening arm] |
| **Overdiagnosis: excess incidence of lung CA from screening among those having cancer diagnosed through screening in the screening arm**<br><br>[{Total no. of lung CA detected by screening in intervention arm/ N enrolled} – {total no. of lung CA detected in control arm / N enrolled}/ total no. diagnosed with cancer via screening in screening arm] |
| **Overdiagnosis: overdiagnosis of screen-detected cancer**<br><br>**Formula from Brodersen, et al.**<br><br>[{Cumulative cancer incidence in intervention arm/N enrolled} - {cumulative cancer incidence in control arm/N enrolled} / {cumulative number of screen-detected cancers in intervention arm/ N enrolled in intervention arm}] |

Brodersen J, Voss T, Martiny F, Siersma V, Barratt A, Heleno B. Overdiagnosis of lung cancer with low-dose computed tomography screening: meta-analysis of the randomised clinical trials. Breathe (Sheff). 2020;16(1):200013. doi: 10.1183/20734735.0013-2020. PMID: 32194774.

### Additional file 2. Additional tables and figures for key question 1

Figure S2.27 Proportion with a false positive at round 1 ... 135
Figure S2.28 Proportion with a false positive over 2/3 rounds ... 136
Figure S2.29 Proportion with a false positive over 4/5 rounds ... 137
Figure S2.30 Proportion with incidental findings at round 1 ... 138
Figure S2.31 Proportion with incidental findings over 2/3 rounds ... 139
Figure S2.32 Proportion with incidental findings over 4/5 rounds ... 140
Figure S2.35 Proportion with clinically significant incidental findings over 4/5 rounds ... 143
Figure S2.36 Proportion with major complications from invasive testing at round 1 ... 144
Figure S2.37 Proportion with major complications from invasive testing over 2/3 rounds ... 145
Figure S2.38 Proportion with major complications from invasive testing over 4/5 rounds ... 146
Figure S2.39 Proportion with major complications from invasive testing among false positives at round 1 ... 147
Figure S2.40 Proportion with major complications from invasive testing among false positives over 2/3 rounds ... 148
Figure S2.41 Proportion with major complications from invasive testing among false positives over 4/5 rounds ... 149
Figure S2.42 Proportion of deaths from invasive testing among false positives at round 1 ... 150
Figure S2.43 Proportion of deaths from invasive testing among false positives over 2/3 rounds ... 151
Figure S2.44 Proportion of deaths from invasive testing among false positives over 4/5 rounds ... 152
Table S2.10 CT screening: Important outcomes sensitivity and subgroup analyses ... 153
Table S2.11 Additional harms outcome data ... 158
Table S2.12 Targeted CT screening: Summary of findings for benefit outcomes ... 162
Figure S2.45 Lung cancer mortality, ECLS trial ... 164
Figure S2.46 Lung cancer mortality, LungSEARCH trial ... 164
Figure S2.47 All-cause mortality, ECLS trial ... 164
Figure S2.48 All-cause mortality, LungSEARCH trial ... 165

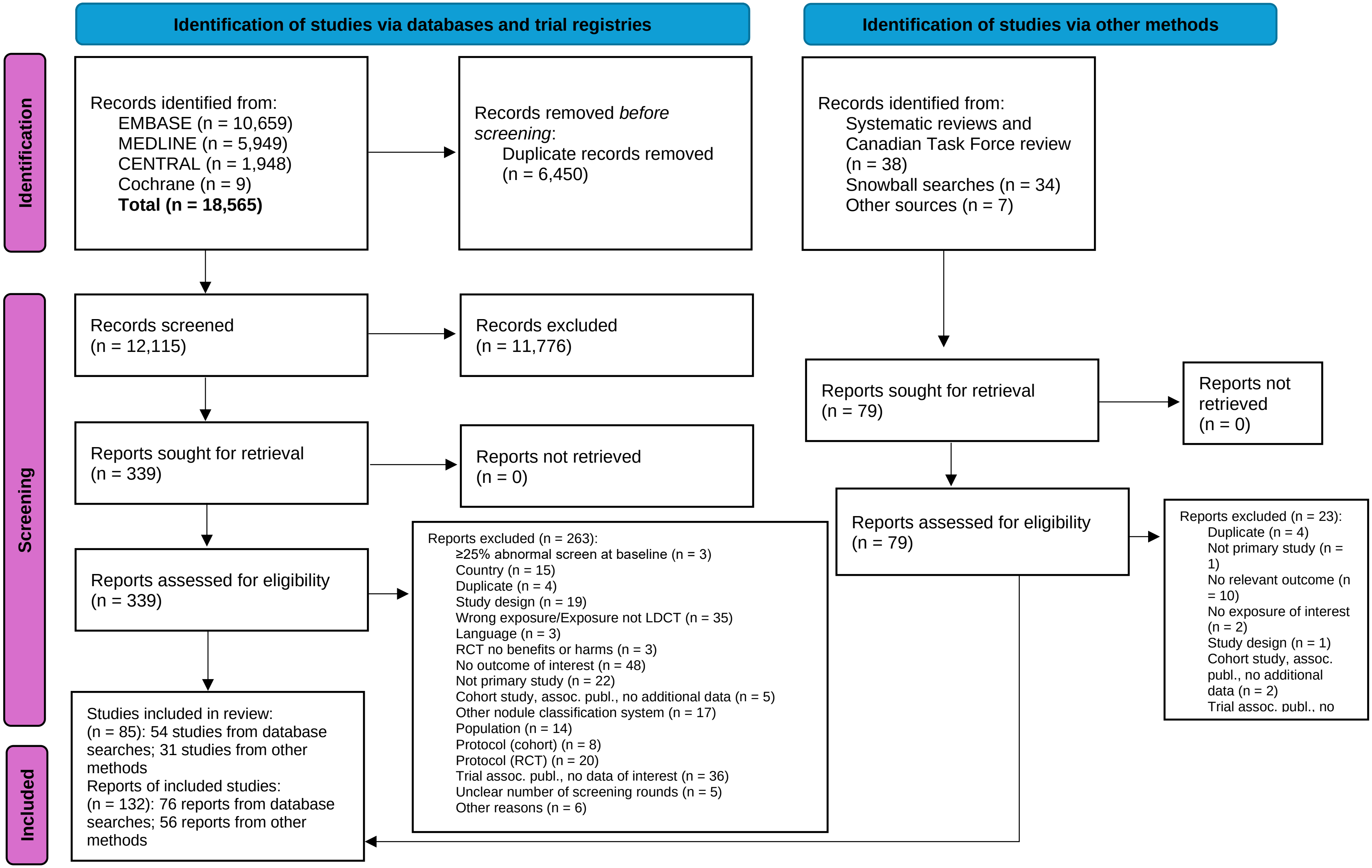

Figure S2.1 KQ1 Literature flow diagram

Table S2.1 Characteristics of the trials on benefits and harms of screening for lung cancer with computed tomography

| Characteristic | NLST | NELSON | DLCST | DANTE | ITALUNG | LUSI | MILD | UKLS | LSS* | ECLS | LungSEARCH |
|---|---|---|---|---|---|---|---|---|---|---|---|
| **Country** | USA | Netherland, Belgium | Denmark | Italy | Italy | Germany | Italy | UK | USA | UK | UK |
| **Eligibility** | | | | | | | | | | | |
| Age range, years | 55-74 | 50-74 | 50-70 | 60-74 | 55-69 | 50-69 | 49-75 | 50-75 | 55-74 | 50-75 | Adults *Mean 63.0 at randomization |
| Smoking history | Current or former (had quit within 15 years) smokers & with ≥30 person years | Current or former (quit ≤10 years ago) smokers who smoked >15 cigarettes/ day for >25 years or >10 cigarettes/day for >30 years | Current or former (quit after the age of 50 years & within 10 years) smokers with ≥20 person years | Current or former male smokers with ≥20 person years who had quit within 10 years | Current or former smokers with ≥20 person years during the preceding 10 years | Current or former smokers (quit within 10 years) with ≥25 years smoking of ≥15 cigarettes/day, or ≥30 years smoking of ≥10 cigarettes/day | Current or former smokers (quit within 10 years) with ≥20 person years | NA but >99% current or former (as per high-risk criteria) | Current or former smokers (had quit within 10 years), **& with ≥30** person years | Current or former smokers with ≥20 person years, or with a history of <20 person years & immediate family history of lung cancer | Current or former smokers (had quit within 8 years) with ≥20 person years and/or had smoked for ≥20 years |
| Other criteria | - | - | FEV1 ≥30% of the predicted value at baseline | - | - | - | - | High risk of lung cancer (i.e., ≥4.5% over 5 years) based on the $LLP_{v2}$ Risk Prediction Model | - | Eastern Cooperative Oncology Group performance status of 0-2 at recruitment | Mild (FEV1/ FVC <70%; FEV1 ≥80% predicted) or moderate (FEV1/FVC <70%; FEV1 50-80% predicted) COPD |
| **Total randomized, N**<br>- Intervention<br>- Comparator | 53,454<br>- 26,722<br>- 26,732 | 15,792<br>- 7,900<br>- 7,892 | 4,104<br>- 2,052<br>- 2,052 | 2,450<br>- 1,264<br>- 1,186 | 3,206<br>- 1,613<br>- 1,593 | 4,052<br>- 2,029<br>- 2,023 | 4,099<br>- 2,376**<br>- 1,723 | 4,055<br>- 2,028<br>- 2,027 | 3,318<br>- 1,660<br>- 1,658 | 12,209<br>- 6,088<br>- 6,121 | 1,568<br>- 785<br>- 783 |
| **Male, %** | 59.0 | 83.6 | 55.2 | 100.0 | 64.7 | 64.7 | 66.3 | 76.0 | 58.6 | 50.9 | 52.2 |
| **Ethnicity, %** | White: 90.8<br>Black: 4.4<br>Others: 4.8 | NR | NR | NR | NR | NR | NR | White: 99.0<br>Non-white: 1.0 | NR | NR | NR |
| **Intervention** | LDCT | LDCT | LDCT | LDCT | LDCT | LDCT | LDCT | LDCT | LDCT | LDCT | LDCT & AFB |

| Characteristic | NLST | NELSON | DLCST | DANTE | ITALUNG | LUSI | MILD | UKLS | LSS* | ECLS | LungSEARCH |
|---|---|---|---|---|---|---|---|---|---|---|---|
| | | | | | | | | | | * If EarlyCDT-Lung test was positive, baseline CXR & LDCT were undertaken | *If sputum cytology was abnormal |
| Number of rounds | 3 | 4 | 5 | 5 | 4 | 5 | 4 or 7 | 1 | 2 | 5 | Variable *Depended on sputum test and nodule size |
| Screening interval(s) | 1-year | 1-, 2-, & 2.5-years (i.e., year 1, 3, & 5.5) | 1-year | 1-year | 1-year | 1-year | 1- or 2-years | NA | 1-year | 6-month | 1-year |
| Screening attendance rate, %‡<br>- Baseline<br>- Final round | - 98.5<br>- 94 | - 95.6<br>- 66.8 | - 99.8<br>- 90.2 | - 100.0<br>- 93.7 | - 87.1<br>- 79.8 | - 99.9<br>- 89.2 | - 100.0<br>- 80.5 | - 98.3<br>- NA | - 96.0<br>- 84.2 | - 97.0<br>- 83.0<br>- (of those assigned to LDCT) | - LDCT uptake at any time: 91.6% |
| **Comparator** | CXR at 1-year intervals for 3 rounds | No screening | No screening | No screening (both arms received CXR at baseline) | No screening | No screening | No screening | No screening | CXR at 1-year intervals for 2 rounds | No screening | No screening |
| **Received by both arms** | - | Computer-tailored smoking cessation for current smokers | Minimal smoking cessation counselling (<5 min); annual visits included spirometry | CXR and 3-day sputum cytology testing at baseline; annual clinical review | Free-access invitation to smoking cessation program | Quit-smoking counseling; blood sample taken at baseline for concomitant biomarker research | Primary prevention program (smoking cessation) | Smoking cessation advice in current smokers | - | Smoking cessation advice | Exit chest radiograph 5 years post-randomization, offered if no lung cancer developed |
| **Image interpretation technique** | Built-in reader functions on clinical workstation (CWS) | Semiautomated software (LungCare, version Somaris/5 VA70C-W) | Cine/slab-viewing supplied with Maximum Intensity Projection | Hard copies of scans complemented by use of workstations later in the | Nodule size, measured manually with electronic callipers on | 2D image representation & subsequent evaluation with 3D | Semi-automatic CAD software (LungCare, Oncology) | Syngo Siemens (Forcheim, Germany) workstation; no computer | NR | NR | Computer workstations with standard lung /soft tissue and bone settings |

| Characteristic | NLST | NELSON | DLCST | DANTE | ITALUNG | LUSI | MILD | UKLS | LSS* | ECLS | LungSEARCH |
|---|---|---|---|---|---|---|---|---|---|---|---|
| | without computer-assisted image analysis | | | study (in 2004) | the work station | depiction using CAD (MEDIAN) | | aided detection software used | | | |
| **Reader(s)** | Radiologists certified by American Board of Radiology or equivalent & had training & experience (including the interpretation of 300 chest CT scans in previous 3 years); comparative interpretations to determine scan results | First two rounds: independent dual reading by radiologists (first reader: 0-20 years of experience, second reader: 6 years of experience) with third reader in case of discrepancy; last two rounds: single reading by radiologist (≥6 years of experience) | Two experienced chest radiologists | Double independent readings by two experienced chest radiologists; consensus readings with local coordinator if in disagreement | Double independent reading by two radiologists (≥5 years of experience); consensus reading in case of disagreement | Two-step evaluation by a trained radiologist & senior radiologist | Reading by two radiologists, one of whom was equipped with the CAD software | Two local radiologists (6 and 25 years' experience); consensus nodule category assigned following central reading at hospital | Radiologists certified by the American Registry of Radiologic Technologists | Panel of experienced thoracic radiologists & respiratory physicians | Double reading at trial beginning which was changed to single reading (due to high concordance); double blind reviews of randomly selected LDCT scans, LDCT case report forms were also audited |
| **Definition of a positive scan at first screening round** | Any NCN ≥4 mm (based on the longest axial perpendicular diameter) ("suspicious for" lung cancer); | Indeterminate (for recall): solid nodules with volume of 50-500 $mm^3$ (4.6-9.8-mm diameter; not defined); pleural-based solid nodules with minimal | Category 3 (indeterminate; for recall): nodules between 5-15 mm (linear measurement of the maximal diameter in axial slices) without | Abnormalities suggestive of malignancy, such as noncalcified pulmonary nodules ≥10mm or smaller (measurement not defined) | At least one non-calcified solid or part-solid (solid component) ≥5 mm (mean diameter) or non-calcified non-solid nodule ≥10 mm (mean diameter) | Newly observed nodules ≥5 mm (in largest diameter) were classified as suspicious implying early recall | Indeterminate (recall): Nodules 60–250 $mm^3$ volume (about 5-8 mm, respectively [not defined])<br><br>Positive (for workup): NCNs > 250 | Category 2: solid with volume 15-49$mm^3$ or 3-4.9 mm (in maximal diameter when volumetric not possible), part solid with a solid | Any NCN ≥3 mm at T0 and ≥4mm T1 scan (measurement not defined), and other findings suspicious of lung cancer | Based on ECLS radiology schema: monitorable abnormalities (pulmonary nodules ≤ 8mm were classified as abnormal scan) | Nodule size ≥9 mm |

| Characteristic | NLST | NELSON | DLCST | DANTE | ITALUNG | LUSI | MILD | UKLS | LSS* | ECLS | LungSEARCH |
|---|---|---|---|---|---|---|---|---|---|---|---|
| | other abnormalities suggestive of lung cancer such as adenopathy or effusion<br><br>See below for recall vs workup | diameter of 5-10 mm; solid nodule with non-solid component or nonsolid with mean diameter of 8+ mm<br><br>Positive (for workup): Solid nodules with volume of >500 $mm^3$ (> 9.8 mm diameter [not defined]); pleural-based solid nodule >10 mm; solid component in partial solid nodule with volume of >500 $mm^3$ or nodule that grows fast i.e., >25% increase in volume (& VDT < 400 days) | benign characteristics<br><br>Category 4: nodules >15 mm & Category 5: all growing nodules, i.e., >25% increase in volume (for workup). | but showing spiculated margins, or non-nodular lesions such as a hilar mass, focal ground-glass opacities, major atelectasis, endobronchial lesions, mediastinal adenopathy, pleural effusion or pleural masses | | | $mm^3$, or part-solid nodules with new or growing solid component; “non-nodular findings” such as non-calcified hilar or mediastinal lymphadenomegaly, atelectasis, consolidation and pleural findings | component <15$mm^3$ or <3mm, nonsolid 3-4.9 mm; Category 3: solid with a volume of 50-500$mm^3$ or 5-9·9 mm, non-solid or part-solid, with the ground-glass component of >5mm, part solid with the solid component of 15-500 $mm^3$ or 5-9·9 mm; Category 4: solid with volume >500$mm^3$ or ≥100mm, part solid with solid component ≥10 mm or volume >500$mm^3$ | | | |
| **Definition of a positive scan at subsequent rounds** | At 3rd round, abnormalities that were stable across the 3 rounds | At round 1: VDT <400 days; new solid component in previously existing non-solid nodule | Growth was defined as an increase in volume of at least 25% | NR | New solid, part-solid or non-solid nodule or growth of solid or non-solid component | Newly observed nodules (same as above) or preexisting nodules with VDT within | Indeterminate nodules with 25% volume growth at the 3-month LDCT | NA | Any NCN ≥4mm, and other findings suspicious of lung cancer | Same as above | Growth of >25% |

| Characteristic | NLST | NELSON | DLCST | DANTE | ITALUNG | LUSI | MILD | UKLS | LSS* | ECLS | LungSEARCH |
|---|---|---|---|---|---|---|---|---|---|---|---|
| | could be classified as minor abnormalities rather than positive results | (GROWCAT C[†])<br>At rounds 2-3: GROWCAT C[†]; new and NODCAT IV[†]<br>At round 4: GROWCAT C[†]; NODCAT IV[†] | | | of a preexisting nodule | 400-600 days or VDT <400 days | | | | | |
| **Management protocol of a positive scan** | (Guideline but not mandated) Nodules 4-10 mm or enlarging nodules <7 mm: repeat LDCT at 3, 6, (3 to 6), 12, or 24 months, depending on size & level of lung cancer suspicion<br>Nodules >10 mm or enlarging nodules ≥7 mm: encouraged to | Indeterminant: recall<br>Positive: Referral to pulmonologist for diagnostic workup followed by treatment according to (inter)national guidelines if lung cancer diagnosed or repeat scan if no lung cancer<br>Growth reviewed at all incident scans; subjects with no growth or new nodule were negative | Category 3: rescanned after 3 months followed by biopsy or repeat scan 9 months later depending on the expansion of the opacity; category 4 or 5: referred to chest physician for diagnostic investigation<br>Opacity growth reviewed at each subsequent screen | Lesion ≥6mm but ≤10mm or >10mm but ≤20 mm: oral antibiotics and repeat HRCT after 6-8 weeks followed by PET scan or evaluation on a case-by-case basis; lesion >20mm: discretional oral antibiotics and HRCT or standard contrast-enhanced CT and PET scan<br>No differences reported between rounds | Based on the International-ELCAP Study: solid component 5-7 mm or non-solid nodule ≥10 mm: follow-up 3 months; ≥10mm suspected Inflammatory disease: follow-up 1 month + antibiotic; solid component 8-10 mm: FDG-PET; solid component ≥10 mm: Immediate diagnostic evaluation<br>Growth reviewed at all incident | Depending upon size of the largest observed nodule: re-invitation with repeat scan after 6 months in case of largest nodule 5-7 mm, after 3 months in case of largest nodule 8-10 mm, and immediate recall with nodules >10 mm<br>Growth reviewed at all incident scans; subjects with no growth or new nodule were negative | Nodules with volume of 60-250mm$^3$: repeat CT after 3 months; > 250mm$^3$: referred for additional workup<br>No differences reported between rounds | Category 2: repeat CT scan at 12 months; Category 3: repeat CT scan at 3 months & (if required) at 12 months; Category 4: immediate referral to multidisciplinary team for work-up & clinical management<br>Growth reviewed at all incident scans; subjects with no growth or new nodule were negative | Referred to their personal HCP for diagnostic follow-up; did not specify a diagnostic algorithm<br>Slight size difference between rounds | Based on the Fleischner Society guidelines; follow-up scan or referred for NHS clinical care as appropriate | Benign nodules: CT at 4, 8, 12 & 24 months; malignant nodules: CT with contrast |

| Characteristic | NLST | NELSON | DLCST | DANTE | ITALUNG | LUSI | MILD | UKLS | LSS* | ECLS | LungSEARCH |
|---|---|---|---|---|---|---|---|---|---|---|---|
| | undergo workup<br><br>Participants negative at 2nd incident scan (3rd screen) if nodules were stable | | | | scans; subjects with no growth or new nodule were negative | | | | | | |
| **Diagnostic workup** | Chest CT with nodule densitometry; FDG-PET or Scintigraphy; biopsy | Baseline NODCAT IV[f], baseline or incidence GROWCATC[f]: biopsy followed by surgical resection, if needed | The first step was normally HRCT; individual diagnostic plan involving a variety of invasive procedures (e.g., bronchoscopy, biopsy) | Varied case-by-case involving bronchoscopy, percutaneous fine-needle or core biopsy, VATS, or PET-scan | FDG-PET, & fine needle aspiration biopsy, surgical removal | At discretion of pulmonologist (CXR, full-dose CT, PET, bronchoscopy, VATS, biopsy, antibiotic treatment) | FDG-PET and/or contrast enhanced CT, biopsy and/or surgery | Further CT, needle biopsy, PET, EBUS, surgical referral | Clinical evaluation, further CT, CXR, pulmonary function test, bronchoscopy, biopsy or resection | CXR, contrast CT scan & bronchoscopy or biopsy | CT with contrast, PET/CT or other investigations for cancer according to local practice |
| **Planned follow-up duration for primary endpoint**<br><br>- Outcome assessment | **Median 6.5 years (through December 31, 2009)**<br>- Annual or semiannual questionnaires and linkage with NDI; lung-cancer mortality evaluated by death | **Minimum of 10 years since randomization**<br>- Blinded clinical expert committee; cause of death established from medical files in first 266 cases, after that (due to high concordance) national | **10 years for mortality; 5 years for incidence**<br>- Cause of death established by blinded local review board & classified based on ICD-10 codes; review of death certificates & national registries data | **5 years from randomization**<br>- Data from local health registries; deaths due to treatment complications were considered as deaths from lung cancer | **≥8 years of average follow up since randomization**<br>- Tuscany Region Registry of causes of death & cancer registry | **Up to 5 years after the last screening round**<br>- Combination of annual follow-up questionnaires (self-reports), & record linkages to cancer & mortality registries; information cross-checked with medical records | **10 years**<br>- Hospital/physician records & periodic inquiry to national registries; vital status determined blindly | **2 years after randomization**<br>- Cancer & death registry data provided by NHS Digital & the National Cancer Registration & Analysis Service who were not aware of participants | **1-year after the final screening**<br>- Abstraction of medical records & study update form to identify any lung cancers diagnosed; a certified tumor registrar confirmed all reported lung cancers | **2 years after randomization**<br>- Linkage with health services data repository & cancer registries | **Median 5 years**<br>- The primary end-point was lung cancers diagnosed at stage I/II or limited disease; linkage with cancer registries & research nurses periodically checked patient records for cancer diagnoses |

| Characteristic | NLST | NELSON | DLCST | DANTE | ITALUNG | LUSI | MILD | UKLS | LSS* | ECLS | LungSEARCH |
|---|---|---|---|---|---|---|---|---|---|---|---|
| | review panel in blinded manner | death registries used | | | | | | ' allocated trial arm | | | |
| **Longest follow-up duration, lung-cancer mortality**<br>- Outcome assessment<br>- Attrition, % | **Median 12.3 years**<br>- Death panel classification for previous deaths & NDI linkage to determine underlying cause of death for subsequent deaths<br>- NR | **11 years**<br>- Unblinded clinical expert committee, through linkage with national registries<br>- <1.0 at 10 years | **Median 11.2 years**<br>- Same as above<br>- 3.0 | **Median 8.35 years**<br>- Death certificates from health registries cross-checked with hospital/physician reports in 78% (of deaths); if several competing causes of death, cases reviewed by a blinded panel | **Median 11.3 years**<br>- Linkage with national registries; causes of deaths reviewed by blinded panel (two experts, & a third expert in case of disagreement)<br>- 20.2% | **Median 9.96 years post randomization**<br>- Linkage with municipal population registers; cross-checked with physician reports; lung-cancer mortality established by blinded endpoint committee<br>- 0.3 | **10 years (>90% with 9 years follow-up)**<br>- Hospital/physician records & periodic inquiry to national registries; vital status determined blindly<br>- 9.0 | **Median 9.9 years**<br>- Death during follow-up period where lung cancer was listed as the underlying cause of death in UK civil registrations data provided by NHS Digital<br>- 10.3 | **Median 5.2 years**<br>- Linkage with NDI; underlying cause of death variable used to classify deaths into lung-cancer- or non-lung cancer-related; no endpoint verification (death review)<br>- NR | **5 years**<br>- Same as above<br>- 10.1 | **8 years since randomization**<br>- Identified through patient records or national cancer registry<br>- 48.6 |
| **Longest follow-up duration, all-cause mortality**<br>- Outcome assessment<br>- Attrition, % | **Median 12.3 years**<br>- Probabilistic linkage with NDI<br>- 2.2 | **Same as above** | **Same as above** | **Same as above** | **Same as above** | **Same as above**<br>- Linkage with municipal population registers; cross-checked with physician reports<br>- 0.3 | **Same as above** | **Median 9.9 years**<br>- NHS digital for mortality from all causes, all cancers, & non-lung cancer causes<br>- NR | **Same as above** | **Same as above** | **Same as above** |
| **Longest follow-up duration, incidence**<br>- Outcome assessment<br>- Attrition, % | Median 11.3 years<br>- Probabilistic linkage with home | 11 years<br>- Medical files & linkage with national registries | Median 9.8 person-years<br>- Linkage with national cancer registries | Same as above | Median 11.3 years<br>- Linkage to the Tuscan Cancer Registry; if multiple | Median 9.77 years<br>- Combination of active follow-up & record linkage | Same as above (median 6.2 years of screening and >90% with 9 years total follow- | Median 9.1 years<br>- Cancer & death registry data provided | 1-year after the final screening<br>- Diagnostic evaluation of all positive | Same as above | 8 years since randomization<br>- Linkage with cancer registries & periodic checking of |

| Characteristic | NLST | NELSON | DLCST | DANTE | ITALUNG | LUSI | MILD | UKLS | LSS* | ECLS | LungSEARCH |
|---|---|---|---|---|---|---|---|---|---|---|---|
| | state cancer registries<br>- 12.4 | - NR | - 0.9 | | lung cancer diagnoses in same subject, only the first is considered<br>- 20.2% | with national registries | up [from randomization]) | by NHS Digital & the National Cancer Registration & Analysis Service (blind to trial arm)<br>- 24.3 | screens tracked by collection & abstraction of medical records<br>- 2.0 | | patient records<br>- 49.0 |
| **Screening outside trial** | CG: 4.3% during active phase (in LSS cohort) | NR ("35 lung cancers were classified as interval cancer, some diagnosed by screening received outside the trial") | IG: 16.5% during follow-up phase;<br>CG: 7.4% during active phase, 12.9% during follow-up phase | IG: 24.3% during active phase;<br>CG: 23.3% during active phase | NR | IG: 0.6% during active phase, 6.5% during follow-up phase;<br>CG: 4.8% during active phase, 8.2% during follow-up phase | IG: NR<br><br>CG: 1.2% overall | NR | IG: 18.0% overall;<br>CG: 3.5% overall<br>*Based on assessment in random sample of 120 LDCT & 240 CXR | NR | NR |
| **Studies used for critical outcomes** [Subgroups] | **All-cause mortality:** (1)[(2)]<br>**Lung cancer mortality:** (1)[(1-8)]<br>**Overdiagnosis:** NA | **All-cause mortality:** (9)[(9)]<br>**Lung cancer mortality:** (9)[(9)]<br>**Overdiagnosis:** (9) | **All-cause mortality:** (10)<br>**Lung cancer mortality:** (10)[(10)]<br>**Overdiagnosis:** (10) | **All-cause mortality:** (11)[(12); DANTE& MILD trials pooled]<br>**Lung cancer mortality:** (11)[(12); DANTE& MILD trials pooled]<br>**Overdiagnosis:** (11) | **All-cause mortality:** (13)<br>**Lung cancer mortality:** (14)<br>**Overdiagnosis:** (14) | **All-cause mortality:** (15)[(16)]<br>**Lung cancer mortality:** (15)[(16)]<br>**Overdiagnosis:** (15)[(15)] | **All-cause mortality:** (17)<br>**Lung cancer mortality:** (17)<br>**Overdiagnosis:** (17) | **All-cause mortality:** (18)[(18)]<br>**Lung cancer mortality:** (18)[(19)]<br>**Overdiagnosis:** (18)[(18); (19) - alternative overdiagnosis definitions] | **All-cause mortality:** (20)<br>**Lung cancer mortality:** (20)<br>**Overdiagnosis:** NA | **All-cause mortality:** (21)<br>**Lung cancer mortality:** (21)<br>**Overdiagnosis:** NA | **All-cause mortality:** (22)<br>**Lung cancer mortality:** (22)<br>**Overdiagnosis:** NA |
| **Studies used for important outcomes** (Subgroups) | **HRQoL:** NR<br>**Major complications from invasive testing:** | **HRQoL:** (29)<br>**Major complications from invasive testing:** (9) | **HRQoL:** NR<br>**Major complications from invasive testing:** NR | **HRQoL:** NR<br>**Major complications from invasive testing:** (11) | **HRQoL:** NR<br>**Major complications from invasive testing:** (38) | **HRQoL:** NR<br>**Major complications from invasive testing:** NR | **HRQoL:** NR<br>**Major complications from invasive testing:** NR | **HRQoL:** NR<br>**Major complications from invasive testing:** NR | **HRQoL:** NR<br>**Major complications from invasive testing:** (46, 47) | **HRQoL:** NR<br><br>**Important outcomes:** NA | **HRQoL:** NR<br><br>**Important outcomes:** NA |

| Characteristic | NLST | NELSON | DLCST | DANTE | ITALUNG | LUSI | MILD | UKLS | LSS* | ECLS | LungSEARCH |
|---|---|---|---|---|---|---|---|---|---|---|---|
| | (23-25)[(24, 25)] **Deaths from invasive testing:** (23) **False positives:** (23)[(3, 25, 26)] **Psychosocial consequences of screening:** (27) **Incidental findings:** (28)[(25)] | **Deaths from invasive testing:** NR **False positives:** (9, 30) **Psychosocial consequences of screening:** (29, 31) **Incidental findings:** NR | **Deaths from invasive testing:** NR **False positives:** (32, 33) **Psychosocial consequences of screening:** (34, 35) **Incidental findings:** NR | **Deaths from invasive testing:** (11) **False positives:** (11, 36) **Psychosocial consequences of screening:** NR **Incidental findings:** (11, 37) | **Deaths from invasive testing:** (39) **False positives:** (38, 40) **Psychosocial consequences of screening:** NR **Incidental findings:** (13, 38) | **Deaths from invasive testing:** NR **False positives:** (16) **Psychosocial consequences of screening:** NR **Incidental findings:** NR | **Deaths from invasive testing:** NR **False positives:** (41, 42) **Psychosocial consequences of screening:** NR **Incidental findings:** NR | **Deaths from invasive testing:** NR **False positives:** (19, 43) **Psychosocial consequences of screening:** (44) **Incidental findings:** (45) | **Deaths from invasive testing:** (46) **False positives:** (46-48) **Psychosocial consequences of screening:** NR **Incidental findings:** NR | | |

AFB: Autofluorescence bronchoscopy; CAD: Computer-aided detection; CG: Comparator group; COPD: Chronic obstructive pulmonary disease; CXR: Chest X-ray; DANTE: Detection and Screening of Early Lung Cancer by Novel Imaging Technology and Molecular Essays; DLCST: Danish Lung Cancer Screening Trial; EBUS: Endobronchial ultrasound; ECLS: Early Detection of Cancer of the Lung Scotland; FEV: Forced expiratory volume; FVC: Forced vital capacity; GROWCAT: growth category; HCP: healthcare practitioner; HRCT: High-resolution CT scan; HRQoL: health-related quality of life; IG: Intervention group; ITALUNG: Italian Lung Cancer Screening Trial; LDCT: Low-dose computed tomography; LLP: Liverpool lung project; LSS: Lung Screening Study; LUSI: Lung Cancer Screening Intervention trial; MILD: Multicentric Italian Lung Detection; NA: Not applicable; NCN: Non-calcified nodule; NDI: National death registry; NELSON: Dutch–Belgian lung-cancer screening trial; NLST: National Lung Screening; Trial; NODCAT: Nodule category; NR: Not reported; PET: Positron Emission Tomography; PY: Pack-years; UKLS: UK Lung Cancer Screening; VATS: Video assisted thoracoscopic surgery; VDT: Volume doubling time

*Pilot trial for NLST

**Annual screening n=1,190; biennial screening n=1,186

[ƚ] NELSON trial nodule categories: NODCAT IV: solid nodules with a volume of >500 mm³, pleural-based solid nodule with a minimal diameter of >10 mm, solid component in a partial solid nodule with a volume of >500 mm³; GROWCAT C: volume doubling time <400 days; new solid component in previously existing non-solid nodule

[‡] Number screened at each round by the number randomized/enrolled at baseline, multiplied by 100.

Table S2.2. Characteristics table, observational studies and trials without benefit data

| **Author, Year**<br>**Country (setting)** | **Inclusion criteria**<br>**Eligible for CT screening, *n***<br>**Enrolled, *n***<br>**Analyzed, *n***<br>**Age, *mean ± SD/median ± IQR (yr)***<br>**Gender, % male**<br>**Smoking status, current vs former smoker vs never smoker**<br>**Pack-years smoking history** | **Screening Strategy:**<br>**Type of CT scan**<br>**Number of rounds**<br>**Nodule management for a positive CT scan**<br>**Other relevant details of screening process** | **Outcomes** |
|---|---|---|---|
| Andersson, 2021<br><br>Sweden (Five sites from Gothenburg, Linköping, Malmö, Stockholm and Umeå) | Inclusion criteria:<br>- 50-64 years old at enrollment,<br>- Ability to speak Swedish<br><br>Eligible, n: 2,842<br>Enrolled, n: 1,604<br>Analyzed, n: 1,604<br><br>Age, Mean (SD): 61.0 (4.56)<br>Gender, % male: 52.6%<br>Current smoker & Former smoker, %: 49.7%<br>Never smoker, %: 50.3%<br>Pack-years smoking history, median (IQR): NR | LDCT<br>One round of LDCT (Referred to pulmonary specialist for diagnostic work-up: 10.1%;<br>One follow-up LDCT: 61.0%)<br><br>Nodule management: Solid nodules ≥300mm3, or high risk for lung cancer; Solid nodules 100-299 mm3 and ≥ 10 pack-years; Non-solid or part-solid nodules, ≥ 5 mm | Psychosocial consequences of screening |
| Bartlett, 2020<br><br>England (17 general practices in West London, England [hospital site or mobile CT scanner]) | Inclusion criteria:<br>- Smokers and ex-smokers aged 60-75 from general practices in West London<br>- LLPv2 5-year lung cancer risk threshold ≥2.0%<br>- PLCOM2012 6-year risk threshold ≥1.51%<br><br>Eligible, n: NR<br>*8366 invitation sent to participate (unclear how many met criteria for LDCT)<br>Enrolled, n: 1,159 (eligible for screening)<br>Analyzed, n: 1,145<br><br>Age, Range: 60-75 years<br>Gender, % male: NR<br>Current smoker, %: NR (included smokers and ex-smokers)<br>Former smoker, %: NR<br>Never smoker, %: NR | LDCT<br>One round of LDCT<br>BTS nodule management guideline (updated for screening). For solid nodules, positive findings included:<br>- Nodule >=300mm3 (>=8mm) + Brock >10% PET CT required<br>- Nodule >=300mm3 (>=8mm) + Brock <10% or nodule >=125mm3-<300mm3 (>=6mm - <8mm) received 3-month follow-up CT<br>- Nodule >=80mm3-<125mm3 (>=5mm - <6mm) received 6-month follow-up<br>- At 3- or 6-month follow-up if volume growth >=25% VDT <=600 days or unequivocal growth and nodule >200mm3 then PET-CT required, or if <=200mm3 CT at 3 months again<br>For sub-solid nodules, positive findings included:<br>- Nodular consolidation >=8mm or suspicious endobronchial lesion, and suspicious features PET CT required (if more likely to be inflammatory appears CT at 3 months and if growing/altered morphology or solid component >=8mm/pGGN>30mm then PET CT required)<br>If any part solid nodule or pGGN >=5mm with altered morphology or pGGN >30mm then CT scan at 3 months (follow-up based on resolved or shrinking nodule or back to annual screening if unchanged) | False positives |

| **Author, Year**<br>**Country (setting)** | **Inclusion criteria**<br>**Eligible for CT screening, *n***<br>**Enrolled, *n***<br>**Analyzed, *n***<br>**Age, *mean ± SD/median ± IQR (yr)***<br>**Gender, % male**<br>**Smoking status, current vs former smoker vs never smoker**<br>**Pack-years smoking history** | **Screening Strategy:**<br>**Type of CT scan**<br>**Number of rounds**<br>**Nodule management for a positive CT scan**<br>**Other relevant details of screening process** | **Outcomes** |
|---|---|---|---|
| | Pack-years smoking history, median (IQR): NR | | |
| Bartlett, 2021 (associated to Bartlett 2020)<br><br>England (17 general practices in West London, England [hospital site or mobile CT scanner]) | Inclusion criteria (from Bartlett, 2020):<br>- Smokers and ex-smokers aged 60-75 from general practices in West London<br>- LLPv2 5-year lung cancer risk threshold ≥2.0%<br>- PLCOM2012 6-year risk threshold ≥1.51%<br><br>Eligible, n: NR<br>*8366 invitation sent to participate (unclear how many met criteria for LDCT)<br>Enrolled, n: 1,159 (eligible for screening)<br>Analyzed, n: 1,145<br><br>Age, Range: 60-75 years<br>Gender, % male: NR<br>Current smoker, %: NR (included smokers and ex-smokers)<br>Former smoker, %: NR<br>Never smoker, %: NR<br>Pack-years smoking history, median (IQR): NR | LDCT<br>One round of LDCT<br>BTS nodule management guideline (updated for screening). For solid nodules, positive findings included:<br>- Nodule >=300mm3 (>=8mm) + Brock >10% PET CT required<br>- Nodule >=300mm3 (>=8mm) + Brock <10% or nodule >=125mm3-<300mm3 (>=6mm - <8mm) received 3 month follow-up CT<br>- Nodule >=80mm3-<125mm3 (>=5mm - <6mm) received 6 month follow-up<br>- At 3 or 6 month follow-up if volume grouwth >=25% VDT <=600 days or unequivocal growth and nodule >200mm3 then PET-CT required, or if <=200mm3 CT at 3 months again<br>For sub-solid nodules, positive findings included:<br>- Nodular consolidation >=8mm or suspicious endobronchial lesion, and suspicious features PET CT required (if more likely to be inflammatory appears CT at 3 months and if growing/altered morphology or solid component >=8mm/pGGN>30mm then PET CT required)<br>If any part solid nodule or pGGN >=5mm with altered morphology or pGGN >30mm then CT scan at 3 months (follow-up based on resolved or shrinking nodule or back to annual screening if unchanged) | Clinically significant incidental findings |
| DEPISCAN trial (Blanchon, 2007)<br><br>France (Practices of participating general and occupational physicians in France) | Inclusion criteria:<br>- Patients aged 50-75 years<br>- Asymptomatic<br>- Current or former smokers (having quit <15 years from enrollment)<br>- ≥15 cigarettes per day for at least 20 years<br><br>Eligible, n: 830<br>Enrolled, n (LDCT arm): 385<br>Analyzed, n (LDCT arm): 330<br><br>Age, Median (range) (LDCT arm): 56.0 (47.0-75.0) | LDCT<br>One round of LDCT<br>Nodule management criteria: Nodules >5 to <10 mm had repeat LDCT 3 months later, and if no change observed, LDCT scans were performed again at 6, 12, and 24 months from baseline. Nodules ≥10 mm had a CT with injection, a PET scan and/or histological diagnosis discussed in multidisciplinary clinic. | 1) False positives<br>2) Incidental findings |

| Author, Year<br>Country (setting) | Inclusion criteria<br>Eligible for CT screening, *n*<br>Enrolled, *n*<br>Analyzed, *n*<br>Age, *mean ± SD/median ± IQR (yr)*<br>Gender, % male<br>Smoking status, current vs former smoker vs never smoker<br>Pack-years smoking history | Screening Strategy:<br>Type of CT scan<br>Number of rounds<br>Nodule management for a positive CT scan<br>Other relevant details<br>of screening process | Outcomes |
|---|---|---|---|
| | Gender, % male (LDCT arm): 71.0%<br>Current smoker, %: 65%<br>Former smoker, %: 35%<br>Never smoker, %: NR<br>Pack per day, median (range):<br>- Current smokers (LDCT arm): 1 (0.25-3)<br>- Former smokers (LDCT arm): 1.5 (0.5-4) | | |
| Byrne, 2008<br><br>USA (Mailed survey; local setting) | Inclusion criteria:<br>- Patients aged 50 to 79 year<br>- No personal history of lung cancer<br>- Smoking history of at least 25 year<br>- No self-reported screening lung CT within 1 year<br>- Weight of less than 400 lb<br><br>Eligible, n: 408<br>Enrolled, n: 400<br>Analyzed, n: 393<br><br>Age, Mean (SD): 60.2 (6.8)<br>Gender, % male: 50.7%<br>Current smoker, %: 59.0%<br>Former smoker, %: NR<br>Never smoker, %: NR<br>Years smoked, mean (SD): 40.8 (7.4) | CT scan<br>Baseline spiral CT screening 1-4 weeks after enrollment and at 1-year follow up<br>Nodule management criteria:<br>Initial CT: Low suspicion: non-calcified nodules of 0.5-0.9 cm average diameter with non-spiculated border. Moderate to high suspicion: 0.5–0.9 cm average diameter with spiculated border or ≥ 1.0 cm average diameter. The radiologist used subjective criteria for noncalcified nodules ≥ 1.0 cm average diameter.<br>Repeat CT: The radiologist used subjective criteria for interpretation. | Psychological consequences of screening |
| Callol, 2007<br><br>Spain (Autonomous community of Madrid) | Inclusion criteria:<br>- over 50 years of age<br>- Active smokers (or who had stopped smoking up to 6 months previously)<br>- Smoked more than 30 cigarettes daily for at least 15 years, or 20 cigarettes for 20 years, or more than 10 pack/years and in contact with asbestos at work<br>- Absence of neoplastic antecedents or tumor symptoms<br>- In good physical condition | LDCT<br>One baseline LDCT and second round of screening at 2-year interval<br>Nodule management criteria: 5-7mm and 8-10mm nodules 6 and 3 months follow up respectively and >10mm receive immediate intervention | 1) False positives<br>2) Incidental findings |

| Author, Year<br>Country (setting) | Inclusion criteria<br>Eligible for CT screening, *n*<br>Enrolled, *n*<br>Analyzed, *n*<br>Age, *mean ± SD/median ± IQR (yr)*<br>Gender, % male<br>Smoking status, current vs former smoker vs never smoker<br>Pack-years smoking history | Screening Strategy:<br>Type of CT scan<br>Number of rounds<br>Nodule management for a positive CT scan<br>Other relevant details<br>of screening process | Outcomes |
|---|---|---|---|
| | Eligible, n: 482<br>Enrolled, n: 466<br>Analyzed, n: Baseline: 466, First annual scan: 406<br><br>Age, Mean: 60.5<br>Gender, % male: 64.7%<br>Current smoker, %: NR<br>Former smoker, %: NR<br>Never smoker, %: NR<br>Pack-years smoking history, mean: Males 40, Females 30 | | |
| Cattaneo, 2018<br><br>USA (Anne Arundel Medical Center, Maryland) | Inclusion criteria: According to NLST criteria,<br>- Age 55-77 years<br>- At least 30 packs per year smoking<br>- Smoking cessation less than 15 years<br>- No cancer symptoms<br><br>Eligible, n: 1,241<br>Enrolled, n: 1,241<br>Analyzed, n: 1,241<br><br>Age, Median: 66 years<br>Gender, % male: 47.5%<br>Current smoker, %: 49.1%<br>Former smoker, %: 48.2%<br>Never smoker, %: NR<br>Pack-years smoking history, median: 40 | LDCT<br>One baseline LDCT scan and reported adherence to follow-up up to 1 year+90 days.<br>Nodule management criteria: Noncalcified nodules with any diameter of at least 4 mm or greater, any noncalcified mass, adenopathy, or effusion were classified as positive, which were suspicious for lung cancer. Management of scans was according to LungRADs criteria.<br>All active smokers were referred to their local smoking cessation program. | Major complications from invasive diagnostic testing |
| Crestanello, 2004<br><br>USA (Mayo medical clinic) | Inclusion criteria:<br>- Asymptomatic men and women 50 years or older<br>- Had a 20 pack-years of smoking history within previous 10 years<br><br>Eligible, n: NR<br>Enrolled, n: 1520 | LDCT<br>One baseline LDCT scan and three annual incidence scans<br>Nodule management criteria: Perform CT scan in 3 months for nodules 4-8mm, perform CT scan as soon as possible and consider a CT nodule enhancement protocol or positron emission tomography, perform biopsy for nodules >20 mm<br>Other relevant details, NR | Clinically significant incidental findings |

| Author, Year<br>Country (setting) | Inclusion criteria<br>Eligible for CT screening, *n*<br>Enrolled, *n*<br>Analyzed, *n*<br>Age, *mean ± SD/median ± IQR (yr)*<br>Gender, % male<br>Smoking status, current vs former smoker vs never smoker<br>Pack-years smoking history | Screening Strategy:<br>Type of CT scan<br>Number of rounds<br>Nodule management for a positive CT scan<br>Other relevant details<br>of screening process | Outcomes |
|---|---|---|---|
| | Analyzed, n: Baseline: 1520<br>First annual incidence: 1478/1520<br>Second annual incidence: 1441/1520<br>Third annual incidence: 1358/1520<br><br>Age, Mean (Range): 59 (50-85)<br>Gender, % male: 51.6%<br>Current smoker, %: 61%<br>Former smoker, %: 39%<br>Never smoker, %: NR<br>Pack-years smoking history, median (Range): 45 (20-230) | | |
| Crosbie, 2019<br><br>UK (Community based LHC) | Inclusion criteria:<br>- Ever smokers (current or former)<br>- Age 55-74 years<br>- Registered at participating general practitioner practices (n=14)<br>- Were invited to community-based LHC<br>- 6-year risk of lung cancer score ≥1.51% (using PLCOM2012 risk model)<br><br>Eligible, n: 1,429<br>Enrolled, n: 1,429<br>Analyzed, n: 1,384<br><br>Age, Mean (SD): 64.7 (5.4)<br>Gender, % male: 49.4%<br>Current smoker, %: 27.9%<br>Former smoker, %: 72.1%<br>Never smoker, %: NR<br>Pack-years smoking history, mean (SD): 51.6 (26.8) | LDCT<br>One baseline LDCT scan and recall in those with intermediate scans<br>BTS nodule management guidelines: A positive scan was defined as any solid nodule ≥8mm with a risk of malignancy ≥10% or any other finding concerning for malignancy requiring immediate assessment. | False positives |
| Darling, 2021<br><br>Canada (Four hospitals in Ontario with level 1 thoracic surgery centers) | Inclusion criteria:<br>- Age 55 to 74 years<br>- Current or former daily cigarette smoker for greater than or equal to 20 years | LDCT<br>One round of LDCT and some receiving recall scans during first year of the program.<br>LungRADS version 1.1: categories 3 or 4 were positive.<br>Current smoker participants were automatically enrolled to smoking cessation program. | Clinically significant incidental findings |

| Author, Year<br>Country (setting) | Inclusion criteria<br>Eligible for CT screening, *n*<br>Enrolled, *n*<br>Analyzed, *n*<br>Age, *mean ± SD/median ± IQR (yr)*<br>Gender, % male<br>Smoking status, current vs former smoker vs never smoker<br>Pack-years smoking history | Screening Strategy:<br>Type of CT scan<br>Number of rounds<br>Nodule management for a positive CT scan<br>Other relevant details<br>of screening process | Outcomes |
|---|---|---|---|
| | - PLCOm2012 risk greater than or equal to 2% in 6 years<br><br>Eligible, n: 2,151 (Originally recruited 4205 but not all eligible for screening)<br>Enrolled, n: 2,151<br>Analyzed, n: 1,624<br><br>Age, Mean (SD): NR<br>Gender, % male: NR<br>Current smoker, %: NR<br>Former smoker, %: NR<br>Never smoker, %: NR<br>Pack-years smoking history, median (IQR): NR | | |
| Erkmen, 2022<br><br>USA (Temple University Hospital [Urban, safety-net health system]) | Inclusion criteria: USPSTF 2013 criteria,<br>- Age 55 to 80 years<br>- Minimum 30 pack-years of smoking<br>- Currently smoking or quit within 15 years<br>- Signed informed consent<br><br>Eligible, n: 1,571<br>Enrolled, n: 1,569<br>Analyzed, n: 1,509<br><br>Age, Mean (SD): NR<br>Gender, % male: 53.0%<br>Current smoker, %: 62.8%<br>Former smoker, %: 37.2%<br>Never smoker, %: NR<br>Pack-years smoking history, median (IQR): 40 (17) | LDCT<br>One baseline LDCT scan<br>LungRADS version 1.0: categories 3 or 4 were positive.<br>Smoking cessation counseling for active smokers. | Major complications from invasive diagnostic testing |
| Fabbrini, 2018<br><br>USA (The Minneapolis Veterans Affairs Health Care System [MVAHCS]) | Inclusion criteria:<br>- Age 55 to 80 years<br>- Having no diagnosis codes in the EHR for hospice care or lung, esophageal, pancreatic, or liver cancer | LDCT<br>One round of LDCT scan<br>Nodule management criteria: Any lung nodule with a diameter of 2 mm or greater as requiring follow-up LDCT. LDCT scans were classified as: diagnostic evaluation needed for possible malignancy (any nodule with | Incidental findings |

| Author, Year<br>Country (setting) | Inclusion criteria<br>Eligible for CT screening, *n*<br>Enrolled, *n*<br>Analyzed, *n*<br>Age, *mean ± SD/median ± IQR (yr)*<br>Gender, % male<br>Smoking status, current vs former smoker vs never smoker<br>Pack-years smoking history | Screening Strategy:<br>Type of CT scan<br>Number of rounds<br>Nodule management for a positive CT scan<br>Other relevant details<br>of screening process | Outcomes |
|---|---|---|---|
| | - No chest CT in the previous year<br>- Not being previously coded in the EHR as not expected to live more than 6 months<br>- ≥30 pack-years and either currently smoking or quit <15 years ago<br><br>Eligible, n: 918<br>Enrolled, n: 178<br>Analyzed, n: 178<br><br>Age, Mean (SD): 64.2 (5.2)<br>Gender, % male: 96.1%<br>Current smoker, %: 48.9%<br>Former smoker, %: 51.1%<br>Never smoker, %: NR<br>Pack-years smoking history, Mean (SD): 54.3 pack-years (23.7) | suspicious features, known to be new, or growing based on prior imaging).<br>Patients were offered to consult with tobacco cessation team. | |
| Fisher-Dortmann, 2024<br><br>Canada (Alberta lung cancer screening project pilot within Alberta primary care networks) | Inclusion criteria:<br>- 50-74 years old<br>- currently smoke or had quit smoking with a six-year risk of lung cancer of ≥ 1.5% using the PLCOm2012 3-race model lung cancer risk calculator<br><br>Eligible, n: 2,686<br>Enrolled, n: 2,101<br>Analyzed: 2, 101<br><br>Age, Mean (SD): NR<br>Gender, % male: NR<br>Current smoker, %: NR<br>Former smoker, %: NR<br>Pack-years smoking history, Mean (SD): NR | LDCT<br>One round of LDCT scan<br>LungRADS version 2022: categories 3 and 4 are considered positive. | Incidental findings |
| Guichet, 2018 | Inclusion criteria: NCCN eligibility criteria,<br>- 55-74 years old | LDCT<br>One round of LDCT | Clinically significant incidental findings |

| **Author, Year**<br>**Country (setting)** | **Inclusion criteria**<br>**Eligible for CT screening, *n***<br>**Enrolled, *n***<br>**Analyzed, *n***<br>**Age, *mean ± SD/median ± IQR (yr)***<br>**Gender, % male**<br>**Smoking status, current vs former smoker vs never smoker**<br>**Pack-years smoking history** | **Screening Strategy:**<br>**Type of CT scan**<br>**Number of rounds**<br>**Nodule management for a positive CT scan**<br>**Other relevant details**<br>**of screening process** | **Outcomes** |
|---|---|---|---|
| USA (Community partner clinics in an underserved region of south Los Angeles County, California) | - Either current or former smokers who had quit smoking with past 15 years<br>- At least a 30-pack-year smoking history<br>- 50-80 years old with 20-pack-year smoking history were also included if they had one additional lung cancer risk factor other than second hand smoke exposure.<br><br>Eligible, n: 329<br>Enrolled, n: 275<br>Analyzed, n: 275<br><br>Age, Mean (range): 59.0 (50-78)<br>Gender, % male: 96.3%<br>Current smoker, %: 81.0%<br>Former smoker, %: NR<br>Never smoker, %: NR<br>Pack-years smoking history, median (range): 40 (20-195) | LungRADS version 1.0: categories 3 and 4 are considered positive. | |
| LSUT trial (secondary analysis: Hall, 2022)<br><br>UK (Two hospitals in London, UK) | Inclusion criteria:<br>- Individuals aged 60 to 75 years<br>- Recorded as 'current smokers' within the seven preceding years were sent an invitation letter from their usual general practice doctor inviting them to an LHC.<br>- ≥30 pack-years<br>- a former smoker had quit ≤15 years ago, or<br>- a lung cancer risk of ≥1.51% as determined by PLCOm2012 criteria, or<br>- ≥2.5% as per the Liverpool Lung Project model<br><br>Eligible, n: NR<br>Enrolled, n: 770 | LDCT<br>One baseline LDCT scan and 11.9% undergoing 3 month and 4.3% undergoing 12-month surveillance for indeterminate nodules<br>BTS guidelines for nodule management: Positive findings: use the Brock model (full, with spiculation) for initial risk assessment of pulmonary nodules (≥8 mm or ≥300 mm3) at presentation in people aged ≥50 who are smokers or former smokers. | Incidental findings |

| Author, Year<br>Country (setting) | Inclusion criteria<br>Eligible for CT screening, *n*<br>Enrolled, *n*<br>Analyzed, *n*<br>Age, *mean ± SD/median ± IQR (yr)*<br>Gender, % male<br>Smoking status, current vs former smoker vs never smoker<br>Pack-years smoking history | Screening Strategy:<br>Type of CT scan<br>Number of rounds<br>Nodule management for a positive CT scan<br>Other relevant details<br>of screening process | Outcomes |
|---|---|---|---|
| | Analyzed, n: 770<br><br>Age, Median (IQR):<br>- Positive CT scan: 67 (63 to 69)<br>- Negative CT scan: 66 (63 to 69)<br>- Other result on CT: 65 (62 to 68)<br>Gender, % male: 55.4%<br>Current smoker, %: 72.4%<br>Former smoker, %: 27.6%<br>Never smoker, %: NR<br>Pack-years smoking history, median (IQR):<br>- Positive CT scan: 41 (27 to 56)<br>- Negative CT scan: 38 (25 to 50)<br>- Other result on CT: 42 (34 to 80) | | |
| Henschke, 2001<br><br>USA (Cornell University Medical College and New York University Medical Center) | Inclusion criteria:<br>- Asymptomatic persons<br>- 60 years of age or older<br>- At least 10 pack-years of cigarette smoking<br>- No prior cancer<br>- Medically fit to undergo thoracic surgery<br><br>Eligible, n: NR<br>Enrolled, n: 1000<br>Analyzed, n: Baseline: 1000<br>First annual repeat scan: 841/1000<br>Second annual repeat scan in those with a first annual scan: 461/841<br><br>Age, Median: 67.0<br>Gender, % male: 54.0%<br>Current smoker, %: NR<br>Former smoker, %: NR<br>Never smoker, %: NR<br>Pack-years smoking history, median: 45 | LDCT<br>One baseline LDCT scan and 2 repeat CT scans at 6-18 months intervals.<br>I-ELCAP nodule management: For NCN 5 mm or less in size (average of length and width), follow-up by HRCT 3 months later, and given no growth, at 6, 12, and 24 months. If no growth was noted over 2 years, the nodule was considered to be benign.<br>B) For NCN 6-10 mm in size, assessment on a case-by-case basis of the possibility of obtaining a biopsy using either percutaneous transthoracic CT-guided fine-needle aspiration or video-assisted thoracoscopic biopsy procedures. | False positives |
| Hirsch, 2023 | Inclusion criteria:<br>-Cohort 1 (not CMS 2015 eligible): Study enrollment was offered to individuals 40– | LDCT<br>One round of LDCT<br>LungRADS version 1.0: categories 3 and 4 are considered positive. | Clinically significant incidental findings |

| Author, Year<br>Country (setting) | Inclusion criteria<br>Eligible for CT screening, *n*<br>Enrolled, *n*<br>Analyzed, *n*<br>Age, *mean ± SD/median ± IQR (yr)*<br>Gender, % male<br>Smoking status, current vs former smoker vs never smoker<br>Pack-years smoking history | Screening Strategy:<br>Type of CT scan<br>Number of rounds<br>Nodule management for a positive CT scan<br>Other relevant details<br>of screening process | Outcomes |
|---|---|---|---|
| USA (University of Colorado Hospital LCS clinic) | 82 years of age with a 6-year risk score of ≥ 1.5% if < 77 years old or > 4% if 78–82 years old (calculated by the PLCOm2012 model) who did not meet 2015 CMS screening criteria.<br>-Cohort 2: CMS eligible: 55-77 years of age, and 30 pack-years smoking history.<br><br>Eligible, n: NR<br>Enrolled, n: 642<br>Analyzed, n: 642<br><br>Non-CMS cohort:<br>Age, Mean (SD): 69.0 (9.0)<br>CMS cohort:<br>Age, Mean (SD): 64.3 (6.0)<br><br>Gender, % male: 56.5%<br><br>Current smoker, %: 51.2%<br>Non-CMS cohort: Median (IQR) pack-years: 33.4 (23.5-45.0)<br>CMS cohort: Median (IQR) pack-years: 41.0 (36.0-54.0) | | |
| Janssen, 2019<br><br>USA (University of Minnesota Health LCS program [community partner practices or university]) | Inclusion criteria:<br>- Individuals who participated in the lung cancer screening program.<br>- 86.9% met USPSTF criteria at first exam<br>95% met NCCN Group 2 lung cancer screening eligibility:<br>i. At least 50 years of age<br>ii. At least 20 pack-years smoking history<br>iii. Additional risk factor<br><br>Eligible, n: 1,715<br>Enrolled, n: 1,715<br>Analyzed, n: 1,715 | LDCT<br>One round of LDCT<br>23.2% undergoing a second scan or more<br>LungRADS version 1.0: categories 3 and 4 are considered positive. | Incidental findings |

| Author, Year<br>Country (setting) | Inclusion criteria<br>Eligible for CT screening, *n*<br>Enrolled, *n*<br>Analyzed, *n*<br>Age, *mean ± SD/median ± IQR (yr)*<br>Gender, % male<br>Smoking status, current vs former smoker vs never smoker<br>Pack-years smoking history | Screening Strategy:<br>Type of CT scan<br>Number of rounds<br>Nodule management for a positive CT scan<br>Other relevant details<br>of screening process | Outcomes |
|---|---|---|---|
| | Age, Median (range): 63.0 (36-81)<br>Gender, % male: 46.4%<br>Current smoker, %: 58.6%<br>Former smoker, %: 39.0%<br>Never smoker, %: NR<br>Pack-years smoking history, median (range): 40.0 (1-260) | | |
| Kapoor, 2020<br><br>USA (Single institution [accredited LCS program]) | Inclusion criteria: USPSTF criteria,<br>- Age between 55 and 77 years<br>- Smoking history 30 pack-years or greater<br>- Being a current smoker or having quit smoking in the past 15 years<br>- Having no symptoms or signs of lung cancer<br><br>Eligible, n: NR<br>Enrolled, n: 1,458<br>Analyzed, n:1,458<br><br>Age, Mean (SD): No actionable IF present (n n=1,320): 64.6 (5.3)<br>Actionable IF present (n=138): 65.7 (5.7)<br>Gender, % male: 55.1%<br>Current smoker, %: 57.8%<br>Former smoker, %: NR<br>Never smoker, %: NR<br>Pack-years smoking history, Mean (SD): No actionable IF present: 47.9 (19.7)<br>Actionable IF present: 48.6 (16.0) | LDCT<br>One round of LDCT<br>LungRADS criteria: Level 3 and 4 lung cancer results were reviewed by the program nurse practitioner, and follow-up care was coordinated in each case on the basis of guidelines. Additional imaging for level 3 and nodule clinic visits with pulmonologist review for level 4. | Incidental findings (+ clinically significant incidental findings) |
| Kim, 2020<br><br>South Korea (The Health Promotion Centre of Seoul National University Bundang Hospital [tertiary center in South Korea]) | Inclusion criteria:<br>- Adults aged ⩾18 years who voluntarily underwent LDCT screening for lung cancer, regardless of their smoking history.<br>- All were asymptomatic at time of first visit.<br><br>Eligible, n: 37,436 | LDCT<br>One round of unenhanced LDCT scan<br>Nodule management criteria: Any noncalcified nodule with a longest diameter of ⩾4 mm was considered as positive. Participants with positive nodules were further classified using LungRADS criteria. Images before 2014 were re-evaluated. | 1) False positives<br>2) Major complications from invasive diagnostic testing |

| Author, Year<br>Country (setting) | Inclusion criteria<br>Eligible for CT screening, *n*<br>Enrolled, *n*<br>Analyzed, *n*<br>Age, *mean ± SD/median ± IQR (yr)*<br>Gender, % male<br>Smoking status, current vs former smoker vs never smoker<br>Pack-years smoking history | Screening Strategy:<br>Type of CT scan<br>Number of rounds<br>Nodule management for a positive CT scan<br>Other relevant details<br>of screening process | Outcomes |
|---|---|---|---|
| | Enrolled, n: 37,436<br>Analyzed, n: 37,436<br><br>Age, Mean (SD): 49.5 (11.2)<br>Gender, % male: 63.6%<br>Ever-smoker %: 52.0%<br>Never-smoker, %: 48.0%<br>Pack-years smoking history, median (IQR): NR | | |
| Kinsinger, 2017<br><br>USA (8 academic Veteran's Health Administration [VHA] hospitals) | Inclusion criteria:<br>- Aged 55 to 80 years<br>- Without a diagnosis of esophageal, liver, or pancreatic or lung cancer<br>- Without a documented estimated life expectancy of less than 6 months<br>- Current or former smokers (less than 15 years ago),<br>- Cigarette smokers who had smoked a minimum of 30 pack-years<br><br>Eligible, n: 4,246<br>Enrolled, n: 2,452<br>Analyzed, n: 2,106<br><br>Age, Mean (SD): 64.9 (5.1)<br>Gender, % male: 61.9%<br>Current smoker, %: 6.5%<br>Former smoker, %: 57.2%<br>Never smoker, %: 36.4%<br>Pack-years smoking history, Mean (SD): 17.1 (25.0) | LDCT<br>One round of LDCT scan<br>Nodule management criteria: “Nodules needing to be tracked” was designated based on Fleischner Society guidelines. These were,<br>- Solid nodules 8 mm or smaller without suspicious features (eg, irregular or spiculated borders)<br>- Not known to be new or growing based on results of prior imaging (if available)<br>- Ground glass nodules larger than 5 mm, or mixed solid and ground glass nodules of any size<br>- Patients with solid nodules 8 mm or smaller with suspicious features or nodules known to be new or growing and those with nodules larger than 8 mm were referred for further evaluation | Clinically significant incidental findings |
| Kucharczyk, 2011<br><br>Canada (Department of Medical Imaging at the Princess Margaret Hospital in Toronto) | Inclusion criteria:<br>- At least 10 pack-years history of smoking<br>- Age between 50 to 80 years<br>- In good health | LDCT<br>One baseline LDCT and some interim and annual repeat scans (details, NR)<br>Nodule management criteria: Solid nodule 8 mm or larger; solid nodule 5 mm or larger) | Clinically significant incidental findings |

| **Author, Year**<br>**Country (setting)** | **Inclusion criteria**<br>**Eligible for CT screening, *n***<br>**Enrolled, *n***<br>**Analyzed, *n***<br>**Age, *mean ± SD/median ± IQR (yr)***<br>**Gender, % male**<br>**Smoking status, current vs former smoker vs never smoker**<br>**Pack-years smoking history** | **Screening Strategy:**<br>**Type of CT scan**<br>**Number of rounds**<br>**Nodule management for a positive CT scan**<br>**Other relevant details**<br>**of screening process** | **Outcomes** |
|---|---|---|---|
| | - No history of cancer except non-melanotic skin cancer<br><br>Eligible, n: NR<br>Enrolled, n: 4,073<br>Analyzed, n: 4,073<br><br>Age, Median (Range): 62 (46-80)<br>Gender, % male: 46.0%<br>Current smoker, %: NR<br>Former smoker, %: NR<br>Never smoker, %: NR<br>Pack-years smoking history, median (Range): 33 (10-189 pack-years) | | |
| LSUT trial (Kummer, 2020)<br><br>United Kingdom (Two hospitals in London) | Inclusion criteria:<br>- Individuals aged 60 to 75 years<br>- Current smokers<br>- ≥30 pack-years<br>- Former smoker had quit ≤15 years ago<br>Or,<br>- A lung cancer risk of ≥1.51% as determined by the Prostate, Lung, Colorectal and Ovarian study<br>Or,<br>- A lung cancer risk of ≥2.5% as per the Liverpool Lung Project model.<br><br>Eligible, n (Screening sample): 1,005<br>Enrolled, n (Screening sample): 1,005<br>Enrolled, n (Community sample): 400<br>Analyzed, n (Screening sample): 744-787 (Number varies responding to each questionnaire)<br>Analyzed, n (Community sample): 383<br><br>Age, Mean (SD) (Screening Sample): 65.75 (4.01)<br>Age, Mean (SD) (Community Sample): 66.24 (4.18) | LDCT<br>One round of LDCT<br>BTS guidelines: Pulmonary nodules ≥8 mm or ≥300 mm3 at presentation in participants aged ≥50 who are smokers or former smokers. | 1) Psychological consequences of screening<br>2) Clinically significant incidental findings |

| Author, Year<br>Country (setting) | Inclusion criteria<br>Eligible for CT screening, *n*<br>Enrolled, *n*<br>Analyzed, *n*<br>Age, *mean ± SD/median ± IQR (yr)*<br>Gender, % male<br>Smoking status, current vs former smoker vs never smoker<br>Pack-years smoking history | Screening Strategy:<br>Type of CT scan<br>Number of rounds<br>Nodule management for a positive CT scan<br>Other relevant details<br>of screening process | Outcomes |
|---|---|---|---|
| | Gender, % male: 54.0%<br>Current smoker, % (*n=1,164): 72.7%<br>Former smoker, % (*n=1,164): 27.3%<br>Never smoker, %: NR<br>Pack-years smoking history, median (IQR): NR | | |
| Laisaar, 2025<br><br>Estonia (92 family physician practices in Tartu County) | Inclusion criteria: Individuals aged 55–74 years who had ever smoked were considered potential study participants. Two LCS inclusion criteria [smoking status and a modified version of the lung cancer (LC) risk prediction model PLCOm2012noRace score] were used in parallel. Participants who met either one or both of the following LCS criteria were recruited:<br>•Currently or ever-smoking individual (quit <15 years ago) who had smoked at least 20 pack-years, and a PLCOm2012noRace risk score >1.5 %/6 years.<br><br>Eligible, n: 3,708<br>Enrolled, n: 3,444<br>Analyzed, n: 3,444<br><br>Age, range: 55-74<br>Gender, % male: NR<br>Current smoker, %: NR<br>Former smoker, %: NR<br>Never smoker, %: NR<br>Pack-years smoking history, median (IQR): NR | LDCT<br>One round of LDCT<br>LungRADS version 1.1: categories 3 and 4 are considered positive. | Incidental findings (+ clinically significant incidental findings) |
| Lam, 2015<br><br>USA (El Camino Hospital, Mountain View, California [large suburban community hospital]) | Inclusion criteria: Eligible participants were,<br>- At least 50 years old<br>- Had a history of cigarette smoking of at least 20 pack-years at the time of study enrollment | LDCT<br>One round LDCT scan (46.1% underwent a diagnostic follow-up scan)<br>Nodule management criteria: Any non-calcified nodule measuring at least 4 mm in any diameter was classified as ‘‘positive’’ (NLST criteria). CT scans with any other abnormalities suspicious for lung cancer, such as adenopathy or effusion, could be classified as positive as well. | 1) False positives<br>2) Incidental findings (+ clinically significant)<br>3) Major complications from invasive diagnostic testing |

| Author, Year<br>Country (setting) | Inclusion criteria<br>Eligible for CT screening, *n*<br>Enrolled, *n*<br>Analyzed, *n*<br>Age, *mean ± SD/median ± IQR (yr)*<br>Gender, % male<br>Smoking status, current vs former smoker vs never smoker<br>Pack-years smoking history | Screening Strategy:<br>Type of CT scan<br>Number of rounds<br>Nodule management for a positive CT scan<br>Other relevant details<br>of screening process | Outcomes |
|---|---|---|---|
| | - All former smokers were eligible regardless of when they had quit<br><br>Eligible, n: NR<br>Enrolled, n: 154<br>Analyzed, n: 154<br><br>Age, Mean (SD): 64.4 (8.0)<br>Gender, % male: 41.6%<br>Current smoker, %: 27.9%<br>Former smoker, %: 72.1%<br>Never smoker, %: NR<br>Pack-years smoking history, Mean (SD): 45.4 (25.0) | Recommendations for diagnostic follow-up of positive scans generally followed International Early Lung Cancer Action Program (I-ELCAP) guidelines.<br>All were also referred to an affiliated smoking cessation program. All received prognostic genomic-based testing but not the focus of this publication. | |
| Ledford, 2016<br><br>USA (Radiology department of community hospital) | Inclusion criteria: Lung cancer screening was ordered when,<br>- The patient was between 55 to 79 years old,<br>- Must had a documented smoking history (30 pack-years) with recent smoking behavior (smoked within 15 years).<br><br>Eligible, n: 72<br>Enrolled, n: 72<br>Analyzed, n: 72<br><br>Age, Mean (SD): 62.48 (5.93)<br>Gender, % male: 73.6%<br>Current smoker, %: NR<br>Former smoker, %: NR<br>Never smoker, %: NR<br>Pack-years smoking history, median (IQR): NR | LDCT<br>One round of LDCT scan<br>Fleischner Society guidelines: Nodules considered positive ranged from <4mm to >8mm, and some categorized as 0-no findings received additional imaging. | Incidental findings (+ clinically significant incidental findings) |
| Lee, 2021<br><br>South Korea (National health screening program or smoking cessation | Inclusion criteria:<br>- Smokers or former smokers (who quit smoking for less than 15 years)<br>- Aged 55-74 years | LDCT<br>One round of LDCT<br>LungRADS criteria: LungRADS criteria category 3 and 4 are considered positive.<br>Mandatory smoking cessation counseling was provided to current smokers by a physician. | 1) Major complications from invasive diagnostic testing<br>2) Deaths from invasive diagnostic testing |

| Author, Year<br>Country (setting) | Inclusion criteria<br>Eligible for CT screening, *n*<br>Enrolled, *n*<br>Analyzed, *n*<br>Age, *mean ± SD/median ± IQR (yr)*<br>Gender, % male<br>Smoking status, current vs former smoker vs never smoker<br>Pack-years smoking history | Screening Strategy:<br>Type of CT scan<br>Number of rounds<br>Nodule management for a positive CT scan<br>Other relevant details<br>of screening process | Outcomes |
|---|---|---|---|
| clinics in South Korea) | - At least 30-pack-years smoking history<br><br>Eligible, n: 6,754<br>Enrolled, n: 5,692<br>Analyzed, n: 5,692<br><br>Age, (Range): 55-74 years<br>Gender, % male: 97.9%<br>Current smoker, %: 53.6%<br>Former smoker, %:46.4%<br>Never smoker, %: NR<br>Pack-years smoking history, median (IQR): NR | | |
| Lutzow, 2022 (associated to Erkmen 2022)<br><br>USA (Temple University Hospital [Urban, safety-net health system]) | Inclusion criteria: USPSTF 2013 criteria,<br>- Age 55 and 80<br>- Minimum of 30 pack-years of smoking and currently smoking or quit within 15 years<br>- Those who signed informed consent<br><br>Eligible, n: NR<br>Enrolled, n: 995<br>Analyzed, n: 995<br><br>Age, Mean (SD): NR<br>Gender, % male: 51.0%<br>Current smoker, %: 62.6%<br>Former smoker, %: 37.4%<br>Never smoker, %: NR<br>Pack-years smoking history, median (IQR): 42 (18) | LDCT<br>One round of LDCT<br>LungRADS version 1.0: categories 3 or 4 are considered positive.<br>Patients discussed smoking cessation at follow-up visit to discuss screening results. | 1) Specific incidental findings<br>2) Deaths from invasive diagnostic testing |
| MacRedmond, 2006<br><br>Ireland (Clinical Research Centre, Beaumont Hospital) | Inclusion criteria:<br>- Aged 50 years or over with a history of at least 10-pack-years smoking<br>- Current smokers at the age of 45<br>- No prior history of cancer | LDCT<br>Annual LDCT for 2 years<br>Nodule management criteria: Non-calcified nodules (NCNs) of >=10 mm in diameter were referred for biopsy; noncalcified nodules < 10 mm in diameter were followed with LDCT scans at 6, 12 and 24 months, larger nodules or those showing interval growth were referred for fine needle aspiration biopsy. | 1) False positives<br>2) Incidental findings (+ clinically significant incidental findings) |

| Author, Year<br>Country (setting) | Inclusion criteria<br>Eligible for CT screening, *n*<br>Enrolled, *n*<br>Analyzed, *n*<br>Age, *mean ± SD/median ± IQR (yr)*<br>Gender, % male<br>Smoking status, current vs former smoker vs never smoker<br>Pack-years smoking history | Screening Strategy:<br>Type of CT scan<br>Number of rounds<br>Nodule management for a positive CT scan<br>Other relevant details<br>of screening process | Outcomes |
|---|---|---|---|
| | - Medically fit for thoracic surgery (no chronic medical condition, not oxygen dependent, and could hold breath for 20 seconds)<br><br>Eligible, n: NR<br>Enrolled, n: 449<br>Analyzed, n:<br>- Baseline: 449<br>- Annual repeat: 413<br><br>Age, Median (Range): 56.4 (50-74)<br>Gender, % male: 49.9%<br>Current smoker, %: 68.4%<br>Former smoker, %: NR<br>Never smoker, %: NR<br>Pack-years smoking history, mean (range): 53.4 (10-160) | Smoking cessation advice was provided to participants at each follow-up. | |
| Marshall, 2015<br><br>Australia (A single tertiary institute, The Prince Charles Hospital) | Inclusion criteria:<br>- Age 60-74 years and 364 days<br>- Current or previous cumulative cigarette smoking history of >30 pack years (packs per day multiplied by the number of years smoked)<br>- Former smokers must have quit smoking within the previous 15 years<br>- With or without asbestos exposure<br><br>Eligible, n: 512<br>Enrolled, n: 256<br>Analyzed, n: T0 (baseline) – 256<br>T1 – 239<br>T2 – 233<br><br>Age, Media: 64.5<br>Gender, % male: 66.8%<br>Current smoker, %: 47.3%<br>Former smoker, %: NR | LDCT<br>One baseline LDCT scan (T0) and two annual incidence scans (T1 & T2)<br>NLST criteria: Baseline scans were considered positive if one or more nodules ≥4 mm diameter were detected; incidence scans were considered positive if one or more new nodules of any size was detected, or a previously identified nodule demonstrated growth. Suspicious non-nodule findings also constituted a positive scan. | 1) False positives<br>2) Major complications from invasive diagnostic testing |

| Author, Year<br>Country (setting) | Inclusion criteria<br>Eligible for CT screening, *n*<br>Enrolled, *n*<br>Analyzed, *n*<br>Age, *mean ± SD/median ± IQR (yr)*<br>Gender, % male<br>Smoking status, current vs former smoker vs never smoker<br>Pack-years smoking history | Screening Strategy:<br>Type of CT scan<br>Number of rounds<br>Nodule management for a positive CT scan<br>Other relevant details<br>of screening process | Outcomes |
|---|---|---|---|
| | Never smoker, %: NR<br>Pack-years smoking history, median (Range): 55 (25-235) | | |
| Mendoza, 2020<br><br>USA (Department of Radiology, Massachusetts General Hospital) | Inclusion criteria:<br>- 55 to 80 years of age<br>- 30 pack-years or more of smoking history<br>- Current or former smokers who have quit smoking in the last 15 years<br><br>Eligible, n: 3,110<br>Enrolled, n: 3,110<br>Analyzed, n: 3,110<br><br>Age, Mean (SD): NR<br>Gender, % male: NR<br>Current smoker, %: NR<br>Former smoker, %: NR<br>Never smoker, %: NR<br>Pack-years smoking history, median (IQR): NR | LDCT<br>One baseline LDCT scan<br>LungRADS criteria: category 3 and 4 are considered positive. | Specific incidental findings |
| Menezes, 2010<br><br>Canada (Department of Medical Imaging at the University Health Network in Toronto) | Inclusion criteria:<br>- Age 50 years or older<br>- Smoking history of at least 10-pack-years<br>- No previous cancer<br>- General good health<br><br>Eligible, n: NR<br>Enrolled, n: 3,352<br>Analyzed, n: 3,352<br><br>Age, Mean (SD): 60.8 (6.7)<br>Gender, % male: 46.0%<br>Current smoker, %: NR<br>Former smoker, %: NR<br>Never smoker, %: NR<br>Pack-years smoking history, median (range): 30 (10-189) | LDCT<br>One LDCT scan at baseline and 5 annual screens at 1-year intervals<br>DLCST criteria: At least one indeterminate non-calcified nodule 5mm or larger in size, or non-solid nodule 8mm or larger in size was identified; annual repeat scans were regarded as positive if a pre-existing nodule was found to have grown or if a new nodule was detected. | False positive |

| **Author, Year**<br>**Country (setting)** | **Inclusion criteria**<br>**Eligible for CT screening, *n***<br>**Enrolled, *n***<br>**Analyzed, *n***<br>**Age, *mean ± SD/median ± IQR (yr)***<br>**Gender, % male**<br>**Smoking status, current vs former smoker vs never smoker**<br>**Pack-years smoking history** | **Screening Strategy:**<br>**Type of CT scan**<br>**Number of rounds**<br>**Nodule management for a positive CT scan**<br>**Other relevant details**<br>**of screening process** | **Outcomes** |
|---|---|---|---|
| Miller, 2016<br><br>USA (Five hospitals, three health perks, 140 outpatient medical offices, and 12 imaging centers within the WellStar Health System) | Inclusion criteria: From 2008-2010 International Early Lung Cancer Action Program (I-ELCAP) screening criteria. From 2011-2013 National Comprehensive Cancer Network (NCCN).<br><br>Eligible, n: 1,267<br>Enrolled, n: 1,267<br>Analyzed, n: 1,267<br><br>Age, Mean (SD): NR<br>Gender, % male: 53.0%<br>Current smoker, %: 32%<br>Former smoker, %: 51%<br>Never smoker, %: 17%<br>Pack-years smoking history, median (IQR): NR | LDCT<br>One baseline LDCT scan<br>Nodule management criteria: Screened positive nodules were defined as greater or equal to 6 mm in LDCT.<br>All the current smoker participants were referred to smoking cessation program. | False positives |
| Morgan, 2017<br><br>USA (Cleveland Clinic Lung Cancer Screening Program) | Inclusion criteria:<br>- Age between 55 to 77 years<br>- 30 pack-years or greater smoking history<br>- Being a current smoker or someone who quit smoking within the past 15 years<br>- Having no symptoms or signs of lung cancer<br>- Being healthy enough to tolerate curative intent treatment of early-stage lung cancer<br>- Not having had a CT scan of the chest within 12 months<br><br>Eligible, n: NR<br>Enrolled, n: 320<br>Analyzed, n: 320<br><br>Age, Mean (SD): 64.4 (5.5)<br>Gender, % male: 61.9%<br>Current smoker, %: 48.1% | LDCT<br>One baseline LDCT scan<br>LungRADS version 1.0: categories 3 or 4 are considered positive. | Incidental findings (+ clinically significant incidental findings) |

| **Author, Year Country (setting)** | **Inclusion criteria Eligible for CT screening, *n* Enrolled, *n* Analyzed, *n* Age, *mean ± SD/median ± IQR (yr)* Gender, % male Smoking status, current vs former smoker vs never smoker Pack-years smoking history** | **Screening Strategy: Type of CT scan Number of rounds Nodule management for a positive CT scan Other relevant details of screening process** | **Outcomes** |
|---|---|---|---|
| | Former smoker, %: 51.9%<br>Never smoker, %: NR<br>Pack-years smoking history, mean (SD): 52.1 (19.8) | | |
| Munoz-Largacha, 2018<br><br>USA (Boston University Medical Center) | Inclusion criteria:<br>- Age between 55 and 80 years<br>- Minimum of 30 pack-years of smoking and currently smoking or quit within 15 years<br>- Signed informed consent<br><br>Eligible, n: 554<br>Enrolled, n: 554<br>Analyzed, n: 554<br><br>Age, Mean (Range): 63 (47-85)<br>Gender, % male: 60.0%<br>Current smoker, %: 52%<br>Former smoker, %: 25%<br>Undetermined, %: 23%<br>Pack-years smoking history, median (IQR): NR | LDCT<br>One baseline LDCT for all participants and three-month follow-up LDCT or PET scans for patients with Lung-RADS 4 criteria.<br>LungRADs criteria: category 4 was considered positive. | Major complications from invasive diagnostic testing |
| Murray, 2016<br><br>Australia (Asbestos Review Programme in Western Australia [single site]) | Inclusion criteria:<br>- Former workers of the Wittenoom crocidolite mine and mill<br>- Ex-residents of the town of Wittenoom (the mine-service town, which was heavily contaminated with mine tailings)<br>- Any worker with greater than 3 months cumulative full-time equivalent occupational exposure to asbestos and/or with evidence of pleural plaques.<br><br>Eligible, n: 1,308<br>Enrolled, n: 906<br>Analyzed, n: 906<br><br>Age, Mean (SD): 68.8 (9.94) | LDCT<br>One baseline LDCT scan and three months repeat CT scan of participants with intermediate nodule<br>Nodule management criteria: Potentially significant nodules (termed 'indeterminate') were defined as solid nodules without benign-type calcification greater than 50mm3 and part-solid or non-solid nodules (for which accurate volumetric assessment is not technically possible) with a mean dimension of at least 5 mm. | 1) Incidental findings (+ clinically significant incidental findings)<br>2) Major complications from invasive diagnostic testing<br>3) False positives |

| **Author, Year**<br>**Country (setting)** | **Inclusion criteria**<br>**Eligible for CT screening, *n***<br>**Enrolled, *n***<br>**Analyzed, *n***<br>**Age, *mean ± SD/median ± IQR (yr)***<br>**Gender, % male**<br>**Smoking status, current vs former smoker vs never smoker**<br>**Pack-years smoking history** | **Screening Strategy:**<br>**Type of CT scan**<br>**Number of rounds**<br>**Nodule management for a positive CT scan**<br>**Other relevant details**<br>**of screening process** | **Outcomes** |
|---|---|---|---|
| | Gender, % male: 83.6%<br>Current smoker, %: 6.5%<br>Former smoker, %: 57.2%<br>Never smoker, %: 36.4%<br>Pack-years smoking history, mean (SD): 17.1 (25.0) | | |
| Nahorecki, 2015<br><br>Poland (A teaching Hospital in Wroclaw, Poland) | Inclusion criteria:<br>- Age 55-70 years<br>- Asymptomatic<br>- Who had not previously suffered from cancer<br>- Smoked at least 10 packs of cigarettes a year<br><br>Eligible, n: NR<br>Enrolled, n: 332<br>Analyzed, n: 332<br><br>Age, Mean (SD): NR<br>Gender, % male: 49.6%<br>Current smoker, %: NR<br>Former smoker, %: NR<br>Never smoker, %: NR<br>Pack-years smoking history, median (IQR): NR | LDCT<br>One baseline LDCT scan<br>Nodule management criteria: The result was considered positive if the identified nodule was non-calcified, solid, non-solid or partly solid and ≥5 mm in diameter, semi-positive if the identified nodule was non-calcified, solid, non-solid or partly solid, and <5 mm in diameter, negative result consisted of no nodules observed (modified I-ELCAP protocol).<br>Participants were referred to a clinic for nicotine-addicts based on the questionnaire. | False positive |
| Nunez, 2022<br><br>USA (Veterans Health Administration locations) | Inclusion criteria:<br>- Veterans 55 to 80 years of age<br>- > 30 years of smoking history<br>- Who underwent initial LCS in any VA facility from January 1, 2013, through December 31, 2019.<br><br>Eligible, n: 82,641<br>Enrolled, n: 82,641<br>Analyzed, n: 82,641<br><br>Age, Mean (SD): 65.2 (5.6)<br>Gender, % male: 95.4%<br>Current smoker, %: NR<br>Former smoker, %: NR | LDCT<br>One baseline LDCT scan<br>Lung-RADS criteria: category 3 or 4 were considered positive. | 1) Major complications from invasive diagnostic testing<br>2) Deaths from invasive diagnostic testing |

| Author, Year<br>Country (setting) | Inclusion criteria<br>Eligible for CT screening, *n*<br>Enrolled, *n*<br>Analyzed, *n*<br>Age, *mean ± SD/median ± IQR (yr)*<br>Gender, % male<br>Smoking status, current vs former smoker vs never smoker<br>Pack-years smoking history | Screening Strategy:<br>Type of CT scan<br>Number of rounds<br>Nodule management for a positive CT scan<br>Other relevant details<br>of screening process | Outcomes |
|---|---|---|---|
| | Never smoker, %: NR<br>Pack-years smoking history, median (IQR): NR<br>>30 years of smoking history: 100% | | |
| Ostrowski, 2019<br><br>Poland (Department of Thoracic Surgery, Medical University of Gdansk) | Inclusion criteria:<br>- Age 50–75 years with performance status 0<br>- Smoking history of >30 pack-years<br><br>Eligible, n: 5,534<br>Enrolled, n: 5,534<br>Analyzed, n: 5,534<br><br>Age, Median (range): 62 (50-79)<br>Gender, % male: 51.0%<br>Current smoker, %: 68.9%<br>Former smoker, %: 31.1%<br>Never smoker, %: NR<br>Pack-years smoking history, median (range): 44.0 (3-200) | LDCT<br>One baseline LDCT scan for all participants and 17.6% underwent more than 1 round of screening. Screening followed a modified UK Lung Screening protocol with 1-year follow-up including 2 screening rounds.<br>Nodule management criteria: All the patients with nodules >500mm3 or with a volume doubling time (VDT) <400 days calculated after the second round were referred for review by the tumor board meeting. After MTB review, all the patients with nodules suggestive of malignancy or with ambiguous appearance underwent a diagnostic work-up.<br>Participants were offered an appointment at a smoking cessation clinic. | 1) False positive<br>2) Incidental findings |
| Pastorino, 2003<br><br>Italy (several screening sites) | Inclusion criteria:<br>- Age 50 years or older<br>- Current or former smokers, with a minimum of 20 pack-years smoking history<br>- No history of malignant disease, and adequate performance status (assessed on the basis of the patient's eligibility to undergo thoracic surgery).<br>Eligible, n: NR<br>Enrolled, n: 1,035<br>Analyzed, n: 1,035<br><br>Age, Median (range): 58 (50-84)<br>Gender, % male: 71.0%<br>Current smoker, %: 86%<br>Former smoker, %: 14%<br>Pack-years smoking history, median: 40 | LDCT<br>One baseline LDCT scan and one follow-up scan at a 1-year interval.<br>Nodule management criteria: Patients with nodules >7mm with contrast enhancement CT but negative on PET, or if the nodule >7mm with no enhancement but positive on PET, they received biopsy. Spiral thin-section CT limited to the area of interest and 3D analysis was done within 1 month in every case of non-calcified lesion >5 mm, with assessment of contrast enhancement in nodular lesions that had density ≥0 Hounsfield Units (HU). Lesions with positive enhancement (>30 HU) or positive PET scan were candidates for biopsy, as well as non-calcified lesions of ≥20 mm, unless unequivocally benign at high-resolution CT. Further examinations for growth assessment were completed within 6 months of baseline CT.<br>At baseline collection of blood, urine, and sputum samples was also done. | False positives |

| **Author, Year**<br>**Country (setting)** | **Inclusion criteria**<br>**Eligible for CT screening, *n***<br>**Enrolled, *n***<br>**Analyzed, *n***<br>**Age, *mean ± SD/median ± IQR (yr)***<br>**Gender, % male**<br>**Smoking status, current vs former smoker vs never smoker**<br>**Pack-years smoking history** | **Screening Strategy:**<br>**Type of CT scan**<br>**Number of rounds**<br>**Nodule management for a positive CT scan**<br>**Other relevant details**<br>**of screening process** | **Outcomes** |
|---|---|---|---|
| | (average tobacco consumption: 26 cigarettes daily for 37 years) | | |
| Priola, 2013<br><br>Italy (The radiology department of San Luigi Gonzaga University Hospital) | Inclusion criteria:<br>- Either sex<br>- Age ≥55 years<br>- Current daily smokers or former (up to 10 years before) smokers of 20 pack-years<br>- Negative personal history of malignancy<br><br>Eligible, n: NR<br>Enrolled, n: 519<br>Analyzed, n: 519<br><br>Age, Mean (SD): 60.4 (4.4)<br>Gender, % male: 73.4%<br>Current smoker, %: 91.1%<br>Former smoker, %: 8.9%<br>Never smoker, %: NR<br>Pack-years smoking history, median (IQR): NR | LDCT<br>One baseline LDCT scan and four follow-up scans at 1-year intervals<br>Nodule management criteria: NR. | Incidental findings (+ clinically significant incidental findings) |
| Raghavan, 2020<br><br>USA (Mobile CT scanner at underserved community clinics that are part of Atrium Health in North and South Carolina [multisite safety-net organization, consisting of more than 40 hospitals]) | Inclusion criteria:<br>- age over 55 years (up to 64 years)<br>- Provided informed consent<br><br>Eligible, n: NR<br>Enrolled, n: 550<br>Analyzed, n: 550<br><br>Age, Mean (Range): 61.0 (55-64)<br>Gender, % male: 52.3%<br>Current smoker, %: NR<br>Former smoker, %: NR<br>Never smoker, %: NR<br>Pack-years smoking history, mean (range): 46.1 (30-220) | LDCT<br>One baseline LDCT scan<br>LungRADs criteria: Lung-RADS categories 3 to 4 constitute positive results.<br>Smoking cessation programs were offered to active smokers. | Specific incidental findings |
| Rampinelli, 2011 | Inclusion criteria:<br>- Age 50 years or older | LDCT<br>Baseline LDCT scan and four follow-up scans at 1-year intervals | Clinically significant incidental findings |

| **Author, Year**<br>**Country (setting)** | **Inclusion criteria**<br>**Eligible for CT screening, *n***<br>**Enrolled, *n***<br>**Analyzed, *n***<br>**Age, *mean ± SD/median ± IQR (yr)***<br>**Gender, % male**<br>**Smoking status, current vs former smoker vs never smoker**<br>**Pack-years smoking history** | **Screening Strategy:**<br>**Type of CT scan**<br>**Number of rounds**<br>**Nodule management for a positive CT scan**<br>**Other relevant details**<br>**of screening process** | **Outcomes** |
|---|---|---|---|
| Italy (Single screening center in Milan) | - Current or former heavy smokers (≥20 pack-years)<br>- Ex-smokers had stopped smoking not more than 10 years previously<br><br>Eligible, n: NR<br>Enrolled, n: 5,203 (from Rampinelli 2017)<br>Analyzed, n: Baseline - 5,201<br>Round 2 – 4,825<br>Round 3 – 4,586<br>Round 4 – 4,389<br>Round 5 – 4,127<br><br>Age, Mean (SD): NR<br>Gender, % male: NR<br>Current smoker, %: NR<br>Former smoker, %: NR<br>Never smoker, %: NR<br>Pack-years smoking history, median (IQR): NR | Nodule management criteria: Patients with nodules between 5.1 and 8 mm were scheduled for repeating CT 3 to 6 months later. Patients with nodules greater than 8.1 mm, or growing lesions less than 8mm after repeated scan, were scheduled for PET-CT. Further investigations (repeat LDCT 6 months later, PET-CT, or surgical biopsy) for patients with growing nodules at subsequent annual screening depended on nodule density (non-solid, solid, or partially solid), growth rate and size. | |
| Rampinelli, 2017 (associated Veronesi 2008; Veronesi 2012, FPs at different timepoints)<br><br>Italy (Single screening center in Milan) | Inclusion criteria:<br>- Asymptomatic participants<br>- Age >50 years<br>- Smoking history with ≥20 pack years<br>- No history of cancer in the past five years<br><br>Eligible, n: 5,203<br>Enrolled, n: 5,203<br>Analyzed, n:<br>- Baseline: 5203<br>- First annual: 4822<br>- Second annual: 4583<br>- Third annual: 4380<br>- Fourth annual: 4120<br>- Fifth annual: 3856<br>- Sixth annual: 3654<br>- Seventh annual: 3449<br>- Eighth annual: 3182 | LDCT<br>One baseline LDCT and 9 follow-up scans at 1-year interval<br>Nodule management criteria: Patients with nodules between 5.1 and 8 mm were scheduled for repeating CT 3 to 6 months later. Patients with nodules greater than 8.1 mm, or growing lesions less than 8mm after repeated scan, were scheduled for PET-CT. Further investigations (repeat LDCT 6 months later, PET-CT, or surgical biopsy) for patients with growing nodules at subsequent annual screening depended on nodule density (non-solid, solid, or partially solid), growth rate and size. | False positives |

| Author, Year<br>Country (setting) | Inclusion criteria<br>Eligible for CT screening, *n*<br>Enrolled, *n*<br>Analyzed, *n*<br>Age, *mean ± SD/median ± IQR (yr)*<br>Gender, % male<br>Smoking status, current vs former smoker vs never smoker<br>Pack-years smoking history | Screening Strategy:<br>Type of CT scan<br>Number of rounds<br>Nodule management for a positive CT scan<br>Other relevant details<br>of screening process | Outcomes |
|---|---|---|---|
| | - Ninth annual: 2732<br><br>Age, Mean (SD): 57.7 (5.6)<br>Gender, % male: 66.0%<br>Current smoker, %: 80.3%<br>Former smoker, %:NR<br>Never smoker, %: NR<br>Pack-years smoking history, median (range): 44 (20-255) | | |
| Regan, 2019<br><br>USA (Genetic Epidemiology of COPD [COPDGene] study at 21 clinical centers) | Inclusion criteria:<br>- Age 55-79 years<br>- Current or former smokers who quit fewer than 15 years age<br>- Had at least a 30 pack-year smoking history<br><br>Eligible, n: 4,078<br>Enrolled, n: 4,078<br>Analyzed, n: varies by specific IF,<br>CAC: 4,078/4,078<br>Emphysema: 3,909/4,078 (95.9%)<br>Osteoporosis: 3,877/4,078 (95.1%)<br>Age, Mean (SD): 63.9 (5.9) (*n=4,055)<br>Gender, % male: 57% (*n=4,055)<br>Current smoker, %: 49%<br>Former smoker, %: NR<br>Never smoker, %: NR<br>Pack-years smoking history, mean (SD): 58.6 (25.0) (*n=4,055) | High dose CT scan<br>One baseline high dose CT scan<br>Nodule management criteria: NR | Incidental findings |
| Regis, 2022<br><br>USA (Single center [Lahey Hospital and Medical Center, Massachusetts]) | Inclusion criteria: Either,<br>- Individuals aged 55 to 74 years with a 30 or more pack-year history of smoking tobacco who currently smoke or, if former smoker, have quit within 15 years; or,<br>- Individuals aged 50 years or older with a 20 or more pack-year history of smoking tobacco and with one additional risk factor | CT<br>One baseline CT scan and 3 follow-up scans at 11+ months (>=335 days) after a negative exam.<br>LungRADS v1.1 criteria: Category 3 and 4 are considered positive. | Incidental findings (+ clinically significant incidental findings) |

| Author, Year<br>Country (setting) | Inclusion criteria<br>Eligible for CT screening, *n*<br>Enrolled, *n*<br>Analyzed, *n*<br>Age, *mean ± SD/median ± IQR (yr)*<br>Gender, % male<br>Smoking status, current vs former smoker vs never smoker<br>Pack-years smoking history | Screening Strategy:<br>Type of CT scan<br>Number of rounds<br>Nodule management for a positive CT scan<br>Other relevant details<br>of screening process | Outcomes |
|---|---|---|---|
| | (These additional risk factors include personal history of cancer or lung disease, family history of lung cancer, radon exposure, and occupational exposure to carcinogens)<br><br>Eligible, n: 4,301<br>Enrolled, n: 4,301<br>Analyzed, n: T0 – 4,301<br>T1 – 2,682<br>T2 – 1,773<br>T3 – 1,144<br>T4+ - 997<br><br>Age, Mean: 63.1<br>Gender, % male: 55.6%<br>Current smoker, %: NR<br>Former smoker, %: 52.6%<br>Never smoker, %: NR<br>Pack-years smoking history, mean: 48.5 pack-years | | |
| Reiter, 2018<br><br>USA (Single center academic university hospital) | Inclusion criteria:<br>- Group one comprises individuals aged 55–77 with 30 or more pack year smoking history and current or former smokers within past 15 years;<br>- Group two includes individuals aged 50 years or older with at least a 20-pack year history and one additional risk factor (family history, asbestosis exposure, etc.).<br><br>Eligible, n: NR<br>Enrolled, n: 581<br>Analyzed, n: 581<br><br>Age, Mean (SD): | LDCT<br>One baseline LDCT scan<br>LungRADs version 1.0: categories 3 and 4 are considered positive. | Incidental findings (+ clinically significant incidental findings) |

| Author, Year<br>Country (setting) | Inclusion criteria<br>Eligible for CT screening, *n*<br>Enrolled, *n*<br>Analyzed, *n*<br>Age, *mean ± SD/median ± IQR (yr)*<br>Gender, % male<br>Smoking status, current vs former smoker vs never smoker<br>Pack-years smoking history | Screening Strategy:<br>Type of CT scan<br>Number of rounds<br>Nodule management for a positive CT scan<br>Other relevant details<br>of screening process | Outcomes |
|---|---|---|---|
| | - LungRADS S modifier negative: 59.9 (6.2)<br>- LungRADS S modifier positive: 61.6 (6.4)<br>Gender, % male: 66.8%<br>Current smoker, %: 43.5%<br>Former smoker, %: NR<br>Never smoker, %: NR<br>Pack-years smoking history, mean (SD):<br>S modifier negative: 45.7 (24.4)<br>S modifier positive: 46.8 (22.8) | | |
| LSUT trial (Ruparel, 2020)<br><br>UK (Two hospitals in London, UK) | Inclusion criteria:<br>- Aged 60 to 75 years<br>- Recorded as current smokers within the seven preceding years were sent an invitation letter from their usual general practice doctor inviting them to an LHC<br>- ≥30 pack-years and if a former smoker had quit ≤15 years ago, or<br>- PLCOm2012 criteria: a lung cancer risk of ≥1.51%<br><br>Eligible, n: 859 (eligible for screening)<br>Enrolled, n: 859<br>Analyzed, n: 768<br><br>Age, Mean (SD): NR<br>Gender, % male: 54.9%<br>Current smoker, %: 70.9%<br>Former smoker, %: 29.1%<br>Never smoker, %: NR<br>Pack-years smoking history, median (IQR):36 (21-50) | LDCT<br>One baseline LDCT scan (11.9% undergoing 3 month and 4.3% undergoing 12-month surveillance for indeterminate nodules).<br>BTS guideline: Used the Brock model (full, with spiculation) for initial risk assessment of pulmonary nodules (≥8 mm or ≥300 mm3) at presentation in people aged ≥50 who are smokers or former smokers. | 1) False positives<br>2) Major complications from invasive diagnostic testing |
| Rzyman, 2015<br><br>Poland (19 radiological centers in Pomerania, Poland) | Inclusion criteria:<br>- Healthy asymptomatic participants<br>- Aged between 50 and 75 years<br>- Smoking history of 20 pack-years | LDCT<br>One baseline LDCT scan with one-year follow-up. 34.7% had annual repeat scan (2 rounds)<br>Nodule management criteria: Nodules with a diameter of 5 to 10 mm required follow-up visits 3, 6, and 12 months after the first LDCT; nodules >10 mm in diameter required further diagnostic workup. | 1) False positive<br>2) Major complications from invasive diagnostic testing<br>3) Incidental findings |

| Author, Year<br>Country (setting) | Inclusion criteria<br>Eligible for CT screening, *n*<br>Enrolled, *n*<br>Analyzed, *n*<br>Age, *mean ± SD/median ± IQR (yr)*<br>Gender, % male<br>Smoking status, current vs former smoker vs never smoker<br>Pack-years smoking history | Screening Strategy:<br>Type of CT scan<br>Number of rounds<br>Nodule management for a positive CT scan<br>Other relevant details<br>of screening process | Outcomes |
|---|---|---|---|
| | - Minimum smoking history of 10 pack-years among participants with family history of lung cancer<br><br>Eligible, n: NR<br>Enrolled, n: 8,649<br>Analyzed, n: 8,649<br><br>Age, Median: 60.6<br>Gender, % male: 52.0%<br>Current smoker, %: 68%<br>Former smoker, %: 32%<br>Never smoker, %: NR<br>Pack-years smoking history, mean (range): 35 (0-200) | | |
| Salvatore, 2016<br><br>USA (Icahn School of Medicine at Mount Sinai, New York) | Inclusion criteria: Participants aged 40-85 years old or older.<br><br>Eligible, n: 951<br>Enrolled, n: 951<br>Analyzed, n: 951<br><br>Age, Mean (SD): NR<br>Gender, % male: 46.7%<br>Current smoker, %: NR<br>Former smoker, %: NR<br>Never smoker, %: NR<br>- Pack-years smoking history, median: 34.8 pack-years | CT<br>Most recent CT scans of all the participants were used.<br>Nodule management criteria: NR | Specific incidental findings |
| Shah, 2024<br><br>USA (Chicago Fire Department Local 2 Union) | Inclusion criteria: Members of the Chicago Fire Department Local 2 Union were invited to undergo lung cancer screening with LDCT scans at a single institution between April 2022 and June 2023 within the pre-existing lung cancer screening program.<br><br>Eligible, n: NR<br>Enrolled, n: 1,347<br>Analyzed, n: 1,347 | LDCT<br>One baseline LDCT scan<br>LungRADs version v2022: category 3 or 4 are considered positive. | Incidental findings (+clinically significant incidental findings) |

| Author, Year<br>Country (setting) | Inclusion criteria<br>Eligible for CT screening, *n*<br>Enrolled, *n*<br>Analyzed, *n*<br>Age, *mean ± SD/median ± IQR (yr)*<br>Gender, % male<br>Smoking status, current vs former smoker vs never smoker<br>Pack-years smoking history | Screening Strategy:<br>Type of CT scan<br>Number of rounds<br>Nodule management for a positive CT scan<br>Other relevant details<br>of screening process | Outcomes |
|---|---|---|---|
| | Age, Median (IQR): 51 (42-58)<br>Gender, % male: 95.8%<br>Current smoker, %: 7.7%<br>Former smoker, %: 25.6%<br>Never smoker, %: 66.7%<br>Pack-years smoking history, median (IQR): 0 (0-4) | | |
| Song, 2018<br><br>South Korea (Hallym University hospital) | Inclusion criteria: Current smokers visiting Hallym University hospital to quit smoking and underwent LDCT, and a control group of non-smokers who underwent LDCT for general health screening.<br><br>Eligible, n: 304<br>Enrolled, n: 304<br>Analyzed, n: 304<br><br>Age, Mean:<br>- Non-smoker: 49.78<br>- Smoker: 54.26<br>Gender, % male: 79.3%<br>Current smoker, %: 57.2%<br>Former smoker, %: NR<br>Never smoker, %: NR<br>Pack-years smoking history, median (IQR): NR | LDCT<br>One baseline LDCT scan<br>Nodule management criteria: NR | Incidental findings |
| Steiger, 2021<br><br>Multiple participating sites including USA, Canada, China, Israel, Italy, Japan, Spain, and Switzerland | Inclusion criteria: Participants were 40-85 years old at the time of enrollment without any symptoms of lung cancer.<br><br>Eligible, n: NR<br>Enrolled, n: 52,726<br>Analyzed, n: 52,726<br><br>Age, Mean (SD): 60.3 (8.8)<br>Gender, % male: 55.8%<br>Current smoker, %: 37.4%<br>Former smoker, %:48.2%<br>Never smoker, %: 14.4% | LDCT<br>One baseline LDCT scan<br>Nodule management criteria: According to I-ELCAP protocol. | Specific incidental findings |

| Author, Year<br>Country (setting) | Inclusion criteria<br>Eligible for CT screening, *n*<br>Enrolled, *n*<br>Analyzed, *n*<br>Age, *mean ± SD/median ± IQR (yr)*<br>Gender, % male<br>Smoking status, current vs former smoker vs never smoker<br>Pack-years smoking history | Screening Strategy:<br>Type of CT scan<br>Number of rounds<br>Nodule management for a positive CT scan<br>Other relevant details<br>of screening process | Outcomes |
|---|---|---|---|
| | Pack-years smoking history, mean (SD): 31.2 (25.3) | | |
| Taghizadeh, 2019<br><br>Canada (8 study centers across Canada [Calgary, Halifax, Hamilton, Laval, Ottawa, St.John's, Toronto, and Vancouver]) | Inclusion criteria:<br>- Current or former smokers<br>- Age between 50 to 75 years<br>- With a 2% or greater lung cancer risk over 6 years using a risk-prediction model developed using PLCO Cancer Screening Trial data<br><br>Eligible, n: NR<br>Enrolled, n: 1,237<br>Analyzed, n: Among all screened,<br>- 1-month post-baseline CT scan: 284/1,237<br>- 12 months after baseline CT scan: 1,066/1,237<br><br>Age, Mean (SD): 62.9(6.1)<br>Gender, % male: 45.1%<br>Current smoker, %: 62.1%<br>Former smoker, %: 37.9%<br>Never smoker, %: NR<br>Pack-years smoking history, median (IQR): 51.3 (21.6) | LDCT<br>One baseline LDCT (and repeat LDCT at year 1 and year 4 in additional to scans as appropriate for nodule follow-up)<br>Nodule management criteria: Participants with any semisolid or solid nodule 5–10mm or ground-glass opacity 8–10mm were to receive an additional LDCT at 3 months, with larger lesions being referred for clinical consultation. Any participant requiring repeat LDCT or investigation for a lung lesion other than a planned 12-month follow-up examination were considered to have a positive screening exam for the purpose of this analysis. | 1) False positives<br>2) Psychosocial consequences of screening |
| Walker, 2015<br><br>USA (Lahey Hospital & Medical Center [single center], Burlington, Massachusetts) | Inclusion criteria: NCCN lung cancer screening guideline group 1 and 2 criteria.<br>Group 1:<br>- 55- 74 years old<br>- current or former smokers who had quit within the past 15 years<br>Group 2:<br>- 50-74 years old<br>- Had a 20 pack-year or more smoking history<br>- Were current or former smokers who had quit for any length of time | LDCT<br>Baseline LDCT scan and recall scans at 3-6 months in 45 individuals.<br>LungRADS criteria: category 4 was considered positive. | Major complications from invasive diagnostic testing |

| Author, Year<br>Country (setting) | Inclusion criteria<br>Eligible for CT screening, *n*<br>Enrolled, *n*<br>Analyzed, *n*<br>Age, *mean ± SD/median ± IQR (yr)*<br>Gender, % male<br>Smoking status, current vs former smoker vs never smoker<br>Pack-years smoking history | Screening Strategy:<br>Type of CT scan<br>Number of rounds<br>Nodule management for a positive CT scan<br>Other relevant details<br>of screening process | Outcomes |
|---|---|---|---|
| | - Had one additional lung cancer risk factor excluding second-hand smoke exposure<br><br>Eligible, n: 1,654<br>Enrolled, n: 1,654<br>Analyzed, n: 1,654<br><br>Age, Mean (SD): NR<br>Gender, % male: NR<br>Current smoker, %: NR<br>Former smoker, %: NR<br>Never smoker, %: NR<br>Pack-years smoking history, median (IQR): NR | | |
| Wilson, 2008<br><br>USA | Inclusion criteria:<br>- Aged 50-79 years<br>- No personal lung cancer history<br>- Non-participation in concurrent lung cancer screening studies<br>- No chest computed tomography within 12 months.<br>- Current or ex-cigarette smoker of at least one-half pack per day for at least 25 years<br>- Quit for no more than 10 years before study enrollment<br>- Body weight less than 400 pounds<br><br>Eligible, n: 5,034<br>Enrolled, n: 3,755<br>Analyzed, n:<br>- Baseline LDCT: 3642/3755 (97.0%)<br>- Repeat LDCT: 3423/3642 (94.0%)<br><br>Age, Mean: 59.0<br>Gender, % male: 51.4%<br>Current smoker, %: 60.2%<br>Former smoker, %: 39.8% | Spiral CT scan<br>One CT scan at baseline and one annual repeat<br>Nodule management criteria:<br>Initial CT: Low suspicion: < 0.5 cm average diameter or 0.5–0.9 cm average diameter with nonspiculated border; Moderate or high suspicion: 0.5–0.9 cm average diameter with speculated border or ≥ 1.0 cm average diameter.<br>Repeat CT: The radiologist applied subjective criteria to interpret temporal change according to a forced choice format. | 1) False positive<br>2) Clinically significant incidental findings |

| Author, Year<br>Country (setting) | Inclusion criteria<br>Eligible for CT screening, *n*<br>Enrolled, *n*<br>Analyzed, *n*<br>Age, *mean ± SD/median ± IQR (yr)*<br>Gender, % male<br>Smoking status, current vs former smoker vs never smoker<br>Pack-years smoking history | Screening Strategy:<br>Type of CT scan<br>Number of rounds<br>Nodule management for a positive CT scan<br>Other relevant details<br>of screening process | Outcomes |
|---|---|---|---|
| | Never smoker, %: NR<br>Pack-years smoking history, median (IQR): NR | | |
| Zhao, 2020<br><br>USA (Centers for Medicare and Medicaid Services) | Inclusion criteria:<br>- Age 55 – 77 years<br>- Asymptomatic (no signs or symptoms of lung cancer);<br>- Tobacco smoking history of at least 30 pack-years (one pack-year = smoking one pack per day for one year; 1 pack = 20 cigarettes);<br>- Current smoker or one who has quit smoking within the last 15 years<br><br>Eligible, n: 18,887<br>Enrolled, n: 18,887<br>Analyzed, n: 18,887<br><br>Age, Median (IQR): 61.0 (58-63)<br>Gender, % male: 52.8%<br>Current smoker or one who has quit smoking within the last 15 year: 100%<br>Tobacco smoking history of at least 30 pack-years: 100% | LDCT<br>One baseline LDCT scan<br>Nodule management criteria: NR | Major complications from invasive diagnostic testing |

Abbreviations: BTS: British Thoracic Society; IQR: interquartile range; LDCT = low dose CT; NR = not reported; SD =standard deviation.

Table S2.3. Risk of bias (comparative outcomes in trials): Mortality, overdiagnosis, HRQoL, and psychosocial consequences, 1-step CT screening

| Author, Year<br>Outcome | Domain 1: Randomization process | Domain 2: Deviations from intended intervention | Domain 3: Missing outcome data | Domain 4: Outcome measurement | Domain 5: Selection bias | Overall risk of bias |
|---|---|---|---|---|---|---|
| **NLST** | | | | | | |
| Pinsky, 2019<br>All-cause mortality | + | ? | + | + | ? | Some concerns |
| Pinsky, 2019<br>Lung cancer mortality | + | ? | + | + | + | Some concerns |
| Gareen, 2014<br>HRQoL | ? | ? | - | ? | + | High |
| **NELSON** | | | | | | |
| De Koning, 2020<br>All-cause mortality | + | ? | + | + | + | Some concerns |
| De Koning, 2020<br>Lung cancer mortality | + | ? | + | ? | + | Some concerns |
| De Koning, 2020<br>Overdiagnosis (men only) | + | ? | + | + | + | Some concerns |
| van den Bergh, 2011<br>HRQoL | + | ? | - | ? | + | High |
| van den Bergh, 2011<br>Psychosocial consequences<br>(Positive screen [before repeat CT or diagnostic results known, screen +ve]);<br>(Before dx results vs. control arm, before randomization]) | + | ? | + | ? | + | Some concerns |
| van den Bergh, 2011<br>Psychosocial consequences<br>(Participating in screening program [screening arm at 1.5 yrs vs. control arm at 2 yrs]);<br>(False positive screening result [>6 months after diagnostic results known vs. control arm]) | + | ? | - | ? | + | High |

| | | | | | | |
|---|---|---|---|---|---|---|
| van den Bergh, 2011<br>Psychosocial consequences (Participating in screening program [screening arm, before randomization vs. control arm before randomization]) | + | + | + | - | + | High |
| **DLCST** | | | | | | |
| Wille, 2016<br>All-cause mortality | ? | ? | + | + | + | Some concerns |
| Wille, 2016<br>Lung cancer mortality | ? | ? | + | + | + | Some concerns |
| Wille, 2016<br>Overdiagnosis | ? | ? | + | + | + | Some concerns |
| Rasmussen, 2015<br>Psychosocial consequences (Participating in screening program [screen arm before each round, 1-5 vs. control arm before each round 1-5]) | ? | ? | + | ? | + | Some concerns |
| Rasmussen, 2020<br>Psychosocial consequences (Positive screen [before repeat CT or diagnostic results known vs. control arm]);<br>False positive screening result [≤6 months after diagnostic results known vs. control arm]);<br>(False positive screening result [>6 months after diagnostic results known vs. control arm]) | ? | ? | + | ? | + | Some concerns |
| **DANTE** | | | | | | |

| | | | | | | |
|---|---|---|---|---|---|---|
| Infante, 2015<br>All-cause mortality (men only) | - | ? | + | + | ? | High |
| Infante, 2015<br>Lung cancer mortality (men only) | - | ? | + | ? | ? | High |
| Overdiagnosis (men only) | - | ? | + | - | ? | High |
| **ITALUNG** | | | | | | |
| Puliti, 2019<br>All-cause mortality | + | ? | + | + | ? | Some concerns |
| Paci, 2021<br>Lung cancer mortality | + | ? | + | ? | ? | Some concerns |
| Overdiagnosis | + | ? | + | + | + | Some concerns |
| **LUSI** | | | | | | |
| Maldonado, 2021<br>All-cause mortality | ? | ? | + | + | + | Some concerns |
| Maldonado, 2021<br>Lung cancer mortality | ? | ? | + | + | + | Some concerns |
| Maldonado, 2021<br>Overdiagnosis | ? | ? | + | ? | + | Some concerns |
| **MILD** | | | | | | |
| Pastorino, 2019<br>All-cause mortality | ? | ? | + | + | + | Some concerns |
| Pastorino, 2019<br>Lung cancer mortality | ? | ? | + | + | + | Some concerns |
| Pastorino, 2019<br>Overdiagnosis | ? | ? | + | - | + | High |
| **UKLS** | | | | | | |
| Davies, 2024<br>All-cause mortality | + | ? | + | + | + | Some concerns |

| | | | | | | |
|---|---|---|---|---|---|---|
| Davies, 2024<br>Lung cancer mortality | + | ? | + | + | + | Some concerns |
| Davies, 2024<br>Overdiagnosis | + | ? | + | + | + | Some concerns |
| Brain, 2016<br>Psychosocial harms (Participating in screening program [screen arm at 2 yrs vs. control arm at 2 yrs]) | + | ? | + | ? | + | Some concerns |
| Brain, 2016<br>Psychosocial harms (False positive screening result [>6 months after diagnostic results known vs. control arm]) | + | ? | - | ? | + | High |
| Brain, 2016<br>Psychosocial harms (Positive screen [before repeat CT or diagnostic results known,<br>vs. control arm at 2 wks]) | + | ? | + | ? | + | Some concerns |
| Brain, 2016<br>Psychosocial harms (Participating in screening program [screen arm, after randomization but before allocation known vs. control arm, after randomization but before allocation known]) | + | ? | + | + | + | Some concerns |
| **LSS** | | | | | | |
| Doroudi, 2018<br>All-cause mortality | + | ? | + | + | ? | Some concerns |
| Doroudi, 2018<br>Lung cancer mortality | + | ? | + | ? | ? | Some concerns |
| **Targeted CT screening trials** | | | | | | |
| **ECLS** | | | | | | |

| | | | | | | |
|---|---|---|---|---|---|---|
| Sullivan, 2025<br>All-cause mortality | + | ? | + | + | + | Some concerns |
| Sullivan, 2025<br>Lung cancer mortality | + | ? | + | ? | + | Some concerns |
| **LungSEARCH** | | | | | | |
| Spiro, 2019<br>All-cause mortality | + | ? | + | + | ? | Some concerns |
| Spiro, 2019<br>Lung cancer mortality | + | ? | + | ? | ? | Some concerns |

Abbreviations: “+”=low risk of bias; “?”=some concerns of bias; “-”=high risk of bias.

Table S2.4. Risk of bias: Harms (screening arm only), trials

| **Author Year Outcome (Screening rounds or timepoints for psychosocial harms)** | **Q1. Clear inclusion criteria?** | **Q2. Exposure (screening method) measured in standard, reliable way?** | **Q3. Valid methods for identification of exposure (screening method)?** | **Q4. Prospective: attempt to enroll all eligible? Retrospective: enroll more than just attenders to screening?** | **Q5a. Complete inclusion of eligible participants?** | **Q5b. Acceptable attrition rate for outcome being assessed?** | **Q6. Clear reporting of demographics?** | **Q7a. Outcome clearly defined and appropriate?** | **Q7b. Structured method of identifying outcomes?** | **Q7c. Follow-up duration adequate?** | **Overall ROB** |
|---|---|---|---|---|---|---|---|---|---|---|---|
| NLST (Aberle 2011) **Any FPs** (round 1) | Y | Y | Y | Y | U | Y | Y | Y | N | Y | High |
| NLST (Aberle 2011) **Any FPs & #Any FPs/person** (2/3 rounds) | Y | Y | Y | Y | U | Y | Y | Y | N | Y | High |
| NLST (Gareen 2023) **Any Ifs & #Any Ifs/person** (2/3 rounds) | Y | Y | Y | Y | U | Y | Y | Y | Y | Y | Some concerns |
| NLST (Gareen 2023) **Clinically significant Ifs** (round 1) | Y | Y | Y | Y | U | Y | Y | Y | Y | Y | Some concerns |
| NLST (Gareen 2023) **Clinically significant Ifs & #Clinically significant Ifs/person** (2/3 rounds) | Y | Y | Y | Y | U | Y | Y | Y | Y | Y | Some concerns |
| NLST (Pinsky 2014) **#Any MCs (FPs only)** | Y | Y | Y | Y | U | U | Y | Y | Y | Y | Some concerns |

| Author Year Outcome (Screening rounds or timepoints for psychosocial harms) | Q1. Clear inclusion criteria? | Q2. Exposure (screening method) measured in standard, reliable way? | Q3. Valid methods for identification of exposure (screening method)? | Q4. Prospective: attempt to enroll all eligible? Retrospective: enroll more than just attenders to screening? | Q5a. Complete inclusion of eligible participants? | Q5b. Acceptable attrition rate for outcome being assessed? | Q6. Clear reporting of demographics? | Q7a. Outcome clearly defined and appropriate? | Q7b. Structured method of identifying outcomes? | Q7c. Follow-up duration adequate? | Overall ROB |
|---|---|---|---|---|---|---|---|---|---|---|---|
| (round 1) | | | | | | | | | | | |
| NLST (Iaccarino 2019; Aberle 2011; Pinsky 2014) **Any MCs & #Any MCs (LCs+FPs)** (2/3 rounds) | Y | Y | Y | Y | U | U | Y | Y | Y | Y | Some concerns |
| NLST (Aberle 2011) **Deaths from IPs** (round 1) | Y | Y | Y | Y | U | U | Y | U | Y | Y | Some concerns |
| NLST (Gareen 2014) **Psychosocial consequences** (Positive screen [before repeat CT or diagnostic results known vs. before randomization [in screen +ve]) | Y | Y | Y | Y | Y | Y | U | Y | Y | Y | Some concerns |
| NLST (Gareen, 2014) **Psychosocial consequences** (False positive screening result [≤6 months after diagnostic results known | Y | Y | Y | Y | Y | N | U | Y | Y | Y | High |

| Author Year Outcome (Screening rounds or timepoints for psychosocial harms) | Q1. Clear inclusion criteria? | Q2. Exposure (screening method) measured in standard, reliable way? | Q3. Valid methods for identification of exposure (screening method)? | Q4. Prospective: attempt to enroll all eligible? Retrospective: enroll more than just attenders to screening? | Q5a. Complete inclusion of eligible participants? | Q5b. Acceptable attrition rate for outcome being assessed? | Q6. Clear reporting of demographics? | Q7a. Outcome clearly defined and appropriate? | Q7b. Structured method of identifying outcomes? | Q7c. Follow-up duration adequate? | Overall ROB |
|---|---|---|---|---|---|---|---|---|---|---|---|
| vs. before randomization [in FPs]) | | | | | | | | | | | |
| NELSON (Horeweg 2013) **Any FPs** (1 round) | Y | Y | Y | Y | Y | Y | U | Y | Y | Y | Some concerns |
| NELSON (Horeweg 2013) Any FPs @ dx (1 round) | Y | Y | Y | Y | Y | Y | U | Y | Y | Y | Some concerns |
| NELSON (Horeweg 2013) **#Any FPs/person** (2/3 rounds) | Y | Y | Y | Y | Y | Y | U | Y | Y | Y | Some concerns |
| NELSON (Horeweg 2013) **Any FPs @ dx & #Any FPs @ dx/person** (2/3 rounds) | Y | Y | Y | Y | Y | U | U | Y | Y | Y | Some concerns |
| NELSON (de Koning 2020: men only) **#Any FPs/person** (4/5 rounds) | Y | Y | Y | Y | Y | U | U | Y | U | Y | Some concerns |
| NELSON (de Koning 2020: men only) **#Any FPs @ dx/person** (4/5 rounds) | Y | Y | Y | Y | Y | U | U | Y | U | Y | Some concerns |

| Author Year Outcome (Screening rounds or timepoints for psychosocial harms) | Q1. Clear inclusion criteria? | Q2. Exposure (screening method) measured in standard, reliable way? | Q3. Valid methods for identification of exposure (screening method)? | Q4. Prospective: attempt to enroll all eligible? Retrospective: enroll more than just attenders to screening? | Q5a. Complete inclusion of eligible participants? | Q5b. Acceptable attrition rate for outcome being assessed? | Q6. Clear reporting of demographics? | Q7a. Outcome clearly defined and appropriate? | Q7b. Structured method of identifying outcomes? | Q7c. Follow-up duration adequate? | Overall ROB |
|---|---|---|---|---|---|---|---|---|---|---|---|
| NELSON (Horeweg 2013) **#Any IFs/person** (2/3 rounds) | Y | Y | Y | Y | Y | Y | U | N | Y | Y | High |
| NELSON (de Koning 2020: men only) **Any MCs** (1 round) | Y | Y | Y | Y | Y | U | U | U | U | Y | Some concerns |
| NELSON (de Koning 2020: men only) **Any MCs** (4/5 rounds) | Y | Y | Y | Y | Y | U | U | U | U | U | Some concerns |
| NELSON (van den Bergh 2010) **Psychosocial consequences of screening** (Undergoing CT scan [after screening, before results are known, screen arm, after screening with scan results unknown vs. before randomization]) | Y | Y | Y | Y | Y | Y | U | Y | Y | Y | Some concerns |
| NELSON (van den Bergh 2011) **Psychosocial consequences** | Y | Y | Y | Y | Y | U | U | Y | Y | Y | Some concerns |

| **Author Year Outcome (Screening rounds or timepoints for psychosocial harms)** | **Q1. Clear inclusion criteria?** | **Q2. Exposure (screening method) measured in standard, reliable way?** | **Q3. Valid methods for identification of exposure (screening method)?** | **Q4. Prospective: attempt to enroll all eligible? Retrospective: enroll more than just attenders to screening?** | **Q5a. Complete inclusion of eligible participants?** | **Q5b. Acceptable attrition rate for outcome being assessed?** | **Q6. Clear reporting of demographics?** | **Q7a. Outcome clearly defined and appropriate?** | **Q7b. Structured method of identifying outcomes?** | **Q7c. Follow-up duration adequate?** | **Overall ROB** |
|---|---|---|---|---|---|---|---|---|---|---|---|
| **of screening** (Positive screen [before repeat CT or diagnostic results known, vs. before randomization]); (False positive screening result [>6 months after diagnostic results known vs. before randomization]) | | | | | | | | | | | |
| ITALUNG (Lopes Pegna 2013) **Any FPs** (1 round) | Y | Y | Y | Y | Y | N | U | Y | Y | Y | High |
| ITALUNG (Lopes Pegna 2013) **Any FPs @ dx** (1 round) | Y | Y | Y | Y | Y | N | U | U | Y | Y | High |
| ITALUNG (Lopes Pegna 2013) **Any FPs & #Any FPs/person** (4/5 rounds) | Y | Y | Y | Y | Y | Y | U | Y | Y | Y | Some concerns |
| ITALUNG (Lopes Pegna 2013) **Any FPs @ dx** | Y | Y | Y | Y | Y | Y | U | U | Y | Y | Some concerns |

| Author Year Outcome (Screening rounds or timepoints for psychosocial harms) | Q1. Clear inclusion criteria? | Q2. Exposure (screening method) measured in standard, reliable way? | Q3. Valid methods for identification of exposure (screening method)? | Q4. Prospective: attempt to enroll all eligible? Retrospective: enroll more than just attenders to screening? | Q5a. Complete inclusion of eligible participants? | Q5b. Acceptable attrition rate for outcome being assessed? | Q6. Clear reporting of demographics? | Q7a. Outcome clearly defined and appropriate? | Q7b. Structured method of identifying outcomes? | Q7c. Follow-up duration adequate? | Overall ROB |
|---|---|---|---|---|---|---|---|---|---|---|---|
| (4/5 rounds) | | | | | | | | | | | |
| ITALUNG (Puliti 2019 & Lopes Pegna 2013) **Any IFs** (4/5 rounds) | Y | Y | Y | Y | Y | U | U | Y | U | Y | Some concerns |
| ITALUNG (Lopes Pegna 2013) **Any MCs** (4/5 rounds) | Y | Y | Y | Y | Y | U | U | U | Y | Y | Some concerns |
| ITALUNG (Paci 2017) **Deaths from IPs** (4/5 rounds) | Y | Y | Y | Y | Y | U | U | Y | Y | Y | Some concerns |
| LUSI (Becker 2020) **Any FPs** (1 round) | Y | Y | Y | Y | Y | U | U | Y | Y | Y | Some concerns |
| LUSI (Becker 2020) **Any FPs @ dx** (1 round) | Y | Y | Y | Y | Y | Y | U | Y | Y | Y | Some concerns |
| LUSI (Becker 2020) **#Any FPs/person** (4/5 rounds) | Y | Y | Y | Y | Y | U | U | Y | Y | Y | Some concerns |
| LUSI (Becker 2020) **#Any FPs @ dx/person** (4/5 rounds) | Y | Y | Y | Y | Y | Y | U | Y | Y | Y | Some concerns |

| Author Year Outcome (Screening rounds or timepoints for psychosocial harms) | Q1. Clear inclusion criteria? | Q2. Exposure (screening method) measured in standard, reliable way? | Q3. Valid methods for identification of exposure (screening method)? | Q4. Prospective: attempt to enroll all eligible? Retrospective: enroll more than just attenders to screening? | Q5a. Complete inclusion of eligible participants? | Q5b. Acceptable attrition rate for outcome being assessed? | Q6. Clear reporting of demographics? | Q7a. Outcome clearly defined and appropriate? | Q7b. Structured method of identifying outcomes? | Q7c. Follow-up duration adequate? | Overall ROB |
|---|---|---|---|---|---|---|---|---|---|---|---|
| DLCST (Pederson 2009) **Any FPs** (1 round) | Y | Y | Y | N | Y | Y | Y | Y | Y | Y | Some concerns |
| DLCST (Pederson 2009) **Any FPs @ dx** (1 round) | Y | Y | Y | N | Y | U | Y | Y | Y | Y | Some concerns |
| DLCST (Saghir 2012) **#Any FPs/person** (4/5 rounds) | Y | Y | Y | N | Y | U | Y | Y | Y | Y | Some concerns |
| DANTE (Infante 2008: men only) **Any FPs** (1 round) | Y | Y | Y | U | Y | Y | Y | Y | Y | Y | Some concerns |
| DANTE (Infante 2008: men only) **Any FPs @ dx** (1 round) | Y | Y | Y | U | Y | Y | Y | Y | Y | Y | Some concerns |
| DANTE (Infante 2015: men only) **Any FPs** (4/5 rounds) | Y | Y | Y | U | Y | U | Y | Y | Y | Y | Some concerns |
| DANTE (Infante 2015: men only) **Any FPs @ dx** (4/5 rounds) | Y | Y | Y | U | Y | U | Y | Y | Y | Y | Some concerns |
| DANTE (Infante 2008: men only) **Any MCs** (1 round) | Y | Y | Y | U | Y | U | Y | U | U | Y | Some concerns |
| DANTE (Infante 2015: men only) | Y | Y | Y | U | Y | U | Y | Y | Y | Y | Some concerns |

| Author Year Outcome (Screening rounds or timepoints for psychosocial harms) | Q1. Clear inclusion criteria? | Q2. Exposure (screening method) measured in standard, reliable way? | Q3. Valid methods for identification of exposure (screening method)? | Q4. Prospective: attempt to enroll all eligible? Retrospective: enroll more than just attenders to screening? | Q5a. Complete inclusion of eligible participants? | Q5b. Acceptable attrition rate for outcome being assessed? | Q6. Clear reporting of demographics? | Q7a. Outcome clearly defined and appropriate? | Q7b. Structured method of identifying outcomes? | Q7c. Follow-up duration adequate? | Overall ROB |
|---|---|---|---|---|---|---|---|---|---|---|---|
| **Deaths from IPs** (1 round) | | | | | | | | | | | |
| DANTE (Infante 2015: men only) **Deaths from IPs** (4/5 rounds) | Y | Y | Y | U | Y | U | Y | Y | Y | Y | Some concerns |
| LSS (Gohagan 2004) **Any FPs** (1 round) | Y | Y | Y | Y | U | Y | U | Y | N | Y | High |
| LSS (Gohagan 2004) **Any FPs @ dx** (1 round) | Y | Y | Y | Y | U | Y | U | Y | N | Y | High |
| LSS (Croswell 2010) **Any FPs & #Any FPs/person** (2/3 rounds) | Y | Y | Y | Y | U | Y | U | Y | N | Y | High |
| LSS (Croswell 2010) **Any FPs @ dx & #Any FPs @ dx/person** (2/3 rounds) | Y | Y | Y | Y | U | Y | U | Y | N | Y | High |
| LSS (Gohagan 2004) **Any MCs & #Any MCs/person** (round 1) | Y | Y | Y | Y | U | U | U | Y | Y | Y | Some concerns |
| LSS (Gohagan 2004) **Deaths from IPs** | Y | Y | Y | Y | U | U | U | Y | Y | Y | Some concerns |

| **Author Year Outcome (Screening rounds or timepoints for psychosocial harms)** | **Q1. Clear inclusion criteria?** | **Q2. Exposure (screening method) measured in standard, reliable way?** | **Q3. Valid methods for identification of exposure (screening method)?** | **Q4. Prospective: attempt to enroll all eligible? Retrospective: enroll more than just attenders to screening?** | **Q5a. Complete inclusion of eligible participants?** | **Q5b. Acceptable attrition rate for outcome being assessed?** | **Q6. Clear reporting of demographics?** | **Q7a. Outcome clearly defined and appropriate?** | **Q7b. Structured method of identifying outcomes?** | **Q7c. Follow-up duration adequate?** | **Overall ROB** |
|---|---|---|---|---|---|---|---|---|---|---|---|
| (1 round) | | | | | | | | | | | |
| LSS (Gohagan 2004 & Gohagan 2005)<br>**#Any MCs/person**<br>(2/3 rounds) | Y | Y | Y | Y | U | U | U | Y | Y | Y | Some concerns |
| UKLS (Mascalchi 2016)<br>**Any FPs**<br>(1 round) | Y | Y | Y | Y | N | U | Y | Y | Y | Y | Some concerns |
| UKLS (Field 2021)<br>**Any FPs @ dx**<br>(1 round) | Y | Y | Y | Y | N | U | Y | Y | Y | Y | Some concerns |
| UKLS (Field 2021)<br>**#Clinically significant Ifs/person**<br>(1 round) | Y | Y | Y | Y | N | U | Y | Y | Y | Y | Some concerns |
| MILD (Pastorino 2012)<br>**Any FPs**<br>(1 round) | Y | Y | Y | Y | U | Y | U | Y | Y | Y | Some concerns |
| MILD (Sverzellati 2016)<br>**Any FPs @ dx**<br>(1 round) | Y | Y | Y | N | U | Y | U | Y | Y | Y | Some concerns |
| MILD (Sverzellati 2016)<br>**#Any FPs/person**<br>(4/5 rounds) | Y | Y | Y | N | U | Y | U | Y | Y | Y | Some concerns |

| Author Year Outcome (Screening rounds or timepoints for psychosocial harms) | Q1. Clear inclusion criteria? | Q2. Exposure (screening method) measured in standard, reliable way? | Q3. Valid methods for identification of exposure (screening method)? | Q4. Prospective: attempt to enroll all eligible? Retrospective: enroll more than just attenders to screening? | Q5a. Complete inclusion of eligible participants? | Q5b. Acceptable attrition rate for outcome being assessed? | Q6. Clear reporting of demographics? | Q7a. Outcome clearly defined and appropriate? | Q7b. Structured method of identifying outcomes? | Q7c. Follow-up duration adequate? | Overall ROB |
|---|---|---|---|---|---|---|---|---|---|---|---|
| MILD (Sverzellati 2016) **#Any FPs @ dx/person** (4/5 rounds) | Y | Y | Y | N | U | Y | U | Y | Y | Y | Some concerns |
| Trials with no benefit data reported | | | | | | | | | | | |
| LSUT (Ruparel 2020) **Any FPs** (1 round) | Y | Y | Y | Y | Y | Y | Y | Y | Y | Y | Low |
| LSUT (Ruparel 2020) **Any FPs @ dx** (1 round) | Y | Y | Y | Y | Y | Y | Y | U | Y | Y | Some concerns |
| LSUT (Ruparel 2020) **Any MCs & #Any MCs/person** (1 round) | Y | Y | Y | Y | Y | Y | Y | Y | Y | U | Some concerns |
| LSUT (Hall 2022) **#Any Ifs/person** (1 round) | Y | Y | Y | N | Y | Y | Y | N | Y | Y | High |
| LSUT (Kummer 2020) **Clinically significant Ifs** (1 round) | Y | Y | Y | Y | Y | N | U | N | U | U | High |
| LSUT (Kummer 2020) **Psychosocial consequences of screening** | Y | Y | Y | Y | U | N | U | Y | Y | Y | High |

| Author Year Outcome (Screening rounds or timepoints for psychosocial harms) | Q1. Clear inclusion criteria? | Q2. Exposure (screening method) measured in standard, reliable way? | Q3. Valid methods for identification of exposure (screening method)? | Q4. Prospective: attempt to enroll all eligible? Retrospective: enroll more than just attenders to screening? | Q5a. Complete inclusion of eligible participants? | Q5b. Acceptable attrition rate for outcome being assessed? | Q6. Clear reporting of demographics? | Q7a. Outcome clearly defined and appropriate? | Q7b. Structured method of identifying outcomes? | Q7c. Follow-up duration adequate? | Overall ROB |
|---|---|---|---|---|---|---|---|---|---|---|---|
| (screen arms [pooled], Undergoing CT scan (after screening, before results are known) vs. after randomization, before screening) | | | | | | | | | | | |
| LSUT (Kummer 2020) **Psychosocial consequences of screening** (False positive screening result [≤6 months after diagnostic results known vs. after randomization, before screening]) | Y | Y | Y | Y | U | N | U | Y | Y | Y | High |
| DEPISCAN (Blanchon 2007) **Any FPs** (1 round) | Y | Y | Y | Y | Y | Y | U | Y | Y | Y | Some concerns |
| DEPISCAN (Blanchon 2007) **#Any Ifs/person** (1 round) | Y | Y | Y | Y | Y | Y | U | N | Y | Y | High |
| DEPISCAN (Blanchon 2007) | Y | Y | Y | Y | Y | Y | U | Y | Y | Y | Some concerns |

| Author Year Outcome (Screening rounds or timepoints for psychosocial harms) | Q1. Clear inclusion criteria? | Q2. Exposure (screening method) measured in standard, reliable way? | Q3. Valid methods for identification of exposure (screening method)? | Q4. Prospective: attempt to enroll all eligible? Retrospective: enroll more than just attenders to screening? | Q5a. Complete inclusion of eligible participants? | Q5b. Acceptable attrition rate for outcome being assessed? | Q6. Clear reporting of demographics? | Q7a. Outcome clearly defined and appropriate? | Q7b. Structured method of identifying outcomes? | Q7c. Follow-up duration adequate? | Overall ROB |
|---|---|---|---|---|---|---|---|---|---|---|---|
| **Any MCs** (1 round) | | | | | | | | | | | |
| DEPISCAN (Blanchon 2007) **Deaths from IPs** (1 round) | Y | Y | Y | Y | Y | Y | U | Y | Y | Y | Some concerns |

Abbreviations: FPs: False positives; IFs: Incidental findings; @dx: At diagnostic follow-up (excluding repeat scans); IPs: invasive diagnostic procedures; MCs: major complications (from invasive diagnostic procedures); N: No; RoB: Risk of bias; U: Unclear; Y: Yes

Notes on RoB:

- Overall RoB was rated as: low if all domains were rated as Y; some concerns if any domains was rated as U; high if at least one of the main domains was rated as N (5b, 7a, and 7b for FP, MC and psychosocial consequences; 7a and 7b for IF).
- Q5a was rated as Y if ≥80% of eligible participants were enrolled.
- Q5b was rated as Y if ≥90% of those who were recalled (for FP) or referred for additional diagnostic workup (for FP@ dx or for IF) attended the subsequent follow-up.
- Q6 was rated as: Y if age, sex, smoking, and at least one comorbidity were reported, U if three of the four variables were reported, and N if less than three of variables were reported.

Table S2.5. Risk of bias: Observational harms studies, case series

| Author Year Outcome (Screening rounds or timepoints for psychosocial harms) | Q1. Clear inclusion criteria? | Q2. Exposure (screening method) measured in standard, reliable way? | Q3. Valid methods for identification of exposure (screening method)? | Q4. Prospective: attempt to enroll all eligible? Retrospective: enroll more than just attenders to screening? | Q5a. Complete inclusion of eligible participants? | Q5b. Acceptable attrition rate for outcome being assessed? | Q6. Clear reporting of demographics? | Q7a. Outcome clearly defined and appropriate? | Q7b. Structured method of identifying outcomes? | Q7c. Follow-up duration adequate? | Overall ROB |
|---|---|---|---|---|---|---|---|---|---|---|---|
| Antonicelli 2024 **Specific Ifs** (1 rounds) | Y | Y | Y | Y | U | Y | Y | Y | Y | Y | Some concerns |
| Bartlett 2020 **Any FPs** (1 round) | Y | Y | Y | U | Y | Y | N | Y | N | Y | High |
| Bartlett 2020 **Any FPs @ dx** (1 round) | Y | Y | Y | U | Y | Y | N | Y | N | Y | High |
| Bartlett 2021 **Clinically significant IF & #Clinically significant IF/person** (1 round) | Y | Y | Y | U | Y | Y | N | N | N | U | High |
| Bhamani 2025 **Specific Ifs** (1 round) | Y | Y | Y | Y | Y | Y | Y | N | U | U | High |
| Bhamani 2025 **Any FPs** (1 round) | Y | Y | Y | Y | Y | Y | Y | Y | U | Y | Some concerns |
| Bonney 2024 **Psychosocial consequences** (1 round) | Y | Y | Y | Y | U | N | U | U | Y | Y | High |
| Bonney 2025 **Any IF** (1 round) | Y | Y | Y | Y | U | Y | Y | U | N | U | High |
| Bonney 2025 **Clinically significant Ifs** (1 round) | Y | Y | Y | Y | U | Y | Y | U | N | U | High |

| **Author Year Outcome (Screening rounds or timepoints for psychosocial harms)** | **Q1. Clear inclusion criteria?** | **Q2. Exposure (screening method) measured in standard, reliable way?** | **Q3. Valid methods for identification of exposure (screening method)?** | **Q4. Prospective: attempt to enroll all eligible? Retrospective: enroll more than just attenders to screening?** | **Q5a. Complete inclusion of eligible participants?** | **Q5b. Acceptable attrition rate for outcome being assessed?** | **Q6. Clear reporting of demographics?** | **Q7a. Outcome clearly defined and appropriate?** | **Q7b. Structured method of identifying outcomes?** | **Q7c. Follow-up duration adequate?** | **Overall ROB** |
|---|---|---|---|---|---|---|---|---|---|---|---|
| Byrne 2008 **Psychosocial consequences** (Positive screen [before repeat CT or diagnostic results known vs. before screening, in screen +ve]); (False positive screening result [≤6 months after diagnostic results known vs. before screening, in FPs]); (False positive screening result [>6 months after diagnostic results known vs. before screening, in FPs]) | Y | Y | Y | Y | Y | Y | U | Y | Y | Y | Some concerns |
| Callol 2007 **Any FPs** (1 round) | Y | Y | Y | U | Y | Y | U | Y | U | Y | Some concerns |
| Callol 2007 **Any FPs @ dx** (1 round) | Y | Y | Y | U | Y | Y | U | Y | U | Y | Some concerns |
| Callol 2007 **Any IFs** (1 round) | Y | Y | Y | U | Y | Y | U | U | U | Y | Some concerns |

| **Author Year Outcome (Screening rounds or timepoints for psychosocial harms)** | **Q1. Clear inclusion criteria?** | **Q2. Exposure (screening method) measured in standard, reliable way?** | **Q3. Valid methods for identification of exposure (screening method)?** | **Q4. Prospective: attempt to enroll all eligible? Retrospective: enroll more than just attenders to screening?** | **Q5a. Complete inclusion of eligible participants?** | **Q5b. Acceptable attrition rate for outcome being assessed?** | **Q6. Clear reporting of demographics?** | **Q7a. Outcome clearly defined and appropriate?** | **Q7b. Structured method of identifying outcomes?** | **Q7c. Follow-up duration adequate?** | **Overall ROB** |
|---|---|---|---|---|---|---|---|---|---|---|---|
| Callol 2007 **Any FPs** (2/3 rounds) | Y | Y | Y | U | Y | Y | U | Y | U | Y | Some concerns |
| Callol 2007 **Any FPs @ dx** (2/3 rounds) | Y | Y | Y | U | Y | Y | U | Y | U | Y | Some concerns |
| Cattaneo 2018 **Any MCs among FPs** (round 1) | U | Y | Y | N | U | U | U | Y | U | Y | Some concerns |
| Crestanello 2004 **#Clinically significant IFs/person** (2/3 rounds) | Y | Y | Y | U | U | Y | U | U | U | Y | Some concerns |
| Crosbie 2019 **Any FPs** (1 round) | Y | Y | Y | Y | Y | Y | Y | Y | Y | U | Some concerns |
| Crosbie 2019 **Any FPs @ dx** (1 round) | Y | Y | Y | Y | Y | Y | Y | Y | Y | U | Some concerns |
| Danila 2025 **Clinically significant Ifs** (1 round) | Y | Y | Y | Y | U | Y | U | U | U | U | Some concerns |
| Darling 2021 **Clinically significant IFs** (1 round) | Y | Y | Y | Y | Y | Y | N | N | U | Y | High |
| Erkmen 2022 **Any MCs & #Any MCs/person** (1 round) | Y | U | Y | Y | Y | Y | Y | Y | Y | U | Some concerns |

| Author Year Outcome (Screening rounds or timepoints for psychosocial harms) | Q1. Clear inclusion criteria? | Q2. Exposure (screening method) measured in standard, reliable way? | Q3. Valid methods for identification of exposure (screening method)? | Q4. Prospective: attempt to enroll all eligible? Retrospective: enroll more than just attenders to screening? | Q5a. Complete inclusion of eligible participants? | Q5b. Acceptable attrition rate for outcome being assessed? | Q6. Clear reporting of demographics? | Q7a. Outcome clearly defined and appropriate? | Q7b. Structured method of identifying outcomes? | Q7c. Follow-up duration adequate? | Overall ROB |
|---|---|---|---|---|---|---|---|---|---|---|---|
| Fabbrini 2018 **Any Ifs & #Any Ifs/person** (1 round) | Y | Y | Y | Y | N | Y | U | N | U | U | High |
| Fisher-Dortmann 2024 **#Any Ifs/person** (1 round) | Y | Y | Y | Y | U | U | N | U | U | Y | Some concerns |
| Guichet 2018 **Clinically significant IFs & #Clinically significant IFs/person** (1 round) | Y | Y | Y | U | Y | Y | Y | N | Y | U | High |
| Hammer 2023 **Specific Ifs** (1 round) | Y | Y | Y | N | Y | Y | Y | N | U | U | High |
| Henderson 2023 **Clinically significant Ifs** (1 round) | U | Y | Y | N | Y | Y | Y | N | U | U | High |
| Henschke 2001 **Any FPs** (1 round) | Y | Y | U | U | U | U | Y | Y | Y | Y | Some concerns |
| Henschke 2001 **Any FPs @ dx** (1 rounds) | Y | Y | U | U | U | U | Y | Y | Y | Y | Some concerns |
| Henschke 2001 **Any FPs** (2/3 roundS) | Y | Y | U | U | U | U | Y | Y | Y | Y | Some concerns |
| Henschke 2001 **Any FPs @ dx** (2/3 rounds) | Y | Y | U | U | U | U | Y | Y | Y | Y | Some concerns |

| **Author Year Outcome (Screening rounds or timepoints for psychosocial harms)** | **Q1. Clear inclusion criteria?** | **Q2. Exposure (screening method) measured in standard, reliable way?** | **Q3. Valid methods for identification of exposure (screening method)?** | **Q4. Prospective: attempt to enroll all eligible? Retrospective: enroll more than just attenders to screening?** | **Q5a. Complete inclusion of eligible participants?** | **Q5b. Acceptable attrition rate for outcome being assessed?** | **Q6. Clear reporting of demographics?** | **Q7a. Outcome clearly defined and appropriate?** | **Q7b. Structured method of identifying outcomes?** | **Q7c. Follow-up duration adequate?** | **Overall ROB** |
|---|---|---|---|---|---|---|---|---|---|---|---|
| Hirsch 2023 **Clinically significant Ifs** (1 round) | Y | Y | Y | Y | U | Y | U | N | U | U | High |
| Janssen 2019 **Any IFs & #Any IFs/person** (1 round) | N | U | Y | N | Y | Y | U | N | U | U | High |
| Kapoor 2020 **#Any IFs/person** (1 round) | Y | Y | Y | N | Y | Y | Y | N | Y | U | High |
| Kapoor 2020 **Clinically significant IFs & #Clinically significant IFs/person** (1 round) | Y | Y | Y | N | Y | Y | Y | N | Y | Y | High |
| Kearney 2023 **Clinically significant Ifs** (1 round) | Y | Y | Y | N | Y | Y | N | N | U | U | High |
| Kim 2020 **Any FPs** (2/3 rounds) | Y | Y | Y | N | Y | U | U | U | U | U | Some concerns |
| Kim 2020 **Any FPs @ dx** (2/3 rounds) | Y | Y | Y | N | Y | U | U | Y | U | U | Some concerns |
| Kim 2020 **#Any MCs/person** (2/3 rounds) | Y | Y | Y | N | Y | U | U | Y | U | Y | Some concerns |
| Kim 2020 **Deaths from IPs** | Y | Y | Y | N | Y | U | U | Y | U | U | Some concerns |

| Author Year Outcome (Screening rounds or timepoints for psychosocial harms) | Q1. Clear inclusion criteria? | Q2. Exposure (screening method) measured in standard, reliable way? | Q3. Valid methods for identification of exposure (screening method)? | Q4. Prospective: attempt to enroll all eligible? Retrospective: enroll more than just attenders to screening? | Q5a. Complete inclusion of eligible participants? | Q5b. Acceptable attrition rate for outcome being assessed? | Q6. Clear reporting of demographics? | Q7a. Outcome clearly defined and appropriate? | Q7b. Structured method of identifying outcomes? | Q7c. Follow-up duration adequate? | Overall ROB |
|---|---|---|---|---|---|---|---|---|---|---|---|
| (1 round) | | | | | | | | | | | |
| Kim 2020<br>**Deaths from IPs**<br>(2/3 rounds) | Y | Y | Y | N | Y | U | U | Y | U | U | Some concerns |
| Kim 2025<br>**Specific Ifs**<br>(1 round) | Y | Y | Y | N | Y | Y | Y | N | U | U | High |
| Kinsinger 2017<br>**Clinically significant IFs & #Clinically significant IFs/person**<br>(1 round) | Y | U | Y | U | Y | Y | U | N | U | U | High |
| Kucharczyk 2011<br>**Clinically significant IFs & #Clinically significant IFs/person**<br>(1 round) | Y | Y | Y | N | U | Y | U | N | Y | U | High |
| Laisaar 2025<br>**Any IFs**<br>(1 round) | Y | Y | Y | Y | Y | Y | N | N | U | Y | High |
| Laisaar 2025<br>**Clinically significant IFs**<br>(1 round) | Y | Y | Y | Y | Y | Y | N | N | U | Y | High |
| Lam 2015<br>**Any IFs**<br>(1 round) | Y | Y | Y | Y | U | Y | U | N | Y | Y | High |
| Lam 2015<br>**Clinically significant IFs** | Y | Y | Y | Y | U | Y | U | N | Y | Y | High |

| Author Year Outcome (Screening rounds or timepoints for psychosocial harms) | Q1. Clear inclusion criteria? | Q2. Exposure (screening method) measured in standard, reliable way? | Q3. Valid methods for identification of exposure (screening method)? | Q4. Prospective: attempt to enroll all eligible? Retrospective: enroll more than just attenders to screening? | Q5a. Complete inclusion of eligible participants? | Q5b. Acceptable attrition rate for outcome being assessed? | Q6. Clear reporting of demographics? | Q7a. Outcome clearly defined and appropriate? | Q7b. Structured method of identifying outcomes? | Q7c. Follow-up duration adequate? | Overall ROB |
|---|---|---|---|---|---|---|---|---|---|---|---|
| (1 rounds) | | | | | | | | | | | |
| Lam 2015<br>**Any FPs**<br>(1 round) | Y | Y | Y | Y | U | N | U | Y | Y | U | High |
| Lam 2015<br>**Any FPs @ dx**<br>(1 round) | Y | Y | Y | Y | U | N | U | Y | Y | U | High |
| Lam 2015<br>**Any MCs**<br>(2/3 rounds) | Y | Y | Y | Y | U | N | U | Y | U | U | High |
| Ledford 2016<br>**Any IFs**<br>(1 round) | Y | U | Y | N | Y | Y | N | N | U | U | High |
| Ledford 2016<br>**Clinically significant IFs**<br>(1 round) | Y | U | Y | N | Y | Y | N | N | U | U | High |
| Lee 2013<br>**Specific Ifs**<br>(1 round) | U | Y | Y | N | Y | Y | N | U | U | U | Some concerns |
| Lee 2021<br>**Any MCs**<br>(1 round) | Y | Y | Y | Y | U | N | Y | Y | U | Y | High |
| Lee 2021<br>**Deaths from IPs**<br>(1 round) | Y | Y | Y | Y | U | N | Y | Y | U | Y | High |
| Leleu 2024<br>**Any FPs**<br>(1 round) | Y | Y | Y | Y | Y | N | U | Y | U | Y | High |
| Lutzow 2022<br>**Deaths from IPs**<br>(1 round) | U | U | Y | U | U | U | U | Y | Y | U | Some concerns |
| Lutzow 2022<br>**Specific IFs** | U | U | Y | U | U | Y | U | N | U | Y | High |

| **Author Year Outcome (Screening rounds or timepoints for psychosocial harms)** | **Q1. Clear inclusion criteria?** | **Q2. Exposure (screening method) measured in standard, reliable way?** | **Q3. Valid methods for identification of exposure (screening method)?** | **Q4. Prospective: attempt to enroll all eligible? Retrospective: enroll more than just attenders to screening?** | **Q5a. Complete inclusion of eligible participants?** | **Q5b. Acceptable attrition rate for outcome being assessed?** | **Q6. Clear reporting of demographics?** | **Q7a. Outcome clearly defined and appropriate?** | **Q7b. Structured method of identifying outcomes?** | **Q7c. Follow-up duration adequate?** | **Overall ROB** |
|---|---|---|---|---|---|---|---|---|---|---|---|
| (1 round) | | | | | | | | | | | |
| MacRedmond 2006 **Any FPs** (1 round) | Y | Y | Y | Y | U | Y | U | Y | Y | Y | Some concerns |
| MacRedmond 2006 **Any FPs @ dx** (1 round) | Y | Y | Y | Y | U | U | U | Y | Y | Y | Some concerns |
| MacRedmond 2006 **Any IFs** (1 round) | Y | Y | Y | Y | U | Y | U | U | Y | Y | Some concerns |
| MacRedmond 2006 **Clinically significant IFs** (1 round) | Y | Y | Y | Y | U | Y | U | U | Y | Y | Some concerns |
| MacRedmond 2006 **Any FPs** (2/3 rounds) | Y | Y | Y | Y | U | Y | U | Y | Y | Y | Some concerns |
| MacRedmond 2006 **Any FPs @ dx** (2/3 rounds) | Y | Y | Y | Y | U | U | U | Y | Y | Y | Some concerns |
| Manyak 2023 **Any MCs** (2/3 rounds) | Y | Y | Y | N | Y | U | Y | U | Y | U | Some concerns |
| Manyak 2023 **Any deaths from Ips** (2/3 rounds) | Y | Y | Y | N | Y | U | Y | U | Y | U | Some concerns |
| Manyak 2023 | Y | Y | Y | N | Y | Y | Y | N | U | U | High |

| Author Year Outcome (Screening rounds or timepoints for psychosocial harms) | Q1. Clear inclusion criteria? | Q2. Exposure (screening method) measured in standard, reliable way? | Q3. Valid methods for identification of exposure (screening method)? | Q4. Prospective: attempt to enroll all eligible? Retrospective: enroll more than just attenders to screening? | Q5a. Complete inclusion of eligible participants? | Q5b. Acceptable attrition rate for outcome being assessed? | Q6. Clear reporting of demographics? | Q7a. Outcome clearly defined and appropriate? | Q7b. Structured method of identifying outcomes? | Q7c. Follow-up duration adequate? | Overall ROB |
|---|---|---|---|---|---|---|---|---|---|---|---|
| **Clinically significant Ifs** (2/3 rounds) | | | | | | | | | | | |
| Marshall 2015 **Any FPs** (1 round) | Y | Y | Y | Y | U | Y | Y | Y | Y | Y | Some concerns |
| Marshall 2015 **Any MCs** (1 round) | Y | Y | Y | Y | U | Y | Y | Y | Y | Y | Some concerns |
| Marshall 2015 **#FPs/person** (2/3 rounds) | Y | Y | Y | Y | U | U | Y | Y | Y | Y | Some concerns |
| Marshall 2015 **Any FPs @ dx** (2/3 rounds) | Y | Y | Y | Y | U | U | Y | Y | Y | Y | Some concerns |
| Marshall 2015 **Any MCs** (2/3 rounds) | Y | Y | Y | Y | U | U | Y | Y | Y | Y | Some concerns |
| Matthews 2024 **Clinically significant Ifs** (1 round) | Y | Y | Y | N | Y | Y | N | N | Y | U | High |
| Mazzei 2017 **Specific IFs** (1 round) | Y | Y | Y | N | U | Y | N | U | Y | U | Some concerns |
| Melzer 2024 **Any Ifs** (1 round) | Y | Y | Y | N | Y | Y | U | N | U | U | High |
| Melzer 2024 **Clinically significant Ifs** (1 round) | Y | Y | Y | N | Y | Y | U | N | U | U | High |
| Mendoza 2020 **Specific IFs** (1 round) | Y | Y | Y | N | Y | Y | N | N | Y | Y | High |

| Author Year Outcome (Screening rounds or timepoints for psychosocial harms) | Q1. Clear inclusion criteria? | Q2. Exposure (screening method) measured in standard, reliable way? | Q3. Valid methods for identification of exposure (screening method)? | Q4. Prospective: attempt to enroll all eligible? Retrospective: enroll more than just attenders to screening? | Q5a. Complete inclusion of eligible participants? | Q5b. Acceptable attrition rate for outcome being assessed? | Q6. Clear reporting of demographics? | Q7a. Outcome clearly defined and appropriate? | Q7b. Structured method of identifying outcomes? | Q7c. Follow-up duration adequate? | Overall ROB |
|---|---|---|---|---|---|---|---|---|---|---|---|
| Menezes 2010 **Any FPs** (1 round) | Y | Y | Y | U | U | Y | Y | Y | U | Y | Some concerns |
| Menezes 2010 **Any FPs @ dx** (1 round) | Y | Y | Y | U | U | Y | Y | Y | U | Y | Some concerns |
| Menezes 2010 **Any FPs & #Any FPs/person** (2/3 rounds) | Y | Y | Y | U | U | Y | Y | Y | U | Y | Some concerns |
| Menezes 2010 **Any FPs @ dx** (2/3 rounds) | Y | Y | Y | U | U | Y | Y | Y | U | Y | Some concerns |
| Miller 2016 **Any FPs** (4/5 rounds) | U | U | Y | N | Y | Y | U | Y | U | U | Some concerns |
| Miller 2016 **Any FPs @ dx** (4/5 rounds) | U | U | Y | N | Y | Y | U | Y | U | U | Some concerns |
| Morgan 2017 **Any IFs & #Any IFs/person** (1 round) | Y | Y | Y | N | U | Y | Y | U | Y | U | Some concerns |
| Morgan 2017 **Clinically significant IFs** (1 round) | Y | Y | Y | N | U | Y | Y | U | Y | U | Some concerns |
| Munoz-Largarcha 2018 **Any MCs** (1 round) | U | U | U | N | Y | Y | U | Y | U | U | Some concerns |
| Murray 2016 **#Any IFs/person** | Y | Y | Y | U | U | Y | Y | N | U | U | High |

| Author Year Outcome (Screening rounds or timepoints for psychosocial harms) | Q1. Clear inclusion criteria? | Q2. Exposure (screening method) measured in standard, reliable way? | Q3. Valid methods for identification of exposure (screening method)? | Q4. Prospective: attempt to enroll all eligible? Retrospective: enroll more than just attenders to screening? | Q5a. Complete inclusion of eligible participants? | Q5b. Acceptable attrition rate for outcome being assessed? | Q6. Clear reporting of demographics? | Q7a. Outcome clearly defined and appropriate? | Q7b. Structured method of identifying outcomes? | Q7c. Follow-up duration adequate? | Overall ROB |
|---|---|---|---|---|---|---|---|---|---|---|---|
| (1 round) | | | | | | | | | | | |
| Murray 2016 **Clinically significant IFs** (1 round) | Y | Y | Y | U | U | Y | Y | N | U | U | High |
| Murray 2016 **Any FPs** (1 round) | Y | Y | Y | U | U | Y | Y | Y | U | U | Some concerns |
| Murray 2016 **Any FPs @ dx** (1 round) | Y | Y | Y | U | U | Y | Y | Y | U | U | Some concerns |
| Murray 2016 **Any MCs & #Any MCs/person** (1 round) | Y | Y | Y | U | U | Y | Y | Y | U | U | Some concerns |
| Nahorecki 2015 **Any FPs** (1 round) | Y | Y | U | U | Y | N | N | Y | Y | Y | High |
| Nahorecki 2015 **Any FPs @ dx** (1 round) | Y | Y | U | U | Y | N | N | Y | Y | Y | High |
| Nunez 2022 **#Any MCs/person** (1 round) | Y | Y | Y | N | Y | U | Y | Y | Y | U | Some concerns |
| Nunez 2022 **Any deaths from IPs** (1 round) | Y | Y | Y | N | Y | U | Y | Y | Y | U | Some concerns |
| Ohmoto-Sekine 2016 **Specific Ifs** (1 round) | U | Y | Y | N | Y | Y | Y | U | U | U | Some concerns |

| Author Year Outcome (Screening rounds or timepoints for psychosocial harms) | Q1. Clear inclusion criteria? | Q2. Exposure (screening method) measured in standard, reliable way? | Q3. Valid methods for identification of exposure (screening method)? | Q4. Prospective: attempt to enroll all eligible? Retrospective: enroll more than just attenders to screening? | Q5a. Complete inclusion of eligible participants? | Q5b. Acceptable attrition rate for outcome being assessed? | Q6. Clear reporting of demographics? | Q7a. Outcome clearly defined and appropriate? | Q7b. Structured method of identifying outcomes? | Q7c. Follow-up duration adequate? | Overall ROB |
|---|---|---|---|---|---|---|---|---|---|---|---|
| Ostrowski 2019 **Any FPs** (1 round) | Y | Y | Y | N | Y | Y | U | Y | Y | Y | Some concerns |
| Ostrowski 2019 **Any FPs @ dx** (1 round) | Y | Y | Y | N | Y | Y | U | Y | Y | Y | Some concerns |
| Ostrowski 2019 **#Any IFs/person** (1 round) | Y | Y | Y | N | Y | Y | U | U | U | Y | Some concerns |
| Park 2024 **Specific Ifs** (1 round) | Y | Y | Y | N | Y | Y | Y | U | Y | U | Some concerns |
| Pastorino 2003 **Any FPs** (1 round) | Y | Y | Y | U | U | Y | U | Y | Y | Y | Some concerns |
| Pastorino 2003 **Any FPs @ dx** (1 round) | Y | Y | Y | U | U | Y | U | Y | Y | Y | Some concerns |
| Pastorino 2003 **#Any FPs/person** (round 2/3) | Y | Y | Y | U | U | Y | U | Y | Y | Y | Some concerns |
| Pastorino 2003 **#Any FPs @ dx/person** (round 2/3) | Y | Y | Y | U | U | Y | U | Y | Y | Y | Some concerns |
| Priola 2013 **Any IFs & #Any IFs/person** (1 round) | Y | Y | Y | U | U | Y | U | N | Y | Y | High |
| Priola 2013 **Clinically significant IFs &Clinically** | Y | Y | Y | U | U | Y | U | N | Y | Y | High |

| Author Year Outcome (Screening rounds or timepoints for psychosocial harms) | Q1. Clear inclusion criteria? | Q2. Exposure (screening method) measured in standard, reliable way? | Q3. Valid methods for identification of exposure (screening method)? | Q4. Prospective: attempt to enroll all eligible? Retrospective: enroll more than just attenders to screening? | Q5a. Complete inclusion of eligible participants? | Q5b. Acceptable attrition rate for outcome being assessed? | Q6. Clear reporting of demographics? | Q7a. Outcome clearly defined and appropriate? | Q7b. Structured method of identifying outcomes? | Q7c. Follow-up duration adequate? | Overall ROB |
|---|---|---|---|---|---|---|---|---|---|---|---|
| **significant IFs/person** (1 round) | | | | | | | | | | | |
| Priola 2013 **Any IFs & #Any IFs/person** (4/5 rounds) | Y | Y | Y | U | U | Y | U | N | Y | Y | High |
| Priola 2013 **Clinically significant IFs & #Clinically significant IFs/person** (4/5 rounds) | Y | Y | Y | U | U | Y | U | N | Y | Y | High |
| Pua 2024 [hospital site] **Clinically significant Ifs** (1 round) | U | Y | Y | Y | U | Y | U | U | U | U | Some concerns |
| Pua 2024 [mobile site] **Clinically significant Ifs** (1 round) | Y | Y | Y | Y | U | Y | U | N | U | U | High |
| Raghaven 2020 **Specific IFs** (1 round) | Y | Y | Y | U | U | U | U | U | Y | N | Some concerns |
| Rampinelli 2011 **#Clinically significant IFs/person** (1 round) | Y | Y | Y | N | Y | U | N | U | Y | Y | Some concerns |
| Rampinelli 2011 **Clinically significant IFs** | Y | Y | Y | N | Y | U | N | U | Y | Y | Some concerns |

| Author Year Outcome (Screening rounds or timepoints for psychosocial harms) | Q1. Clear inclusion criteria? | Q2. Exposure (screening method) measured in standard, reliable way? | Q3. Valid methods for identification of exposure (screening method)? | Q4. Prospective: attempt to enroll all eligible? Retrospective: enroll more than just attenders to screening? | Q5a. Complete inclusion of eligible participants? | Q5b. Acceptable attrition rate for outcome being assessed? | Q6. Clear reporting of demographics? | Q7a. Outcome clearly defined and appropriate? | Q7b. Structured method of identifying outcomes? | Q7c. Follow-up duration adequate? | Overall ROB |
|---|---|---|---|---|---|---|---|---|---|---|---|
| **& #Clinically significant IFs/person** (4/5 rounds) | | | | | | | | | | | |
| Rampinelli 2017 **#Any FPs/person** (4/5 rounds) | Y | Y | Y | N | Y | Y | U | U | Y | Y | Some concerns |
| Regan 2019 **#Any IFs/person** (1 round) | Y | Y | Y | U | Y | Y | Y | N | U | U | High |
| Regis 2022 **#Any IFs/person** (1 round) | U | Y | Y | N | Y | Y | U | N | U | Y | High |
| Regis 2022 **Clinically significant IFs** (1 round) | U | Y | Y | N | Y | Y | U | N | U | Y | High |
| Regis 2022 **#Any IFs/person** (4/5 rounds) | U | Y | Y | N | Y | Y | U | N | U | Y | High |
| Regis 2022 **Clinically significant IFs** (4/5 rounds) | U | Y | Y | N | Y | Y | U | N | U | Y | High |
| Reiter 2018 **Any IFs & #Any IFs/person** (1 round) | Y | Y | Y | N | Y | Y | Y | N | N | U | High |
| Reiter 2018 **Clinically significant IFs** | Y | Y | Y | N | Y | Y | Y | N | N | U | High |

| Author Year Outcome (Screening rounds or timepoints for psychosocial harms) | Q1. Clear inclusion criteria? | Q2. Exposure (screening method) measured in standard, reliable way? | Q3. Valid methods for identification of exposure (screening method)? | Q4. Prospective: attempt to enroll all eligible? Retrospective: enroll more than just attenders to screening? | Q5a. Complete inclusion of eligible participants? | Q5b. Acceptable attrition rate for outcome being assessed? | Q6. Clear reporting of demographics? | Q7a. Outcome clearly defined and appropriate? | Q7b. Structured method of identifying outcomes? | Q7c. Follow-up duration adequate? | Overall ROB |
|---|---|---|---|---|---|---|---|---|---|---|---|
| **& #Clinically significant IFs/person**<br>(1 round) | | | | | | | | | | | |
| Rendle 2024<br>**Any MCs**<br>(1 round) | Y | Y | Y | N | Y | U | Y | Y | Y | Y | Some concerns |
| Rode 2025<br>**Clinically significant Ifs**<br>(1 round) | Y | Y | Y | Y | U | Y | Y | N | U | Y | High |
| Rode 2025<br>**Any MCs**<br>(4/5 rounds) | Y | Y | Y | Y | U | U | Y | Y | Y | Y | Some concerns |
| Rzyman 2015<br>**Any FPs @ dx**<br>(1 round) | Y | Y | Y | Y | U | Y | U | Y | Y | Y | Some concerns |
| Rzyman 2015<br>**#Any IFs/person**<br>(2/3 rounds) | Y | Y | Y | Y | U | Y | U | U | Y | Y | Some concerns |
| Rzyman 2015<br>**Any FPs**<br>(2/3 rounds) | Y | Y | Y | Y | U | Y | U | Y | Y | Y | Some concerns |
| Rzyman 2015<br>**Any FPs @ dx**<br>(2/3 rounds) | Y | Y | Y | Y | U | Y | U | Y | Y | Y | Some concerns |
| Rzyman 2015<br>**Any MCs**<br>(2/3 rounds) | Y | Y | Y | Y | U | Y | U | Y | U | U | Some concerns |
| Salvatore 2016<br>**Specific IFs & #Specific IFs/person**<br>(1 round) | N | Y | U | N | Y | Y | U | N | Y | U | High |

| Author Year Outcome (Screening rounds or timepoints for psychosocial harms) | Q1. Clear inclusion criteria? | Q2. Exposure (screening method) measured in standard, reliable way? | Q3. Valid methods for identification of exposure (screening method)? | Q4. Prospective: attempt to enroll all eligible? Retrospective: enroll more than just attenders to screening? | Q5a. Complete inclusion of eligible participants? | Q5b. Acceptable attrition rate for outcome being assessed? | Q6. Clear reporting of demographics? | Q7a. Outcome clearly defined and appropriate? | Q7b. Structured method of identifying outcomes? | Q7c. Follow-up duration adequate? | Overall ROB |
|---|---|---|---|---|---|---|---|---|---|---|---|
| Seo 2023 **Specific IFs** (1 round) | Y | Y | Y | N | Y | Y | U | U | Y | U | Some concerns |
| Shah 2024 **Any IFs** (1 round) | Y | Y | Y | Y | U | Y | U | N | U | U | High |
| Shah 2024 **Clinically significant IFs** (1 round) | Y | Y | Y | Y | U | Y | U | N | U | U | High |
| Song 2018 **#Any IF/person** (1 round) | U | Y | Y | N | Y | Y | U | N | Y | U | High |
| Steiger 2021 **Specific IFs** (1 round) | Y | U | U | N | Y | Y | Y | N | Y | N | High |
| Taghizadeh 2019 **Any FPs** (1 round) | Y | Y | Y | U | U | Y | Y | Y | Y | Y | Some concerns |
| Taghizadeh 2019 **Any FPs @ dx** (1 round) | Y | Y | Y | U | U | Y | Y | Y | Y | Y | Some concerns |
| Taghizadeh 2019 **Psychosocial consequences** (Positive screen [before repeat CT or diagnostic results known vs. before screening, in screen +ve]); | Y | Y | Y | U | U | Y | Y | Y | Y | Y | Some concerns |

| Author Year Outcome (Screening rounds or timepoints for psychosocial harms) | Q1. Clear inclusion criteria? | Q2. Exposure (screening method) measured in standard, reliable way? | Q3. Valid methods for identification of exposure (screening method)? | Q4. Prospective: attempt to enroll all eligible? Retrospective: enroll more than just attenders to screening? | Q5a. Complete inclusion of eligible participants? | Q5b. Acceptable attrition rate for outcome being assessed? | Q6. Clear reporting of demographics? | Q7a. Outcome clearly defined and appropriate? | Q7b. Structured method of identifying outcomes? | Q7c. Follow-up duration adequate? | Overall ROB |
|---|---|---|---|---|---|---|---|---|---|---|---|
| (False positive screening result [>6 months after diagnostic results known vs. before screening, in FPs]) | | | | | | | | | | | |
| Taghizadeh 2019 **Psychosocial consequences** (False positive screening result [≤6 months after diagnostic results known, vs. before screening, in FPs]) | Y | Y | Y | U | U | N | Y | Y | Y | Y | High |
| Tisi 2023 **Specific Ifs** (1 round) | Y | Y | Y | Y | Y | Y | Y | N | U | U | High |
| Triplette 2024 **Clinically significant Ifs** (1 round) | Y | Y | Y | N | U | Y | U | N | U | U | High |
| van Gerwen 2024 **Specific Ifs** (1 round) | Y | Y | Y | N | Y | Y | Y | N | U | U | High |
| van Gerwen 2024 **Specific Ifs** (2/3 rounds) | Y | Y | Y | N | Y | Y | Y | N | N | U | High |

| **Author Year Outcome (Screening rounds or timepoints for psychosocial harms)** | **Q1. Clear inclusion criteria?** | **Q2. Exposure (screening method) measured in standard, reliable way?** | **Q3. Valid methods for identification of exposure (screening method)?** | **Q4. Prospective: attempt to enroll all eligible? Retrospective: enroll more than just attenders to screening?** | **Q5a. Complete inclusion of eligible participants?** | **Q5b. Acceptable attrition rate for outcome being assessed?** | **Q6. Clear reporting of demographics?** | **Q7a. Outcome clearly defined and appropriate?** | **Q7b. Structured method of identifying outcomes?** | **Q7c. Follow-up duration adequate?** | **Overall ROB** |
|---|---|---|---|---|---|---|---|---|---|---|---|
| Veronesi 2008 **Any FPs** (1 round) | Y | Y | Y | N | Y | Y | U | Y | Y | Y | Some concerns |
| Veronesi 2008 **Any FPs @ dx** (1 round) | Y | Y | Y | N | Y | Y | U | Y | Y | Y | Some concerns |
| Veronesi 2012 **Any FPs @ dx** (4/5 rounds) | Y | Y | Y | N | Y | Y | U | Y | Y | Y | Some concerns |
| Walker 2015 **Any MCs & #Any MCs/person** (1 round) | Y | Y | Y | N | Y | Y | N | Y | Y | U | Some concerns |
| Wichmann 2025 **Any FPs** (1 round) | Y | Y | Y | Y | U | Y | Y | Y | Y | Y | Some concerns |
| Wilson 2008 **Any FPs** (1 round) | Y | Y | Y | Y | U | Y | Y | Y | Y | Y | Some concerns |
| Wilson 2008 **Any FPs @ dx** (1 round) | Y | Y | Y | Y | U | Y | Y | Y | Y | Y | Some concerns |
| Wilson 2008 **#Clinically significant IFs/person** (1 round) | Y | Y | Y | Y | U | Y | Y | N | Y | Y | High |
| Wilson 2008 **Any FPs** (2/3 rounds) | Y | Y | Y | Y | U | Y | Y | Y | Y | Y | Some concerns |
| Wilson 2008 **Any FPs** (2/3 rounds) | Y | Y | Y | Y | U | Y | Y | Y | Y | Y | Some concerns |

| Author Year Outcome (Screening rounds or timepoints for psychosocial harms) | Q1. Clear inclusion criteria? | Q2. Exposure (screening method) measured in standard, reliable way? | Q3. Valid methods for identification of exposure (screening method)? | Q4. Prospective: attempt to enroll all eligible? Retrospective: enroll more than just attenders to screening? | Q5a. Complete inclusion of eligible participants? | Q5b. Acceptable attrition rate for outcome being assessed? | Q6. Clear reporting of demographics? | Q7a. Outcome clearly defined and appropriate? | Q7b. Structured method of identifying outcomes? | Q7c. Follow-up duration adequate? | Overall ROB |
|---|---|---|---|---|---|---|---|---|---|---|---|
| Wilson 2008 **Any FPs @ dx** (2/3 rounds) | Y | Y | Y | Y | U | Y | Y | Y | Y | Y | Some concerns |
| Wilson 2008 **#Clinically significant IFs/person** (2/3 rounds) | Y | Y | Y | Y | U | Y | Y | U | Y | Y | Some concerns |
| Zhao 2020 **#Any MCs/person** (1 round) | Y | Y | Y | N | U | N | N | Y | U | U | High |

Abbreviations: FPs = false positives; IFs= incidental findings; MC = major complication; N = No; U = Unclear; Y=Yes.

Notes on ROB:

- Overall ROB was rated as: low if all domains were rated as Y; some concerns if any domains were rated as U; high if at least one of the main domains was rated as N (5b, 7a, and 7b for FP, MC and psychosocial consequences; 7a and 7b for IF).
- Q5a was rated as: Y if ≥80% of eligible participants were enrolled.
- Q5b was rated as: Y if ≥90% of those who were recalled (for FP) or referred for additional diagnostic workup (for FP@ dx or for IF) attended the subsequent follow-up.
- Q6 was rated as: Y if age, sex, smoking, and at least one comorbidity were reported, U if three of the four variables were reported, and N if less than three of variables were reported.

Table S2.6. Risk of bias: Observational harms studies, cohort

| Author Year Outcome (Timepoints for psychosocial harms) | Q1. Groups similar and from same population? | Q2. Exposures measured similarly? | Q3. Exposure measured in valid and reliable way? | Q4. Confounding factors identified? | Q5. Strategies for confounding stated? | Q6. Participants free of outcome at moment of exposure? | Q7. Outcomes measured valid and reliable? | Q8. Follow-up time sufficient for outcomes to occur? | Q9. Follow-up complete? | Q10. Strategies to address incomplete follow-up used? | Q11. Appropriate statistical analysis? | Overall ROB |
|---|---|---|---|---|---|---|---|---|---|---|---|---|
| Andersson, 2021 **Psychosocial consequences** (False positive screening result [>6 months after diagnostic results known vs. control group]) | Y | Y | Y | Y | Y | Y | Y | U | U | Y | Y | Some concerns |
| LSUT (Kummer, 2020) (Undergoing CT scan [after screening, before results are known in pooled screening arms vs. community control]); (False positive screening result [≤6 months after diagnostic results known vs. community control]) | N | Y | Y | U | N | Y | Y | U | N | N | U | High |

Abbreviations: N = No; U = Unclear; Y=Yes.

Notes on ROB:

- Overall ROB was rated as: low if all domains were rated as Y; some concerns if any domains were rated as U; high if at least one of the main domains was rated as N (7, 8, 9, 11)
- Q4 was rated as: Y if age, sex, smoking, and at least one comorbidity were reported, U if three of the four variables were reported, and N if less than three of variables were reported.
- Q9 was rated as: Y if ≥90% of those who enrolled had complete follow-up.

Table S2.7 CT screening: Critical outcome sensitivity and subgroup analyses

| Comparison | N trials | n | RR (95% CI) | Mean control event rate | $I^2$ | Absolute difference (95% CI) per 1,000 |
|---|---|---|---|---|---|---|
| **Lung cancer mortality** | | | | | | |
| Longest follow-up vs longest follow-up <10 years | | | | | | |
| Longest follow-up | 9 (1, 9-11, 14, 15, 17, 18, 20) | 94,530 | 0.87 [0.79, 0.96] | 0.03053 | 14% | -3.97 [-6.41, -1.22] |
| Longest follow-up <10 years for NLST (6.5 yrs follow-up) | 9 (2, 9-11, 14, 15, 17, 18, 20) | 94,530 | 0.83 [0.76, 0.91] | 0.02769 | 0% | -4.71 [-6.65, -2.49] |
| Sensitivity analysis: risk of bias (Fig. S2.2) | | | | | | |
| All trials | 9 (1, 9-11, 14, 15, 17, 18, 20) | 94,530 | 0.87 [0.79, 0.96] | 0.03053 | 14% | -3.97 [-6.41, -1.22] |
| Some concerns of bias trials | 8 (1, 9, 10, 14, 15, 17, 18, 20) | 92,080 | 0.86 [0.77, 0.95] | 0.02855 | 21% | -4.00 [-6.57, -1.43] |
| Between study subgroup: comparator strategy (test for subgroup differences, p=0.07) (Fig. S2.3) | | | | | | |
| No screening | 7 (9-11, 14, 15, 17, 18) | 37,758 | 0.80 [0.71, 0.90] | 0.03041 | 0% | -6.08 [-8.82, -3.04] |
| CXR screening | 2 (1, 20) | 56,772 | 0.95 [0.82, 1.10] | 0.03096 | 11% | -1.55 [-5.57, 3.10] |
| Between study subgroup: number of screening rounds (test for subgroup differences, p=0.20) (Fig. S2.4) | | | | | | |
| 1 round | 1 (18) | 4,055 | 0.76 [0.51, 1.15] | 0.02516 | NA | -6.04 [-12.33, 3.77] |
| 2-3 rounds | 2 (1, 20) | 56,772 | 0.95 [0.82, 1.10] | 0.03096 | 11% | -1.55 [-5.57, 3.10] |
| 4+ rounds | 6 (9-11, 14, 15, 17) | 33,703 | 0.80 [0.70, 0.91] | 0.03129 | 0% | -6.26 [-9.39, -2.82] |
| Between study subgroup: screening interval (test for subgroup differences, p=0.08) (Fig. S2.5) | | | | | | |
| Annual | 7 (1, 10, 11, 14, 15, 17, 20) | 72,636 | 0.92 [0.86, 0.99] | 0.03117 | 0% | -2.49 [-4.36, -0.31] |
| >Annual | 2 (9, 17) | 17,840 | 0.75 [0.63, 0.90] | 0.02731 | 0% | -6.83 [-10.11, -2.73] |
| Only one round | 1 (18) | 4,055 | 0.76 [0.51, 1.15] | 0.02516 | NA | -6.04 [-12.33, 3.77] |
| Between study subgroup: ≤2% vs >2% 6-year lung cancer detection rate in comparator group (test for subgroup differences, p=0.004) (Fig. S2.6) | | | | | | |
| >2% | 8 (1, 9, 11, 14, 15, 17, 18, 20) | 90,426 | 0.86 [0.78, 0.95] | 0.03204 | 22% | -4.49 [-7.05, -1.60] |
| ≤2% | 1 (10) | 4,104 | 1.03 [0.66, 1.60] | 0.01852 | NA | 0.56 [-6.30, 11.11] |
| Between study subgroup: ≤3% vs >3% 6-year lung cancer detection rate in comparator group (test for subgroup differences, p=0.02) (Fig. S2.7) | | | | | | |
| >3% | 4 (1, 11, 14, 20) | 62,428 | 0.77 [0.67, 0.89] | 0.03869 | 0% | -8.90 [-12.77, -4.26] |
| ≤3% | 5 (9, 10, 15, 17, 18) | 32,102 | 0.93 [0.86, 1.00] | 0.02401 | 0% | -1.68 [-3.36, 0.00] |
| Between study subgroup: nodule management strategy (test for subgroup differences, p=0.07) (Fig. S2.8) | | | | | | |
| Most sensitive (3 to <5mm) | 2 (1, 20) | 56,772 | 0.95 [0.82, 1.10] | 0.03096 | 11% | -1.55 [-5.57, 3.10] |
| Less sensitive (>5mm) | 7 (9-11, 14, 15, 17, 18) | 37,758 | 0.80 [0.71, 0.90] | 0.03041 | 0% | -6.08 [-8.82, -3.04] |
| Between study subgroup: sex (test for subgroup differences, p=0.31) (Fig. S2.9) | | | | | | |
| Men | 5 (1, 9, 11, 16, 19) | 52,833 | 0.88 [0.76, 1.02] | 0.03385 | 37% | -4.06 [-8.12, 0.68] |

| Comparison | N trials | n | RR (95% CI) | Mean control event rate | $I^2$ | Absolute difference (95% CI) per 1,000 |
|---|---|---|---|---|---|---|
| Women | 4 (1, 9, 16, 19) | 26,965 | 0.75 [0.56, 1.00] | 0.02871 | 30% | -7.18 [-12.63, 0.00] |
| Within study subgroup: sex (test for subgroup differences, p=0.14) (Fig. S2.10) | | | | | | |
| Men vs. women | 4 (1, 9, 16, 19) | 77,348 | Ratio of RRs: 1.21 [0.93, 1.57] | NA | 45.4% | NA |
| Within study subgroup: age | | | | | | |
| <65 years | 1 (NLST) (1) | 39,234 | 0.87 [0.78, 0.96] | 0.03766 | NA | -4.90 [-8.29, -1.51] |
| ≥65 years | 1 (NLST) (1) | 14,218 | 1.02 [0.90, 1.15]<br><br>Test for subgroup differences: p=0.05 | 0.06992 | NA | 1.40 [-6.99, 10.49] |
| ≤60 years | 2 (DANTE+MILD pooled) (12) | 3,158 | Adjusted HR: 0.52 [0.23, 1.21] | NR | NA | NR |
| >60 years | 2 (DANTE+MILD pooled) (12) | 3,445 | Adjusted HR: 0.88 [0.64, 1.23]<br><br>Test for subgroup differences: p=0.24 | NR | NA | NR |
| Within study subgroup: age in men only (test for subgroup differences, p=0.81) | | | | | | |
| Men, 50-54 years | 1 (NELSON) (9) | 3,305 | 0.85 [0.50, 1.43] | 0.018300 | NA | -2.74 [-9.15, 7.87] |
| Men, 55-59 years | 1 (NELSON) (9) | 4,457 | 0.70 [0.46, 1.08] | 0.02241 | NA | -6.72 [-12.10, 1.79] |
| Men, 60-64 years | 1 (NELSON) (9) | 3,029 | 0.83 [0.57, 1.21] | 0.03797 | NA | -6.45 [-16.33, 7.97] |
| Men, 65-69 years | 1 (NELSON) (9) | 1,578 | 0.59 [0.37, 0.95] | 0.05506 | NA | -22.57 [-34.69, -2.75] |
| Men, 70-74 years | 1 (NELSON) (9) | 666 | 0.81 [0.45, 1.45] | 0.07122 | NA | -13.53 [-39.17, 32.05] |
| Within study subgroup: smoking status | | | | | | |
| Current smoker | 1 (NLST) (1) | 25,760 | 0.89 [0.81, 0.98] | 0.06341 | NA | -4.90 [-8.29, -1.51] |
| Former smoker | 1 (NLST) (1) | 27,692 | 1.01 [0.88, 1.15]<br><br>Test for subgroup differences: p=0.13 | 0.03022 | NA | 1.40 [-6.99, 10.49] |
| Current smoker | 2 (DANTE+MILD pooled) (12) | 4,508 | Adjusted HR: 0.78 [0.54, 1.11] | NR | NA | NR |
| Former smoker | 2 (DANTE+MILD pooled) (12) | 2,041 | Adjusted HR: 1.32 [0.63, 2.75] | NR | NA | NR |

| Comparison | N trials | n | RR (95% CI) | Mean control event rate | $I^2$ | Absolute difference (95% CI) per 1,000 |
|---|---|---|---|---|---|---|
| | | | Test for subgroup differences: p=0.21 | | | |
| Within study subgroup: COPD status | | | | | | |
| COPD | 1 (DLCST) (10) | 1,750 | 0.85 [0.49, 1.47] | 0.03037 | NA | -4.56 [-15.49, 14.28] |
| No COPD | 1 (DLCST) (10) | 2,353 | 1.38 [0.65, 2.90]<br><br>Test for subgroup differences: p=0.30 | 0.01003 | NA | 3.81 [-3.51, 19.06] |
| COPD | 1 (NLST) (8) | 6,160 | Mortality reduction ratio: 0.83 [0.63, 1.09] | NR | NA | NR |
| No COPD | 1 (NLST) (8) | 12,303 | Mortality reduction ratio: 0.74 [0.55, 1.00]<br><br>Test for subgroup differences: p=0.52 | NR | NA | NR |
| Within study subgroup: pack-years smoking history | | | | | | |
| ≥35 pack-years smoking history | 1 (DLCST) (10) | 1,955 | 0.92 [0.54, 1.54] | 0.02941 | NA | -2.35 [-13.53, 15.88] |
| <35 pack-years smoking history | 1 (DLCST) (10) | 2,148 | 1.26 [0.55, 2.90]<br><br>Test for subgroup differences: p=0.52 | 0.00909 | NA | 2.36 [-4.09, 17.27] |
| ≥40 pack-years smoking history | 2 (DANTE+MILD pooled) (12) | 3,743 | Adjusted HR: 0.94 [0.66, 1.33] | NR | NA | NR |
| <40 pack-years smoking history | 2 (DANTE+MILD pooled) (12) | 2,806 | Adjusted HR: 0.53 [0.27, 1.04]<br><br>Test for subgroup differences: p=0.14 | NR | NA | NR |
| Within study subgroup: COPD and pack-years smoking history (test for subgroup differences: p=0.76) | | | | | | |
| COPD and ≥35 pack-years smoking history | 1 (DLCST) (10) | 803 | 0.81 [0.43, 1.52] | 0.04338 | NA | -8.24 [-24.73, 22.56] |
| No COPD and ≥35 pack-years smoking history | 1 (DLCST) (10) | 1,345 | 1.19 [0.47, 2.98] | 0.01629 | NA | 3.10 [-8.64, 32.26] |
| COPD and <35 pack-years smoking history | 1 (DLCST) (10) | 947 | 0.97 [0.31, 2.98] | 0.01519 | NA | -0.46 [-10.48, 30.08] |
| No COPD and <35 pack-years smoking history | 1 (DLCST) (10) | 1,008 | 1.65 [0.47, 5.83] | 0.00567 | NA | 3.69 [-3.01, 27.40] |
| Within study subgroup: Race (test for subgroup differences: p=0.29) | | | | | | |

| Comparison | N trials | n | RR (95% CI) | Mean control event rate | $I^2$ | Absolute difference (95% CI) per 1,000 |
|---|---|---|---|---|---|---|
| White | 1 (NLST) (7) | 47,902 | Adjusted HR: 0.86 [0.75, 0.99] | NR | NA | NR |
| Black | 1 (NLST) (7) | 2,361 | Adjusted HR: 0.61 [0.37, 1.01] | NR | NA | NR |
| Other/missing race | 1 (NLST) (7) | 3,189 | Adjusted HR: 0.72 [0.53, 0.98] | NR | NA | NR |
| Within study subgroup: Index of multiple deprivation (IMD) quintile (test for subgroup differences: p=0.81) | | | | | | |
| Quintile 1-2 | 1 (UKLS) (18) | 1,540 | 0.82 [0.48, 1.39] | 0.03846 | NA | -6.92 [-20.00, 15.00] |
| Quintile 3-5 | 1 (UKLS) (18) | 2,430 | 0.74 [0.38, 1.43] | 0.01699 | NA | -4.42 [-10.53, 7.31] |
| Within study subgroup: Co-existing pulmonary conditions | | | | | | |
| ≥2 vs. 1 vs. no co-existing pulmonary conditions | 1 (NLST) (3) | 53,158 | NR | NR | NA | CT-prevented lung cancer deaths per 10,000-person years (95% CI):<br><br>6.2 (1.7 to 10.7) with no pulmonary condition;<br><br>9.6 (1.5 to 17.7) with 1 condition;<br><br>-0.5 (-15.4 to 14.3) with ≥2 conditions |
| Within study subgroup: NLST-eligibility (test for subgroup differences: p=0.82) | | | | | | |
| NLST-eligible | 2 (DANTE+MILD pooled) (12) | 2,338 | Adjusted HR: 0.82 [0.59, 1.14] | NR | NA | NR |
| Not NLST-eligible | 2 (DANTE+MILD pooled) (12) | 4,211 | Adjusted HR: 0.91 [0.40, 2.06] | NR | NA | NR |
| Within study subgroup: sex in those NLST-eligible only (test for subgroup differences: p=0.25) | | | | | | |
| NLST-eligible men | 1 (NELSON) (9) | NR | Cumulative rate ratio: 0.82 [0.64, 1.05] | NR | NA | NR |
| NLST-eligible women | 1 (NELSON) (9) | NR | Cumulative rate ratio: 0.61 [0.30, 1.21] | NR | NA | NR |
| Within study subgroup: smoking status in those NLST-eligible only (test for subgroup differences: p=0.07) | | | | | | |
| NLST-eligible and current smoker | 2 (DANTE+MILD pooled) (12) | 2,579 | Adjusted HR: 0.71 [0.48, 1.06] | NR | NA | NR |
| NLST-eligible and former smoker | 2 (DANTE+MILD pooled) (12) | 1,445 | Adjusted HR: 1.59 [0.74, 3.39] | NR | NA | NR |
| Within study subgroup: 5-year lung cancer death risk quintile (test for trend, p=0.80) | | | | | | |

| Comparison | N trials | n | RR (95% CI) | Mean control event rate | $I^2$ | Absolute difference (95% CI) per 1,000 |
|---|---|---|---|---|---|---|
| Risk quintile 1 to 5<br><br>5-year risk of lung-cancer death: 0.15 to 0.55% in quintile 1 to more than 2.00% in quintile 5<br><br>N~10,000 per quintile | 1 (NLST) (3) | 53,158 | Lung cancer mortality ratio (ratio of expected to observed lung cancer deaths): 0.97 in quintile 1, 0.78 in quintile 2, 0.75 in quin tile 3, 0.70 in quintile 4, and 0.84 in quintile 5; p=0.80 for trend<br><br>Overall rate of death from lung cancer: 24.6 per 10,000 py vs. 30.9 per 10,000py<br><br>77 of 88 CT-prevented lung-cancer deaths (88%) occurred among the 60% of participants with a 5-year risk of lung-cancer death of 0.85% or more (i.e., in quintiles 3 through 5), whereas only 1% of prevented lung-cancer deaths occurred among the 20% of participants at lowest risk (i.e., in quintile 1).<br><br>Relative reduction of 20.4% in the CT group (RR: 0.80; 95% CI: 0.69 to 0.92; p=0.001) and 6.3 fewer lung-cancer deaths per 10,000 person-years (95% CI: 2.4 to 10.1; p=0.001)<br><br>Relative reduction of 20.4% in the CT group (RR: 0.80; 95% CI: 0.69 to 0.92; p=0.001) and 6.3 fewer lung-cancer deaths per 10,000 person-years (95% CI: 2.4 to 10.1; p=0.001)<br>NNS: 5276 in quintile 1, 531 in quintile 2, 415 in quintile 3, 171 in quintile 4, and 161 in quintile 5; p<0.001 for trend | NR | NA | Per 10,000 py:<br>0.2 in quintile 1, 3.5 in quintile 2, 5.1 in quintile 3, 11.0 in quintile 4, and 12.0 in quintile 5; p=0.01 for trend) |
| Within study subgroup: PLCO 6-year cancer risk (test for interaction, p=0.61) | | | | | | |

| Comparison | N trials | n | RR (95% CI) | Mean control event rate | $I^2$ | Absolute difference (95% CI) per 1,000 |
|---|---|---|---|---|---|---|
| Quartiles 1 to 4<br><br>Quartiles based on $PLCO_M2012$ **6-year risk for cancer** (roughly equally sized groups; risks not reported) | 1 (NLST) (6) | 53,202 | Hazard ratios (by increasing risk):<br><br>Quintile 1: 0.86 (95% CI, 0.50 to 1.48);<br>Quintile 2: 0.71 (95% CI, 0.49 to 1.04);<br>Quintile 3: 0.70 (95% CI, 0.53 to 0.91);<br>Quntile 4: 0.88 (95% CI, 0.73 to 1.06); p-value for interaction=0.61<br><br>At all four levels of risk, the screening effect was protective. Random variation may explain differences in hazard ratios according to risk quartiles. | NR | NA | NR |
| Within study subgroup: Sex, and PLCO 6-year cancer risk (test for interaction, p=NR) | | | | | | |
| Women vs men, quintiles 1 to 5<br><br>Quintiles based on $PLCO_M2012$ **6-year risk for cancer** | 1 (NLST) (4) | 53,452 | Odds ratios (by increasing risk, and men as reference group), p-value:<br>Quintile 1: 1.63 (95% CI 0.79 to 2.49), p=0.15<br>Quintile 2: 0.78 (95% CI 0.59 to 1.37), p=0.46<br>Quintile 3: 1.38 (95% CI 0.93 to 1.82), p=0.11<br>Quintile 4: 1.09 (95% CI 0.73 to 1.47), p=0.62<br>Quintile 5: 1.45 (95% CI 1.17 to 1.72), p=0.002<br><br>OR (95% CI) data estimated from a figure | NR | NA | NR |
| Within study subgroup: PLCO lung cancer risk percentile (test for subgroup difference, p=NR) | | | | | | |
| Risk percentile: 0 to 10 | 1 (NLST) (5) | 1 | NA (zero in denominator) | NR | NA | NR |
| Risk percentile: >10 to 20 | 1 (NLST) (5) | 8 | NA (zero in denominator) | NR | NA | NR |
| Risk percentile: >20 to 30 | 1 (NLST) (5) | 154 | NA (zero in denominator) | NR | NA | NR |
| Risk percentile: >30 to 40 | 1 (NLST) (5) | 997 | 1.76 [0.09, 103.71] | NR | NA | 2.48 [-7.74, 12.68] per 10,000 py |

| Comparison | N trials | n | RR (95% CI) | Mean control event rate | $I^2$ | Absolute difference (95% CI) per 1,000 |
|---|---|---|---|---|---|---|
| Risk percentile: >40 to 50 | 1 (NLST) (5) | 3,094 | 0.73 [0.18, 2.68] | NR | NA | -1.82 [-8.50, 4.87] per 10,000 py |
| Risk percentile: >50 to 60 | 1 (NLST) (5) | 5,940 | 0.62 [0.28, 1.36] | NR | NA | -3.68 [-9.31, 1.95] per 10,000 py |
| Risk percentile: >60 to 70 | 1 (NLST) (5) | 8,581 | 1.01 [0.61, 1.66] | NR | NA | 0.14 [-5.74, 6.01] per 10,000 py |
| Risk percentile: >70 to 80 | 1 (NLST) (5) | 10,819 | 0.74 [0.52, 1.04] | NR | NA | -6.06 [-12.74, 0.62] per 10,000 py |
| Risk percentile: >80 to 90 | 1 (NLST) (5) | 10,997 | 0.76 [0.58, 0.99] | NR | NA | -9.38 [-18.07, -0.70] per 10,000 py |
| Risk percentile: >90 to 100 | 1 (NLST) (5) | 11,374 | 0.92 [0.76, 1.10] | NR | NA | -5.94 [-18.17, 6.30] per 10,000 py |
| At **PLCOm2012 risk ≥0.0151 (1.5%), the 65th percentile of risk, the NLST CT arm mortality rates are consistently below the CXR arm's rates**. The NNS to prevent one lung cancer death in the 65th to 100th percentile risk group is 255 (95% CI 143 to 1,184; ARD 6.43 per 10,000 person-years), and in the 30th to <65th percentile risk group is 963 (95% CI 291 to 2754; ARD 1.60 fewer per 10,000 person-years); **the NNS could not be estimated in the <30th percentile risk group because of absence of lung cancer deaths (5)**. | | | | | | |
| **All-cause mortality** | | | | | | |
| Sensitivity analysis: risk of bias (Fig. S2.11) | | | | | | |
| All trials | 9 (1, 9-11, 13, 15, 17, 18, 20) | 94,530 | 0.97 [0.93, 1.01] | 0.1218 | 15% | -3.66 [-8.53, 1.22] |
| Some concerns of bias trials | 8 (1, 9, 10, 13, 15, 17, 18, 20) | 92,080 | 0.97 [0.92, 1.02] | 0.1185 | 25% | -3.56 [-9.48, 2.37] |
| Between study subgroup: comparator strategy (test for subgroup differences, p=0.34) (Fig S2.12) | | | | | | |
| X-ray screening | 2 (1, 20) | 56,722 | 1.04 [0.87, 1.26] | 0.13535 | 63% | 5.41 [-17.60, 35.19] |
| No screening | 7 (9-11, 13, 15, 17, 18) | 37,758 | 0.95 [0.90, 1.01] | 0.11798 | 0% | -5.90 [-11.80, 1.18] |
| Between study subgroup: number of screening rounds (test for subgroup differences, p=0.40) (Fig. S2.13) | | | | | | |
| 1 round | 1 (18) | 4,055 | 0.89 [0.78, 1.03] | 0.1727 | NA | -18.99 [-37.99, 5.18] |
| 2-3 rounds | 2 (1, 20) | 56,772 | 1.04 [0.87, 1.26] | 0.13535 | 63% | 5.41 [-17.60, 35.19] |
| 4+ rounds | 6 (9-11, 13, 15, 17) | 33,703 | 0.96 [0.91, 1.02] | 0.10887 | 0% | -4.35 [-9.80, 2.18] |
| Between study subgroup: screening interval (test for subgroup differences, p=0.49) (Fig. S2.14) | | | | | | |
| Annual | 7 (1, 10, 11, 13, 15, 17, 20) | 72,636 | 0.98 [0.92, 1.05] | 0.11435 | 23% | -2.29 [-9.15, 5.72] |
| >Annual | 2 (9, 17) | 17,840 | 0.98 [0.90, 1.06] | 0.09245 | 0% | -1.85 [-9.25, 5.55] |
| Only one round | 1 (18) | 4,055 | 0.89 [0.78, 1.03] | 0.1727 | NA | -18.99 [-37.99, 5.18] |

| Comparison | N trials | n | RR (95% CI) | Mean control event rate | $I^2$ | Absolute difference (95% CI) per 1,000 |
|---|---|---|---|---|---|---|
| Between study subgroup: ≤2% vs >2% 6-year lung cancer detection rate in comparator group (test for subgroup differences, p=0.68) (Fig. S2.15) | | | | | | |
| ≤2% | 1 (10) | 4,104 | 1.01 [0.82, 1.25] | 0.07943 | NA | 0.79 [-14.30, 19.86] |
| >2% | 8 (1, 9, 11, 13, 15, 17, 18, 20) | 90,426 | 0.97 [0.92, 1.02] | 0.1271 | 24% | -3.81 [-10.17, 2.54] |
| Between study subgroup: ≤3% vs >3% 6-year lung cancer detection rate in comparator group (test for subgroup differences, p=0.94) (Fig. S2.16) | | | | | | |
| ≤3% | 5 (9, 10, 15, 17, 18) | 32,102 | 0.97 [0.91, 1.03] | 0.1046 | 0% | -3.14 [-9.41, 3.14] |
| >3% | 4 (1, 11, 13, 20) | 62,428 | 0.97 [0.86, 1.08] | 0.1434 | 58% | -4.30 [-20.07, 11.47] |
| Between study subgroup: nodule management strategy (test for subgroup differences, p=0.34) (Fig. S2.17) | | | | | | |
| Most sensitive (3 to <5mm) | 2 (1, 20) | 56,722 | 1.04 [0.87, 1.26] | 0.13535 | 63% | 5.41 [-17.60, 35.19] |
| Less sensitive (>5mm) | 7 (9-11, 13, 15, 17, 18) | 37,758 | 0.95 [0.90, 1.01] | 0.11798 | 0% | -5.90 [-11.80, 1.18] |
| Between study subgroup: sex (test for subgroup differences, p=0.19) (Fig. S2.18) | | | | | | |
| Men | 4 (2, 9, 11, 16) | 49,797 | 0.97 [0.92, 1.02] | 0.11567 | 0% | -3.47 [-9.25, 2.31] |
| Women | 3 (2, 9, 16) | 25,946 | 0.90 [0.81, 1.00] | 0.06114 | 0% | -6.11 [-11.62, 0.00] |
| Within study subgroup: sex (test for subgroup differences, p=0.03) (Fig. S2.19) | | | | | | |
| Men vs. women | 3 (2, 9, 16) | 73,293 | Ratio of RRs: 1.15 [0.94, 1.42] | NA | 72% | NA |
| Within study subgroup: age | | | | | | |
| <65 years | 1 (NLST) (2) | 39,234 | 0.95 [0.87, 1.03] | 0.05693 | NA | -2.85 [-7.40, 1.71] |
| ≥65 years | 1 (NLST) (2) | 14,218 | 0.92 [0.85, 1.01]<br><br>Test for subgroup differences: p=0.68 | 0.12971 | NA | -10.38 [-19.46, 1.30] |
| ≤60 years | 2 (DANTE+MILD pooled) (12) | 3,158 | Adjusted HR: 0.86 [0.55, 1.33] | NR | NA | NR |
| >60 years | 2 (DANTE+MILD pooled) (12) | 3,445 | Adjusted HR: 0.90 [0.74, 1.08]<br><br>Test for subgroup differences: p=0.86 | NR | NA | NR |
| Within study subgroup: smoking status | | | | | | |
| Current smoker | 1 (NLST) (2) | 25,760 | 0.95 [0.88, 1.03] | 0.09349 | NA | -4.67 [-11.22, 2.80] |

| Comparison | N trials | n | RR (95% CI) | Mean control event rate | $I^2$ | Absolute difference (95% CI) per 1,000 |
|---|---|---|---|---|---|---|
| Former smoker | 1 (NLST) (2) | 27,692 | 0.92 [0.83, 1.01]<br><br>Test for subgroup differences: p=0.54 | 0.06023 | NA | -4.82 [-10.24, 0.60] |
| Current smoker | 2 (DANTE+MILD pooled) (12) | 4,508 | Adjusted HR: 0.87 [0.70, 1.07] | NR | NA | NR |
| Former smoker | 2 (DANTE+MILD pooled) (12) | 2,041 | Adjusted HR: 1.21 [0.84, 1.74]<br><br>Test for subgroup differences: p=0.13 | NR | NA | NR |
| Within study subgroup: pack-years smoking history (test for subgroup differences: p=0.35) | | | | | | |
| <40 pack-years | 2 (DANTE+MILD pooled) (12) | 2,806 | Adjusted HR: 1.03 [0.73, 1.45] | NR | NA | NR |
| ≥40 pack-years | 2 (DANTE+MILD pooled) (12) | 3,743 | Adjusted HR: 0.85 [0.69, 1.05] | NR | NA | NR |
| Within study subgroup: race (test for subgroup differences: p=0.13) | | | | | | |
| White | 1 (NLST) (7) | 47,902 | Adjusted HR: 0.95 [0.89, 1.02] | NR | NA | NR |
| Black | 1 (NLST) (7) | 2,361 | Adjusted HR: 0.81 [0.65, 1.00] | NR | NA | NR |
| Other/missing race | 1 (NLST) (7) | 3,189 | Adjusted HR: 0.78 [0.62, 0.99] | NR | NA | NR |
| Within study subgroup: Index of multiple deprivation (IMD) quintile (test for subgroup differences: p=0.36) | | | | | | |
| Quintile 1-2 | 1 (UKLS) (18) | 1,540 | 0.86 [0.69, 1.06] | 0.20128 | NA | -28.18 [-62.40, 12.08] |
| Quintile 3-5 | 1 (UKLS) (18) | 2,430 | 0.98 [0.81, 1.18] | 0.15615 | NA | -3.12 [-29.67, 28.11] |
| Within study subgroup: NLST-eligibility (test for subgroup differences: p=0.04) | | | | | | |
| NLST-eligible | 2 (DANTE+MILD pooled) (12) | 4,211 | Adjusted HR: 0.80 [0.66, 0.97] | NR | NA | NR |
| Not NLST-eligible | 2 (DANTE+MILD pooled) (12) | 2,338 | Adjusted HR: 1.31 [0.85, 2.02] | NR | NA | NR |
| Within study subgroup: smoking status in those NLST-eligible (test for subgroup differences: p=0.01) | | | | | | |
| Current smokers in those NLST-eligible | 2 (DANTE+MILD pooled) (12) | 2,579 | Adjusted HR: 0.73 [0.57, 0.93] | NR | NA | NR |
| Ex-smokers in those NLST-eligible | 2 (DANTE+MILD pooled) (12) | 1,445 | Adjusted HR: 1.27 [0.88, 1.84] | NR | NA | NR |
| **Overdiagnosis** | | | | | | |
| Sensitivity analysis: risk of bias and comparator (Fig. S2.20) | | | | | | |

| Comparison | N trials | n | RR (95% CI) | Mean control event rate | $I^2$ | Absolute difference (95% CI) per 1,000 |
|---|---|---|---|---|---|---|
| All trials | 7 (9-11, 14, 15, 17, 18) | 35,161 | 1.19 [1.03, 1.37] | 0.044160 | 50% | 8.39 [1.32, 16.34] |
| Some concerns of bias trials with no X-ray at baseline | 5 (9, 10, 14, 15, 18) | 28,612 | 1.17 [0.97, 1.42] | 0.042717 | 63% | 7.26 [-1.28, 17.94] |
| Sensitivity analysis: X-ray comparator at each screening round | | | | | | |
| All trials | 7 (9-11, 14, 15, 17, 18) | 35,161 | 1.19 [1.03, 1.37] | 0.044160 | 50% | 8.39 [1.32, 16.34] |
| All trials + NLST with X-ray comparator | 8 (7, 9-11, 14, 15, 17, 18) | 88,615 | 1.15 [1,02, 1.30] | 0.046503 | 62% | 6.98 [0.93, 13.95] |
| Between study subgroup: number of screening rounds (test for subgroup differences, p=0.41) (Fig. S2.21) | | | | | | |
| 1 round | 1 (18) | 4,055 | 1.06 [0.79, 1.41] | 0.04243 | NA | 2.55 [-8.91, 17.40] |
| 4+ rounds | 6 (9-11, 14, 15, 17) | 31,106 | 1.22 [1.03, 1.43] | 0.04444 | 56% | 9.78 [1.33, 19.11] |
| Between study subgroup: screening interval (test for subgroup differences, p=0.67) (Fig. S2.22) | | | | | | |
| Annual | 4 (10, 11, 14, 15) | 13,812 | 1.27 [0.96, 1.68] | 0.04647 | 72% | 12.55 [-1.86, 31.60] |
| >Annual | 2 (9, 17) | 17,294 | 1.15 [1.00, 1.31] | 0.04040 | 0% | 6.06 [0.00, 12.52] |
| Only one round | 1 (18) | 4,055 | 1.06 [0.79, 1.41] | 0.04243 | NA | 2.55 [-8.91, 17.40] |
| Between study subgroup: ≤2% vs >2% 6-year lung cancer detection rate in comparator group (test for subgroup differences, p=0.007) (Fig. S2.23) | | | | | | |
| ≤2% | 1 (10) | 4,104 | 1.81 [1.30, 2.52] | 0.02583 | NA | 20.92 [7.75, 39.26] |
| >2% | 6 (9, 11, 14, 15, 17, 18) | 31,057 | 1.13 [1.02, 1.25] | 0.04817 | 0% | 6.26 [0.96, 12.04] |
| Between study subgroup: ≤3% vs >3% 6-year lung cancer detection rate in comparator group (test for subgroup differences, p=0.62) (Fig. S2.24) | | | | | | |
| ≤3% | 5 (9, 10, 15, 17, 18) | 29,505 | 1.23 [1.05, 1.44] | 0.03995 | 45% | 9.19 [2.00, 17.58] |
| >3% | 2 (11, 14) | 5,656 | 1.10 [0.74, 1.65] | 0.06174 | 75% | 6.17 [-16.05, 40.13] |
| Between study subgroup: sex (test for subgroup differences, p=0.80) (Fig. S2.25) | | | | | | |
| Men | 4 (9, 11, 15, 19) | 21,305 | 1.17 [1.04, 1.32] | 0.04568 | 0% | 7.76 [1.83, 14.62] |
| Women | 2 (15, 19) | 2,449 | 1.24 [0.83, 1.85] | 0.03407 | 0% | 8.18 [-5.79, 28.96] |
| Within study subgroup: sex (test for subgroup differences, p=0.28) (Fig. S2.26) | | | | | | |

| Comparison | N trials | n | RR (95% CI) | Mean control event rate | $I^2$ | Absolute difference (95% CI) per 1,000 |
|---|---|---|---|---|---|---|
| Men vs. women | 2 (15, 19) | 8,107 | Ratio of RRs: 0.93 [0.65, 1.33] | NA | 14% | NA |
| Within study subgroup: Index of multiple deprivation (IMD) quintile (test for subgroup differences: p=0.64) | | | | | | |
| Quintile 1-2 | 1 (UKLS) (18) | 1,540 | 1.03 [0.70, 1.50] | 0.06410 | NA | 1.92 [-19.23, 32.05] |
| Quintile 3-5 | 1 (UKLS) (18) | 2,430 | 1.18 [0.76, 1.83] | 0.02913 | NA | 5.24 [-6.99, 24.17] |

| Study or Subgroup | LDCT Events | LDCT Total | Control Events | Control Total | Weight | Risk Ratio M-H, Random, 95% CI |
|---|---|---|---|---|---|---|
| **4.1.1 Some concerns** | | | | | | |
| DLCST (Wille 2016, median 9.8 y) | 39 | 2052 | 38 | 2052 | 5.2% | 1.03 [0.66, 1.60] |
| ITALUNG (Paci 2021, median 11.3 y) | 58 | 1613 | 74 | 1593 | 8.4% | 0.77 [0.55, 1.08] |
| LSS (Doroudi 2018, median 5.2 y) | 32 | 1660 | 26 | 1658 | 3.9% | 1.23 [0.74, 2.05] |
| LUSI (Maldonado 2021, median 9.77 y) | 33 | 2029 | 44 | 2023 | 5.1% | 0.75 [0.48, 1.17] |
| MILD (Pastorino 2019, 9 years follow-up) | 40 | 2376 | 40 | 1723 | 5.4% | 0.73 [0.47, 1.12] |
| NELSON (de Koning 2020, 10y or to censoring) | 186 | 7900 | 248 | 7892 | 20.8% | 0.75 [0.62, 0.90] |
| NLST (Pinsky 2019, median 12.3 y) | 1147 | 26722 | 1236 | 26732 | 45.3% | 0.93 [0.86, 1.00] |
| UKLS (Davies 2024, median 9.9 y) | 39 | 2028 | 51 | 2027 | 5.9% | 0.76 [0.51, 1.15] |
| **Subtotal (95% CI)** | | **46380** | | **45700** | **100.0%** | **0.86 [0.77, 0.95]** |
| Total events | 1574 | | 1757 | | | |
| Heterogeneity: Tau² = 0.00; Chi² = 8.89, df = 7 (P = 0.26); I² = 21% | | | | | | |
| Test for overall effect: Z = 2.86 (P = 0.004) | | | | | | |
| **Total (95% CI)** | | **46380** | | **45700** | **100.0%** | **0.86 [0.77, 0.95]** |
| Total events | 1574 | | 1757 | | | |
| Heterogeneity: Tau² = 0.00; Chi² = 8.89, df = 7 (P = 0.26); I² = 21% | | | | | | |
| Test for overall effect: Z = 2.86 (P = 0.004) | | | | | | |
| Test for subgroup differences: Not applicable | | | | | | |

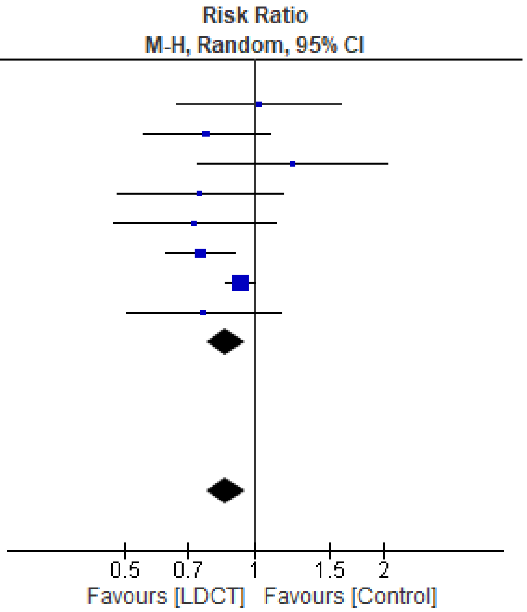

Figure S2.2 Lung cancer mortality, sensitivity analysis excluding high ROB studies (DANTE trial)

| Study or Subgroup | LDCT Events | LDCT Total | Control Events | Control Total | Weight | Risk Ratio M-H, Random, 95% CI |
|---|---|---|---|---|---|---|
| **4.2.1 No screening comparator** | | | | | | |
| DANTE (Infante 2015, median 8.35 y) | 59 | 1264 | 55 | 1186 | 6.1% | 1.01 [0.70, 1.44] |
| DLCST (Wille 2016, median 9.8 y) | 39 | 2052 | 38 | 2052 | 4.1% | 1.03 [0.66, 1.60] |
| ITALUNG (Paci 2021, median 11.3 y) | 58 | 1613 | 74 | 1593 | 6.9% | 0.77 [0.55, 1.08] |
| LUSI (Maldonado 2021, median 9.77 y) | 33 | 2029 | 44 | 2023 | 4.1% | 0.75 [0.48, 1.17] |
| MILD (Pastorino 2019, 9 years follow-up) | 40 | 2376 | 40 | 1723 | 4.3% | 0.73 [0.47, 1.12] |
| NELSON (de Koning 2020, 10y or to censoring) | 186 | 7900 | 248 | 7892 | 18.4% | 0.75 [0.62, 0.90] |
| UKLS (Davies 2024, median 9.9 y) | 39 | 2028 | 51 | 2027 | 4.7% | 0.76 [0.51, 1.15] |
| **Subtotal (95% CI)** | | **19262** | | **18496** | **48.6%** | **0.80 [0.71, 0.90]** |
| Total events | 454 | | 550 | | | |
| Heterogeneity: Tau² = 0.00; Chi² = 3.63, df = 6 (P = 0.73); I² = 0% | | | | | | |
| Test for overall effect: Z = 3.62 (P = 0.0003) | | | | | | |
| **4.2.2 X-ray screening comparator** | | | | | | |
| LSS (Doroudi 2018, median 5.2 y) | 32 | 1660 | 26 | 1658 | 3.1% | 1.23 [0.74, 2.05] |
| NLST (Pinsky 2019, median 12.3 y) | 1147 | 26722 | 1236 | 26732 | 48.3% | 0.93 [0.86, 1.00] |
| **Subtotal (95% CI)** | | **28382** | | **28390** | **51.4%** | **0.95 [0.82, 1.10]** |
| Total events | 1179 | | 1262 | | | |
| Heterogeneity: Tau² = 0.00; Chi² = 1.12, df = 1 (P = 0.29); I² = 11% | | | | | | |
| Test for overall effect: Z = 0.71 (P = 0.47) | | | | | | |
| **Total (95% CI)** | | **47644** | | **46886** | **100.0%** | **0.87 [0.79, 0.96]** |
| Total events | 1633 | | 1812 | | | |
| Heterogeneity: Tau² = 0.00; Chi² = 9.33, df = 8 (P = 0.32); I² = 14% | | | | | | |
| Test for overall effect: Z = 2.89 (P = 0.004) | | | | | | |
| Test for subgroup differences: Chi² = 3.19, df = 1 (P = 0.07), I² = 68.6% | | | | | | |

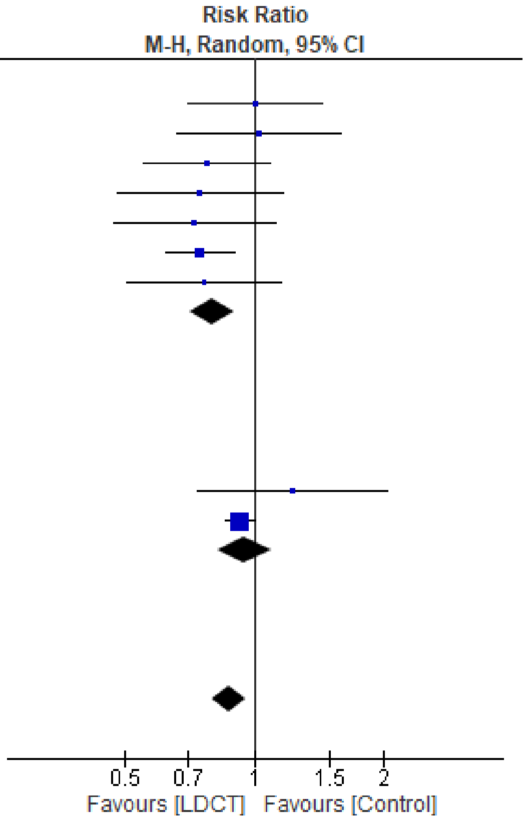

Figure S2.3 Lung cancer mortality, by comparator strategy

| Study or Subgroup | LDCT Events | LDCT Total | Control Events | Control Total | Weight | Risk Ratio M-H, Random, 95% CI |
|---|---|---|---|---|---|---|
| **4.3.1 1 round** | | | | | | |
| UKLS (Davies 2024, median 9.9 y) | 39 | 2028 | 51 | 2027 | 4.7% | 0.76 [0.51, 1.15] |
| **Subtotal (95% CI)** | | **2028** | | **2027** | **4.7%** | **0.76 [0.51, 1.15]** |
| Total events | 39 | | 51 | | | |
| Heterogeneity: Not applicable | | | | | | |
| Test for overall effect: Z = 1.28 (P = 0.20) | | | | | | |
| **4.3.2 2-3 rounds** | | | | | | |
| LSS (Doroudi 2018, median 5.2 y) | 32 | 1660 | 26 | 1658 | 3.1% | 1.23 [0.74, 2.05] |
| NLST (Pinsky 2019, median 12.3 y) | 1147 | 26722 | 1236 | 26732 | 48.3% | 0.93 [0.86, 1.00] |
| **Subtotal (95% CI)** | | **28382** | | **28390** | **51.4%** | **0.95 [0.82, 1.10]** |
| Total events | 1179 | | 1262 | | | |
| Heterogeneity: Tau² = 0.00; Chi² = 1.12, df = 1 (P = 0.29); I² = 11% | | | | | | |
| Test for overall effect: Z = 0.71 (P = 0.47) | | | | | | |
| **4.3.3 4-5 rounds** | | | | | | |
| DANTE (Infante 2015, median 8.35 y) | 59 | 1264 | 55 | 1186 | 6.1% | 1.01 [0.70, 1.44] |
| DLCST (Wille 2016, median 9.8 y) | 39 | 2052 | 38 | 2052 | 4.1% | 1.03 [0.66, 1.60] |
| ITALUNG (Paci 2021, median 11.3 y) | 58 | 1613 | 74 | 1593 | 6.9% | 0.77 [0.55, 1.08] |
| LUSI (Maldonado 2021, median 9.77 y) | 33 | 2029 | 44 | 2023 | 4.1% | 0.75 [0.48, 1.17] |
| MILD (Pastorino 2019, 9 years follow-up) | 40 | 2376 | 40 | 1723 | 4.3% | 0.73 [0.47, 1.12] |
| NELSON (de Koning 2020, 10y or to censoring) | 186 | 7900 | 248 | 7892 | 18.4% | 0.75 [0.62, 0.90] |
| **Subtotal (95% CI)** | | **17234** | | **16469** | **43.9%** | **0.80 [0.70, 0.91]** |
| Total events | 415 | | 499 | | | |
| Heterogeneity: Tau² = 0.00; Chi² = 3.58, df = 5 (P = 0.61); I² = 0% | | | | | | |
| Test for overall effect: Z = 3.39 (P = 0.0007) | | | | | | |
| **Total (95% CI)** | | **47644** | | **46886** | **100.0%** | **0.87 [0.79, 0.96]** |
| Total events | 1633 | | 1812 | | | |
| Heterogeneity: Tau² = 0.00; Chi² = 9.33, df = 8 (P = 0.32); I² = 14% | | | | | | |
| Test for overall effect: Z = 2.89 (P = 0.004) | | | | | | |
| Test for subgroup differences: Chi² = 3.23, df = 2 (P = 0.20), I² = 38.1% | | | | | | |

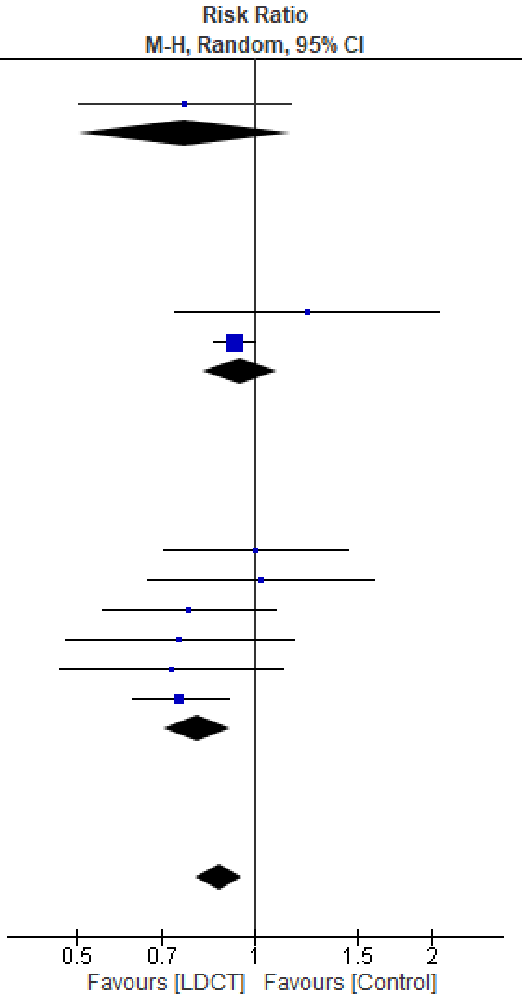

Figure S2.4 Lung cancer mortality, by number of screening rounds

| Study or Subgroup | LDCT Events | LDCT Total | Control Events | Control Total | Weight | Risk Ratio M-H, Random, 95% CI |
|---|---|---|---|---|---|---|
| **4.4.1 Annual screening interval** | | | | | | |
| DANTE (Infante 2015, median 8.35 y) | 59 | 1264 | 55 | 1186 | 4.4% | 1.01 [0.70, 1.44] |
| DLCST (Wille 2016, median 9.8 y) | 39 | 2052 | 38 | 2052 | 2.9% | 1.03 [0.66, 1.60] |
| ITALUNG (Paci 2021, median 11.3 y) | 58 | 1613 | 74 | 1593 | 5.0% | 0.77 [0.55, 1.08] |
| LSS (Doroudi 2018, median 5.2 y) | 32 | 1660 | 26 | 1658 | 2.2% | 1.23 [0.74, 2.05] |
| LUSI (Maldonado 2021, median 9.77 y) | 33 | 2029 | 44 | 2023 | 2.9% | 0.75 [0.48, 1.17] |
| MILD, annual (Pastorino 2019, 9 years follow-up) | 19 | 1190 | 20 | 862 | 1.5% | 0.69 [0.37, 1.28] |
| NLST (Pinsky 2019, median 12.3 y) | 1147 | 26722 | 1236 | 26732 | 61.2% | 0.93 [0.86, 1.00] |
| **Subtotal (95% CI)** | | **36530** | | **36106** | **80.0%** | **0.92 [0.86, 0.99]** |
| Total events | 1387 | | 1493 | | | |
| Heterogeneity: $Tau^2 = 0.00$; $Chi^2 = 4.42$, df = 6 (P = 0.62); $I^2 = 0\%$ | | | | | | |
| Test for overall effect: Z = 2.21 (P = 0.03) | | | | | | |
| **4.4.2 >Annual screening interval (on average)** | | | | | | |
| MILD, biennial (Pastorino 2019, 9 years follow-up) | 21 | 1186 | 20 | 862 | 1.6% | 0.76 [0.42, 1.40] |
| NELSON (de Koning 2020, 10y or to censoring) | 186 | 7900 | 248 | 7892 | 15.1% | 0.75 [0.62, 0.90] |
| **Subtotal (95% CI)** | | **9086** | | **8754** | **16.6%** | **0.75 [0.63, 0.90]** |
| Total events | 207 | | 268 | | | |
| Heterogeneity: $Tau^2 = 0.00$; $Chi^2 = 0.00$, df = 1 (P = 0.95); $I^2 = 0\%$ | | | | | | |
| Test for overall effect: Z = 3.14 (P = 0.002) | | | | | | |
| **4.4.3 One screen only** | | | | | | |
| UKLS (Davies 2024, median 9.9 y) | 39 | 2028 | 51 | 2027 | 3.3% | 0.76 [0.51, 1.15] |
| **Subtotal (95% CI)** | | **2028** | | **2027** | **3.3%** | **0.76 [0.51, 1.15]** |
| Total events | 39 | | 51 | | | |
| Heterogeneity: Not applicable | | | | | | |
| Test for overall effect: Z = 1.28 (P = 0.20) | | | | | | |
| **Total (95% CI)** | | **47644** | | **46887** | **100.0%** | **0.88 [0.82, 0.95]** |
| Total events | 1633 | | 1812 | | | |
| Heterogeneity: $Tau^2 = 0.00$; $Chi^2 = 9.38$, df = 9 (P = 0.40); $I^2 = 4\%$ | | | | | | |
| Test for overall effect: Z = 3.17 (P = 0.002) | | | | | | |
| Test for subgroup differences: $Chi^2 = 4.95$, df = 2 (P = 0.08), $I^2 = 59.6\%$ | | | | | | |

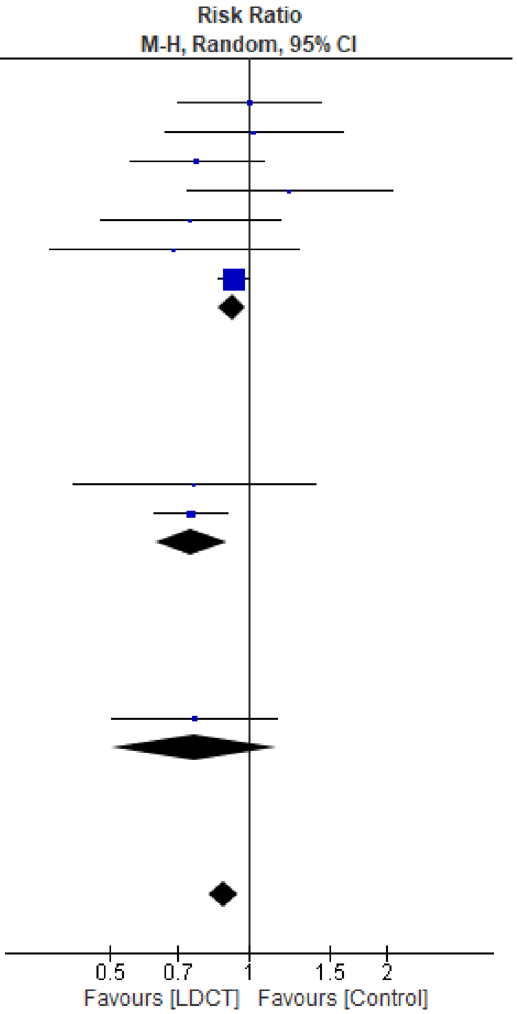

Figure S2.5 Lung cancer mortality, by screening interval

| Study or Subgroup | LDCT Events | LDCT Total | Control Events | Control Total | Weight | Risk Ratio M-H, Random, 95% CI |
|---|---|---|---|---|---|---|
| **4.5.1 >2 percent per year cancer detection rate in control group** | | | | | | |
| DANTE (Infante 2015, median 8.35 y) | 59 | 1264 | 55 | 1186 | 6.1% | 1.01 [0.70, 1.44] |
| ITALUNG (Paci 2021, median 11.3 y) | 58 | 1613 | 74 | 1593 | 6.9% | 0.77 [0.55, 1.08] |
| LSS (Doroudi 2018, median 5.2 y) | 32 | 1660 | 26 | 1658 | 3.1% | 1.23 [0.74, 2.05] |
| LUSI (Maldonado 2021, median 9.77 y) | 33 | 2029 | 44 | 2023 | 4.1% | 0.75 [0.48, 1.17] |
| MILD (Pastorino 2019, 9 years follow-up) | 40 | 2376 | 40 | 1723 | 4.3% | 0.73 [0.47, 1.12] |
| NELSON (de Koning 2020, 10y or to censoring) | 186 | 7900 | 248 | 7892 | 18.4% | 0.75 [0.62, 0.90] |
| NLST (Pinsky 2019, median 12.3 y) | 1147 | 26722 | 1236 | 26732 | 48.3% | 0.93 [0.86, 1.00] |
| UKLS (Davies 2024, median 9.9 y) | 39 | 2028 | 51 | 2027 | 4.7% | 0.76 [0.51, 1.15] |
| **Subtotal (95% CI)** | | **45592** | | **44834** | **95.9%** | **0.86 [0.78, 0.95]** |
| Total events | 1594 | | 1774 | | | |
| Heterogeneity: Tau² = 0.00; Chi² = 8.94, df = 7 (P = 0.26); I² = 22% | | | | | | |
| Test for overall effect: Z = 2.84 (P = 0.004) | | | | | | |
| **4.5.2 <=2 percent per year cancer detection rate in control group** | | | | | | |
| DLCST (Wille 2016, median 9.8 y) | 39 | 2052 | 38 | 2052 | 4.1% | 1.03 [0.66, 1.60] |
| **Subtotal (95% CI)** | | **2052** | | **2052** | **4.1%** | **1.03 [0.66, 1.60]** |
| Total events | 39 | | 38 | | | |
| Heterogeneity: Not applicable | | | | | | |
| Test for overall effect: Z = 0.12 (P = 0.91) | | | | | | |
| **Total (95% CI)** | | **47644** | | **46886** | **100.0%** | **0.87 [0.79, 0.96]** |
| Total events | 1633 | | 1812 | | | |
| Heterogeneity: Tau² = 0.00; Chi² = 9.33, df = 8 (P = 0.32); I² = 14% | | | | | | |
| Test for overall effect: Z = 2.89 (P = 0.004) | | | | | | |
| Test for subgroup differences: Chi² = 0.58, df = 1 (P = 0.45), I² = 0% | | | | | | |

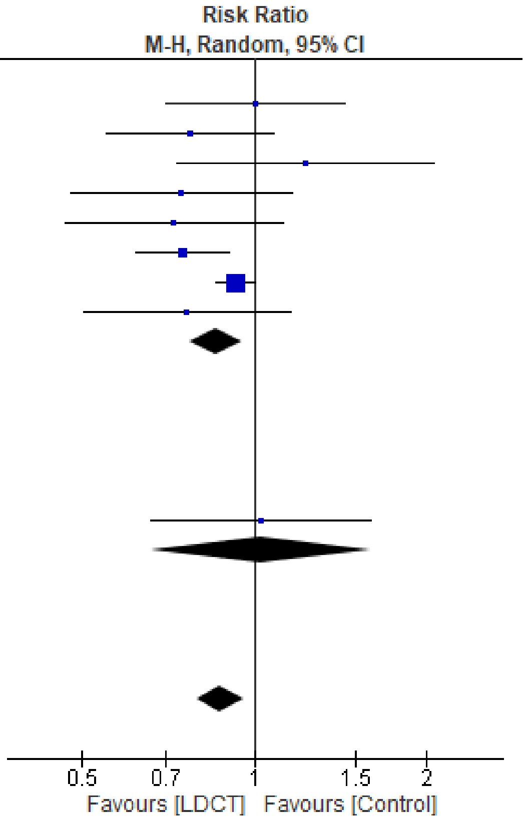

Figure S2.6 Lung cancer mortality, by 6-year lung cancer detection rate in comparator (≤2% vs >2%)

| Study or Subgroup | LDCT Events | LDCT Total | Control Events | Control Total | Weight | Risk Ratio M-H, Random, 95% CI |
|---|---|---|---|---|---|---|
| **4.6.1 >3 percent 6-year cancer detection rate in control group** | | | | | | |
| DANTE (Infante 2015, median 8.35 y) | 59 | 1264 | 55 | 1186 | 6.1% | 1.01 [0.70, 1.44] |
| ITALUNG (Paci 2021, median 11.3 y) | 58 | 1613 | 74 | 1593 | 6.9% | 0.77 [0.55, 1.08] |
| LSS (Doroudi 2018, median 5.2 y) | 32 | 1660 | 26 | 1658 | 3.1% | 1.23 [0.74, 2.05] |
| NLST (Pinsky 2019, median 12.3 y) | 1147 | 26722 | 1236 | 26732 | 48.3% | 0.93 [0.86, 1.00] |
| **Subtotal (95% CI)** | | **31259** | | **31169** | **64.4%** | **0.93 [0.86, 1.00]** |
| Total events | 1296 | | 1391 | | | |
| Heterogeneity: Tau² = 0.00; Chi² = 2.47, df = 3 (P = 0.48); I² = 0% | | | | | | |
| Test for overall effect: Z = 1.96 (P = 0.05) | | | | | | |
| **4.6.2 ≤3 percent 6-year cancer detection rate in control group** | | | | | | |
| DLCST (Wille 2016, median 9.8 y) | 39 | 2052 | 38 | 2052 | 4.1% | 1.03 [0.66, 1.60] |
| LUSI (Maldonado 2021, median 9.77 y) | 33 | 2029 | 44 | 2023 | 4.1% | 0.75 [0.48, 1.17] |
| MILD (Pastorino 2019, 9 years follow-up) | 40 | 2376 | 40 | 1723 | 4.3% | 0.73 [0.47, 1.12] |
| NELSON (de Koning 2020, 10y or to censoring) | 186 | 7900 | 248 | 7892 | 18.4% | 0.75 [0.62, 0.90] |
| UKLS (Davies 2024, median 9.9 y) | 39 | 2028 | 51 | 2027 | 4.7% | 0.76 [0.51, 1.15] |
| **Subtotal (95% CI)** | | **16385** | | **15717** | **35.6%** | **0.77 [0.67, 0.89]** |
| Total events | 337 | | 421 | | | |
| Heterogeneity: Tau² = 0.00; Chi² = 1.79, df = 4 (P = 0.77); I² = 0% | | | | | | |
| Test for overall effect: Z = 3.56 (P = 0.0004) | | | | | | |
| **Total (95% CI)** | | **47644** | | **46886** | **100.0%** | **0.87 [0.79, 0.96]** |
| Total events | 1633 | | 1812 | | | |
| Heterogeneity: Tau² = 0.00; Chi² = 9.33, df = 8 (P = 0.32); I² = 14% | | | | | | |
| Test for overall effect: Z = 2.89 (P = 0.004) | | | | | | |
| Test for subgroup differences: Chi² = 5.08, df = 1 (P = 0.02), I² = 80.3% | | | | | | |

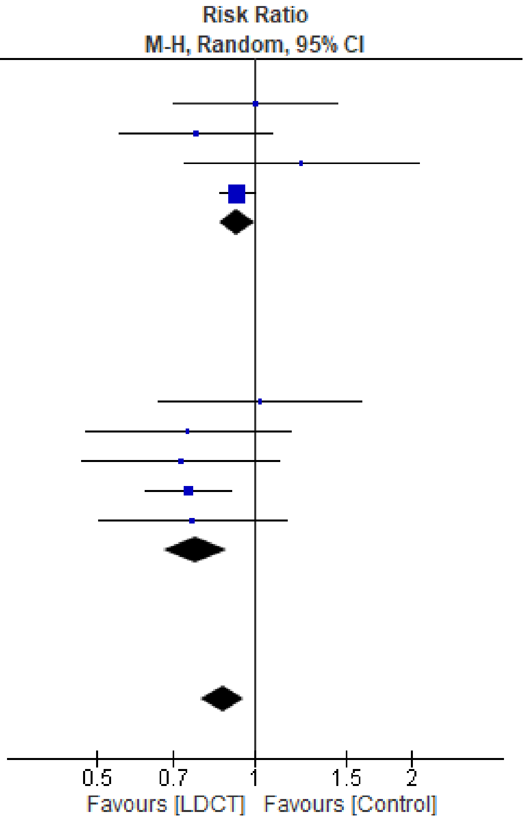

Figure S2.7 Lung cancer mortality, by 6-year lung cancer detection rate in comparator (≤3% vs >3%)

| Study or Subgroup | LDCT Events | LDCT Total | Control Events | Control Total | Weight | Risk Ratio M-H, Random, 95% CI |
|---|---|---|---|---|---|---|
| **4.7.1 Less sensitive (>=5mm)** | | | | | | |
| DANTE (Infante 2015, median 8.35 y) | 59 | 1264 | 55 | 1186 | 6.1% | 1.01 [0.70, 1.44] |
| DLCST (Wille 2016, median 9.8 y) | 39 | 2052 | 38 | 2052 | 4.1% | 1.03 [0.66, 1.60] |
| ITALUNG (Paci 2021, median 11.3 y) | 58 | 1613 | 74 | 1593 | 6.9% | 0.77 [0.55, 1.08] |
| LUSI (Maldonado 2021, median 9.77 y) | 33 | 2029 | 44 | 2023 | 4.1% | 0.75 [0.48, 1.17] |
| MILD (Pastorino 2019, 9 years follow-up) | 40 | 2376 | 40 | 1723 | 4.3% | 0.73 [0.47, 1.12] |
| NELSON (de Koning 2020, 10y or to censoring) | 186 | 7900 | 248 | 7892 | 18.4% | 0.75 [0.62, 0.90] |
| UKLS (Davies 2024, median 9.9 y) | 39 | 2028 | 51 | 2027 | 4.7% | 0.76 [0.51, 1.15] |
| **Subtotal (95% CI)** | | **19262** | | **18496** | **48.6%** | **0.80 [0.71, 0.90]** |
| Total events | 454 | | 550 | | | |
| Heterogeneity: Tau² = 0.00; Chi² = 3.63, df = 6 (P = 0.73); I² = 0% | | | | | | |
| Test for overall effect: Z = 3.62 (P = 0.0003) | | | | | | |
| **4.7.2 Most sensitive (3 to <5mm)** | | | | | | |
| LSS (Doroudi 2018, median 5.2 y) | 32 | 1660 | 26 | 1658 | 3.1% | 1.23 [0.74, 2.05] |
| NLST (Pinsky 2019, median 12.3 y) | 1147 | 26722 | 1236 | 26732 | 48.3% | 0.93 [0.86, 1.00] |
| **Subtotal (95% CI)** | | **28382** | | **28390** | **51.4%** | **0.95 [0.82, 1.10]** |
| Total events | 1179 | | 1262 | | | |
| Heterogeneity: Tau² = 0.00; Chi² = 1.12, df = 1 (P = 0.29); I² = 11% | | | | | | |
| Test for overall effect: Z = 0.71 (P = 0.47) | | | | | | |
| **Total (95% CI)** | | **47644** | | **46886** | **100.0%** | **0.87 [0.79, 0.96]** |
| Total events | 1633 | | 1812 | | | |
| Heterogeneity: Tau² = 0.00; Chi² = 9.33, df = 8 (P = 0.32); I² = 14% | | | | | | |
| Test for overall effect: Z = 2.89 (P = 0.004) | | | | | | |
| Test for subgroup differences: Chi² = 3.19, df = 1 (P = 0.07), I² = 68.6% | | | | | | |

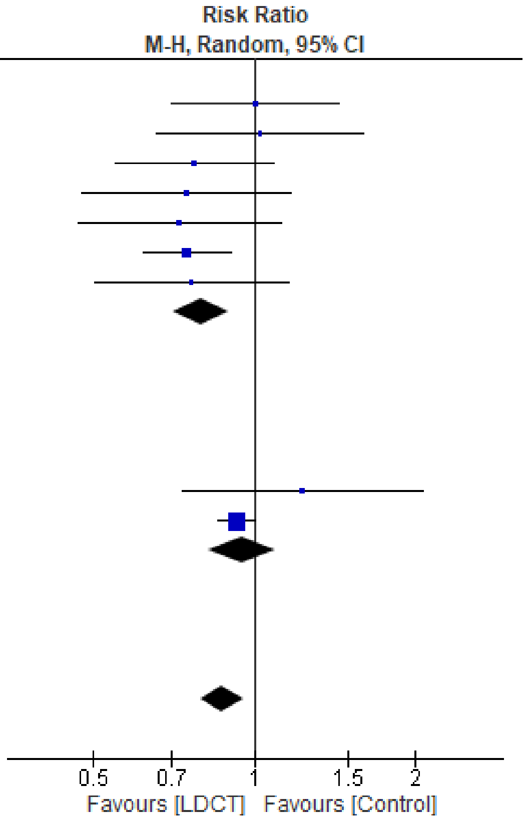

Figure S2.8 Lung cancer mortality, by nodule management strategy

| Study or Subgroup | LDCT Events | LDCT Total | Control Events | Control Total | Weight | Risk Ratio M-H, Random, 95% CI |
|---|---|---|---|---|---|---|
| **10.5.1 Men** | | | | | | |
| DANTE (Infante 2015, median 8.35 y) | 59 | 1264 | 55 | 1186 | 8.2% | 1.01 [0.70, 1.44] |
| LUSI (Becker 2020, mean 8.8 y) | 25 | 1315 | 27 | 1307 | 4.1% | 0.92 [0.54, 1.58] |
| NELSON (de Koning 2020, 10y or to censoring) | 160 | 6583 | 210 | 6612 | 17.9% | 0.77 [0.62, 0.94] |
| NLST (Pinsky 2019, median 12.3 y) | 733 | 15769 | 755 | 15761 | 31.3% | 0.97 [0.88, 1.07] |
| UKLS (Field 2021, median 7.3 y) | 22 | 1529 | 34 | 1507 | 4.2% | 0.64 [0.37, 1.09] |
| **Subtotal (95% CI)** | | **26460** | | **26373** | **65.6%** | **0.88 [0.76, 1.02]** |
| Total events | 999 | | 1081 | | | |
| Heterogeneity: Tau² = 0.01; Chi² = 6.34, df = 4 (P = 0.18); I² = 37% | | | | | | |
| Test for overall effect: Z = 1.63 (P = 0.10) | | | | | | |
| **10.5.2 Women** | | | | | | |
| LUSI (Becker 2020, mean 8.8 y) | 4 | 714 | 13 | 716 | 1.0% | 0.31 [0.10, 0.94] |
| NELSON (de Koning 2020, 10y or to censoring) | 26 | 1317 | 38 | 1277 | 4.8% | 0.66 [0.41, 1.09] |
| NLST (Pinsky 2019, median 12.3 y) | 414 | 10953 | 481 | 10969 | 27.0% | 0.86 [0.76, 0.98] |
| UKLS (Field 2021, median 7.3 y) | 8 | 499 | 12 | 520 | 1.6% | 0.69 [0.29, 1.69] |
| **Subtotal (95% CI)** | | **13483** | | **13482** | **34.4%** | **0.75 [0.56, 1.00]** |
| Total events | 452 | | 544 | | | |
| Heterogeneity: Tau² = 0.03; Chi² = 4.30, df = 3 (P = 0.23); I² = 30% | | | | | | |
| Test for overall effect: Z = 1.99 (P = 0.05) | | | | | | |
| **Total (95% CI)** | | **39943** | | **39855** | **100.0%** | **0.86 [0.76, 0.96]** |
| Total events | 1451 | | 1625 | | | |
| Heterogeneity: Tau² = 0.01; Chi² = 12.38, df = 8 (P = 0.13); I² = 35% | | | | | | |
| Test for overall effect: Z = 2.68 (P = 0.007) | | | | | | |
| Test for subgroup differences: Chi² = 1.03, df = 1 (P = 0.31), I² = 2.6% | | | | | | |

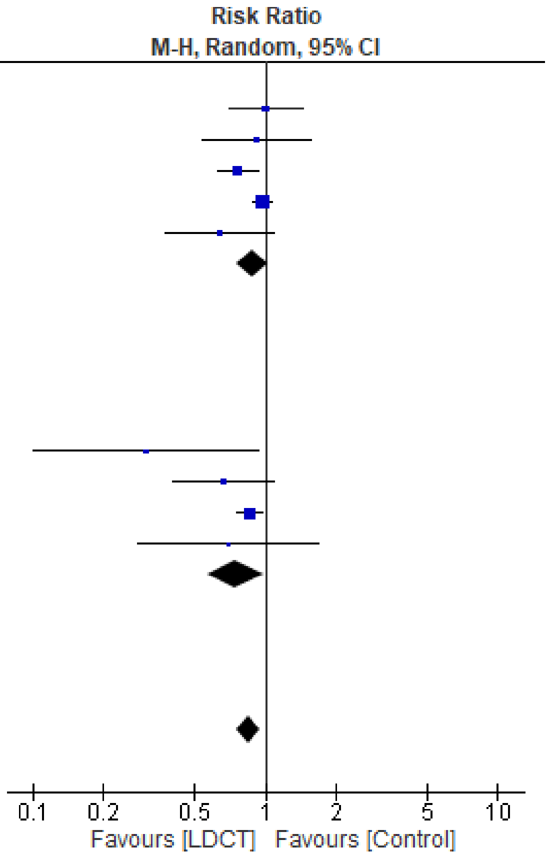

Figure S2.9 Lung cancer mortality, by sex

| Study or Subgroup | log[Risk Ratio] | SE | Weight | Risk Ratio IV, Random, 95% CI |
|---|---|---|---|---|
| **11.3.1 NLST** | | | | |
| NLST (Pinsky 2019, median 12.3 y) | 0.1204 | 0.0574 | 52.1% | 1.13 [1.01, 1.26] |
| **Subtotal (95% CI)** | | | **52.1%** | **1.13 [1.01, 1.26]** |
| Heterogeneity: Not applicable | | | | |
| Test for overall effect: Z = 2.10 (P = 0.04) | | | | |
| **11.3.2 NELSON** | | | | |
| NELSON (de Koning 2020, 10y or to censoring) | 0.1542 | 0.1778 | 28.5% | 1.17 [0.82, 1.65] |
| **Subtotal (95% CI)** | | | **28.5%** | **1.17 [0.82, 1.65]** |
| Heterogeneity: Not applicable | | | | |
| Test for overall effect: Z = 0.87 (P = 0.39) | | | | |
| **11.3.3 LUSI** | | | | |
| LUSI (Becker 2020, mean 8.8 y) | 1.0878 | 0.4227 | 8.5% | 2.97 [1.30, 6.80] |
| **Subtotal (95% CI)** | | | **8.5%** | **2.97 [1.30, 6.80]** |
| Heterogeneity: Not applicable | | | | |
| Test for overall effect: Z = 2.57 (P = 0.01) | | | | |
| **11.3.4 UKLS** | | | | |
| UKLS (Field 2021, median 7.3 y) | -0.0752 | 0.3626 | 11.0% | 0.93 [0.46, 1.89] |
| **Subtotal (95% CI)** | | | **11.0%** | **0.93 [0.46, 1.89]** |
| Heterogeneity: Not applicable | | | | |
| Test for overall effect: Z = 0.21 (P = 0.84) | | | | |
| **Total (95% CI)** | | | **100.0%** | **1.21 [0.93, 1.57]** |
| Heterogeneity: Tau² = 0.03; Chi² = 5.49, df = 3 (P = 0.14); I² = 45% | | | | |
| Test for overall effect: Z = 1.43 (P = 0.15) | | | | |
| Test for subgroup differences: Chi² = 5.49, df = 3 (P = 0.14), I² = 45.4% | | | | |

Risk Ratio
IV, Random, 95% CI

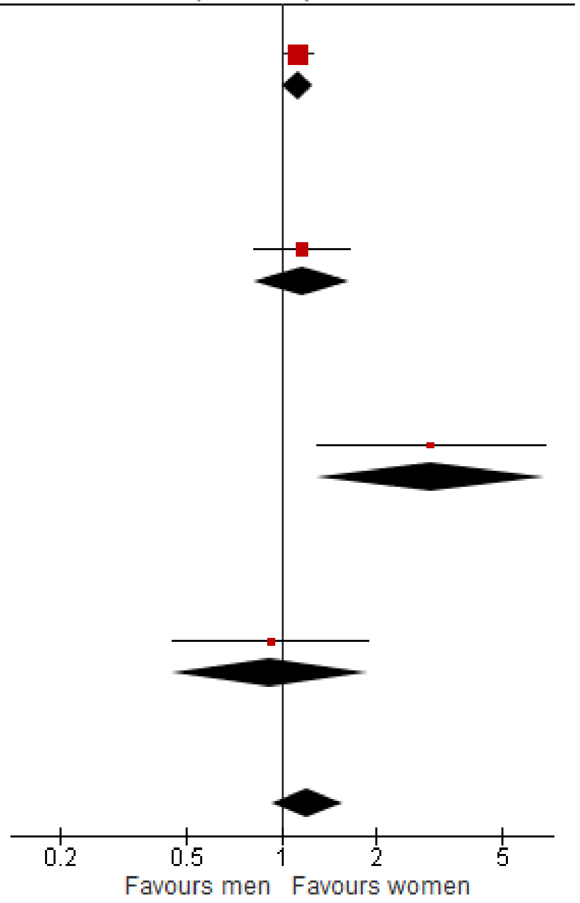

Figure S2.10 Lung cancer mortality, within study ratio of ratios by sex

| Study or Subgroup | LDCT Events | LDCT Total | Control Events | Control Total | Weight | Risk Ratio M-H, Random, 95% CI |
|---|---|---|---|---|---|---|
| **2.1.1 Some concerns** | | | | | | |
| DLCST (Wille 2016, median 9.8 y) | 165 | 2052 | 163 | 2052 | 5.1% | 1.01 [0.82, 1.25] |
| ITALUNG (Puliti 2019, median 11.3 y) | 203 | 1613 | 246 | 1593 | 7.2% | 0.81 [0.69, 0.97] |
| LSS (Doroudi 2018, median 5.2 y) | 139 | 1660 | 116 | 1658 | 4.0% | 1.20 [0.94, 1.52] |
| LUSI (Maldonado 2021, median 9.77 y) | 183 | 2029 | 174 | 2023 | 5.6% | 1.05 [0.86, 1.28] |
| MILD (Pastorino 2019, 9 years follow-up) | 137 | 2376 | 106 | 1723 | 3.8% | 0.94 [0.73, 1.20] |
| NELSON (de Koning 2020, 10y or to censoring) | 959 | 7900 | 974 | 7892 | 21.3% | 0.98 [0.90, 1.07] |
| NLST (Pinsky 2019, median 12.3 y) | 5253 | 26722 | 5366 | 26732 | 42.8% | 0.98 [0.95, 1.01] |
| UKLS (Davies 2024, median 9.9 y) | 313 | 2028 | 350 | 2027 | 10.2% | 0.89 [0.78, 1.03] |
| **Subtotal (95% CI)** | | **46380** | | **45700** | **100.0%** | **0.97 [0.92, 1.02]** |
| Total events | 7352 | | 7495 | | | |
| Heterogeneity: Tau² = 0.00; Chi² = 9.37, df = 7 (P = 0.23); I² = 25% | | | | | | |
| Test for overall effect: Z = 1.20 (P = 0.23) | | | | | | |
| **Total (95% CI)** | | **46380** | | **45700** | **100.0%** | **0.97 [0.92, 1.02]** |
| Total events | 7352 | | 7495 | | | |
| Heterogeneity: Tau² = 0.00; Chi² = 9.37, df = 7 (P = 0.23); I² = 25% | | | | | | |
| Test for overall effect: Z = 1.20 (P = 0.23) | | | | | | |
| Test for subgroup differences: Not applicable | | | | | | |

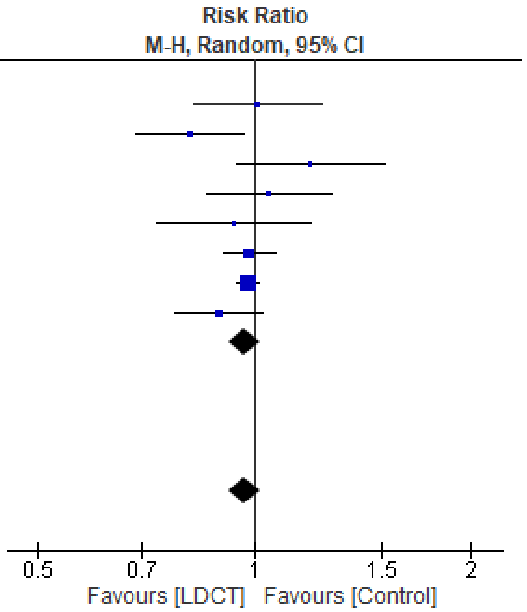

Figure S2.11 All-cause mortality, sensitivity analysis excluding high ROB studies (DANTE trial)

| Study or Subgroup | LDCT Events | LDCT Total | Control Events | Control Total | Weight | Risk Ratio M-H, Random, 95% CI |
|---|---|---|---|---|---|---|
| **2.2.1 X-ray screening comparator** | | | | | | |
| LSS (Doroudi 2018, median 5.2 y) | 139 | 1660 | 116 | 1658 | 3.0% | 1.20 [0.94, 1.52] |
| NLST (Pinsky 2019, median 12.3 y) | 5253 | 26722 | 5366 | 26732 | 48.9% | 0.98 [0.95, 1.01] |
| **Subtotal (95% CI)** | | **28382** | | **28390** | **51.9%** | **1.04 [0.87, 1.26]** |
| Total events | 5392 | | 5482 | | | |
| Heterogeneity: Tau² = 0.01; Chi² = 2.70, df = 1 (P = 0.10); I² = 63% | | | | | | |
| Test for overall effect: Z = 0.47 (P = 0.64) | | | | | | |
| **2.2.2 No screening comparator** | | | | | | |
| DANTE (Infante 2015, median 8.35 y) | 180 | 1264 | 176 | 1186 | 4.5% | 0.96 [0.79, 1.16] |
| DLCST (Wille 2016, median 9.8 y) | 165 | 2052 | 163 | 2052 | 3.9% | 1.01 [0.82, 1.25] |
| ITALUNG (Puliti 2019, median 11.3 y) | 203 | 1613 | 246 | 1593 | 5.5% | 0.81 [0.69, 0.97] |
| LUSI (Maldonado 2021, median 9.77 y) | 183 | 2029 | 174 | 2023 | 4.3% | 1.05 [0.86, 1.28] |
| MILD (Pastorino 2019, 9 years follow-up) | 137 | 2376 | 106 | 1723 | 2.8% | 0.94 [0.73, 1.20] |
| NELSON (de Koning 2020, 10y or to censoring) | 959 | 7900 | 974 | 7892 | 18.9% | 0.98 [0.90, 1.07] |
| UKLS (Davies 2024, median 9.9 y) | 313 | 2028 | 350 | 2027 | 8.1% | 0.89 [0.78, 1.03] |
| **Subtotal (95% CI)** | | **19262** | | **18496** | **48.1%** | **0.95 [0.90, 1.01]** |
| Total events | 2140 | | 2189 | | | |
| Heterogeneity: Tau² = 0.00; Chi² = 5.76, df = 6 (P = 0.45); I² = 0% | | | | | | |
| Test for overall effect: Z = 1.72 (P = 0.09) | | | | | | |
| **Total (95% CI)** | | **47644** | | **46886** | **100.0%** | **0.97 [0.93, 1.01]** |
| Total events | 7532 | | 7671 | | | |
| Heterogeneity: Tau² = 0.00; Chi² = 9.39, df = 8 (P = 0.31); I² = 15% | | | | | | |
| Test for overall effect: Z = 1.37 (P = 0.17) | | | | | | |
| Test for subgroup differences: Chi² = 0.90, df = 1 (P = 0.34), I² = 0% | | | | | | |

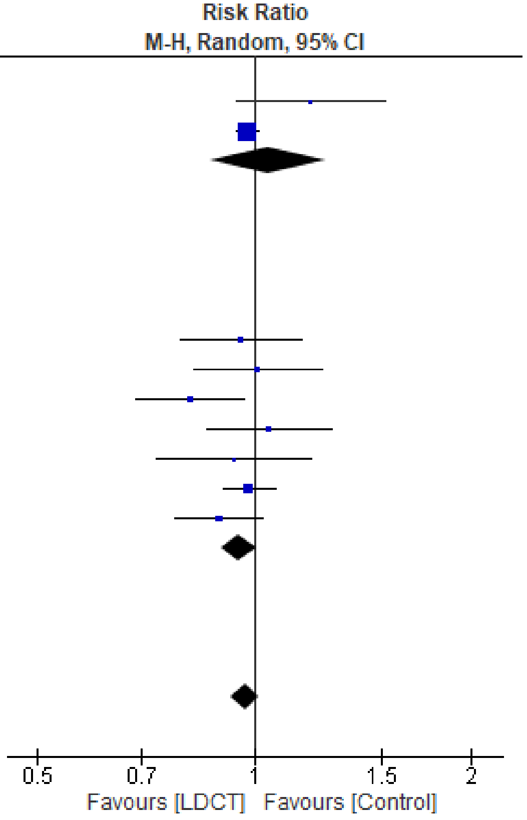

Figure S2.12 All-cause mortality, by comparator strategy

| Study or Subgroup | LDCT Events | LDCT Total | Control Events | Control Total | Weight | Risk Ratio M-H, Random, 95% CI |
|---|---|---|---|---|---|---|
| **2.3.1 1 round** | | | | | | |
| UKLS (Davies 2024, median 9.9 y) | 313 | 2028 | 350 | 2027 | 8.1% | 0.89 [0.78, 1.03] |
| **Subtotal (95% CI)** | | **2028** | | **2027** | **8.1%** | **0.89 [0.78, 1.03]** |
| Total events | 313 | | 350 | | | |
| Heterogeneity: Not applicable | | | | | | |
| Test for overall effect: Z = 1.58 (P = 0.11) | | | | | | |
| **2.3.2 2-3 rounds** | | | | | | |
| LSS (Doroudi 2018, median 5.2 y) | 139 | 1660 | 116 | 1658 | 3.0% | 1.20 [0.94, 1.52] |
| NLST (Pinsky 2019, median 12.3 y) | 5253 | 26722 | 5366 | 26732 | 48.9% | 0.98 [0.95, 1.01] |
| **Subtotal (95% CI)** | | **28382** | | **28390** | **51.9%** | **1.04 [0.87, 1.26]** |
| Total events | 5392 | | 5482 | | | |
| Heterogeneity: Tau² = 0.01; Chi² = 2.70, df = 1 (P = 0.10); I² = 63% | | | | | | |
| Test for overall effect: Z = 0.47 (P = 0.64) | | | | | | |
| **2.3.3 4+ rounds** | | | | | | |
| DANTE (Infante 2015, median 8.35 y) | 180 | 1264 | 176 | 1186 | 4.5% | 0.96 [0.79, 1.16] |
| DLCST (Wille 2016, median 9.8 y) | 165 | 2052 | 163 | 2052 | 3.9% | 1.01 [0.82, 1.25] |
| ITALUNG (Puliti 2019, median 11.3 y) | 203 | 1613 | 246 | 1593 | 5.5% | 0.81 [0.69, 0.97] |
| LUSI (Maldonado 2021, median 9.77 y) | 183 | 2029 | 174 | 2023 | 4.3% | 1.05 [0.86, 1.28] |
| MILD (Pastorino 2019, 9 years follow-up) | 137 | 2376 | 106 | 1723 | 2.8% | 0.94 [0.73, 1.20] |
| NELSON (de Koning 2020, 10y or to censoring) | 959 | 7900 | 974 | 7892 | 18.9% | 0.98 [0.90, 1.07] |
| **Subtotal (95% CI)** | | **17234** | | **16469** | **40.0%** | **0.96 [0.91, 1.02]** |
| Total events | 1827 | | 1839 | | | |
| Heterogeneity: Tau² = 0.00; Chi² = 4.82, df = 5 (P = 0.44); I² = 0% | | | | | | |
| Test for overall effect: Z = 1.19 (P = 0.23) | | | | | | |
| **Total (95% CI)** | | **47644** | | **46886** | **100.0%** | **0.97 [0.93, 1.01]** |
| Total events | 7532 | | 7671 | | | |
| Heterogeneity: Tau² = 0.00; Chi² = 9.39, df = 8 (P = 0.31); I² = 15% | | | | | | |
| Test for overall effect: Z = 1.37 (P = 0.17) | | | | | | |
| Test for subgroup differences: Chi² = 1.84, df = 2 (P = 0.40), I² = 0% | | | | | | |

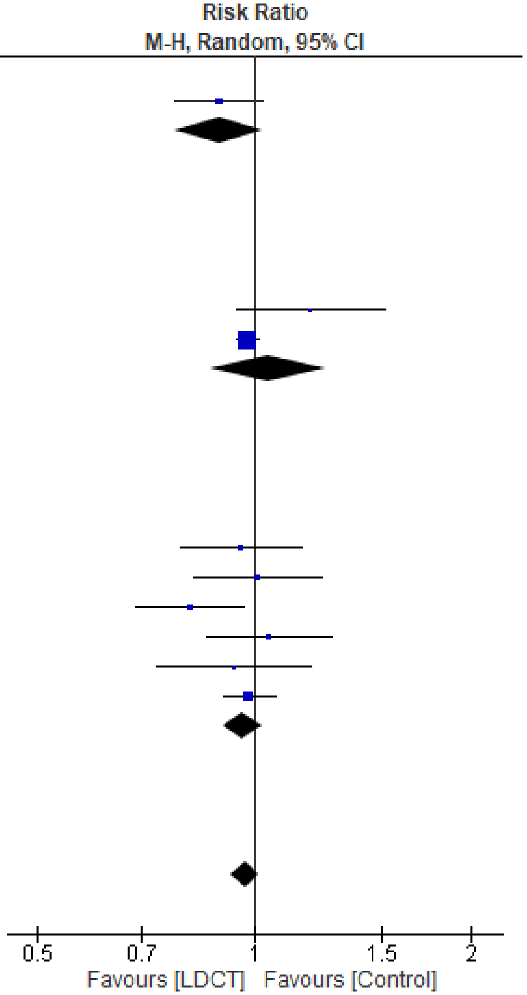

Figure S2.13 All-cause mortality, by number of screening rounds

| Study or Subgroup | LDCT Events | LDCT Total | Control Events | Control Total | Weight | Risk Ratio M-H, Random, 95% CI |
|---|---|---|---|---|---|---|
| **2.4.1 Annual screening interval** | | | | | | |
| DANTE (Infante 2015, median 8.35 y) | 180 | 1264 | 176 | 1186 | 4.2% | 0.96 [0.79, 1.16] |
| DLCST (Wille 2016, median 9.8 y) | 165 | 2052 | 163 | 2052 | 3.6% | 1.01 [0.82, 1.25] |
| ITALUNG (Puliti 2019, median 11.3 y) | 203 | 1613 | 246 | 1593 | 5.2% | 0.81 [0.69, 0.97] |
| LSS (Doroudi 2018, median 5.2 y) | 139 | 1660 | 116 | 1658 | 2.8% | 1.20 [0.94, 1.52] |
| LUSI (Maldonado 2021, median 9.77 y) | 183 | 2029 | 174 | 2023 | 4.0% | 1.05 [0.86, 1.28] |
| MILD, annual (Pastorino 2019, 9 years follow-up) | 76 | 1190 | 53 | 862 | 1.4% | 1.04 [0.74, 1.46] |
| NLST (Pinsky 2019, median 12.3 y) | 5253 | 26722 | 5366 | 26732 | 51.6% | 0.98 [0.95, 1.01] |
| **Subtotal (95% CI)** | | **36530** | | **36106** | **72.8%** | **0.98 [0.92, 1.05]** |
| Total events | 6199 | | 6294 | | | |
| Heterogeneity: Tau² = 0.00; Chi² = 7.83, df = 6 (P = 0.25); I² = 23% | | | | | | |
| Test for overall effect: Z = 0.58 (P = 0.56) | | | | | | |
| **2.4.2 >Annual screening interval (on average)** | | | | | | |
| MILD, biennial (Pastorino 2019, 9 years follow-up) | 61 | 1186 | 53 | 862 | 1.3% | 0.84 [0.59, 1.20] |
| NELSON (de Koning 2020, 10y or to censoring) | 959 | 7900 | 974 | 7892 | 18.3% | 0.98 [0.90, 1.07] |
| **Subtotal (95% CI)** | | **9086** | | **8754** | **19.5%** | **0.98 [0.90, 1.06]** |
| Total events | 1020 | | 1027 | | | |
| Heterogeneity: Tau² = 0.00; Chi² = 0.75, df = 1 (P = 0.39); I² = 0% | | | | | | |
| Test for overall effect: Z = 0.60 (P = 0.55) | | | | | | |
| **2.4.3 One screen only** | | | | | | |
| UKLS (Davies 2024, median 9.9 y) | 313 | 2028 | 350 | 2027 | 7.7% | 0.89 [0.78, 1.03] |
| **Subtotal (95% CI)** | | **2028** | | **2027** | **7.7%** | **0.89 [0.78, 1.03]** |
| Total events | 313 | | 350 | | | |
| Heterogeneity: Not applicable | | | | | | |
| Test for overall effect: Z = 1.58 (P = 0.11) | | | | | | |
| **Total (95% CI)** | | **47644** | | **46887** | **100.0%** | **0.97 [0.93, 1.01]** |
| Total events | 7532 | | 7671 | | | |
| Heterogeneity: Tau² = 0.00; Chi² = 10.13, df = 9 (P = 0.34); I² = 11% | | | | | | |
| Test for overall effect: Z = 1.40 (P = 0.16) | | | | | | |
| Test for subgroup differences: Chi² = 1.44, df = 2 (P = 0.49), I² = 0% | | | | | | |

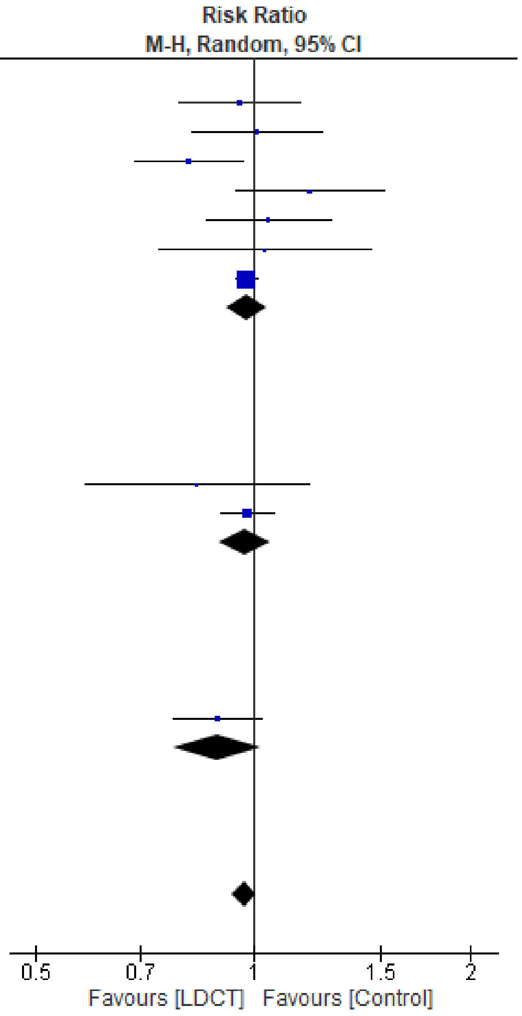

Figure S2.14 All-cause mortality, by screening interval

| Study or Subgroup | LDCT Events | LDCT Total | Control Events | Control Total | Weight | Risk Ratio M-H, Random, 95% CI |
|---|---|---|---|---|---|---|
| **2.5.1 ≤2 percent 6-year cancer detection rate in control group** | | | | | | |
| DLCST (Wille 2016, median 9.8 y) | 165 | 2052 | 163 | 2052 | 3.9% | 1.01 [0.82, 1.25] |
| **Subtotal (95% CI)** | | **2052** | | **2052** | **3.9%** | **1.01 [0.82, 1.25]** |
| Total events | 165 | | 163 | | | |
| Heterogeneity: Not applicable | | | | | | |
| Test for overall effect: Z = 0.12 (P = 0.91) | | | | | | |
| **2.5.2 >2 percent 6-year cancer detection rate in control group** | | | | | | |
| DANTE (Infante 2015, median 8.35 y) | 180 | 1264 | 176 | 1186 | 4.5% | 0.96 [0.79, 1.16] |
| ITALUNG (Puliti 2019, median 11.3 y) | 203 | 1613 | 246 | 1593 | 5.5% | 0.81 [0.69, 0.97] |
| LSS (Doroudi 2018, median 5.2 y) | 139 | 1660 | 116 | 1658 | 3.0% | 1.20 [0.94, 1.52] |
| LUSI (Maldonado 2021, median 9.77 y) | 183 | 2029 | 174 | 2023 | 4.3% | 1.05 [0.86, 1.28] |
| MILD (Pastorino 2019, 9 years follow-up) | 137 | 2376 | 106 | 1723 | 2.8% | 0.94 [0.73, 1.20] |
| NELSON (de Koning 2020, 10y or to censoring) | 959 | 7900 | 974 | 7892 | 18.9% | 0.98 [0.90, 1.07] |
| NLST (Pinsky 2019, median 12.3 y) | 5253 | 26722 | 5366 | 26732 | 48.9% | 0.98 [0.95, 1.01] |
| UKLS (Davies 2024, median 9.9 y) | 313 | 2028 | 350 | 2027 | 8.1% | 0.89 [0.78, 1.03] |
| **Subtotal (95% CI)** | | **45592** | | **44834** | **96.1%** | **0.97 [0.92, 1.02]** |
| Total events | 7367 | | 7508 | | | |
| Heterogeneity: Tau² = 0.00; Chi² = 9.27, df = 7 (P = 0.23); I² = 24% | | | | | | |
| Test for overall effect: Z = 1.33 (P = 0.18) | | | | | | |
| **Total (95% CI)** | | **47644** | | **46886** | **100.0%** | **0.97 [0.93, 1.01]** |
| Total events | 7532 | | 7671 | | | |
| Heterogeneity: Tau² = 0.00; Chi² = 9.39, df = 8 (P = 0.31); I² = 15% | | | | | | |
| Test for overall effect: Z = 1.37 (P = 0.17) | | | | | | |
| Test for subgroup differences: Chi² = 0.17, df = 1 (P = 0.68), I² = 0% | | | | | | |

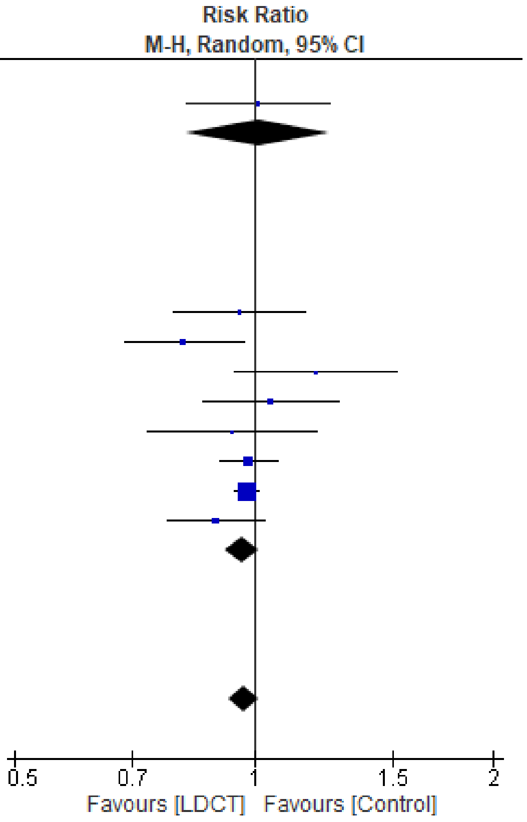

Figure S2.15 All-cause mortality, by 6-year lung cancer detection rate in comparator (≤2% vs >2%)

| Study or Subgroup | LDCT Events | LDCT Total | Control Events | Control Total | Weight | Risk Ratio M-H, Random, 95% CI |
|---|---|---|---|---|---|---|
| **2.6.1 ≤3 percent 6-year cancer detection rate in control group** | | | | | | |
| DLCST (Wille 2016, median 9.8 y) | 165 | 2052 | 163 | 2052 | 3.9% | 1.01 [0.82, 1.25] |
| LUSI (Maldonado 2021, median 9.77 y) | 183 | 2029 | 174 | 2023 | 4.3% | 1.05 [0.86, 1.28] |
| MILD (Pastorino 2019, 9 years follow-up) | 137 | 2376 | 106 | 1723 | 2.8% | 0.94 [0.73, 1.20] |
| NELSON (de Koning 2020, 10y or to censoring) | 959 | 7900 | 974 | 7892 | 18.9% | 0.98 [0.90, 1.07] |
| UKLS (Davies 2024, median 9.9 y) | 313 | 2028 | 350 | 2027 | 8.1% | 0.89 [0.78, 1.03] |
| **Subtotal (95% CI)** | | **16385** | | **15717** | **38.0%** | **0.97 [0.91, 1.03]** |
| Total events | 1757 | | 1767 | | | |
| Heterogeneity: Tau² = 0.00; Chi² = 2.26, df = 4 (P = 0.69); I² = 0% | | | | | | |
| Test for overall effect: Z = 0.94 (P = 0.35) | | | | | | |
| **2.6.2 >3 percent 6-year cancer detection rate in control group** | | | | | | |
| DANTE (Infante 2015, median 8.35 y) | 180 | 1264 | 176 | 1186 | 4.5% | 0.96 [0.79, 1.16] |
| ITALUNG (Puliti 2019, median 11.3 y) | 203 | 1613 | 246 | 1593 | 5.5% | 0.81 [0.69, 0.97] |
| LSS (Doroudi 2018, median 5.2 y) | 139 | 1660 | 116 | 1658 | 3.0% | 1.20 [0.94, 1.52] |
| NLST (Pinsky 2019, median 12.3 y) | 5253 | 26722 | 5366 | 26732 | 48.9% | 0.98 [0.95, 1.01] |
| **Subtotal (95% CI)** | | **31259** | | **31169** | **62.0%** | **0.97 [0.86, 1.08]** |
| Total events | 5775 | | 5904 | | | |
| Heterogeneity: Tau² = 0.01; Chi² = 7.11, df = 3 (P = 0.07); I² = 58% | | | | | | |
| Test for overall effect: Z = 0.61 (P = 0.54) | | | | | | |
| **Total (95% CI)** | | **47644** | | **46886** | **100.0%** | **0.97 [0.93, 1.01]** |
| Total events | 7532 | | 7671 | | | |
| Heterogeneity: Tau² = 0.00; Chi² = 9.39, df = 8 (P = 0.31); I² = 15% | | | | | | |
| Test for overall effect: Z = 1.37 (P = 0.17) | | | | | | |
| Test for subgroup differences: Chi² = 0.01, df = 1 (P = 0.94), I² = 0% | | | | | | |

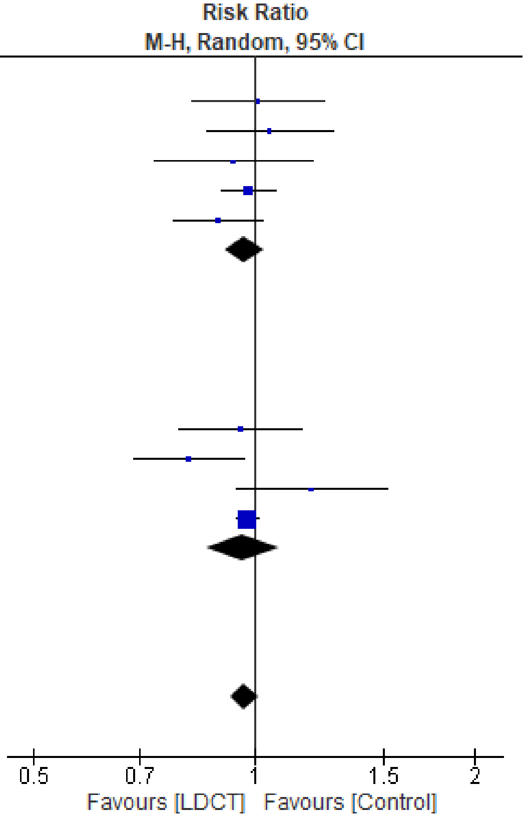

Figure S2.16 All-cause mortality, by 6-year lung cancer detection rate in comparator (≤3% vs >3%)

| Study or Subgroup | LDCT Events | LDCT Total | Control Events | Control Total | Weight | Risk Ratio M-H, Random, 95% CI |
|---|---|---|---|---|---|---|
| **2.7.1 Most sensitive** | | | | | | |
| LSS (Doroudi 2018, median 5.2 y) | 139 | 1660 | 116 | 1658 | 3.0% | 1.20 [0.94, 1.52] |
| NLST (Pinsky 2019, median 12.3 y) | 5253 | 26722 | 5366 | 26732 | 48.9% | 0.98 [0.95, 1.01] |
| **Subtotal (95% CI)** | | **28382** | | **28390** | **51.9%** | **1.04 [0.87, 1.26]** |
| Total events | 5392 | | 5482 | | | |
| Heterogeneity: Tau² = 0.01; Chi² = 2.70, df = 1 (P = 0.10); I² = 63% | | | | | | |
| Test for overall effect: Z = 0.47 (P = 0.64) | | | | | | |
| **2.7.2 Less sensitive** | | | | | | |
| DANTE (Infante 2015, median 8.35 y) | 180 | 1264 | 176 | 1186 | 4.5% | 0.96 [0.79, 1.16] |
| DLCST (Wille 2016, median 9.8 y) | 165 | 2052 | 163 | 2052 | 3.9% | 1.01 [0.82, 1.25] |
| ITALUNG (Puliti 2019, median 11.3 y) | 203 | 1613 | 246 | 1593 | 5.5% | 0.81 [0.69, 0.97] |
| LUSI (Maldonado 2021, median 9.77 y) | 183 | 2029 | 174 | 2023 | 4.3% | 1.05 [0.86, 1.28] |
| MILD (Pastorino 2019, 9 years follow-up) | 137 | 2376 | 106 | 1723 | 2.8% | 0.94 [0.73, 1.20] |
| NELSON (de Koning 2020, 10y or to censoring) | 959 | 7900 | 974 | 7892 | 18.9% | 0.98 [0.90, 1.07] |
| UKLS (Davies 2024, median 9.9 y) | 313 | 2028 | 350 | 2027 | 8.1% | 0.89 [0.78, 1.03] |
| **Subtotal (95% CI)** | | **19262** | | **18496** | **48.1%** | **0.95 [0.90, 1.01]** |
| Total events | 2140 | | 2189 | | | |
| Heterogeneity: Tau² = 0.00; Chi² = 5.76, df = 6 (P = 0.45); I² = 0% | | | | | | |
| Test for overall effect: Z = 1.72 (P = 0.09) | | | | | | |
| **Total (95% CI)** | | **47644** | | **46886** | **100.0%** | **0.97 [0.93, 1.01]** |
| Total events | 7532 | | 7671 | | | |
| Heterogeneity: Tau² = 0.00; Chi² = 9.39, df = 8 (P = 0.31); I² = 15% | | | | | | |
| Test for overall effect: Z = 1.37 (P = 0.17) | | | | | | |
| Test for subgroup differences: Chi² = 0.90, df = 1 (P = 0.34), I² = 0% | | | | | | |

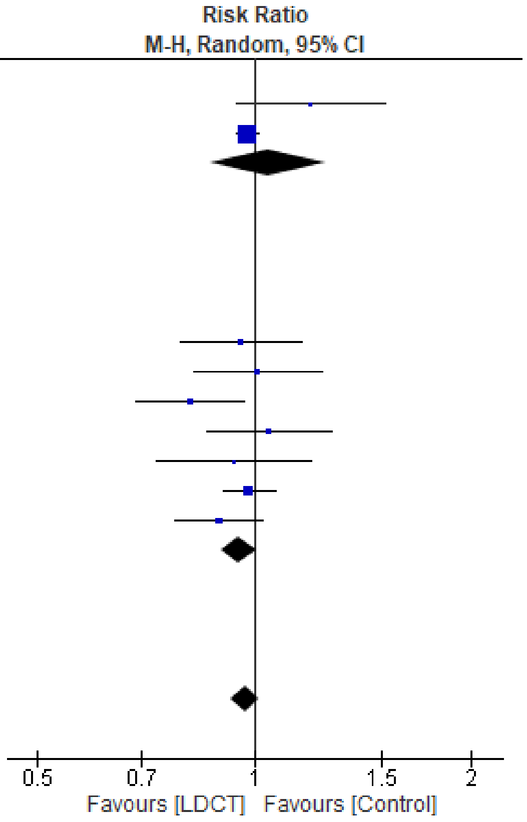

Figure S2.17 All-cause mortality, by nodule management strategy

| Study or Subgroup | LDCT Events | LDCT Total | Control Events | Control Total | Weight | Risk Ratio M-H, Random, 95% CI |
|---|---|---|---|---|---|---|
| **9.1.1 Men** | | | | | | |
| DANTE (Infante 2015, median 8.35 y) | 180 | 1264 | 176 | 1186 | 5.8% | 0.96 [0.79, 1.16] |
| LUSI (Becker 2020, mean 8.8 y) | 126 | 1315 | 123 | 1307 | 3.8% | 1.02 [0.80, 1.29] |
| NELSON (de Koning 2020, 10y or to censoring) | 868 | 6583 | 860 | 6612 | 27.5% | 1.01 [0.93, 1.11] |
| NLST (Pinsky 2013, median 6.5 y) | 1338 | 15769 | 1420 | 15761 | 41.8% | 0.94 [0.88, 1.01] |
| **Subtotal (95% CI)** | | **24931** | | **24866** | **78.8%** | **0.97 [0.92, 1.02]** |
| Total events | 2512 | | 2579 | | | |
| Heterogeneity: Tau² = 0.00; Chi² = 1.80, df = 3 (P = 0.62); I² = 0% | | | | | | |
| Test for overall effect: Z = 1.10 (P = 0.27) | | | | | | |
| **9.1.2 Women** | | | | | | |
| LUSI (Becker 2020, mean 8.8 y) | 22 | 714 | 27 | 716 | 0.7% | 0.82 [0.47, 1.42] |
| NELSON (de Koning 2020, 10y or to censoring) | 91 | 1317 | 114 | 1277 | 3.0% | 0.77 [0.59, 1.01] |
| NLST (Pinsky 2013, median 6.5 y) | 574 | 10953 | 619 | 10969 | 17.4% | 0.93 [0.83, 1.04] |
| **Subtotal (95% CI)** | | **12984** | | **12962** | **21.2%** | **0.90 [0.81, 1.00]** |
| Total events | 687 | | 760 | | | |
| Heterogeneity: Tau² = 0.00; Chi² = 1.67, df = 2 (P = 0.43); I² = 0% | | | | | | |
| Test for overall effect: Z = 2.04 (P = 0.04) | | | | | | |
| **Total (95% CI)** | | **37915** | | **37828** | **100.0%** | **0.96 [0.91, 1.00]** |
| Total events | 3199 | | 3339 | | | |
| Heterogeneity: Tau² = 0.00; Chi² = 5.18, df = 6 (P = 0.52); I² = 0% | | | | | | |
| Test for overall effect: Z = 1.92 (P = 0.06) | | | | | | |
| Test for subgroup differences: Chi² = 1.71, df = 1 (P = 0.19), I² = 41.4% | | | | | | |

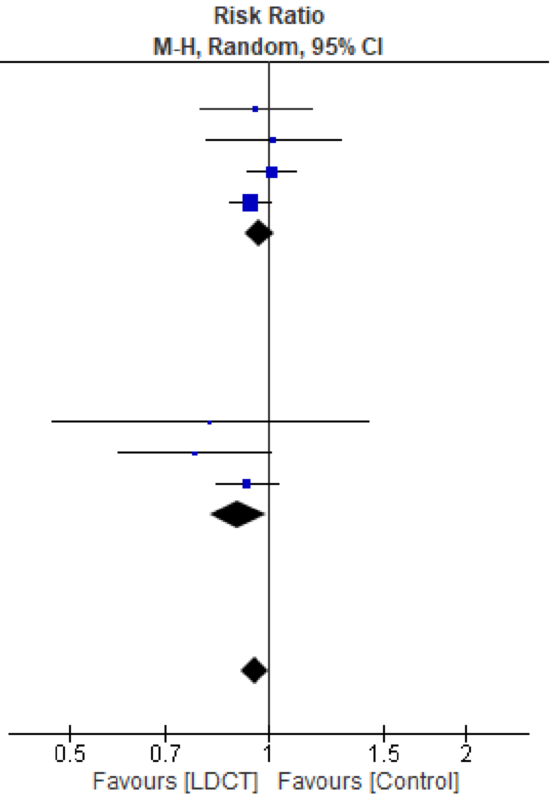

Figure S2.18 All-cause mortality, by sex

| Study or Subgroup | log[Risk Ratio] | SE | Weight | Risk Ratio IV, Random, 95% CI |
|---|---|---|---|---|
| **11.2.1 NLST** | | | | |
| NLST (Pinsky 2013, median 6.5 y) | 0.0106953 | 0.0463436 | 45.9% | 1.01 [0.92, 1.11] |
| **Subtotal (95% CI)** | | | **45.9%** | **1.01 [0.92, 1.11]** |
| Heterogeneity: Not applicable | | | | |
| Test for overall effect: Z = 0.23 (P = 0.82) | | | | |
| **11.2.2 NELSON** | | | | |
| NELSON (de Koning 2020, 10y or to censoring) | 0.2713151 | 0.091137 | 36.5% | 1.31 [1.10, 1.57] |
| **Subtotal (95% CI)** | | | **36.5%** | **1.31 [1.10, 1.57]** |
| Heterogeneity: Not applicable | | | | |
| Test for overall effect: Z = 2.98 (P = 0.003) | | | | |
| **11.2.3 LUSI** | | | | |
| LUSI (Becker 2020, mean 8.8 y) | 0.2182536 | 0.2019726 | 17.6% | 1.24 [0.84, 1.85] |
| **Subtotal (95% CI)** | | | **17.6%** | **1.24 [0.84, 1.85]** |
| Heterogeneity: Not applicable | | | | |
| Test for overall effect: Z = 1.08 (P = 0.28) | | | | |
| **Total (95% CI)** | | | **100.0%** | **1.15 [0.94, 1.42]** |
| Heterogeneity: $Tau^2 = 0.02$; $Chi^2 = 7.06$, df = 2 (P = 0.03); $I^2 = 72\%$ | | | | |
| Test for overall effect: Z = 1.36 (P = 0.18) | | | | |
| Test for subgroup differences: $Chi^2 = 7.06$, df = 2 (P = 0.03), $I^2 = 71.7\%$ | | | | |

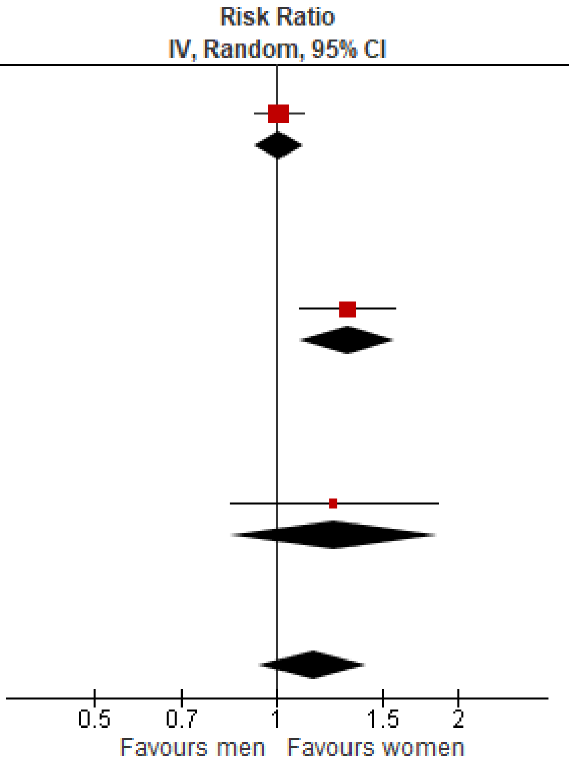

Figure S2.19 All-cause mortality, within study ratio of ratios by sex

| Study or Subgroup | LDCT Events | LDCT Total | Control Events | Control Total | Weight | Risk Ratio M-H, Random, 95% CI |
|---|---|---|---|---|---|---|
| **6.1.2 Some concerns of bias** | | | | | | |
| DLCST (Wille 2016, median 9.8 y) | 96 | 2052 | 53 | 2052 | 16.5% | 1.81 [1.30, 2.52] |
| ITALUNG (Paci 2021, median 11.3 y) | 91 | 1613 | 100 | 1593 | 19.5% | 0.90 [0.68, 1.18] |
| LUSI (Maldonado 2021, median 9.77 y) | 90 | 2029 | 74 | 2023 | 18.0% | 1.21 [0.90, 1.64] |
| NELSON (de Koning 2020, 10y or to censoring) | 344 | 6583 | 304 | 6612 | 27.2% | 1.14 [0.98, 1.32] |
| UKLS (Davies 2024, median 9.1 y) | 91 | 2028 | 86 | 2027 | 18.7% | 1.06 [0.79, 1.41] |
| **Subtotal (95% CI)** | | **14305** | | **14307** | **100.0%** | **1.17 [0.97, 1.42]** |
| Total events | 712 | | 617 | | | |
| Heterogeneity: Tau² = 0.03; Chi² = 10.84, df = 4 (P = 0.03); I² = 63% | | | | | | |
| Test for overall effect: Z = 1.62 (P = 0.10) | | | | | | |
| **Total (95% CI)** | | **14305** | | **14307** | **100.0%** | **1.17 [0.97, 1.42]** |
| Total events | 712 | | 617 | | | |
| Heterogeneity: Tau² = 0.03; Chi² = 10.84, df = 4 (P = 0.03); I² = 63% | | | | | | |
| Test for overall effect: Z = 1.62 (P = 0.10) | | | | | | |
| Test for subgroup differences: Not applicable | | | | | | |

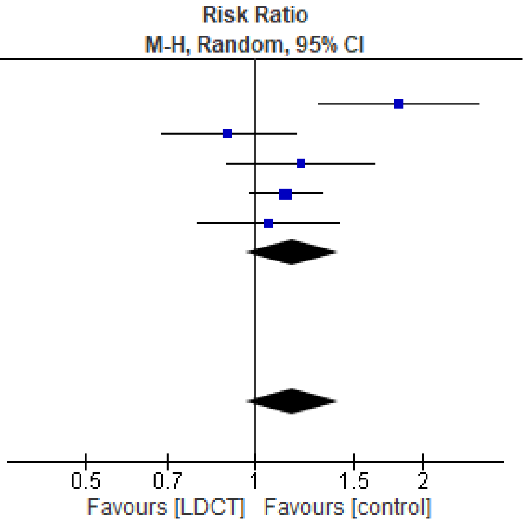

Figure S2.20 Overdiagnosis, sensitivity analysis excluding high ROB studies and those with an X-ray screen at baseline (DANTE trial)

| Study or Subgroup | LDCT Events | LDCT Total | Control Events | Control Total | Weight | Risk Ratio M-H, Random, 95% CI |
|---|---|---|---|---|---|---|
| **6.2.1 1 round** | | | | | | |
| UKLS (Davies 2024, median 9.1 y) | 91 | 2028 | 86 | 2027 | 13.5% | 1.06 [0.79, 1.41] |
| **Subtotal (95% CI)** | | **2028** | | **2027** | **13.5%** | **1.06 [0.79, 1.41]** |
| Total events | 91 | | 86 | | | |
| Heterogeneity: Not applicable | | | | | | |
| Test for overall effect: Z = 0.38 (P = 0.70) | | | | | | |
| **6.2.2 4+ rounds** | | | | | | |
| DANTE (Infante 2015, median 8.35 y) | 104 | 1264 | 72 | 1186 | 13.4% | 1.36 [1.01, 1.81] |
| DLCST (Wille 2016, median 9.8 y) | 96 | 2052 | 53 | 2052 | 11.5% | 1.81 [1.30, 2.52] |
| ITALUNG (Paci 2021, median 11.3 y) | 91 | 1613 | 100 | 1593 | 14.2% | 0.90 [0.68, 1.18] |
| LUSI (Maldonado 2021, median 9.77 y) | 90 | 2029 | 74 | 2023 | 12.8% | 1.21 [0.90, 1.64] |
| MILD (Pastorino 2019, 9 years follow-up) | 98 | 2376 | 60 | 1723 | 12.2% | 1.18 [0.86, 1.62] |
| NELSON (de Koning 2020, 10y or to censoring) | 344 | 6583 | 304 | 6612 | 22.5% | 1.14 [0.98, 1.32] |
| **Subtotal (95% CI)** | | **15917** | | **15189** | **86.5%** | **1.22 [1.03, 1.43]** |
| Total events | 823 | | 663 | | | |
| Heterogeneity: Tau² = 0.02; Chi² = 11.38, df = 5 (P = 0.04); I² = 56% | | | | | | |
| Test for overall effect: Z = 2.35 (P = 0.02) | | | | | | |
| **Total (95% CI)** | | **17945** | | **17216** | **100.0%** | **1.19 [1.03, 1.37]** |
| Total events | 914 | | 749 | | | |
| Heterogeneity: Tau² = 0.02; Chi² = 11.93, df = 6 (P = 0.06); I² = 50% | | | | | | |
| Test for overall effect: Z = 2.41 (P = 0.02) | | | | | | |
| Test for subgroup differences: Chi² = 0.69, df = 1 (P = 0.41), I² = 0% | | | | | | |

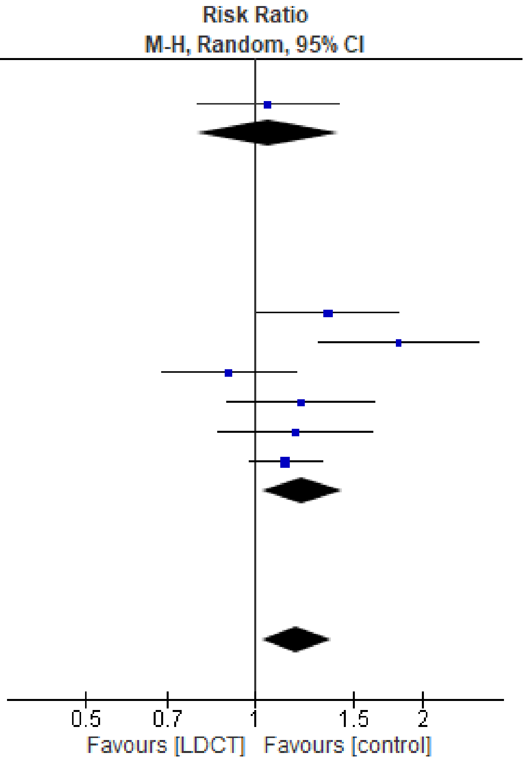

Figure S2.21 Overdiagnosis, by number of screening rounds

| Study or Subgroup | LDCT Events | LDCT Total | Control Events | Control Total | Weight | Risk Ratio M-H, Random, 95% CI |
|---|---|---|---|---|---|---|
| **6.3.1 Annual screening interval** | | | | | | |
| DANTE (Infante 2015, median 8.35 y) | 104 | 1264 | 72 | 1186 | 13.4% | 1.36 [1.01, 1.81] |
| DLCST (Wille 2016, median 9.8 y) | 96 | 2052 | 53 | 2052 | 11.5% | 1.81 [1.30, 2.52] |
| ITALUNG (Paci 2021, median 11.3 y) | 91 | 1613 | 100 | 1593 | 14.2% | 0.90 [0.68, 1.18] |
| LUSI (Maldonado 2021, median 9.77 y) | 90 | 2029 | 74 | 2023 | 12.8% | 1.21 [0.90, 1.64] |
| **Subtotal (95% CI)** | | **6958** | | **6854** | **51.9%** | **1.27 [0.96, 1.68]** |
| Total events | 381 | | 299 | | | |
| Heterogeneity: Tau² = 0.06; Chi² = 10.71, df = 3 (P = 0.01); I² = 72% | | | | | | |
| Test for overall effect: Z = 1.65 (P = 0.10) | | | | | | |
| **6.3.2 >Annual screening interval (on average)** | | | | | | |
| MILD (Pastorino 2019, 9 years follow-up) | 98 | 2376 | 60 | 1723 | 12.2% | 1.18 [0.86, 1.62] |
| NELSON (de Koning 2020, 10y or to censoring) | 344 | 6583 | 304 | 6612 | 22.5% | 1.14 [0.98, 1.32] |
| **Subtotal (95% CI)** | | **8959** | | **8335** | **34.6%** | **1.15 [1.00, 1.31]** |
| Total events | 442 | | 364 | | | |
| Heterogeneity: Tau² = 0.00; Chi² = 0.05, df = 1 (P = 0.82); I² = 0% | | | | | | |
| Test for overall effect: Z = 1.96 (P = 0.05) | | | | | | |
| **6.3.3 One screen only** | | | | | | |
| UKLS (Davies 2024, median 9.1 y) | 91 | 2028 | 86 | 2027 | 13.5% | 1.06 [0.79, 1.41] |
| **Subtotal (95% CI)** | | **2028** | | **2027** | **13.5%** | **1.06 [0.79, 1.41]** |
| Total events | 91 | | 86 | | | |
| Heterogeneity: Not applicable | | | | | | |
| Test for overall effect: Z = 0.38 (P = 0.70) | | | | | | |
| **Total (95% CI)** | | **17945** | | **17216** | **100.0%** | **1.19 [1.03, 1.37]** |
| Total events | 914 | | 749 | | | |
| Heterogeneity: Tau² = 0.02; Chi² = 11.93, df = 6 (P = 0.06); I² = 50% | | | | | | |
| Test for overall effect: Z = 2.41 (P = 0.02) | | | | | | |
| Test for subgroup differences: Chi² = 0.79, df = 2 (P = 0.67), I² = 0% | | | | | | |

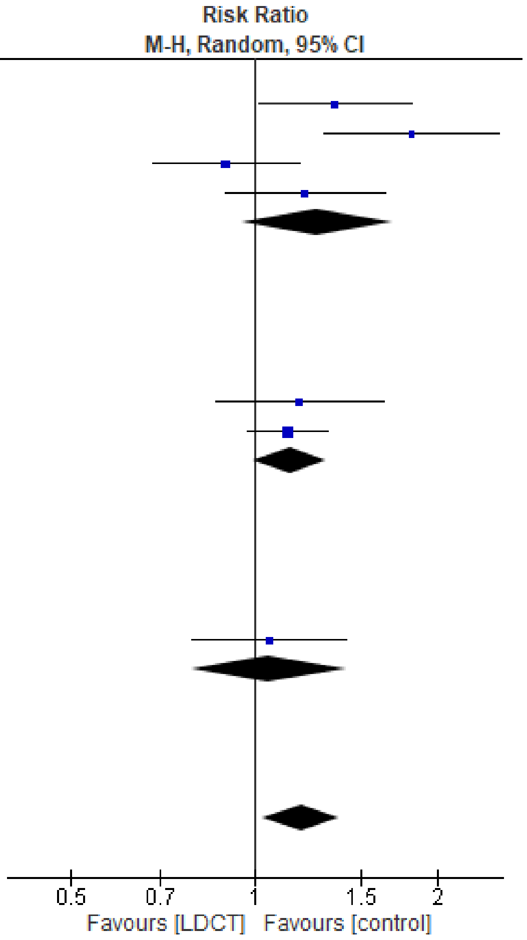

Figure S2.22 Overdiagnosis, by screening interval

| Study or Subgroup | LDCT Events | LDCT Total | Control Events | Control Total | Weight | Risk Ratio M-H, Random, 95% CI |
|---|---|---|---|---|---|---|
| **6.4.1 ≤2 percent 6-year cancer detection rate in control group** | | | | | | |
| DLCST (Wille 2016, median 9.8 y) | 96 | 2052 | 53 | 2052 | 11.5% | 1.81 [1.30, 2.52] |
| **Subtotal (95% CI)** | | **2052** | | **2052** | **11.5%** | **1.81 [1.30, 2.52]** |
| Total events | 96 | | 53 | | | |
| Heterogeneity: Not applicable | | | | | | |
| Test for overall effect: Z = 3.53 (P = 0.0004) | | | | | | |
| **6.4.2 >2 percent 6-year cancer detection rate in control group** | | | | | | |
| DANTE (Infante 2015, median 8.35 y) | 104 | 1264 | 72 | 1186 | 13.4% | 1.36 [1.01, 1.81] |
| ITALUNG (Paci 2021, median 11.3 y) | 91 | 1613 | 100 | 1593 | 14.2% | 0.90 [0.68, 1.18] |
| LUSI (Maldonado 2021, median 9.77 y) | 90 | 2029 | 74 | 2023 | 12.8% | 1.21 [0.90, 1.64] |
| MILD (Pastorino 2019, 9 years follow-up) | 98 | 2376 | 60 | 1723 | 12.2% | 1.18 [0.86, 1.62] |
| NELSON (de Koning 2020, 10y or to censoring) | 344 | 6583 | 304 | 6612 | 22.5% | 1.14 [0.98, 1.32] |
| UKLS (Davies 2024, median 9.1 y) | 91 | 2028 | 86 | 2027 | 13.5% | 1.06 [0.79, 1.41] |
| **Subtotal (95% CI)** | | **15893** | | **15164** | **88.5%** | **1.13 [1.02, 1.25]** |
| Total events | 818 | | 696 | | | |
| Heterogeneity: Tau² = 0.00; Chi² = 4.67, df = 5 (P = 0.46); I² = 0% | | | | | | |
| Test for overall effect: Z = 2.40 (P = 0.02) | | | | | | |
| **Total (95% CI)** | | **17945** | | **17216** | **100.0%** | **1.19 [1.03, 1.37]** |
| Total events | 914 | | 749 | | | |
| Heterogeneity: Tau² = 0.02; Chi² = 11.93, df = 6 (P = 0.06); I² = 50% | | | | | | |
| Test for overall effect: Z = 2.41 (P = 0.02) | | | | | | |
| Test for subgroup differences: Chi² = 7.26, df = 1 (P = 0.007), I² = 86.2% | | | | | | |

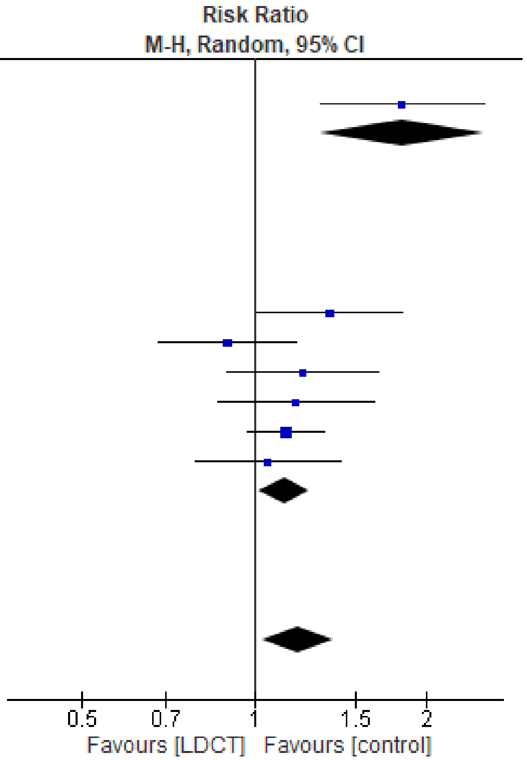

Figure S2.23 Overdiagnosis, by 6-year lung cancer detection rate in comparator (≤2% vs >2%)

| Study or Subgroup | LDCT Events | LDCT Total | Control Events | Control Total | Weight | Risk Ratio M-H, Random, 95% CI |
|---|---|---|---|---|---|---|
| **6.5.1 ≤3 percent 6-year cancer detection rate in control group** | | | | | | |
| DLCST (Wille 2016, median 9.8 y) | 96 | 2052 | 53 | 2052 | 11.5% | 1.81 [1.30, 2.52] |
| LUSI (Maldonado 2021, median 9.77 y) | 90 | 2029 | 74 | 2023 | 12.8% | 1.21 [0.90, 1.64] |
| MILD (Pastorino 2019, 9 years follow-up) | 98 | 2376 | 60 | 1723 | 12.2% | 1.18 [0.86, 1.62] |
| NELSON (de Koning 2020, 10y or to censoring) | 344 | 6583 | 304 | 6612 | 22.5% | 1.14 [0.98, 1.32] |
| UKLS (Davies 2024, median 9.1 y) | 91 | 2028 | 86 | 2027 | 13.5% | 1.06 [0.79, 1.41] |
| **Subtotal (95% CI)** | | **15068** | | **14437** | **72.4%** | **1.23 [1.05, 1.44]** |
| Total events | 719 | | 577 | | | |
| Heterogeneity: Tau² = 0.01; Chi² = 7.24, df = 4 (P = 0.12); I² = 45% | | | | | | |
| Test for overall effect: Z = 2.52 (P = 0.01) | | | | | | |
| **6.5.2 >3 percent 6-year cancer detection rate in control group** | | | | | | |
| DANTE (Infante 2015, median 8.35 y) | 104 | 1264 | 72 | 1186 | 13.4% | 1.36 [1.01, 1.81] |
| ITALUNG (Paci 2021, median 11.3 y) | 91 | 1613 | 100 | 1593 | 14.2% | 0.90 [0.68, 1.18] |
| **Subtotal (95% CI)** | | **2877** | | **2779** | **27.6%** | **1.10 [0.74, 1.65]** |
| Total events | 195 | | 172 | | | |
| Heterogeneity: Tau² = 0.06; Chi² = 4.06, df = 1 (P = 0.04); I² = 75% | | | | | | |
| Test for overall effect: Z = 0.47 (P = 0.64) | | | | | | |
| **Total (95% CI)** | | **17945** | | **17216** | **100.0%** | **1.19 [1.03, 1.37]** |
| Total events | 914 | | 749 | | | |
| Heterogeneity: Tau² = 0.02; Chi² = 11.93, df = 6 (P = 0.06); I² = 50% | | | | | | |
| Test for overall effect: Z = 2.41 (P = 0.02) | | | | | | |
| Test for subgroup differences: Chi² = 0.24, df = 1 (P = 0.62), I² = 0% | | | | | | |

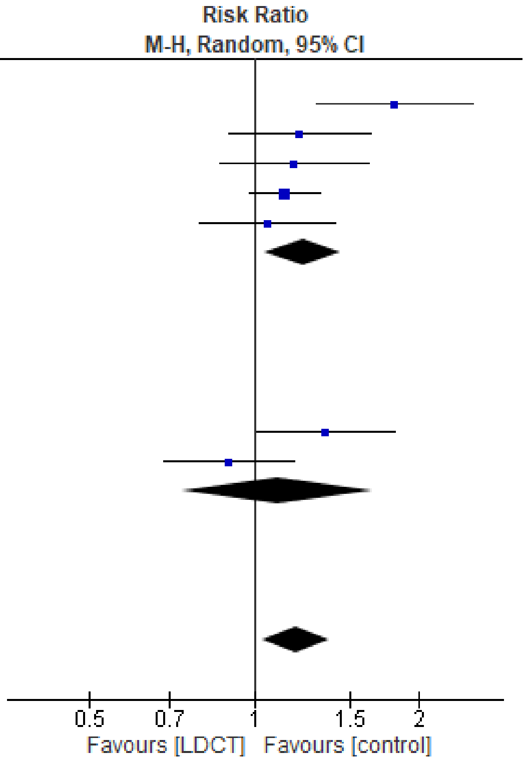

Figure S2.24 Overdiagnosis, by 6-year lung cancer detection rate in comparator (≤3% vs >3%)

| Study or Subgroup | Experimental Events | Experimental Total | Control Events | Control Total | Weight | Risk Ratio M-H, Random, 95% CI |
|---|---|---|---|---|---|---|
| **8.1.1 Men** | | | | | | |
| DANTE (Infante 2015, median 8.35 y) | 104 | 1264 | 72 | 1186 | 15.3% | 1.36 [1.01, 1.81] |
| LUSI (Maldonado 2021, median 9.77 y) | 63 | 1315 | 50 | 1309 | 9.7% | 1.25 [0.87, 1.80] |
| NELSON (de Koning 2020, 10y or to censoring) | 344 | 6583 | 304 | 6612 | 56.7% | 1.14 [0.98, 1.32] |
| UKLS (Field 2021, median 7.3 y) | 62 | 1529 | 57 | 1507 | 10.3% | 1.07 [0.75, 1.53] |
| **Subtotal (95% CI)** | | **10691** | | **10614** | **92.0%** | **1.17 [1.04, 1.32]** |
| Total events | 573 | | 483 | | | |
| Heterogeneity: Tau² = 0.00; Chi² = 1.50, df = 3 (P = 0.68); I² = 0% | | | | | | |
| Test for overall effect: Z = 2.67 (P = 0.008) | | | | | | |
| **8.1.2 Women** | | | | | | |
| LUSI (Maldonado 2021, median 9.77 y) | 27 | 714 | 24 | 716 | 4.4% | 1.13 [0.66, 1.94] |
| UKLS (Field 2021, median 7.3 y) | 24 | 499 | 18 | 520 | 3.6% | 1.39 [0.76, 2.53] |
| **Subtotal (95% CI)** | | **1213** | | **1236** | **8.0%** | **1.24 [0.83, 1.85]** |
| Total events | 51 | | 42 | | | |
| Heterogeneity: Tau² = 0.00; Chi² = 0.26, df = 1 (P = 0.61); I² = 0% | | | | | | |
| Test for overall effect: Z = 1.05 (P = 0.30) | | | | | | |
| **Total (95% CI)** | | **11904** | | **11850** | **100.0%** | **1.18 [1.05, 1.32]** |
| Total events | 624 | | 525 | | | |
| Heterogeneity: Tau² = 0.00; Chi² = 1.82, df = 5 (P = 0.87); I² = 0% | | | | | | |
| Test for overall effect: Z = 2.86 (P = 0.004) | | | | | | |
| Test for subgroup differences: Chi² = 0.06, df = 1 (P = 0.80), I² = 0% | | | | | | |

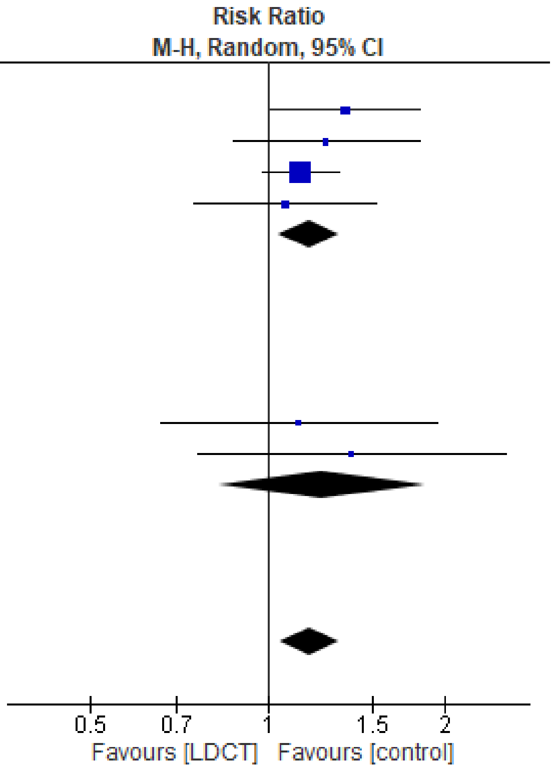

Figure S2.25 Overdiagnosis, by sex

| Study or Subgroup | log[Risk Ratio] | SE | Weight | Risk Ratio IV, Random, 95% CI |
|---|---|---|---|---|
| **11.4.1 LUSI** | | | | |
| LUSI (Maldonado 2021, median 9.77 y) | 0.1009 | 0.2303 | 52.5% | 1.11 [0.70, 1.74] |
| **Subtotal (95% CI)** | | | **52.5%** | **1.11 [0.70, 1.74]** |
| Heterogeneity: Not applicable | | | | |
| Test for overall effect: Z = 0.44 (P = 0.66) | | | | |
| **11.4.2 UKLS** | | | | |
| UKLS (Field 2021, median 7.3 y) | -0.2616 | 0.2443 | 47.5% | 0.77 [0.48, 1.24] |
| **Subtotal (95% CI)** | | | **47.5%** | **0.77 [0.48, 1.24]** |
| Heterogeneity: Not applicable | | | | |
| Test for overall effect: Z = 1.07 (P = 0.28) | | | | |
| **Total (95% CI)** | | | **100.0%** | **0.93 [0.65, 1.33]** |
| Heterogeneity: Tau² = 0.01; Chi² = 1.17, df = 1 (P = 0.28); I² = 14% | | | | |
| Test for overall effect: Z = 0.39 (P = 0.69) | | | | |
| Test for subgroup differences: Chi² = 1.17, df = 1 (P = 0.28), I² = 14.2% | | | | |

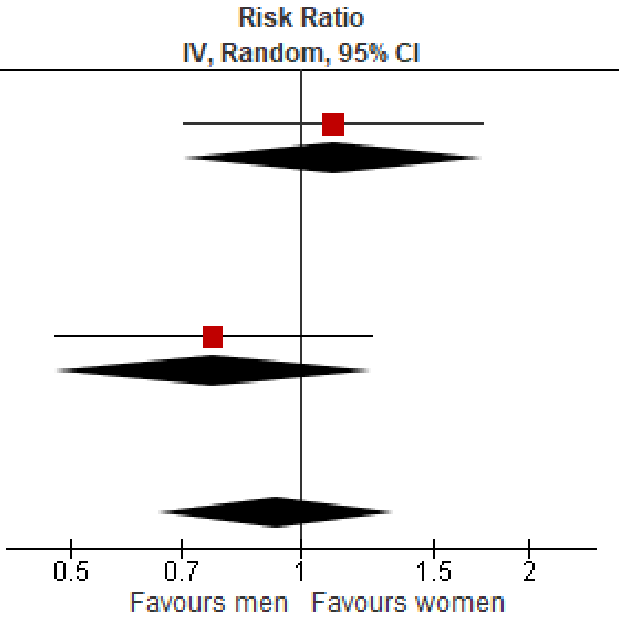

Figure S2.26 Overdiagnosis, within study ratio of ratios by sex

Table S2.8 Additional overdiagnosis definitions and findings

| Number of studies; Number enrolled/randomized to CT screening | Overdiagnosis calculations (excess incidence or fraction overdiagnosed) |
|---|---|
| Overdiagnosis: excess incidence from screening | |
| Trials: 7 studies; N=35,161 (9-11, 14, 15, 17, 19) | Excess incidence:<br>NELSON: -1.51<br>DANTE: -0.85<br>LUSI: -0.55<br>ITALUNG: -3.92<br>DLCST: 0.73<br>UKLS: -1.63<br>MILD: -0.49 |
| Overdiagnosis: excess incidence of lung cancer from screening among those having cancer diagnosed in the screening arm | |
| Trials: 7 studies; N=35,161 (9-11, 14, 15, 17, 19) | Excess incidence:<br>NELSON: -0.000044<br>DANTE: -0.000082<br>LUSI: -0.000061<br>ITALUNG: -0.00043<br>DLCST: 0.000076<br>UKLS: -0.00019<br>MILD: -0.000050 |
| Overdiagnosis: excess incidence of lung cancer from screening among those having cancer diagnosed through screening in the screening arm | |
| Trials: 7 studies; N=35,161 (9-11, 14, 15, 17, 19) | Excess incidence:<br>NELSON: -0.000075<br>DANTE: -0.00013<br>LUSI: -0.000088<br>ITALUNG: -0.0010<br>DLCST: 0.00011<br>UKLS: -0.00039<br>MILD: -0.000070 |
| Overdiagnosis: overdiagnosis of screen-detected cancer | |
| Trials: 7 studies; N=35,161 (9-11, 14, 15, 17, 19) | Fraction of overdiagnosed cases:<br>**Weighted average (multiplied by 100%): 24% (range 20% to 63%)**<br>NELSON: 0.20<br>DANTE: 0.41<br>LUSI: 0.25<br>ITALUNG: -0.27<br>DLCST: 0.63<br>UKLS: 0.26<br>MILD: 0.22 |

Table S2.9 CT screening: Summary of findings for important outcomes

| Number of studies; Number enrolled/randomized to CT screening | Absolute events (95% CI) per 1000 | Certainty of evidence (GRADE[i]) | What happens? |
|---|---|---|---|
| Proportion with a false positive (threshold 75 per 1000) | | | |
| Round 1 (Fig. S2.27):<br>Trials: 11 studies; N=48,888<br>Observational: 18 studies; N=40,757 | 190 [160, 220] | **High**<br>⨁⨁⨁⨁[a] | Among people 50-74 years of age who are current or former smokers, screening with low-dose CT 3-4 times causes at least 75 false positives per 1000 screened.<br><br>Our certainty would be moderate for effects of at least 225 false positives per 1000 screened. |
| Over 2/3 rounds (Fig. S2.28):<br>Trials: 2 studies; N=28,382<br>Observational: 8 studies; N=55,308 | 310 [180, 440] | | |
| Over 4/5 rounds (Fig. S2.29):<br>Trials: 2 studies; N=2,877<br>Observational: 1 study; N=1,267 | 350 [220, 480] | | |
| Proportion with incidental findings (threshold 100 or 150 per 1000) | | | |
| Round 1 (Fig. S2.30):<br>Trials: 0 studies<br>Observational: 13 studies; N=14,549 | 550 [370, 730] | **High**<br>⨁⨁⨁⨁[b] | Among people 50-74 years of age who are current or former smokers, screening with low-dose CT 3-4 times causes at least 150 incidental findings per 1000 screened.<br><br>Our certainty would be moderate for effects of at least 450 incidental findings per 1000 screened. |
| Over 2/3 rounds (Fig. S2.31):<br>Trials: 1 study; N=26,722<br>Observational: 0 studies | 570 [560, 570] | | |
| Over 4/5 rounds (Fig. S2.32):<br>Trials: 2 studies; N=2,877<br>Observational: 1 study; N=519 | 400 [50, 750] | | |
| Proportion with clinically significant incidental findings (threshold 50 or 100 per 1000) | | | |
| Round 1 (Fig. S2.33):<br>Trials: 2 studies; N=27,582<br>Observational: 26 studies; N=81,843 | 220 [180, 260] | **Threshold: 50 per 1000**<br>**High**<br>⨁⨁⨁⨁ [c]<br><br>**Threshold: 100 per 1000:**<br>**Moderate** | Among people 50-74 years of age who are current or former smokers, screening with low-dose CT 3-4 times |
| Over 2/3 rounds (Fig. S2.34):<br>Trials: 1 study; N=26,722 | 340 [330, 340] | | |

| | | | |
|---|---|---|---|
| Observational: 0 studies | | ⊕⊕⊕⊖[c]<br>Due to inconsistency | causes at least 50, and probably causes at least 100 clinically significant incidental findings per 1000 screened. |
| Over 4/5 rounds (Fig. S2.35):<br>Trials: 0 studies<br>Observational: 4 studies; N=12,026 | 80 [30, 120] | | |
| Proportion with major complications from invasive testing (lung cancers and false positives) (threshold 2.5 per 1000) | | | |
| Round 1 (Fig. S2.36):<br>Trials: 4 studies; N=9,829<br>Observational: 7 studies; N=19,795 | 0.70 [0.00, 2.40] | **Low**<br>⊕⊕⊖⊖[d]<br>Due to very serious inconsistency | Among people 50-74 years of age who are current or former smokers, screening with low-dose CT 3-4 times may not cause at least 2.5 major complications from invasive testing among those with and without lung cancer per 1000 screened. |
| Over 2/3 rounds (Fig. S2.37):<br>Trials: 1 study; N=26,722<br>Observational: 4 studies; N=48,344 | 3.00 [1.30, 5.10] | | |
| Over 4/5 rounds (Fig. S2.38):<br>Trials: 2 studies; N=8,196<br>Observational: 1 study; N=2,471 | 1.80 [0.00, 7.90] | | |
| Proportion with major complications from invasive testing among false positives (threshold 2.5 per 1000) | | | |
| Round 1 (Fig. S2.39):<br>Trials: 5 studies; N=10,751<br>Observational: 6 studies; N=13,040 | 0.00 [0.00, 0.30] | **High**<br>⊕⊕⊕⊕[e] | Among people 50-74 years of age who are current or former smokers, screening with low-dose CT 3-4 times does not cause at least 2.5 major complications from invasive testing for those without cancer per 1000 screened.<br><br>Our certainty would be moderate for effects of as low as 1 per 1000 screened. |
| Over 2/3 rounds (Fig. S2.40):<br>Trials: 1 study; N=26,722<br>Observational: 2 studies; N=2,259 | 0.00 [0.00, 0.20] | | |
| Over 4/5 rounds (Fig. S2.41):<br>Trials: 1 study; N=6,583<br>Observational: 1 study; N=2,471 | 0.10 [0.00, 0.40] | | |
| Proportion of deaths from invasive testing among false positives (threshold 0.1 per 1000) | | | |
| Round 1 (Fig. S2.42):<br>Trials: 3 studies; N=3,309 | 0.00 [0.00, 0.00] | **Moderate** | Among people 50-74 years of age |

| | | | | | |
|---|---|---|---|---|---|
| Observational: 3 studies; N=44,123 | | | ⊕⊕⊕⊖[f]<br>Due to inconsistency | | who are current or former smokers, screening with low-dose CT 3-4 times may not cause 0.1 deaths from invasive testing among those without cancer per 1000 screened. |
| Over 2/3 rounds (Fig. S2.43):<br>Trials: 1 study; N=26,722<br>Observational: 1 study; N=2,003 | 0.10 [0.00, 0.30] | | | | |
| Over 4/5 rounds (Fig. S2.44):<br>Trials: 1 study; N=1,264 | 0.00 [0.00, 2.91] | | | | |
| **Number of studies; Number enrolled/randomized to CT screening** | **Mean difference (95% CI)** | | Findings | **Certainty of evidence (GRADE[i])** | **What happens?** |
| Health-related quality of life (thresholds: small=5 to <10% difference; moderate=10 to 20% difference) | | | | | |
| Follow-up: 6 months and 1.5 years<br><br>N=3,937 (subsamples of 2 RCTs) | Difference in change scores between groups:<br><br>NELSON [(29); N=1,125]: mean scores in each arm were adjusted for age, gender, education, smoking status, and smoking pack-years:<br>EQ-VAS (scale 0-100): 1.39 (-0.46, 3.24); p=0.14<br>**1.4% increase**<br>SF-12 PCS (scale 0-100): 1.05 (-0.13, 2.23); p=0.08<br>**1.1% increase**<br>SF-12 MCS (scale 0-100): 0.87 (-0.58, 2.32); p=0.24<br>**0.9% increase**<br><br>NLST [(27); N=2,812]: adjusted for days since baseline evaluation, baseline age, sex, years of education, marital status, smoking status, race, ethnicity, number of prior suspicious for lung cancer screens, number of prior SIFs screens<br>SF-36 PCS (scale 0-100): 0.50 (-0.06, 1.07)<br>**0.5% increase**<br>SF-36 MCS (scale 0-100): 0.07 (-0.61, 0.74)<br>**0.1% increase** | | Subsamples from the NELSON (N=1,125) and NLST (N=2,812) trials found little-to-no difference between groups in health-related quality of life scores at 6-month and 1.5-year follow-up after randomization | **Very low**<br>⊕⊖⊖⊖[g]<br>Due to ROB, and very serious indirectness | Among people 50-74 years of age who are current or former smokers, we are very uncertain whether screening with low-dose CT 3-4 times impacts health-related quality of life. |
| Psychosocial consequences of screening (thresholds: small=5 to <10% difference; moderate=10 to 20% difference) | | | | | |
| Participating in screening program<br><br>3 studies (N=9,205) | NELSON (29)<br>Spielberger State-trait anxiety (scale 20-80)<br>N=1,125; Comparative with unscreened control arm | Before randomization and screening:<br>-0.48 [-1.64, 0.68]<br>**0.8% difference**<br><br>After first screening round (6 months into 2nd screening round), | Three studies (N=9,205) comparing screened and unscreened participants at different timepoints during the screening program found little-to-no difference across measures of psychosocial | **Moderate**<br>⊕⊕⊕⊖[h]<br>Due to indirectness | Among people 50-74 years of age who are current or former smokers, screening with low-dose CT 3-4 times probably causes |

| | | | | | |
|---|---|---|---|---|---|
| | ROB, before randomization: Some concerns<br>ROB, after first screening round: High | difference in change score:<br>-0.27 [-1.47, 0.93]<br>**0.5% decrease** | harm. The applicability of the findings to 3-4 rounds of screening is limited. | | little-to-no psychosocial harm. |
| | NELSON (29)<br>Impact of events scale - LC specific (scale 0-75)<br>N=1,125; Comparative with unscreened control arm<br><br>ROB, before randomization: Some concerns<br>ROB, after first screening round: High | Before randomization and screening: 0.03 [-0.88, 0.94]<br>**0.0% difference**<br><br>After first screening round (6 months into 2nd screening round), difference in change score:<br>-0.34 [-1.29, 0.61]<br>**0.5% decrease** | | | |
| | UKLS (44)<br>Cancer worry (scale 6-24)<br>N=4,037; Comparative with unscreened control arm<br><br>ROB, at both timepoints: Some concerns | After randomization (allocation unknown) and before screening:<br>0.01 [-0.07, 0.09]<br>**0.1% difference**<br><br>2 weeks after screening or 2-week follow-up in unscreened arm (N=3,232), difference in change score:<br>0.11 [-0.00, 0.20]<br>**0.6% increase**<br><br>Up to 2 years from screening (N=2,855), difference in change score:<br>0.10 [-0.00, 0.20]<br>**0.6% increase** | | | |
| | DLCST (35)<br>Consequences of LC subscales: anxiety (scale 0-21), behavior (scale 0-21), dejection (scale 0-18), negative impact on sleep (scale 0-12) | Before each screening round:<br><br>Anxiety:<br>Round 1 (n=3,988):<br>0.14 [0.00, 0.28]<br>**0.7% increase**;<br>Round 2 (n=3,352):<br>0.27 [0.07, 0.47]<br>**1.3% increase**; | | | |

| | | |
|---|---|---|
| | N=4,043; Comparative with unscreened control arm<br><br>ROB, at all timepoints: Some concerns | Round 3 (n=3,555):<br>0.34 [0.14, 0.54]<br>**1.6% increase**;<br>Round 4 (n=3,017):<br>0.26 [0.03, 0.49]<br>**1.2% increase**;<br>Round 5 (n=3,185):<br>0.65 [0.41, 0.89]<br>**3.1% increase**<br><br>Behavior:<br>Round 1 (n=4,030):<br>0.13 [0.01, 0.25]<br>**0.7% increase**;<br>Round 2 (n=3,337):<br>0.33 [0.12, 0.55]<br>**1.6% increase**;<br>Round 3 (n=3,540):<br>0.61 [0.38, 0.84]<br>Round 4 (n=3,010):<br>0.40 [0.16, 0.64]<br>**1.9% increase**;<br>Round 5 (n=3,180):<br>0.72 [0.45, 1.00]<br>**3.4% increase**<br><br>Dejection:<br>Round 1 (n=4,035):<br>0.15 [0.02, 0.28]<br>**0.8% increase**;<br>Round 2 (n=3,377):<br>0.31 [0.10, 0.51]<br>**1.7% increase**;<br>Round 3 (n=3,573):<br>0.45 [0.24, 0.67]<br>**2.5% increase**;<br>Round 4 (n=3,032):<br>0.28 [0.05, 0.52]<br>**1.5% increase**;<br>Round 5 (n=3,195):<br>0.73 [0.47, 0.99]<br>**4.1% increase**<br><br>Negative impact on sleep:<br>Round 1 (n=4,043):<br>0.08 [-0.02, 0.18]<br>**0.7% increase**; |

| | | | | | |
|---|---|---|---|---|---|
| | | Round 2 (n=3,389):<br>0.22 [0.04, 0.40]<br>**1.8% increase**;<br>Round 3 (n=3,577):<br>0.43 [0.24, 0.61]<br>**3.6% increase**;<br>Round 4 (n=3,029):<br>0.34 [0.14, 0.55]<br>**2.8% increase**;<br>Round 5 (n=3,198):<br>0.78 [0.53, 1.03]<br>**6.5% increase**<br><br>*The adjusted analyses had very similar results to the unadjusted analysis, therefore only the unadjusted analysis was reported | | | |
| Undergoing CT scan (after screening, before results are known)<br><br>2 non-randomized studies (N=1,645) | LSUT (49)<br>LC worry (scale 7-29)<br>N=1,014; Comparative with unscreened community sample, matched for age and smoking history<br><br>ROB: High | 1.76 [1.27, 2.25]<br>**8.0% increase** | Study data found there may be little-to-no difference before and after screening, although there was some inconsistency. One study (N=1,014) comparing a screening group to an unscreened community sample, matched for age and smoking history found a small increase in lung cancer worry right after receiving a CT scan. Among those being screened in this study (N=631), and in the NELSON trial (N=630), there was little-to-no change across various measures from before to after screening. | **Low**<br>⊕⊕⊖⊖[i]<br>Due to ROB and inconsistency | Among people 50-74 years of age who are current or former smokers, undergoing a low-dose CT scan may cause little-to-no psychosocial harms. |
| | LSUT (49)<br>LC worry (scale 7-29)<br>N=631; Pre-post<br><br>ROB: High | -0.35 [-0.66, -0.04]<br>**1.6% decrease** | | | |
| | NELSON (31)(only indeterminate and negative)<br>Spielberger State-trait anxiety (scale 20-80)<br>N=630; Pre-post<br><br>ROB: Some concerns | -0.49 [-1.17, 0.19]<br>**0.8% decrease**<br><br>Subgroup (van den Bergh 2008, n=324):<br>Men: -1.40 [-2.58, -0.22]<br>Women: -1.3 (-2.62, 0.02] | | | |
| | NELSON (31)(only indeterminate and negative)<br>Impact of events scale - LC specific (scale 0-75)<br>N=630; Pre-post | 0.40 [-0.19, 0.99]<br>**0.5% increase**<br><br>Subgroup (van den Bergh 2008, n=324):<br>Men: -0.90 [-2.09, 0.29] | | | |

| | | | | | |
|---|---|---|---|---|---|
| | ROB: Some concerns | Women: -1.40 [-3.06, 0.26] | | | |
| Positive screen (before repeat CT or diagnostic results known)<br><br>7 non-randomized studies (N=4,794) | NELSON (29)<br>Spielberger State-trait anxiety (scale 20-80)<br>N=595; Comparative with unscreened control arm<br><br>ROB: Some concerns | To recall:<br>0.80 [-1.04, 2.64]<br>**1.3% increase** | Data from seven non-randomized studies (N=4,794) suggested that receiving a positive screening result may cause a small psychosocial harm, particularly when measured by scales specific to lung cancer. | **Low**<br>⊕⊕⊖⊖[j]<br>Due to ROB, and inconsistency | Among people 50-74 years of age who are current or former smokers, undergoing a low-dose CT scan and receiving a positive screening result may cause small psychosocial harm. |
| | NELSON (29)<br>Impact of events scale - LC specific (scale 0-75)<br>N=595; Comparative with unscreened control arm<br><br>ROB: Some concerns | To recall:<br>3.78 [2.32, 5.24]<br>**5.0% increase** | | | |
| | UKLS (44)<br>Cancer worry (scale 6-24)<br>N=2,415; Comparative with unscreened control arm<br><br>ROB: Some concerns | 0.89 [0.72, 1.06]<br>**4.9% increase** | | | |
| | DLCST (34)<br>Consequences of LC subscales: anxiety (scale 0-21), behavior (scale 0-21), dejection (scale 0-18), negative impact on sleep (scale 0-12)<br>N=295; Comparative with unscreened control arm, matched 1:2 on sex, age, and time of clinic visit<br><br>ROB: Some concerns | Anxiety:<br>FPs: 2.77 (1.63 to 3.92)<br>**13.2% increase**;<br>TPs: 1.89 (0.01 to 3.78)<br>**9.0% increase**;<br>Behavior:<br>FPs: 1.21 (0.19 to 2.23)<br>**5.8% increase**;<br>TPs: 0.65 (0.65 to 1.94)<br>**3.1% increase**;<br>Dejection:<br>FPs: 2.63 (1.53 to 3.74)<br>**14.6% increase**;<br>TPs: 2.20 (0.28 to 4.12)<br>**12.2% increase**;<br>Negative impact on sleep:<br>FPs: 0.65 (-0.14 to 1.44)<br>**5.4% increase**; | | | |

| | | |
|---|---|---|
| | | TPs: 0.47 (-0.61 to 1.54)<br>**3.9% increase**<br>All MDs are adjusted |
| | NELSON (29)<br>Spielberger State-trait anxiety (scale 20-80)<br>N=135; Pre-post<br><br>ROB: Some concerns | To recall:<br>1.27 [-0.34, 2.88]<br>**2.1% increase** |
| | NELSON (29)<br>Impact of events scale - LC specific (scale 0-75)<br>N=135; Pre-post<br><br>ROB: Some concerns | To recall:<br>3.80 [2.55, 5.05]<br>**5.1% increase** |
| | NLST (27)<br>Short form-36 mental component subscale (scale 0 to 100)<br>N=617 (CT arm); N=266 (CXR arm); Pre-post<br><br>ROB: Some concerns | LDCT arm: -1.56 [-2.39, -0.73]<br>**1.6% decrease;**<br>CXR arm: -0.99 [-2.17, 0.19]<br>**1.0% decrease**<br><br>**Higher score = better mental health** |
| | (50)Consequences of LC subscales:<br>anxiety (scale 0-21), behavior (scale 0-21), dejection (scale 0-18), negative impact on sleep (scale 0-12);<br>N=203; Pre-post<br>Adjusted for age, sex, smoking status, pack-years, education, employment, PLCOm2012 score and baseline responses<br><br>ROB: High | To recall (3 months):<br>Anxiety: 0.29 [-0.22, 0.80]<br>**1.38% increase**<br>Behavior:<br>0.53 [0.03, 1.02]<br>**2.52% increase**<br>Dejection:<br>0.30 [-0.19, 0.78]<br>**1.67% increase**<br>Negative impact on sleep:<br>0.29 [-0.08, 0.66]<br>**2.42% increase**<br><br>To recall (6 months):<br>Anxiety:<br>0.45 [-0.28, 1.17]<br>**2.14% increase**<br>Behavior:<br>0.67 [-0.36, 1.71]<br>**3.19% increase** |

| | | | | | |
|---|---|---|---|---|---|
| | | Dejection:<br>0.44 [-0.27, 1.15]<br>**2.44% increase**<br>Negative impact on sleep:<br>0.07 [-0.75, 0.89]<br>**0.58% increase**<br>All MDs are adjusted | | | |
| | (51) State Trait Anxiety: State anxiety subscore (scale 20-80)<br>N=165; Pre-post<br><br>ROB: Some concerns | 3.65 [1.63, 5.67]<br>**6.1% increase** | | | |
| | (51) State Trait Anxiety: Trait anxiety subscore (scale 20-80)<br>N=165; Pre-post<br><br>ROB: Some concerns | 1.08 [-0.73, 2.89]<br>**1.8% increase** | | | |
| | (51) Fear of LC (adapted from psychosocial consequences questionnaire) (scale 3-15)<br>N=165; Pre-post<br><br>ROB: Some concerns | 0.57 [0.15, 0.99]<br>**4.8% increase** | | | |
| | (52)<br>Spielberger State-trait anxiety (scale 20-80)<br>N=238; Pre-post<br><br>ROB: Some concerns | 3.28 [-0.42, 6.97]<br>**5.5% increase**<br><br>Adjusted p-value:<br>p>0.05 | | | |
| Positive screen, requiring diagnostic follow-up (before diagnostic results known)<br><br>2 non-randomized studies (N=1,651) | UKLS (44)<br>Cancer worry (scale 6-24)<br>N=1,627; Comparative with unscreened control arm<br><br>ROB: Some concerns | 3.28 [2.52, 4.04]<br>**18.2% increase** | Data from two non-randomized studies (N=1,651) suggested that receiving a positive screening result may cause moderate psychosocial harm, particularly when measured by scales specific to lung cancer. | Small harm:<br>Moderate<br>⨁⨁⨁⊖[k]<br>Due to ROB<br><br>Moderate harm:<br>Low<br>⨁⨁⊖⊖[k]<br>Due to ROB, and inconsistency | Among people 50-74 years of age who are current or former smokers, undergoing a low-dose CT scan and receiving a positive screening result requiring diagnostic follow-up probably causes small, and |
| | (51)<br>State Trait Anxiety: State anxiety subscore (scale 20-80) | 5.70 [0.01, 11.39]<br>**9.5% increase** | | | |

| | | | | | |
|---|---|---|---|---|---|
| | N=24; Pre-post<br><br>ROB: Some concerns | | | | may cause moderate psychosocial harm. |
| | Byrne, 2008<br>State Trait Anxiety: Trait anxiety subscore (scale 20-80)<br>N=24; Pre-post<br><br>ROB: Some concerns | 2.70 [-1.76, 7.16]<br>**4.5% increase** | | | |
| | (51)<br>Fear of LC (adapted from psychosocial consequences questionnaire) (scale 3-15)<br>N=24; Pre-post<br><br>ROB: Some concerns | 2.10 [1.69, 2.51]<br>**17.5% increase** | | | |
| False positive screening result (≤6 months after diagnostic results known)<br><br>6 non-randomized studies (N=1,860) | DLCST (34)<br>Consequences of LC subscales: anxiety (scale 0-21), behavior (scale 0-21), dejection (scale 0-18), negative impact on sleep (scale 0-12)<br>N=277; Comparative with unscreened control arm, matched 1:2 on sex, age, and time of clinic visit<br><br>ROB: Some concerns | Anxiety: -0.25 [-1.25, 0.75]<br>**1.2% decrease**;<br>Behavior: -0.09 [-1.34, 1.16]<br>**0.4% decrease**;<br>Dejection: -0.09 [-1.21, 1.04]<br>**0.5% decrease**;<br>Negative impact on sleep: -0.10 [-1.01, 0.81]<br>**0.8% decrease**<br><br>All MDs are adjusted | Findings across six non-randomized studies (N=1,860) indicated little-to-no difference on average, but there was inconsistency with two of the studies finding at least a small amount of harm among a sample that likely included some lung cancer patients. | Low<br>⊕⊕⊖⊖<br>Due to ROB, and inconsistency[l] | Among people 50-74 years of age who are current or former smokers, undergoing a low-dose CT scan and receiving a false positive screening result may cause little-to-no psychosocial harm under 6 months after the diagnostic results are known. |
| | LSUT (49)<br>LC worry (scale 7-29)<br>N=514; Comparative with unscreened community sample, matched on age and smoking history. Unclear if some participants had lung cancer<br><br>ROB: High | Any FPs:<br>3.65 [2.71, 4.59]<br>**16.6% increase** | | | |
| | LSUT (49) | Any FPs: | | | |

| | | |
|---|---|---|
| | LC worry (scale 7-29)<br>N=131; Pre-post<br><br>ROB: High | 1.71 [0.93, 2.49]<br>**7.8% increase** |
| | NLST (27)<br>Short form-36 mental component subscale (scale 0 to 100)<br>N=489 (CT arm); N=214 (CXR arm); Pre-post<br><br>ROB: High | LDCT arm: -1.38 [-2.33, -0.43]<br>**1.4% decrease**<br><br>CXR arm: -0.55 [-1.57, 0.47]<br>**0.6% decrease**<br><br>**Higher score = better mental health** |
| | (52)<br>Spielberger State-trait anxiety (scale 20-80)<br>N=168; Pre-post<br>2.8% had lung cancer<br><br>ROB: High | 2.42 [-1.14, 5.99]<br>**4.0% increase** |
| | (50)<br>Consequences of LC subscales:<br>anxiety (scale 0-21), behavior (scale 0-21), dejection (scale 0-18), negative impact on sleep (scale 0-12);<br>N=33; Pre-post<br>Adjusted for age, sex, smoking status, pack-years, education, employment, PLCOm2012 score and baseline responses.<br>Unclear if some participants had lung cancer<br><br>ROB: High | High malignancy risk (after biopsy results):<br>Anxiety:<br>0.50 [-1.58, 2.58]<br>**2.38% increase**<br>Behavior:<br>1.22 [-0.44, 2.87]<br>**5.81% increase**<br>Dejection:<br>-0.39 [-1.47, 0.70]<br>**2.17% decrease**<br>Negative impact on sleep:<br>0.89 [-0.08, 1.85]<br>**7.42% increase**<br><br>Suspicious for lung cancer (after biopsy results):<br>Anxiety:<br>0.36 [-0.26, 0.97]<br>**1.71% increase**<br>Behavior:<br>1.25 [-0.15, 2.66]<br>**5.95% increase**<br>Dejection: |

| | | | | | |
|---|---|---|---|---|---|
| | | 0.47 [-0.10, 1.04]<br>**2.61% increase**<br>Negative impact on sleep:<br>1.52 [0.28, 2.76]<br>**12.67% increase** | | | |
| | (51)<br>State Trait Anxiety: State anxiety subscore (scale 20-80)<br>N=165; Pre-post<br><br>ROB: Some concerns | Any FPs:<br>2.48 [0.57, 4.39]<br>**4.1% increase** | | | |
| | (51)<br>State Trait Anxiety: Trait anxiety subscore (scale 20-80)<br>N=165; Pre-post<br><br>ROB: Some concerns | Any FPs:<br>0.22 [-1.57, 2.01]<br>**0.4% increase** | | | |
| | (51)<br>Fear of LC (adapted from psychosocial consequences questionnaire) (scale 3-15)<br>N=165; Pre-post<br><br>ROB: Some concerns | Any FPs:<br>0.06 [-0.35, 0.47]<br>**0.5% increase** | | | |
| False positive screening result (>6 months after diagnostic results known)<br><br><br>6 non-randomized studies (N=4,553) | DLCST (34)<br>Consequences of LC subscales: anxiety (scale 0-21), behavior (scale 0-21), dejection (scale 0-18), negative impact on sleep (scale 0-12)<br>N=277; Comparative with unscreened control arm, matched 1:2 on sex, age, and time of clinic visit<br><br>ROB: Some concerns | Any FPs:<br>Anxiety: -0.26 [-1.16, 0.65]<br>**1.2% decrease**;<br>Behavior: -0.07 [-1.30, 1.17]<br>**0.3% decrease**;<br>Dejection: -0.01 [-1.04, 1.03]<br>**0.1% decrease**;<br>Negative impact on sleep: 0.11 [-0.68, 0.90]<br>**0.9% increase** | Findings across 6 non-randomized studies (N=4,553) consistently showed little-to-no impact on psychosocial harms. | Moderate<br>⊕⊕⊕⊖[m]<br>Due to ROB | Among people 50-74 years of age who are current or former smokers, undergoing a low-dose CT scan and receiving a false positive screening result probably cause little-to-no psychosocial harm over 6 months after the diagnostic results are known. |
| | NELSON (29)<br>Spielberger State-trait anxiety (scale 20-80) | To recall:<br>Difference in change score: | | | |

| | | |
|---|---|---|
| | N=595; Comparative with unscreened control arm<br><br>ROB: High | 0.29 [-1.55, 2.13]<br>**0.5% increase** |
| | NELSON (29)<br>Impact of events scale - LC specific (scale 0-75)<br>N=595; Comparative with unscreened control arm<br><br>ROB: High | To recall:<br>Difference in change score:<br>0.54 [-0.89, 1.97]<br>**0.7% increase** |
| | UKLS (44)<br>Cancer worry (scale 6-24)<br>N=1,747; Comparative with unscreened control arm<br><br>ROB: High | Any FPs:<br>-0.07 [-0.26, 0.12]<br>**0.4% decrease** |
| | (53)<br>Consequences of LC (scores dichotomized: 0=no consequences; >0=psychosocial consequences)<br>N=1,523; Comparative with matched by CT date, gender and site<br><br>ROB: Some concerns | Odds ratio (95% CI):<br>Sense of dejection: 1.35 [1.06, 1.72]<br>Anxiety: 1.22 [0.97, 1.54]<br>Behavioral: 1.21 [0.95, 1.54]<br>Sleep: 1.17 [0.93, 1.45] |
| | NELSON (29)<br>Spielberger State-trait anxiety (scale 20-80)<br>N=135; Pre-post<br><br>ROB: Some concerns | To recall:<br>-0.04 [-1.64, 1.56]<br>**0.1% decrease** |
| | NELSON (29)<br>Impact of events scale - LC specific (scale 0-75)<br>N=135; Pre-post<br><br>ROB: Some concerns | To recall:<br>0.55 [-0.68, 1.78]<br>**0.7% increase** |
| | (52)<br>Spielberger State-trait anxiety (scale 20-80) | Any FPs:<br>1.79 [-0.62, 4.19]<br>**3.0% increase** |

| | | |
|---|---|---|
| | N=246; Pre-post<br><br>ROB: Some concerns | |
| | (51)<br>State Trait Anxiety: State anxiety subscore (scale 20-80)<br>N=165; Pre-post<br><br>ROB: Some concerns | Any FPs:<br>1.13 [-0.92, 3.18]<br>**1.9% increase** |
| | (51)<br>State Trait Anxiety: Trait anxiety subscore (scale 20-80)<br>N=165; Pre-post<br><br>ROB: Some concerns | Any FPs:<br>-0.18 [-2.08, 1.72]<br>**0.3% decrease** |
| | (51)<br>Fear of LC (adapted from psychosocial consequences questionnaire) (scale 3-15)<br>N=165; Pre-post<br><br>ROB: Some concerns | Any FPs:<br>0.02 [-0.40, 0.44]<br>**0.2% increase** |

**[ǂ]GRADE ratings (all outcomes started at high certainty):**

[a] False positives: ***No serious concerns about risk of bias:*** 6 studies at round one, two studies at 2-3 rounds, and all studies at 4-5 rounds of screening were at high risk of bias, but findings did not change when excluded in sensitivity analysis. ***No serious concerns about inconsistency:*** Findings were consistent across rounds of screening and subgroup findings. ***No serious concerns about indirectness:*** The study data was direct for this outcome; findings are applicable to contemporary nodule management approaches, which are less sensitive than used in the NLST trial. ***No serious concerns about imprecision:*** The confidence intervals are precise for our threshold of effects over 3-4 screening rounds. ***No serious concerns about reporting bias:*** No serious concerns about reporting bias. ***Our certainty would be moderate for effects of at least 225 false positives per 1000 as there would be serious concerns about inconsistency among studies contributing to analyses at both round one and over 2/3 rounds of screening (figures S2.27 and S2.28).***

[b] Incidental findings: ***No serious concerns about risk of bias:*** 8 studies at round one, and three studies at 4-5 rounds of screening were at high risk of bias, but findings did not change when excluded in sensitivity analysis. ***No serious concerns about inconsistency:*** Findings were consistent across rounds of screening and subgroup findings. ***No serious concerns about indirectness:*** The study data was direct for this outcome. ***No serious concerns about imprecision:*** The confidence intervals are precise for our threshold of effects over 3-4 screening rounds. ***No serious concerns about reporting bias:*** No serious concerns about reporting bias. ***Our certainty would be the same for the lower threshold of 100 per 1000. Our certainty would be moderate for effects of at least 450 incidental findings per 1000 as there would be serious concerns of inconsistency among studies contributing to analyses at both round one and over 4/5 rounds of screening (figures S2.30 and S2.32).***

[c] Clinically significant incidental findings: ***No serious concerns about risk of bias:*** 20 studies at round one, and two studies at 4-5 rounds of screening were at high risk of bias (usually due to a poorly defined or ascertained outcome), but findings did not change when excluded in sensitivity analysis. ***No serious concerns about inconsistency:*** Findings were consistent across rounds of screening and subgroup findings. ***No serious concerns about indirectness:*** The study data was direct for this outcome. ***No serious concerns about imprecision:*** The confidence intervals are precise for our threshold of effects over 3-4 screening rounds. ***No serious concerns about reporting bias:*** No serious concerns

about reporting bias. ***Our certainty would be moderate for effects of at least 100 clinically significant incidental findings per 1000 screened as there would be serious concerns about inconsistency among studies contributing to analyses at both round one and over 4/5 rounds of screening (figures S2.33 and S2.35).***

[d] Major complications from invasive testing (lung cancers and false positives): ***No serious concerns about risk of bias:*** Two studies at round one of screening were at high risk of bias, but findings were similar among studies not at high risk of bias. ***Very serious concerns about inconsistency:*** There were very serious concerns of inconsistency among studies contributing to each analysis (figures S2.36, S2.37, and S2.38). ***No serious concerns about indirectness:*** The study data was direct for this outcome. ***No serious concerns about imprecision:*** The confidence intervals are reasonably precise for our threshold of effects over 3-4 screening rounds. ***No serious concerns about reporting bias:*** No serious concerns about reporting bias.

[e] Major complications from invasive testing (false positives): ***No serious concerns about risk of bias:*** One study at round one of screening was at high risk of bias, but findings were similar among studies not at serious risk of bias. ***No serious concerns about inconsistency:*** Findings were consistent across rounds of screening and subgroup findings. ***No serious concerns about indirectness:*** The study data was direct for this outcome. ***No serious concerns about imprecision:*** The confidence intervals are precise for our threshold of effects over 3-4 screening rounds. ***No serious concerns about reporting bias:*** No serious concerns about reporting bias. ***Our certainty would be moderate for effects as low as 1 major complication from invasive testing among false positives per 1000 screened as there would be serious concerns about inconsistency among studies contributing to round 1 of screening (figure S2.39).***

[f] Deaths from invasive testing among false positives: ***No serious concerns about risk of bias:*** One study at round one of screening was at high risk of bias, but findings were similar among studies not at high risk of bias. ***Serious concerns about inconsistency:*** One large trial at 2-3 rounds of screening is inconsistent with findings at round one, and 4-5 rounds of screening. ***No serious concerns about indirectness:*** The study data was direct for this outcome. ***No serious concerns about imprecision:*** The confidence intervals are precise for our threshold of effects over 3-4 screening rounds. ***No serious concerns about reporting bias:*** No serious concerns about reporting bias.

[g] Health-related quality of life: ***Serious concerns about risk of bias:*** both studies included for this outcome were non-randomized subsamples of the original trials. ***No serious concerns about inconsistency:*** Both studies had consistent findings. ***Very serious concerns about indirectness:*** both studies were indirect for 3-4 rounds of screening (insufficient follow-up). Also, one study (NLST) is comparing CT screening to screening with a chest X-ray, rather than an unscreened comparator. ***No serious concerns about imprecision:*** The confidence intervals were precise for little-to-no difference. ***No serious concerns about reporting bias:*** No serious concerns about reporting bias.

[h] Psychosocial harms: Participating in screening program: ***No serious concerns about risk of bias:*** Only one of the three studies was a non-randomized sample (subsample of NELSON trial, and smallest sample in analysis), at high risk of bias. ***No serious concerns about inconsistency:*** All three studies had consistent findings. ***Serious concerns about indirectness:*** two of the three studies were less than 3-4 rounds of screening; one study with 5 rounds of screening indicated a trend for increasing effects over time but effects, on average, still did not meet our threshold so we did not judge this as very serious concerns. ***No serious concerns about imprecision:*** The confidence intervals were precise for little-to-no difference. ***No serious concerns about reporting bias:*** No serious concerns about reporting bias.

[i] Psychosocial harms: Undergoing CT scan (after screening, before results are known): ***Serious concerns about risk of bias:*** One of the two studies are at high risk of bias (the single comparative study was high risk of bias), and the majority of data came from pre-post analyses. ***Serious concerns of inconsistency:*** Findings were inconsistent between studies, and based on the comparator. ***No serious concerns about indirectness:*** Findings are from one round of screening but there were no serious concerns the results would differ over 3-4 screening rounds. One study (sample from the NELSON trial) included only those with indeterminate or negative screening results, but the sample is still applicable as results were not yet known. ***No serious concerns about imprecision:*** The confidence intervals were generally precise for little-to-no difference. ***No serious concerns about reporting bias:*** No serious concerns about reporting bias.

[j] Psychosocial harms: Positive screen (before repeat CT or diagnostic results known): ***Serious concerns about risk of bias:*** One study had high risk of bias, and all other studies had some concerns of risk of bias. ***Serious concerns about inconsistency:*** There was serious concerns of inconsistency across tools and comparisons. ***No serious concerns of indirectness:*** There were no serious concerns of indirectness. Two studies included only those with screening results requiring a recall scan, however lung-cancer specific tool from the trial subset (sample from the NELSON trial) and observational study (50) showed similar results to other studies including both those requiring recall scans and diagnostic workup. ***No serious concerns about imprecision:*** The confidence intervals were generally precise for a small increase. ***No serious concerns about reporting bias:*** No serious concerns about reporting bias.

[k] Psychosocial harms: Positive screen, requiring diagnostic follow-up (before diagnostic results known): ***Small effect threshold, Serious concerns about risk of bias:*** Both studies had some concerns of risk of bias. ***No serious concerns about inconsistency:*** Both studies had consistent findings, when considering this small effect. ***No serious concerns about***

***indirectness:*** The study data was direct for this outcome. ***No serious concerns about imprecision:*** The confidence intervals were generally precise for a moderate increase. ***No serious concerns about reporting bias:*** No serious concerns about reporting bias. ***For a moderate effect threshold, we had serious concerns about inconsistency in effects between studies, and heavy reliance on one study with adequate sample size.***

[l] Psychosocial harms: False positive screening result (≤6 months after diagnostic results known): ***Serious concerns about risk of bias:*** Four of the six studies are at high risk of bias (one of two comparative studies were high risk of bias), and the majority of data came from pre-post analyses. ***Serious concerns about inconsistency:*** There were serious concerns about inconsistency. Four of the six studies showed little-to-no difference, whereas two studies with an unknown number of lung cancer patients found at least a small increase in the overall score (LSUT trial) or in two of the four subscores (Bonney, 2024). ***No serious concerns about indirectness:*** The study data was direct for this outcome. ***No serious concerns about imprecision:*** The confidence intervals were generally precise for little-to-no difference. ***No serious concerns about reporting bias:*** No serious concerns about reporting bias.

[m] Psychosocial harms: False positive screening result (>6 months after diagnostic results known): ***Serious concerns about risk of bias:*** Two of the six included studies were at high risk of bias (two of four comparative studies were high risk of bias). ***No serious concerns about inconsistency:*** All six studies had consistent findings. ***No serious concerns about indirectness:*** The study data was direct for this outcome. ***No serious concerns about imprecision:*** The confidence intervals were generally precise for little-to-no difference. ***No serious concerns about reporting bias:*** No serious concerns about reporting bias.

| Study or Subgroup | Proportion | SE | Weight | Proportion IV, Random, 95% CI |
|---|---|---|---|---|
| **1.1.2 Observational** | | | | |
| Bartlett 2020 | 0.132 | 0.00994 | 3.5% | 0.13 [0.11, 0.15] |
| Bhamani 2025 | 0.176 | 0.00334 | 3.5% | 0.18 [0.17, 0.18] |
| Callol 2007 | 0.0665 | 0.0115 | 3.5% | 0.07 [0.04, 0.09] |
| Crosbie 2019 | 0.139 | 0.00916 | 3.5% | 0.14 [0.12, 0.16] |
| Henschke 2001 | 0.206 | 0.0128 | 3.5% | 0.21 [0.18, 0.23] |
| Lam 2015 | 0.695 | 0.0371 | 2.9% | 0.69 [0.62, 0.77] |
| Leleu 2024 | 0.158 | 0.0103 | 3.5% | 0.16 [0.14, 0.18] |
| Macredmond 2006 | 0.203 | 0.019 | 3.3% | 0.20 [0.17, 0.24] |
| Marshall 2015 | 0.48 | 0.0312 | 3.0% | 0.48 [0.42, 0.54] |
| Menezes 2010 | 0.166 | 0.00642 | 3.5% | 0.17 [0.15, 0.18] |
| Murray 2016 | 0.0728 | 0.00863 | 3.5% | 0.07 [0.06, 0.09] |
| Nahorecki 2015 | 0.172 | 0.0207 | 3.3% | 0.17 [0.13, 0.21] |
| Ostrowski 2019 | 0.157 | 0.00489 | 3.5% | 0.16 [0.15, 0.17] |
| Pastorino 2003 | 0.182 | 0.012 | 3.5% | 0.18 [0.16, 0.21] |
| Taghizadeh 2019 | 0.197 | 0.0113 | 3.5% | 0.20 [0.17, 0.22] |
| Veronesi 2008 | 0.0863 | 0.00389 | 3.5% | 0.09 [0.08, 0.09] |
| Wichmann 2025 | 0.0796 | 0.0191 | 3.3% | 0.08 [0.04, 0.12] |
| Wilson 2008 | 0.384 | 0.00794 | 3.5% | 0.38 [0.37, 0.40] |
| **Subtotal (95% CI)** | | | **61.5%** | **0.20 [0.17, 0.24]** |
| Heterogeneity: Tau² = 0.01; Chi² = 1721.12, df = 17 (P < 0.00001); I² = 99% | | | | |
| Test for overall effect: Z = 10.63 (P < 0.00001) | | | | |
| **1.1.3 Trials** | | | | |
| DANTE (Infante 2008) | 0.0902 | 0.00806 | 3.5% | 0.09 [0.07, 0.11] |
| DEPISCAN (Blanchon 2007) | 0.19 | 0.02 | 3.3% | 0.19 [0.15, 0.23] |
| DLCST (Pedersen 2009) | 0.0789 | 0.00595 | 3.5% | 0.08 [0.07, 0.09] |
| ITALUNG (Lopes Pegna 2013) | 0.252 | 0.0108 | 3.5% | 0.25 [0.23, 0.27] |
| LSS (Gohagan 2004) | 0.178 | 0.00938 | 3.5% | 0.18 [0.16, 0.20] |
| LSUT (Ruparel 2020) | 0.142 | 0.0119 | 3.5% | 0.14 [0.12, 0.17] |
| LUSI (Becker 2020) | 0.21 | 0.00905 | 3.5% | 0.21 [0.19, 0.23] |
| MILD (Pastorino 2012) | 0.134 | 0.00699 | 3.5% | 0.13 [0.12, 0.15] |
| NELSON (Horeweg 2013) | 0.19 | 0.00441 | 3.5% | 0.19 [0.18, 0.20] |
| NLST (Aberle 2011) | 0.259 | 0.00268 | 3.6% | 0.26 [0.25, 0.26] |
| UKLS (Mascalchi 2016) | 0.244 | 0.00953 | 3.5% | 0.24 [0.23, 0.26] |
| **Subtotal (95% CI)** | | | **38.5%** | **0.18 [0.14, 0.22]** |
| Heterogeneity: Tau² = 0.01; Chi² = 1245.61, df = 10 (P < 0.00001); I² = 99% | | | | |
| Test for overall effect: Z = 8.01 (P < 0.00001) | | | | |
| **Total (95% CI)** | | | **100.0%** | **0.19 [0.16, 0.22]** |
| Heterogeneity: Tau² = 0.01; Chi² = 3330.04, df = 28 (P < 0.00001); I² = 99% | | | | |
| Test for overall effect: Z = 13.39 (P < 0.00001) | | | | |
| Test for subgroup differences: Chi² = 0.64, df = 1 (P = 0.42), I² = 0% | | | | |

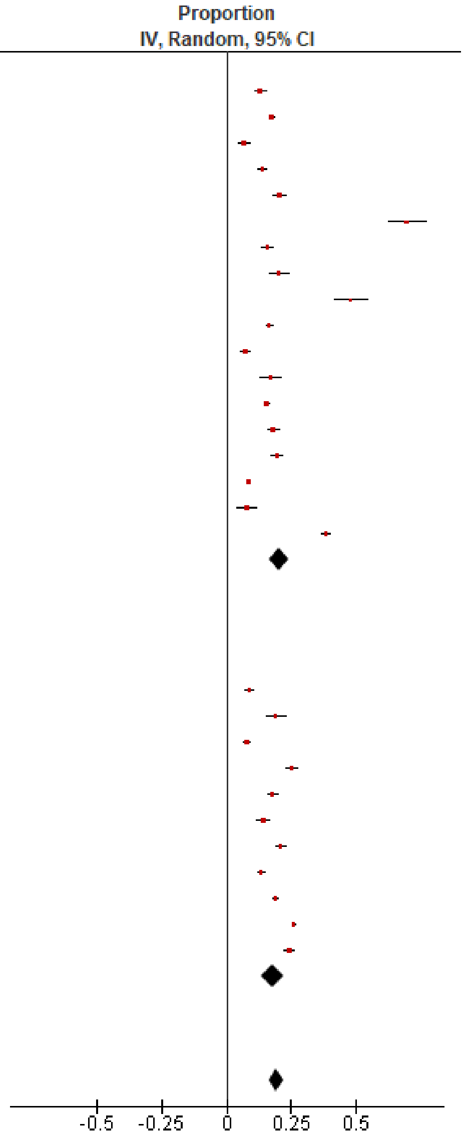

Figure S2.27 Proportion with a false positive at round 1

| Study or Subgroup | Proportion | SE | Weight | Proportion IV, Random, 95% CI |
|---|---|---|---|---|
| **1.3.2 Observational** | | | | |
| Callol 2007 | 0.0773 | 0.0124 | 10.0% | 0.08 [0.05, 0.10] |
| Henschke 2001 | 0.262 | 0.0139 | 10.0% | 0.26 [0.23, 0.29] |
| Kim 2020 | 0.157 | 0.00188 | 10.0% | 0.16 [0.15, 0.16] |
| Macredmond 2006 | 0.241 | 0.0202 | 9.9% | 0.24 [0.20, 0.28] |
| Menezes 2010 | 0.25 | 0.00748 | 10.0% | 0.25 [0.24, 0.26] |
| Rzyman 2015 | 0.53 | 0.00537 | 10.0% | 0.53 [0.52, 0.54] |
| Wichmann 2025 | 0.129 | 0.0237 | 9.9% | 0.13 [0.08, 0.18] |
| Wilson 2008 | 0.758 | 0.00699 | 10.0% | 0.76 [0.74, 0.77] |
| **Subtotal (95% CI)** | | | **80.0%** | **0.30 [0.12, 0.48]** |
| Heterogeneity: Tau² = 0.07; Chi² = 10517.06, df = 7 (P < 0.00001); I² = 100% | | | | |
| Test for overall effect: Z = 3.22 (P = 0.001) | | | | |
| **1.3.3 Trials** | | | | |
| LSS (Croswell 2010) | 0.305 | 0.0113 | 10.0% | 0.30 [0.28, 0.33] |
| NLST (Aberle 2011) | 0.377 | 0.00296 | 10.0% | 0.38 [0.37, 0.38] |
| **Subtotal (95% CI)** | | | **20.0%** | **0.34 [0.27, 0.41]** |
| Heterogeneity: Tau² = 0.00; Chi² = 37.99, df = 1 (P < 0.00001); I² = 97% | | | | |
| Test for overall effect: Z = 9.50 (P < 0.00001) | | | | |
| **Total (95% CI)** | | | **100.0%** | **0.31 [0.18, 0.44]** |
| Heterogeneity: Tau² = 0.04; Chi² = 12421.54, df = 9 (P < 0.00001); I² = 100% | | | | |
| Test for overall effect: Z = 4.80 (P < 0.00001) | | | | |
| Test for subgroup differences: Chi² = 0.17, df = 1 (P = 0.68), I² = 0% | | | | |

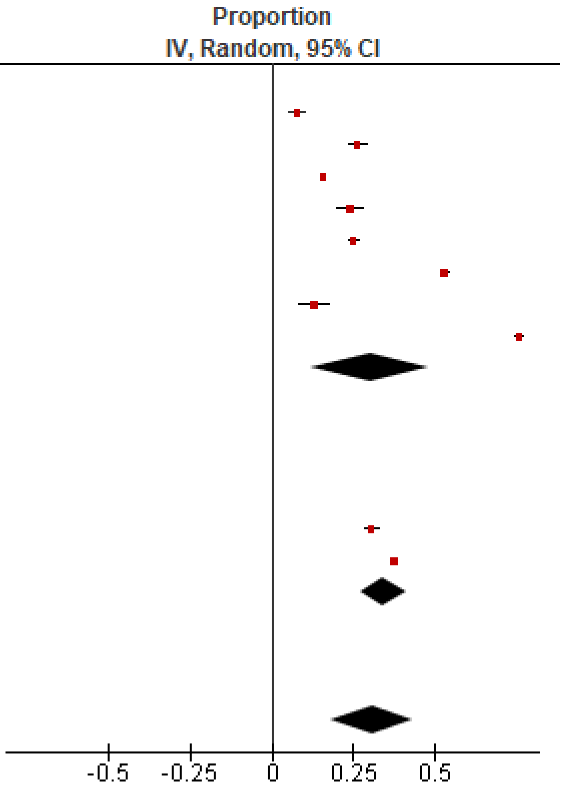

Figure S2.28 Proportion with a false positive over 2/3 rounds

| Study or Subgroup | Proportion | SE | Weight | Proportion IV, Random, 95% CI |
|---|---|---|---|---|
| **1.4.1 Observational** | | | | |
| Miller 2016 | 0.387 | 0.0137 | 33.3% | 0.39 [0.36, 0.41] |
| **Subtotal (95% CI)** | | | **33.3%** | **0.39 [0.36, 0.41]** |
| Heterogeneity: Not applicable | | | | |
| Test for overall effect: Z = 28.25 (P < 0.00001) | | | | |
| **1.4.3 Trials** | | | | |
| DANTE (Infante 2015) | 0.229 | 0.0118 | 33.4% | 0.23 [0.21, 0.25] |
| ITALUNG (Lopes Pegna 2013) | 0.438 | 0.0124 | 33.3% | 0.44 [0.41, 0.46] |
| **Subtotal (95% CI)** | | | **66.7%** | **0.33 [0.13, 0.54]** |
| Heterogeneity: Tau² = 0.02; Chi² = 149.08, df = 1 (P < 0.00001); I² = 99% | | | | |
| Test for overall effect: Z = 3.19 (P = 0.001) | | | | |
| **Total (95% CI)** | | | **100.0%** | **0.35 [0.22, 0.48]** |
| Heterogeneity: Tau² = 0.01; Chi² = 162.29, df = 2 (P < 0.00001); I² = 99% | | | | |
| Test for overall effect: Z = 5.36 (P < 0.00001) | | | | |
| Test for subgroup differences: Chi² = 0.26, df = 1 (P = 0.61), I² = 0% | | | | |

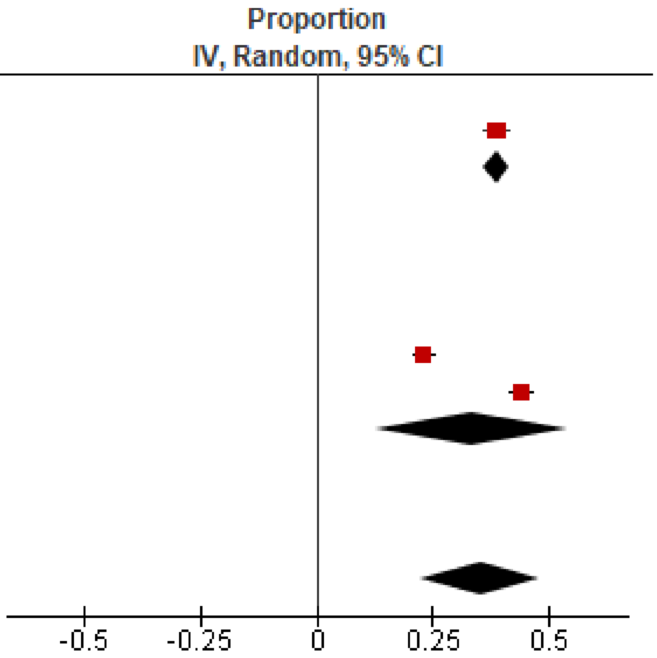

Figure S2.29 Proportion with a false positive over 4/5 rounds

| Study or Subgroup | Proportion | SE | Weight | Proportion IV, Random, 95% CI |
|---|---|---|---|---|
| **1.1.2 Observational** | | | | |
| Bonney 2025 | 0.732 | 0.00667 | 7.7% | 0.73 [0.72, 0.75] |
| Callol 2007 | 0.0558 | 0.0106 | 7.7% | 0.06 [0.04, 0.08] |
| Fabbrini 2018 | 0.652 | 0.0357 | 7.6% | 0.65 [0.58, 0.72] |
| Janssen 2019 | 0.438 | 0.012 | 7.7% | 0.44 [0.41, 0.46] |
| Laisaar 2025 | 0.969 | 0.00296 | 7.7% | 0.97 [0.96, 0.97] |
| Lam 2015 | 0.156 | 0.0292 | 7.7% | 0.16 [0.10, 0.21] |
| Ledford 2016 | 0.181 | 0.0453 | 7.6% | 0.18 [0.09, 0.27] |
| MacRedmond 2006 | 0.617 | 0.0229 | 7.7% | 0.62 [0.57, 0.66] |
| Melzer 2024 | 0.939 | 0.00798 | 7.7% | 0.94 [0.92, 0.95] |
| Morgan 2017 | 0.947 | 0.0125 | 7.7% | 0.95 [0.92, 0.97] |
| Priola 2013 | 0.57 | 0.0217 | 7.7% | 0.57 [0.53, 0.61] |
| Reiter 2018 | 0.692 | 0.0192 | 7.7% | 0.69 [0.65, 0.73] |
| Shah 2024 | 0.206 | 0.011 | 7.7% | 0.21 [0.18, 0.23] |
| **Subtotal (95% CI)** | | | **100.0%** | **0.55 [0.37, 0.73]** |
| Heterogeneity: Tau² = 0.11; Chi² = 13351.65, df = 12 (P < 0.00001); I² = 100% | | | | |
| Test for overall effect: Z = 5.92 (P < 0.00001) | | | | |
| **Total (95% CI)** | | | **100.0%** | **0.55 [0.37, 0.73]** |
| Heterogeneity: Tau² = 0.11; Chi² = 13351.65, df = 12 (P < 0.00001); I² = 100% | | | | |
| Test for overall effect: Z = 5.92 (P < 0.00001) | | | | |
| Test for subgroup differences: Not applicable | | | | |

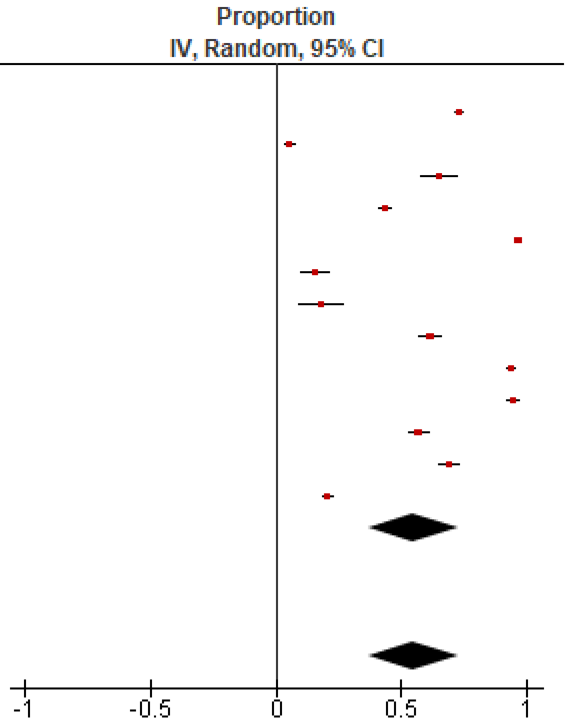

Figure S2.30 Proportion with incidental findings at round 1

| Study or Subgroup | Proportion | SE | Weight | Proportion IV, Random, 95% CI |
|---|---|---|---|---|
| **1.3.3 Trials** | | | | |
| NLST (Gareen 2023) | 0.569 | 0.00303 | 100.0% | 0.57 [0.56, 0.57] |
| **Subtotal (95% CI)** | | | **100.0%** | **0.57 [0.56, 0.57]** |
| Heterogeneity: Not applicable | | | | |
| Test for overall effect: Z = 187.79 (P < 0.00001) | | | | |
| **Total (95% CI)** | | | **100.0%** | **0.57 [0.56, 0.57]** |
| Heterogeneity: Not applicable | | | | |
| Test for overall effect: Z = 187.79 (P < 0.00001) | | | | |
| Test for subgroup differences: Not applicable | | | | |

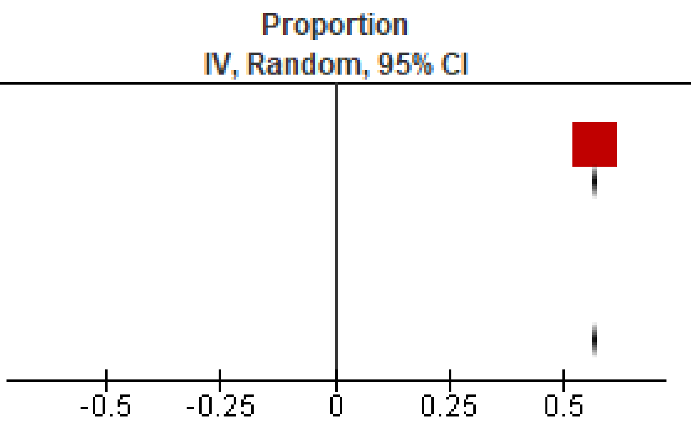

Figure S2.31 Proportion with incidental findings over 2/3 rounds

| Study or Subgroup | Proportion | SE | Weight | Proportion IV, Random, 95% CI |
|---|---|---|---|---|
| **1.4.2 Observational** | | | | |
| Priola 2013 | 0.732 | 0.0194 | 33.3% | 0.73 [0.69, 0.77] |
| **Subtotal (95% CI)** | | | **33.3%** | **0.73 [0.69, 0.77]** |
| Heterogeneity: Not applicable | | | | |
| Test for overall effect: Z = 37.73 (P < 0.00001) | | | | |
| **1.4.3 Trials** | | | | |
| DANTE (Infante 2015) | 0.0688 | 0.00712 | 33.4% | 0.07 [0.05, 0.08] |
| ITALUNG (Puliti 2019 & Lopes Pegna 2013) | 0.39 | 0.0121 | 33.3% | 0.39 [0.37, 0.41] |
| **Subtotal (95% CI)** | | | **66.7%** | **0.23 [-0.09, 0.54]** |
| Heterogeneity: Tau² = 0.05; Chi² = 523.43, df = 1 (P < 0.00001); I² = 100% | | | | |
| Test for overall effect: Z = 1.43 (P = 0.15) | | | | |
| **Total (95% CI)** | | | **100.0%** | **0.40 [0.05, 0.75]** |
| Heterogeneity: Tau² = 0.10; Chi² = 1337.60, df = 2 (P < 0.00001); I² = 100% | | | | |
| Test for overall effect: Z = 2.23 (P = 0.03) | | | | |
| Test for subgroup differences: Chi² = 9.66, df = 1 (P = 0.002), I² = 89.6% | | | | |

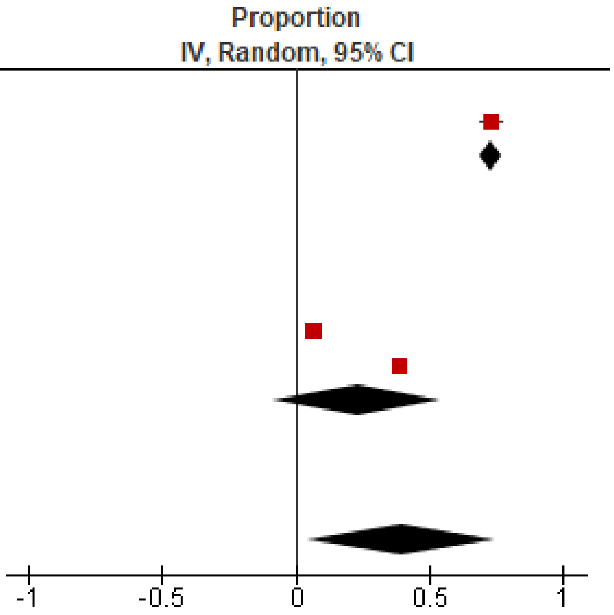

Figure S2.32 Proportion with incidental findings over 4/5 rounds

| Study or Subgroup | Proportion | SE | Weight | Proportion IV, Random, 95% CI |
|---|---|---|---|---|
| **2.1.1 Observational** | | | | |
| Bartlett 2021 | 0.0785 | 0.0079 | 3.8% | 0.08 [0.06, 0.09] |
| Danila 2025 | 0.202 | 0.012 | 3.8% | 0.20 [0.18, 0.23] |
| Darling 2021 | 0.184 | 0.00835 | 3.8% | 0.18 [0.17, 0.20] |
| Guichet 2018 | 0.291 | 0.0274 | 3.5% | 0.29 [0.24, 0.34] |
| Hirsch 2023 | 0.263 | 0.017 | 3.7% | 0.26 [0.23, 0.30] |
| Kapoor 2020 | 0.0947 | 0.00767 | 3.8% | 0.09 [0.08, 0.11] |
| Kearney 2023 | 0.0576 | 0.0012 | 3.8% | 0.06 [0.06, 0.06] |
| Kinsinger 2017 | 0.35 | 0.00963 | 3.8% | 0.35 [0.33, 0.37] |
| Kucharczyk 2011 | 0.192 | 0.00617 | 3.8% | 0.19 [0.18, 0.20] |
| Laisaar 2025 | 0.0828 | 0.00469 | 3.8% | 0.08 [0.07, 0.09] |
| Lam 2015 | 0.039 | 0.0156 | 3.7% | 0.04 [0.01, 0.07] |
| Ledford 2016 | 0.0833 | 0.0326 | 3.4% | 0.08 [0.02, 0.15] |
| MacRedmond 2006 | 0.494 | 0.0236 | 3.6% | 0.49 [0.45, 0.54] |
| Matthews 2024 | 0.0995 | 0.00398 | 3.8% | 0.10 [0.09, 0.11] |
| Melzer 2024 | 0.625 | 0.0161 | 3.7% | 0.63 [0.59, 0.66] |
| Morgan 2017 | 0.288 | 0.0253 | 3.6% | 0.29 [0.24, 0.34] |
| Murray 2016 | 0.0938 | 0.00969 | 3.8% | 0.09 [0.07, 0.11] |
| Priola 2013 | 0.1 | 0.0132 | 3.8% | 0.10 [0.07, 0.13] |
| Pua 2024 (hospital site) | 0.282 | 0.0306 | 3.5% | 0.28 [0.22, 0.34] |
| Pua 2024 (mobile site) | 0.367 | 0.0426 | 3.2% | 0.37 [0.28, 0.45] |
| Regis 2022 | 0.064 | 0.00373 | 3.8% | 0.06 [0.06, 0.07] |
| Reiter 2018 | 0.449 | 0.0206 | 3.7% | 0.45 [0.41, 0.49] |
| Rode 2025 | 0.187 | 0.00784 | 3.8% | 0.19 [0.17, 0.20] |
| Shah 2024 | 0.188 | 0.0106 | 3.8% | 0.19 [0.17, 0.21] |
| Triplette 2024 | 0.213 | 0.027 | 3.6% | 0.21 [0.16, 0.27] |
| **Subtotal (95% CI)** | | | **92.5%** | **0.21 [0.18, 0.25]** |
| Heterogeneity: Tau² = 0.01; Chi² = 4198.96, df = 24 (P < 0.00001); I² = 99% | | | | |
| Test for overall effect: Z = 11.90 (P < 0.00001) | | | | |
| **2.1.3 Trials** | | | | |
| LSUT (Kummer 2020) | 0.434 | 0.0169 | 3.7% | 0.43 [0.40, 0.47] |
| NLST (Gareen 2023) | 0.22 | 0.00253 | 3.8% | 0.22 [0.22, 0.22] |
| **Subtotal (95% CI)** | | | **7.5%** | **0.33 [0.12, 0.54]** |
| Heterogeneity: Tau² = 0.02; Chi² = 156.83, df = 1 (P < 0.00001); I² = 99% | | | | |
| Test for overall effect: Z = 3.05 (P = 0.002) | | | | |
| **Total (95% CI)** | | | **100.0%** | **0.22 [0.18, 0.26]** |
| Heterogeneity: Tau² = 0.01; Chi² = 7197.56, df = 26 (P < 0.00001); I² = 100% | | | | |
| Test for overall effect: Z = 11.57 (P < 0.00001) | | | | |
| Test for subgroup differences: Chi² = 1.11, df = 1 (P = 0.29), I² = 10.2% | | | | |

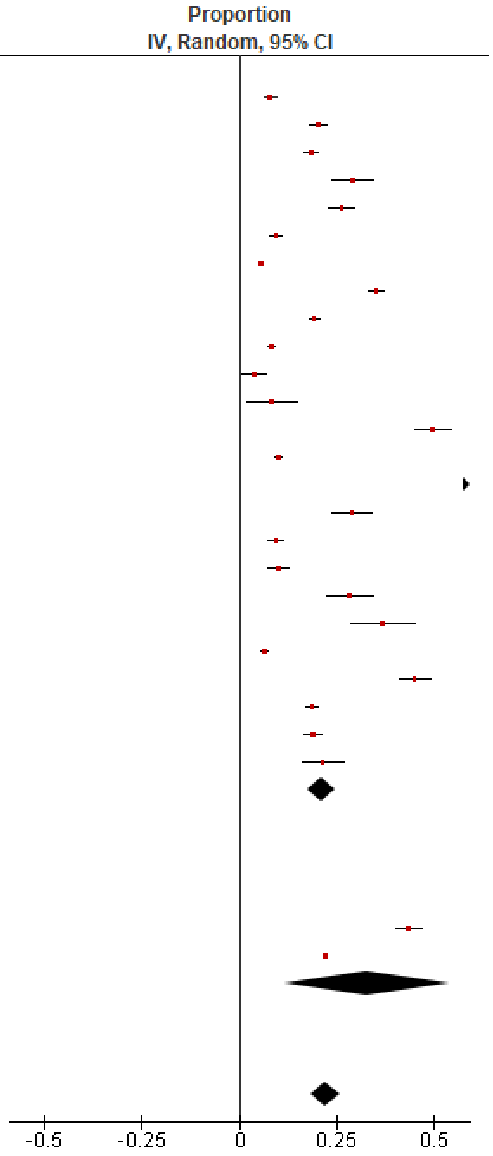

Figure S2.33 Proportion with clinically significant incidental findings at round 1

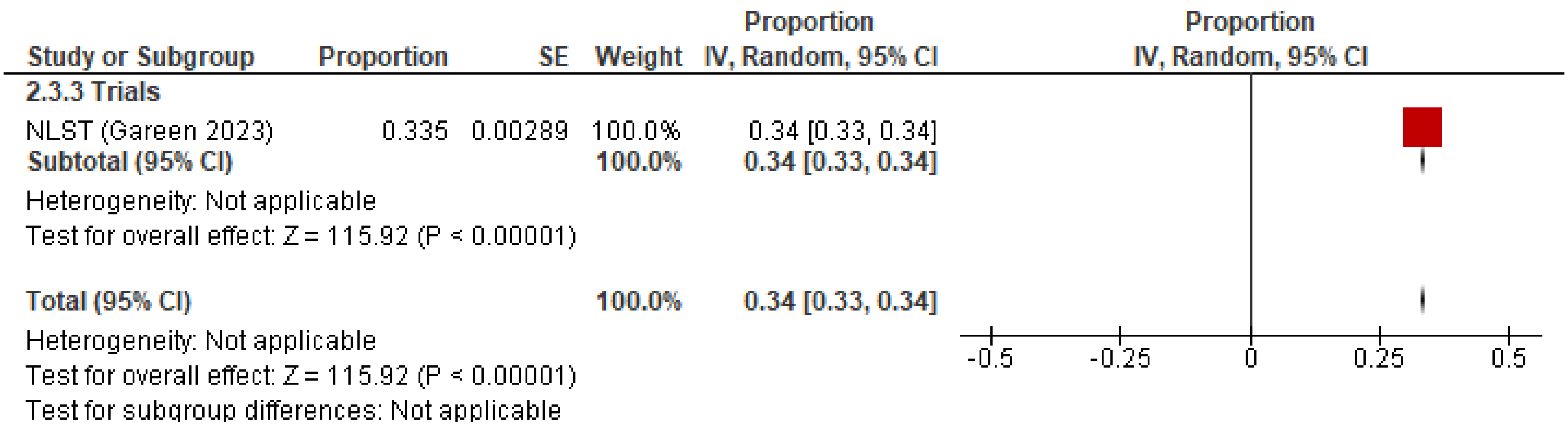

Figure S2.34 Proportion with clinically significant incidental findings over 2/3 rounds

| Study or Subgroup | Proportion | SE | Weight | Proportion IV, Random, 95% CI |
|---|---|---|---|---|
| **2.4.1 Observational** | | | | |
| Manyak 2023 | 0.016 | 0.0028 | 25.6% | 0.02 [0.01, 0.02] |
| Priola 2013 | 0.119 | 0.0142 | 23.4% | 0.12 [0.09, 0.15] |
| Rampinelli 2011 | 0.0773 | 0.0037 | 25.5% | 0.08 [0.07, 0.08] |
| Regis 2022 | 0.0988 | 0.00455 | 25.4% | 0.10 [0.09, 0.11] |
| **Subtotal (95% CI)** | | | **100.0%** | **0.08 [0.03, 0.12]** |

Heterogeneity: Tau² = 0.00; Chi² = 340.03, df = 3 (P < 0.00001); I² = 99%
Test for overall effect: Z = 3.30 (P = 0.0010)

| | | | | |
|---|---|---|---|---|
| **Total (95% CI)** | | | **100.0%** | **0.08 [0.03, 0.12]** |

Heterogeneity: Tau² = 0.00; Chi² = 340.03, df = 3 (P < 0.00001); I² = 99%
Test for overall effect: Z = 3.30 (P = 0.0010)
Test for subgroup differences: Not applicable

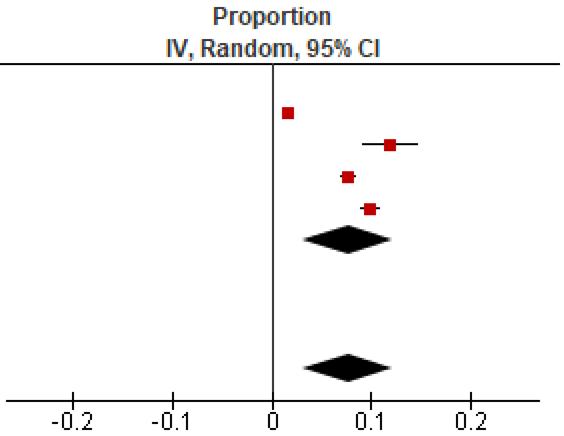

Figure S2.35 Proportion with clinically significant incidental findings over 4/5 rounds

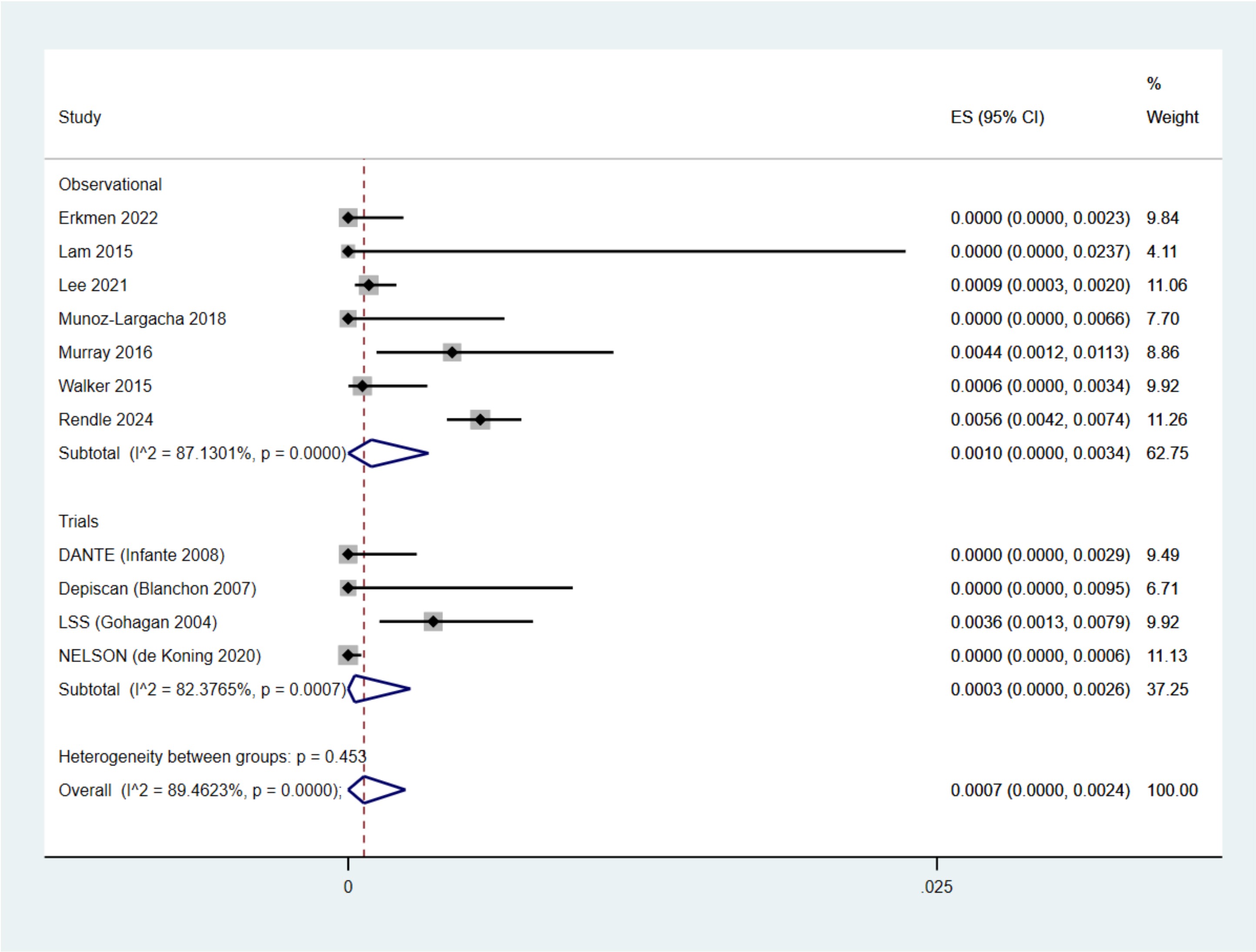

Figure S2.36 Proportion with major complications from invasive testing at round 1

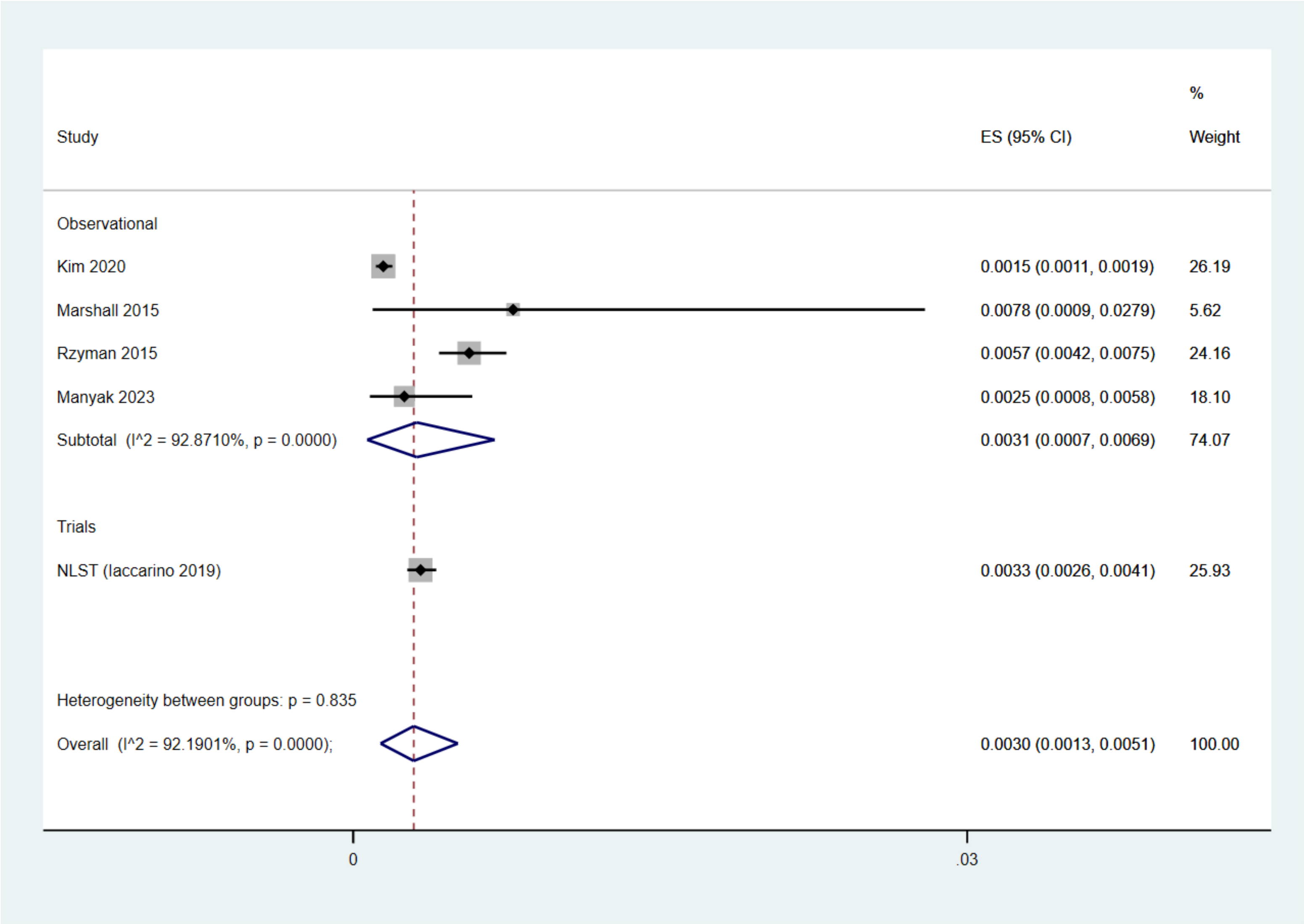

Figure S2.37 Proportion with major complications from invasive testing over 2/3 rounds

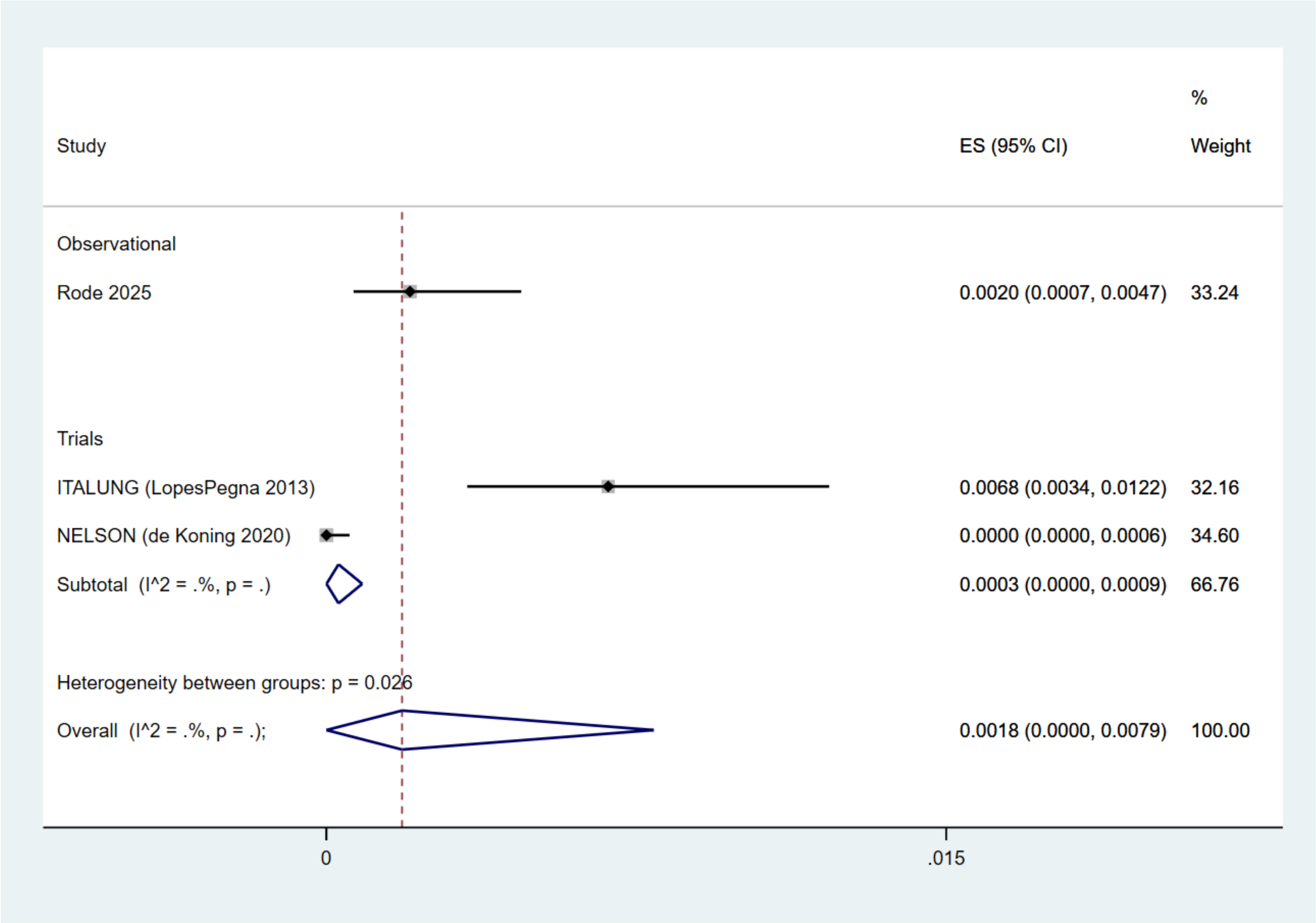

Figure S2.38 Proportion with major complications from invasive testing over 4/5 rounds

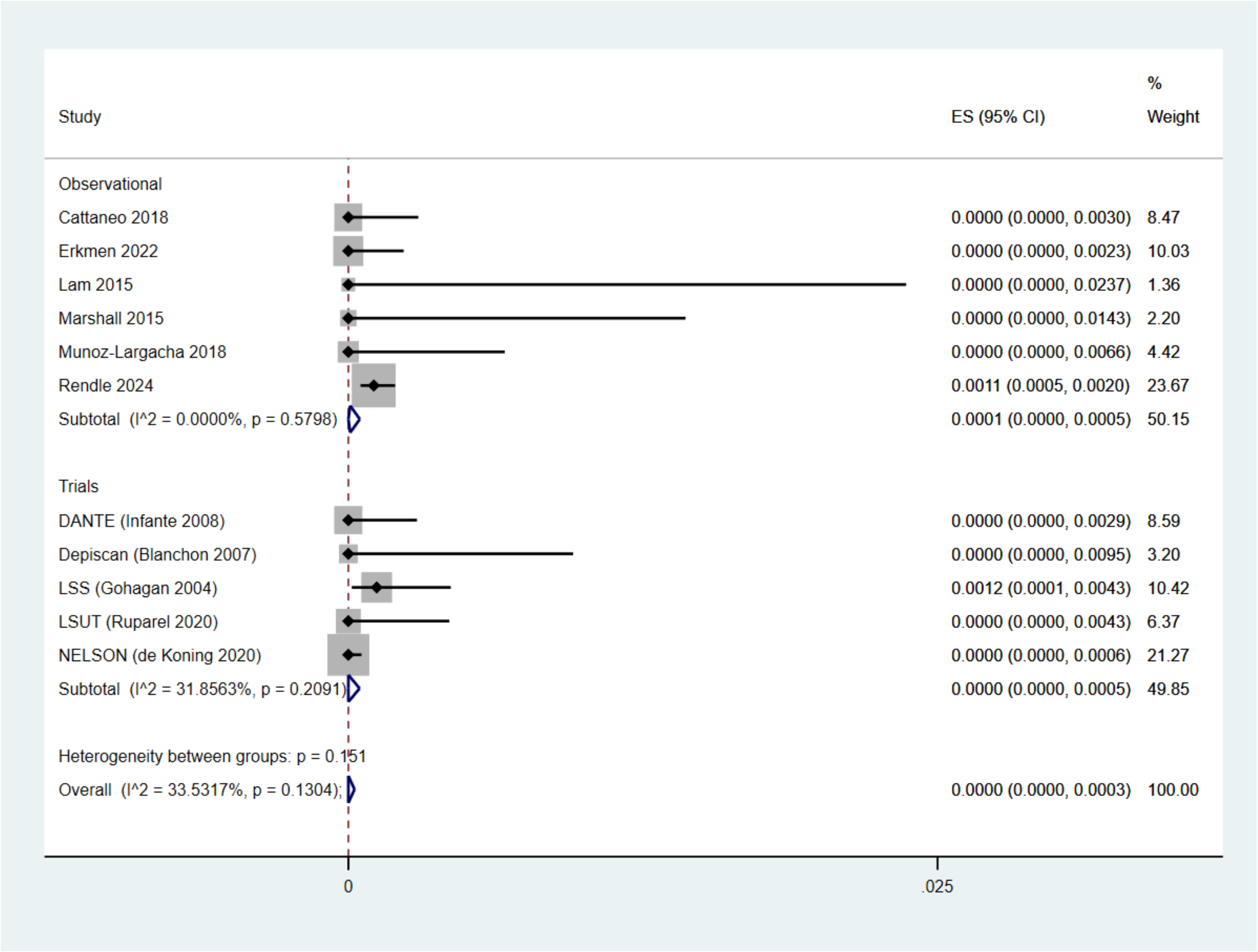

Figure S2.39 Proportion with major complications from invasive testing among false positives at round 1

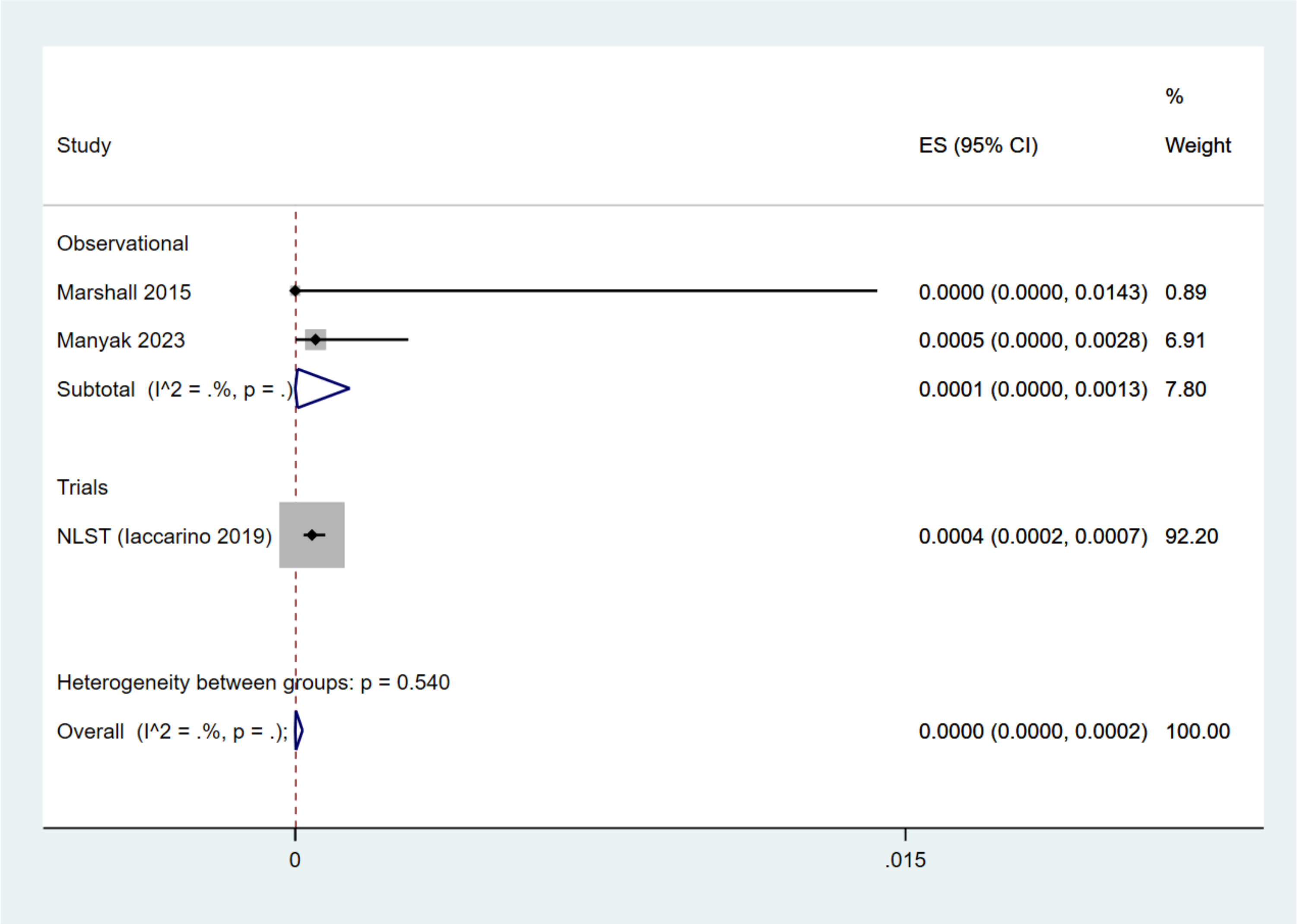

Figure S2.40 Proportion with major complications from invasive testing among false positives over 2/3 rounds

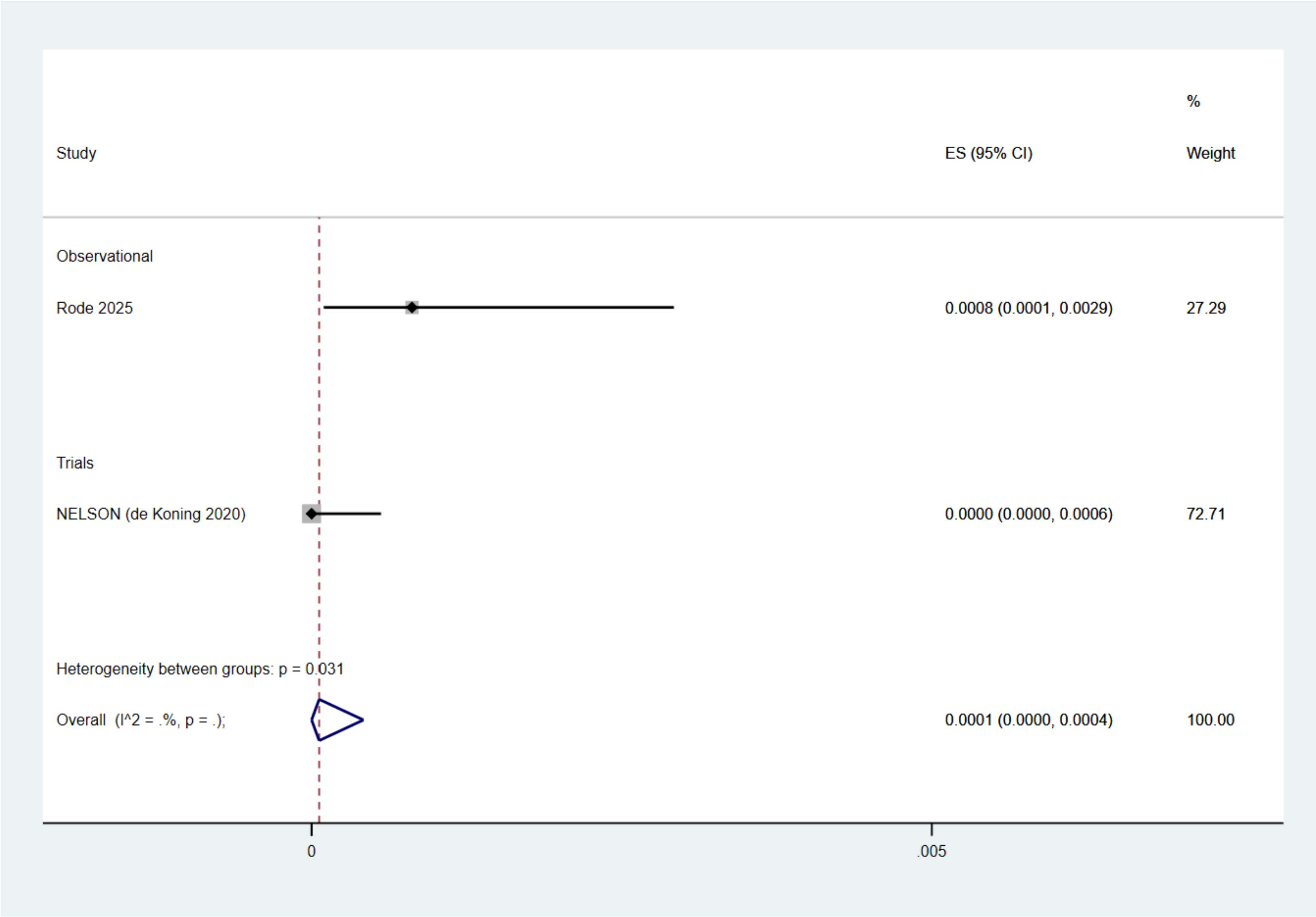

Figure S2.41 Proportion with major complications from invasive testing among false positives over 4/5 rounds

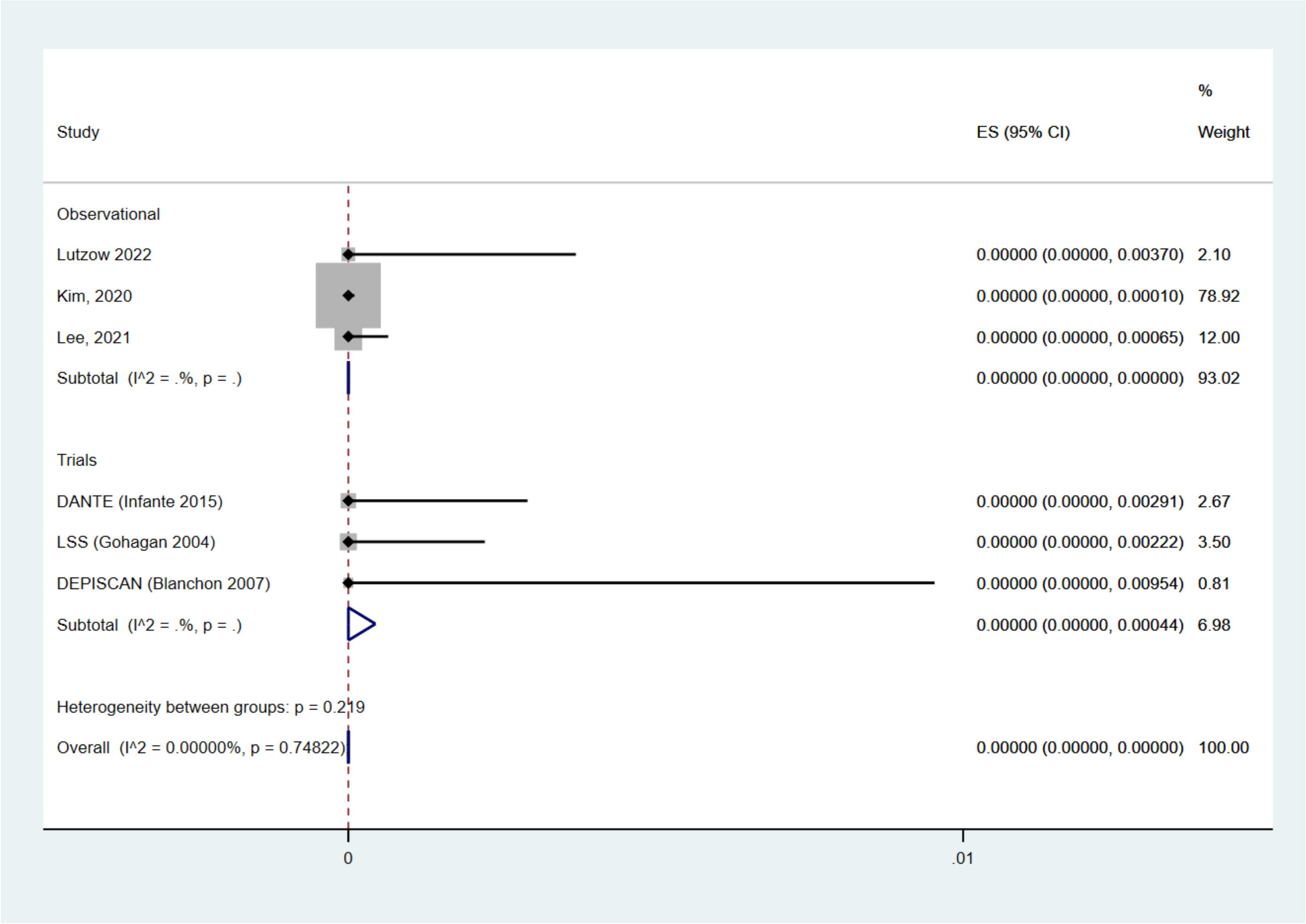

Figure S2.42 Proportion of deaths from invasive testing among false positives at round 1

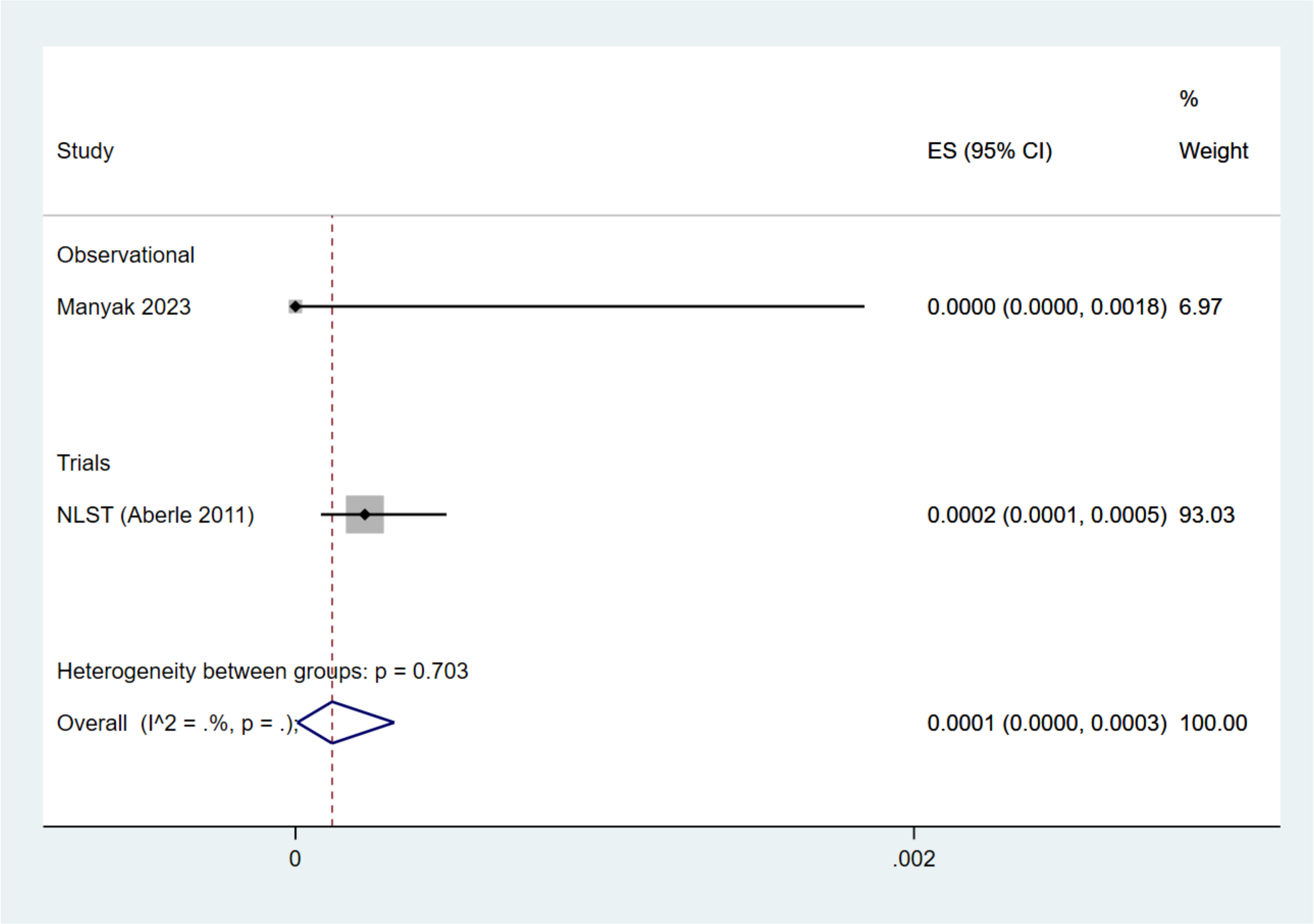

Figure S2.43 Proportion of deaths from invasive testing among false positives over 2/3 rounds

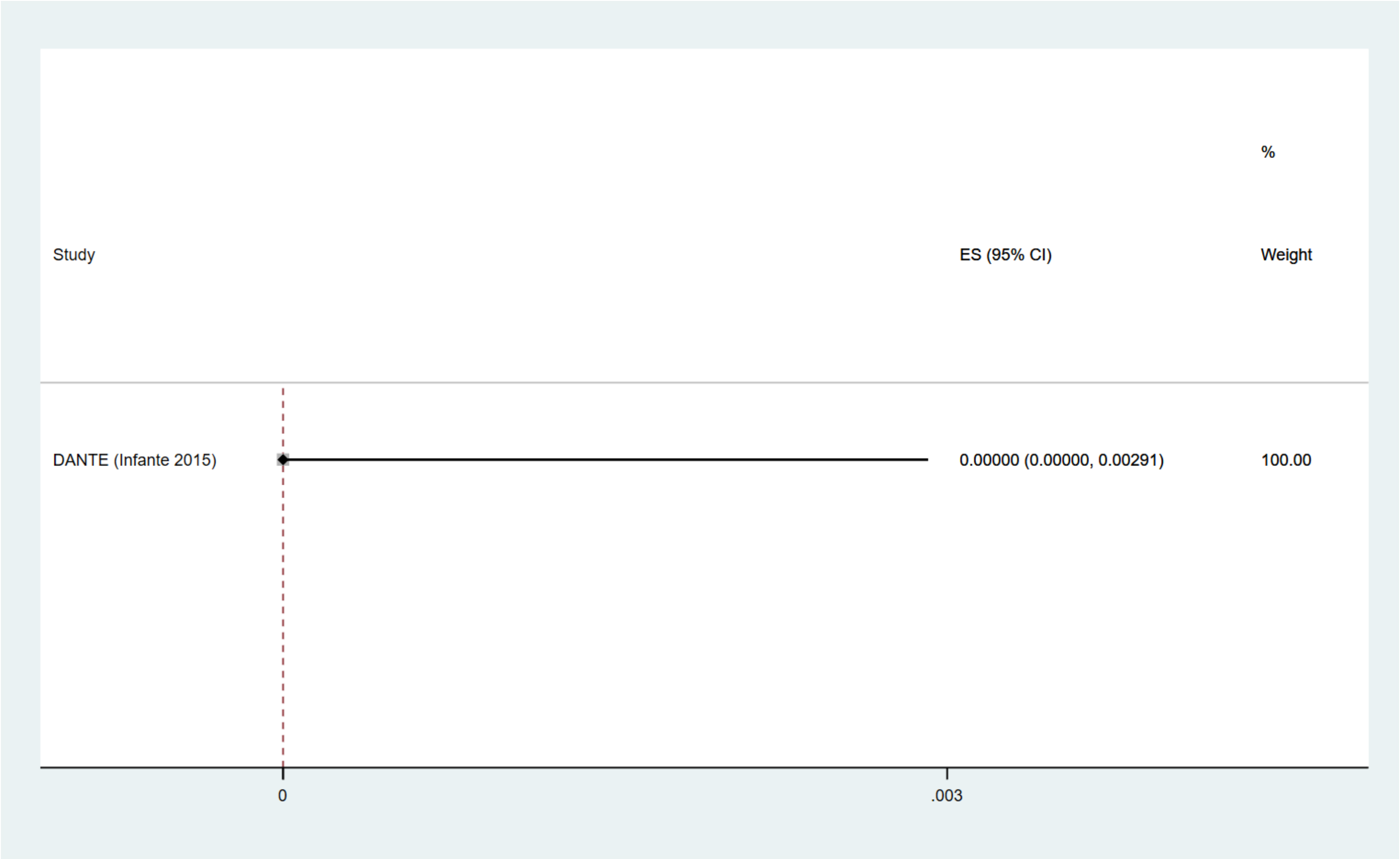

Figure S2.44 Proportion of deaths from invasive testing among false positives over 4/5 rounds

Table S2.10 CT screening: Important outcomes sensitivity and subgroup analyses

| Comparison | N studies | n | $I^2$ | Absolute number (95% CI) per 1,000 |
|---|---|---|---|---|
| **False positives** | | | | |
| Between study subgroup: Round 1: Proportion with a false positive, by study design (test for subgroup differences, p=0.42) | | | | |
| Observational | 18 (52, 54-70) | 40,757 | 99% | 200 [170, 240] |
| Trials | 11 (16, 23, 30, 32, 36, 38, 41, 43, 46, 71, 72) | 48,888 | 99% | 180 [140, 220] |
| Sensitivity analysis: Round 1: Proportion with a false positive, excluding high risk of bias studies | | | | |
| All studies | 29 (16, 23, 30, 32, 36, 38, 41, 43, 46, 52, 54-72) | 89,645 | 99% | 200 [170, 240] |
| Not high risk of bias | 22 (16, 30, 32, 36, 41, 43, 52, 55-58, 61-64, 66-72) | 56,751 | 99% | 170 [140, 200] |
| Between study subgroup: Round 1: Proportion with a false positive, by nodule management (test for subgroup differences, p<0.0001) | | | | |
| Highly sensitive | 4 (23, 46, 59, 62) | 28,792 | 99% | 390 [290, 500] |
| Less sensitive | 25 (16, 30, 32, 36, 38, 41, 43, 52, 54-58, 60, 61, 63-72) | 60,853 | 99% | 160 [140, 190] |
| Within study subgroup: Round 1: Proportion with a false positive, by age (test for subgroup differences, p<0.00001) | | | | |
| <65 years | Trials: 1 (25) | 19,612 | NA | 250 [240, 250] |
| ≥65 years | Trials: 1 (25) | 7,110 | NA | 300 [290, 310] |
| Within study subgroup: Round 1: Proportion with a false positive, by 5-year lung cancer death risk quintile (test for subgroup differences, p<0.00001) | | | | |
| Risk quintile 1 | Trials: 1 (3) | 5,276 | NA | 220 [210, 230] |
| Risk quintile 2 | Trials: 1 (3) | 5,310 | NA | 240 [230, 250] |
| Risk quintile 3 | Trials: 1 (3) | 5,396 | NA | 250 [240, 260] |
| Risk quintile 4 | Trials: 1 (3) | 5,314 | NA | 270 [260, 280] |
| Risk quintile 5 | Trials: 1 (3) | 5,308 | NA | 290 [280, 310] |
| Between study subgroup: 2/3 Rounds: Proportion with a false positive, by study design (test for subgroup differences, p=0.68) | | | | |
| Observational | 8 (56, 58, 61, 63, 69, 70, 73, 74) | 55,308 | 100% | 300 [120, 480] |
| Trials | 2 (23, 48) | 28,382 | 97% | 340 [270, 410] |
| Sensitivity analysis: 2/3 Rounds: Proportion with a false positive, excluding high risk of bias studies | | | | |
| All studies | 10 (23, 48, 56, 58, 61, 63, 69, 70, 73, 74) | 83,680 | 100% | 310 [180, 440] |
| Not high risk of bias | 8 (56, 58, 61, 63, 69, 70, 73, 74) | 55,308 | 100% | 300 [120, 480] |
| Between study subgroup: 2/3 Rounds: Proportion with a false positive, by nodule management (test for subgroup differences, p=0.75) | | | | |
| Highly sensitive | 3 (23, 48, 73) | 65,818 | 100% | 280 [110, 450] |
| Less sensitive | 7 (56, 58, 61, 63, 69, 70, 74) | 17,872 | 100% | 320 [130, 510] |
| Within study subgroup: 2/3 Rounds: Proportion with a false positive, by age (test for subgroup differences, p<0.00001) | | | | |

| Comparison | N studies | n | $I^2$ | Absolute number (95% CI) per 1,000 |
|---|---|---|---|---|
| <65 years | Trials: 1 (25) | 19,612 | NA | 350 [340, 350] |
| ≥65 years | Trials: 1 (25) | 7,110 | NA | 410 [400, 430] |
| Within study subgroup: 2/3 Rounds: Proportion with a false positive, by smoking status (test for subgroup differences, p=0.79) | | | | |
| Never-smoker | Observational: 1 (73) | 17,968 | NA | 160 [150, 160] |
| Ever-smoker | Observational: 1 (73) | 19,468 | NA | 160 [150, 160] |
| Within study subgroup: 2/3 Rounds: Proportion with a false positive, by 5-year lung cancer death risk quintile (test for subgroup differences, p<0.00001) | | | | |
| Risk quintile 1 | Trials: 1 (3) | 5,276 | NA | 310 [300, 320] |
| Risk quintile 2 | Trials: 1 (3) | 5,310 | NA | 340 [330, 350] |
| Risk quintile 3 | Trials: 1 (3) | 5,396 | NA | 350 [340, 370] |
| Risk quintile 4 | Trials: 1 (3) | 5,314 | NA | 370 [360, 380] |
| Risk quintile 5 | Trials: 1 (3) | 5,308 | NA | 400 [390, 420] |
| Within study subgroup: 2/3 Rounds: Proportion with a false positive, by 6-year lung cancer risk prediction decile (test for subgroup differences, p=NR) | | | | |
| 1 trial; N=26,309 overall (NLST): "Cumulative false-positive proportion increased with risk decile, ranging from 29.7% in the lowest to 41.4% in the highest decile." (26) | | | | |
| Between study subgroup: 4/5 Rounds: Proportion with a false positive, by study design (test for subgroup differences, p=0.61) | | | | |
| Observational | 1 (75) | 1,267 | NA | 390 [360, 410] |
| Trials | 2 (11, 38) | 2,877 | 99% | 330 [130, 540] |
| Sensitivity analysis: 4/5 Rounds: Proportion with a false positive, excluding high risk of bias studies | | | | |
| All studies | 3 (11, 38, 75) | 4,144 | 99% | 350 [220, 480] |
| Not high risk of bias | NA (None at high risk of bias) | NA | NA | NA |
| Between study subgroup: 4/5 Rounds: Proportion with a false positive, by nodule management (test for subgroup differences, p=NA) | | | | |
| Highly sensitive | None (All less sensitive nodule management) | NA | NA | NA |
| Less sensitive | 3 (11, 38, 75) | 4,144 | 99% | 350 [220, 480] |
| **Incidental findings** | | | | |
| Between study subgroup: Round 1: Proportion with an incidental finding, by study design (test for subgroup differences, p=NA) | | | | |
| Observational | 13 (56, 59, 61, 76-85) | 14,549 | 100% | 550 [370, 730] |
| Trials | None (all observational studies) | NA | NA | NA |
| Sensitivity analysis: Round 1: Proportion with an incidental finding, excluding high risk of bias studies | | | | |
| All studies | 13 (56, 59, 61, 76-85) | 14,549 | 100% | 550 [370, 730] |
| Not high risk of bias | 3 (56, 61, 82) | 1,235 | 100% | 540 [0, 1000] |
| Between study subgroup: 2/3 Rounds: Proportion of incidental findings, by study design (test for subgroup differences, p=NA) | | | | |

| Comparison | N studies | n | $I^2$ | Absolute number (95% CI) per 1,000 |
|---|---|---|---|---|
| Observational | None (all trials) | NA | NA | NA |
| Trials | 1 (28) | 26,722 | NA | 570 [560, 570] |
| Sensitivity analysis: 2/3 Rounds: Proportion of incidental findings, excluding high risk of bias studies | | | | |
| All studies | 1 (28) | 26,722 | NA | 570 [560, 570] |
| Not high risk of bias | NA (no high risk of bias studies) | NA | NA | NA |
| Between study subgroup: 4/5 Rounds: Proportion of incidental findings, by study design (test for subgroup differences, p=0.002) | | | | |
| Observational | 1 (83) | 519 | NA | 730 [690, 770] |
| Trials | 2 (11, 13, 38) (2 publications for 1 trial used for this outcome) | 2,877 | 100% | 230 [0, 540] |
| Sensitivity analysis: 4/5 Rounds: Proportion of incidental findings, excluding high risk of bias studies | | | | |
| All studies | 3 (11, 13, 38, 83) | 3,396 | 100% | 400 [50, 750] |
| Not high risk of bias | 2 (11, 13, 38) | 2,877 | 100% | 230 [0, 540] |
| **Clinically significant incidental findings** | | | | |
| Between study subgroup: Round 1: Proportion of clinically significant incidental findings, by study design (test for subgroup differences, p=0.29) | | | | |
| Observational | 26 (59, 61, 64, 79-99) | 81,843 | 99% | 210 [180, 250] |
| Trials | 2 (28, 49) | 27,582 | 99% | 330 [120, 540] |
| Sensitivity analysis: Round 1: Proportion of clinically significant incidental findings, excluding high risk of bias | | | | |
| All studies | 28 (28, 49, 59, 61, 64, 79-99) | 109,425 | 100% | 220 [180, 260] |
| Not high risk of bias | 5 (28, 49, 61, 87, 96) | 28,821 | 97% | 290 [220, 370] |
| Within study subgroup: Round 1: Proportion of clinically significant incidental findings, by sex (test for subgroup differences, p<0.00001) | | | | |
| Men | Observational: 1 (94) | 1,874 | NA | 230 [210, 250] |
| Women | Observational: 1 (94) | 2,199 | NA | 160 [140, 170] |
| Within study subgroup: Round 1: Proportion of clinically significant incidental findings, by asbestos exposure (test for subgroup differences, p=0.0005) | | | | |
| Asbestos exposure (>1 year) | Trials: 1 (100) | 1,228 | NA | 130 [110, 150] |
| No asbestos exposure | Trials: 1 (100) | 25,290 | NA | 100 [100, 100] |
| Between study subgroup: 2/3 Rounds: Proportion of clinically significant incidental findings, by study design (test for subgroup differences, p=NA) | | | | |
| Observational | None (only a trial) | NA | NA | NA |
| Trials | 1 (28) | 26,722 | NA | 340 [330, 340] |
| Sensitivity analysis: 2/3 Rounds: Proportion of clinically significant incidental findings, excluding high risk of bias studies | | | | |

| Comparison | N studies | n | $I^2$ | Absolute number (95% CI) per 1,000 |
|---|---|---|---|---|
| All studies | 1 (28) | 26,722 | NA | 340 [330, 340] |
| Not high risk of bias | None (no high risk of bias studies) | NA | NA | NA |
| Within study subgroup: 2/3 Rounds: Proportion of clinically significant incidental findings, by age (test for subgroup differences, p<0.00001) | | | | |
| <65 years | Trials: 1 (25) | 19,612 | NA | 150 [140, 150] |
| ≥65 years | Trials: 1 (25) | 7,110 | NA | 190 [180, 200] |
| Between study subgroup: 4/5 Rounds: Proportion of clinically significant incidental findings, by study design (test for subgroup differences, p=NA) | | | | |
| Observational | 4 (83, 97, 101, 102) | 12,026 | | 80 [30, 120] |
| Trials | None (all observational studies) | NA | NA | NA |
| Sensitivity analysis: 4/5 Rounds: Proportion of clinically significant incidental findings, excluding high risk of bias studies | | | | |
| All studies | 4 (83, 97, 101, 102) | 12,026 | 99% | 80 [30, 120] |
| Not high risk of bias | 1 (102) | 5,203 | NA | 80 [70, 80] |
| **Major complications from invasive diagnostic testing** | | | | |
| Between study subgroup: Round 1: Proportion of major complications from invasive testing (LCs+FPs), by study design (test for subgroup differences, p=0.453) | | | | |
| Observational | 7 (59, 64, 103-107) | 19,795 | 87% | 1.00 [0.00, 3.4] |
| Trials | 4 (9, 36, 46, 71) | 9,892 | 82% | 0.30 [0.00, 2.60] |
| Between study subgroup: Round 1: Proportion of major complications from invasive testing among false positives, by study design (test for subgroup differences, p=0.151) | | | | |
| Observational | 6 (59, 62, 103, 105, 106, 108) | 13,040 | 0% | 0.10 [0.00, 0.50] |
| Trials | 5 (9, 36, 46, 71, 72) | 10,751 | 32% | 0.00 [0.00, 0.50] |
| Between study subgroup: 2/3 Rounds: Proportion of major complications from invasive testing (LCs+FPs), by study design (test for subgroup differences, p=0.835) | | | | |
| Observational | 4 (62, 73, 74, 101) | 48,344 | 93% | 3.10 [0.70, 6.90] |
| Trials | 1 (24) | 26,722 | NA | 3.30 [0.70, 4.10] |
| Within study subgroup: 2/3 Rounds: Proportion of major complications from invasive testing (LCs+FPs), by COPD status (test for subgroup differences, p=0.001) | | | | |
| COPD | Trials: 1 (24) | 4,632 | NA | 6.26 [4.20, 8.98] |
| No COPD | Trials: 1 (24) | 21,821 | NA | 2.70 [2.06, 3.49] |
| Within study subgroup: 2/3 Rounds: Proportion of major complications from invasive testing (LCs+FPs), by smoking status (test for subgroup differences, p=0.084) | | | | |
| Never-smoker | Observational: 1 (73) | 17,968 | NA | 1.11 [0.68, 1.72] |

| Comparison | N studies | n | $I^2$ | Absolute number (95% CI) per 1,000 |
|---|---|---|---|---|
| Ever-smoker | Observational: 1 (73) | 19,468 | NA | 1.80 [1.25, 2.50] |
| Between study subgroup: 2/3 Rounds: Proportion of major complications from invasive testing among false positives, by study design (test for subgroup differences, p=0.540) | | | | |
| Observational | 2 (62, 101) | 2,259 | NA | 0.10 [0.00, 1.30] |
| Trials | 1 (24) | 26,722 | NA | 0.40 [0.20, 0.70] |
| Between study subgroup: 4/5 Rounds: Proportion of major complications from invasive testing (LCs+FPs), by study design (test for subgroup differences, p=0.026) | | | | |
| Observational | 1 (98) | 2,471 | NA | 2.00 [0.70, 4.70] |
| Trials | 2 (9, 38) | 8,196 | NA | 0.30 [0.00, 0.90] |
| Between study subgroup: 4/5 Rounds: Proportion of major complications from invasive testing among false positives, by study design (test for subgroup differences, p=0.031) | | | | |
| Observational | 1 (98) | 2,471 | NA | 0.80 [0.10, 2.90] |
| Trials | 1 (9) | 6,583 | NA | 0.00 [0.00, 0.60] |
| **Death from invasive diagnostic testing** | | | | |
| Between study subgroup: Round 1: Proportion of deaths from invasive testing among false positives, by study design (test for subgroup differences, p=0.219) | | | | |
| Observational | 3 (73, 104, 109) | 44,123 | NA | 0.00 [0.00, 0.00] |
| Trials | 3 (11, 46, 71) | 3,309 | NA | 0.00 [0.00, 0.44] |
| Between study subgroup: 2/3 Rounds: Proportion of deaths from invasive testing among false positives, by study design (test for subgroup differences, p=0.703) | | | | |
| Observational | 1 (101) | 2,003 | NA | 0.00 [0.00, 1.80] |
| Trials | 1 (23) | 26,722 | NA | 0.20 [0.10, 0.50] |
| Between study subgroup: 4/5 Rounds: Proportion of deaths from invasive testing among false positives, by study design (test for subgroup differences, p=NA) | | | | |
| Observational | None (only a single trial) | NA | NA | NA |
| Trials | 1 (11) | 1,264 | NA | 0.00 [0.00, 2.91] |

Table S2.11 Additional harms outcome data

| Number of studies; Number enrolled/randomized to CT screening | Absolute numbers (95% CI) per 1000 |
|---|---|
| Number of false positive events | |
| Over 2/3 rounds: Trials: 3 studies; N=34,622 (23, 30, 46, 47) (2 publications from 1 trial were used for this outcome)<br>Observational: 3 studies; N=4,643 (62, 63, 67) | 500 [330, 680] |
| Over 4/5 rounds: Trials: 5 studies; N=14,653 (9, 16, 33, 38, 42)<br>Observational: 1 study; N=5,203 (110) | 350 [270, 420] |
| Proportion of false positive requiring diagnostic follow-up | |
| Round 1: Trials: 9 studies; N=21,781 (16, 19, 30, 32, 36, 40, 42, 46, 72)<br>Observational: 15 studies; N=34,660 (52, 54, 56-59, 61, 63-68, 70, 74) | 20 [10, 20] |
| Over 2/3 rounds: Trials: 2 studies; N=4,780 (30, 48)<br>Observational: 8 studies; N=55,363 (56, 58, 61-63, 70, 73, 74) | 20 [10, 30] |
| Over 4/5 rounds: Trials: 2 studies; N=2,877 (11, 38)<br>Observational: 2 studies; N=6,470 (75, 111) | 30 [10, 50] |
| Number of false positive events requiring diagnostic follow-up | |
| Over 2/3 rounds: Trials: 1 study; N=1,660 (46, 47) (2 publications from 1 trial were used for this outcome)<br>Observational: 1 study; N=1,035 (67) | 30 [20, 40] |
| Over 4/5 rounds: Trials: 3 studies; N=10,988 (9, 16, 42)<br>Observational: 0 studies | 40 [30, 50] |
| Number of incidental finding events | |
| Round 1: Trials: 2 studies; N=1,155 (71, 112)<br>Observational: 12 studies; n=21,995 (64, 66, 77, 78, 82, 83, 91, 97, 113-115) | 790 [630, 950] |
| Over 2/3 rounds: Trials: 1 study; N=26,722 (28)<br>Observational: 1 study; N=8,649 (74) | 500 [10, 1000] |
| Over 4/5 rounds: Trials: 0 studies<br>Observational: 2 studies; N=4,820 (83, 97) | 720 [0, 1570] |
| Number of clinically significant incidental finding events | |
| Round 1: Trials: 1 study; N=2,028 (19)<br>Observational: 9 studies; N=19,475 (70, 83, 84, 86, 89, 91, 93, 94, 102) | 200 [140, 250] |

| Number of studies; Number enrolled/randomized to CT screening | Absolute numbers (95% CI) per 1000 |
|---|---|
| Over 2/3 rounds: Trials: 1 study; N=26,722 (28)<br>Observational: 2 studies; N=5,275 (70, 116) | 350 [0, 700] |
| Over 4/5 rounds: Trials: 0 studies<br>Observational: 2 studies; N=5,722 (83, 102) | 100 [60, 140] |
| Proportion[‡] with specific incidental finding – coronary artery calcification | |
| Round 1: Trials: 1 study; N=770 (112)<br>Observational: 24 observational; N=54,665 (61, 76, 78, 79, 81-84, 86, 87, 89-91, 96, 114, 115, 117-124) | 340 [230, 450] |
| Over 2/3 rounds: Trials: 1 study; N=26,722 (28)<br>Observational: 0 studies | 70 [60, 70] |
| Over 4/5 rounds: Trials: 1 study; N=1,613 (13)<br>Observational: 1 study; N=519 (83) | 240 [0, 520] |
| Proportion with specific incidental finding - emphysema | |
| Round 1: Trials: 3 studies; N=3,183 (45, 71, 112)<br>Observational: 18 studies; N=242,526 (55, 61, 73, 76, 79-83, 87, 89, 96, 114, 115, 118, 124-126) | 240 [180, 290] |
| Over 2/3 rounds: Trials: 1 study; N=26,722 (28)<br>Observational: 0 studies | 210 [210, 220] |
| Over 4/5 rounds: Trials: 0 studies<br>Observational: 1 study; N=519 (83) | 120 [90, 140] |
| Proportion with specific incidental finding – thyroid abnormalities[‖] | |
| Round 1: Trials: 1 study; N=2,028 (45)<br>Observational: 18 studies; N=28,340 (56, 61, 64, 76, 78, 81-84, 87, 91, 93, 94, 96, 118, 124, 127, 128) | 15.9 [0.0, 22.2] |
| Over 2/3 rounds: Trials: 1 study; N=26,722 (28)<br>Observational: 1 study; N=2,309 (124) | 9.0 [8.0, 10.0] |
| Over 4/5 rounds: Trials: 0 studies<br>Observational: 1 study; N=519 (83) | 52.0 [36.0, 74.6] |
| Proportion with specific incidental finding – lymph node abnormalities[‖] | |
| Round 1: Trials: 1 study; N=2,028 (45)<br>Observational: 10 studies; N=14,559 (64, 70, 76, 78, 81, 82, 84, 91, 96, 115) | 19.0 [11.0, 29.0] |
| Over 2/3 rounds: Trials: 1 study; N=26,722 (28)<br>Observational: 3 studies; N=13,924 (70, 74, 116) | 50.7 [7.4, 129.3] |
| Over 4/5 rounds: no data | No data |
| Proportion with specific incidental finding – aortic aneurysm[‖] | |
| Round 1: Trials: 0 studies<br>Observational: 10 studies; N=20,023 (56, 61, 64, 78, 81, 83, 86, 89, 93, 123) | 14.0 [7.0, 22.0] |
| Over 2/3 rounds: Trials: 1 study; N=26,722 (28)<br>Observational: 1 study; N=1,520 (116) | 9.3 [8.2, 10.5] |
| Over 4/5 rounds: Trials: 0 studies<br>Observational: 1 study; N=519 (83) | 11.6 [5.3, 25.0] |

| Number of studies; Number enrolled/randomized to CT screening | Absolute numbers (95% CI) per 1000 |
|---|---|
| Proportion with specific incidental finding – cancer (excluding primary lung cancer)‖ | |
| Round 1: Trials: 1 study; N=1,264 (37)<br>Observational: 8 studies; N=162,874 (64, 81-83, 102, 109, 129, 130)<br><br><u>Specific cancers:</u><br>Thymoma (n=3)<br>Renal cell cancer (n=9)<br>Gastric cancer (n=3)<br>Thyroid cancer (n=3)<br>Malignant lymphoma (n=1)<br>Malignant mesothelioma (n=6)<br>Urinary tract malignancy (n=1)<br>Adrenal gland malignancy (n=1)<br>Metastases (n=5)<br>Non-LC malignancy (details NR, n=136)<br>**Total cancers**: **168** | 5.0 [1.0, 11.0] |
| Over 2/3 rounds: Trials: 0 studies<br>Observational: 2 studies; N=5,275 (70, 116)<br><br><u>Specific cancers:</u><br>Hodgkin's disease (n=2)<br>Lymphoma (n=4)<br>Metastasis (n=6)<br>Renal cell cancer (n=4)<br>Bronchial cancer (n=2)<br>Ovarian cancer (n=1)<br>Pancreatic cancer (n=1)<br>Breast cancer (n=3)<br>**Total cancers: 23** | 4.2 [2.6, 6.2] |
| Over 4/5 rounds: Trials: 1 study; N=1,613 (38)<br>Observational: 3 studies; N=7,725 (83, 101, 102)<br><br><u>Specific cancers:</u><br>Thymoma (n=5)<br>Metastases (n=1)<br>Malignant lymphoma (n=7)<br>Malignant schwannoma (n=1)<br>Hepatocellular cancer (n=1)<br>Gastrointestinal cancer (n=1),<br>Pancreatic cancer (n=2)<br>Prostate cancer (n=1)<br>Renal cancer (n=12)<br>Urinary tract malignancy (n=1)<br>Thyroid cancer (n=3) | 5.0 [3.0, 8.0] |

| Number of studies; Number enrolled/randomized to CT screening | Absolute numbers (95% CI) per 1000 |
|---|---|
| Breast cancer (n=1)<br>Adrenal gland malignancy (n=1)<br>Ovarian cancer (n=1)<br>Malignant mesothelioma (n=2)<br>Breast cancer (n=2)<br>Kidney cancer (n=1)<br>Esophageal cancer (n=2)<br>Head and neck cancer (n=1)<br>**Total cancers: 46** | |
| Number of major complication events from invasive testing (LCs+FPs)[ǁ] | |
| Round 1: Trials: 4 studies; N=9,892 (9, 36, 46, 71)<br>Observational: 8 studies; N=112,057 (59, 64, 103-105, 107, 131, 132) | 1.41 [0.00, 5.10] |
| Over 2/3 rounds: Trials: 2 studies; N=28,382 (23, 46)<br>Observational: 2 studies; N=37,692 (62, 73) | 5.34 [1.19, 12.00] |
| Over 4/5 rounds: Trials: 1 study; N=6,583 (9)<br>Observational: 0 studies | 0.00 [0.00, 0.56] |
| Number of major complication events from invasive testing among FPs[ǁ] | |
| Round 1: Trials: 5 studies; N=35,813 (9, 25, 36, 71, 72)<br>Observational: 5 studies; N=3,774 (59, 62, 103, 105, 108) | 0.00 [0.00, 0.00] |
| Over 2/3 rounds: Trials: 2 studies; N=28,382 (23, 46)<br>Observational: 1 study; N=256 (62) | 0.85 [0.47, 1.31] |
| Over 4/5 rounds: no evidence | No evidence. |
| Proportion with deaths from invasive testing (LCs+FPs)[ǁ] | |
| Round 1: Trials: 2 studies; N=2,045 (46, 71)<br>Observational: 3 studies; N=125,769 (73, 104, 131) | 0.00 [0.00, 0.12] |
| Over 2/3 rounds: Trials: 1 study; N=26,722 (23)<br>Observational: 1 study; N=37,436 (73) | 0.13 [0.05, 0.23] |
| Over 4/5 rounds: Trials: 1 study; N=1,613 (39)<br>Observational: 0 studies | 3.72 [1.37, 8.08] |

[ǂ] Specific incidental findings were assumed to be proportions (proportions and events are pooled together).

[ǁ] Rare events were analyzed in STATA with the exact method.

Table S2.12 Targeted CT screening: Summary of findings for benefit outcomes

| **Outcome**<br><br>**Trial**<br><br>**Number of participants; follow-up**<br><br>**Threshold** | **Relative effects, RR (95% CI)** | **Anticipated absolute effects** | | | **Certainty of evidence (GRADE[ǂ])** | **What happens?** |
|---|---|---|---|---|---|---|
| | | **With screening** | **Without screening** | **Difference** | | |
| Lung cancer mortality (Fig. S2.45)<br><br>ECLS (Early-CDT test autoantibodies (CT for positive screen and x-ray normal/negative every 6 mos x 4)<br><br>N=12,209; 5 years follow-up<br><br>Threshold: **1 fewer per 1000** | 0.82 [0.60, 1.11] | 12.1 per 1000 (8.8 to 16.3) | 14.7 per 1000 | 2.7 fewer (5.9 fewer to 1.6 more) per 1000 | **Very low**<br>⨁⊖⊖⊖[a]<br>Due to very serious indirectness (timing insufficient), and very serious imprecision | We are very uncertain about the effects from targeted screening strategies on lung-cancer mortality. |
| Lung cancer mortality (Fig. S2.46)<br><br>LungSEARCH (Annual sputum cytology/cytometry (CT and autofluorescence bronchoscopy if positive), all COPD<br><br>N=1,568; 5 years follow-up<br><br>Threshold: **1 fewer per 1000** | 0.76 [0.40, 1.45] | 20.4 per 1000 (10.7 to 38.9) | 26.8 per 1000 | 6.44 fewer (16.09 fewer to 12.07 more) per 1000 | **Very low**<br>⨁⊖⊖⊖[b]<br>Due to very serious indirectness (timing insufficient and population), and very serious imprecision | |
| All-cause mortality (Fig. S2.47)<br><br>ECLS<br><br>N=12,209; 5 years follow-up<br><br>Thresholds: **0.5 and 1 fewer per 1000** | 0.89 [0.77, 1.03] | 56.4 per 1000 (48.8 to 65.3) | 63.4 per 1000 | 7.0 fewer (14.6 fewer to 1.9 more) per 1000 | **Very low**<br>⨁⊖⊖⊖[c]<br>Due to very serious indirectness (timing insufficient), and very serious imprecision | We are very uncertain about the effects from targeted screening strategies on all-cause mortality. |
| All-cause mortality (Fig. S2.48)<br><br>LungSEARCH | 0.73 [0.54, 0.97] | 89.50 per 1000 (66.2 to 118.9) | 122.6 per 1000 | 33.1 fewer (56.4 to 3.7 fewer) per 1000 | **Very low**<br>⨁⊖⊖⊖[d]<br>Due to very serious indirectness (timing | |

| | | | | | |
|---|---|---|---|---|---|
| N=1,568; 5 years follow-up<br><br>Thresholds: **0.5 and 1 fewer per 1000** | | | | | insufficient and population), and imprecision |

[ǂ]**GRADE ratings:**

[a] <u>Lung cancer mortality (ECLS trial)</u>: ***No serious concerns about risk of bias:*** The ECLS trial was not high risk of bias for this outcome. ***No serious concerns about inconsistency:*** No evidence of inconsistency (a single trial contributed to this outcome). ***Very serious concerns about indirectness:*** There were serious concerns about insufficient timing for this outcome (up to 5 years follow-up). ***Very serious concerns about imprecision:*** The upper confidence interval crosses two thresholds (1 fewer and 1 more). ***No serious concerns about reporting bias:*** No serious concerns about reporting bias.

[b] <u>Lung cancer mortality (LungSEARCH trial)</u>: ***No serious concerns about risk of bias:*** The LungSEARCH trial was not high risk of bias for this outcome. ***No serious concerns about inconsistency:*** No evidence of inconsistency (a single trial contributed to this outcome). ***Very serious concerns about indirectness:*** There were very serious concerns about insufficient timing for this outcome (median 5 years follow-up), and the population all has COPD. ***Very serious concerns about imprecision:*** The upper confidence interval crosses the threshold. ***No serious concerns about reporting bias:*** No serious concerns about reporting bias.

[c] <u>All-cause mortality (ECLS trial)</u>: ***No serious concerns about risk of bias:*** The ECLS trial was not high risk of bias for this outcome. ***No serious concerns about inconsistency:*** No evidence of inconsistency (a single trial contributed to this outcome). ***Very serious concerns about indirectness:*** There were very serious concerns about insufficient timing for this outcome (up to 5 years follow-up). ***Very serious concerns about imprecision:*** The upper confidence interval crosses two thresholds (1 fewer and 1 more). ***No serious concerns about reporting bias:*** No serious concerns about reporting bias.

[d] <u>All-cause mortality (LungSEARCH trial)</u>: ***No serious concerns about risk of bias:*** The LungSEARCH trial was not high risk of bias for this outcome. ***No serious concerns about inconsistency:*** No evidence of inconsistency (a single trial contributed to this outcome). ***Very serious concerns about indirectness:*** There were very serious concerns about insufficient timing for this outcome (median 5 years follow-up), and the population all has COPD. ***Serious concerns about imprecision:*** The trial sample size was inadequate (as per GRADE guideline 11). ***No serious concerns about reporting bias:*** No serious concerns about reporting bias.

| Study or Subgroup | LDCT Events | LDCT Total | Control Events | Control Total | Weight | Risk Ratio M-H, Random, 95% CI |
|---|---|---|---|---|---|---|
| ECLS (Sullivan 2025, maximum 5 y) | 73 | 6088 | 90 | 6121 | 100.0% | 0.82 [0.60, 1.11] |
| **Total (95% CI)** | | **6088** | | **6121** | **100.0%** | **0.82 [0.60, 1.11]** |
| Total events | 73 | | 90 | | | |
| Heterogeneity: Not applicable | | | | | | |
| Test for overall effect: Z = 1.30 (P = 0.19) | | | | | | |

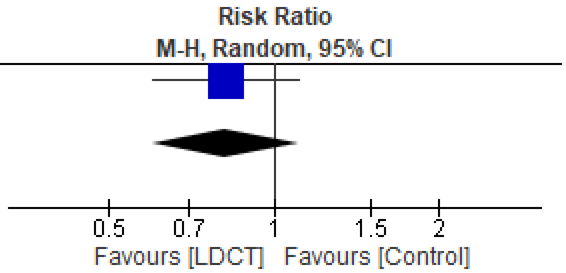

Figure S2.45 Lung cancer mortality, ECLS trial

| Study or Subgroup | LDCT Events | LDCT Total | Control Events | Control Total | Weight | Risk Ratio M-H, Random, 95% CI |
|---|---|---|---|---|---|---|
| LungSEARCH (Spiro 2019, median 5 y) | 16 | 785 | 21 | 783 | 100.0% | 0.76 [0.40, 1.45] |
| **Total (95% CI)** | | **785** | | **783** | **100.0%** | **0.76 [0.40, 1.45]** |
| Total events | 16 | | 21 | | | |
| Heterogeneity: Not applicable | | | | | | |
| Test for overall effect: Z = 0.84 (P = 0.40) | | | | | | |

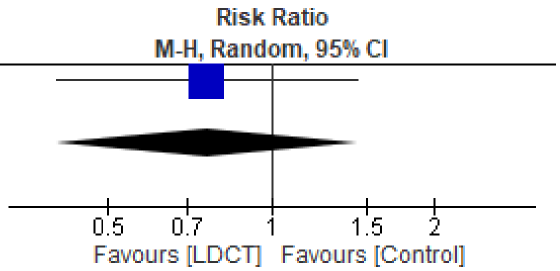

Figure S2.46 Lung cancer mortality, LungSEARCH trial

| Study or Subgroup | LDCT Events | LDCT Total | Control Events | Control Total | Weight | Risk Ratio M-H, Random, 95% CI |
|---|---|---|---|---|---|---|
| ECLS (Sullivan 2025, maximum 5 y) | 344 | 6088 | 388 | 6121 | 100.0% | 0.89 [0.77, 1.03] |
| **Total (95% CI)** | | **6088** | | **6121** | **100.0%** | **0.89 [0.77, 1.03]** |
| Total events | 344 | | 388 | | | |
| Heterogeneity: Not applicable | | | | | | |
| Test for overall effect: Z = 1.60 (P = 0.11) | | | | | | |

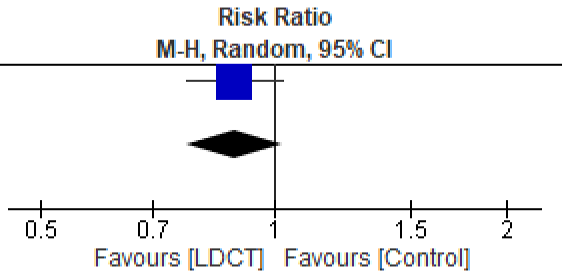

Figure S2.47 All-cause mortality, ECLS trial

| Study or Subgroup | LDCT Events | LDCT Total | Control Events | Control Total | Weight | Risk Ratio M-H, Random, 95% CI |
|---|---|---|---|---|---|---|
| LungSEARCH (Spiro 2019, median 5 y) | 70 | 785 | 96 | 783 | 100.0% | 0.73 [0.54, 0.97] |
| **Total (95% CI)** | | **785** | | **783** | **100.0%** | **0.73 [0.54, 0.97]** |
| Total events | 70 | | 96 | | | |
| Heterogeneity: Not applicable | | | | | | |
| Test for overall effect: Z = 2.14 (P = 0.03) | | | | | | |

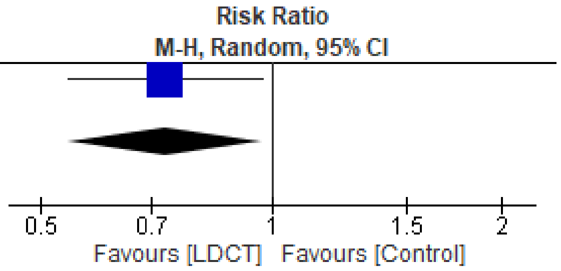

Figure S2.48 All-cause mortality, LungSEARCH trial

### Additional file 3. Additional tables and figures for key question 2

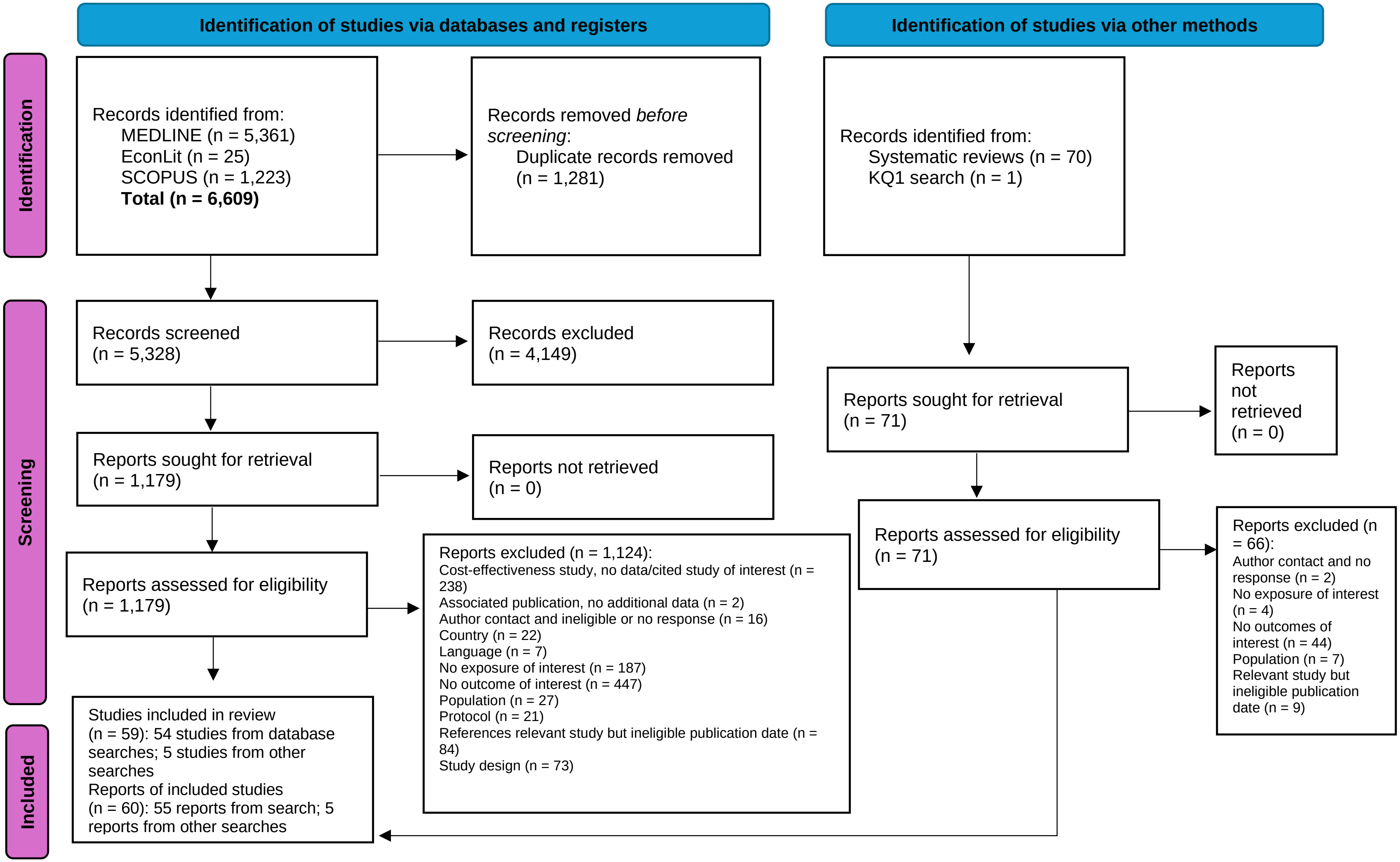
Identification of studies via databases and registers
Identification of studies via other methods
Identification
Screening
Included
Records identified from:
MEDLINE (n = 5,361)
EconLit (n = 25)
SCOPUS (n = 1,223)
Total (n = 6,609)
Records removed before screening:
Duplicate records removed (n = 1,281)
Records identified from:
Systematic reviews (n = 70)
KQ1 search (n = 1)
Records screened (n = 5,328)
Records excluded (n = 4,149)
Reports sought for retrieval (n = 1,179)
Reports not retrieved (n = 0)
Reports sought for retrieval (n = 71)
Reports not retrieved (n = 0)
Reports assessed for eligibility (n = 1,179)
Reports excluded (n = 1,124):
Cost-effectiveness study, no data/cited study of interest (n = 238)
Associated publication, no additional data (n = 2)
Author contact and ineligible or no response (n = 16)
Country (n = 22)
Language (n = 7)
No exposure of interest (n = 187)
No outcome of interest (n = 447)
Population (n = 27)
Protocol (n = 21)
References relevant study but ineligible publication date (n = 84)
Study design (n = 73)
Reports assessed for eligibility (n = 71)
Reports excluded (n = 66):
Author contact and no response (n = 2)
No exposure of interest (n = 4)
No outcomes of interest (n = 44)
Population (n = 7)
Relevant study but ineligible publication date (n = 9)
Studies included in review (n = 59): 54 studies from database searches; 5 studies from other searches
Reports of included studies (n = 60): 55 reports from search; 5 reports from other searches

Figure S3.1 Literature flow diagram

Table S3.1 Characteristics of health state utility studies

| Study<br><br>Country (setting) | Inclusion criteria<br>Age (mean/median)<br>Sex (% female)<br>Smoking status<br>Comorbidities<br>Race/ethnicity<br>LC risk factors<br>Stages included<br>Type of LC included | Number of participants | Methods for preferences;<br>Experienced vs hypothetical health states | Relevant heath states valued (timepoint used) | Subgroups |
|---|---|---|---|---|---|
| Bendixen 2019<br><br>Denmark (Public university-based cardiothoracic surgery department) | Patients who were scheduled for lobectomy for stage I non-small-cell lung cancer with curative intent. Only patients with performance status 0–1 were eligible, and co morbidities were not a reason for exclusion, except for alcoholism or ability to ambulate freely.<br><br>Age, median (IQR):<br>VATs arm: 66 (41-83)<br>Anterolateral thoracotomy arm: 65 (48-85)<br><br>Female: 51.7%<br><br>Smoker: 83.6%<br>Non-smoker: 16.4%<br><br>Comorbidities: NR<br><br>Race/ethnicity: NR<br><br>LC risk factors: NR<br><br>Stage I: 100%<br><br>Non-small cell: 100% | 201 (99 VATs arm, 102 anterolateral thoracotomy arm)<br>*both timepoints, with imputation | EQ-5D (Danish tariff; TTO); 0 to 1<br><br>In order to obtain a measure for the quality of life, each EQ 5D-3L health state was assigned a single-index score anchored at 0 for a state equivalent to being dead. The maximum index score is 1 for full health. The index score is <0 for health states regarded as worse than being dead. The index score is calculated using a formula that attaches weights to each of the levels in each dimension based on valuations by the general Danish population [13]. The Danish valuation set is based on the time trade-off technique [13]. Multiple imputation (MI) with 10 iterations was used to impute all missing values for the quality of life. We used 4 cycles of imputed data, and the variables used were the EQ-5D-3l index scores.<br><br>Experienced (patients) | **Stage I non-small-cell lung cancer:**<br>T1: Pre-treatment<br>T3: 2 weeks post-surgery | None. |
| Bertolaccini 2023<br><br>Italy (Italian VATS lobectomy database [50 participating centers]) | The inclusion criteria were: VATS lobectomy procedures intended as a primary procedure for pulmonary lobectomy; (performed consecutively during study timeframe.<br><br>Age, mean (SD): 67.9 (10.0)<br><br>Female: 40.2%<br><br>Current smoker: 66.8% | 11,772 | EQ-5D-5L (NR); range NR<br><br>No additional information provided on scale, tariff used, etc.<br><br>Experienced (patients) | **Stage I-IIIA non-small cell lung cancer:**<br>T3: At hospital discharge for surgery | None. |

| Study<br><br>Country (setting) | Inclusion criteria<br>Age (mean/median)<br>Sex (% female)<br>Smoking status<br>Comorbidities<br>Race/ethnicity<br>LC risk factors<br>Stages included<br>Type of LC included | Number of participants | Methods for preferences;<br>Experienced vs hypothetical health states | Relevant heath states valued (timepoint used) | Subgroups |
|---|---|---|---|---|---|
| | Packages per year, mean (SD): 39.64 (44.33)<br><br>Comorbidities: NR<br><br>Race/ethnicity: NR<br><br>LC risk factors: NR<br><br>In situ: 0.2%<br>Stage IA: 25.6%<br>Stage IB: 29.5%<br>Stage IC: 13.7%<br>Stage II: 0.8%<br>Stage IIA: 18.1%<br>Stage IIB: 5.2%<br>Stage III: 5.4%<br>Stage IV: 1.4%<br><br>Non-small cell: 100% | | | | |
| Black 2014<br><br>USA (16 of 23 ACRIN screening sites in the NLST trial) | Subset of NLST trial participants: To estimate quality adjusted life expectancy for each screening strategy, we used utilities derived from the Short Form (SF)-6D, a subset of the SF-36, which was collected at 16 of 23 ACRIN screening sites.<br><br>Age: NR<br><br>Female: NR<br><br>Pack-years smoking history: 30 pack-years (inclusion criteria)<br><br>Comorbidities: NR<br><br>Race/ethnicity: NR<br><br>LC risk factors: NR<br><br>Stage: NA | 11,696 | SF-6D (UK tariff; SG); range NR<br><br>To estimate quality adjusted life expectancy for each screening strategy, we used utilities derived from the Short Form (SF)-6D, a subset of the SF-36, which was collected at 16 of 23 ACRIN screening sites<br><br>Experienced (patients) | **Comparator, NLST participants:**<br>Before screening | None. |

| Study<br><br>Country (setting) | Inclusion criteria<br>Age (mean/median)<br>Sex (% female)<br>Smoking status<br>Comorbidities<br>Race/ethnicity<br>LC risk factors<br>Stages included<br>Type of LC included | Number of participants | Methods for preferences;<br>Experienced vs hypothetical health states | Relevant heath states valued (timepoint used) | Subgroups |
|---|---|---|---|---|---|
| | Type of LC: NA | | | | |
| Chouaid 2013<br><br>8 countries (all VHDI sites) | Patients were included if they had an ECOG performance status of 0 to 2, and were aged 18 years or older. Patients should have been receiving first-, second-, or third-/fourth-line pharmacotherapy or best supportive care (BSC; BSC patients were required to have failed at least 2 lines of pharmacotherapy) and had to have received two cycles of their current pharmacotherapy (equivalent to 6–8 weeks ±3 days of therapy) or equivalent duration of BSC and have previously been diagnosed with advanced NSCLC (stage IIIB/IV).<br><br>Age: NR for sample of interest<br><br>Female: NR for sample of interest<br><br>Smoking status: NR for sample of interest<br><br>Comorbidities: NR for sample of interest<br><br>Race/ethnicity: NR<br><br>LC risk factors: NR<br><br>Stage IIIB or IV: 100%<br><br>Non-small cell: 100% | 111 (first-line, progression-free) | EQ-5D (UK tariff; TTO); range NR<br><br>The EQ-5D measures five dimensions of health (mobility, self-care, usual activities, pain/ discomfort, anxiety/depression) within three levels of severity (no/some/extreme problems). The quality-of-life scores in the EQ-5D were transformed into utility values by using modeling valuations developed by Dolan et al.12 with EQ-5D value weights elicited from a U.K. population.<br><br>Experienced (patients) | **Stage IIIB or IV non-small-cell lung cancer:**<br>T2: On first-line therapy | None. |
| Erbaycu 2018<br><br>Western Turkey (inpatient setting or at the outpatient clinics of five comprehensiv e cancer | Stage IIIB or IV cases (including all histological types) diagnosed with primary lung cancer; cases that received no previous treatment and are planned to be given chemotherapy (CT), chemoradiotherapy (CRT) or radiotherapy (RT); cases who were literate, could read and fill in the test forms and accepted to participate in the study and to come for examinations; cases who signed the written consent form for voluntary participation.<br><br>Age, mean (SD): 61.35 (9.57) | 266 | EQ-5D (UK tariff; TTO); range NR<br><br>EQ-5D index scale; consist of five dimensions namely; mobility, self-care, usual activities, pain/discomfort and anxiety/depression. Each item on the scale is assessed on three levels (Likert type). As well as obtaining response distributions of these dimensions; health availability index scores can be calculated. EQ-5D questionnaire was filled in by the patient | **Stage IIIB or IV lung cancer:**<br>T1: At diagnosis | **Subgroup:** gender; smoking status; age groups; tumor cell type (7 types); stage of disease (IIIB vs IV) |

| Study<br><br>Country (setting) | Inclusion criteria<br>Age (mean/median)<br>Sex (% female)<br>Smoking status<br>Comorbidities<br>Race/ethnicity<br>LC risk factors<br>Stages included<br>Type of LC included | Number of participants | Methods for preferences;<br>Experienced vs hypothetical health states | Relevant heath states valued (timepoint used) | Subgroups |
|---|---|---|---|---|---|
| centers [AKAYAK-1 Multicenter Project]) | Female: 6.8%<br><br>Smoker: 19.5%<br>Past smoker ("gave up"): 75.6%<br><br>Comorbidities: 35.3%<br><br>Race/ethnicity: NR<br><br>LC risk factors: NR<br><br>Stage IIIB: 22.3%<br>Stage IV: 67.7%<br><br>Small-cell: 21.0%<br>Non-small cell: 77.1%<br>Non-specified: 1.9% | | prior to the doctor's examination, following a detailed explanation in an empty and quiet room (no further details).<br><br>Experienced (patients) | | |
| Galetta 2015<br><br>Italy (14 Italian centers) | ECOG performance status (PS) of 0 to 1; age 18 to 70 years; measurable disease according to Response Evaluation Criteria in Solid Tumors (RECIST; version 1.1)10 that had not been previously irradiated; a life expectancy longer than 12 weeks; adequate bone marrow, renal, and hepatic functions; and coagulation parameters with International Normalized Ratio <= 1.5 and Partial Thromboplastine Time <= 1.5 times the upper normal laboratory range.<br><br>Age, median (range): 62 (35-72)<br><br>Female: 26.0%<br><br>Smoker: 65.0%<br>Non-smoker: 25.0%<br>Unknown smoking status: 10.0%<br><br>Comorbidities: NR<br><br>Race/ethnicity: NR | T1: 118;<br>T2: 96;<br>T3: 52 | EQ-5D-3L (NR); range NR<br><br>The EQ5D consists of 2 pages: the EQ5D descriptive system and the EQ5D-VAS. The EQ5D descriptive system comprises 5 dimensions: mobility, self-care, usual activities, pain/discomfort, and anxiety/depression. Each dimension has 3 levels: no problems, some problems, and severe problems. The respondent was asked to indicate his/her health state by placing a cross in the box against the most appropriate statement in each of the 5 dimensions. A summary index with a maximum score of 1 was derived from these 5 dimensions by conversion with the table of scores by Rabin and de Charro.6<br><br>Experienced (patients) | **Stage IV non-small cell lung cancer:**<br>T1: Prior to first-line therapy<br>T2: 6th cycle of first-line therapy<br>T3: Week 12 and 18 of maintenance therapy | None. |

| Study<br><br>Country (setting) | Inclusion criteria<br>Age (mean/median)<br>Sex (% female)<br>Smoking status<br>Comorbidities<br>Race/ethnicity<br>LC risk factors<br>Stages included<br>Type of LC included | Number of participants | Methods for preferences;<br>Experienced vs hypothetical health states | Relevant heath states valued (timepoint used) | Subgroups |
|---|---|---|---|---|---|
| | LC risk factors: NR<br><br>Stage IIIB: 6.0%<br>Stage IV: 94.0%<br><br>Non-small cell: 100% | | | | |
| Ganti 2025<br><br>United States (National Cancer Institute National Clinical Trials Network) | Age requirement was ≥ 18 years of age. Histologically or cytologically documented small cell lung cancer (SCLC). Limited-stage disease patients with disease restricted to one hemithorax with regional lymph node metastases, including ipsilateral hilar, ipsilateral and contralateral mediastinal, and ipsilateral supraclavicular lymph nodes. Patients with disease involvement of the contralateral hilar or supraclavicular lymph nodes are not eligible. Patients with pleural effusions that are visible on plain chest radiographs, whether cytologically positive or not are not eligible unless they have a negative thoracentesis. Patients with cytologically positive pleural or pericardial fluid, regardless of the appearance on plain x-ray are not eligible. Measurable disease - Patients must have measurable disease, which includes lesions that can be accurately measured in at least one dimension (longest diameter to be recorded) as ≥ 2 cm with conventional techniques OR ≥ 1 cm by spiral CT scan (additional treatment requirements reported in trial registry: NCT00632853).<br><br>Age, median (IQR)(n=383): 63.0 (37.0, 81.0)<br><br>Female: 51.5%<br><br>Smoking status: NR<br><br>Comorbidities: NR<br><br>Race:<br>White: 87.0%<br>Black or African American: 8.9%<br>Asian: 0.3% | T2: 209 | EQ-5D (NR); range 0 to 1<br><br>No further details.<br><br>Experienced (patients) | **Limited stage small-cell lung cancer:**<br>T2: Week 5 or 7 of first-line therapy | None. |

| Study<br><br>Country (setting) | Inclusion criteria<br>Age (mean/median)<br>Sex (% female)<br>Smoking status<br>Comorbidities<br>Race/ethnicity<br>LC risk factors<br>Stages included<br>Type of LC included | Number of participants | Methods for preferences;<br>Experienced vs hypothetical health states | Relevant heath states valued (timepoint used) | Subgroups |
|---|---|---|---|---|---|
| | American Indian or Alaskan Native: 0.9%<br>Unknown or NR: 3.0%<br><br>Ethnicity:<br>Hispanic or Latino: 3.3%<br>Non-Hispanic: 92.3%<br>Unknown or NR: 4.4%<br><br>LC risk factors: NR<br><br>Limited stage: 100%<br><br>Small-cell: 100% | | | | |
| Gridelli 2012<br><br>16 countries (83 investigational sites) | To receive induction treatment, patients were required to have a histologic or cytologic diagnosis of advanced (stage IIIB/IV) nonsquamous NSCLC based on the Lung Cancer Staging System version 5.13 Additional eligibility criteria included no prior systemic chemotherapy for lung cancer, including adjuvant; an Eastern Cooperative Oncology Group (ECOG) performance status of 0 or 114; one or more measurable lesion(s) per Response Evaluation Criteria in Solid Tumors (1.0); adequate organ function; and age of 18 years or older. Patients receiving concurrent administration of other antitumor therapy were ineligible for the study.<br>After completing four cycles of induction treatment, patients were eligible to receive maintenance therapy if they had an ECOG performance status of 0 or 1 and documented radiographic evidence of complete response (CR), partial response (PR), or stable disease (SD). The response did not have to be confirmed for the patient to be randomized.<br><br>Age, median (IQR) induction phase (n=939): 61 (24-83)<br>Age, median (IQR) maintenance phase (n=539):<br>Pemetrexed arm: 61 (32-79)<br>Placebo arm: 62 (35-83)<br><br>Female: | T1: 445;<br>T2: 445;<br>T3: 397 | EQ-5D-3L (UK; TTO); -0.59 to 1<br><br>The EQ-5D is a self-administered health-status questionnaire consisting of two parts.12 The first part includes five descriptive questions with which the patient rates his/her health regarding mobility, self care, usual activities, pain/discomfort, and anxiety/depression. Each of the five attributes has three levels: no problem, some problems, and major problems, thus defining 243 possible health states, to which the EuroQol committee added unconscious and dead for a total of 245. The possible values for health utility ranges from −0.59 (severe problems in all 5 dimensions) to 1.0 (no problem in any dimension) on a scale in which 0 represents death and 1 represents the best possible health state.<br>All patients who provided a baseline assessment and at least one subsequent assessment were included in the EQ-5D analysis. Compliance was defined as the number of completed EQ-5D assessments | **Stage IIIB or IV non-small cell lung cancer:**<br>T1: Pre-treatment<br>T2: On first-line therapy (cycle 1)<br>T3: Maintenance therapy after first-line therapy (cycle 1) | None. |

| Study<br><br>Country (setting) | Inclusion criteria<br>Age (mean/median)<br>Sex (% female)<br>Smoking status<br>Comorbidities<br>Race/ethnicity<br>LC risk factors<br>Stages included<br>Type of LC included | Number of participants | Methods for preferences;<br>Experienced vs hypothetical health states | Relevant heath states valued (timepoint used) | Subgroups |
|---|---|---|---|---|---|
| | Induction phase: 39.0%<br>Maintenance subsample: 41.9%<br><br>Induction phase:<br>Ever smoker: 81%<br>Never smoker: 19%<br>Unknown: 0.7%<br><br>Maintenance subsample:<br>Ever smoker: 77.7%<br>Never smoker: 21.5%<br>Unknown: 0.7%<br><br>Comorbidities: NR<br><br>Induction phase:<br>Asian: 6%<br>African: 0.7%<br>White: 93%<br>Multiple: 0.2%<br><br>Maintenance subsample:<br>Asian: 4.5%<br>African: 0.9%<br>White: 94.6%<br>Multiple: 0.0%<br><br>LC risk factors: NR<br><br>Induction phase:<br>Stage IIIB: 11%<br>Stage IV: 89%<br><br>Maintenance subsample:<br>Stage IIIB: 9.3%<br>Stage IV: 90.7% | | divided by the number of expected EQ-5D assessments (i.e., patients still on study at that time).<br><br>Experienced (patients) | | |

| Study<br><br>Country (setting) | Inclusion criteria<br>Age (mean/median)<br>Sex (% female)<br>Smoking status<br>Comorbidities<br>Race/ethnicity<br>LC risk factors<br>Stages included<br>Type of LC included | Number of participants | Methods for preferences;<br>Experienced vs hypothetical health states | Relevant heath states valued (timepoint used) | Subgroups |
|---|---|---|---|---|---|
| | Non-small cell: 100% | | | | |
| Iyer 2013<br><br>France & Germany | Patients with advanced (stage IIIB/IV) NSCLC in France and Germany, who were receiving drug treatment for lung cancer in non-clinical trial settings.<br><br>Age, mean: NR by sample of interest<br><br>Female: NR by sample of interest<br><br>Smoking status: NR by sample of interest<br><br>Comorbidities: NR<br><br>Race/ethnicity: NR by sample of interest<br><br>LC risk factors: NR<br><br>Stage IIIB or IV: 100%<br><br>Non-small cell: 100% | 435 | EQ-5D-3L (NR); -0.59 to 1<br><br>Generic QOL and general health status were assessed using the EuroQol five-dimensional questionnaire (EQ-5D) [14]. The EQ 5D consists of a 5-item descriptive system (including mobility, self-care, usual activities, pain/discomfort and anxiety/depression), with each item rated as no problems, some problems or severe problems. EQ-5D utility index scores were calculated using the recommended algorithm and the range between −0.59 and 1, where 0 represents 'death' and a score of 1 indicates perfect health. Negative values indicate a state worse than death.<br><br>Experienced (patients) | **Stage IIIB or IV non-small-cell lung cancer:**<br>T2: On first-line therapy | None. |
| Jeppesen 2018<br><br>Denmark (Department of Oncology, Odense University Hospital) | The study included all consecutive patients with histologically/cytologically proven T1-2N0M0 NSCLC referred to the Department of Oncology, OUH for treatment with SBRT between January 2015 and June 2016. The patients had either refused surgery, or were considered poor surgical candidates based on a multidisciplinary assessment. Since most patients were considered too frail for surgery there was no age limit for inclusion.<br><br>Age, median (range): 72 (52-87)<br><br>Female: 47%<br><br>Pack-years smoking history, median (range): 43 (0-100)<br><br>Charlson Comorbidity Index: | T1: 51<br>T3: 41 | EQ-5D-5L (Denmark; TTO); -0.624 to 1<br><br>The EQ-5D questionnaire was used [22]. It consists of an objective and a subjective component. The objective health index score is based on five questions of the EQ-5D and each question represents one dimension of health-related QoL (mobility, self-care, usual activities, pain/discomfort and anxiety/depression). In each dimension a respondent can belong to one of five categories. Combinations of these categories result in 3125 (55) objective health index scores, which was mapped to a QoL index using a validated method for a Danish population [23]. The index ranging from −0.624 to 1, where 1 represents best possible health | **Stage I non-small cell lung cancer:**<br>T1: Prior to first-line therapy<br>T3: On maintenance therapy after first-line therapy | None. |

| Study<br><br>Country (setting) | Inclusion criteria<br>Age (mean/median)<br>Sex (% female)<br>Smoking status<br>Comorbidities<br>Race/ethnicity<br>LC risk factors<br>Stages included<br>Type of LC included | Number of participants | Methods for preferences;<br>Experienced vs hypothetical health states | Relevant heath states valued (timepoint used) | Subgroups |
|---|---|---|---|---|---|
| | 0-1: 43%<br>2-3: 45%<br>4+: 12%<br><br>Race/ethnicity: NR<br><br>LC risk factors: NR<br><br>Stage IA 78%<br>Stage IB: 20%<br>Synchronous cancers: 2%<br><br>Non-small cell: 100% | | and 0 represents dead. Health state b0 is considered worse than being death.<br><br>Experienced (patients) | | |
| Kim 2018<br><br>South Korea (except Jeju island) | The target population comprised adults aged ≥19 years who lived in Korea (except Jeju Island) using a multi-stage stratified quota sampling based on region, sex, and age group.<br><br>Age, mean (SD): 45.7 (13.9)<br><br>Female: 51.1%<br><br>Smoking history: NR<br><br>Morbidity: 11.5%<br><br>Race/ethnicity: NR<br><br>LC risk factors: NR<br><br>Stage: NA (hypotheticals are presented by stage for stage I-IV, and a pulmonary nodule during screening)<br><br>Type of LC: NA (hypotheticals are non-small cell) | 515 | SG (choice-based in general population); 0 to 1<br><br>Participants were surveyed through face-to-face computer-assisted interviews. Participants used the SG method to evaluate the six health states in random order. The health state descriptions were printed on cards to facilitate participants with limited vision. Before conducting the survey, the interviewers were educated about the purpose of the study and the descriptions of the six health states and the total education time was approximately 2.5 h. For the SG, the participants were asked to select the better of two choices: either a health state or death. If the participant chose the proposed health state as the better option, he/she was asked to determine the point of indifference between two hypothetical options: remaining in the proposed health state for the remaining life time, or receiving treatment that might lead to either a full recovery of health | **Screening health states (in those with positive screening results only):**<br>1) After a positive screening result (diagnosis not known)<br><br>**Stage I-IIIA non-small cell lung cancer:**<br>T1: At diagnosis (describes what treatments will be received and prognosis)<br><br>**Stage IIIB-IV non-small cell lung cancer:** | Subgroups (by participants evaluating the health states):<br><br>Age (19-29y; 30-39y; 40-49y; 50-59y; ≥60y)<br><br>Gender (men vs. women) |

| Study<br><br>Country (setting) | Inclusion criteria<br>Age (mean/median)<br>Sex (% female)<br>Smoking status<br>Comorbidities<br>Race/ethnicity<br>LC risk factors<br>Stages included<br>Type of LC included | Number of participants | Methods for preferences;<br>Experienced vs hypothetical health states | Relevant heath states valued (timepoint used) | Subgroups |
|---|---|---|---|---|---|
| | | | (probability 'p') or immediate death (probability '1-p'). The probability 'p' increased or decreased by 5% according to the previous participants' choice. The participants continued to select options until their preferences for the options became equal. The initial probability 'p' was 50%. If the participant selected death as a preferred state, the next SG question was presented.<br><br>Hypotheticals (general population) | T1: At diagnosis (describes what treatments will be received and prognosis) | |
| Koide 2019 | Consecutive NSCLC patients who underwent VATS were recruited prospectively for this study after providing informed consent and procedures securing respondents' anonymity were applied.<br><br>Age, mean (SD): 68.0 (9.1)<br><br>Female: 58.3%<br><br>Smoking status: NR<br><br>Comorbidities: NR<br><br>Race/ethnicity: NR<br><br>LC risk factor: NR<br><br>Stage I: 75.0%<br>Stage II: 16.7%<br>Stage III: 8.3%<br><br>Non-small cell: 100% | 24 | EQ-5D-5L (Japan; NR0; 0 to 1<br><br>The EQ-5D-5L questionnaire, the Japanese version of which has been validated in a previous work, was used with face-to-face interviews to estimate the utility values of QOL. The five dimensions assessed by the EQ-5D-5L are mobility (MO), self-care (SC), usual activities (UA), pain/discomfort (PD), and anxiety/depression (AD), each of which has five levels of severity. The level 1 is able to do without any difficulty or to feel comfortable. The level 5 is hard to do any task or feel uncomfortable. Using the scoring function from Japan, these health state parameters were transformed into a utility value ranging from 0 to 1, in which 0 represented death and 1 indicated full health.<br><br>Experienced (patients) | **Stage I-IIIA non-small cell lung cancer:**<br>T1: Pre-treatment<br>T3: At hospital discharge for surgery | None. |
| Kuehne 2022 | Eligibility criteria were histologically diagnosed SCLC, ability to consent, and ability to complete questionnaires in English. Patients were approached and enrolled at any point during the | T1: 34<br>T2: 65 | EQ-5D-5L (Canada; TTO); range -0.148 to 0.949 | **Limited stage small-cell lung cancer:**<br>T1: Newly diagnosed and treatment naive | None. |

| Study<br><br>Country (setting) | Inclusion criteria<br>Age (mean/median)<br>Sex (% female)<br>Smoking status<br>Comorbidities<br>Race/ethnicity<br>LC risk factors<br>Stages included<br>Type of LC included | Number of participants | Methods for preferences;<br>Experienced vs hypothetical health states | Relevant heath states valued (timepoint used) | Subgroups |
|---|---|---|---|---|---|
| Canada (Toronto, Ontario Princess Margaret Cancer Centre) | disease course, including while on treatment, and during follow-up.<br><br>Age:<br>Limited stage, median (range): 65 (32-88)<br>Extensive stage, median (range): 66 (48-91)<br><br>Female: 42.3%<br><br>Never smoker: 7.2%<br>Past smoker: 63.1%<br>Current smoker: 29.7%<br>Limited stage, mean (SD): 48.5 (27)<br>Extensive stage, mean (SD): 55.0 (35.0)<br><br>Charlson Comorbidity Index:<br>0: 45.0%<br>1: 32.4%<br>2: 11.7%<br>3: 7.2%<br>4+: 3.6%<br><br>Ethnicity:<br>Caucasian: 84.7%<br>East/South-East Asian: 9.9%<br>Other: 5.4%<br><br>LC risk factors: NR<br><br>Limited stage: 40.4%<br>Extensive stage: 59.6%<br><br>Small cell: 100% | | Participants completed a survey containing the EQ-5D-5L, which was used to generate Canadian-referenced health utility scores. HUS were generated according to Canadian reference values.19 HUS values were calculated for every available encounter with an enrolled patient, and thus, multiple HUS values could be derived from a patient on a given treatment.<br><br>Experienced (patients) | T2: On first-line therapy<br><br>**Extensive stage small-cell lung cancer:**<br>T1: Newly diagnosed and treatment naive<br>T2: On first-line therapy | |
| Kunikane 2019<br><br>Japan | The eligibility criteria included newly-diagnosed patients with stage IIIB or IV non-small cell lung cancer (NSCLC) or small cell lung cancer (SCLC) in | T1: 264;<br>T2: 212 | EQ-5D (Japanese tariff; TTO); range NR<br><br>No additional information provided on scale, levels, etc. | **Stage IIIB-IV non-small-cell or extensive small-cell lung cancer:** | None. |

| Study<br><br>Country (setting) | Inclusion criteria<br>Age (mean/median)<br>Sex (% female)<br>Smoking status<br>Comorbidities<br>Race/ethnicity<br>LC risk factors<br>Stages included<br>Type of LC included | Number of participants | Methods for preferences;<br>Experienced vs hypothetical health states | Relevant heath states valued (timepoint used) | Subgroups |
|---|---|---|---|---|---|
| | any stage, whose ages were over 20-years old, and who had provided written informed consent.<br><br>Age, median (range): 68 (35-89)<br><br>Female: 29.6%<br><br>Smoking status: NR<br><br>Comorbidities: NR<br><br>Race/ethnicity: NR<br><br>LC risk factors: NR<br><br>Stage IIIB:26.6%<br>Stage IV: 45.3%<br>Limited (SCLC): 10.9%<br>Extensive (SCLC): 17.2%<br><br>Non-small cell: 71.9%<br>Small-cell: 28.1% | | Experienced (patients) | T1: Newly diagnosed<br>T2: On first-line therapy | |
| Lal 2015<br><br>UK and Sweden (7 centers from the UK and 2 from Sweden) | Adult patients with advanced non-squamous NSCLC (stage IIIB or IV) that had not progressed after 4 cycles of platinum-based doublet treatment and who had an ECOG PS of 0/1 were eligible.<br><br>Age, median (range): 66 (27-82.5)<br><br>Female: 50%<br><br>Current or ex-smoker: 94%<br>Never smoked: 6%<br><br>Comorbidities: NR<br><br>Race/ethnicity: NR | 46 | EQ-5D-3L (UK; TTO); -0.59 to 1<br><br>The 5 individual dimensions (mobility, self-care, usual activities, pain/discomfort, and anxiety/depression; with 3 levels each: no, some, or major problems) were recorded and the health utility index was calculated using the UK population-based weights.<br><br>Experienced (patients) | **Stage IIIB or IV non-small cell lung cancer:**<br>T3: Prior to maintenance therapy after first-line therapy completion | None. |

| Study<br><br>Country (setting) | Inclusion criteria<br>Age (mean/median)<br>Sex (% female)<br>Smoking status<br>Comorbidities<br>Race/ethnicity<br>LC risk factors<br>Stages included<br>Type of LC included | Number of participants | Methods for preferences;<br>Experienced vs hypothetical health states | Relevant heath states valued (timepoint used) | Subgroups |
|---|---|---|---|---|---|
| | LC risk factors: NR<br><br>Stage IIIA: 2%<br>Stage IIIB: 6%<br>Stage IV: 92%<br><br>Non-small cell: 100% | | | | |
| Lim 2022<br><br>UK (NHS affiliated hospitals) | Aged ≥ 16 years, undergoing lobectomy or bi-lobectomy for treatment of known or suspected primary lung cancer beyond a lobar orifice, or undergoing frozen section biopsy with the intention to proceed with<br>lobectomy or bi-lobectomy if primary lung cancer beyond a lobar orifice is confirmed. (Note that for bi-lobectomy, the distance for the lobar orifice is in reference to the bronchus intermedius.) Cancer staging using TMN8, multidisciplinary team (MDT) consider the disease is suitable for both VATS lobectomy and lobectomy via open surgery, and ability to give written informed consent.<br><br>Age, mean (SD): 69 (8.8)<br><br>Female: 50.4%<br><br>Smoking history: NR<br><br>Comorbidities: NR<br><br>Race/ethnicity: NR<br><br>LC risk factors: NR<br><br>Stage IA: 8.2%<br>Stage IB: 32.5%<br>Stage IC: 26.7%<br>Stage IIA: 19.3%<br>Stage IIB: 5.8%<br>Stage III: 7.6% | 502 | EQ-5D-5L (UK; TTO); 0 to 1<br><br>The EQ-5D is a generic measure of health outcome, covering five dimensions:<br>(1) mobility, (2) self-care, (3) usual activities, (4) pain/discomfort and (5) anxiety/depression. The EQ-5D-5L<br>was completed by participants at six time points: (1) baseline (pre-randomization), (2) 2 weeks, (3) 5 weeks, (4) 3 months, (5) 6 months and (6) 1-year post randomization. Although data were gathered using the<br>EQ-5D-5L (the five-level version, with five possible responses for each dimension), responses recorded on the instrument were converted into a single index value using the original three-level UK valuation<br>set.<br><br>Experienced (patients) | **Stage I-IIIA lung cancer:**<br>T1: pre-treatment<br>T3: 2 weeks post-surgery | None. |

| Study<br><br>Country (setting) | Inclusion criteria<br>Age (mean/median)<br>Sex (% female)<br>Smoking status<br>Comorbidities<br>Race/ethnicity<br>LC risk factors<br>Stages included<br>Type of LC included | Number of participants | Methods for preferences;<br>Experienced vs hypothetical health states | Relevant heath states valued (timepoint used) | Subgroups |
|---|---|---|---|---|---|
| | Type of LC:<br>Non-small cell: 51.6%<br>Small cell: 0.2%<br>Histology not confirmed: 48.2% | | | | |
| Limwattananon 2018<br><br>Thailand (university hospital) | The target population was defined as patients with stage IIIB/IV, advanced NSCLC, who previously had neither chemotherapy nor TKIs. During January–June 2017, a total of 135 patients, aged 18 years or older who were hospitalized with NSCLC in a university hospital.<br><br>Age: NR<br><br>Female: NR<br><br>Smoking status: NR<br><br>Comorbidities: NR<br><br>Race/ethnicity: NR<br><br>LC risk factors: NR<br><br>Stage IIIB or IV: 100%<br><br>Non-small cell: 100% | T2: 82;<br>T3: 28 | EQ-5D (Thailand tariff; TTO); 0 to 1<br><br>Values on the five EQ5D health states were converted into 0–1 utility scores by an equation developed from the time trade-off method in a sample of the general Thai population.<br><br>Experienced (patients) | **Stage IIIB or IV non-small cell lung cancer:**<br>T2: On first-line therapy<br>T3: Stable disease after first-line therapy | None. |
| Matter-Walstra 2014<br><br>Germany (CESAR CEPAC-TDM Oncology trial) | Eligible patients were age 18–75 years with pathologically confirmed NSCLC stage IIIB or IV not amenable to curative treatment, ECOG ≤2, measurable tumor by RECIST v.1.1 [11], neutrophil count >1.5 G/l, platelet count >100 G/l, total bilirubin ≤2× upper limit of normal, AST and ALT ≤2.5× upper limit of normal (≤5× upper limit of normal in the case of liver metastases), creatinine clearance (according to the Cockcroft–Gault formula) ≥60 ml/min and no prior systemic treatment for advanced NSCLC. Patients with asymptomatic brain metastases were eligible in case steroids were not indicated. | 154 | EQ-5D (German tariff; TTO); range NR<br><br>Patients completed the EQ-5D according to the clinical visit schedule of each study. In addition, only forms of patients who had completed a minimum of two forms were included in the analyses. For each completed EQ-5D questionnaire, utilities were calculated using the German (EQGE utility), EU (EQEU utility), French (EQFR utility) and UK | **Stage IIIB or IV non-small-cell lung cancer:**<br>T2: On first-line therapy | None. |

| Study<br><br>Country (setting) | Inclusion criteria<br>Age (mean/median)<br>Sex (% female)<br>Smoking status<br>Comorbidities<br>Race/ethnicity<br>LC risk factors<br>Stages included<br>Type of LC included | Number of participants | Methods for preferences;<br>Experienced vs hypothetical health states | Relevant heath states valued (timepoint used) | Subgroups |
|---|---|---|---|---|---|
| | Age, mean (SD): 62.1 (7.9)<br><br>Female: NR by sample of interest<br><br>Smoking status: NR<br><br>Comorbidities: NR<br><br>Race/ethnicity: NR<br><br>LC risk factors: NR<br><br>Stage IIIB or IV: 100%<br><br>Non-small cell: 100% | | (EQUK utility) value sets and calculation algorithms as described elsewhere.<br><br>Experienced (patients) | | |
| Matza, 2024<br><br>United Kingdom | To be eligible for this study, participants were required to be over 18 years of age, a UK resident, able to understand the assessments as judged by the investigator, able and willing to give electronic informed consent, and able to complete the protocol requirements. Because the sample was intended to match the demographics of the UK general population, there were no eligibility criteria for specific clinical characteristics.<br><br>Age, mean (SD): 42.0 (13.8)<br><br>Female: 48.8%<br><br>Smoking status: NR<br><br>Comorbidities: NR<br><br>Ethnicity:<br>White: 79.8%<br>Black: 3.9%<br>Asian: 8.9%<br>Mixed: 5.4%<br>Other: 2.0% | 203 | TTO (choice-based in general population); 0 to 1<br>Trained interviewers conducted one-on-one in-person interviews in private offices, following a semi-structured interview guide. Participants were first introduced to the health states. Health state A was presented first followed by the groups of false-positive health states. The four groups of false-positive health states (i.e., the B, C, D, and E health states) were presented in random order. Each group was presented as a set. For example, when presenting the three lung cancer health states in group B, all three were presented on the table simultaneously, but the three were presented in random order. The health states were not presented with the organized lettering/numbering system. Instead, health states were labeled with different letters that would not provide any indication of the organizational structure or order of severity. For example, health states B1 and B2 were labeled | **Hypotheticals, describing screening health states:**<br>1) A false positive screening result (pooled results from:<br>a. False positive for lung cancer without head or neck involvement;<br>b. False positive for lung cancer with possible head or neck involvement;<br>c) False positive for lung cancer with a 6-month follow-up scan) | NR |

| Study<br><br>Country (setting) | Inclusion criteria<br>Age (mean/median)<br>Sex (% female)<br>Smoking status<br>Comorbidities<br>Race/ethnicity<br>LC risk factors<br>Stages included<br>Type of LC included | Number of participants | Methods for preferences;<br>Experienced vs hypothetical health states | Relevant heath states valued (timepoint used) | Subgroups |
|---|---|---|---|---|---|
| | LC risk factors: NR<br><br>Stage and type of LC: NA (hypothetical health states in general population) | | as T and Y. Interviewers reviewed every health state in detail with the participants, who were then given an opportunity to read the materials independently and ask questions. Participants were then asked to rank the health states from most preferable to least preferable. After completing the ranking, participants valued the health states in a TTO task with a 1-year time horizon. TTO methods have been described extensively in previous publications [30]. For each of the health states, participants were offered a series of choices between spending a 12-month period in the health state versus spending varying amounts of time in full health. Choices were presented in a booklet with bars of varying length illustrating duration of the two choices. Time in full health varied in 1-month increments, alternating between longer and shorter periods of time in full health (i.e., 12 months, 0 months [dead], 11 months, 1 month, 10 months, 2 months…). Each health state received a utility value (u) on a scale with anchors of dead (0) and full health (1) based on the choice in which the respondent was indifferent between 12 months in the health state and x months in full health. The resulting utility estimate (u) is calculated as u = x/12.<br><br>Hypotheticals (general public) | | |
| Mazzone 2013<br><br>USA (Cleveland Clinic Health System) | Ages 40-75 years, able to return for annual follow-up screening, willing to sign a medical release form, and at least one of the following criteria:<br>(a) a current or ex-smoker with at least a 10 pack years history;<br>(b) has first degree family member (parent, sibling, or child) with a history of lung cancer;<br>(c) has a clinical diagnosis of chronic obstructive pulmonary disease (COPD). | 1,424 (25 with a positive screening result) | EQ-5D-3L (US tariff; TTO); 0 to 1<br><br>The Euro-Qol – 5 dimensions (EQ-5D) is a standardized non-disease specific instrument for describing and valuing health related quality of life. 5 dimensions are included in the measure – mobility, self-care, usual activities, pain/discomfort, | **Screening health states (in those with positive screening results only):**<br>1) After a positive screening result (diagnosis not known) | None. |

| Study<br><br>Country (setting) | Inclusion criteria<br>Age (mean/median)<br>Sex (% female)<br>Smoking status<br>Comorbidities<br>Race/ethnicity<br>LC risk factors<br>Stages included<br>Type of LC included | Number of participants | Methods for preferences;<br>Experienced vs hypothetical health states | Relevant heath states valued (timepoint used) | Subgroups |
|---|---|---|---|---|---|
| | Age:<br>Screening group, mean (range): 59.9 (42.5-77.8)<br>Control group, mean (range): 60.2 (41.5-77.7)<br><br>Female: 54.2%<br><br>Active smokers: 51.1%<br>Never smokers: 1.7%<br>Former smokers: 47.3%<br>Number of pack-years:<br><20: 17.1%<br>20-40: 42.1%<br>40-60: 25.6%<br>80+: 4.4%<br><br>Comorbidities: NR<br><br>Race/ethnicity: NR<br><br>LC risk factors:<br>Family history of LC: 25.6%<br><br>Stage: NA<br><br>Type of LC: NA | | and anxiety/depression. For each dimension the subject can answer no problem, some or moderate problems, and extreme problems. The information obtained can be converted into a single index by applying one of several developed valuation measures. We used the US population preference algorithm for our study.<br><br>Experienced (patients) | **Comparator, screening health states:**<br>Before screening | |
| Nafees 2017<br><br>Australia, France, South Korea, United Kingdom (recruited from commercial patient panels) | Respondents were eligible if they were over 18 years of age, currently resident in the study country, able to understand the study procedures, and able to give informed consent.<br><br>Age: NR<br><br>Female: 56.0%<br><br>Smoking status: NR<br><br>Comorbidities: NR | T2: 300<br>T3: 75 (Korea only) | TTO (choice-based in general population); 0 to 1<br><br>All interviews were conducted by trained individuals in the target language. The TTO interview included two tasks; a visual analogue scale (VAS) assessment and the TTO interview. In the VAS exercise, respondents were asked to rate each health state on a 100 point scale whereby 0 represented “dead” and 100 represented full health. This task is used to | **Hypotheticals, stage IV non-small cell lung cancer:**<br>T2: On first-line therapy (excluding progression)<br>T3: On maintenance therapy after first-line therapy | |

| Study<br><br>Country (setting) | Inclusion criteria<br>Age (mean/median)<br>Sex (% female)<br>Smoking status<br>Comorbidities<br>Race/ethnicity<br>LC risk factors<br>Stages included<br>Type of LC included | Number of participants | Methods for preferences;<br>Experienced vs hypothetical health states | Relevant heath states valued (timepoint used) | Subgroups |
|---|---|---|---|---|---|
| | Ethnicity:<br>Caucasian: 41.0%<br>Black: 0.3%<br>South Asian (India, Pakistan): 3.3%<br>East Asian (China, Taiwan, South Korea): 27.3%<br>South East Asian (Indonesia, Malaysia): 1.0%<br>Mixed race: 0.7%<br>Other: 1.0%<br><br>LC risk factors: NR<br><br>Stage: NA (hypotheticals are for stage IV)<br><br>Type of LC: NA (hypotheticals are non-small cell) | | familiarise people with the descriptions and to provide additional ratings for each state. The TTO method derives utility values based upon subjects' responses and was developed specifically for use in health care.12 The TTO exercise is a standardized process for the valuation of health states.12 For each health state, respondents chose between (1) remaining in the health state without improvement for 10 years or (2) giving up× number of years of life in favor of "full health." The process incorporated a "ping-pong" approach until the participant was indifferent between the options presented. The data collected for each state were in the form of a utility value ranging from 0 [dead] to 1 [full health].<br><br>Hypotheticals (general public) | | |
| Naik 2017<br><br>Canada (outpatient clinics at the Princess Margaret Cancer Centre (PM), Toronto, Ontario) | The eligible population was comprised of ambulatory cancer survivors who were at least 18 years of age, were able to communicate in English, and had no significant cognitive impairment.<br><br>Age: NR by sample of interest<br><br>Female: NR by sample of interest<br><br>Smoking status: NR<br><br>Comorbidities: NR<br><br>Race/ethnicity: NR<br><br>LC risk factors: NR<br><br>Stage: | 149 | EQ-5D-3L (Canadian tariff; TTO); range NR<br><br>The EQ-5D-3L descriptive system (DS) consists of five questions, each representing a health dimension (mobility, self-care, usual activities, pain/discomfort, and anxiety/ depression). For each dimension, participants indicate one of three levels (no problems, moderate problems, or severe problems), allowing for 243 permutations of unique health states. From the health states reported by respondents on the EQ-5D, Canadian HU scores were calculated using the preference weight algorithm developed by Bansback et al.<br><br>Experienced (patients) | **Local/regional lung cancer:**<br>T3: Survivors after first-line therapy<br><br>**Distant/metastatic lung cancer:**<br>T3: Survivors after first-line therapy | None. |

| Study<br><br>Country (setting) | Inclusion criteria<br>Age (mean/median)<br>Sex (% female)<br>Smoking status<br>Comorbidities<br>Race/ethnicity<br>LC risk factors<br>Stages included<br>Type of LC included | Number of participants | Methods for preferences;<br>Experienced vs hypothetical health states | Relevant heath states valued (timepoint used) | Subgroups |
|---|---|---|---|---|---|
| | Local/regional: 59.7%<br>Distant/metastatic: 40.3%<br><br>Type of LC: NR | | | | |
| Ngo 2022<br><br>Australia (New South Wales, Sub-sample of the 45 and Up Study from Service Australia [eligible SEER questionnaire respondents]) | The study sample consisted of participants with a history of smoking<br>(current or former) aged 50–80 years with no prior diagnosis of lung<br>cancer.<br><br>Age, mean: 63.5<br><br>Female: 46.6%<br><br>Former smoker: 87.7%<br>Current smoker: 12.3%<br>Pack-year history:<br><20: 48.6%<br>20-29: 14.2%<br>30+: 27.4%<br>Missing: 9.8%<br><br>Comorbidities: NR<br><br>Ethnicity (authors determined by country of birth):<br>Asian: 1.4%<br>Pacific Islander: 0.2%<br>White: 98.4%<br><br>LC risk factors:<br>Family history of LC: 11.2%<br>Personal history of any cancer: 21.2%<br><br>Stage: NA (general population sample)<br><br>Type of LC: NA | 4,918 (varies by eligibility; range 3,036 to 4,918) | SF-6D (UK tariff; SG); range NR<br><br>The SF-6D utility instrument was derived from the SF-12 which was embedded in the SEEF questionnaire. To account for missing data, multiple imputation by chained equations (MICE) was performed to impute missing responses to the SF-12 and the SF-36 physical functioning scale using proportional odds logistic regression (with these variables used as predictors in the imputation model) in 30 imputed datasets. SF-6D utility values were obtained using the scoring algorithm created by Brazier et al. [26]. Selecting participants with up to two imputed dimensions in the SF-6D provided utilities for 98% of the sample. | **Comparator, screen-eligible general population (5 different, validated criteria):**<br>1) PLCOm2012 (≥1.51% risk score & aged 50–80 years with a history of smoking)<br>2) PLCOm2012 ≥2.00% risk score & aged 50–80 years with a history of smoking)<br>3) USPSTF 2013 (aged 55–80 years with a history of smoking [current or quit ≤ 15 years] and ≥ 30 pack-years of smoking,<br>where a pack was assumed to be 20 cigarettes/pipes/cigar s)<br>4) USPSTF 2021 (aged 50–80 years with a history of smoking [current or quit ≤ 15 years], and ≥ 20 pack-years of smoking) | None. |

| Study<br><br>Country (setting) | Inclusion criteria<br>Age (mean/median)<br>Sex (% female)<br>Smoking status<br>Comorbidities<br>Race/ethnicity<br>LC risk factors<br>Stages included<br>Type of LC included | Number of participants | Methods for preferences;<br>Experienced vs hypothetical health states | Relevant heath states valued (timepoint used) | Subgroups |
|---|---|---|---|---|---|
| | | | | 5) NELSON criteria (aged 50–74 years with a history of smoking [current or quit ≤ 10 years], and had smoked > 15 cigarettes<br>a day for > 25 years or > 10 cigarettes a day for > 30 years) | |
| Patel 2023<br><br>International sites in Canada, the United States, and France (St. Joseph's Healthcare Hamilton [Canada]; Toronto General Hospital (Canada); UF Health Shands Hospital [USA]; and CHU-Hôpitaux de Rouen [France]) | Inclusion criteria were age 18 years and above; clinical stage I, II, or IIIa NSCLC; and candidate for minimally invasive pulmonary lobectomy, as determined by the operating surgeon. Patients received neoadjuvant induction therapy when clinically indicated.<br><br>Age, median (IQR)(n=164): 68 (60-74)<br><br>Female: 67.1%<br><br>Smoker: 23.7%<br>Former smoker: 57.7%<br>Never smoker: 18.6%<br><br>1+ comorbidities: 89.0%<br><br>Race/ethnicity: NR<br><br>LC risk factor: NR<br><br>Stage I-IIIA: 92.7%<br><br>Non-small cell: 95.7% | T1: 157<br>T3: 155 | EQ-5D-5L (Canada; TTO); 0 to 1<br><br>The research team administered the EuroQol 5 Dimension 5 Level (EQ-5D-5L) questionnaire13 directly to the patients. The EQ-5D-5L was chosen because it is validated for a number of chronic conditions, including cardiovascular and respiratory disease, and it assesses mobility, self-care, usual activities, pain/ discomfort, and anxiety/depression. The data was collected and managed using REDCap. This score is typically interpreted along a continuum where 1 represents full health and 0 represents dead. A MID of 0.07 and SD of 0.16 is considered clinically significant for this patient population. HU scores were calculated using the Canadian EQ-5D-5L value set.<br><br>Experienced (patients) | **Stage I-IIIA non-small cell lung cancer:**<br>T1: Pre-treatment<br>T3: 3 weeks post-surgery | None. |
| Rauma 2015<br><br>Finland (Helsinki University | Between January 2000 and June 2009, patients undergoing operations for NSCLC at Helsinki University Central Hospital. Patients had to not speak in a foreign language and be alive as of June 2011 to be sent a questionnaire. | Patients: 230<br>General population (age-and gender- | 15D (NR); 0 to 1<br><br>The 15D is a standardized self-administered generic HRQoL instrument containing 15 dimensions with 5 levels each: moving, | **Stage I-IIIA non-small cell lung cancer:**<br>T3: Survivors after therapy | None. |

| Study<br><br>Country (setting) | Inclusion criteria<br>Age (mean/median)<br>Sex (% female)<br>Smoking status<br>Comorbidities<br>Race/ethnicity<br>LC risk factors<br>Stages included<br>Type of LC included | Number of participants | Methods for preferences;<br>Experienced vs hypothetical health states | Relevant heath states valued (timepoint used) | Subgroups |
|---|---|---|---|---|---|
| Central Hospital) | Patients only:<br>Age, mean (SD): 63.0 (8.7)<br><br>Female: 47.0%<br><br>Current smoker: 60.4%<br>Former smoker: 26.1%<br>Never-smoker: 13.5%<br>Mean (SD) smoke pack-years: 19.2%<br><br>Charlson Comorbidity Index:<br>0: 59.1%<br>1: 23.9%<br>2: 12.6%<br>3: 3.9%<br>4+: 0.4%<br><br>Race/ethnicity: NR<br><br>LC risk factors: NR<br><br>Stage IA: 39.6%<br>Stage IB: 21.3%<br>Stage IA: 2.2%<br>Stage IIB: 20.0%<br>Stage IIIA: 8.7%<br>Stage IIIB: 8.3%<br>Stage IV: 0.0%<br><br>Non-small cell: 100% | standardized): 4,191 | seeing, hearing, breathing, sleeping, eating, speech, excretion, usual activities, mental function, discomfort and symptoms, depression, distress, vitality, and sexual activity. The single index score (15D score) representing the overall HRQoL on a 0 to 1 scale (1 = full health, 0 = dead) and dimension level values reflecting the goodness of the levels relative to no problems on the dimension (= 1) and to being dead (= 0) are calculated from a health state-descriptive system by a set of population-based preference or utility weights.<br>A change of 0.02 to 0.03 in the 15D score is considered clinically important.<br>The 15D data for our general population came from the National Health 2000 Health Examination Surveys representing the Finnish population aged 30 and over.21 For this analysis, 4191 individuals were matched. This sample was weighted to reflect patients' age and gender distribution.<br><br>Experienced (patients) | **Comparator:** An age- and gender-standardized sample of the general population (National Health 2000 Health Examination Surveys representing the Finnish population aged 30 and over). | |
| Taghizadeh 2019<br><br>Canada (eight centres across Canada in the | Current or former smokers aged between 50 and 75 years and with a 2% or greater lung cancer risk over 6 years using a risk-prediction model developed using Prostate, Lung, Colorectal and Ovarian Cancer Screening Trial data.<br><br>Age, mean (SD): 62.9 (6.1) | 1,237 (279 with positive screening results; 238 and 168 at follow-up) | EQ-5D-3L (Canadian tariff; TTO); -0.34 to 1<br><br>The EQ-5D-3L consists of a preference-based index score; the index scores were derived from the current Canadian tariff15 (a maximum (best) value of 1 (for health state 11111) and a minimum value of −0.34 (for 33333)). | **Screening health states (in those with positive screening results only):**<br>1) After a positive screening result (diagnosis not known) | None. |

| Study<br><br>Country (setting) | Inclusion criteria<br>Age (mean/median)<br>Sex (% female)<br>Smoking status<br>Comorbidities<br>Race/ethnicity<br>LC risk factors<br>Stages included<br>Type of LC included | Number of participants | Methods for preferences;<br>Experienced vs hypothetical health states | Relevant heath states valued (timepoint used) | Subgroups |
|---|---|---|---|---|---|
| Pan-Canadian Early Detection of Lung Cancer Study) | Female: 54.9%<br><br>Current smokers: 62.1%<br>Former smokers: 37.9%<br>Pack-years, median (IQR; range): 51.3 (21.6; 2.2-230)<br><br>Comorbidities: NR<br><br>Race:<br>Caucasian: 97.1%<br>Black or African Canadian: 0.6%<br>Aboriginal: 0.3%<br>Pacific Islander: 0.0%<br>Other: 0.8%<br>Missing: 0.2%<br><br>LC risk factors:<br>Family history of LC: 31.7%<br>LC risk score, median (IQR; range): 3.5 (2.9; 2.0-33.5)<br><br>Stage: NR at last follow-up of 12% with LC diagnosis)<br><br>Type of LC: NR | | Experienced (patients) | 2) After a false positive result (diagnostic results known; 12% true positive and diagnosed with lung cancer)<br><br>**Comparator, screening health states:**<br>Before screening (and in those positive on screening only) | |
| Takamiya 2023<br><br>Japan (Showa University Northern Yokohama) | Patients who underwent lung tumor surgery at Showa University Northern Yokohama Hospital from 19 October 2020 to 24 September 2021.<br><br>Age, mean (SD): 68.0 (10.1)<br><br>Female: 35.3%<br><br>Pack-years smoking history (smoking index), mean (SD): 578.5 (636.4)<br><br>Charlson Comorbidity Index:<br>0: 60.8%<br>1: 15.7% | 51 | EQ-5D-5L (NR); 0 to 1<br><br>It consists of five dimensions: mobility, self-care, usual activities, pain/discomfort, and anxiety/depression. Each dimension is rated on a 5-point scale ranging from no problem (:1) to extreme problem (:5).<br><br>Experienced (patients) | **Stage I-IIIA lung cancer:**<br>T1: Pre-treatment<br>T3: At hospital discharge for surgery | NR |

| Study<br><br>Country (setting) | Inclusion criteria<br>Age (mean/median)<br>Sex (% female)<br>Smoking status<br>Comorbidities<br>Race/ethnicity<br>LC risk factors<br>Stages included<br>Type of LC included | Number of participants | Methods for preferences;<br>Experienced vs hypothetical health states | Relevant heath states valued (timepoint used) | Subgroups |
|---|---|---|---|---|---|
| | 2: 13.7%<br>≥3: 9.8%<br><br>Race/ethnicity: NR<br><br>LC risk factors: NR<br><br>Clinical stage:<br>Stage I: 80.4%<br>Stage II: 5.9%<br>Stage III: 7.8%<br>Stage IV: 2.0%<br><br>Type of LC: NR | | | | |
| Teramatsu 2022<br><br>Japan (University Hospital of Occupational and Environmental Health, Department of Occupational Medicine) | Employed patients (aged 20 years or older) who wished to resume work after discharge among the patients admitted to our hospital for lung resection surgery between August 2018 and October 2019.<br><br>Age, mean (SD): 63.1 (10.4)<br><br>Female: 37.3%<br><br>Smoking history: NR<br><br>Comorbidities:<br>0: 40.7%<br>1-2: 37.3%<br>3-4: 15.3%<br>≥5: 6.8%<br><br>Race/ethnicity: NR<br><br>LC risk factors: NR<br><br>Stage 0: 6.8%<br>Stage I: 71.2% | 59 | EQ-5D-3L (Japan; TTO); range NR<br><br>No further details on scale, etc.<br><br>Experienced (patients) | **Stage I-IIIA lung cancer:**<br>T3: 3 months post-surgery/treatment (10% with adjuvant therapy after surgery) | None. |

| Study<br><br>Country (setting) | Inclusion criteria<br>Age (mean/median)<br>Sex (% female)<br>Smoking status<br>Comorbidities<br>Race/ethnicity<br>LC risk factors<br>Stages included<br>Type of LC included | Number of participants | Methods for preferences;<br>Experienced vs hypothetical health states | Relevant heath states valued (timepoint used) | Subgroups |
|---|---|---|---|---|---|
| | Stage II: 6.8%<br>Stage III: 10.2%<br>Unknown: 5.1% | | | | |
| Tramontano 2015<br><br>USA (one of 5 large health maintenance organizations or 15 Veterans Administration Health Care System [VA] study sites) | Patients initially diagnosed with lung cancer in 2003-2005 in Northern California, Los Angeles County, North Carolina, Iowa, or Alabama, or who received care in one of 5 large health maintenance organizations or 15 Veterans Administration Health Care System (VA) study sites.<br><br>Age: NR<br><br>Female: NR by sample of interest<br><br>Smoking status: NR<br><br>Comorbidities: NR by sample of interest<br><br>Race/ethnicity: NR by sample of interest<br><br>Stage I: 46.6%<br>Stage II: 14.4%<br>Stage IV: 39.0%<br><br>Non-small cell or small-cell: 100% | 1,610 | EQ-5D-3L (USA tariff; TTO); -0.11 to 1<br><br>We calculated community-weight utilities for each of the 243 possible health states using the patient-provided scores for each of the 5 EQ-5D attributes and the algorithm provided by U.S. Agency for Healthcare Research and Quality (AHRQ).(10) Each EQ-5D-3L version attribute (mobility, self-care, usual activities, pain/ discomfort, anxiety/ depression) is scored as 1, 2, or 3 to indicate the level of severity on that attribute; for example, a score of '22222' would indicate some problems in each attribute. The AHRQ-provided algorithm was based on a random effects model (specifically, the 'D1 model') fit to time-tradeoff values provided in 2002 by a probability sample of U.S. adults (civilian, non-institutionalized).(8, 11) The range of utilities possible from this model is -0.11 to 1.00 (i.e., includes states worse than dead). The EQ-5D asks respondents about health "today".<br><br>Experienced (patients) | **Stage I-II lung cancer:**<br>T1: Treatment naïve at 4-6 months after diagnosis<br>T2: On first-line therapy<br>T3: >30 days after first-line therapy completion<br><br>**Stage IV lung cancer:**<br>T1: Treatment naïve at 4-6 months after diagnosis<br>T2: On first-line therapy<br>T3: >30 days after first-line therapy completion | None. |
| Tse 2020<br><br>Canada (Toronto, Ontario Princess Margaret Cancer Centre) | Eligible clinical outpatients had metastatic, histologically confirmed ALK+ NSCLC and were capable of providing informed consent. Patients were required to be fluent in 1 of the more than 24 languages of the HUS assessment tool EQ-5D-5L.<br><br>Age: NR for sample of interest<br><br>Female: NR for sample of interest<br><br>Smoking status: NR for sample of interest | NR explicitly (T1: 38; T2: 38) | EQ-5D-5L (Canada; TTO); range NR<br><br>The 5-question EQ-5D-5L survey that was used to generate health utility scores based on Canadian reference values.<br><br>Experienced (patients) | **Stage IV non-small cell lung cancer:**<br>T1: Treatment naïve/newly diagnosed<br>T2: On first-line therapy | None. |

| Study<br><br>Country (setting) | Inclusion criteria<br>Age (mean/median)<br>Sex (% female)<br>Smoking status<br>Comorbidities<br>Race/ethnicity<br>LC risk factors<br>Stages included<br>Type of LC included | Number of participants | Methods for preferences;<br>Experienced vs hypothetical health states | Relevant heath states valued (timepoint used) | Subgroups |
|---|---|---|---|---|---|
| | Comorbidities: NR<br><br>Race/ethnicity: NR for sample of interest<br><br>LC risk factors: NR<br><br>Stage IV: 100%<br><br>Non-small cell: 100% | | | | |
| Yalcin Balcik 2016<br><br>Turkey (Ankara, Hacettepe University Oncology Hospital of Ankara University, Ibni Sina Hospital and GATA Hospital) | Advanced stage NSCLC patients receiving gemcitabine/cisplatin and pemetrexed/cisplatin treatment at the Hacettepe University Oncology Hospital of Ankara University, Cebeci Hospital of Ankara University, Ibni Sina Hospital, and GATA Hospital.<br><br>Age: NR for sample of interest<br><br>Female: NR for sample of interest<br><br>Smoking status: NR<br><br>Comorbidities: NR<br><br>Race/ethnicity: NR<br><br>LC risk factors: NR<br><br>Advanced stage: 100%<br><br>Non-small cell: 100% | 30 | EQ-5D (Netherlands; NR); range NR | **Advanced stage non-small cell lung cancer:**<br>T2: On first-line therapy | None. |
| Yang 2023<br><br>International multicohort study | Adults with advanced epidermal growth factor receptor (EGFR)/anaplastic lymphoma kinase wild-type NSCLC with METex14 skipping, who were treatment-naïve or who had received up to 2 prior lines of therapy, were enrolled from September 2016.<br><br>Age: NR for sample of interest | 117 | EQ-5D-3L (US; TTO); -0.573 to 1<br><br><br>EQ-5D-5L responses were used to derive EQ-5D-5L utilities based on value sets for the United States using the “eq5d” package31 in | **Advanced non-small cell lung cancer:**<br>T1: Treatment naive | None. |

| Study<br><br>Country (setting) | Inclusion criteria<br>Age (mean/median)<br>Sex (% female)<br>Smoking status<br>Comorbidities<br>Race/ethnicity<br>LC risk factors<br>Stages included<br>Type of LC included | Number of participants | Methods for preferences;<br>Experienced vs hypothetical health states | Relevant heath states valued (timepoint used) | Subgroups |
|---|---|---|---|---|---|
| | Female: NR for sample of interest<br><br>Smoking status: NR for sample of interest<br><br>Comorbidities: NR<br><br>Race/ethnicity: NR for sample of interest<br><br>LC risk factors: NR<br><br>Advanced stage: 100%<br><br>Non-small cell: 100% | | the statistical software R (version 4.0.3; R Project for Statistical Computing).<br><br>Experienced (patients) | | |
| Yoh 2020<br><br>13 countries (110 investigative centers) | Previously untreated Stage IV NSCLC; EGFR exon 19-del or 21_L858R mutation; Eastern Cooperative Oncology Group (ECOG) performance status (PS) of 0 or 119; measurable disease as assessed by Response Evaluation Criteria in Solid Tumors (RECIST), version 1.120; and adequate hematological and organ function.<br><br>Age:<br>RamErL arm, median (IQR): 65 (57-71)<br>PboErL arm, median (IQR): 64 (56-70)<br><br>Female: 63.0%<br><br>Ever smoker: 30.5%<br>Never smoker: 60.8%<br>Unknown or missing: 8.7%<br><br>Comorbidities: NR<br><br>Race:<br>Asian: 77.1%<br>White: 22.3%<br>Other: 0.2% | T1: 449<br>T2: NR explicitly (90)<br>T3: 261 | EQ-5D-5L (UK; TTO); 0 to 1<br><br>The EQ-5D-5L was used to examine the impact of treatment on general health status, using five measures or “dimensions”: mobility, self-care, usual activities, pain or discomfort, and anxiety or depression. For each measure, patients recorded one of five levels of severity: having no problems, having slight problems, having moderate problems, having severe problems, and unable to do or having extreme problems. The Index Score, a summary score of the dimensions, was calculated from the United Kingdom valuation set of item weights to derive a score of 0 to 1, with 1 representing the best health status21. The Index Score was not computed for a patient if he/she had one or more missing values for the 5 items.<br><br>Experienced (patients) | **Stage IV non-small cell lung cancer:**<br>T1: Treatment naïve<br>T2: On first-line therapy (cycle 48 and 60)<br>T3: 30 days after first-line therapy | None. |

| Study<br><br>Country (setting) | Inclusion criteria<br>Age (mean/median)<br>Sex (% female)<br>Smoking status<br>Comorbidities<br>Race/ethnicity<br>LC risk factors<br>Stages included<br>Type of LC included | Number of participants | Methods for preferences;<br>Experienced vs hypothetical health states | Relevant heath states valued (timepoint used) | Subgroups |
|---|---|---|---|---|---|
| | LC risk factors: NR<br><br>Stage IV: 100%<br><br>Non-small cell: 100% | | | | |

Abbreviations: ECOG = Eastern Cooperative Oncology Group performance; IQR = interquartile range; NR = not reported; NSCLC = non-small-cell lung cancer; SD= standard deviation; SG = standard gamble; VATs = video-assisted thoracoscopic surgery; VHDI = very high development index; y = year(s).

Table S3.2. Risk of bias, utility studies

| Author, Year Tool (Health state if multiple) | 1.1 Selection of participants | Domain 1 | 2.1 Participation rate | 2.2 Responders similar to non-responders | 2.3 Attrition and missing data | Domain 2 | 3.1 Instrument valid and reliable | 3.2 Instrument administered in the intended way | 3.3 Valid representation of health state | 3.4 Understanding of instrument | Domain 3 | 4.1 Results analyzed appropriately | Domain 4 | Overall ROB |
|---|---|---|---|---|---|---|---|---|---|---|---|---|---|---|
| Bendixen 2019 EQ-5D (T1) | Y/PY | Low | Y/PY | NI | N/PN | High | Y/PY | Y/PY | Y/PY | Y/PY | Low | Y/PY | Low | High |
| Bendixen 2019 EQ-5D (T3) | Y/PY | Low | Y/PY | NI | N/PN | Some concerns | Y/PY | Y/PY | Y/PY | Y/PY | Low | Y/PY | Low | Some concerns |
| Bertolaccini 2023 EQ-5D | Y/PY | Low | Y/PY | Y/PY | Y/PY | Low | Y/PY | Y/PY | Y/PY | Y/PY | Low | Y/PY | Low | Low |

| | | | | | | | | | | | | | | |
|---|---|---|---|---|---|---|---|---|---|---|---|---|---|---|
| Black 2014 SF-6D | Y/PY | Low | NI | NI | NI | Some concerns | Y/PY | Y/PY | Y/PY | Y/PY | Low | N/PN | High | High |
| Chouaid 2013 EQ-5D | Y/PY | Low | NI | NI | NI | Some concerns | Y/PY | Y/PY | Y/PY | Y/PY | Low | Y/PY | Low | Some concerns |
| Erbaycu 2018 EQ-5D | NI | Some concerns | Y/PY | Y/PY | Y/PY | Low | Y/PY | Y/PY | Y/PY | Y/PY | Low | Y/PY | Low | Some concerns |
| Galetta 2015 EQ-5D (T1 & T2) | Y/PY | Low | NI | NI | Y/PY | Some concerns | Y/PY | Y/PY | Y/PY | Y/PY | Low | Y/PY | Low | Some concerns |
| Galetta 2015 EQ-5D (T3) | Y/PY | Low | NI | NI | N/PN | High | Y/PY | Y/PY | Y/PY | Y/PY | Low | Y/PY | Low | High |
| Ganti 2025 EQ-5D | Y/PY | Low | Y/PY | NI | NI | Some concerns | Y/PY | Y/PY | Y/PY | Y/PY | Low | N/PN | High | High |
| Iyer 2013 EQ-5D | Y/PY | Low | Y/PY | NI | Y/PY | Some concerns | Y/PY | Y/PY | Y/PY | Y/PY | Low | Y/PY | Low | Some concerns |
| Jeppesen 2018 EQ-5D | Y/PY | Low | Y/PY | Y/PY | Y/PY | Low | Y/PY | Y/PY | Y/PY | Y/PY | Low | Y/PY | Low | Low |
| Kim 2018 SG | Y/PY | Low | NI | NI | Y/PY | Some concerns | Y/PY | Y/PY | N/PN | N/PN | High | Y/PY | Low | High |
| Koide 2019 EQ-5D | Y/PY | Low | NI | NI | Y/PY | Some concerns | Y/PY | Y/PY | Y/PY | Y/PY | Low | Y/PY | Low | Some concerns |
| Kuehne 2022 EQ-5D | Y/PY | Low | NI | NI | Y/PY | Some concerns | Y/PY | Y/PY | Y/PY | Y/PY | Low | Y/PY | Low | Some concerns |
| Kunikane 2019 EQ-5D (T1) | NI | Some concerns | NI | NI | Y/PY | Some concerns | Y/PY | Y/PY | Y/PY | Y/PY | Low | Y/PY | Low | Some concerns |
| Kunikane 2019 EQ-5D (T2) | NI | Some concerns | NI | NI | N/PN | Some concerns | Y/PY | Y/PY | Y/PY | Y/PY | Low | Y/PY | Low | Some concerns |
| Lal 2015 EQ-5D | NI | Some concerns | Y/PY | Y/PY | Y/PY | Low | Y/PY | Y/PY | Y/PY | Y/PY | Low | N/PN | High | High |
| Lim 2022 EQ-5D | Y/PY | Low | Y/PY | NI | Y/PY | Some concerns | Y/PY | Y/PY | Y/PY | Y/PY | Low | Y/PY | Low | Some concerns |
| Limwattana non 2018 EQ-5D | NI | Some concerns | NI | NI | Y/PY | Some concerns | Y/PY | Y/PY | Y/PY | Y/PY | Low | Y/PY | Low | Some concerns |
| Matter-Walstra 2014 EQ-5D | N/PN | Some concerns | N/PN | NI | Y/PY | Some concerns | Y/PY | Y/PY | Y/PY | Y/PY | Low | Y/PY | Low | Some concerns |
| Matza 2024 TTO | Y/PY | Low | NI | NI | Y/PY | Some concerns | Y/PY | Y/PY | Y/PY | Y/PY | Low | Y/PY | Low | Some concerns |

| | | | | | | | | | | | | | | |
|---|---|---|---|---|---|---|---|---|---|---|---|---|---|---|
| Mazzone 2013 EQ-5D Comparator | Y/PY | Low | NI | NI | Y/PY | Some concerns | Y/PY | Y/PY | Y/PY | Y/PY | Low | N/PN | High | High |
| Mazzone 2013 EQ-5D CT +ve Within-study comparison | Y/PY | Low | NI | NI | Y/PY | Some concerns | Y/PY | Y/PY | Y/PY | Y/PY | Low | Y/PY | Low | Some concerns |
| Mazzone 2013 EQ-5D CT +ve Between-study comparison | Y/PY | Low | NI | NI | Y/PY | Some concerns | Y/PY | Y/PY | Y/PY | Y/PY | Low | N/PN | High | High |
| Nafees 2017 TTO | N/PN | Some concerns | NI | NI | Y/PY | Some concerns | Y/PY | Y/PY | Y/PY | Y/PY | Low | N/PN | High | High |
| Naik 2017 EQ-5D | N/PN | Some concerns | NI | NI | NI | Some concerns | Y/PY | Y/PY | Y/PY | Y/PY | Low | Y/PY | Low | Some concerns |
| Ngo 2022 EQ-5D | Y/PY | Low | Y/PY | Y/PY | Y/PY | Low | Y/PY | Y/PY | Y/PY | Y/PY | Low | Y/PY | Low | Low |
| Patel 2023 EQ-5D | Y/PY | Low | Y/PY | NI | Y/PY | Some concerns | Y/PY | Y/PY | Y/PY | Y/PY | Low | Y/PY | High | Some concerns |
| Rauma 2015 EQ-5D | Y/PY | Low | Y/PY | Y/PY | Y/PY | Low | Y/PY | Y/PY | Y/PY | Y/PY | Low | Y/PY | Low | Low |
| Taghizadeh 2019 EQ-5D Comparator | N/PN | Some concerns | NI | NI | Y/PY | Some concerns | Y/PY | Y/PY | Y/PY | Y/PY | Low | Y/PY | Low | Some concerns |
| Taghizadeh 2019 EQ-5D (CT +ve: 12-month follow-up) | N/PN | Some concerns | NI | NI | Y/PY | Some concerns | Y/PY | Y/PY | Y/PY | Y/PY | Low | Y/PY | Low | Some concerns |
| Taghizadeh 2019 EQ-5D (CT +ve: 1-month follow-up) | N/PN | Some concerns | NI | NI | N/PN | High | Y/PY | Y/PY | Y/PY | Y/PY | Low | Y/PY | Low | High |
| Takamiya 2023 EQ-5D | Y/PY | Low | Y/PY | NI | Y/PY | Low | Y/PY | Y/PY | Y/PY | Y/PY | Low | Y/PY | Low | Low |
| Teramatsu 2022 EQ-5D | Y/PY | Low | Y/PY | Y/PY | N/PN | Some concerns | Y/PY | Y/PY | Y/PY | Y/PY | Low | Y/PY | Low | Some concerns |
| Tramontano 2015 | Y/PY | Low | N/PN | N/PN | Y/PY | Some concerns | Y/PY | Y/PY | Y/PY | Y/PY | Low | Y/PY | Low | Some concerns |

| | | | | | | | | | | | | | | |
|---|---|---|---|---|---|---|---|---|---|---|---|---|---|---|
| EQ-5D | | | | | | | | | | | | | | |
| Tse 2020 EQ-5D | Y/PY | Low | NI | NI | NI | Some concerns | Y/PY | Y/PY | Y/PY | Y/PY | Low | Y/PY | Low | Some concerns |
| Yalcin Balcik 2016 EQ-5D | NI | Some concerns | NI | NI | Y/PY | Some concerns | Y/PY | Y/PY | Y/PY | Y/PY | Low | N/PN | High | High |
| Yang 2023 EQ-5D | Y/PY | Low | NI | NI | Y/PY | Some concerns | Y/PY | Y/PY | Y/PY | Y/PY | Low | Y/PY | Low | Some concerns |
| Yoh 2020 EQ-5D (T1) | Y/PY | Low | Y/PY | Y/PY | Y/PY | Low | Y/PY | Y/PY | Y/PY | Y/PY | Low | Y/PY | Low | Some concerns |
| Yoh 2020 EQ-5D (T2 & T3) | Y/PY | Low | Y/PY | Y/PY | N/PN | High | Y/PY | Y/PY | Y/PY | Y/PY | Low | Y/PY | Low | High |

Abbreviations: CT +ve = patients who receiving a positive result on CT scan; N/PN = no/partial no; NI = no information; Y/PY = yes/partial yes; T1=timepoint 1 (at diagnosis/pre-treatment); T2=timepoint 2 (on first-line treatment); T3=timepoint 3 (after first-line treatment completion/on maintenance therapy/survivors).

Notes on ROB assessments:

- Overall ROB was rated as: low if all domains were rated as low; some concerns if one or more domains was rated as some concerns and none as high; high if one or more domains was rated as high.
- Domain 2 was rated as: low if questions 2.1 to 2.3 were rated as Y/PY; moderate if any or all of the three questions in 2.1 to 2.3 were rated as NI; and serious if either 2.1 and 2.3 were rated as N/PN, or if 2.3 was rated as N/PN and <70% enrolled had complete data. Question 2.1 was rated as Y/PY if ≥50% of those eligible were enrolled. Question 2.3 was rated as Y/PY if ≥80% enrolled had compete data.
- Domain 3 was rated as: low if questions 3.1 to 3.4 were rated as Y/PY; moderate if any or all of the four questions were NI and/or if only one was N/PN; and serious of any two or more questions were rated as N/PN.

Table S3.3. Summary of Findings: Health state utility studies

| Number of included studies; Sample size | Findings | GRADE* | What does the evidence say? |
|---|---|---|---|
| **Unaffected eligible population comparator (FOR USE TO CALCULATE DISUTILITIES OF OTHER HEALTH STATES)** | | | |
| 5 studies (1-5);<br>N=23,466<br>(EQ-5D: 2 studies, N=1,237; SF-6D: 2 studies, N=4,918; 15D: 1 study, n=4,191)<br><br>Alberta norms; n=19,859<br>*Al Sayah et al. Qual Life Res. 2025. 34:219-30.* | **Pooled utilities (unweighted average):** 0.81 (range: 0.74 to 0.91)<br><br>**Sensitivity analysis, excluding high ROB studies (unweighted average, N=3 studies; N=10,346):** 0.82 (range: 0.77 to 0.86)<br><br>Included studies for comparator utility measure:<br>EQ-5D: 2 studies: Taghizadeh 2019 (n=1,237): participants before undergoing screening; Mazzone 2013 (n=1,424, **high ROB**): participants before screening.<br><br>SF-6D: 2 studies: Ngo 2022 (n=4,918): average across four eligibility criteria for screening but not actively undergoing screening; Black 2014 (n=11,696, **high ROB**): subsample of NLST trial participants before undergoing screening.<br><br>15D: Rauma 2015 (n=4,191): general population sample, matched to lung cancer patients on age and gender.<br><br>**Alberta EQ-5D utility norms (45 to 75+ year), mean (SD):**<br>Average 45-75 years: 0.828<br>45-54 years: 0.837 (0.152); 55-64 years: 0.827 (0.153); 65-74 years: 0.820 (0.148) | Low[a]<br>⊕⊕⊖⊖<br>Inconsistency, and indirectness | The utility value for an unaffected eligible population comparator may be 0.81. |
| **Disutility of positive screening result (results not known; among those without cancer)** | | | |

| Number of included studies;<br>Sample size | Findings | GRADE* | What does the evidence say? |
|---|---|---|---|
| Within study:<br>EQ-5D: 2 studies (2, 5); N=263<br><br>Between study:<br>3 studies (2, 5, 6); N=778 (EQ-5D: 2 studies, N=263; SG: 1 study, n=515) | **Within study pooled disutilities (95% CI) (Figure S3.2):** 0.01 (-0.02, 0.03)<br>*neither high ROB<br><br>**Between study pooled utilities (95% CI); $I^2$ (Figure S3.3):** 0.84 (0.81, 0.86); 10%<br><br>**Disutility (95% CI) from pooled comparator:** 0.81 – 0.84 (0.81, 0.86) = -0.03 (-0.05, 0.00)<br><br>**Sensitivity analysis, excluding high ROB studies, disutility (95% CI) (N=1 study; N=24):** 0.81 – 0.88 (0.82, 0.94) = -0.07 (-0.01, -0.13)<br><br>**Sensitivity analysis, excluding SG utility study, disutility (95% CI) (N=2 studies; N=263):** 0.81 – 0.86 (0.82, 0.90) = -0.05 (-0.09, -0.01)<br><br>**Within study subgroup: Standard gamble (N=1 study; N=515 participants):**<br>Age at evaluation of hypotheticals, utilities:<br>19-29y: 0.83<br>30-39y: 0.83<br>40-49y: 0.82<br>50-59y: 0.82<br>≥60y: 0.85<br><br>Gender, utilities:<br>Men: 0.82<br>Women: 0.83 | Moderate[b]<br>⊕⊕⊕⊖<br>Imprecision | There is probably little-to-no disutility (0.01) of receiving a positive screening result. |
| **Disutility of false positive result (results known)** | | | |
| Within study:<br>EQ-5D: 1 study (5); N=246<br><br>Between study:<br>2 studies (5, 7); N=371 (EQ-5D: 1 study, N=168; TTO: 1 study; N=203) | **Within study disutilities (95% CI), EQ-5D:**<br>1 month after diagnostic work-up (n=168): 0.00 (-0.03, 0.04)<br>*High ROB at this timepoint; includes those with a cancer diagnosis (12% of sample)<br>12 months after screening (NR timeframe after diagnostic work-up) (n=246): -0.01 (-0.05, 0.02)<br>*Includes those with a cancer diagnosis (12% of sample)<br><br>**Between study pooled utilities (95% CI); $I^2$ (Figure S3.4):** 0.86 (0.84, 0.88); 70%<br>Up to 1 month after diagnostic work-up<br><br>**Disutility (95% CI) from pooled comparator:** 0.81 - 0.86 (0.84, 0.88) = -0.05 (-0.07, -0.03)<br><br>**Sensitivity analysis, excluding high ROB study, disutility (95% CI) (1 study; N=203):** 0.81 – 0.87 (0.85, 0.89) = -0.06 (-0.08, -0.04) | Low[c]<br>⊕⊕⊖⊖<br>Indirectness, imprecision | There may be little-to-no disutility (0.00) of a false positive result. |

| Number of included studies;<br>Sample size | Findings | GRADE* | What does the evidence say? |
|---|---|---|---|
| | **Sensitivity analysis, excluding time trade off utility study, disutility (95% CI) (N=1 study; N=168):** 0.81 – 0.85 (0.83, 0.87) = -0.04 (-0.06, -0.02) | | |
| **Disutility of lung cancer at diagnosis/treatment naive, stage I-IIIA** | | | |
| Between study:<br>9 studies (6, 8-15); N=1,572<br>(EQ-5D: 8 studies, N=1,057; SG: 1 study, N=515) | **Within study disutilities (95% CI):** no evidence.<br><br>**Between study pooled utilities (95% CI) EQ-5D; $I^2$ (Figure S3.5):** 0.82 (0.77, 0.87); 94%<br>*Timing from diagnosis not reported by most studies.<br><br>**Disutility (95% CI) from pooled comparator:** 0.81 – 0.82 (0.77, 0.87) = -0.01 (-0.06, 0.04)<br><br>**Sensitivity analysis, excluding high ROB studies, disutility (95% CI) (N=6 studies; N=856):** 0.81 – 0.81 (0.76, 0.86) = 0.00 (-0.05, 0.05)<br><br>**Between study subgroup, by stage:**<br>Stage I (N=2 studies; N=252): 0.81 (0.68, 0.95); $I^2$=95%<br>Stage I-II (N=1 study; N=52): 0.79 (0.74, 0.84); $I^2$=NA<br>Stage I-IIIA (N=5 studies; N=734): 0.83 (0.77, 0.89); $I^2$=95%<br>P-value for subgroup differences: p=0.63<br><br>**Standard gamble utility (95% CI):** 0.56 (0.54, 0.58)<br>**Disutility (95% CI) from pooled comparator:** 0.81 - 0.56 (0.54, 0.58) = 0.25 (0.23, 0.27)<br>*high ROB<br><br>**Within study subgroup: Standard gamble (N=1 study; N=515 participants):**<br>Age at evaluation of hypotheticals, utilities:<br>19-29y: 0.57<br>30-39y: 0.54<br>40-49y: 0.56<br>50-59y: 0.55<br>≥60y: 0.56<br><br>Gender, utilities:<br>Men: 0.55<br>Women: 0.56<br><br>No study identified that the lung cancer was screen-detected. | Low[d]<br>⊕⊕⊖⊖<br>Indirectness, and imprecision | There may be little-to-no disutility (-0.01) for stage I-IIIA lung cancer at diagnosis/treatment naïve. |
| **Disutility of lung cancer at diagnosis/treatment naive, stage IIIB-IV** | | | |
| Between study:<br>10 studies (6, 11, 15-22); N=2,243 | **Within study disutilities (95% CI):** no evidence.<br><br>**Between study pooled utilities (95% CI) EQ-5D; $I^2$ (Figure S3.6):** 0.74 (0.67, 0.80); 96% | Low[e]<br>⊕⊕⊖⊖ | There may be a small, but important disutility (0.07) for stage IIIB-IV |

| Number of included studies; Sample size | Findings | GRADE* | What does the evidence say? |
|---|---|---|---|
| (EQ-5D: 9 studies, N=1,728;<br><br>SG: 1 study, n=515) | *Timing from diagnosis not reported by most studies.<br><br>**Disutility (95% CI) from pooled comparator:** 0.81 – 0.74 (0.67, 0.80) = 0.07 (0.01, 0.14)<br><br>**Sensitivity analysis, excluding high ROB studies, disutility (95% CI) (N=8 studies; N=1,283):** 0.81 **–** 0.74 (0.67, 0.81) = 0.07 (0.00, 0.14)<br><br>**Between study subgroup, by stage:**<br>Stage IIIB-IV (N=4 studies; N=662): 0.71 (0.65, 0.77); $I^2$ =82%<br>Stage IV (N=5 studies; N=1,066): 0.76 (0.66, 0.85); $I^2$ =97%<br>P-value for subgroup differences: p=0.44<br><br>**Within study subgroups: EQ-5D (N=1 study; N=266 participants):**<br>Age, utilities:<br><40y: 0.82<br>40-49y: 0.74<br>50-59y: 0.71<br>60-69y: 0.66<br>≥70y: 0.54<br><br>Gender, utilities (95% CI):<br>Men: 0.66 (0.62, 0.71)<br>Women: 0.71 (0.55, 0.87)<br><br>Smoking status, utilities (95% CI):<br>Never smoked: 0.66 (0.45, 0.87)<br>Smoker: 0.44 (0.27, 0.61)<br>“Gave up” smoking: 0.65 (0.60, 0.70)<br><br>**Standard gamble utility (95% CI):** 0.35 (0.33, 0.37)<br>**Disutility (95% CI) from pooled comparator:** 0.81 - 0.35 (0.33, 0.37) = 0.46 (0.44, 0.48)<br>*high ROB<br><br>**Within study subgroups: Standard gamble (N=1 study; N=515 participants):**<br>Age at evaluation of hypotheticals, utilities:<br>19-29y: 0.34<br>30-39y: 0.34<br>40-49y: 0.33<br>50-59y: 0.36<br>>=60y: 0.36<br><br>Gender, utilities: | Indirectness, and imprecision | lung cancer at diagnosis/treatment naïve. |

| Number of included studies; Sample size | Findings | GRADE* | What does the evidence say? |
|---|---|---|---|
| | Men: 0.35<br>Women: 0.34<br><br>No study identified that the lung cancer was screen-detected. | | |
| **Disutility of lung cancer, on first-line adjuvant treatment (none only receiving surgery), stage I-IIIA** | | | |
| Between study:<br>EQ-5D: 3 studies (11, 15, 23); N=609 | **Within study disutilities (95% CI):** no evidence.<br><br>**Between study pooled utilities (95% CI) EQ-5D; $I^2$ (Figure S3.7):** 0.76 (0.73, 0.79); $I^2$ =75%<br>*One study 4-6 months from diagnosis, one "newly diagnosed" at enrollment, one NR.<br><br>**Disutility (95% CI) from pooled comparator:** 0.81 – 0.76 (0.73, 0.79) = 0.05 (0.02, 0.08)<br><br>**Sensitivity analysis, excluding high ROB studies, disutility (95% CI) (N=2 studies; N=400):** 0.81 – 0.77 (0.73, 0.80) = 0.04 (0.01, 0.08)<br><br>**Between study subgroup, by stage:**<br>Stage I-II (N=1 study; N=373): 0.78 (0.76, 0.80); $I^2$ =NA<br>Stage I-IIIA (N=2 studies; N=236): 0.76 (0.73, 0.79); $I^2$ =0%<br>P-value for subgroup differences: p=0.004 | Low[f]<br>⊕⊕⊖⊖<br>Indirectness, and imprecision | There may be a small, but important disutility (0.05) for stage I-IIIA lung cancer during first-line treatment. |
| **Disutility of lung cancer, on first-line adjuvant treatment, stage IIIB-IV** | | | |
| Between study:<br>13 studies (11, 15, 17-20, 22, 24-29); N=2,635 (EQ-5D: 12 studies, N=2,335; TTO: 1 study, n=300) | **Within study disutilities (95% CI):** no evidence.<br><br>**Between study pooled utilities (95% CI); $I^2$ (Figure S3.8):** 0.72 (0.67, 0.76); 96%<br>*Timing from diagnosis not reported by most studies.<br><br>**Disutility (95% CI) from pooled comparator:** 0.81 – 0.72 (0.67, 0.76) = 0.09 (0.05, 0.14)<br><br>**Sensitivity analysis, excluding high ROB studies, disutility (95% CI) (N=9 studies; N=2,070):** 0.81 - 0.72 (0.67, 0.77) = 0.09 (0.04, 0.14)<br><br>**Sensitivity analysis, excluding time trade off utility studies, disutility (95% CI) (N=12 studies; N=2,335):** 0.81 - 0.73 (0.69, 0.77) = 0.08 (0.04, 0.12)<br><br>**Between study subgroup, by stage:**<br>Stage IIIB-IV (N=7 studies; N=1,091): 0.71 (0.63, 0.79); $I^2$ =97%<br>Stage IV (N=6 studies; N=1,544): 0.72 (0.66, 0.79); $I^2$ =97%<br>P-value for subgroup differences: p=0.81 | Moderate[g]<br>⊕⊕⊕⊖<br>Indirectness<br><br>Low[g]<br>⊕⊕⊖⊖<br>Inconsistency and indirectness | There is probably a small, but important disutility (at least 0.04) for stage IIIB-IV lung cancer on first-line treatment.<br><br>There may be a moderate disutility (0.09) for stage IIIB-IV lung cancer on first-line treatment. |
| **Disutility of lung cancer, after first-line treatment completion and non-progressive (post-surgery, on maintenance therapy, or survivors), stage I-IIIA** | | | |

| Number of included studies;<br>Sample size | Findings | GRADE* | What does the evidence say? |
|---|---|---|---|
| Within study:<br>15D: 1 study (4); n=230<br><br>Between study:<br>11 studies (4, 8-10, 12-15, 30-32);<br>N=13,848 (EQ-5D: 10 studies, N=13,618;<br>15D: 1 study, n=230) | **Within study disutilities (95% CI):**<br>15D (n=230): 0.03 (0.01, 0.04)<br><br>**Between study pooled utilities (95% CI); $I^2$ (Figure S3.9):** 0.78 (0.73, 0.83); $I^2$ =99%<br>*Timing from diagnosis not reported by most studies.<br><br>**Disutility (95% CI) from pooled comparator:** 0.81 – 0.78 (0.73, 0.83) = 0.03 (-0.02, 0.08)<br>*No studies at high ROB<br>**Sensitivity analysis, excluding 15D utility studies, disutility (95% CI) (N=10 studies; N=13,618):** 0.81 - 0.77 (0.71, 0.83) = 0.04 (-0.02, 0.10)<br><br>**Between study subgroup, by treatment type:**<br>After surgery (N=7 studies; N=12,764): 0.78 (0.69, 0.86); $I^2$ =99%<br>After both surgery and adjuvant therapy (N=4 studies; N=1,084): 0.79 (0.75, 0.82); $I^2$ =90%<br>P-value for subgroup differences: p=0.85<br><br>**Between study subgroup, by stage:**<br>Stage I (N=2 studies; N=242): 0.73 (0.64, 0.82); $I^2$ =87%<br>Stage I-II (N=4 studies; N=13,022): 0.79 (0.70, 0.88); $I^2$ =99%<br>Stage I-IIIA (N=5 studies; N=13,228): 0.78 (0.74, 0.83); $I^2$ =90%<br>P-value for subgroup differences: p=0.53 | Low[h]<br>⊕⊕⊖⊖<br>Inconsistency and indirectness | There may be little-to-no disutility (0.03) for stage I-IIIA lung cancer after first-line treatment completion without progression. |
| **Disutility of lung cancer, after first-line treatment completion and non-progressive (on maintenance therapy, or survivors), stage IIIB-IV** | | | |
| Between study:<br>8 studies (15, 17, 18, 22, 26, 28, 31, 33); N=1,158<br>(EQ-5D: 7 studies, N=1,083;<br>TTO: 1 study, n=75) | **Within study disutilities (95% CI):** no evidence.<br><br>**Between study pooled utilities (95% CI); $I^2$ (Figure S3.10):** 0.75 (0.71, 0.78); 78%<br>*Timing from diagnosis not reported by most studies.<br><br>**Disutility (95% CI) from pooled comparator:** 0.81 – 0.75 (0.71, 0.78) = 0.06 (0.03, 0.10)<br><br>**Sensitivity analysis, excluding high ROB studies, disutility (95% CI) (N=3 studies; N=327):** 0.81 – 0.74 (0.69, 0.79) = 0.07 (0.02, 0.12)<br><br>**Sensitivity analysis, excluding time trade off utility studies, disutility (95% CI) (N=7 studies; N=1,083):** 0.81 - 0.74 (0.71, 0.78) = 0.07 (0.03, 0.10)<br><br>**Subgroup, by stage:**<br>Stage IIIB-IV (N=1 study; N=28): 0.68 (0.62, 0.74); $I^2$ =NA<br>Stage IV (N=7 studies; N=1,130): 0.76 (0.73, 0.79); $I^2$ =75%<br>P-value for subgroup differences: p=0.02 | Low[i]<br>⊕⊕⊖⊖<br>Indirectness and imprecision | There may be a small, but important disutility (0.06) for stage IIIB-IV lung cancer after first-line treatment completion without progression. |

***GRADE ratings:**

[a]**Unaffected eligible population comparator (FOR USE TO CALCULATE DISUTILITIES OF OTHER HEALTH STATES):**

***No serious concerns about risk of bias:*** The pooled utility (unweighted average) did not differ when excluding high risk of bias studies. ***Serious concerns about inconsistency:*** There was serious inconsistency in the measured utilities across different tools. ***Serious concerns about indirectness:*** The population is indirect as they are primarily those attending lung cancer screening. ***No serious concerns about imprecision:*** The range of utility measures across studies was attributed to inconsistency. ***No serious concerns about reporting bias:*** No serious concerns about reporting bias.

[b]**Disutility of positive screening result (results not known, among those without cancer):**

***No serious concerns about risk of bias:*** The within-study disutilities were not at high risk of bias. Two of three studies in the between-study analysis were at high risk of bias, but we relied on within-study findings as only one small study (n=24) not at high risk for between-study analysis had findings that disagreed with within-study results. ***No serious concerns about inconsistency:*** The measured utilities were consistent. ***No serious concerns of indirectness:*** Within-study analysis agrees with between study data. Some participants (12%) in one study had cancer but removing these would be unlikely to influence a finding of little-to-no disutility. ***Serious concerns of imprecision:*** The sample for the within study comparison is small (n=263), and the 95% CI around the between-study calculation is imprecise, showing the possibility of a small disutility or utility. ***No serious concerns about reporting bias:*** No serious concerns about reporting bias.

[c]**Disutility of false positive result (results known):**

***No serious concerns about risk of bias:*** A single study contributed to the within study analysis, which at the one-month timepoint was at high risk of bias, but had similar results to the 12-month follow-up timepoint. The additional study that contributed to the between study analysis was not at high risk of bias. ***No serious concerns about inconsistency:*** The study contributing to the within study analysis did not find a disutility, and neither did the two studies in the between-study analysis. ***Serious concerns of indirectness:*** 12% of the sample in the within-study analysis had lung cancer to inform the screening health state of false positives. ***Serious concerns of imprecision:*** The study used in the within study comparison was small (n=168 and n=246 at each timepoint), and the upper limit of the 95% CI at the one-month timepoint suggests a possibly small, but important disutility, whereas the lower limit of the 95% CI at 12-month timepoint indicates a possibility a small, but important utility. ***No serious concerns about reporting bias:*** No serious concerns about reporting bias.

[d]**Disutility of lung cancer, at diagnosis/treatment naive, stage I-IIIA:**

***No serious concerns about risk of bias:*** The between study disutilities did not differ when excluding high risk of bias studies. ***No serious concerns of inconsistency:*** The measured utilities were consistent. ***Serious concerns about indirectness:*** No within-study comparator for disutility calculation. ***Serious concerns about imprecision:*** The upper and lower bound of the confidence intervals for the EQ-5D disutility indicate a small utility (-0.06) and disutility (0.04). ***No serious concerns about reporting bias:*** No serious concerns about reporting bias.

[e]**Disutility of lung cancer, at diagnosis/treatment naive, stage IIIB-IV:**

***No serious concerns about risk of bias:*** The between study disutilities did not differ when excluding high risk of bias studies. ***No serious concerns of inconsistency:*** The measured utilities were consistent. ***Serious concerns about indirectness:*** No within-study comparator for disutility calculation. ***Serious concerns about imprecision:*** The lower limit of the 95% CI indicates little-to-no disutility. ***No serious concerns about reporting bias:*** No serious concerns about reporting bias.

[f]**Disutility of lung cancer, on first-line adjuvant treatment (none only receiving surgery), stage I-IIIA:**

***No serious concerns about risk of bias:*** the between study disutilities did not substantially differ when excluding the high risk of bias study. ***No serious concerns of inconsistency:*** The measured utilities were consistent. Some evidence suggested lower stages (I-II) may not reach a small disutility, but this came from between study subgroups and a single study. It is unclear what percent of lung cancers were stage IIIA from two studies suggesting a small disutility. ***Serious concerns about indirectness:*** No within-study comparator for disutility calculation. ***Serious concerns about imprecision:*** The lower limit of the 95% CI indicates little-to-no disutility. ***No serious concerns about reporting bias:*** No serious concerns about reporting bias.

[g]**Disutility of lung cancer, on first-line treatment, stage IIIB-IV:**

Small, but important disutility:

***No serious concerns about risk of bias:*** The between study disutilities did not differ when excluding high risk of bias studies. ***No serious concerns of inconsistency:*** The measured utilities were relatively consistent for a small, but important disutility. Three of 13 studies had utilities of 0.56, 0.58 and 0.63, but contributed 23.4% of the total weight in the analysis. ***Serious concerns about indirectness:*** No within-study comparator for disutility calculation. ***No serious concerns about imprecision:*** The 96% CI is precise for a small, but important disutility. ***No serious concerns about reporting bias:*** No serious concerns about reporting bias.

Moderate disutility:

***No serious concerns about risk of bias:*** The between study disutilities did not differ when excluding high risk of bias studies. ***Serious concerns about inconsistency:*** There is serious inconsistency for a moderate disutility, which was not explained by treatment type, stage or measurement tool. ***Serious concerns about indirectness:*** No within-study comparator for disutility calculation. ***No serious concerns about imprecision:*** The 95% CI does not have any serious concerns of imprecision (lower limit of 0.05 indicating a small, but important difference is attributed to inconsistency). ***No serious concerns about reporting bias:*** No serious concerns about reporting bias.

##### [h]Disutility of lung cancer, after first-line treatment completion and non-progressive (post-surgery, on maintenance therapy, or survivors), stage I-IIIA

***No serious concerns about risk of bias:*** The within and between study disutilities were not at high risk of bias. ***Serious concerns about inconsistency:*** There was serious inconsistency in the measured utilities (6 of 11 point estimates indicated a small disutility), which was not explained by treatment type, stage, or measurement tool. ***Serious concerns about indirectness:*** Only one of 11 studies had a direct comparison using the 15D measurement tool, 10 studies required an indirect comparison. ***No serious concerns about imprecision:*** The 95% CI does not have any serious imprecision around little-to-no disutility (upper limit 0.08 indicating a small, but important disutility was attributed to inconsistency). ***No serious concerns about reporting bias:*** No serious concerns about reporting bias.

##### [i]Disutility of lung cancer, after first-line treatment completion and non-progressive (on maintenance therapy, or survivors) stage IIIB-IV:

***No serious concerns about risk of bias:*** The between study disutilities did not differ when excluding high risk of bias studies. ***No serious concerns about inconsistency:*** The measured utilities were consistent. ***Serious concerns about indirectness:*** No within study comparator for disutility calculation, as well as the populations were largely stage IV across all studies. ***Serious concerns about imprecision:*** The lower limit of the 95% CI indicates little-to-no disutility. ***No serious concerns about reporting bias:*** No serious concerns about reporting bias.

| Study or Subgroup | Disutility | SE | Weight | Disutility IV, Random, 95% CI |
|---|---|---|---|---|
| Mazzone 2013 | 0.03 | 0.0306 | 20.6% | 0.03 [-0.03, 0.09] |
| Taghizadeh 2019 | 0 | 0.0156 | 79.4% | 0.00 [-0.03, 0.03] |
| **Total (95% CI)** | | | **100.0%** | **0.01 [-0.02, 0.03]** |

Heterogeneity: Tau² = 0.00; Chi² = 0.76, df = 1 (P = 0.38); I² = 0%
Test for overall effect: Z = 0.45 (P = 0.66)

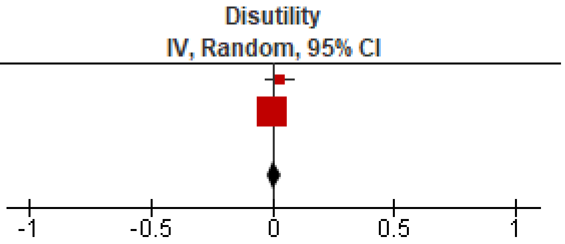

Figure S3.2 Within study disutility of positive screening result (results not known; among those without cancer)

| Study or Subgroup | Utility | SE | Weight | Utility IV, Random, 95% CI |
|---|---|---|---|---|
| Kim 2018 | 0.83 | 0.0106 | 75.4% | 0.83 [0.81, 0.85] |
| Mazzone 2013 | 0.88 | 0.032 | 12.3% | 0.88 [0.82, 0.94] |
| Taghizadeh 2019 | 0.84 | 0.032 | 12.3% | 0.84 [0.78, 0.90] |
| **Total (95% CI)** | | | **100.0%** | **0.84 [0.81, 0.86]** |

Heterogeneity: $Tau^2 = 0.00$; $Chi^2 = 2.22$, $df = 2$ ($P = 0.33$); $I^2 = 10\%$
Test for overall effect: $Z = 72.25$ ($P < 0.00001$)

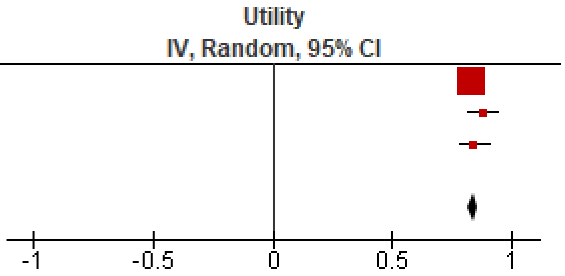

Figure S3.3 Pooled utilities of positive screening result (results not known; among those without cancer)

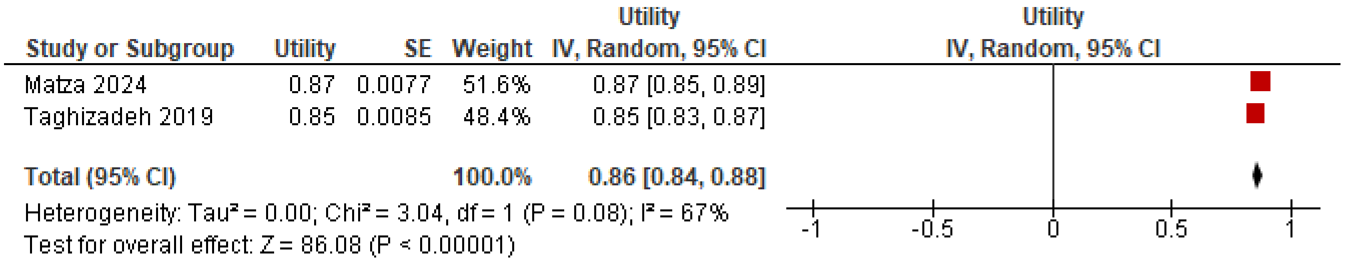

Figure S3.4 Pooled utilities of false positive screening result (up to 1 month after diagnostic workup)

| Study or Subgroup | Utilities | SE | Weight | Utilities IV, Random, 95% CI |
|---|---|---|---|---|
| Bendixen 2019 | 0.88 | 0.01 | 13.7% | 0.88 [0.86, 0.90] |
| Jeppesen 2018 | 0.74 | 0.028 | 11.7% | 0.74 [0.69, 0.79] |
| Koide 2019 | 0.81 | 0.039 | 10.1% | 0.81 [0.73, 0.89] |
| Kuehne 2022 | 0.85 | 0.028 | 11.7% | 0.85 [0.80, 0.90] |
| Lim 2022 | 0.75 | 0.01 | 13.7% | 0.75 [0.73, 0.77] |
| Patel 2023 | 0.83 | 0.011 | 13.6% | 0.83 [0.81, 0.85] |
| Takamiya 2023 | 0.9 | 0.015 | 13.3% | 0.90 [0.87, 0.93] |
| Tramontano 2015 | 0.79 | 0.025 | 12.1% | 0.79 [0.74, 0.84] |
| **Total (95% CI)** | | | **100.0%** | **0.82 [0.77, 0.87]** |

Heterogeneity: Tau² = 0.00; Chi² = 123.80, df = 7 (P < 0.00001); I² = 94%
Test for overall effect: Z = 34.88 (P < 0.00001)

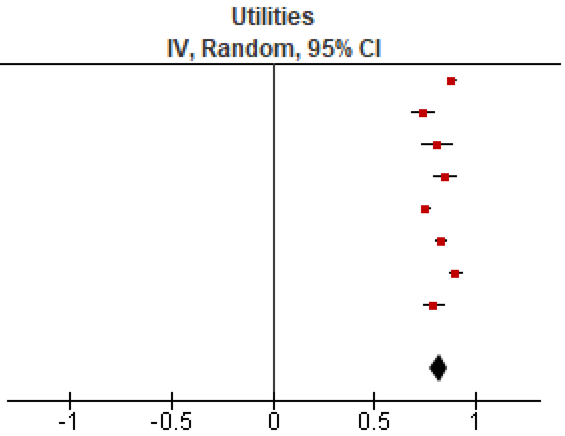

Figure S3.5 Pooled utilities of lung cancer, at diagnosis/treatment naive, stage I-IIIA

| Study or Subgroup | Utilities | SE | Weight | Utilities IV, Random, 95% CI |
|---|---|---|---|---|
| Erbaycu 2018 | 0.66 | 0.022 | 11.6% | 0.66 [0.62, 0.70] |
| Galetta 2015 | 0.69 | 0.026 | 11.4% | 0.69 [0.64, 0.74] |
| Gridelli 2012 | 0.7 | 0.017 | 11.8% | 0.70 [0.67, 0.73] |
| Kuehne 2022 | 0.72 | 0.046 | 10.0% | 0.72 [0.63, 0.81] |
| Kunikane 2019 | 0.78 | 0.021 | 11.6% | 0.78 [0.74, 0.82] |
| Tramontano 2015 | 0.79 | 0.03 | 11.1% | 0.79 [0.73, 0.85] |
| Tse 2020 | 0.73 | 0.057 | 9.1% | 0.73 [0.62, 0.84] |
| Yang 2023 | 0.69 | 0.026 | 11.4% | 0.69 [0.64, 0.74] |
| Yoh 2020 | 0.86 | 0.006 | 12.1% | 0.86 [0.85, 0.87] |
| **Total (95% CI)** | | | **100.0%** | **0.74 [0.67, 0.80]** |

Heterogeneity: $Tau^2 = 0.01$; $Chi^2 = 207.83$, df = 8 ($P < 0.00001$); $I^2 = 96\%$
Test for overall effect: $Z = 21.48$ ($P < 0.00001$)

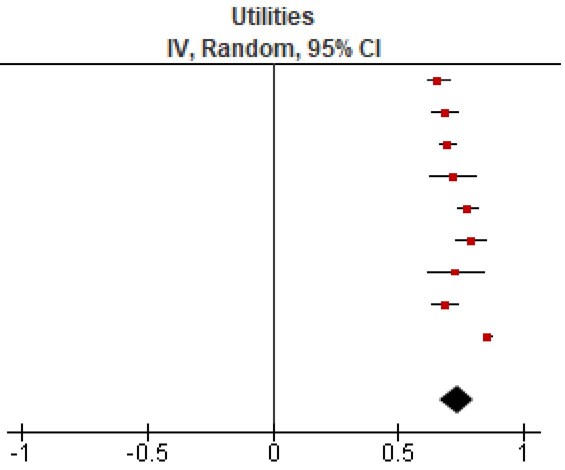

Figure S3.6 Pooled utilities of lung cancer patients at diagnosis/treatment naive, stage IIIB-IV

| Study or Subgroup | Utilities | SE | Weight | Utilities IV, Random, 95% CI |
|---|---|---|---|---|
| Ganti 2025 | 0.74 | 0.012 | 37.2% | 0.74 [0.72, 0.76] |
| Kuehne 2022 | 0.74 | 0.025 | 21.9% | 0.74 [0.69, 0.79] |
| Tramontano 2015 | 0.78 | 0.009 | 40.9% | 0.78 [0.76, 0.80] |
| **Total (95% CI)** | | | **100.0%** | **0.76 [0.73, 0.79]** |

Heterogeneity: $Tau^2 = 0.00$; $Chi^2 = 8.08$, $df = 2$ ($P = 0.02$); $I^2 = 75\%$
Test for overall effect: $Z = 47.27$ ($P < 0.00001$)

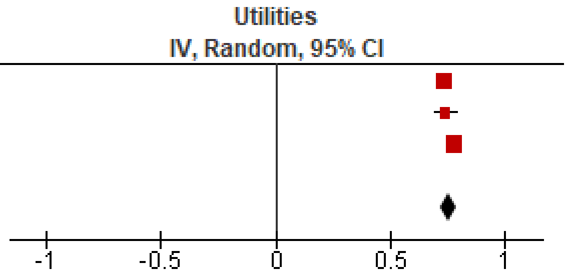

Figure S3.7 Pooled utilities of lung cancer patients on first-line adjuvant treatment (none only receiving surgery), stage I-IIIA

| Study or Subgroup | Utilities | SE | Weight | Utilities IV, Random, 95% CI |
|---|---|---|---|---|
| Chouaid 2013 | 0.71 | 0.023 | 7.7% | 0.71 [0.66, 0.76] |
| Galetta 2015 | 0.65 | 0.033 | 7.1% | 0.65 [0.59, 0.71] |
| Gridelli 2012 | 0.7 | 0.015 | 8.0% | 0.70 [0.67, 0.73] |
| Iyer 2012 | 0.63 | 0.015 | 8.0% | 0.63 [0.60, 0.66] |
| Kuehne 2022 | 0.71 | 0.017 | 8.0% | 0.71 [0.68, 0.74] |
| Kunikane 2019 | 0.76 | 0.016 | 8.0% | 0.76 [0.73, 0.79] |
| Limwattananon 2018 | 0.56 | 0.027 | 7.5% | 0.56 [0.51, 0.61] |
| Matter-Walstra 2014 | 0.85 | 0.013 | 8.1% | 0.85 [0.82, 0.88] |
| Nafees 2017 | 0.58 | 0.018 | 7.9% | 0.58 [0.54, 0.62] |
| Tramontano 2015 | 0.77 | 0.008 | 8.2% | 0.77 [0.75, 0.79] |
| Tse 2020 | 0.78 | 0.013 | 8.1% | 0.78 [0.75, 0.81] |
| Yalcin Balcik 2016 | 0.76 | 0.058 | 5.5% | 0.76 [0.65, 0.87] |
| Yoh 2020 | 0.85 | 0.019 | 7.9% | 0.85 [0.81, 0.89] |
| **Total (95% CI)** | | | **100.0%** | **0.72 [0.67, 0.76]** |

Heterogeneity: Tau² = 0.01; Chi² = 326.76, df = 12 (P < 0.00001); I² = 96%
Test for overall effect: Z = 30.27 (P < 0.00001)

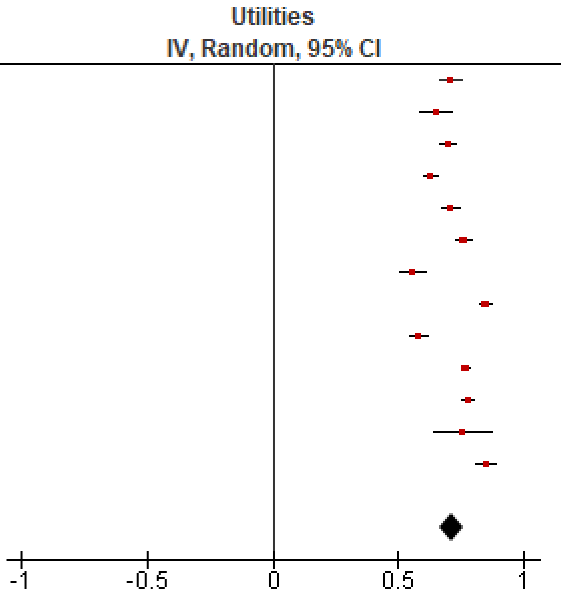

Figure S3.8 Pooled utilities of lung cancer patients on first-line treatment, stage IIIB-IV

| Study or Subgroup | Utilities | SE | Weight | Utilities IV, Random, 95% CI |
|---|---|---|---|---|
| Bendixen 2019 | 0.77 | 0.011 | 9.3% | 0.77 [0.75, 0.79] |
| Bertolaccini 2023 | 0.88 | 0.002 | 9.4% | 0.88 [0.88, 0.88] |
| Jeppesen 2018 | 0.68 | 0.031 | 8.3% | 0.68 [0.62, 0.74] |
| Koide 2019 | 0.74 | 0.022 | 8.8% | 0.74 [0.70, 0.78] |
| Lim 2022 | 0.58 | 0.013 | 9.2% | 0.58 [0.55, 0.61] |
| Naik 2017 | 0.78 | 0.02 | 8.9% | 0.78 [0.74, 0.82] |
| Patel 2023 | 0.76 | 0.014 | 9.2% | 0.76 [0.73, 0.79] |
| Rauma 2015 | 0.83 | 0.007 | 9.4% | 0.83 [0.82, 0.84] |
| Takamiya 2023 | 0.76 | 0.018 | 9.0% | 0.76 [0.72, 0.80] |
| Teramatsu 2022 | 0.94 | 0.016 | 9.1% | 0.94 [0.91, 0.97] |
| Tramontano 2015 | 0.8 | 0.006 | 9.4% | 0.80 [0.79, 0.81] |
| **Total (95% CI)** | | | **100.0%** | **0.78 [0.73, 0.83]** |

Heterogeneity: $Tau^2 = 0.01$; $Chi^2 = 933.52$, df = 10 ($P < 0.00001$); $I^2 = 99\%$
Test for overall effect: $Z = 30.70$ ($P < 0.00001$)

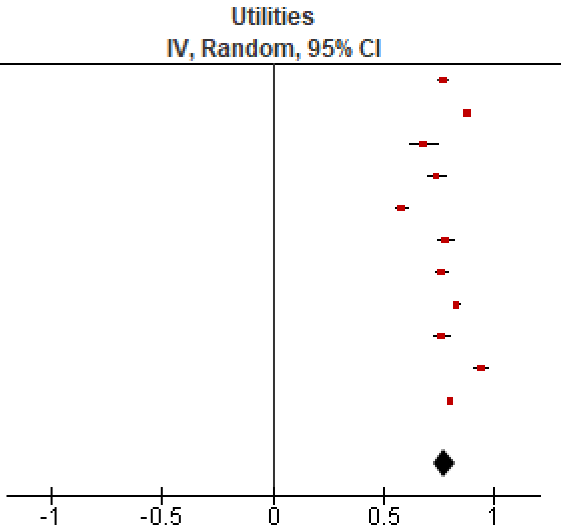

Figure S3.9 Pooled utilities of lung cancer patients, after first-line treatment completion and non-progressive (post-surgery, on maintenance therapy, or survivors), stage I-IIIA

| Study or Subgroup | Utilities | SE | Weight | Utilities IV, Random, 95% CI |
|---|---|---|---|---|
| Galetta 2015 | 0.61 | 0.044 | 8.7% | 0.61 [0.52, 0.70] |
| Gridelli 2012 | 0.77 | 0.016 | 16.0% | 0.77 [0.74, 0.80] |
| Lal 2015 | 0.72 | 0.047 | 8.1% | 0.72 [0.63, 0.81] |
| Limwattananon 2018 | 0.68 | 0.031 | 11.8% | 0.68 [0.62, 0.74] |
| Nafees 2017 | 0.8 | 0.037 | 10.2% | 0.80 [0.73, 0.87] |
| Naik 2017 | 0.77 | 0.03 | 12.1% | 0.77 [0.71, 0.83] |
| Tramontano 2015 | 0.76 | 0.013 | 16.7% | 0.76 [0.73, 0.79] |
| Yoh 2020 | 0.81 | 0.014 | 16.5% | 0.81 [0.78, 0.84] |
| **Total (95% CI)** | | | **100.0%** | **0.75 [0.71, 0.78]** |

Heterogeneity: Tau² = 0.00; Chi² = 32.14, df = 7 (P < 0.0001); I² = 78%
Test for overall effect: Z = 42.07 (P < 0.00001)

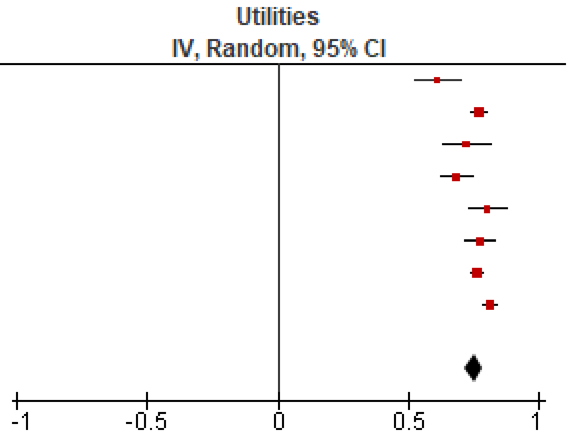

Figure S3.10 Pooled utilities of lung cancer patients, after first-line treatment completion and non-progressive (on maintenance therapy, or survivors) stage IIIB-IV

Table S3.4 Study characteristics, other preference data, direct comparison

| Study, Country<br>Study description<br>Eligibility and Recruitment<br>Sample size; Age; Female; Insured; White (other race/ethnicities); Smoking history; other LC risk factors<br>Screening history<br>History of FP or lung surgery<br>Risk of bias (ROB) | Study data e.g. communicated outcomes of screening, attributes and their levels<br>Format of information and definitions provided<br>Data collection<br>Other information provided | Findings (including relevant subgroup findings) | Knowledge and understanding (including relevant subgroup findings) |
|---|---|---|---|
| **Lung-cancer mortality versus false positives** | | | |
| **Community/public sample** | | | |
| Byrne 2019, U.S.<br><br>Online survey asking individuals to complete a conjoint valuation exercise on hypothetical LCS scenarios.<br><br>Eligibility and Recruitment: 45+ yr current and former smokers with at least 20 pack-yr history via public survey panel (Knowledge Networks); targeted a sample that was approximately 50% female; 25% African American and 25% Hispanic; and 25% rural dwelling<br><br>N=210; 45+ yrs (mean ± SD 61.1 ± 8.5); 52% Female; about half with medical insurance through an employer (% NR); 46% White-non-Hispanic (25% Black, non-Hispanic, 29% Hispanic); 41% current smoker and mean ± SD pack-years: 40 ± 20; NR<br><br>Screening history: NR<br><br>History of FP or lung surgery: NR<br><br>ROB: Some risk | **Communicated Outcomes of Screening & Attribute levels:**<br>• LC mortality reduction by screening: 1%, 10% or 20%<br>• FPs: 10%, 25%, or 40%<br><br>Format & definitions: 22 hypothetical scenarios with 5 attributes and different levels of attributes. Part-worth utilities reflect the desirability (or lack of undesirability) of a given attribute relative to other levels for the same attribute, which sum to 0. Relative importance scores were a measure of how much influence each individual characteristic had on an individual's willingness to be screened. Relative importance scores sum to 100. Unclear if FPs defined.<br><br>Data collection: online survey using conjoint valuation exercise to create unique scenarios, where respondents rate each scenario on a 9-point Likert type rating scale.<br><br>Other attributes:<br>Out-of-pocket costs: $100, $300, or $500<br>HCP recommendation: do not get screened, do get screened, you should make the decision, or you do not discuss screening with your doctor<br>Ease of access: center in a convenient location and open evenings/weekends, convenient location and open 9-5pm weekdays only, inconvenient location but open evenings/weekends, or inconvenient location and open 9-5pm weekdays only | **Part-worth utilities for attribute levels**, means sum to 0 per attribute:<br>FP rate:<br>10%: 21.1<br>25% 2.6<br>40% -23.7<br>"10% much more desirable than a 40%"<br>LC mortality reduction:<br>1%: -31.1<br>10%: 13.5<br>20%: 17.6<br><br>**Relative importance of the attributes** of the LCS scenarios, mean (SD; range), relative importance scores for each individual sum to 100:<br>LC mortality reduction: 17.2 (8.9; 1.8 to 49.3)<br>FP rate: 15.8 (10.4; 0.8 to 49.3)<br>Higher for costs (27.3 ±17.7)<br>("Large variation with income and costs only significant association") | None |
| **Select sample** | | | |
| Fraenkel 2016, U.S. | **Communicated Outcomes of Screening & Attribute levels:** | **Attitudes:** | Knowledge about FPs: |

| | | | |
|---|---|---|---|
| RCT of 3 risk formats on LC screening, to evaluate the effects on screening-related knowledge, beliefs and screening endorsement.<br><br>Eligibility and Recruitment: outpatient pulmonary practices during reminder telephone calls (not for a pulmonary nodule or previous cancer).<br><br>N=254 (Numbers only=84; Numbers+Icon array=86; Numbers+slide presentation=83); age range NR (mean ± SD: 60.9 ± 8.8 yrs); 54.2% Female; NR; race NR; 56.3% current or past smoker of whom 28.7% had 30+ pack-year smoking history and 43.7% never smokers; NR<br><br>Screening history: NR<br><br>History of FP or lung surgery: NR<br><br>ROB: Some risk | • LC mortality: 3 of 250 will die of LC despite getting screened<br>• LC mortality reduction by screening: 1 of 250 will have their life saved from LC by being screened and having cancer detected<br>• FPs: 91 of 250<br><br>Format & definitions: patients were randomized to receive risk information with 1) numbers only, 2) numbers and corresponding icon arrays, or 3) numbers and experienced format (a set of slides illustrating LDCT scans with normal, FP and LC results). The risk information took 4 minutes total to view, at 1 slide per second. FPs: "A false alarm means that the CT scan shows a small spot on the lung that after doing more tests turns out NOT to be cancer. It may take a biopsy or up to two years of repeat CT scans to be sure that a spot on the lung is not cancer"<br><br>**No personalized risk or outcomes presented.**<br><br>Data collection: patients viewed the risk information in 1 of the 3 formats they were randomized to on a computer in a quiet room while a medical assistant directly observed them. Participants were then asked to complete outcome questions.<br><br>Other information:<br>Process of LC screening. | Based on the information given to you, do you think heavy smokers should have screening LDCT scans? (Responding "yes" as endorsement of screening)?<br>Numbers only group: 86.9%<br>Numbers and icon array group: 82.6%<br>Numbers and experience group: 89.3%<br>Entire sample: 85.8% | The percent of participants responding correctly to "Compared to the number of people with lung cancers found by screening, how many people will have a false alarm?" 56.3% (higher for those randomized to the numbers + icon array and numbers + experienced format compared to the numbers only format [39.3%]).<br><br>The percent of participants responding correctly to the "Of all the people screened, about how many will have a false alarm?" 38.6% (higher only for those randomized to the numbers + icon array format [52.3%] compared to the numbers only format [31%]).<br><br>Objective numeracy did not impact findings. |
| **Lung-cancer mortality versus multiple harms** | | | |
| **Community/public sample** | | | |
| Clark 2022, U.S.<br><br>Online, parallel, double-blinded RCT survey of video DA with or without incidental findings information.<br><br>Eligibility and Recruitment: Screen-eligible (USPSTF 2013) via public survey panel (Qualtrics); through quotas ensured 25% Black/African American or Hispanic/Latino, and 25% less than college educated | **Communicated Outcomes of Screening & Attribute levels (over 7 years):**<br>• LC mortality without screening: 21 in 1000<br>• LC mortality with screening: 18 (3 fewer in 1000)<br>• FPs: 206 in 1000<br>• FPs with biopsy: 14 in 1000<br>• Major complications: <1 in 1000 (among those without cancer)<br>• Overdiagnosis: 4 in 1000<br>• Anxiety: "Screening can also cause anxiety from worrying about lung cancer, especially when the CT scan finds nodules."<br>• Incidental findings (for intervention group): 200 in 1000 | **Rating of LCS benefit and harms, mean (SD), scale 1-5 (1=not very important, 5=very important):**<br>Intervention:<br>LC mortality: 3.7 (1.3)<br>FPs: 4.2 (1.0)<br>FPs with biopsy: 4.2 (1.0)<br>Overdiagnosis: 4.2 (1.0)<br>Incidental findings: 4.0 (1.1)<br><br>Control:<br>LC mortality: 3.7 (1.3)<br>FPs: 4.2 (1.1)<br>FP with biopsy: 4.1 (1.1) | Knowledge test (6 questions; 5 of them conceptual 1 on magnitude of LC mortality reduction): mean correct screening knowledge scores increased across groups after viewing the DA (pre 2.6 vs post 5.2 out of 6, diff + 2.7, 95% CI 2.5, 2.8, $p < 0.01$).<br><br>At baseline, only 8.6% of participants correctly |

| | | | |
|---|---|---|---|
| N=348 (I=173; C=175); 55-80 yrs (mean ± SD: 65.4 ± 6.3); 51.4% Female; 93.4% known insurance coverage (private, medicare, Medicaid, military or other); 73.0% White (17.5% Black or African American; 7.8% Hispanic or Latino; 1.7% Other); 70.4% current smoker and pack-years of smoking, mean ± SD: 51.6 (24.2) in current smokers and 61.4 (33.3) in former smokers; NR<br><br>Screening history: NR<br><br>History of FP or lung surgery: NR<br><br>ROB: Low risk | Format & definitions: 4.5 min video DA, with text, images and diagrams (icon arrays of screened and not screened): but also including a 31s long description of information on incidental findings in the intervention group.<br><br>FPs: "CT scans find many nodules or "spots" in the lungs that turn out NOT to be cancer after more tests are done. People with nodules found by screening usually need additional scans to monitor the nodules.<br>FP biopsies: "Some types of biopsies include needles inserted into the chest and surgery to remove part of a lung. Screening can lead to a biopsy in people who do NOT have lung cancer.<br>Major complications: "Serious complications from biopsies can include bleeding, infection, collapsed lung, or rarely death. This means that serious complications can result from biopsies even for people who do not have cancer."<br>Overdiagnosis: "Screening can sometimes find slow growing or harmless "cancers" that never would have caused a problem to the person. This is called over-detection. Doctors can't tell which cancers found by screening will cause problems and which will not, so everyone who is diagnosed with lung cancer will be treated the same way - with surgery, radiation, or chemotherapy. This means that people with harmless "cancers" will be treated needlessly. This is called over-treatment."<br>Incidental findings: "Screening also shows areas outside of the lungs and can find things in places such as the heart, kidneys, and thyroid not related to lung cancer. What we find is often benign and would never cause a problem, but may lead to further testing or a visit with a specialist to find out if it might become a problem. This testing may require additional cost to you, including co-pays and deductibles."<br>Anxiety: "Screening can also cause anxiety from worrying about lung cancer, especially when the CT scan finds nodules."<br>Incidental findings: "Screening also shows areas outside of the lungs and can find things in places such as the heart, kidneys, and thyroid not related to lung cancer. What we find is often benign and would never cause a problem, but may lead to further testing or a visit with a specialist to find out if it might become a problem. This testing may require additional cost to you, including co-pays and deductibles."<br><br>**No personalized risk or outcomes presented.**<br><br>Data collection: participants were randomized to receive the decision aid with or without incidental findings and were then asked to complete knowledge questionnaires before receiving the DA, then after DA completed outcome questions. | Overdiagnosis: 4.2 (1.2)<br>Incidental findings: 3.9 (1.2)<br><br>**Ranking (among 6 outcomes with costs as one)**: Proportion of 1st ranks & mean rank:<br>Intervention:<br>LC mortality 52.0% & 2.5 (1.9)<br>FPs 13.3% & 3.7 (1.7)<br>FPs with biopsy 11% & 3.2 (1.5)<br>Incidental findings 9.2% & 3.6 (1.5)<br>Overdiagnosis 5.8% & 3.7 (1.3)<br><br>Control:<br>LC mortality 56.6%<br>FPs 9.1%<br>FPs with biopsy 10.9%<br>Incidental findings 9.1%<br>Overdiagnosis 6.3%<br><br>A substantial minority (45%) of participants ranked one of harms attributes as most important, even above mortality benefit. | estimated the approximate likelihood of benefitting from screening, with most greatly overestimating the probability. Similarly, only 13.2% of participants correctly answered the knowledge item about overdiagnosis.<br><br>For incidental findings, after viewing the DA, the intervention group had a higher percentage of individuals choosing the correct answer compared with the control group (164/173, 94.8% vs 129/175, 73.7%, 95% CI − 28.4 to − 13.8%, $p < 0.01$). |

| | | | |
|---|---|---|---|
| | Other information:<br>Radiation: “First, when a person gets a CT scan, he or she is exposed to radiation. Rarely, this can cause cancer 10 to 20 years later.”<br>Costs: “While the initial screening test is covered by insurance plans, you may be responsible for copays and deductibles for follow up tests and procedures.” | | |
| Clark 2021, U.S.<br><br>An online pre-post study decision aid for LC screening.<br><br>Eligibility and Recruitment: screen-eligible (USPSTF 2013) via public survey panel (Qualtrics); through quotas ensured 20% Black/African American or Hispanic/Latino, and 25% less than college educated<br><br>N=219; 55-80 yrs (mean ± SD 64.7 ± 6.1); 57.5% Female; 95% Insured (Private, Medicare, Medicaid, Military, or other); 75.4% White (17.4% Black or African American, 5.9% Hispanic or Latino, 1.4% other); 67.1% current smoker and in current smokers, mean ± SD pack-years: 47.2 ± 17.2, former smokers 63.1 ± 30.1; NR<br><br>Screening history: NR<br><br>History of FP or lung surgery: NR<br><br>ROB: Serious risk (for comparisons with overdiagnosis); some risk for other comparisons | **Communicated Outcomes of Screening & Attribute levels (over 7 years):**<br>• LC mortality: 21 in 1000 over 7 yrs<br>• LC mortality reduction by screening: 3 in 1000 (18 still die of LC)<br>• FPs: 365 in 1000<br>• FPs with biopsy: 25 in 1000<br>• Major complications: 3 in 1000<br>• Overdiagnosis: 4 in 1000<br>• Anxiety: “Screening can also cause anxiety from worrying about lung cancer, especially when the CT scan finds nodules.”<br><br>Format & definitions: 3.5 min video DA, with text, images and diagrams (icon arrays of screened and not screened): https://www.youtube.com/watch?v=kNerT-OWKPw.<br><br>**No personalized risk or outcomes presented**.<br><br>See Clark 2022 for definitions.<br><br>Data collection: participants were asked to complete a knowledge questionnaire (7 of 10 questions focused on concepts of overdiagnosis) then received the DA and subsequently completed outcome questions.<br><br>Other information:<br>Radiation: “When a person gets a CT scan, he or she is exposed to radiation. Rarely, this can cause cancer 10 to 20 years later.”<br>Costs: “While the initial screening test is covered by insurance plans, you may be responsible for copays and deductibles for follow up tests and procedures.” | **Rating of LCS attributes, mean (SD), scale 1-5 (1=not very important, 5=very important):**<br>LC mortality: 4.3 (1.0)<br>Complications: 3.7 (1.3)<br>Overdiagnosis (“Reduce risk of diagnosis with LC”: 3.6 (1.2)<br>FPs with biopsy: 3.6 (1.2)<br>FPs: 3.3 (1.2)<br><br>Black/African-American respondents were more likely to rate ‘reducing the chance of diagnosis’ higher than whites (4.0 vs 3.5, diff +0.5, 95% CI 0.1 to 0.9; p=0.02). No significant differences by sex.<br><br>**Ranking (among 6 outcomes with costs as one)**: Proportion of 1st ranks (mean ranks NR):<br>LC mortality 59.4%<br>FPs 7.3%<br>FPs with biopsies 7.3%<br>Overdiagnosis (“Risk of diagnosis with lung cancer) 6.8%<br>Complications 5.9%<br><br>34% ranked one of the harms highest<br><br>≥65 years were more likely to rank ‘reducing the chance of death’ first compared with those <65 (67.0% vs 51.4%, p=0.02).<br>Black/African-American were more likely to rank ‘reducing the chance of diagnosis’ first compared with whites (18.8% vs 5.1%, p=0.02). | Knowledge score (10 questions): pre 2.7±1.7, post 5.8±2.3 (mean diff +3.0, 95%CI 2.7 to 3.3, p<0.001).<br><br>Proportions of respondents able to correctly identify the likelihood of benefiting from screening increased after decision aid viewing (pre 16.4% vs post 38.4%, p<0.001), as did the proportion who accurately estimated the likelihood of being overdiagnosed with screening (24.7% vs 46.1%, p<0.001).<br><br>No difference between demographic groups for both unadjusted and adjusted analyses.<br><br>Individuals indicating they were ‘not likely’ to pursue screening, had a greater improvement in pre–post knowledge score than those indicating that they were ‘very likely’ to pursue screening (+3.5 vs +2.6, diff +0.9; 95% CI 0.1 to 1.8; p=0.023). |
| **False positives versus overdiagnosis** | | | |

| **Select populations** | | | |
|---|---|---|---|
| Hoffman 2018, U.S.<br><br>A pre-post study of online DA<br><br>Eligibility and Recruitment: 55-80 yrs, current or former smokers who quit within the past 15 yrs (no pack-year minimum), members of the Tobacco Treatment Program at University of Texas Cancer Center, seeking smoking cessation services<br><br>N=30; 55-80 yrs (mean: 61.5 yrs); 50% Female; 100% insured (Medicare, Medicaid, job/private insurance, or other); 63.3% White (30.0% Black/African American, 3.3% Hispanic/Latino, 3.3% other); 60.7% current smoker and mean (SD) pack-year history: 30.4 (18.9); NR<br><br>Screening history: NR<br><br>History of FP or lung surgery: NR<br><br>ROB: High risk | **Communicated Outcomes of Screening & Attribute levels (annually for 3 years; follow-up over 6.5 years):**<br>• LC mortality: 21 in 1000<br>• LC mortality reduction by screening: 3 in 1000 (18 still die of LC)<br>• FPs: 365 in 1000 (additional scan)<br>• FPs: 18 in 1000 (test or surgery)<br>• Major complications: 3 in 1000<br>• Overdiagnosis: "presented as cancers that would never become life threatening, and the video explains that some people may be treated for cancer that would never have harmed them."<br><br>Format & definitions: Video decision aid "Lung Cancer screening: Is it Right for Me?", covering benefits and harms of screening with text, images and diagrams (icon arrays of screened and not screened). **USPSTF 2013 "recommends"**. Emphasizes importance of considering this personal decision with their doctor.<br><br>**No personalized risk or outcomes presented.**<br>**Independent review of material.**<br><br>Data collection: patients completed an online baseline questionnaire including items on knowledge of LCS with LDCT. Patients then viewed a video DA, and completed a post-decision aid questionnaire.<br><br>Other information:<br>TPs: 24 in 1000 individuals will have LC<br>Radiation exposure: shown pictorially with other radiation exposures to contextualize: from a flight from NY to LA (0.03 mSv) as lowest exposure to highest for living in the US (3.00 mSv). LDCT scan was presented as 1.50 mSv.<br>The video also describes the natural history of smoking and LC (emphasis of importance of quitting smoking), the USPSTF screening recommendation, the importance of an informed decision, and what the procedure of screening is. | **Relative importance:**<br><br>How concerned are you that your scan may *say you have cancer* when you do not (in other words, a false alarm)? (0 = Not at all concerned; 10 = Extremely concerned), median (IQR; range): 7.0 (5; 2-10)<br><br>How concerned are you about being treated for a lung cancer that never would have harmed you (in other words, overdiagnosis)? (0 = Not at all concerned; 10 = Extremely concerned), median (IQR; range): 6.0 (5; 1-10)<br><br>**Data used:**<br>FPs slightly more important than overdiagnosis | Mean knowledge scores (12-item measure): pre, 5.7 (47.3% correct), post 9.6 (80.3% correct); 3.9 mean difference (SD = 2.9, 95% CI 2.9–5.0; p<0.001). |
| Volk 2020, U.S.<br>(Data from associated paper Maki 2021)<br><br>Randomized controlled trial of a patient DA versus standard education materials (SEM) | **Communicated Outcomes of Screening & Attribute levels:**<br>• LC mortality without screening: 21 in 1000<br>• LC mortality reduction by screening: 3 fewer in 1000 (18 still die of LC)<br>• FPs: 356 in 1000<br>• FPs invasive procedures: 18 in 1000 (test or surgery)<br>• Major complications: 3 in 1000<br>• Overdiagnosis: "presented as cancers that would never become life threatening, and the video explains | **Relative importance** ("How concerned are you about…? 11-point scale: 0-4 no/some risk; 5-8 concerned; 9-10 very concerned):<br>False positives: 81.4% concerned/very concerned<br>Overdiagnosis: 83.8% concerned/very concerned | Correct responses to LCS knowledge, correct response, mean % (95% CI)<br><br>1 week<br>Intervention (n=235): 57.5% (54.7-60.3) |

| | | | |
|---|---|---|---|
| Eligibility and Recruitment: clients aged 55-77 years who reported a 30-plus pack-year smoking history via 13 state tobacco quit lines and having an interest in screening<br><br>N=204; mean ± SD age 61.4 ± 4.9; 59% Female; 91.7%; 69.6% White (1.5% American Indian or Alaska Native, 26.5% Black, 2.5% Hispanic or Latino); mean pack-year history (SD) 53.6 (23.8); NR<br><br>Screening history: NR<br><br>History of FP or lung surgery: NR<br><br>ROB: High risk | that some people may be treated for cancer that would never have harmed them."<br><br>Format & definitions: PDA arm: 9.5-minute narrated video, Lung Cancer Screening: Is It Right for Me? The narrated DA included information about (1) eligibility for lung cancer screening and a calculation of tobacco pack-year smoking history, (2) lung cancer epidemiology and risk factors, (3) a video of a patient in a CT scanner, (4) icon arrays to graphically depict the magnitude of mortality reduction, false-positive results, and harms from invasive diagnostic procedures, and (5) radiation exposure depicted within the context of other sources of radiation (e.g., a screening mammogram). Smoking cessation was emphasized throughout the DA. The DA was updated according to USPSTF 2014 recommendations. USPSTF "recommends"<br><br>**No personalized risk or outcomes presented. Independent review of material.**<br><br>**Judgement of Net benefit: Moderate**<br><br>SEM arm: The SEM material was a 2-page brochure from a lung cancer advocacy group and included structured questions a patient can ask a physician about lung cancer screening. Benefits and harms were described but no probabilities of outcomes were included. (not eligible for this review)<br><br>Data collection: Baseline, 1-week, 3-month, and 6-month questionnaires via telephone or mailed if not reachable by phone.<br><br>Other information:<br>TPs: 24 in 1000 individuals will have LC<br>Radiation exposure: shown pictorially with other radiation exposures to contextualize: from a flight from NY to LA (0.03 mSv) as lowest exposure to highest for living in the US (3.00 mSv). LDCT scan was presented as 1.50 mSv.<br>The video also describes the natural history of smoking and LC (emphasis of importance of quitting smoking), the USPSTF screening recommendation, the importance of an informed decision, and what the procedure of screening is. | Factors associated with obtaining LCS at 6-months, OR (95% CI) (Maki 2021):<br><br>False alarm: Very important 1.98 (0.55-7.13)<br>False alarm: Important 2.25 (0.65-7.78)<br>Over diagnosis: Very important (9-10 = 1): 0.17 (0.04 – 0.65), p<0.05<br>Over diagnosis: Important (5-8 = 1): 0.15 (0.05 – 0.53), p>0.001<br><br>**Data used:**<br>Asking about concern, FPs are of similar importance as overdiagnosis. When associated to screening uptake, the information on FPs nonsignificantly associated with higher uptake whereas the information on overdiagnosis significantly associated with lower uptake, to a large degree. Overdiagnosis appeared to be more important than FPs. | Control (n=233): 40.1% (37.9-42.3)<br>Difference: 17.4% (13.9-21.0), p<0.001<br><br>6 months<br>Intervention (n=218): 49.9% (47.5-52.3)<br>Control (n=225): 40.0% (37.6-42.4)<br>Difference: 9.9% (6.5-13,3), p<0.001<br><br>Tests were adjusted for age, sex, race/ethnicity, educational level, insurance status, quitline service provider, and recruitment method |
| **False positives vs. psychosocial harms from screening** | | | |
| **Primary/secondary care** | | | |
| Lillie 2017, U.S. | **Communicated Outcomes of Screening & Attribute levels (at 6.5 years):**<br>• LC mortality: 21 in 1000 | **Relative importance of outcomes (1-5 (1='not at all important', 2='slightly** | None |

| | | |
|---|---|---|
| RCT with individuals participating in the Veterans Health Administration LCS Demonstration Project. Participants randomized in 2:1 ratio to 1) direct LCS invitation with DA, or 2) Usual care under PCP with annual smoking cessation clinical reminders.<br><br>Eligibility and Recruitment: screen-eligible (USPSTF 2013), members of the Minneapolis VA Health Care System<br><br>N=384 in intervention group; 55-80 yrs (mean ± SD: 65.1 ± 5.4); 4.9% Female; 100% insured (VHA patients); 90.6% White (7.3% African American, 0.3% Hispanic/Latino only, 1.3% Native American/Alaskan Native); 44% current smoker and median (IQR) pack-year history: 48 (20); Spouse or family member with LC: 32.0%<br><br>Screening history: NR<br><br>History of FP or lung surgery: NR<br><br>ROB: High risk | • LC mortality reduction by screening: 3 in 1000 (18 still die of LC)<br>• FPs: 365 in 1000<br>• FP invasive procedures: 25 in 1000<br>• Major complications among FPs: 3 in 1000<br>• Overdiagnosis: Sometimes screening tests find cancers that would have never caused problems. This is called overdiagnosis. Unfortunately, it is often impossible to tell which cancers fall into this category. So, there is a very small chance someone may be treated unnecessarily for a cancer that would not have harmed them<br>• Stress/Anxiety: It is normal to feel stressed or anxious while waiting for your results or if you have something that is suspicious for lung cancer. Most patients with suspicious findings are reassured when they learn that most of these are false positives.<br><br>**Format & definitions:** the intervention group received a mailed LCS invitation that had a letter to call the LCS coordinator for a discussion about screening and to schedule an appointment with or without additional phone calls. They also received a paper copy of the VHA-developed DA. The control group only received annual smoking cessation clinical reminders. All patients had access to the comprehensive MVAHCS Tobacco Cessation Program.<br><br>FPs: "Screening for lung cancer by LDCT scan may find something that is suspicious but, after further testing, turns out not to be cancer. This is called a "false positive" … "Most false positive results are resolved with further LDCT testing."<br>FPs (invasive testing): "extra testing that involved putting a tube in the body or having surgery (these are called "invasive procedures")."<br>Major complications: "Complications can include bleeding, infections, or rarely, a collapsed lung."<br><br>**No personalized risk or outcomes presented.**<br>**Independent review of material.**<br><br>**Data collection:** surveys were administered 3 months after randomization and receipt of the DA. A survey packet was mailed with a study invitation letter, a self-administered survey and a postage-paid return envelope. After 21 days non-respondents received a second survey packet. Intervention group only: LCS completion dates were taken from the VHA medical records. LCS completion was an LDCT completed before the survey return date.<br><br>**Other information:** | **important', 3='neutral', 4='very important', 5='extremely important')**<br>Very or extremely important:<br>False positive results: 30.8%<br>Anxiety of waiting for the results: 28.0%<br><br>(Incidental findings were measured but there was no data or information on these in the decision aid) |

| | |
|---|---|
| | TPs: "26 people would get a true positive (they did have lung cancer)."<br>Radiation: "Exposure to radiation from LDCT scans increases your risk of cancer by a very small amount. We want to keep your chances of getting cancer from radiation very low. We do that by using "low-dose" CTs that use much less radiation than a standard CT."<br>Cost: "If you are charged co-pays for your VA visits, you will be charged a $50.00 co-pay for the day you have the low-dose chest Computed Tomography scan (LDCT). Talk with the Lung Cancer Screening coordinator if you are charged co-pays. Scheduling the scan on the same day as another visit may decrease the total charges."<br>The decision aid also included information on what lung cancer is, the lung cancer screening process, and eligibility. Both groups had access to the MVAHCS Tobacco Cessation Program which included assessments of smoking status via a clinical reminder, patient education materials, individual and group behavioral therapy and pharmacotherapy. |

Table S3.5. Study characteristics, other preference data, indirect comparisons

| **Study, Country, Study ID**<br>**Study description**<br>**Recruitment setting**<br>**Sample size; Age; Female Gender; Insured; White (other race/ethnicities); Smoking history; other LC risk factors**<br>**Screening history**<br>**History of FP or lung surgery**<br>**Risk of bias (ROB)** | **Study data e.g. communicated outcomes of screening, attributes and their levels**<br>**Format of information and definitions provided**<br>**Judgement of Net Benefit**<br>**Data collection**<br>**Other information provided** | **Findings reported by authors (including relevant subgroup findings) & Data used in review analysis** | **Knowledge and understanding (including relevant subgroup findings)** |
|---|---|---|---|
| **Community/public samples of those eligible to screen** | | | |
| Lau 2021, U.S.<br>Study #: 1<br><br>Evaluation of a web-based, patient-facing DA for LCS among African Americans<br><br>Eligibility and Recruitment: current/former smokers, aged 45 −77 years, did not have a history of lung cancer, via community-based organizations on the east side of Detroit that served the African American community. | **Communicated Outcomes of Screening & Attribute levels:**<br>• **Benefits and harms based on personalized 6-year risk of cancer via modelling**:<br>• LC mortality reduction by screening (per 1000): 10% risk: 20 fewer; 6% risk: 11 fewer; 5% risk: 10 fewer; 4% risk; 8 fewer; 3% risk 6 fewer: 2% risk: 4 fewer; 1%: 2 fewer; 0.5%: 1 fewer (1000-person arrays of screened vs. not screened)<br>• FPs: 365 in 1000<br>• FP invasive procedures: 18 in 1000<br>• Major complications: 3 in 1000<br>• Overdiagnosed: 1 in 10 cancers found by screening<br><br>Format & definitions: A modified version of shouldiscreen.com, a web-based DA, such as adding information about insurance | **Preference/intentions for screening (if free)**:<br>33% among 15 eligible<br>64% among 59 not eligible<br>51% among all<br><br>**Screening uptake** (n=15) at 6-8 months:<br>Only 1 eligible under USPSTF and underwent screening; 2 were not eligible, but still given screening recommendation and 1 underwent screening. | Knowledge (overall (max=13): before mean (SD) 5.69 (1.94); after 7.09 (2.3); p <0.001<br>Possible benefits (max 3): before 1.38 (0.84) after 1.61 (0.89)<br>Possible harms (max 3): before 1.20 (0.79) after 1.47 (0.95)<br><br>60.8% of the participants had trouble understanding written health information |

| Study, Country, Study ID<br>Study description<br>Recruitment setting<br>Sample size; Age; Female Gender; Insured; White (other race/ethnicities); Smoking history; other LC risk factors<br>Screening history<br>History of FP or lung surgery<br>Risk of bias (ROB) | Study data e.g. communicated outcomes of screening, attributes and their levels<br>Format of information and definitions provided<br>Judgement of Net Benefit<br>Data collection<br>Other information provided | Findings reported by authors (including relevant subgroup findings) & Data used in review analysis | Knowledge and understanding (including relevant subgroup findings) |
|---|---|---|---|
| N=74; 45-77 (mean ± SD 62.7 ± 6.84); 49% Female; NR; 100% African American; 69% current smokers, 25% reported smoking history of 30 pack-years or more; NR<br><br>Screening history: NR<br><br>History of FP or lung surgery: NR<br><br>ROB: Some risk (intentions); high risk (uptake) | coverage and eliminating icon arrays. Includes USPSTF eligibility criteria.<br><br>FPs: Screening works by finding lung nodules, some of which are cancer, but most nodules detected on the low-dose CT scan for lung cancer screening are not cancer. False positive findings can cause anxiety. However, understanding that most lung nodules found from the CT screening do not represent cancer helps most avoid significant anxiety while they wait for follow-up testing.<br>Overdiagnosis: There are times when screening will find a nodule that is cancer but was never going to cause problems. This can lead to treatments such as surgery, chemotherapy, and radiation which are unnecessary and considered overtreatment. Overtreatment can cause side effects that make you feel sick.<br>Major complications: Sometimes further testing for a finding on lung cancer screening leads to the need for additional testing that can include invasive procedures (biopsies or surgery). These tests are risky and can cause harm in people that end up not having had cancer. Invasive procedures can result in minor complications such as bleeding and infections. They can also result in more major complications such as a collapsed lung and even death.<br>**Personalized risk and outcomes presented.**<br>**Independent review of material at study site.**<br><br>**Judgement of Net benefit: High**<br><br>Data collection: Independent review of DA. Paper or online pre and post surveys after viewing the website. After 6 month, participants were contracted up to 3 times to complete a follow-up phone survey on screening completion. Knowledge questions NR.<br><br>Other information:<br>Radiation exposure (6 months of radiation from every day environment) | **Data used:**<br>Intentions: 33% (eligible)<br>Uptake: 100% (1/1) | The concept of risk as a probability was not well understood among the participants. |
| Lau 2015, U.S.<br>Study #: 2 | **Communicated Outcomes of Screening & Attribute levels:**<br>• **Benefits and harms based on personalized 6-year risk of cancer via modelling**: | **Attitudes:** | Knowledge (overall) (max score 14): Before mean |

| **Study, Country, Study ID**<br>**Study description**<br>**Recruitment setting**<br>**Sample size; Age; Female Gender; Insured; White (other race/ethnicities); Smoking history; other LC risk factors**<br>**Screening history**<br>**History of FP or lung surgery**<br>**Risk of bias (ROB)** | **Study data e.g. communicated outcomes of screening, attributes and their levels**<br>**Format of information and definitions provided**<br>**Judgement of Net Benefit**<br>**Data collection**<br>**Other information provided** | **Findings reported by authors (including relevant subgroup findings) & Data used in review analysis** | **Knowledge and understanding (including relevant subgroup findings)** |
|---|---|---|---|
| Before-after study of a web-based DA<br><br>Eligibility and Recruitment: volunteers, current/former smokers, aged 45–80 years, with no previous history of lung cancer and no CT in the previous year through the University of Michigan<br><br>N=60; 45-80 (mean ± SD 60.6 ±7.3); 50% Female; NR; 88% White (12% Black); 27% current smoker, 50% quit > 15 years ago; 24 ± 24 pack-years; mean 6-year risk 1.2%; 63% college+ educated<br><br>Screening history: 0% in previous year<br><br>History of FP or lung surgery: NR<br><br>ROB: Low risk | • LC mortality reduction by screening (per 1000): 10% risk: 20 fewer; 6% risk: 11 fewer; 5% risk: 10 fewer; 4% risk; 8 fewer; 3% risk 6 fewer: 2% risk: 4 fewer; 1%: 2 fewer; 0.5%: 1 fewer (1000-person arrays of screened vs. not screened)<br>• FPs: 365 in 1000<br>• FP invasive procedures: 18 in 1000<br>• Major complications: 3 in 1000<br>• Overdiagnosed: 1 in 10 cancers found by screening<br><br>Format & definitions: Independent review of DA. A web-based decision aid (shouldiscreen.com) that provided information about LDCT screening. The DA computed baseline lung cancer risk and an individual's chances of benefiting from, and risk of being harmed by, screening (using the PLCOm2012 model).<br><br>FPs: Screening works by finding lung nodules, some of which are cancer, but most nodules detected on the low-dose CT scan for lung cancer screening are not cancer. False positive findings can cause anxiety. However, understanding that most lung nodules found from the CT screening do not represent cancer helps most avoid significant anxiety while they wait for follow-up testing.<br>Overdiagnosis: There are times when screening will find a nodule that is cancer but was never going to cause problems. This can lead to treatments such as surgery, chemotherapy, and radiation which are unnecessary and considered overtreatment. Overtreatment can cause side effects that make you feel sick.<br>Major complications: Sometimes further testing for a finding on lung cancer screening leads to the need for additional testing that can include invasive procedures (biopsies or surgery). These tests are risky and can cause harm in people that end up not having had cancer. Invasive procedures can result in minor complications such as bleeding and infections. They can also result in more major complications such as a collapsed lung and even death.<br><br>**Personalized risk and outcomes presented.**<br>**Independent review of material.** | Considering both the risks and benefits of CT screening, how good a choice is CT screening as a way to reduce your risk of dying from cancer? (1=not effective; 10=extremely effective)<br>8.09 (2.07) among 11 eligible<br>6.32 (3.20) among ineligible<br><br>**Intentions:**<br>Preference for screening (if free):<br>100% among 11 eligible<br>69% among 49 ineligble<br><br>**Data used:**<br>**Attitudes:**<br>(1=not effective; 10=extremely effective) 8.09 (2.07) among 11 eligible ("at least 75% think screening is effective")<br>**Intentions:**<br>100% among 11 eligible | (SD) 7.52 (1.89); After 10.93 (2.19); p<0.0001<br><br>Possible benefits (max 3): before 1.95 (0.77) after 2.70 (0.56)<br>Possible harms (max 3): before 1.20 (0.99) after 2.23 (1.0) |

| Study, Country, Study ID<br>Study description<br>Recruitment setting<br>Sample size; Age; Female Gender; Insured; White (other race/ethnicities); Smoking history; other LC risk factors<br>Screening history<br>History of FP or lung surgery<br>Risk of bias (ROB) | Study data e.g. communicated outcomes of screening, attributes and their levels<br>Format of information and definitions provided<br>Judgement of Net Benefit<br>Data collection<br>Other information provided | Findings reported by authors (including relevant subgroup findings) & Data used in review analysis | Knowledge and understanding (including relevant subgroup findings) |
|---|---|---|---|
| | **Judgement of Net benefit: High**<br><br>Data collection: Online survey before and after viewing the shoudliscfreen.com website<br><br>Other information:<br>$100-$400 if uninsured<br>Radiation exposure: 6 months of radiation from every day environment | | |
| Behr 2024, France, Germany, Italy and the Netherlands<br>Study#: 3<br><br>Online cross-sectional survey aimed at willingness of individuals to participate in LCS, with or without additional screening for COPD and CVD<br><br>Eligibility and Recruitment: (former) smokers aged 50–75 years (no smoking duration/intensity criteria) recruited by online market research firm via various mechanisms; convenience sampling<br><br>N= 1470 (France = 391, Germany n = 338, Italy n = 399, and the Netherlands n = 342); 50-75 (median age category 55-60), 48.8% Female; NR; NR; 54% current smokers & 8.2% occasional smokers | **Communicated Outcomes of Screening & Attribute levels: (three screens)**<br><br>• LC mortality: 3 fewer per 1000<br>• FPs: 193 per 1000<br>• Invasive procedures: 13 (of 193) per 1000<br>• Overdiagnosis: 4 per 1000 (among 41 diagnosed cancers)<br>• Incidental findings: 350 will have a finding suggesting emphysema; after further testing, 313 will be treated<br><br>Format & definitions: Online survey included a 1000-person pictogram; FPs: need an extra scan but will not have lung cancer; Invasive procedure (biopsy) to rule out lung cancer; Overdiagnosis: cancers that would never have caused the person harm (overdiagosed); Incidental findings presented as possible benefit.<br><br>**No personalized risk and outcomes presented**.<br>**Independent review of material.**<br><br>**Judgement of Net benefit: Moderate**<br><br>Data collection: Online survey asked How likely would you be to participate in lung cancer screening? And presented a scale between 0-100 with deciles shown. “somewhat likely” was stated for 60-80 | **Screening intentions:**<br>Likelihood to screen mean 71.2 out of 100 (SD 24.1) (68.5 to 72.5 across countries)<br><br>Subgroups:<br>Current vs. former smokers: 73% vs 69%<br><br>**Data used:**<br>Intentions 25-50% | None |

| Study, Country, Study ID<br>Study description<br>Recruitment setting<br>Sample size; Age; Female Gender; Insured; White (other race/ethnicities); Smoking history; other LC risk factors<br>Screening history<br>History of FP or lung surgery<br>Risk of bias (ROB) | Study data e.g. communicated outcomes of screening, attributes and their levels<br>Format of information and definitions provided<br>Judgement of Net Benefit<br>Data collection<br>Other information provided | Findings reported by authors (including relevant subgroup findings) & Data used in review analysis | Knowledge and understanding (including relevant subgroup findings) |
|---|---|---|---|
| Screening history: NR (but screening not available in these countries)<br><br>History of FP or lung surgery: NR<br><br>ROB: Some concerns | Other information:<br>Data for other screening tests, but not considered for this review. | | |
| Clark 2022, U.S.<br>Study #: 4<br><br>Online, parallel, double-blinded RCT survey on video DA with or without incidental findings information.<br><br>Eligibility and Recruitment: Screen-eligible (USPSTF 2013) via public survey panel (Qualtrics); through quotas ensured 25% Black/African American or Hispanic/Latino, and 25% less than college educated<br><br>N=348 (I=173; C=175); 55-80 yrs (mean ± SD: 64.5 ± 6.3); 51.4% Female; 93.4% known insurance coverage (private, Medicare, Medicaid, military or other); 73.0% White (17.5% Black or African American; 7.8% Hispanic or Latino; 1.7% Other); 70.4% current smoker and pack-years of smoking, mean ± SD: 51.6 (24.2) | **Communicated Outcomes of Screening & Attribute levels (over 7 years):**<br>• LC mortality without screening: 21 in 1000<br>• LC mortality with screening: 18 (3 fewer in 1000)<br>• FPs: 206 in 1000<br>• FPs with biopsy: 14 in 1000<br>• Major complications: <1 in 1000<br>• Overdiagnosis: 4 in 1000<br>• Anxiety: "Screening can also cause anxiety from worrying about lung cancer, especially when the CT scan finds nodules."<br>• Incidental findings (for intervention group): 200 in 1000<br><br>Format & definitions: 4.5 min video DA, with text, images and diagrams (icon arrays of screened and not screened): but also including a 31s long description of information on incidental findings in the intervention group.<br><br>FPs: "CT scans find many nodules or "spots" in the lungs that turn out NOT to be cancer after more tests are done. People with nodules found by screening usually need additional scans to monitor the nodules.<br>FP biopsies: "Some types of biopsies include needles inserted into the chest and surgery to remove part of a lung. Screening can lead to a biopsy in people who do NOT have lung cancer.<br>Major complications: "Serious complications from biopsies can include bleeding, infection, collapsed lung, or rarely death. This | **Screening intentions:**<br>Intervention:<br>Strongly agree: 12.1%<br>Agree: 28.3%<br>Disagree: 37.6%<br>Strongly disagree: 22.0%<br><br>Control:<br>Strongly agree: 12.0%<br>Agree: 29.7%<br>Disagree: 40.6%<br>Strongly disagree: 17.7%<br><br>**Data used:**<br>Intentions across both groups: 41.1% | Knowledge test (6 questions; 5 of them conceptual 1 on magnitude of LC mortality reduction): mean correct screening knowledge scores increased across groups before and after viewing the DA (pre 2.6 vs post 5.2 out of 6, diff + 2.7, 95% CI 2.5, 2.8, p < 0.01).<br><br>At baseline, only 8.6% of participants correctly estimated the approximate likelihood of benefitting from screening, with most greatly overestimating the probability. Similarly, only 13.2% of participants correctly answered the knowledge item about overdiagnosis. |

| Study, Country, Study ID<br>Study description<br>Recruitment setting<br>Sample size; Age; Female Gender; Insured; White (other race/ethnicities); Smoking history; other LC risk factors<br>Screening history<br>History of FP or lung surgery<br>Risk of bias (ROB) | Study data e.g. communicated outcomes of screening, attributes and their levels<br>Format of information and definitions provided<br>Judgement of Net Benefit<br>Data collection<br>Other information provided | Findings reported by authors (including relevant subgroup findings) & Data used in review analysis | Knowledge and understanding (including relevant subgroup findings) |
|---|---|---|---|
| in current smokers and 61.4 (33.3) in former smokers; NR<br><br>Screening history: NR<br><br>History of FP or lung surgery: NR<br><br>ROB: Low risk | means that serious complications can result from biopsies even for people who do not have cancer."<br>Overdiagnosis: "Screening can sometimes find slow growing or harmless "cancers" that never would have caused a problem to the person. This is called over-detection. Doctors can't tell which cancers found by screening will cause problems and which will not, so everyone who is diagnosed with lung cancer will be treated the same way - with surgery, radiation, or chemotherapy. This means that people with harmless "cancers" will be treated needlessly. This is called over-treatment."<br>Anxiety: "Screening can also cause anxiety from worrying about lung cancer, especially when the CT scan finds nodules."<br>Incidental findings: "Screening also shows areas outside of the lungs and can find things in places such as the heart, kidneys, and thyroid not related to lung cancer. What we find is often benign and would never cause a problem, but may lead to further testing or a visit with a specialist to find out if it might become a problem. This testing may require additional cost to you, including co-pays and deductibles."<br><br>**No personalized risk or outcomes presented.**<br>**Independent review of material.**<br><br>**Judgement of Net benefit: Low (both groups)**<br><br>Data collection: participants were randomized to receive the decision aid with or without incidental findings and were then asked to complete knowledge questionnaires before receiving the DA, then after DA completed outcome questions.<br><br>Other information:<br>TPs: no data<br>Radiation: "First, when a person gets a CT scan, he or she is exposed to radiation. Rarely, this can cause cancer 10 to 20 years later." | | For incidental findings, after viewing the DA, the intervention group had a higher percentage of individuals choosing the correct answer compared with the control group (164/173, 94.8% vs 129/175, 73.7%, 95% CI − 28.4 to − 13.8%, p < 0.01). |

| Study, Country, Study ID<br>Study description<br>Recruitment setting<br>Sample size; Age; Female Gender; Insured; White (other race/ethnicities); Smoking history; other LC risk factors<br>Screening history<br>History of FP or lung surgery<br>Risk of bias (ROB) | Study data e.g. communicated outcomes of screening, attributes and their levels<br>Format of information and definitions provided<br>Judgement of Net Benefit<br>Data collection<br>Other information provided | Findings reported by authors (including relevant subgroup findings) & Data used in review analysis | Knowledge and understanding (including relevant subgroup findings) |
|---|---|---|---|
| | Costs: "While the initial screening test is covered by insurance plans, you may be responsible for copays and deductibles for follow up tests and procedures." | | |
| Clark 2021, U.S.<br>Study #: 5<br><br>An online pre-post study decision aid for LC screening.<br><br>Eligibility and Recruitment: screen-eligible (USPSTF 2013) via public survey panel (Qualtrics); through quotas ensured 20% Black/African American or Hispanic/Latino, and 25% less than college educated<br><br>N=219; 55-80 yrs (mean ± SD 64.7 ± 6.1); 57.5% Female; 95% Insured (Private, Medicare, Medicaid, Military, or other); 75.4% White (17.4% Black or African American, 5.9% Hispanic or Latino, 1.4% other); 67.1% current smoker and in current smokers, mean ± SD pack-years: 47.2 ± 17.2, former smokers 63.1 ± 30.1; NR<br><br>Screening history: NR<br><br>History of FP or lung surgery: NR<br><br>ROB: Some risk | **Communicated Outcomes of Screening & Attribute levels (over 7 years):**<br>• LC mortality: 21 in 1000<br>• LC mortality reduction by screening: 3 in 1000 (18 still die of LC)<br>• FPs: 365 in 1000<br>• FPs with biopsy: 25 in 1000<br>• Major complications: 3 in 1000<br>• Overdiagnosis: 4 in 1000<br>• Anxiety: "Screening can also cause anxiety from worrying about lung cancer, especially when the CT scan finds nodules."<br><br>Format & definitions: 3.5 min video DA from a prior study, with text, images and diagrams (icon arrays of screened and not screened): https://www.youtube.com/watch?v=kNerT-OWKPw.<br>See Clark 2022 for definitions.<br><br>**No personalized risk or outcomes presented.**<br>**Independent review of material.**<br><br>**Judgement of Net benefit: Low**<br><br>Data collection: participants were asked to complete a knowledge questionnaire (7 of 10 questions focused on concepts of overdiagnosis) then received the DA and subsequently completed outcome questions.<br><br>Other information:<br>TPs: no data<br>Radiation: "When a person gets a CT scan, he or she is exposed to radiation. Rarely, this can cause cancer 10 to 20 years later." | **Intentions, scale 1-4 (1=not likely, 4=very likely):**<br>1: 21.0%<br>2: 22.4%<br>3: 29.7%<br>4: 26.9%<br><br>**Subgroups:** There were no differences between age, sex or race.<br><br>**Data used:**<br>Intentions: 56.6% | Knowledge score (10 questions): pre 2.7±1.7, post 5.8±2.3 (mean diff +3.0, 95%CI 2.7 to 3.3, p<0.001).<br><br>Proportions of respondents able to correctly identify the likelihood of benefiting from screening increased after decision aid viewing (pre 16.4% vs post 38.4%, p<0.001), as did the proportion who accurately estimated the likelihood of being overdiagnosed with screening (24.7% vs 46.1%, p<0.001).<br><br>No difference between demographic groups for both unadjusted and adjusted analyses.<br><br>Individuals indicating they were 'not likely' to pursue screening, had a greater improvement in pre–post knowledge score than those indicating that they were 'very likely' to pursue |

| Study, Country, Study ID<br>Study description<br>Recruitment setting<br>Sample size; Age; Female Gender; Insured; White (other race/ethnicities); Smoking history; other LC risk factors<br>Screening history<br>History of FP or lung surgery<br>Risk of bias (ROB) | Study data e.g. communicated outcomes of screening, attributes and their levels<br>Format of information and definitions provided<br>Judgement of Net Benefit<br>Data collection<br>Other information provided | Findings reported by authors (including relevant subgroup findings) & Data used in review analysis | Knowledge and understanding (including relevant subgroup findings) |
|---|---|---|---|
| | Costs: "While the initial screening test is covered by insurance plans, you may be responsible for copays and deductibles for follow up tests and procedures." | | screening (+3.5 vs +2.6, diff +0.9; 95% CI 0.1 to 1.8; p=0.023). |
| **Primary/secondary care population** | | | |
| Manners 2020, Australia<br>Study #: 6<br><br>Quasi-experimental pre-post LCS patient DA exposure for those eligible for screening (>1.5% 6-year cancer risk)<br><br>Eligibility and Recruitment: ever smokers 55-79 years via St John of God Midland Hospital general medical outpatient clinics<br><br>N=36 eligible for screening [>1.5% 6-year cancer risk] viewing DA); 55-79 (mean ± SD 67.7 ± 6.6); 42% Female; NR; NR; 78% former smokers, 59.1 ± 31.6 smoking pack-years; NR; 6-year cancer risk 3.1% (2.1%–4.4%)<br><br>Screening history: NR<br><br>History of FP or lung surgery: NR<br><br>ROB: Some risk | **Communicated Outcomes of Screening & Attribute levels (for those with 5% LC risk; others for 2%, 3% and 8%, over 3 screens unless noted):**<br>• LC mortality reduction by screening: 10 in 1000<br>• LC mortality (screened): 40 in 1000<br>• LC mortality (not screened): 50 in 1000<br>• FPs: over 50% in text; in icon arrays (numbers for first screen); 273 in 1000 have nodules that require another scan or biopsy with 10 cancers diagnosed<br>• FP biopsies: 9 in 1000 have biopsy without cancer diagnosed<br>• Over-diagnosis: 2 in 10 cancers diagnosed among 1000 after first screen<br>• Stress, anxiety and worry: Having a CT scan, waiting for the results or waiting several months for a follow up scan on a nodule can lead to stress and worry for some people.<br>• Incidental findings: If an abnormality is found on the CT scan outside the lungs, you may need to have more appointments or tests. In this case your GP will be notified, and they will follow up with these tests. Sometimes this is a benefit of lung cancer screening because other problems can be picked up earlier. Sometimes this is harmful because further tests show this was also a false alarm or the abnormality was over-diagnosed.<br><br>Format & definitions: The lung cancer screening recruitment pamphlet was developed as a tri-fold A4 pamphlet suitable for mass postal distribution. It contains brief information about lung cancer screening and a self-administered eligibility assessment, determined by PLCOm2012 lung cancer risk. Then, those eligible received a DA to review independently. Four variations of the PtDA | Screening preference, p=0.76:<br>Prefer: 78% pre, 72% post<br>Unsure: 19% pre, 22% post<br>Prefer not: 3% pre, 6% post<br><br>**Data used**:<br>Intentions:<br>72% (83% for sensitivity [assuming 50% of unsure actually prefer screening]) | Knowledge (% correct):<br>"Screening can reduce LC mortality": 53% pre, 89% post, p=0.001<br>"Screening lowers the chance of developing nodules" ("No"): 25% pre, 53% post, p<0.01<br>"How many current or former smokers will have nodules?": 47% pre, 81% post, p<0.001<br>"How many nodules will turn out to be lung cancer?": 47% pre, 81% post, p<0.001<br><br>No significant change in participants' understanding of relevant medical screening terms including over-diagnosis (~30% correct), false alarms, incidental findings, and false-negatives |

| Study, Country, Study ID<br>Study description<br>Recruitment setting<br>Sample size; Age; Female Gender; Insured; White (other race/ethnicities); Smoking history; other LC risk factors<br>Screening history<br>History of FP or lung surgery<br>Risk of bias (ROB) | Study data e.g. communicated outcomes of screening, attributes and their levels<br>Format of information and definitions provided<br>Judgement of Net Benefit<br>Data collection<br>Other information provided | Findings reported by authors (including relevant subgroup findings) & Data used in review analysis | Knowledge and understanding (including relevant subgroup findings) |
|---|---|---|---|
| | were developed corresponding to arbitrarily determined PLCOm2012 risk estimates of 2%, 3%, 5%, and 8%. Estimates of lung cancer mortality reduction were calculated using a 20% relative risk reduction of the individual's baseline PLCOm2012 risk and presented using opposing icon arrays.<br><br>The group ineligible for screening received an educational pamphlet to persuade them to not prefer screening; icon arrays showing no benefit from screening but no harms data.<br><br>Overdiagnosis: When doctors screen for cancers, some of the cancers will be slow growing. If those small slow growing cancers were not found, they would not cause that person any harm. Treatment is recommended for all cancers found by screening because doctors cannot tell which cancers are aggressive and likely to cause harm, and which ones grow slowly. It is better to treat all cancers than to potentially miss a cancer that would cause harm. However, this may result in unnecessary treatment and possible complications.<br><br>**Personalized risk and outcomes presented.**<br>**Independent review of material.**<br><br>**Judgement of Net benefit: High**<br><br>Data collection: Participants completed questionnaires immediately before or after their medical appointment. LCS eligible participants exposed to the DA and completed pre and post-pamphlet questionnaires<br><br>Other information:<br>Exposure to radiation; missed cancers (1 in 1000); financial costs; time commitment<br>Additional benefit of reassurance of no cancer. | | |

| Study, Country, Study ID<br>Study description<br>Recruitment setting<br>Sample size; Age; Female Gender; Insured; White (other race/ethnicities); Smoking history; other LC risk factors<br>Screening history<br>History of FP or lung surgery<br>Risk of bias (ROB) | Study data e.g. communicated outcomes of screening, attributes and their levels<br>Format of information and definitions provided<br>Judgement of Net Benefit<br>Data collection<br>Other information provided | Findings reported by authors (including relevant subgroup findings) & Data used in review analysis | Knowledge and understanding (including relevant subgroup findings) |
|---|---|---|---|
| Bittner-Fagan 2024, US<br>Study#: 7<br><br>Single-arm trial of online DA program with telephone SDM counsellor<br><br>Eligibility and Recruitment: six selected primary care practices using health system electronic medical records, screen eligible (USPSTF 2013), no LCS in past year and without limited life expectancy or prohibitive comorbid conditions<br><br>N= 64 (intervention group); mean 64.2 (SD 6.1); 50% Female; 97%; 79.6% White (17.2% Black; 1.6% Asian); current smokers 100%; 44.2 mean pack-year history<br><br>Screening history: NR (not within 12 months)<br><br>History of FP or lung surgery: NR<br><br>ROB: Low risk | **Communicated Outcomes of Screening & Attribute levels (annually for 3 years):**<br>• LC mortality without screening: 21 in 1000<br>• LC mortality reduction by screening: 3 in 1000 (18 still die of LC) (lowers risk by 20%)<br>• All-cause mortality without screening: 75 in 1000<br>• All-cause mortality with screening: 70 in 1000<br>• FPs: 365 in 1000<br>• FP biopsy: 18 in 1000<br>• Biopsy complication: <1 in 1000<br>• Overdiagnosis: 1 to 2 in 10 screen-detected cancers<br>• Incidental finding: descriptive<br><br>**Format & definitions:** Online DA (developed by Agency of Healthcare Research and Quality) reviewed via telephone counselling. Mentions that USPSTF recommends LCS. DA contains text and icon arrays for those screened and not screened. Patients interested in LCS were directly referred to screening program.<br>FPs: a person has a positive screening test but does not actually have lung cancer<br>Major complication: bleeding in the lung, a collapsed lung, or an infection<br>Overdiagnosis: Lung cancer screening may find a lung cancer that would not have ever caused symptoms or harmed the patient in his or her lifetime if the cancer had not been found. This could lead to treatment of people who do not really need treatment. At the time of diagnosis, there is no way for health care professionals to know if the lung cancer will cause health problems over a lifetime. For this reason, almost all people who are diagnosed with lung cancer are treated.<br>Incidental finding: Screening can find heart disease or thickened tissue in the lungs from scarring. Researchers do not know the possible benefits or harms of finding other things about your health through lung cancer screening.<br><br>**No personalized risk or outcomes presented.** | **Attitudes about screening (preference scores 0-1.0 with 0.51-1.00 “favor”):**<br>81%<br><br>**Screening completion in 1 year:**<br>45%<br><br>Multivariable regression: higher pack years and males had significantly (p value NR) more test ordering and completion<br><br>**Data used:**<br>45% screening completion | None |

| Study, Country, Study ID<br>Study description<br>Recruitment setting<br>Sample size; Age; Female Gender; Insured; White (other race/ethnicities); Smoking history; other LC risk factors<br>Screening history<br>History of FP or lung surgery<br>Risk of bias (ROB) | Study data e.g. communicated outcomes of screening, attributes and their levels<br>Format of information and definitions provided<br>Judgement of Net Benefit<br>Data collection<br>Other information provided | Findings reported by authors (including relevant subgroup findings) & Data used in review analysis | Knowledge and understanding (including relevant subgroup findings) |
|---|---|---|---|
| | **Review of material during SDM, via telephone.**<br><br>**Judgement of Net benefit: Moderate**<br><br>**Data collection:** Survey during telephone SDM**.**<br><br>**Other information:**<br>Radiation: Exposure to radiation increases a person's chance of developing cancer. LDCT screening for lung cancer exposes a person to radiation. If the screening test is positive, additional testing may involve higher doses of radiation. Level of radiation in comparison with other procedures eg about 30-40% average background radiation over 1 year | | |
| Lillie 2017, U.S.<br>Study #: 8<br><br>RCT with individuals participating in the Veterans Health Administration LCS Demonstration Project. Participants randomized in 2:1 ratio to 1) direct LCS invitation with DA, or 2) Usual care under PCP with annual smoking cessation clinical reminders.<br><br>Eligibility and Recruitment: screen-eligible (USPSTF 2013), members of the Minneapolis VA Health Care System<br><br>N=384 in intervention group; 55-80 yrs (mean ± SD: 65.1 ± 5.4); 4.9% Female; 100% insured (VHA patients); 90.6% White (7.3% African American, 0.3% Hispanic/Latino only, 1.3% Native | **Communicated Outcomes of Screening & Attribute levels (at 6.5 years):**<br>• LC mortality: 21 in 1000<br>• LC mortality reduction by screening: 3 in 1000 (18 still die of LC)<br>• FPs: 365 in 1000<br>• FP invasive procedures: 25 in 1000<br>• Major complications among FPs: 3 in 1000<br>• Overdiagnosis: Sometimes screening tests find cancers that would have never caused problems. This is called overdiagnosis. Unfortunately, it is often impossible to tell which cancers fall into this category. So, there is a very small chance someone may be treated unnecessarily for a cancer that would not have harmed them<br>• Stress/Anxiety: It is normal to feel stressed or anxious while waiting for your results or if you have something that is suspicious for lung cancer. Most patients with suspicious findings are reassured when they learn that most of these are false positives.<br><br>**Format & definitions:** the intervention group received a mailed LCS invitation that had a letter to call the LCS coordinator for a discussion about screening and to schedule an appointment with or | **Completion of screening (intervention group) at 3 months:**<br>22.7%<br><br>**Data used:**<br>Uptake: 22.7% | None |

| Study, Country, Study ID<br>Study description<br>Recruitment setting<br>Sample size; Age; Female Gender; Insured; White (other race/ethnicities); Smoking history; other LC risk factors<br>Screening history<br>History of FP or lung surgery<br>Risk of bias (ROB) | Study data e.g. communicated outcomes of screening, attributes and their levels<br>Format of information and definitions provided<br>Judgement of Net Benefit<br>Data collection<br>Other information provided | Findings reported by authors (including relevant subgroup findings) & Data used in review analysis | Knowledge and understanding (including relevant subgroup findings) |
|---|---|---|---|
| American/Alaskan Native); 44% current smoker and median (IQR) pack-year history: 48 (20); Spouse or family member with LC: 32.0%<br><br>Screening history: NR<br><br>History of FP or lung surgery: NR<br><br>ROB: High risk | without additional phone calls. They also received a paper copy of the VHA-developed DA. The control group only received annual smoking cessation clinical reminders. All patients had access to the comprehensive MVAHCS Tobacco Cessation Program.<br><br>FPs: "Screening for lung cancer by LDCT scan may find something that is suspicious but, after further testing, turns out not to be cancer. This is called a "false positive" … "Most false positive results are resolved with further LDCT testing."<br>FPs (invasive testing): "extra testing that involved putting a tube in the body or having surgery (these are called "invasive procedures")."<br>Major complications: "Complications can include bleeding, infections, or rarely, a collapsed lung."<br><br>**No personalized risk or outcomes presented.**<br>**Independent review of mailed material.**<br><br>**Judgement of Net benefit: Moderate**<br><br>**Data collection:** surveys were administered 3 months after randomization and receipt of the DA. A survey packet was mailed with a study invitation letter, a self-administered survey and a postage-paid return envelope. After 21 days non-respondents received a second survey packet. Intervention group only: LCS completion dates were taken from the VHA medical records. LCS completion was an LDCT completed before the survey return date.<br><br>Other information:<br>TPs: "26 people would get a true positive (they did have lung cancer)."<br>Radiation: "Exposure to radiation from LDCT scans increases your risk of cancer by a very small amount. We want to keep your chances of getting cancer from radiation very low. We do that by using "low-dose" CTs that use much less radiation than a standard CT." | | |

| Study, Country, Study ID<br>Study description<br>Recruitment setting<br>Sample size; Age; Female Gender; Insured; White (other race/ethnicities); Smoking history; other LC risk factors<br>Screening history<br>History of FP or lung surgery<br>Risk of bias (ROB) | Study data e.g. communicated outcomes of screening, attributes and their levels<br>Format of information and definitions provided<br>Judgement of Net Benefit<br>Data collection<br>Other information provided | Findings reported by authors (including relevant subgroup findings) & Data used in review analysis | Knowledge and understanding (including relevant subgroup findings) |
|---|---|---|---|
| | Cost: "If you are charged co-pays for your VA visits, you will be charged a $50.00 co-pay for the day you have the low-dose chest Computed Tomography scan (LDCT). Talk with the Lung Cancer Screening coordinator if you are charged co-pays. Scheduling the scan on the same day as another visit may decrease the total charges."<br>The decision aid also included information on what lung cancer is, the lung cancer screening process, and eligibility. Both groups had access to the MVAHCS Tobacco Cessation Program which included assessments of smoking status via a clinical reminder, patient education materials, individual and group behavioral therapy and pharmacotherapy. | | |
| Dharod 2019, U.S.<br>Study #: 9<br><br>Single-arm pragmatic study where individuals potentially eligible for LCS (and recent users of EHR patient portals) were invited to an interactive website assessing screening eligibility (USPSTF 2013) and included a DA.<br><br>Eligibility and Recruitment: 55-77 year-olds with record of any smoking from large academic health system including 4 hospitals and a network of over 70 community-based clinics in 19 North Carolina counties, houses large centralized LCS program.<br><br>N=81 with data NR (for 1000 enrolled: 55-77 (mean ± SD 65 ± NR); 51% Female; 45% commercial insurance, 51% | **Communicated Outcomes of Screening & Attribute levels (annually for 3 years):**<br>• LC mortality: 18 in 1000 (screened)<br>• LC mortality reduction by screening: 3 in 1000 (some given personalized benefits based on risk for cancer)<br>• FPs: 210 in 1000 (false alarm)<br>• Invasive procedures e.g., biopsy/surgery (from FP): 15 in 210<br>• Major complications (from FP + invasive procedure): less than 1 in 15<br><br>Format & definitions: An EHR algorithm identified potential candidates for LCS based on age and smoking history and used patient portal to send patients link to interactive website (mPATH-Lung (mobile Patient Technology for Health-Lung) with a DA used with those eligible for screening. Immediately after the video, the decision aid displayed the personalized risk-benefit screening information in an infographic, based on the $PLCO_{2012}$ and NLST results. 1000- person diagram in screened population in DA. Definitions NR only screen shot of 1000-diagram available.<br><br>**If participants answered questions to calculate 6-year risk for cancer, also showed personalized benefit-risk estimates. Independent review of material on website.** | **Screening intentions** (n=81 **of 99 eligible for screening**)<br>30% Yes<br>44% Unsure<br>26% No<br><br>Median (range) risk for developing lung cancer for participants who wanted screening was 4.2% (1.5–13.7%), for those who stated "Unsure" was 3.5% (0.4–21.5%), and for those who stated "No" was 2.1% (0.6–13.4%), p=0.09.<br><br>**Screening uptake over 4 months:**<br>10/81 (12.3%)<br><br>**Data used:**<br>Intentions: 30% (52% for sensitivity analysis [assuming 50% unsure actually prefer screening])<br>Uptake: 12.3% | None. |

| Study, Country, Study ID<br>Study description<br>Recruitment setting<br>Sample size; Age; Female Gender; Insured; White (other race/ethnicities); Smoking history; other LC risk factors<br>Screening history<br>History of FP or lung surgery<br>Risk of bias (ROB) | Study data e.g. communicated outcomes of screening, attributes and their levels<br>Format of information and definitions provided<br>Judgement of Net Benefit<br>Data collection<br>Other information provided | Findings reported by authors (including relevant subgroup findings) & Data used in review analysis | Knowledge and understanding (including relevant subgroup findings) |
|---|---|---|---|
| Medicare, 2% Medicaid, 2% uninsured; 84% non-Hispanic White (14% non-Hispanic Black, 1% Hispanic, 1% others); 19% current smokers, 81% former smokers; NR<br><br>Screening history: NR<br><br>History of FP or lung surgery: NR<br><br>ROB: Some risk (Intentions); high risk (Uptake) | **Judgement of Net benefit: Moderate**<br><br>Data collection: Within the online portal and after viewing the video, participants answered 8 demographic and medical history items. After being presented with personalized risk-benefits, participants were asked if they would like to receive screening. Screening completion within 4 months via records.<br><br>Other information: NR | | |
| Kinsinger 2017, U.S.<br>Study #: 10<br><br>Population-based screening demonstration project<br><br>Eligibility and Recruitment: patients screen-eligible (USPSTF 2013) and appropriate for screening visiting 8 academic medical sites supported by The Veterans Health Administration (VHA)<br><br>N= 4246; data on those who underwent screening: mean ± SD: 64.9 ± 5.1, 3.7% Female, NR; 72.2% White (14.8% Black, 93.8% Non-Hispanic or Latino, 1.3% Hispanic or Latino); 56.6% current smoker, ≥30 pack-years; 43.4% former smoker; NR | **Communicated Outcomes of Screening & Attribute levels (annually for three years with 6.5 year follow-up):**<br><br>• Mortality (Screened): 18 out of 1000<br>• Mortality (Not Screened): 21 out of 1000<br>• LC mortality reduction by screening: 3 fewer out of 1000<br>• FP: 365 in 1000<br>• Invasive procedure among FPs: 25 in 1000<br>• Major complication from invasive procedure: 3 out of 25<br><br>Format & definitions: SDM during scheduled clinic visits for most participants. Patient education materials were developed by NCP for use by providers and coordinators with SDM to help patients make decisions about whether they wanted to be screened (NR how many patients had SDM with primary care physician vs. program coordinator). The main brochure, Screening for Lung Cancer, was an SDM tool, which gave information about the benefits and harms of screening, and helped patients think through their values and preferences about screening. Included icon arrays (1000-people) for those screened and not screened. Data is from supplement figure; no access to full DA for definitions.<br><br>**No personalized risk or outcomes presented.**<br>**Review of material during SDM.** | **Screening intentions**:<br>Candidates eligible for LCS who agreed to undergo screening: 2452/4246 (57.7%)<br><br>**Screening uptake**:<br>Candidates eligible for LCS who underwent screening (within 3 months of data cut-off): 2106/4246 (49.6%)<br><br>**Data used:**<br>Intentions: 57.7%<br>Uptake: 49.6% | None |

| Study, Country, Study ID<br>Study description<br>Recruitment setting<br>Sample size; Age; Female Gender; Insured; White (other race/ethnicities); Smoking history; other LC risk factors<br>Screening history<br>History of FP or lung surgery<br>Risk of bias (ROB) | Study data e.g. communicated outcomes of screening, attributes and their levels<br>Format of information and definitions provided<br>Judgement of Net Benefit<br>Data collection<br>Other information provided | Findings reported by authors (including relevant subgroup findings) & Data used in review analysis | Knowledge and understanding (including relevant subgroup findings) |
|---|---|---|---|
| Screening history: 0% chest CT in previous year<br><br>History of FP or lung surgery: NR<br><br>ROB: Some risk (intentions); high risk (uptake) | **Judgement of Net benefit: Moderate**<br><br>Data collection: LCS program documented agreement to screening and completion within 3 months before data cut-off.<br><br>Other information: Data is from supplement figure; no access to full DA. | | |
| Schapira 2023, US<br>Study#: 11<br><br>Randomized trial among patients in VA centres comparing a web-based patient- and clinician-facing decision support tool with an attention control<br><br>Eligibility and Recruitment: screen eligible (USPSTF 2013; 55 -80 yrs) individuals who had not screened in past 15 mos and an upcoming appointment in primary care within 3 weeks, and a life expectancy of >2 years<br><br>N= 69; 55-80 (mean: 64); 12% Female; 100% VA patients; 38% Caucasian (61% Black, 1.4% Hispanic); current smokers 74%; mean pack-years 40.5<br><br>Screening history: NR (not in past year)<br><br>History of FP or lung surgery: NR | **Communicated Outcomes of Screening & Attribute levels (annually for 3 years):**<br>• LC mortality without screening: 21 in 1000<br>• LC mortality reduction by screening: 3 in 1000 (18 still die of LC) (lowers risk by 20%)<br>• FPs: 365 in 1000<br>• FP biopsy: 25 in 1000<br>• Biopsy complications: 3 in 1000<br>• Overdiagnosis: 1 in 5 cancers found by screening<br>• Incidental finding: 137 in 1000<br><br>**Format & definitions:** Web-based tool available day before clinic visit (98% logged on) and to clinician during visit. Opportunity to discuss DA in clinic visit but not per protocol and frequency NR. Text with definitions and numerics, and a pictogram with mortality, FPs and FP biopsies presented.<br>FPs: an abnormal finding on a CT scan (a nodule or spot) that with further testing turns out not to be cancer<br>Overdiagnosis: a lung cancer that is found by screening but is so slow growing that it would never have been found or caused you harm if you did not have the screening test... it can lead to cancer treatment that was not necessary (also called overtreatment).<br>Incidental finding: an abnormality that is not in the lung (such as heart disease or a thyroid nodule). These findings may then lead to additional tests. | **Screening completion at 9 months by EHR review**<br>31 (45%)<br><br>Black/African American: 36%<br><br>**Data used:**<br>Uptake 45% | 12-item LCS questionnaire<br><br>Mean knowledge scores:<br><br>Immediately after intervention: 7.0 [95% CI, 6.3-7.7] vs. 4.9 [95% CI, 4.3-5.5]; P < .001)<br>1 month: 6.3 [95% CI, 5.7-6.8] vs. 5.2 [95% CI, 4.5-5.8]; P = .03)<br>3 months: 6.2 [95% CI, 5.6-6.8] vs. 5.1 [95% CI, 4.4-5.8]; P = .01) |

| Study, Country, Study ID<br>Study description<br>Recruitment setting<br>Sample size; Age; Female Gender; Insured; White (other race/ethnicities); Smoking history; other LC risk factors<br>Screening history<br>History of FP or lung surgery<br>Risk of bias (ROB) | Study data e.g. communicated outcomes of screening, attributes and their levels<br>Format of information and definitions provided<br>Judgement of Net Benefit<br>Data collection<br>Other information provided | Findings reported by authors (including relevant subgroup findings) & Data used in review analysis | Knowledge and understanding (including relevant subgroup findings) |
|---|---|---|---|
| ROB: Some concerns | **No personalized risk or outcomes presented.**<br>**Mainly independent review of material.**<br><br>**Judgement of Net benefit: Low**<br><br>**Data collection:** Survey on decision making and knowledge. Screening records for 9 months.<br><br>**Other information:**<br>Radiation: 5 in 10,000 develop cancer from radiation | | |
| Reuland 2018, U.S.<br>Study #: 12<br><br>A pre-post study of a decision aid for LC screening in primary care.<br><br>Eligibility and Recruitment: eligible for screening (USPSTF 2013) at an academic primary care clinic<br><br>N=50; 55-80 yrs (mean 63 yrs); 48% Female; 92% Insured (Private, Medicare, Medicaid, or Medicare + other insurer); 58% White (30% Black, 12% Other); 46% current smoker and mean pack-years smoked: 52; NR<br><br>Screening history: 0% chest CT within 18 months (eligibility)<br><br>History of FP or lung surgery: NR<br><br>ROB: High risk | **Communicated Outcomes of Screening & Attribute levels (screening annually for 3 years; follow-up 7 years):**<br>• LC mortality: 21 in 1000<br>• LC mortality reduction by screening: 3 in 1000 (18 still die of LC)<br>• FPs: 365 in 1000<br>• FPs with biopsy: 25 in 1000<br>• Major complications: 3 in 1000<br>• Overdiagnosis: 4 in 1000<br>• Anxiety: "Screening can also cause anxiety from worrying about lung cancer, especially when the CT scan finds nodules."<br><br>Format & definitions: Six-minute video decision aid covering benefits and harms of screening with text, images and diagrams (icon arrays of screened and not screened): https://goo.gl/1f7XIY See Clark 2022 for definitions.<br><br>**No personalized risk or outcomes presented.**<br>**Independent review of material.**<br><br>**Judgement of Net benefit: Low**<br><br>Data collection: at the enrollment clinic visit a questionnaire was filled out, then individuals received the video DA, and afterwards completed the post-viewing survey. A scan of EHR records was done at 3 months follow-up for screening behavior. | **Screening intentions, "I plan to have a lung cancer screening test in the near future" (5-point Likert scale: strongly agree to strongly disagree):**<br>Pre-test: 27/50 (54%) agree or strongly agree<br>Post-test: 25/50 (50%) agree or strongly agree<br>(% "neutral/undecided" NR)<br><br>8/27 (30%) who preferred screening at baseline changed and did not prefer screening after viewing the decision aid.<br><br>6/23 (26%) who did not prefer screening at baseline changed and did prefer screening post-decision aid.<br><br>**Screening behavior at 3 months:**<br>36/50 (72.0%) participants had a clinic visit within 3 months post-enrollment. Overall, 13/36 (36%) had a test ordered, and 10/36 (28%) completed LDCT. (26% and 20% among whole sample) | Mean knowledge score (9 questions [7 related to overdiagnosis]) increased after viewing the decision aid from 2.6 to 5.5 (difference 2.8; 95% CI 2.1, 3.6, $p < 0.001$).<br><br>Before decision aid viewing, the most common response was that 401–700 individuals benefit per 1000 screened.<br>After decision aid viewing, the most common response category was 1–5 per 1000.<br><br>There was an observed inverse relationship between post-decision aid knowledge and screening preference (odds ratio = 0.73; 95% CI 0.54, 0.98; p = 0.03). For each point increase in post-decision |

| Study, Country, Study ID<br>Study description<br>Recruitment setting<br>Sample size; Age; Female Gender; Insured; White (other race/ethnicities); Smoking history; other LC risk factors<br>Screening history<br>History of FP or lung surgery<br>Risk of bias (ROB) | Study data e.g. communicated outcomes of screening, attributes and their levels<br>Format of information and definitions provided<br>Judgement of Net Benefit<br>Data collection<br>Other information provided | Findings reported by authors (including relevant subgroup findings) & Data used in review analysis | Knowledge and understanding (including relevant subgroup findings) |
|---|---|---|---|
| | Other information:<br>Radiation: "First, when a person gets a CT scan, he or she is exposed to radiation. Rarely, this can cause cancer 10 to 20 years later." <1 related cancer.<br>Costs: "Finally, while the initial screening test is covered by insurance plans, you may be responsible for copays and deductibles for follow up tests and procedures."<br>The video decision aid also included the rationale for screening, eligibility, and the LDCT procedure. | Of 18 who preferred screening, 8 (44%) had a test ordered and 6 (33%) completed LDCT.<br>Of 18 not preferring screening, 5 (28%) had a test ordered and 4 (22%) completed LDCT.<br><br>**Data used:**<br>Intentions: 50%<br>Uptake: 10/50 (20%) | aid knowledge score there was an estimated 27% reduction in the odds of preferring screening. |
| **Select population** | | | |
| Webster 2023, US<br>Study#: 13<br><br>Randomized trial comparing online and print DA among tobacco quitline callers<br><br>Eligibility and Recruitment: quitline callers eligible (USPSTF 2021) for LCS recruited via quitline providers with referral to study staff, no LCS in past year<br><br>N= 146 (Web group); 50-80 mean 61.6 (6.8); 64% Female; 40% White, 52% Black or African American, 1% Hispanic or Latino); 94% insured; 68% daily smokers; 64 mean pack-year history; 41% family history of lung cancer<br><br>Screening history: NR | **Communicated Outcomes of Screening & Attribute levels:**<br><br>**Should I Screen?**<br><br>• **Main text:**<br>• 20% reduction in lung cancer mortality<br>• 10-12% false positive rate<br>• **Benefits and harms based on personalized 6-year risk of cancer via modelling**:<br>• LC mortality reduction by screening (per 1000): 10% risk: 20 fewer; 6% risk: 11 fewer; 5% risk: 10 fewer; 4% risk; 8 fewer; 3% risk 6 fewer: 2% risk: 4 fewer; 1%: 2 fewer; 0.5%: 1 fewer (1000 arrays of screened vs. not screened)<br>• FPs: 120 in 1000<br>• FP invasive procedures: 13 in 1000<br>• Major complications: <1 in 1000<br>• Overdiagnosis: <1 in 10 cancers found by screening<br><br>Format & definitions: Web and print versions used in the study, however only data from web used in this analysis. Online DA includes a calculator consisting of 12 questions to calculate the risks of lung cancer. Print version used graphics to display risk based on sex, age and pack years, however does not provide enough numerical information on benefits and harms to merit | **Intention to undergo screening with a provider's recommendation:**<br>**Definitely** 75.2%<br>**Probably** 19.3%<br><br>**Screening uptake at 4 months:**<br>**Scheduled:** 4.7%<br>**Completed:** 11.4%<br><br>**Data used:**<br>Intentions 94.5%<br>Uptake: 11.4% | Knowledge assessments using 9 questions asking conceptual aspects.<br><br>One question attempting to indicate understanding of overdiagnosis: "It is sometimes hard for doctors to tell when an abnormality will grow quickly and needs to be treated vs. when it is unlikely to grow and can be left untreated"<br><br>Mean % correct answers at 1 and 4 months were 65-67% in both groups (similar at baseline) |

| Study, Country, Study ID<br>Study description<br>Recruitment setting<br>Sample size; Age; Female Gender; Insured; White (other race/ethnicities); Smoking history; other LC risk factors<br>Screening history<br>History of FP or lung surgery<br>Risk of bias (ROB) | Study data e.g. communicated outcomes of screening, attributes and their levels<br>Format of information and definitions provided<br>Judgement of Net Benefit<br>Data collection<br>Other information provided | Findings reported by authors (including relevant subgroup findings) & Data used in review analysis | Knowledge and understanding (including relevant subgroup findings) |
|---|---|---|---|
| History of FP or lung surgery: NR<br><br>ROB: High risk (intentions and uptake) | inclusion. Both modalities highlighted that an SDM visit with a provider is needed for a screening referral. List of screening sites was provided.<br><br>**Personalized risk and outcomes presented** (supplementary information reports few actually used the risk calculator in the web version, which provided the personalized risk information). **Individualized review of DA.**<br><br>**Judgement of Net benefit: High**<br><br>FPs: Screening works by finding lung nodules, some of which are cancer, but most nodules detected on the low-dose CT scan for lung cancer screening are not cancer. False positive findings can cause anxiety. However, understanding that most lung nodules found from the CT screening do not represent cancer helps most avoid significant anxiety while they wait for follow-up testing.<br>Overdiagnosis: These overdiagnosed cases of lung cancer are ones that may not have caused harm had they not been found, and a patient may not have felt any symptoms. However, by virtue of finding this kind of cancer, a patient may receive treatment that is unnecessary, with potential for complications. Note that once a cancer is found, it is not possible to know if it had been an overdiagnosis or not.<br>Invasive procedures: Sometimes further testing for a finding on lung cancer screening leads to the need for additional testing that can include invasive procedures (biopsies or surgery). These tests are risky and can cause harm in people that end up not having had cancer. Invasive procedures can result in minor complications such as bleeding and infections. They can also result in more major complications such as a collapsed lung and even death.<br><br>**Data collection:** Self-reported outcomes using surveys.<br><br>**Other information:** | | |

| Study, Country, Study ID<br>Study description<br>Recruitment setting<br>Sample size; Age; Female Gender; Insured; White (other race/ethnicities); Smoking history; other LC risk factors<br>Screening history<br>History of FP or lung surgery<br>Risk of bias (ROB) | Study data e.g. communicated outcomes of screening, attributes and their levels<br>Format of information and definitions provided<br>Judgement of Net Benefit<br>Data collection<br>Other information provided | Findings reported by authors (including relevant subgroup findings) & Data used in review analysis | Knowledge and understanding (including relevant subgroup findings) |
|---|---|---|---|
| | Radiation dose is equal to about 6 months of radiation from your every day environment. The additional risk of developing fatal lung cancer from one adult exam is considered to be extremely low | | |
| Tanner 2019, U.S.<br>Study #: 14<br><br>Prospective observational study with SDM and pre and post-surveys.<br><br>Eligibility and Recruitment: screen-eligible (USPSTF 2013) patients of Medical University of South Carolina<br><br>N=69; 55-80 (mean ± SD 64.1 ± 6.0); 52% Female; $0-$50 copay for Veterans; 64.2% non-Hispanic white (28.5% non-Hispanic Black, 5.1% Hispanic, 0 American Indian, 1.5% Other); NR; 23.2% family history of lung cancer; PLCO >2% risk 75.3%<br><br>Screening history: NR<br><br>History of FP or lung surgery: NR<br><br>ROB: Some risk | **Communicated Outcomes of Screening & Attribute levels:**<br>• **Benefits and harms based on personalized 6-year risk of cancer via modelling:**<br>• LC mortality reduction by screening: 10% risk: 20 fewer per 1000; 6% risk: 11 fewer; 5% risk: 10 fewer; 4% risk; 8 fewer; 3% risk 6 fewer: 2% risk: 4 fewer; 1%: 2 fewer; 0.5%: 1 fewer (1000 arrays of screened vs. not screened)<br>• FPs: 365 in 1000<br>• FP invasive procedures: 18 in 1000<br>• Major complications: 3 in 1000<br>• Overdiagnosed: 1 in 10 cancers found by screening<br><br>*about half of original study sample are not used for this review because of unclear information provided in a mailed DA.<br><br>Format & definitions: Participants underwent an in-person SDM by LCS providers. The visit started with explanatory counseling about the benefits and potential harms using the decision aid (shouldiscreen.com). Patients then received a personalized risk assessment for developing LC over the next 6 years using the PLCO 2012 2012 calculator. The in-person visit also included a web-based interactive decision aid (as described here).<br><br>FPs: Screening works by finding lung nodules, some of which are cancer, but most nodules detected on the low-dose CT scan for lung cancer screening are not cancer. False positive findings can cause anxiety. However, understanding that most lung nodules found from the CT screening do not represent cancer helps most avoid significant anxiety while they wait for follow-up testing. Overdiagnosis: There are times when screening will find a nodule that is cancer but was never going to cause problems. This can lead to treatments such as surgery, chemotherapy, and radiation which | **Screening completion:** 88.4%<br><br>**Data used**:<br>Uptake: 88.4% | None. |

| Study, Country, Study ID<br>Study description<br>Recruitment setting<br>Sample size; Age; Female Gender; Insured; White (other race/ethnicities); Smoking history; other LC risk factors<br>Screening history<br>History of FP or lung surgery<br>Risk of bias (ROB) | Study data e.g. communicated outcomes of screening, attributes and their levels<br>Format of information and definitions provided<br>Judgement of Net Benefit<br>Data collection<br>Other information provided | Findings reported by authors (including relevant subgroup findings) & Data used in review analysis | Knowledge and understanding (including relevant subgroup findings) |
|---|---|---|---|
| | are unnecessary and considered overtreatment. Overtreatment can cause side effects that make you feel sick.<br>Major complications: Sometimes further testing for a finding on lung cancer screening leads to the need for additional testing that can include invasive procedures (biopsies or surgery). These tests are risky and can cause harm in people that end up not having had cancer. Invasive procedures can result in minor complications such as bleeding and infections. They can also result in more major complications such as a collapsed lung and even death.<br><br>**Personalized risk for outcomes presented**.<br>**Review of material during SDM.**<br><br>**Judgement of Net benefit: High**<br><br>Data collection: Self-administered baseline survey and follow-up telephone survey 1 month after SDM visit. Screening uptake used medical records (follow-up duration NR)<br><br>Other information:<br>$100-$400 if uninsured<br>Radiation exposure (6 months of radiation from every day environment) | | |
| Djuric 2023, Italy<br>Study#: 15<br><br>Multi-center survey among people attending smoking cessation programs<br><br>Eligibility and Recruitment: participants seeking smoking cessation, without age or smoking eligibility criteria but data reported by eligibility | **Communicated Outcomes of Screening & Attribute levels (annually for 3 years):**<br>• LC mortality without screening: 21 in 1000<br>• LC mortality reduction by screening: 3 in 1000 (18 still die of LC) (lowers risk by 20%)<br>• All-cause mortality without screening: 75 in 1000<br>• All-cause mortality with screening: 70 in 1000<br>• FPs: 356 in 1000<br>• FP biopsy: 18 in 1000<br>• Biopsy complication: <1 in 1000<br>• Overdiagnosis: 1 to 2 in 10 screen-detected cancers<br>• Incidental finding: descriptive | **Attitudes about screening (acceptability of screening; quite/a lot in favor):**<br>90.2% | None |

| Study, Country, Study ID<br>Study description<br>Recruitment setting<br>Sample size; Age; Female Gender; Insured; White (other race/ethnicities); Smoking history; other LC risk factors<br>Screening history<br>History of FP or lung surgery<br>Risk of bias (ROB) | Study data e.g. communicated outcomes of screening, attributes and their levels<br>Format of information and definitions provided<br>Judgement of Net Benefit<br>Data collection<br>Other information provided | Findings reported by authors (including relevant subgroup findings) & Data used in review analysis | Knowledge and understanding (including relevant subgroup findings) |
|---|---|---|---|
| N= 144; median 58 (IQR 53-63); 54% Female; NR; NR; current smokers 100%; 79% ≥20 cigarettes per day; 47.8 median pack-year history<br><br>Screening history: NR (not offered in Italy)<br><br>History of FP or lung surgery: NR<br><br>ROB: High | **Format & definitions:** DA developed by Agency of Healthcare Research and Quality was translated. Mentions that USPSTF recommends LCS. Explained by educator during session. DA contains text and icon arrays for those screened and not screened.<br>FPs: a person has a positive screening test but does not actually have lung cancer<br>Major complication: bleeding in the lung, a collapsed lung, or an infection<br>Overdiagnosis: Lung cancer screening may find a lung cancer that would not have ever caused symptoms or harmed the patient in his or her lifetime if the cancer had not been found. This could lead to treatment of people who do not really need treatment. At the time of diagnosis, there is no way for health care professionals to know if the lung cancer will cause health problems over a lifetime. For this reason, almost all people who are diagnosed with lung cancer are treated.<br>Incidental finding: Screening can find heart disease or thickened tissue in the lungs from scarring. Researchers do not know the possible benefits or harms of finding other things about your health through lung cancer screening.<br><br>**No personalized risk or outcomes presented.**<br>**DA explained by educator.**<br><br>**Judgement of Net benefit: Moderate**<br><br>**Data collection:** Survey during later smoking cessation sessions.<br><br>**Other information:**<br>Radiation: Exposure to radiation increases a person’s chance of developing cancer. LDCT screening for lung cancer exposes a person to radiation. If the screening test is positive, additional testing may involve higher doses of radiation. Level of radiation in comparison with other procedures eg about 30-40% average background radiation over 1 year | | |

| **Study, Country, Study ID**<br>**Study description**<br>**Recruitment setting**<br>**Sample size; Age; Female Gender; Insured; White (other race/ethnicities); Smoking history; other LC risk factors**<br>**Screening history**<br>**History of FP or lung surgery**<br>**Risk of bias (ROB)** | **Study data e.g. communicated outcomes of screening, attributes and their levels**<br>**Format of information and definitions provided**<br>**Judgement of Net Benefit**<br>**Data collection**<br>**Other information provided** | **Findings reported by authors (including relevant subgroup findings) & Data used in review analysis** | **Knowledge and understanding (including relevant subgroup findings)** |
|---|---|---|---|
| Fraenkel 2016, U.S.<br>Study #: 16<br><br>RCT of three risk formats on LC screening, to evaluate the effects on screening-related knowledge, beliefs and screening endorsement.<br><br>Eligibility and Recruitment: outpatient pulmonary practices during reminder telephone calls (not for a pulmonary nodule or previous cancer).<br><br>N=254 (Numbers only=84; Numbers+Icon array=86; Numbers+slide presentation=83); age range NR (mean ± SD: 60.9 ± 8.8 yrs); 54.2% Female; NR; race NR; 56.3% current or past smoker of whom 28.7% had 30+ pack-year smoking history and 43.7% never smokers; NR<br><br>Screening history: NR<br><br>History of FP or lung surgery: NR<br><br>ROB: High risk | **Communicated Outcomes of Screening & Attribute levels:**<br>• LC mortality: 3 of 250 will die of LC despite getting screened<br>• LC mortality reduction by screening: 1 of 250 will have their life saved from LC by being screened and having cancer detected<br>• FPs: 91 of 250<br><br>Format & definitions: patients were randomized to receive risk information with 1) numbers only, 2) numbers and corresponding icon arrays, or 3) numbers and experienced format (a set of slides illustrating LDCT scans with normal, FP and LC results). The risk information took 4 minutes total to view, at 1 slide per second.<br>FPs: "A false alarm means that the CT scan shows a small spot on the lung that after doing more tests turns out NOT to be cancer. It may take a biopsy or up to two years of repeat CT scans to be sure that a spot on the lung is not cancer"<br><br>**No personalized risk or outcomes presented. Independent review of material.**<br><br>**Judgement of Net benefit: Moderate (all groups)**<br><br>Data collection: patients viewed the risk information in 1 of the 3 formats they were randomized to on a computer in a quiet room while a medical assistant directly observed them. Participants were then asked to complete outcome questions.<br><br>Other information:<br>TN: 155 of 250 will have a normal CT scan and told they do not have lung cancer.<br>The materials also described the process of LC screening. | **Intentions (preferences for screening):**<br>What do you think right now about having a CT scan to screen for lung cancer each year? (for never smokers "Imagine that you were a heavy smoker"; 11-point numeric scale anchored by "I am certain that I don't want to be screened" and "I am certain that I do want to be screened" with unsure if the middle, mean (SE):<br>Numbers only group: 7.9 (0.3)<br>Numbers and icon array group: 7.6 (0.3)<br>Numbers and experience group: 8.1 (0.3)<br><br>**Data used**:<br>Intentions:<br>Average of means = 7.9 ("51-74% intend to screen") | Knowledge about FPs:<br><br>The percent of participants responding correctly to "Compared to the number of people with lung cancers found by screening, how many people will have a false alarm?" 56.3% (higher for those randomized to the numbers + icon array and numbers + experienced format compared to the numbers only format [39.3%]).<br><br>The percent of participants responding correctly to the "Of all the people screened, about how many will have a false alarm?" 38.6% (higher only for those randomized to the numbers + icon array format [52.3%] compared to the numbers only format [31%]).<br><br>Objective numeracy did not impact findings.<br><br>Perceived chance of developing LC (11-point |

| Study, Country, Study ID<br>Study description<br>Recruitment setting<br>Sample size; Age; Female Gender; Insured; White (other race/ethnicities); Smoking history; other LC risk factors<br>Screening history<br>History of FP or lung surgery<br>Risk of bias (ROB) | Study data e.g. communicated outcomes of screening, attributes and their levels<br>Format of information and definitions provided<br>Judgement of Net Benefit<br>Data collection<br>Other information provided | Findings reported by authors (including relevant subgroup findings) & Data used in review analysis | Knowledge and understanding (including relevant subgroup findings) |
|---|---|---|---|
| | | | scale [not described]; mean [SD]):<br>Numbers only group: 5.0 (2.8)<br>Numbers and icon array group: 4.9 (2.9)<br>Numbers and experience group: 5.1 (2.9) |
| Hoffman 2018, U.S.<br>Study #: 17<br><br>A pre-post study of online DA<br><br>Eligibility and Recruitment: 55-80 yrs, current or former smokers who quit within the past 15 yrs (no pack-year minimum), members of the Tobacco Treatment Program at University of Texas Cancer Center, seeking smoking cessation services<br><br>N=30; 55-80 yrs (mean: 61.5 yrs); 50% Female; 100% insured (Medicare, Medicaid, job/private insurance, or other); 63.3% White (30.0% Black/African American, 3.3% Hispanic/Latino, 3.3% other); 60.7% current smoker and mean (SD) pack-year history: 30.4 (18.9); NR<br><br>Screening history: NR<br><br>History of FP or lung surgery: NR | **Communicated Outcomes of Screening & Attribute levels (annually for 3 years; follow-up over 6.5 years):**<br>• LC mortality: 21 in 1000<br>• LC mortality reduction by screening: 3 in 1000 (18 still die of LC)<br>• FPs: 356 in 1000 (additional scan)<br>• FPs: 18 in 1000 (test or surgery)<br>• Major complications: 3 in 1000<br>• Overdiagnosis: "presented as cancers that would never become life threatening, and the video explains that some people may be treated for cancer that would never have harmed them."<br><br>Format & definitions: Video decision aid "Lung Cancer screening: Is it Right for Me?", covering benefits and harms of screening with text, images and diagrams (icon arrays of screened and not screened). **USPSTF 2013 "recommends"**. Emphasizes importance of considering this personal decision with their doctor.<br><br>**No personalized risk or outcomes presented.**<br>**Independent review of material.**<br><br>**Judgement of Net benefit: Moderate**<br><br>Data collection: patients completed an online baseline questionnaire including items on knowledge of LCS with LDCT. Patients then viewed a video DA, and completed a post-decision aid questionnaire. | **Intentions:** Interest in screening in the next year: 63% definitely/probably (% unsure NR)<br>(only 12 [40%] eligible for screening, and participants were aware of eligibility criteria)<br><br>**Data used:**<br>Intentions: 63% | Mean knowledge scores (12-item measure): pre, 5.7 (47.3% correct), post 9.6 (80.3% correct); 3.9 mean difference (SD = 2.9, 95% CI 2.9–5.0; p<0.001). |

| Study, Country, Study ID<br>Study description<br>Recruitment setting<br>Sample size; Age; Female Gender; Insured; White (other race/ethnicities); Smoking history; other LC risk factors<br>Screening history<br>History of FP or lung surgery<br>Risk of bias (ROB) | Study data e.g. communicated outcomes of screening, attributes and their levels<br>Format of information and definitions provided<br>Judgement of Net Benefit<br>Data collection<br>Other information provided | Findings reported by authors (including relevant subgroup findings) & Data used in review analysis | Knowledge and understanding (including relevant subgroup findings) |
|---|---|---|---|
| ROB: High risk | Other information:<br>TPs: 24 in 1000 individuals will have LC<br>Radiation exposure: shown pictorially with other radiation exposures to contextualize: from a flight from NY to LA (0.03 mSv) as lowest exposure to highest for living in the US (3.00 mSv). LDCT scan was presented as 1.50 mSv.<br>The video also describes the natural history of smoking and LC (emphasis of importance of quitting smoking), the **USPSTF screening recommendation**, the importance of an informed decision, and what the procedure of screening is. | | |
| Volk 2020, U.S.<br>Study #: 18<br><br>Randomized controlled trial of a patient DA versus standard education materials (SEM)<br><br>Eligibility and Recruitment: clients aged 55-77 years who reported a 30-plus pack-year smoking history via 13 state tobacco quit lines and having an interest in screening<br><br>N=237 data in intervention group (232 in CG); 73.4% <65 yrs; 60.6% Female; NR; 71.4% White (0.8% American Indian or Alaska Native, 23.9% Black, 0% Native Hawaiian or other Pacific Islander, 0.4% more than 1 category, 0.8% other); median years smoked (IQR) 43.0 (40.0-50.0), median cigarettes smoked per day (IQR) 20.0 (20.0-30.0), median pack- | **Communicated Outcomes of Screening & Attribute levels:**<br>• LC mortality without screening: 21 in 1000<br>• LC mortality reduction by screening: 3 fewer in 1000 (18 still die of LC)<br>• FPs: 356 in 1000<br>• FPs invasive procedures: 18 in 1000 (test or surgery)<br>• Major complications: 3 in 1000<br>• Overdiagnosis: "presented as cancers that would never become life threatening, and the video explains that some people may be treated for cancer that would never have harmed them."<br><br>Format & definitions: PDA arm: 9.5-minute narrated video, Lung Cancer Screening: Is It Right for Me? The narrated DA included information about (1) eligibility for lung cancer screening and a calculation of tobacco pack-year smoking history, (2) lung cancer epidemiology and risk factors, (3) a video of a patient in a CT scanner, (4) icon arrays to graphically depict the magnitude of mortality reduction, false-positive results, and harms from invasive diagnostic procedures, and (5) radiation exposure depicted within the context of other sources of radiation (e.g., a screening mammogram). Smoking cessation was emphasized throughout the DA. The DA was updated according to USPSTF 2014 recommendations. "**USPSTF "recommends"** | **Intentions** (intent to be screening within 1 yr, at 1-week assessment):<br>Intervention (n=233): 70.8%<br><br>**Screening behavior:**<br>Screened for LC by 6 month follow up:<br>Intervention (n=237): 24.1%<br><br>**Data used:**<br>Intentions: 70.8%<br>Uptake: 24.1% | Correct responses to LCS knowledge, correct response, mean % (95% CI)<br><br>1 week<br>Intervention (n=235): 57.5% (54.7-60.3)<br>Control (n=233): 40.1% (37.9-42.3)<br>Difference: 17.4% (13.9-21.0), p<0.001<br><br>6 months<br>Intervention (n=218): 49.9% (47.5-52.3)<br>Control (n=225): 40.0% (37.6-42.4)<br>Difference: 9.9% (6.5-13,3), p<0.001<br><br>Tests were adjusted for age, sex, race/ethnicity, educational level, insurance status, quitline |

| Study, Country, Study ID<br>Study description<br>Recruitment setting<br>Sample size; Age; Female Gender; Insured; White (other race/ethnicities); Smoking history; other LC risk factors<br>Screening history<br>History of FP or lung surgery<br>Risk of bias (ROB) | Study data e.g. communicated outcomes of screening, attributes and their levels<br>Format of information and definitions provided<br>Judgement of Net Benefit<br>Data collection<br>Other information provided | Findings reported by authors (including relevant subgroup findings) & Data used in review analysis | Knowledge and understanding (including relevant subgroup findings) |
|---|---|---|---|
| year history (IQR) 48.0 (40.0-63.0); NR<br><br>Screening history: NR<br><br>History of FP or lung surgery: NR<br><br>ROB: Some risk | SEM arm: The SEM material was a 2-page brochure from a lung cancer advocacy group and included structured questions a patient can ask a physician about lung cancer screening. Benefits and harms were described but no probabilities of outcomes were included. (not eligible for this review)<br><br>**No personalized risk or outcomes presented. Independent review of material.**<br><br>**Judgement of Net benefit: Moderate**<br><br>Data collection: Baseline, 1-week, 3-month, and 6-month questionnaires via telephone or mailed if not reachable by phone.<br><br>Other information:<br>TPs: 24 in 1000 individuals will have LC<br>Radiation exposure: shown pictorially with other radiation exposures to contextualize: from a flight from NY to LA (0.03 mSv) as lowest exposure to highest for living in the US (3.00 mSv). LDCT scan was presented as 1.50 mSv.<br>The video also describes the natural history of smoking and LC (emphasis of importance of quitting smoking), the **USPSTF screening recommendation**, the importance of an informed decision, and what the procedure of screening is. | | service provider, and recruitment method |
| Kathuria 2022, U.S.<br>Study #: 19a ,19b<br><br>Two pilot RCTs using a DA with or without SDM<br><br>Eligibility and Recruitment: screen-eligible (USPSTF 2013), current smokers, and hospitalized at largest safety-net hospital in New England, Boston Medical Center, | **Communicated Outcomes of Screening & Attribute levels (annually for 3 years):**<br>• LC mortality: 21 in 1000<br>• LC mortality reduction by screening: 3 in 1000 (18 still die of LC)<br>• All-cause mortality: 75 in 1000<br>• All-cause mortality reduction by screening: 5 in 1000 (70 still die of any cause)<br>• FPs: 356 in 1000<br>• FPs with biopsy: 18 in 1000 (biopsy/invasive procedure)<br>• Major complication: 1 in 1000<br>• Overdiagnosis: 1-2 of 10 diagnosed with LC | **Screening uptake at 3 months:**<br>**Pilot 1**<br>DA alone:<br>1/48 (2%) completed LCS<br>DA with SDM:<br>2/52 (4%) completed LCS<br><br>No difference between study arms (p=0.6). | None. |

| Study, Country, Study ID<br>Study description<br>Recruitment setting<br>Sample size; Age; Female Gender; Insured; White (other race/ethnicities); Smoking history; other LC risk factors<br>Screening history<br>History of FP or lung surgery<br>Risk of bias (ROB) | Study data e.g. communicated outcomes of screening, attributes and their levels<br>Format of information and definitions provided<br>Judgement of Net Benefit<br>Data collection<br>Other information provided | Findings reported by authors (including relevant subgroup findings) & Data used in review analysis | Knowledge and understanding (including relevant subgroup findings) |
|---|---|---|---|
| and referred to inpatient Tobacco Treatment Consult service.<br><br>RCT Pilot 1: N=100 (I=52; C=48); 55-80 yrs (mean ± SD, I: 62.1 ± 5.0; mean ± SD, C: 61.6 ± 5.8); 35% female; 86.0% Medicaid-insured; 30.0% White (60.0% Black, 2.0% multi-race, 8.0% other/unknown, 5.0% Hispanic or Latino ethnicity); 100% current smokers; mean ± SD yrs smoked, I: 42.2 ± 9.8; mean ± SD yrs smoked, C: 44.4 ± 9.4; NR<br><br>RCT Pilot 2: N=21 (I=10; C=11); 55-80 yrs (mean ± SD, I: 64.0 ± 6.1; mean ± SD, C: 64.0 ± 5.62); 29% female; 85.7% Medicaid-insured; 23.8% White (66.7% Black, 9.5% other/unknown, 9.5% Hispanic or Latino ethnicity); 100% current smokers; mean ± SD yrs smoked, I: 46.4 ± 9.7; mean ± SD yrs smoked, C: 44.0 ± 8.9; NR<br><br>Screening history: 0% chest CT in the past year (exclusion criteria)<br><br>History of FP or lung surgery: NR<br><br>ROB: High risk | • Incidental findings: Screening can find heart disease or thickened tissue in the lungs from scarring.<br><br>Format & definitions: RCT pilot 1: a nurse practitioner initiated inpatient SDM for LCS with a DA that they reviewed together with the participant and notification to PCPs to refer their patient (intervention group), or enhanced usual care with a DA given to the patients to review on their own (control group). RCT pilot 2: same intervention as RCT pilot 1, but with an added community health worker (CHW) to address barriers to LCS completion, and a nurse practitioner ordering the LCS CT. **USPSTF 2013 "recommends"**<br><br>Overdiagnosis: "Lung cancer screening may find a lung cancer that would not have ever caused symptoms or harmed the patient in his or her lifetime if the cancer had not been found. This could lead to treatment of people who do not really need treatment. At the time of diagnosis, there is no way for health care professionals to know if the lung cancer will cause health problems over a lifetime. For this reason, almost all people who are diagnosed with lung cancer are treated."<br>Incidental findings: "Researchers do not know the possible benefits or harms of finding other things about your health through lung cancer screening."<br><br>**No personalized risk or outcomes presented.**<br>**Review of material independently (n=59) or during SDM (n=62).**<br><br>**Judgement of Net benefit: Low (all groups)**<br><br>Data collection: LCS orders and completion at 3 months was measured by chart review.<br><br>Other information:<br>Radiation: "Exposure to radiation increases a person's chance of developing cancer. LDCT screening for lung cancer exposes a person to radiation. If the screening test is positive, additional testing may involve higher doses of radiation. Researchers | Primary care providers noted that competing priorities and patient factors (e.g., social barriers to keeping appointments) prevented the intervention from working as intended.<br><br>**Pilot 2**<br>DA alone:<br>1/11 (9%) completed LCS<br>DA with SDM and CHW-nav<br>5/10 (50%) completed LCS<br><br>LCS completion was higher in the intervention arm ($p<0.05$).<br><br>**Data used:**<br>Uptake:<br>Completed LCS among the DA and DA with SDM (n=111) = 3.6% (19a)<br>Completed LCS SDM + CHW intervention (n=10) = 50% (19b) | |

| Study, Country, Study ID<br>Study description<br>Recruitment setting<br>Sample size; Age; Female Gender; Insured; White (other race/ethnicities); Smoking history; other LC risk factors<br>Screening history<br>History of FP or lung surgery<br>Risk of bias (ROB) | Study data e.g. communicated outcomes of screening, attributes and their levels<br>Format of information and definitions provided<br>Judgement of Net Benefit<br>Data collection<br>Other information provided | Findings reported by authors (including relevant subgroup findings) & Data used in review analysis | Knowledge and understanding (including relevant subgroup findings) |
|---|---|---|---|
| | do not know how being exposed to radiation from LDCT scans and additional diagnostic imaging tests may affect people". A diagram of different levels of radiation.<br>The decision aid also reports in what LC is, the LDCT screening process, signs & symptoms of LC, eligibility, and smoking cessation information. | | |
| **Attending screening program** | | | |
| Mazzone 2017, U.S.<br>Study#: 20<br><br>Pre/post study of a counselling and SDM visit for LCS<br><br>Eligibility and Recruitment: eligible patients referred to LCS program<br><br>N=411 eligible (n=125 answering other questions; 55-77 (mean ± SD 64.4 ± NR); 33.9% Female; NR; NR; 45.2% active smokers, mean pack-years smoked 53.0 (range 30-112); 40.3% COPD<br><br>Screening history: NR<br><br>History of FP or lung surgery: NR<br><br>ROB: High risk | **Communicated Outcomes of Screening & Attribute levels:**<br>• **Benefits and harms based on personalized 6-year risk of cancer via modelling:**<br>• LC mortality reduction by screening (per 1000): 10% risk: 20 fewer; 6% risk: 11 fewer; 5% risk: 10 fewer; 4% risk; 8 fewer; 3% risk 6 fewer: 2% risk: 4 fewer; 1%: 2 fewer; 0.5%: 1 fewer (1000 arrays of screened vs. not screened)<br>• FPs: 365 in 1000<br>• FP invasive procedures: 18 in 1000<br>• Major complications: 3 in 1000<br>• Overdiagnosed: 1 in 10 cancers found by screening<br><br>*Contents of video NR<br><br>Format & definitions: All eligible patients were scheduled for their counseling and SDM visit, with a LDCT examination coordinated after the visit. The counseling and SDM visits were usually staffed by a nurse practitioner and less often by a physician assistant or physician. The counseling and SDM visit began with a review of patient eligibility criteria. This was followed by the presentation of an approximately 6-min narrated video slide show (developed for our program) describing the benefits and harms of lung cancer screening, the use of a decision aid (http://www.shouldiscreen.com) to individualize the discussion of benefits and risks, and data collection. Patients had an opportunity to ask questions throughout the visit, including a prompt for questions from the provider at the end of the video slide show.<br>FPs: Screening works by finding lung nodules, some of which are cancer, but most nodules detected on the low-dose CT scan for lung cancer screening are not cancer. False positive findings can | **Screening uptake**:400 /411 eligible (97%)<br><br>**Data used:**<br>Uptake: 97% | N=125<br>64% correct about benefits of early detection (but 8% correct that LCS reduced the chance of lung cancer mortality)<br><br>95% could identify one harm |

| Study, Country, Study ID<br>Study description<br>Recruitment setting<br>Sample size; Age; Female Gender; Insured; White (other race/ethnicities); Smoking history; other LC risk factors<br>Screening history<br>History of FP or lung surgery<br>Risk of bias (ROB) | Study data e.g. communicated outcomes of screening, attributes and their levels<br>Format of information and definitions provided<br>Judgement of Net Benefit<br>Data collection<br>Other information provided | Findings reported by authors (including relevant subgroup findings) & Data used in review analysis | Knowledge and understanding (including relevant subgroup findings) |
|---|---|---|---|
| | cause anxiety. However, understanding that most lung nodules found from the CT screening do not represent cancer helps most avoid significant anxiety while they wait for follow-up testing.<br>Overdiagnosis: There are times when screening will find a nodule that is cancer but was never going to cause problems. This can lead to treatments such as surgery, chemotherapy, and radiation which are unnecessary and considered overtreatment. Overtreatment can cause side effects that make you feel sick.<br>Major complications: Sometimes further testing for a finding on lung cancer screening leads to the need for additional testing that can include invasive procedures (biopsies or surgery). These tests are risky and can cause harm in people that end up not having had cancer. Invasive procedures can result in minor complications such as bleeding and infections. They can also result in more major complications such as a collapsed lung and even death.<br><br>**Personalized risk and outcomes presented**.<br>**Review of material during SDM.**<br><br>**Judgement of Net benefit: High**<br><br>Data collection: Participants completed a pre-visit survey, immediate post-visit survey, and one-month post-visit survey<br><br>Other information: $100-$400 if uninsured<br>Overtreatments such as surgery, chemotherapy, radiation<br>Follow up scans at 6 to 12-month intervals<br>Radiation exposure (6 months of radiation from every day environment) | | |
| Quaife 2020, U.K.<br>Study #: 21<br><br>A two-arm, blinded RCT that tested the effect of intervention invitation materials on uptake of a prescheduled Lung Health Check | **Communicated Outcomes of Screening & Attribute levels:**<br>• LC mortality reduction CT **vs. chest X-ray**: 20% (as per associated study Ruparel 2019, 120 people received film stating 320 needed to screen to prevent 1 LC death)<br>• FPs: 23 in 100<br>• Incidental findings: “This means there are signs of other problems on the scan that may need treatment or medical advice. If you already have a lung problem, this might be | **Screening uptake (of those eligible for LDCT via NLST criteria or risk tools e.g. PLCO, n=845):**<br>Overall: 91.2%<br>Intervention (n=416): 92.8%<br>Control (n=429): 89.7% | No |

| Study, Country, Study ID<br>Study description<br>Recruitment setting<br>Sample size; Age; Female Gender; Insured; White (other race/ethnicities); Smoking history; other LC risk factors<br>Screening history<br>History of FP or lung surgery<br>Risk of bias (ROB) | Study data e.g. communicated outcomes of screening, attributes and their levels<br>Format of information and definitions provided<br>Judgement of Net Benefit<br>Data collection<br>Other information provided | Findings reported by authors (including relevant subgroup findings) & Data used in review analysis | Knowledge and understanding (including relevant subgroup findings) |
|---|---|---|---|
| appointment at which LDCT screening might be offered. Data among those attending Health Checks.<br><br>Eligibility and Recruitment: 60-75 yrs, recorded as smokers since 2010 – within 7 yrs of invitation and without CT in past year from 16 primary care practices in London<br><br>N=2,012 randomized (I=1,006; C=1,006); mean ± SD: 66.0 ± 4.3 years; 46.3% Female; NR; 79.7% White (9.6% Black, 2.1% Asian, 1.7% Mixed, 2.9% Other; 4.0% Not stated); 74.5% current smoker (0.6% never smoked tobacco, 0.1% refused/not stated); NR<br>***N=1,005 attending Health Checks and consenting to study (I=494; C=511)**<br><br>Screening history: 0% past year thoracic CT scans<br><br>History of FP or lung surgery: NR<br><br>ROB: High risk | why and you may not need any extra care. You may be advised to make contact with your GP to make an appointment to find out more.”<br>• Overdiagnosis: “In some cases, people will be diagnosed and treated for lung cancer that would never have caused the person harm. If they had not been screened, they would never have known about the cancer or have had any treatment.”<br>• Worry: “Waiting for the results of these tests can be worrying. People with an unclear result will need to be monitored and have a further scan. This can be a worrying time and in most cases they will not have lung cancer.”<br><br>Format & definitions: individuals were invited to pre-scheduled Lung Health Check (LHC) appointments, with randomization to the intervention group that received “M.O.T. For Your Lungs” leaflet (no benefits or harms described, targets psychological barriers to attendance) and controls that received an information booklet mimicking the facts booklet of NHS cancer screening programs (includes benefits and harms of screening). Both groups received a **pre-invitation letter from their PCP**, an invitation letter with scheduled appointment, and a reminder letter with a second scheduled appointment. Letters were identical except 1) the intervention group referred to “ever smokers” and the control referred to “current and former smokers”, and 2) the intervention group’s invitation letter included a bullet-pointed summary of the LHC, including LDCT scan offer, on the reverse side. **Full information via the control groups leaflet was given to the intervention group at the appointment**. **The LHC had a nurse for discussions.**<br>FPs & worry: “Waiting for the results of these tests can be worrying. People with an unclear result will need to be monitored and have a further scan. This can be a worrying time and in most cases they will not have lung cancer.” | **Subgroups:** Sex and age were not associated with LDCT uptake. For ethnicity, Asian ethnicity predicted lower odds of uptake compared with white ethnicity (53.8% vs. 91.3%; aOR, 0.09; 95% CI, 0.02–0.31), but there were few Asian participants (n = 13). There was no association with black ethnicity (92.7%) and too few noncases within the other ethnic groups. Deprivation was not associated with LDCT uptake. In unadjusted analyses, current smokers were less likely to opt for the LDCT scan than former smokers (90% vs 94.6%), but the association was not statistically significant in adjusted analyses (aOR, 0.52; 95% CI, 0.27–1.01).<br><br>**Willingness to screen of those *ineligible* for screening (n=160):**<br>Intervention (n=78):<br>Yes, definitely: 71.8%<br>Yes, probably: 10.3%<br>Probably not: 1.3%<br>Definitely not: 5.1%<br>Missing: 11.5%<br><br>Control (n=82):<br>Yes, definitely: 62.2%<br>Yes, probably: 19.5%<br>Probably not: 6.1%<br>Definitely not: 2.4%<br>Missing: 9.8%<br><br>**Data used:** | |

| Study, Country, Study ID<br>Study description<br>Recruitment setting<br>Sample size; Age; Female Gender; Insured; White (other race/ethnicities); Smoking history; other LC risk factors<br>Screening history<br>History of FP or lung surgery<br>Risk of bias (ROB) | Study data e.g. communicated outcomes of screening, attributes and their levels<br>Format of information and definitions provided<br>Judgement of Net Benefit<br>Data collection<br>Other information provided | Findings reported by authors (including relevant subgroup findings) & Data used in review analysis | Knowledge and understanding (including relevant subgroup findings) |
|---|---|---|---|
| | **Personalized risk likely presented due to screening eligibility criteria using risk prediction tool.**<br>**Review during discussions.**<br><br>**Judgement of Net benefit: High (both groups)**<br><br>Data collection: participants were randomized to the intervention and control arms of the Lung Health Check and of those who attended (n=1,058), those who consent to take part in the LSUT (n=1,005) were assessed for their willingness to scan if ineligible, and their uptake of screening for those eligible.<br><br>Other information:<br>TPs: 2 in 100<br>FN: "Like all cancer screening tests, lung cancer screening is not completely accurate and some cancers will be missed. Nodules found in the middle of the chest and some small cancers are harder to see. Some cancers start to grow after screening."<br>Radiation: "The low dose CT scan will expose you to a small amount of radiation. It is the same as about one year's worth of radiation from the natural environment. The risk of a CT scan causing a cancer is very low compared with the benefits of detecting lung cancer early. If a further CT scan is needed then this will expose you to more radiation." | Uptake: 91.2% | |
| Daskalakis 2023, U.S.<br>Study #: 22<br><br>Case series with use of a standardized decision counseling protocol with patient education handout during visit to LCS program.<br><br>Eligibility and Recruitment: invited all patients referred to centralized | **Communicated Outcomes of Screening & Attribute levels:**<br>• LC mortality: 21 in 1000 (not screened)<br>• LC mortality: 18 in 1000 (screened); 300 people need to be screened to save one life (in text benefits for higher risk [6 fewer/160 needed to screen] and lower risk [0.2 fewer/5000 needed to screen] are presented<br>• FPs: 365 in 1000<br>• Invasive procedures e.g., biopsy/surgery (from FPs): 25 in 365<br>• Major complications (from FP + invasive procedure: 3 in 25<br>• Overdiagnosis: 1 in 5 cancers detected by screening (in text; not icon arrays) | **Screening uptake**:<br>89% screened (73% on same day; 20% within 30 days)<br><br>Subgroups: 93% among White participants, 84% among African American participants, and 90% among Asian participants. The difference between African American and White participants remained after adjustment for participant characteristics (OR 0.41; 95% CI, 0.21–0.81; P = 0.009) | None. |

| Study, Country, Study ID<br>Study description<br>Recruitment setting<br>Sample size; Age; Female Gender; Insured; White (other race/ethnicities); Smoking history; other LC risk factors<br>Screening history<br>History of FP or lung surgery<br>Risk of bias (ROB) | Study data e.g. communicated outcomes of screening, attributes and their levels<br>Format of information and definitions provided<br>Judgement of Net Benefit<br>Data collection<br>Other information provided | Findings reported by authors (including relevant subgroup findings) & Data used in review analysis | Knowledge and understanding (including relevant subgroup findings) |
|---|---|---|---|
| LCS program in a large urban health system<br><br>N=419; 55-78 yrs (mean ± SD NR); 54% female; NR; 53% White/Caucasian (45% Black/African American, 2% Asian); 63% current smoker; 30% COPD, 18% family history of LC<br><br>Screening history: NR<br><br>History of FP or lung surgery: NR<br><br>ROB: High risk | • Stress and anxious: It is normal to have increased stress after these findings (FPs). It can be very stressful waiting for several months to see if your nodule has grown.<br><br>Format & definitions: 20-page DA with icon arrays for most outcomes (not overdiagnosis) and text with numerical data for all outcomes. Text for data on those with highest and lower risk. A counselor followed a standardized script *to elicit factors* influencing participant`s preference regarding LCS, as well as quantitative ratings of the strength of each factor in favor or against LCS and of the relative importance placed on each factor.<br>FPs: Nodules are "spots" in the lung that are sometimes very early lung cancer. However, most of the time they do not turn out to be cancer (sometimes referred to as benign nodules). Most often, we recommend people with nodules get another CT scan in 3 to 12 months to make sure the nodule is not growing.<br>Overdiagnosis: Treatment for a slow-growing lung cancer that would not have caused problems even if you never got treatment (sometimes called "overdiagnosis" or "overtreatment").<br><br>**Personalized risk offered in DA but NR if part of protocol; overview of differing benefits by risk level given.**<br>**Review of material during SDM.**<br><br>**Judgement of Net benefit: Moderate**<br><br>Data collection: Participants completed a standardized decision counseling protocol and completed handout on reasons to be screened or not screened (1-3 factors) and rated how important they are (0 [not important at all] -10 [very important]). LCS completion was determined using LCS Program Registry.<br><br>Other information:<br>Radiation exposure: Radiation can cause cancer. The amount of radiation from screening CTs is low, so the risk of dying from lung cancer caused by the radiation is very low (much lower than the number of people who benefit). | Factors reported to influence decision preference (n=682):<br>29% worries and risk<br>27% social influence and support<br>20% susceptibility<br>11% salience and coherence<br>11% response efficacy<br>2% other<br><br>**Data used**:<br>Uptake: 89% | |

| Study, Country, Study ID<br>Study description<br>Recruitment setting<br>Sample size; Age; Female Gender; Insured; White (other race/ethnicities); Smoking history; other LC risk factors<br>Screening history<br>History of FP or lung surgery<br>Risk of bias (ROB) | Study data e.g. communicated outcomes of screening, attributes and their levels<br>Format of information and definitions provided<br>Judgement of Net Benefit<br>Data collection<br>Other information provided | Findings reported by authors (including relevant subgroup findings) & Data used in review analysis | Knowledge and understanding (including relevant subgroup findings) |
|---|---|---|---|
| Han 2019, U.S.<br>Study #: 23<br><br>Pre-post study using surveys administered to patients before and after provision of personalized cancer risk information in SDM consultations at LCS centre<br><br>Eligibility and Recruitment: patients eligible (NLST) for screening and referred by primary care to a centralized, specialty-based LDCT screening program a centralized, specialty-based LDCT screening program<br><br>N=60; 55-80 (mean ± SD 63.2 ± 5.2); 41% Female; NR; NR; 51% current smokers; NR; 6-year predicted risk mean 4.5%<br><br>Screening history: NR<br><br>History of FP or lung surgery: NR<br><br>ROB: High risk | **Communicated Outcomes of Screening & Attribute levels:**<br>• LC mortality reduction by screening: screening can reduce a person's chances of dying from lung cancer by about 3/1000 or <1% (next 6-7 years)<br>• FPs: 25%<br>• Invasive procedures: 2%<br>• Major complication (from invasive procedure): <1%<br><br>Format & definitions: The program's prescreening SDM counseling was provided by 2 pulmonary physicians during 40-minute consultation visits, guided by a brief 1-page DA with Q&A format. Modified for this study to calculate and share the patient' 6-year cancer risk via PLCOm2012.<br>FP: "Falsely abnormal result from LDCT—that is, the LDCT will indicate they may have lung cancer or other problems, even when they are healthy"<br><br>**Personalized risk but not outcomes presented**.<br>**Review of material during SDM.**<br><br>**Judgement of Net benefit: Moderate**<br><br>Data collection: Pre/post quantitative surveys. Screening completion by medical records.<br><br>Other information:<br>FNs: 5%<br>Radiation exposure: "LDCT exposes people to a small amount of radiation that might slightly increase the risk of cancer" | **Intentions**:<br>Interest in LC screening (n=47; 5-point Likert scale: 1 = not interested, 5 = extremely interested): pre 4.37 (95% CI 4.20, 4.53); post 4.70 (4.53, 4.88); P <0.001<br><br>**Screening completion**: 100%<br><br>**Data used**:<br>Intentions: >75%<br>Uptake: 100% | Perceived lifetime LC risk (interval scale 0% to 100%, in 5% increments: pre 52.1% (45.59, 58.60); post 32.8% (26.37, 39.31); p <0.0001<br><br>From interviews, most participants expressed uncertainty about the validity of objective, model-estimated risks, which they traced to various sources. |
| Sferra 2021, U.S.<br>Study #: 24<br><br><br>A prospective, randomized study evaluating a web-based randomization tool to receive | **Communicated Outcomes of Screening & Attribute levels:**<br><br>**Option Grid:**<br>• No numerical data on LC mortality, just speaks to better treatment at earlier stage<br>• FPs: 25 in 1000 (25%) with more tests done for up to 2 years | **Screening uptake**: 100% (none decided not to screen at CT appointment)<br><br>**Data used**:<br>Uptake 100% | Overall knowledge was equivalent, with a mean correct score of 64.7% with Option Grids and 62.4% with Shouldiscreen.com (P = 0.43). The only statistically significant |

| Study, Country, Study ID<br>Study description<br>Recruitment setting<br>Sample size; Age; Female Gender; Insured; White (other race/ethnicities); Smoking history; other LC risk factors<br>Screening history<br>History of FP or lung surgery<br>Risk of bias (ROB) | Study data e.g. communicated outcomes of screening, attributes and their levels<br>Format of information and definitions provided<br>Judgement of Net Benefit<br>Data collection<br>Other information provided | Findings reported by authors (including relevant subgroup findings) & Data used in review analysis | Knowledge and understanding (including relevant subgroup findings) |
|---|---|---|---|
| either a directed SDM discussion utilizing Option Grids, found at www.optiongrid.org or the online decision aid, www.shouldiscreen.com.<br><br>Eligibility and Recruitment: screen-eligible patients visiting Temple University Hospital's LCS program for CT scan<br><br>N= 237; 55-80 (mean: 64), 51.5% Female; NR; 29.1% Caucasian (61.6% African American, 7.2% Hispanic and 1.3% Asian/-Pacific Islander); NR<br><br>Screening history: NR<br><br>History of FP or lung surgery: NR<br><br>ROB: High risk | • Overdiagnosis: some suspicious nodules may be treated even though they may never cause problems<br>• Incidental findings: 10 in 100 (10%); findings in other parts of the body that may lead to more tests and possible treatment.<br><br>**Should I Screen?:**<br><br>• **Benefits and harms based on personalized 6-year risk of cancer via modelling**:<br>• LC mortality reduction by screening (per 1000): 10% risk: 20 fewer; 6% risk: 11 fewer; 5% risk: 10 fewer; 4% risk; 8 fewer; 3% risk 6 fewer: 2% risk: 4 fewer; 1%: 2 fewer; 0.5%: 1 fewer (1000 arrays of screened vs. not screened)<br>• FPs: 365 in 1000<br>• FP invasive procedures: 18 in 1000<br>• Major complications: 3 in 1000<br>• Overdiagnosed: 1 in 10 cancers found by screening<br><br>Format & definitions: Option Grids is a brief information sheet to guide a physician–patient encounter in which patients and providers can select compare lung cancer screening options and made a decision. Shouldiscreen.com includes a web-based calculator consisting of 12 questions to calculate the risks of lung cancer. The physician navigated the patient through the website. Patients of both cohorts had an opportunity for further discussion with a physician prior to deciding if they would pursue screening.<br>ShouldIScreen:<br>FPs: Screening works by finding lung nodules, some of which are cancer, but most nodules detected on the low-dose CT scan for lung cancer screening are not cancer. False positive findings can cause anxiety. However, understanding that most lung nodules found from the CT screening do not represent cancer helps most avoid significant anxiety while they wait for follow-up testing.<br>Overdiagnosis: These overdiagnosed cases of lung cancer are ones that may not have caused harm had they not been found, and a patient may not have felt any symptoms. However, by virtue of | | difference was knowledge about potential complications from the work-up of a positive LDCT scan (Option Grids versus Shouldiscreen.com; 84.8% correct versus 63.4% correct; P = 0.02)<br><br>No correlation between education level and knowledge retention in patients who used Option Grids (r = 0.09; P = 0.61) or Shouldiscreen.com (r= 0.01; P = 0.95). |

| Study, Country, Study ID<br>Study description<br>Recruitment setting<br>Sample size; Age; Female Gender; Insured; White (other race/ethnicities); Smoking history; other LC risk factors<br>Screening history<br>History of FP or lung surgery<br>Risk of bias (ROB) | Study data e.g. communicated outcomes of screening, attributes and their levels<br>Format of information and definitions provided<br>Judgement of Net Benefit<br>Data collection<br>Other information provided | Findings reported by authors (including relevant subgroup findings) & Data used in review analysis | Knowledge and understanding (including relevant subgroup findings) |
|---|---|---|---|
| | finding this kind of cancer, a patient may receive treatment that is unnecessary, with potential for complications. Note that once a cancer is found, it is not possible to know if it had been an overdiagnosis or not.<br>Invasive procedures: Sometimes further testing for a finding on lung cancer screening leads to the need for additional testing that can include invasive procedures (biopsies or surgery). These tests are risky and can cause harm in people that end up not having had cancer. Invasive procedures can result in minor complications such as bleeding and infections. They can also result in more major complications such as a collapsed lung and even death.<br><br>**Personalized risk and outcomes presented (for 1 group)**.<br>**Review of material during SDM.**<br><br>**Judgement of Net benefit: Moderate (on average across groups)**<br><br>Data collection: For eligible patients, a single visit consisting of an SDM visit, LDCT scan and follow-up visit on the same day.<br><br>Other information:<br>Radiation dose is equal to about 6 months of radiation from your every day environment. | | |
| Pignone 2025, US<br>Study#: 25<br><br>Prospective case series of a navigator-centered LCS program for high-risk patients in a large, urban Texas Federally Qualified Healthcare system<br><br>Eligibility and Recruitment: screen-eligible (USPSTF 2013 and 2021) patients who were recent FQHC | **Communicated Outcomes of Screening & Attribute levels: (screening annually)**<br><br>• LC mortality (over 7 years): 18 in 1000 (screened) vs. 21 in 1000 (not screened) cancer deaths; 3 cancer deaths prevented<br>• FPs: 206 in 1000<br>• FP biopsies: 14 in 1000<br>• Complications from biopsies: 0-1 in 1000<br>• Overdiagnosis: 3 in 1000<br>• Anxiety and distress: Screening can cause anxiety and distress from worrying about lung cancer, especially when the CT scan finds nodules | **Intentions:** 636 (97.8%) agreed<br><br>**Screening completion:** 528 (81%) completed<br><br>No statistically significant differences in completion by age (≤ vs > 60 yrs), gender, race/ethnicity, or insurance status (commercial vs. not).<br><br>Spanish-speaking patients were more likely to complete LCS than English | None |

| Study, Country, Study ID<br>Study description<br>Recruitment setting<br>Sample size; Age; Female Gender; Insured; White (other race/ethnicities); Smoking history; other LC risk factors<br>Screening history<br>History of FP or lung surgery<br>Risk of bias (ROB) | Study data e.g. communicated outcomes of screening, attributes and their levels<br>Format of information and definitions provided<br>Judgement of Net Benefit<br>Data collection<br>Other information provided | Findings reported by authors (including relevant subgroup findings) & Data used in review analysis | Knowledge and understanding (including relevant subgroup findings) |
|---|---|---|---|
| attenders and agreeing to SDM for LCS program via mailed outreach materials (7.4% response rate among current/formers smokers and 66% of eligible enrolled) or direct provider referrals (75% response rate and 89% of sample)<br><br>N= 650; 55-77 (mean 61.7), 38% Female; 100% (insured or covered for screening and work-up; ~90% insured for treatment); 37% Caucasian (18% African American, 36% Hispanic and 2.5% other); 76% current smokers; NR<br><br>Screening history: NR<br><br>History of FP or lung surgery: NR<br><br>ROB: High | Format & definitions: One-page print DA with text definitions and absolute risks for screened and not screened. Discussed during SDM via telephone with LCS scans booked for those interested and reminders/transportation.<br>FPs: need for more scans; FP biopsies: biopsies in people who do NOT have lung cancer; Complications from biopsies: Serious complications from biopsies in people who do NOT have lung cancer; Overdiagnosis: Over-detected and treated cancers and Over-detection and over-treatment of harmless "cancers"<br><br>**No personalized risk and outcomes presented**.<br>**Review of material during SDM, mostly via telephone with navigator.**<br><br>**Judgement of Net benefit: Low**<br><br>Data collection: For eligible patients, mostly telephone SDM in English or Spanish with LDCT scans booked. Reminders were sent and transportation provided for LCS. LCS and diagnostic work-up was paid for by grant, for those uninsured. Data via bookings and records of screening.<br><br>Other information:<br>Radiation exposure: 0-1 Radiation-related cancer (10-20 years later)<br>Costs: The CT screening test is covered by your insurance or our program. Our team will work with you if you require additional tests. | speakers (90% vs. 81%; OR 2.22, 95% CI=1.22, 4.41)<br>Former smokers were more likely to complete LCS than current smokers (89% vs.81%; OR 1.93, 95%CI=1.12, 3.51).<br><br>**Data used:**<br>Intentions 98%<br>Uptake 81% | |

Table S3.6 Risk of bias, other preference data, direct and indirect comparisons

| Author, Year Outcome | 1.1 Selection of participants | **Domain 1** | 2.1 Participation rate (>50%) | 2.2 Responders similar to non-responders | 2.3 Attrition and missing data (<30%) | **Domain 2** | 3.1 Outcome valid and reliable (e.g. uptake over >6 months) | 3.2 Instrument administered in the intended way | 3.3 Valid representation/ description of outcomes | 3.4 Understanding of instrument | **Domain 3** | 4.1 Results reported and analyzed appropriately | **Domain 4** | **Overall ROB** |
|---|---|---|---|---|---|---|---|---|---|---|---|---|---|---|
| Behr (2024); Intentions to screen | N/PN | Some concerns | NI | NI | NI | Some concerns | Y/PY | Y/PY | Y/PY | Y/PY | Low | Y/PY | Low | Some concerns |
| Bittner-Fagan 2024; Screening attitudes and screening uptake | Y/PY | Low | Y/PY | NI | Y/PY | Low | Y/PY | Y/PY | Y/PY | Y/PY | Low | Y/PY | Low | Low |
| Byrne (2019); Direct data | Y/PY | Low | Y/PY | Y/PY | Y/PY | Low | Y/PY | Y/PY | NI | Y/PY | Some concerns | Y/PY | Low | Some concerns |
| Clark (2021); Intention to screen | Y/PY | Low | Y/PY | Y/PY | Y/PY | Low | Y/PY | Y/PY | Y/PY | N/PN | Some concerns | Y/PY | Low | Some concerns |
| Clark (2021); Direct data (comparisons with overdiagnosis (“reduced risk of diagnosis with lung cancer”)) | Y/PY | Low | Y/PY | Y/PY | Y/PY | Low | N/PN | Y/PY | Y/PY | N/PN | High | Y/PY | Low | High |
| Clark (2021); Direct data (comparisons limited to other outcomes) | Y/PY | Low | Y/PY | Y/PY | Y/PY | Low | Y/PY | Y/PY | Y/PY | N/PN | Some concerns | Y/PY | Low | Some concerns |
| Clark (2022); Direct data & Intention to screen | Y/PY | Low | Y/PY | Y/PY | Y/PY | Low | Y/PY | Y/PY | Y/PY | Y/PY | Low | Y/PY | Low | Low |
| Daskalakis (2023); Screening uptake | N/PN | High | Y/PY | Y/PY | Y/PY | Low | Y/PY | Y/PY | Y/PY | Y/PY | Low | Y/PY | Low | High |
| Dharod (2019); Intention to screen | Y/PY | Low | NI | NI | Y/PY | Some concerns | Y/PY | Y/PY | NI | Y/PY | Some concerns | Y/PY | Low | Some concerns |
| Dharod (2019); Screening uptake | Y/PY | Low | NI | NI | Y/PY | Some concerns | N/PN | Y/PY | NI | Y/PY | High | Y/PY | Low | High |

| Author, Year Outcome | 1.1 Selection of participants | **Domain 1** | 2.1 Participation rate (>50%) | 2.2 Responders similar to non-responders | 2.3 Attrition and missing data (<30%) | **Domain 2** | 3.1 Outcome valid and reliable (e.g. uptake over >6 months) | 3.2 Instrument administered in the intended way | 3.3 Valid representation/ description of outcomes | 3.4 Understanding of instrument | **Domain 3** | 4.1 Results reported and analyzed appropriately | **Domain 4** | **Overall ROB** |
|---|---|---|---|---|---|---|---|---|---|---|---|---|---|---|
| Djuirc 2023; Screening attitudes and relative importance of harms | N/PN | Some concerns | NI | NI | N/PN | High | Y/PY | Y/PY | Y/PY | Y/PY | Low | Y/PY | Low | High |
| Fraenkel (2016); Intention to screen | N/PN | High | Y/PY | Y/PY | Y/PY | Low | Y/PY | Y/PY | N/PN | Y/PY | Some concerns | Y/PY | Low | High |
| Fraenkel (2016); Direct data | N/PN | Some concerns | Y/PY | Y/PY | Y/PY | Low | Y/PY | Y/PY | Y/PY | Y/PY | Low | Y/PY | Low | Some concerns |
| Han (2019); Intention to screen | N/PN | High | Y/PY | Y/PY | Y/PY | Low | Y/PY | Y/PY | Y/PY | Y/PY | Low | Y/PY | Low | High |
| Han (2019); Screening uptake | N/PN | High | Y/PY | Y/PY | Y/PY | Low | Y/PY | Y/PY | Y/PY | Y/PY | Low | Y/PY | Low | High |
| Hoffman (2018) Intention to screen | N/PN | High | NI | NI | Y/PY | Some concerns | Y/PY | Y/PY | N/PY | Y/PY | Some concerns | N/PN | High | High |
| Hoffman (2018) Direct data | N/PN | High | NI | NI | Y/PY | Some concerns | Y/PY | Y/PY | Y/PY | Y/PY | Low | N/PN | High | High |
| Kathuria (2022); Screening uptake | N/PN | Some concerns | Y/PY | Y/PY | Y/PY | Low | N/PN | Y/PY | Y/PY | Y/PY | High | Y/PY | Low | High |
| Kinsinger (2017); Intention to screen | Y/PY | Low | N/PN | NI | Y/PY | Some concerns | Y/PY | Y/PY | Y/PY | Y/PY | Low | Y/PY | Low | Some concerns |
| Kinsinger (2017); Screening uptake | Y/PY | Low | N/PN | NI | Y/PY | Some concerns | N/PN | Y/PY | Y/PY | Y/PY | High | Y/PY | Low | High |
| Lau (2015); Intention/attitudes to screen | Y/PY | Low | Y/PY | Y/PY | Y/PY | Low | Y/PY | Y/PY | Y/PY | Y/PY | Low | Y/PY | Low | Low |
| Lau (2021); Intention to screen | Y/PY | Low | NI | NI | Y/PY | Some concerns | Y/PY | Y/PY | Y/PY | N/PN | Some concerns | Y/PY | Low | Some concerns |
| Lau (2021); Screening uptake | Y/PY | Low | NI | NI | N/PN | High | Y/PY | Y/PY | Y/PY | N/PN | Some concerns | Y/PY | Low | High |
| Lillie (2017); Screening uptake | Y/PY | Low | Y/PY | Y/PY | N/PN | High | N/PN | Y/PY | Y/PY | Y/PY | High | Y/PY | Low | High |
| Lillie (2017); Direct data | Y/PY | Low | Y/PY | Y/PY | N/PN | High | N/PN | Y/PY | Y/PY | Y/PY | High | Y/PY | Low | High |
| Manners (2020); Intentions to screen | Y/PY | Low | N/PN | NI | Y/PY | Some concerns | Y/PY | Y/PY | Y/PY | N/PN | Some concerns | Y/PY | Low | Some concerns |
| Mazzone (2017); Screening uptake | N/PN | High | Y/PY | Y/PY | Y/PY | Low | Y/PY | Y/PY | NI | N/PN | Some concerns | Y/PY | Low | High |
| Pignone (2025); Intentions to screen and screening uptake | N/PN | High | N/PN | N/PN | Y/PY | High | Y/PY | Y/PY | N/PN | NI | Some | Y/PY | Low | High |

| Author, Year Outcome | 1.1 Selection of participants | **Domain 1** | 2.1 Participation rate (>50%) | 2.2 Responders similar to non-responders | 2.3 Attrition and missing data (<30%) | **Domain 2** | 3.1 Outcome valid and reliable (e.g. uptake over >6 months) | 3.2 Instrument administered in the intended way | 3.3 Valid representation/ description of outcomes | 3.4 Understanding of instrument | **Domain 3** | 4.1 Results reported and analyzed appropriately | **Domain 4** | **Overall ROB** |
|---|---|---|---|---|---|---|---|---|---|---|---|---|---|---|
| Quaife (2020); Screening uptake | N/PN | High | Y/PY | Y/PY | Y/PY | Low | Y/PY | Y/PY | N/PN | N/PN | High | Y/PY | Low | High |
| Reuland (2018); Intention to screen | Y/PY | Low | Y/PY | Y/PY | Y/PY | Low | Y/PY | Y/PY | Y/PY | Y/PY | Low | N/PN | High | High |
| Reuland (2018); Screening uptake | Y/PY | Low | Y/PY | Y/PY | Y/PY | Low | N/PN | Y/PY | Y/PY | Y/PY | High | Y/PY | Low | High |
| Schapira 2023; Screening uptake | Y/PY | Low | NI | NI | Y/PY | Some concerns | Y/PY | Y/PY | Y/PY | Y/PY | Low | Y/PY | Low | Some concerns |
| Sferra (2021); Screening uptake | N/PN | High | NI | NI | Y/PY | Some concerns | Y/PY | Y/PY | N/PN | Y/PY | Some concerns | Y/PY | Low | High |
| Tanner (2019); Screening uptake | N/PN | Some concerns | N/PN | NI | Y/PY | Some concerns | NI | Y/PY | Y/PY | Y/PY | Some concerns | Y/PY | Low | Some concerns |
| Volk (2014); Screening attitudes | N/PN | Some concerns | N/PN | NI | NI | Some concerns | N/PN | Y/PY | N/PN | Y/PY | High | Y/PY | Low | High |
| Volk (2020); Intention to screen, screening uptake | N/PN | Some concerns | Y/PY | Y/PY | Y/PY | Low | Y/PY | Y/PY | N/PN | Y/PY | Some concerns | Y/PY | Low | Some concerns |
| Volk (2020); Direct data | N/PN | Some concerns | Y/PY | Y/PY | Y/PY | Low | Y/PY | Y/PY | N/PN | Y/PY | High | Y/PY | Low | High |
| Webster (2023); Screening intentions | N/PN | Some concerns | Y/PY | NI | Y/PY | Low | N/PN | Y/PY | Y/PY | Y/PY | High | Y/PY | Low | High |
| Webster (2023); Screening uptake | N/PN | Some concerns | Y/PY | NI | N/PN | High | N/PN | Y/PY | Y/PY | Y/PY | High | Y/PY | Low | High |

Notes on ROB assessments:

Domain 3.1 revised to make about outcome validity eg duration follow-up for uptake; wording of questions eg “avoiding risk for cancer “ when rating/ranking overdiagnosis; 3.3. only 1 harm presented for intention/uptake; benefits described as mortality for CT vs x-ray, “USPSTF recommends” 3.4 no piloting of questions and/or poor knowledge on test

Domain 1: Y/PY=low; NI=some concerns; N/PN some concerns with select samples or some ineligible for direct data; but high risk for screening attendees or substantial portion (>25%) are not eligible for screening (intentions/uptake data); overall High if high

Domain 2: overall Low: 2.1 and 2.3 Y/PY; some if 2.1 N/PN and 2.2 Y/PY and 2.3 Y/PY (or if NI for 2.1/2.2 and 2.3); High: 2.1 and 2.2 are N/PN OR 2.3 N/PN

Domain 3: overall: Low Y/PY for all; some if 1-2 NI or 1 N/PN; high if 3 NI or 2 NI & 1N/PN or 3.1 is N/PN

Domain 4: Y/PY=low; NI=some concerns; N/PY=high (when % undecided was NR for intentions (and may impact findings; or no SDs/variance measures)

### Additional file 4. Additional tables and figures for key question 3

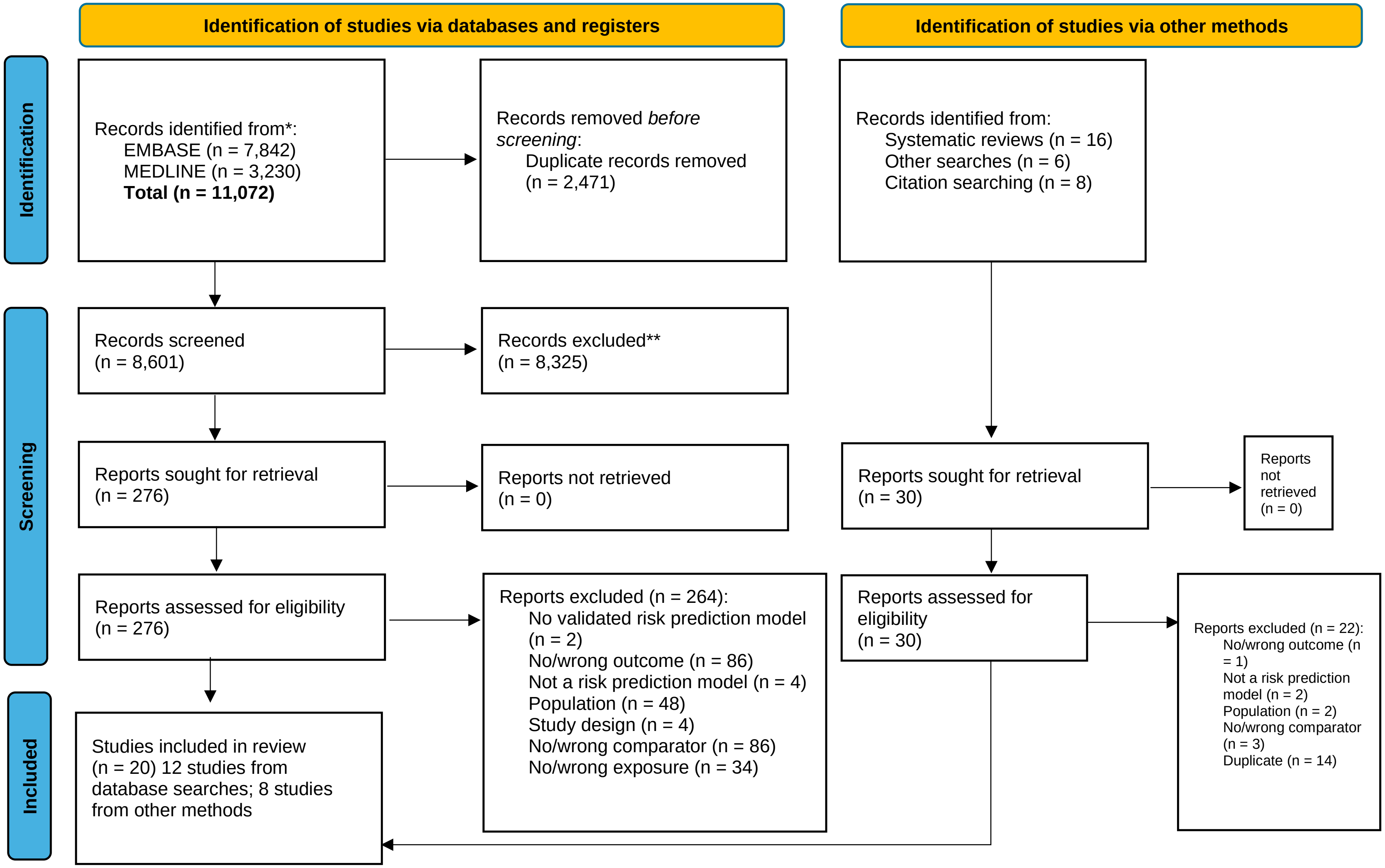

Figure S4.1 Literature flow diagram

Table S4.1 Characteristics table, NLST and LungRADs nodule management, NLST-eligible

| **Author, Year**<br>**Country (setting)** | **Inclusion criteria**<br>**Eligible for CT screening, *n***<br>**Enrolled, *n***<br>**Analyzed, *n***<br>**Age, *mean ± SD/median ± IQR (yr)***<br>**Gender, % male**<br>**Smoking status, current vs former smoker vs never smoker**<br>**Pack-years smoking history** | **Screening Strategy:**<br>**Type of CT scan**<br>**Number of rounds**<br>**Nodule management for a positive CT scan**<br>**Other relevant details**<br>**of screening process** |
|---|---|---|
| Cattaneo, 2018<br><br>USA (Anne Arundel Medical Center, 383-bed acute care community hospital) | We initiated an unfunded registration trial for LDCT lung cancer screening by adhering to the patient population, radiologic technique, and follow-up protocol used in the NLST...In 2014, after the USPSTF grade B evaluation of LDCT lung cancer screening [5], we ended the registration trial and created the pathway for other physicians to order the test on appropriate patients.<br><br>Eligible, n: 1,241<br>Enrolled, n: 1,241<br>Analyzed, n: 1,241<br><br>Age, median: 66.0<br>Gender, % male: 47.5%<br>Current smoker, %:49.1%<br>Former smoker, %: 48.2%<br>Not reported, %: 2.7%<br>Pack-years smoking history, median: 40.0 | LDCT<br>One baseline LDCT scan.<br>LungRADs version 1.0: categories 3 or 4 were positive.<br>Active smokers referred to local smoking cessation program. |
| Erkmen, 2022<br><br>USA (Temple University Hospital, Urban, safety-net health system) | Inclusion criteria: USPSTF 2013 criteria,<br>- Age 55 to 80 years<br>- Minimum 30 pack-years of smoking<br>- Currently smoking or quit within 15 years<br>- Signed informed consent<br><br>Eligible, n: 1,571<br>Enrolled, n: 1,569<br>Analyzed, n: 1,509<br><br>Age, Mean (SD): NR<br>Gender, % male: 53.0%<br>Current smoker, %: 62.8%<br>Former smoker, %: 37.2%<br>Never smoker, %: NR<br>Pack-years smoking history, median (IQR): 40 (17) | LDCT<br>One baseline LDCT scan.<br>LungRADS version 1.0: categories 3 or 4 were positive.<br>Smoking cessation counseling for active smokers. |

| **Author, Year**<br>**Country (setting)** | **Inclusion criteria**<br>**Eligible for CT screening, *n***<br>**Enrolled, *n***<br>**Analyzed, *n***<br>**Age, *mean ± SD/median ± IQR (yr)***<br>**Gender, % male**<br>**Smoking status, current vs former smoker vs never smoker**<br>**Pack-years smoking history** | **Screening Strategy:**<br>**Type of CT scan**<br>**Number of rounds**<br>**Nodule management for a positive CT scan**<br>**Other relevant details**<br>**of screening process** |
|---|---|---|
| Gohagan, 2004<br><br>USA (6 screening centers) | An individual had to be between 55 and 74 years old at the time of randomization, to be either a current cigarette smoker or a former smoker who had quit within the previous 10 years, and to have at least a 30-pack-year history of cigarette smoking.<br><br>Eligible, n: 4,828<br>Enrolled, n: 3,318 (LDCT arm: 1,660)<br>Analyzed, n: 1,586<br><br>Age, mean (SD): NR<br>Gender, % male: 58.1%<br>Current smoker, %: 57.9%<br>Former smoker, %: 42.1%<br>Never smoker, %: 0%<br>Pack-years smoking history, mean (SD): NR | LDCT<br>One baseline LDCT scan.<br>NLST criteria: ≥4mm nodules. |
| Hirsch, 2020<br><br>USA (University of Colorado Hospital LCS clinic) | We retrospectively identified all patients who underwent a lung cancer screening CT between July 1, 2014 and June 30, 2018 at University of Colorado Hospital. Subjects were identified from the clinical database used to manage the LCS program (Primordial Design, Nuance Communications, Burlington, MA).Those who meet CMS LCS criteria (age 55–77, ≥30 pack year smoking history) were eligible.<br><br>Eligible, n: 624<br>Enrolled, n: 624<br>Analyzed, n: 624<br><br>Age, mean (SD): 64.3 (5.7)<br>Gender, % male: 56.9%<br>Current smoker, %: 51.4%<br>Former smoker, %: 48.6%<br>Never smoker, %:<br>Pack-years smoking history, median (IQR):<br>From specialist referral: 45.0 (40.0-59.0)<br>From PCP referral: 40.0 (33.0-51.0) | LDCT<br>One baseline LDCT scan.<br>LungRADs (version NR, timeframe suggestive of version 1.0): category 3 or 4 were positive. |

| Author, Year<br>Country (setting) | Inclusion criteria<br>Eligible for CT screening, *n*<br>Enrolled, *n*<br>Analyzed, *n*<br>Age, *mean ± SD/median ± IQR (yr)*<br>Gender, % male<br>Smoking status, current vs former smoker vs never smoker<br>Pack-years smoking history | Screening Strategy:<br>Type of CT scan<br>Number of rounds<br>Nodule management for a positive CT scan<br>Other relevant details<br>of screening process |
|---|---|---|
| Jacobs, 2017<br><br>USA (Gundersen Health System) | Asymptomatic patients aged 55 to 77 years who had smoked within the last 15 years and had a ≥30 pack-year smoking history.<br><br>Eligible, n: NR<br>Enrolled, n: 680<br>Analyzed, n: 680<br><br>Age, median (range): 64 (55-77)<br>Gender, % male: 55.3%<br>Current smoker, %: 48.3%<br>Former smoker, %: 51.7%<br>Never smoker, %: 0%<br>Pack-years smoking history, median (IQR): 44.5 (30.0-125.0) | LDCT<br>One baseline LDCT scan.<br>LungRADs version 1.0: category 3 and 4 were positive. |
| Kaminetzky, 2019<br><br>USA (Academic multisite LCS program) | All patients who underwent LDCT as part of our academic multisite lung cancer screening program; NLST criteria<br><br>Eligible, n: 1,285<br>Enrolled, n: 1,181<br>Analyzed, n: 1,181<br><br>Age, mean (SD): 64 (16.4)<br>Gender, % male: 48%<br>Current smoker, %: 71%<br>Former smoker, %: 29%<br>Never smoker, %: 0%<br>Pack-years smoking history, median (IQR): 45 (40 to 55) | LDCT<br>One baseline LDCT scan, and yearly LDCT scans for 3 additional rounds.<br>LungRADs version 1.0: category 3 and 4 were positive. |
| Lee, 2021<br><br>South Korea (National health screening program and smoking cessation clinics) | K-LUCAS targets high-risk smokers or former smokers (who quit smoking for less than 15 years) aged 55–74 years with an at least 30-pack-year smoking history.<br><br>Eligible, n: 6,754<br>Enrolled, n: 5,692<br>Analyzed, n: 5,692<br><br>Age, range: 55-74 | LDCT<br>One baseline LDCT scan.<br>LungRADs version 1.0: category 3 and 4 were positive.<br>Smoking cessation counselling was provided. |

| Author, Year<br>Country (setting) | Inclusion criteria<br>Eligible for CT screening, *n*<br>Enrolled, *n*<br>Analyzed, *n*<br>Age, *mean ± SD/median ± IQR (yr)*<br>Gender, % male<br>Smoking status, current vs former smoker vs never smoker<br>Pack-years smoking history | Screening Strategy:<br>Type of CT scan<br>Number of rounds<br>Nodule management for a positive CT scan<br>Other relevant details<br>of screening process |
|---|---|---|
| | Gender, % male: 97.9%<br>Current smoker, %: 53.6%<br>Former smoker, %: 46.4%<br>Never smoker, %:0%<br>Pack-years smoking history:<br>30-34 PY: 33.2%<br>35-39 PY: 16.0%<br>40-44 PY: 22.5% | |
| Marshall, 2017<br><br>Australia (The Queensland Lung Cancer Screening Study, single site) | Age 60–74 years and 364 days current or previous cumulative cigarette smoking history of >30 pack years (packs per day multiplied by the number of years smoked), and former smokers must have quit smoking within the previous 15 years, with or without asbestos exposure.<br><br>Eligible, n: 256<br>Enrolled, n: 256<br>Analyzed, n: 256<br><br>Age, median: 64.5<br>Gender, % male: 66.8%<br>Current smoker, %: 47.3%<br>Former smoker, %: 52.7%<br>Never smoker, %: 0%<br>Pack-years smoking history, median (IQR): 55 (25-235) | LDCT<br>One baseline LDCT scan, and yearly LDCT scans for 2 additional rounds.<br>NLST criteria: ≥4mm nodules were positive, and scans were reanalyzed for within-study comparison according to LungRADs version 1.0: category 3 and 4 were positive. |
| Morgan, 2017<br><br>USA (Cleveland Clinic LCS Program) | Being between ages 55 and 77 years, having a 30 pack-years or greater smoking history, being a current smoker or someone who quit smoking within the past 15 years, having no symptoms or signs of lung cancer, being healthy enough to tolerate curative intent treatment of early-stage lung cancer, and not having had a CT scan of the chest within 12 months.<br><br>Eligible, n: NR<br>Enrolled, n: 320<br>Analyzed, n: 320 | LDCT<br>One baseline LDCT scan.<br>LungRADs version 1.0: category 3 and 4 were positive. |

| Author, Year<br>Country (setting) | Inclusion criteria<br>Eligible for CT screening, *n*<br>Enrolled, *n*<br>Analyzed, *n*<br>Age, *mean ± SD/median ± IQR (yr)*<br>Gender, % male<br>Smoking status, current vs former smoker vs never smoker<br>Pack-years smoking history | Screening Strategy:<br>Type of CT scan<br>Number of rounds<br>Nodule management for a positive CT scan<br>Other relevant details<br>of screening process |
|---|---|---|
| | Age, mean (SD): 64.4 (5.5)<br>Gender, % male: 61.9%<br>Current smoker, %: 48.1%<br>Former smoker, %: 51.9%<br>Never smoker, %: 0%<br>Pack-years smoking history, mean (SD): 52.1 (19.8) | |
| Percac-Lima, 2018<br><br>USA (Five community health centers affiliated with Massachusetts General Hospital) | Eligibility criteria included patients aged 55–77 years old who were identified as current smokers with ≥30 pack-years smoking history in the electronic medical record (EMR). Since the majority of our community health center patients have public insurance, we used the upper age criteria set by CMS.<br><br>Eligible, n: NR<br>Enrolled, n: 346<br>Analyzed, n: 163<br><br>Age, mean (SD): 62.3 (5.6)<br>Gender, % male: 52.5%<br>Current smoker, %: 100% (inclusion criteria)<br>Former smoker, %: 0%<br>Never smoker, %: 0%<br>Pack-years smoking history, mean (SD): NR | LDCT<br>One baseline LDCT scan.<br>LungRADs version 1.0: category 3 and 4 were positive.<br>Brief smoking cessation counselling and resources for quitting smoking was given. |
| Pinsky, 2015<br><br>USA (National Lung Cancer Screening Trial [NLST] substudy, 33 centers) | Aged 55 to 74 years, had at least a 30–pack-year history of smoking, and were current smokers or had quit within the past 15 years.<br><br>Eligible, n: 26,722<br>Enrolled, n: 26,722<br>Analyzed, n: 26,455<br><br>Age, range: 55-74<br>Gender, % male: 59.0%<br>Current smoker, %: 48.1%<br>Former smoker, %: 51.9%<br>Never smoker, %: 0%<br>Pack-years smoking history, mean (SD): NR | LDCT<br>One baseline LDCT scan, and yearly LDCT for two additional rounds.<br>NLST criteria: ≥4mm nodules, and scans were reanalyzed for within-study comparison according to LungRADs version 1.0: category 3 and 4 were positive. |

| **Author, Year**<br>**Country (setting)** | **Inclusion criteria**<br>**Eligible for CT screening, *n***<br>**Enrolled, *n***<br>**Analyzed, *n***<br>**Age, *mean ± SD/median ± IQR (yr)***<br>**Gender, % male**<br>**Smoking status, current vs former smoker vs never smoker**<br>**Pack-years smoking history** | **Screening Strategy:**<br>**Type of CT scan**<br>**Number of rounds**<br>**Nodule management for a positive CT scan**<br>**Other relevant details**<br>**of screening process** |
|---|---|---|
| Raghavan, 2020<br><br>USA (Mobile LCS unit) | Significant current or recent smoking history, age over 55 years and up to 64 years.<br><br>Eligible, n:<br>Enrolled, n:<br>Analyzed, n:<br><br>Age, mean (range): 61 (55-64)<br>Gender, % male: 52%<br>Current smoker, %: NR<br>Former smoker, %: NR<br>Never smoker, %: 0% (inclusion criteria)<br>Pack-years smoking history, mean (range): 46.1 (30-220) | LDCT<br>One baseline LDCT scan.<br>LungRADs version 1.0: category 3 and 4 were positive. |
| Rendle, 2024<br><br>USA (PROSPR [Population-based Research to Optimize the Screening Process] Lung Consortium; 5 health care systems) | Patients aged 55 to 80 years who completed at least 1 LDCT scan for LCS (LCS-LDCT) between 1 January 2014 and 30 September 2018 (“screening cohort”)<br><br>Eligible, n: 9,266<br>Enrolled, n: 9,266<br>Analyzed, n: 9,266<br><br>Age, mean (SD): NR (6.6% age 74-80)<br>Gender, % male: 53.3%<br>Current smoker, %: 55.2%<br>Former smoker, %: 44.8%<br>Never smoker, %: 0%<br>Pack-years smoking history, mean (SD): NR (9.7% <30 pack-years) | LDCT<br>One baseline LDCT scan.<br>LungRADs version 1.1: category 3 and 4 were positive. |
| Silva, 2021<br><br>USA (Multicentric Italian Lung Detection trial [MILD] substudy) | We extracted a higher-risk population from the MILD study by applying the NLST eligibility criteria to the intervention arm: age ≥ 55 years and pack-years ≥ 30.<br><br>Eligible, n: 1,248<br>Enrolled, n: 1,248<br>Analyzed, n: 1,248<br><br>Age, mean (SD): NR | LDCT<br>One baseline LDCT scan.<br>LungRADs version 1.1: category 3 and 4 were positive. |

| Author, Year<br>Country (setting) | Inclusion criteria<br>Eligible for CT screening, *n*<br>Enrolled, *n*<br>Analyzed, *n*<br>Age, *mean ± SD/median ± IQR (yr)*<br>Gender, % male<br>Smoking status, current vs former smoker vs never smoker<br>Pack-years smoking history | Screening Strategy:<br>Type of CT scan<br>Number of rounds<br>Nodule management for a positive CT scan<br>Other relevant details<br>of screening process |
|---|---|---|
| | Gender, % male: 72%<br>Current smoker, %: 66.3%<br>Former smoker, %: 33.7%<br>Never smoker, %: 0%<br>Pack-years smoking history, mean (SD): NR | |
| Sylvestri, 2023<br><br>USA (3,625 facilities within the ACR LCS registry) | People who were eligible to receive screening according to the 2013 USPSTF recommendations (aged 55-80 years, current tobacco use with ≥30 pack-years of smoking or prior tobacco use with ≥30 pack-years of smoking and quit within the past 15 years).<br><br>Eligible, n: 1,052,591<br>Enrolled, n: 1,052,591<br>Analyzed, n: 1,052,591<br><br>Age, mean (SD): NR (6.5% 75-80)<br>Gender, % male: 51.7%<br>Current smoker, %: 61.4%<br>Former smoker, %: 38.6%<br>Never smoker, %: 0%<br>Pack-years smoking history, mean (SD): NR | LDCT<br>One baseline LDCT scan, and yearly LDCT for one additional round.<br>LungRADs (version NR, timeframe suggestive of version 1.0): category 3 and 4 were positive. |

Abbreviations: ACR=American College of Radiology; IQR=interquartile range; LCS=lung cancer screening; LDCT=low dose computed tomography; NR=not reported; SD=standard deviation.

Table S4.2 Risk of bias, case series screening arm by NLST eligibility and LungRADs or NLST nodule management criteria

| Author Year Outcome (Screening rounds) | Q1. Clear inclusion criteria? | Q2. Exposure (screening method) measured in standard, reliable way? | Q3. Valid methods for identification of exposure (screening method)? | Q4. Prospective: attempt to enroll all eligible? Retrospective: enroll more than just attenders to screening? | Q5a. Complete inclusion of eligible participants? | Q5b. Acceptable attrition rate for outcome being assessed? | Q6. Clear reporting of demographics? | Q7a. Outcome clearly defined and appropriate? | Q7b. Structured method of identifying outcomes? | Q7c. Follow-up duration adequate? | Overall ROB |
|---|---|---|---|---|---|---|---|---|---|---|---|
| **LungRADs nodule management criteria** | | | | | | | | | | | |
| Cattaneo 2018 **Any FPs** (round 1) | Y | Y | Y | N | Y | N | U | Y | N | Y | High |
| Erkmen 2022 **Any FPs** (round 1) | Y | Y | Y | Y | Y | Y | Y | Y | Y | Y | Low |
| Hirsch 2020 **Any FPs** (round 1) | Y | Y | Y | N | Y | U | U | Y | U | U | Some concerns |
| Jacobs 2017 **Any FPs** (round 1) | Y | Y | Y | N | Y | N | U | Y | U | N | High |
| Kaminetzky 2019 **Any FPs** (round 1) | Y | Y | Y | Y | Y | U | Y | Y | U | Y | Some concerns |
| Lee 2021 **Any FPs** (round 1) | Y | Y | Y | Y | Y | N | Y | Y | Y | Y | High |
| Marshall 2017 **Any FPs** (round 1) | Y | Y | Y | N | Y | Y | Y | Y | U | Y | Some concerns |
| Morgan 2017 **Any FPs** (round 1) | Y | Y | Y | N | U | U | Y | Y | U | Y | Some concerns |
| NLST (Pinsky 2015) **Any FPs** (round 1) | Y | Y | Y | N | Y | Y | Y | Y | U | Y | Some concerns |
| Percac-Lima 2018 **Any FPs** (round 1) | Y | Y | Y | Y | U | N | U | Y | U | U | High |
| Raghaven 2020 **Any FPs** | Y | Y | Y | U | U | U | U | Y | Y | U | Some concerns |

| | | | | | | | | | | | |
|---|---|---|---|---|---|---|---|---|---|---|---|
| (round 1) | | | | | | | | | | | |
| Rendle 2024 **Any FPs** (round 1) | Y | Y | Y | N | Y | N | Y | Y | U | U | High |
| Silva 2021 **Any FPs** (round 1) | Y | Y | Y | N | Y | Y | U | Y | U | Y | Some concerns |
| Silvestri 2023 **Any FPs** (round 1) | Y | Y | Y | N | Y | U | U | Y | U | Y | Some concerns |
| **NLST nodule management criteria** | | | | | | | | | | | |
| LSS (Gohagan 2004) **Any FPs** (round 1) | Y | Y | Y | Y | U | Y | U | Y | N | Y | High |
| Marshall 2017 **Any FPs** (round 1) | Y | Y | Y | N | Y | Y | Y | Y | U | Y | Some concerns |
| NLST (Pinsky 2015) **Any FPs** (round 1) | Y | Y | Y | N | Y | Y | Y | Y | N | Y | High |

Notes on ROB:

- Overall ROB was rated as: low if all domains were rated as Y; some concern if any domains were rated as U; high if at least one of the main domains was rated as N (5b, 7a, and 7b).
- Q5a was rated as Y if ≥50% of eligible participants were enrolled.
- Q5b was rated as Y if ≥90% of those who were recalled (for FP) or referred for additional diagnostic workup attended the subsequent follow-up.
- Q6 was rated as: Y if age, sex, smoking, and at least one comorbidity was reported, U if three of the four variables were reported, and N if less than three of variables were reported.

| Study or Subgroup | Proportion | SE | Weight | Proportion IV, Random, 95% CI |
|---|---|---|---|---|
| LSS (Gohagan 2004) | 0.178 | 0.00938 | 34.8% | 0.18 [0.16, 0.20] |
| Marshall 2017 | 0.48 | 0.0312 | 29.9% | 0.48 [0.42, 0.54] |
| NLST (Pinsky 2015) | 0.26 | 0.00268 | 35.3% | 0.26 [0.25, 0.27] |
| **Total (95% CI)** | | | **100.0%** | **0.30 [0.21, 0.38]** |

Heterogeneity: $Tau^2 = 0.01$; $Chi^2 = 122.86$, df = 2 ($P < 0.00001$); $I^2 = 98\%$
Test for overall effect: Z = 6.88 ($P < 0.00001$)

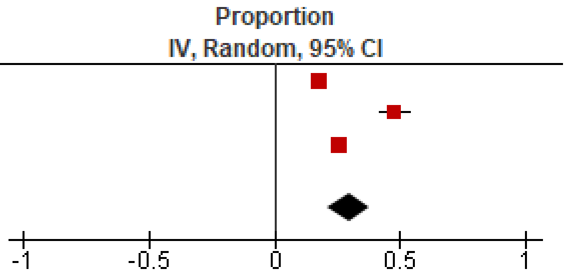

Figure S4.2 Round 1: Proportion of false positives, NLST eligibility and nodule management

| Study or Subgroup | LungRADs Events | LungRADs Total | NLST Events | NLST Total | Weight | Risk Ratio M-H, Random, 95% CI |
|---|---|---|---|---|---|---|
| Marshall 2017 | 51 | 256 | 123 | 256 | 6.6% | 0.41 [0.31, 0.55] |
| NLST (Pinsky 2015) | 3343 | 26722 | 6939 | 26722 | 93.4% | 0.48 [0.46, 0.50] |
| **Total (95% CI)** | | **26978** | | **26978** | **100.0%** | **0.48 [0.44, 0.51]** |
| Total events | 3394 | | 7062 | | | |

Heterogeneity: $Tau^2 = 0.00$; $Chi^2 = 1.11$, df = 1 ($P = 0.29$); $I^2 = 10\%$
Test for overall effect: Z = 19.88 ($P < 0.00001$)

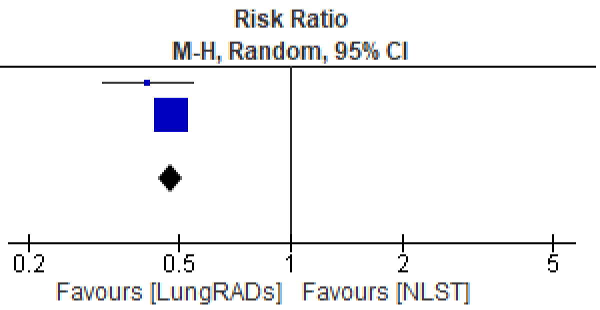

Figure S4.3 Within study comparison, Round 1: Proportion with a false positive, LungRADs vs NLST nodule management criteria

| Study or Subgroup | Proportion | SE | Weight | Proportion IV, Random, 95% CI |
|---|---|---|---|---|
| Cattaneo 2018 | 0.193 | 0.0112 | 7.2% | 0.19 [0.17, 0.21] |
| Erkmen 2022 | 0.0886 | 0.00717 | 7.8% | 0.09 [0.07, 0.10] |
| Hirsch 2020 | 0.107 | 0.0124 | 7.0% | 0.11 [0.08, 0.13] |
| Jacobs 2017 | 0.172 | 0.0145 | 6.7% | 0.17 [0.14, 0.20] |
| Kaminetzky 2019 | 0.106 | 0.00895 | 7.6% | 0.11 [0.09, 0.12] |
| Lee 2021 | 0.145 | 0.00467 | 8.0% | 0.14 [0.14, 0.15] |
| Marshall 2017 | 0.199 | 0.025 | 4.9% | 0.20 [0.15, 0.25] |
| Morgan 2017 | 0.166 | 0.0208 | 5.6% | 0.17 [0.13, 0.21] |
| NLST (Pinsky 2015) | 0.125 | 0.00202 | 8.2% | 0.13 [0.12, 0.13] |
| Percac-Lima 2018 | 0.0551 | 0.0123 | 7.0% | 0.06 [0.03, 0.08] |
| Raghavan 2020 | 0.16 | 0.0156 | 6.5% | 0.16 [0.13, 0.19] |
| Rendle 2024 | 0.144 | 0.00364 | 8.1% | 0.14 [0.14, 0.15] |
| Silva 2021 | 0.183 | 0.0109 | 7.3% | 0.18 [0.16, 0.20] |
| Silvestri 2023 | 0.168 | 0.000364 | 8.2% | 0.17 [0.17, 0.17] |
| **Total (95% CI)** | | | **100.0%** | **0.14 [0.12, 0.16]** |

Heterogeneity: Tau² = 0.00; Chi² = 774.59, df = 13 (P < 0.00001); I² = 98%
Test for overall effect: Z = 16.48 (P < 0.00001)

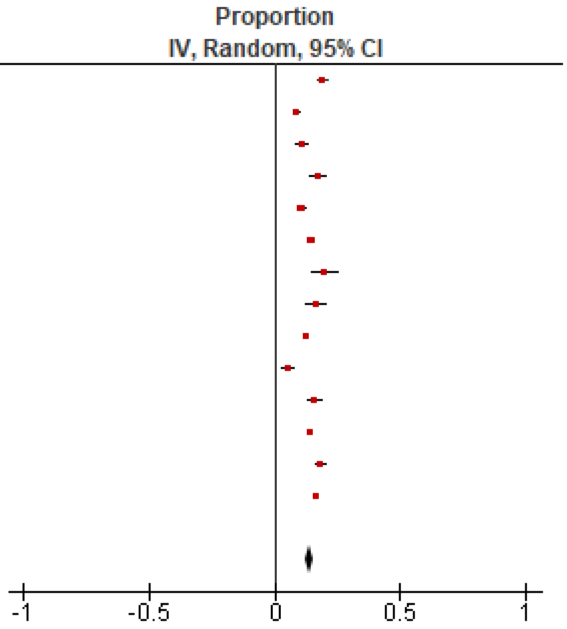

Figure S4.4. Round 1: Proportion of false positives, NLST eligibility and LungRADs nodule management
